# Sedentary behaviour and cancer risk: a World Cancer Research Fund International Global Cancer Update Programme (CUP Global) systematic literature review and meta-analysis

**DOI:** 10.64898/2026.06.23.26355971

**Authors:** Georgios Markozannes, Ahmad Jayedi, Margarita Cariolou, Eirini Pagkalidou, Sayada Zartasha Kazmi, Katia Balducci, Sonia Kiss, Rita Vieira, Sofia Cividini, Dagfinn Aune, Darren C. Greenwood, Amanda J. Cross, Marc J. Gunter, Shalini Jayasekar Zürn, Christian C. Abnet, Vanessa L.Z. Gordon-Dseagu, Kristy Maskell, Christelle Clary, Helen Croker, Panagiota Mitrou, Elio Riboli, Monica Baskin, Rajiv Chowdhury, Mia Gaudet, Edward Giovannucci, Ellen Kampman, Sarah J. Lewis, Anne M. May, Yikyung Park, Tobias Pischon, Gianluca Severi, Lynette Hill, Matty P. Weijenberg, John Krebs, Konstantinos K. Tsilidis, Doris S.M. Chan

## Abstract

**Background:** High levels of sedentary behaviour are an emerging global public health concern, but its impact on cancer risk remains unclear.

**Methods:** Within the Global Cancer Update Programme (CUP Global), we systematically searched the literature in PubMed and Embase until September 2024 for observational cohort studies on sedentary behaviour and adult cancer risk. Using dose-response meta-analyses, we investigated sedentary behaviour domains (total, occupational, recreational, transportation, and/or other) and dimensions (duration, frequency), and additionally pooled across domains. The quality of evidence was graded by the CUP Global Expert Panel (protocol registration: https://osf.io/7utbm/).

**Findings:** We identified 62 publications from 27 cohorts comprising 162,902 incident cancer cases across 19 anatomical sites. There was evidence for a probable causal positive association between sedentary time (mixed definitions) and breast (RR_per 2 hours/day_=1.03; 95%CI=1.02–1.05; I^2^=10%; n=11 studies) and colon (RR=1.05; 95%CI=1.03–1.07; I^2^=0%; n=9) cancer risk, and between television watching time and colon cancer risk (RR_per 2 hours/day_=1.08; 95%CI=1.05–1.11; I^2^=0%; n=6). Limited suggestive evidence supported positive associations between sedentary time (mixed definitions) and lung (RR=1.04; 95%CI=1.00–1.09; I^2^=69%; n=7), ovarian (RR=1.06; 95%CI=1.01–1.10; I^2^=0%; n=7), premenopausal (RR=1.03; 95%CI=0.99–1.08; I^2^=20%; n=7) and postmenopausal breast (RR=1.02; 95%CI=1.00–1.04; I^2^=7%; n=11), and colorectal (RR=1.02; 95%CI=1.00–1.04; I^2^=47%; n=11) cancers, and between occupational sitting time and breast (RR_per 2 hours/day_=1.05; 95%CI=1.01–1.09; I^2^=0%; n=4) and colon (RR=1.08; 95%CI=1.02–1.15; I^2^=28%; n=2) cancers. An interactive evidence platform is available at: https://teacup.cc.ic.ac.uk/sedentary-behaviour-cancer.html.

**Interpretation:** Evidence supports that prolonged sedentary behaviour is probably a cause of breast and colon cancers, while limited suggestive evidence supports positive associations for several other exposure–cancer pairs, including lung and ovarian cancers. This evidence should lead to revised cancer prevention recommendations. Future research should focus on device-based exposure assessments, repeated measurements, exposure substitution models, inclusion of diverse populations and investigation of biological mechanisms.

**Funding:** World Cancer Research Fund network of charities (American Institute for Cancer Research; World Cancer Research Fund; Wereld Kanker Onderzoek Fonds).

## Introduction

Prolonged sedentary behaviour (over 2 hours continuously) has become increasingly prevalent worldwide, with notable increases observed over the past few decades,^1–3^ possibly reflecting shifts in occupational, recreational, and transportation habits. The worldwide average sitting time is estimated (through self-report) as 4.7 [interquartile range, IQR: 3.5–5.1] hours daily, varying by economic development (2.7 [IQR: 2.6–3.3] hours in low-income countries to 4.9 [IQR: 4.7–5.3] hours in high-income ones).^4^ Sedentary behaviour is characterised as any activity resulting in an energy expenditure of less than or equal to 1.5 metabolic equivalents of task (METs) while in a sitting, reclining, or lying posture.^5,6^ Importantly, sedentary behaviour is distinct from, and not simply the opposite of, physical activity.^5,6^ A person may spend prolonged periods sedentary while also meeting or exceeding the physical activity guidelines. Sedentary behaviour has an adverse impact on several health outcomes, including obesity, type 2 diabetes, cardiovascular disease, and all-cause mortality.^7–10^ In contrast, studies suggest that even small reductions in sedentary behaviour through replacement with physical activity, may reduce the risk of all-cause mortality.^11,12^

While international guidelines now recommend limiting sedentary time to improve health outcomes,^13,14^ the recommendations remain general (“limit the amount of time spent being sedentary” and “replacing sedentary time with physical activity of any intensity provides health benefits”). The lack of clearly defined behavioural targets, compounded by modern work environments and entertainment options, has contributed to increasingly persistent sedentary lifestyles worldwide.^1–3^ The World Health Organization (WHO) recommends “To help reduce the detrimental effects of high levels of sedentary behaviour on health, adults should aim to do more than the recommended levels of moderate-to-vigorous physical activity”. Still, the WHO physical activity guideline development group has highlighted major gaps in understanding the effect of sedentary behaviour in human health including dose–response relationships, domain-specific effects, and exposure measurement, underscoring the need for more refined epidemiological evidence.^15^

The role of physical activity on cancer aetiology has been widely recognised and incorporated in cancer prevention guidelines,^16,17^ yet the role of sedentary behaviour on cancer risk is still elusive. Previous meta-analyses of cohort studies have suggested positive associations between sedentary behaviours and the risk of cancer at different sites such as breast and colon.^18^ However, associations with other cancer sites and with specific domains of behaviour still remain elusive. The underlying biological mechanisms remain poorly understood but may involve chronic inflammation and metabolic dysregulation.^19^ The Physical Activity Guidelines Advisory Committee (PAGAC) has concluded that there is moderate evidence for a positive association between sedentary behaviour and cancer risk.^16^ The World Cancer Research Fund/American Institute for Cancer Research (WCRF/AICR) Third Expert Report in 2018 concluded that there was limited but suggestive evidence that sedentary behaviour (mixed definitions) increases the risk for endometrial cancer and limited no conclusion for other investigated cancers.^17^ Since the publication of the Third Expert Report, new evidence from observational studies has emerged.

To address these gaps, the WCRF International Global Cancer Update Programme (CUP Global) prioritised sedentary behaviour^20^ for an updated systematic literature review (SLR) to complement the conclusions of the Third Expert Report. This work presents the results of a SLR of observational studies examining the associations between different domains, dimensions, and types of sedentary behaviours and the risks of site-specific cancer incidence and mortality in adults. This SLR, alongside a companion review on sugary drinks, juices, and cancer risk,(Jayedi et al, unpublished) contributes to a series of CUP Global reviews examining lifestyle-related exposures and cancer risk that will inform the Fourth Expert Report which will be developed by WCRF International in the coming years.

## Methods

This SLR has been conducted in accordance with a pre-registered CUP Global standard protocol (https://osf.io/7utbm/) and is reported in alignment with the Preferred Reporting Items for Systematic Reviews and Meta-Analyses (PRISMA) checklist^21^ (**Supplementary Table 1**).

### Search strategy and selection criteria

We searched PubMed and Embase until 01 September 2024 using a pre-specified standard CUP Global strategy (**Supplementary Text 1**), supplemented by reference screening. Eligible studies included randomised controlled trials (RCTs), cohort studies, or pooled analyses of such studies reporting multivariable-adjusted risk estimates for cancer incidence or mortality in adulthood (**Supplementary Text 2, Supplementary Table 2**). We also included Mendelian randomisation (MR) studies investigating associations of genetically predicted sedentary behaviour and cancer risk **(Supplementary Text 2).**

We categorised sedentary behaviour by domain (total, occupational, recreational, transportational, other) and dimension (duration, frequency), and by specific comparable definitions (sedentary, sitting, computer screening, television watching time). Additional analyses were conducted by pooling studies across domains to reflect overall sedentary behaviour. When multiple estimates were reported in a study, we selected a single estimate for inclusion in the mixed-definitions analyses (in order: total, recreational, non-recreational, occupational, transportational, other sitting time). When possible, we used the maximally-adjusted estimates that were not adjusted for body mass index (BMI) or other adiposity measures, as adiposity was considered a potential mediator in the association between sedentary behaviour and cancer risk.^22,23^ When studies reported results by sub-populations or cancer subtypes, we pooled results to an overall estimate using established methods^24^ (**Supplementary Text 2**).

Risk of bias was assessed using a modified version of the Risk of Bias for Nutrition Observational Studies (RoB-NObs) tool,^25^ originally developed by the U.S Department of Agriculture (USDA) Nutrition Evidence Systematic Review based on a modification of the Cochrane’s Risk of Bias In Non-randomised Studies of Interventions (ROBINS-I) tool.^26^ The CUP Global team further adapted the tool focusing on lifestyle-cancer associations. The tool assesses bias across seven domains: confounding, participant selection, exposure classification, departures from intended exposures, missing data, outcome measurement, and selective reporting (**Supplementary Tables 3, 4**). Article selection, data extraction, and risk of bias assessment were double checked in at least 10% of the records by a second reviewer. The review coordinator resolved any disagreements. All data were extracted into the CUP Global database.

### Data analysis

Meta-analyses were conducted separately for incidence (primary) and mortality (secondary) outcomes. For cancers with high mortality rate (pancreas, liver, lung, gallbladder), we further combined incidence and mortality outcomes. Our primary analyses were linear dose-response meta-analyses when at least two studies provided sufficient information (**Supplementary Text 2**). When dose-response meta-analyses were not possible or when more than a third of the included studies did not have sufficient information for inclusion, categorical high versus low analyses were conducted. When meta-analyses were not possible, the individual studies were descriptively reviewed. DerSimonian-Laird random effects models ^27^ were used for categorical and linear dose-response meta-analyses. Non-linear dose-response meta-analyses were conducted by one-stage mixed effects models^28,29^ when at least five primary studies provided sufficient information (**Supplementary Text 2**). Two hours/day of sedentary time was selected as the increment unit for the linear meta-analyses and as the referent point of exposure for the non-linear meta-analyses, consistent with the previous CUP Global SLRs.

The proportion of total variability in effect estimates attributable to heterogeneity between studies was assessed by the I^2^ metric.^30^ Potential sources of heterogeneity were explored by pre-specified subgroup analyses including participant and study characteristics when at least two primary studies reported results for the same subgroup. The impact of individual studies on the summary estimate was assessed in leave-one-out analyses.^31^ The low number of studies per meta-analysis (less than 10) precluded the investigation of small study effects by Egger’s regression test^32^ or by visual inspection of the funnel plots. Analyses were conducted in Stata version 16.1.

### Evidence grading criteria

An Expert Panel convened by WCRF International interpreted and graded the quality of evidence as *convincing, probable, limited suggestive, limited no conclusion,* or *substantial effect on risk unlikely*, following pre-defined evidence grading criteria designed to evaluate the likelihood of causality (**Supplementary Table 5**) based on the volume and type, precision, consistency and quality of the evidence. Assessment of biological plausibility in humans is formally undertaken within CUP Global using a structured framework,(REF) and it has the potential to strengthen evidence judgements.

However, because challenges were anticipated in disentangling biological mechanisms specific to sedentary behaviours from those related to other exposures such as physical activity, exploration of biological mechanisms was not prioritised as part of this work.

## Results

The search identified 14,470 records, of which 136 were potentially eligible for full text review (**Figure 1**). Of those, 78 were excluded based on reasons presented in **Supplementary Table 6**. We identified another 4 publications from the CUP Global database. In total, 62 publications from 27 observational cohort studies were included in the systematic review^33–94^ (**Supplementary Table 7**). Twelve studies (44%) (31 publications) were from the North America,^36–41,44,46,48–53,55–61,63,69,71,73,75,80,84,85,87,89^ 8 studies (30%) (13 publications) were from Southeast/East Asia,^42,47,54,62,64,65,74,76,77,79,81,86,91^ 6 studies (22%) (17 publications) were from Europe,^33–35,43,45,67,68,70,72,78,82,83,88,90,92–94^ and one was from Australia.^66^ All studies investigated cancer incidence across at least one of 19 major anatomical sites comprising 162,902 cases. Four studies (6 publications)^50,53,54,59,69,81^ investigated cancer mortality across 5 major anatomical sites comprising 5,985 cancer deaths (**Supplementary Tables 8-10**). A detailed list of exposure-cancer pairs assessed in the 27 cohort studies is presented in (**excel file: Sedentary_Behaviour_Cohorts**). Except for one study that assessed accelerometer-based sedentary activity,^88^ all studies reported results on different self-reported domains, dimensions, or types of sedentary behaviour. We additionally identified and included 15 publications of MR studies. No RCT was identified.

**Figure 1:**
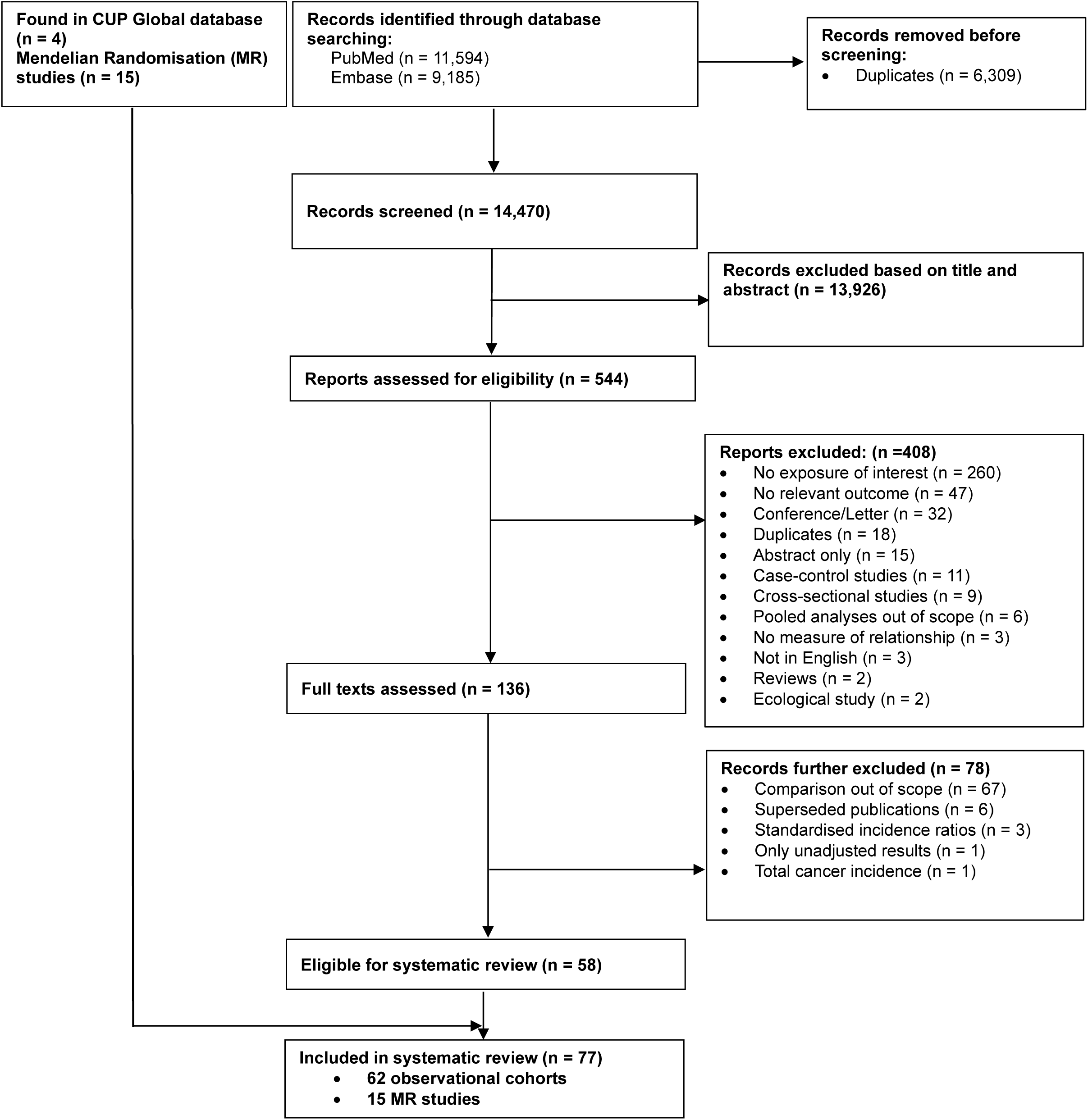
Flowchart of study selection process.

Summaries of all meta-analysis results for the associations investigated across various exposure groups, including results for cancer subtypes and subsites and cancer mortality outcomes, are presented in **Supplementary Figures 1-7**. Supporting results of the categorical highest versus lowest comparisons are presented in **Supplementary Figures 8-20**. Detailed results of the meta-analyses and descriptive syntheses are presented in **Supplementary Text 3** and **Supplementary Tables 11-14**. A brief overview of the main results is provided below.

For sitting time, we identified 21 studies (41 publications)^33,34,36,38–46,48,49,51–53,55–63,65,66,70–73,75–77,84–86,89,90,94^ with 119,737 cases of cancer incidence at 19 anatomical sites and 15,899 cases of cancer mortality at two anatomical sites **(Supplementary Tables 7, 9)**. Meta-analyses were possible for total and occupational sitting **(Supplementary Figures 21-51)**. Total sitting time was linearly associated with higher risk of breast (RR_per 2 hours/day_ 1.04, 95%CI 1.01-1.07, I^2^ 0%, 4 studies, 3,658 cases)^46,55,60,85^ **(Supplementary Figure 23)**, postmenopausal breast (RR 1.03, 95%CI 1.00-1.05, I^2^ 0%, 4 studies, 8,627 cases)^39,55,60,63,85^ **(Supplementary Figure 32)** and colon cancer incidence (RR 1.03, 95%CI 1.01-1.06, I^2^ 0%, 3 studies, 2,396 cases)^36,61,66^ **(Supplementary Figure 39)**. For prostate cancer, only a high versus low meta-analysis was possible without evidence of an association **(Supplementary Figure 25)**. Occupational sitting time was positively linearly associated with the risk of breast (RR 1.05, 95%CI 1.01-1.09, I^2^ 0%, 4 studies, 3,334 cases)^42,46,60,77^ **(Supplementary Figure 28)**, ovarian (RR 1.25, 95%CI 1.12-1.40, I^2^ 3.7%, 3 studies [including one pooled analysis of 2 cohorts], 2 publications, 908 cases)^77,84^ **(Supplementary Figure 29)**, and colon cancer incidence (RR 1.08, 95%CI 1.02-1.15, I^2^ 28.2%, 2 studies, 1,554 cases)^45,77^ **(Supplementary Figure 40)**. The results of categorical high versus low analyses for breast and colorectal cancers supported the linear analysis findings **(Supplementary Figures 31, 38)**. There was little evidence of heterogeneity across subgroups **(Supplementary Table 13; Supplementary Figures 32-37, 41-45)**. In general, individual studies did not influence the summary estimates **(Supplementary Figures 46-51)**.

No meta-analyses were conducted for total and non-occupational sedentary time (2 studies, 4 publications, 3 different anatomical sites)^83,88,91,92^ or for recreational (one study, 5 publications),^44,51,56,57,89^ transportational (3 studies, each on different cancer),^46,58,90^ or other sitting time (5 studies reporting incomparable definitions)^46,62,71,77,84^. The only study that investigated objectively measured sedentary time and non-melanoma skin cancer in UK Biobank indicated an inverse association, but the 95%CI crossed the null (RR_Per 20 mins/day_: 0.99; 95%CI: 0.97-1.00; 1,734 cases). A substitution model where sedentary time was replaced with equivalent time of light or moderate-to-vigorous physical activity indicated no association^88^.

For screen time, we identified 14 studies (33 publications)^35–39,41,46–50,52–55,58,60,62,64,67–69,71,73,74,78–82,84,87,93^ with 60,171 cases of cancer incidence at 16 anatomical sites and 5,171 cases of cancer mortality at 5 anatomical sites. Definitions included recreational screen time,^68,93^ recreational computer screen time^67^, TV watching time,^35–39,41,46–50,52–55,58,60,62,64,67,69,71,73,74,78,79,81,82,84,87^ and TV watching frequency^80^ **(Supplementary Tables 7, 10)**. Meta-analyses were only possible for TV watching time **(Supplementary Tables 7, 14; Supplementary Figures 52-95)**. TV watching time was positively associated with colorectal (RR_per 2 hours/day_ 1.03, 95%CI 1.01-1.06, I^2^ 0%, 6 studies, 5 publications, 11,619 cases)^58,62,71,73,78^ **(Supplementary Figure 54)**, colon (RR 1.08, 95%CI 1.05-1.11, I^2^ 0%, 5 studies, 4 publications, 5,553 cases)^36,58,62,78^ **(Supplementary Figure 65)**, and lung (RR 1.05, 95%CI 1.01-1.10, I^2^ 0%, 3 studies, 3,079 cases)^47,48,78^ cancer incidence **(Supplementary Figure 55)**. Examination of non-linearity was possible in meta-analyses for colorectal, colon, and breast cancers, without evidence of non-linearity **(Supplementary Figures 61-63)**. For bladder cancer, only a categorical high versus low analysis was possible, without evidence of an association **(Supplementary Figure 64)**. The results of the subgroup and sensitivity analyses were consistent with those of the main analyses **(Supplementary Table 14; Supplementary Figures 65-87)**. There was little evidence for between-subgroup heterogeneity. In general, the results remained consistent in leave-one-out analyses **(Supplementary Figures 88-92)**. No association was observed with colorectal, prostate, and breast cancer mortality **(Supplementary Figures 93-95)**.

Additional meta-analyses on mixed definitions of sedentary time were largely consistent with the main analyses on total and occupational sitting time **(Supplementary Figures 96-111)**. Positive associations were observed for colon (RR_per 2 hours/day_ 1.05, 95%CI 1.03-1.07, I^2^ 0%, 9 studies, 8,834 cases^36,45,58,61,62,66,77,78^) **(Supplementary Figure 99)** and breast cancers (RR 1.03, 95%CI 1.02-1.05, I^2^ 9.8%, 11 studies, 17,433 cases^42,46,55,56,60,74,77,82,85,91,93^) **(Supplementary Figure 104)**. Meta-analyses were possible for liver, advanced prostate, and bladder cancer incidence using mixed definitions of sedentary time, for which meta-analyses on specific sedentary domains/definitions were not possible; all these associations were largely null. Finally, a linear dose-response meta-analysis on non-occupational sedentary time and melanoma without a main analysis on specific types of sitting time was further included with no association observed. We did not identify any uniquely or unduly influential studies in the leave-one-out sensitivity analysis **(Supplementary Figures 112-121)**.

Risk of bias assessments are summarised in **Supplementary Figure 122** and detailed in **Supplementary Text 4**. Most publications were at serious (43%) or critical (46%) risk of bias due to confounding. Those at critical risk did not adjust for important confounders such as age, sex, smoking (for smoking-related cancers), alcohol, physical activity, and reproductive factors (for reproductive cancers) **(Supplementary Table 4)**, while those at serious risk often relied on self-reported confounder assessments without further validation. Although 83% of publications adjusted for adiposity (a potential mediator of the investigated associations), this was not considered in the risk of bias assessment. Estimates were similar across models but with slight attenuation after adjusting for adiposity. For TV watching time, we further assessed adjustment for education and dietary habits. Only three studies (four examined associations; lung,^47^ ovarian,^84^ endometrial cancer incidence in main and sensitivity analyses^38^) lacked adjustments for either education or diet. Exposure assessment was primarily self-reported, and around half of publications did not report validity or reliability metrics, resulting in 57% being at low/moderate, 41% at serious, and 2% at critical risk of bias.

The systematic review of MR studies identified 15 publications ^95–109^ **(Supplementary Tables 15-17)** reporting results on 191 MR analyses of sedentary time with cancer risk. Genetically predicted self-reported TV watching time showed positive association with lung cancer, aligning with the observational findings. Results for other exposure definitions, including measured sedentary time, and cancer sites were mixed or largely null. Detailed results are presented in **Supplementary Text 5**.

A summary of the associations that received evidence grading is presented in **Figure 2** and a summary of the evidence grading agreed by the Expert Panel is presented in the **Panel: grading the evidence**. There was evidence for a probable causal positive association between sedentary time (mixed definitions) and breast and colon cancers, and between television watching time and colon cancer. This reflects the large number of prospective studies and cancer cases and the consistency of positive findings with little between-study heterogeneity, also supported by the broadly consistent findings across specific sedentary behaviour definitions.

**Figure 2:**
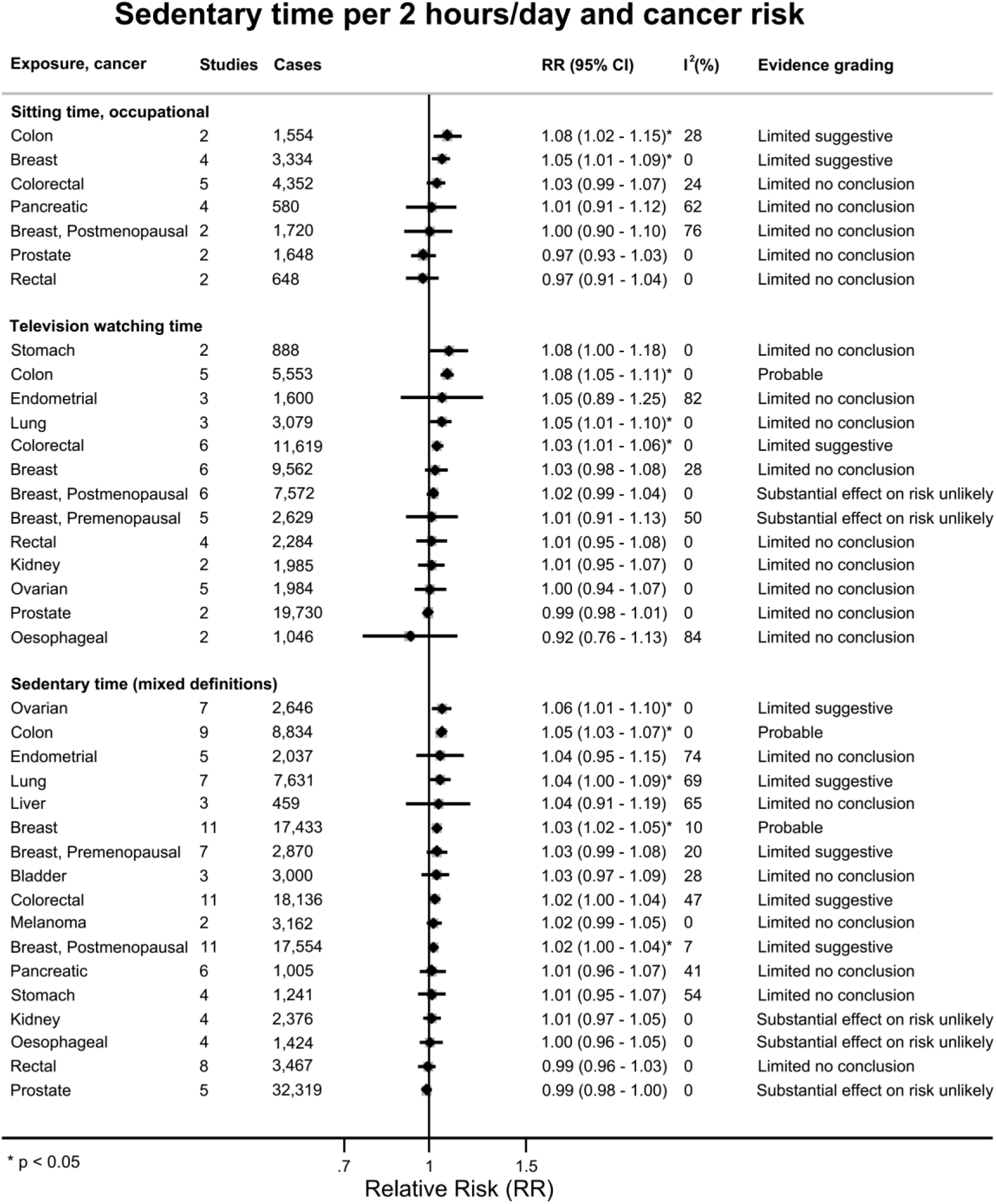
Summary forest plot showing the linear dose-response associations between sedentary time and cancer risk. Forest plot shows the summary results across the linear dose-response (per 2 hours/day increase of sedentary time) inverse variance DerSimonian-Laird random-effects model meta-analyses. Each square and the horizontal line across the square represent the relative risk (RR) estimate of the corresponding meta-analysis and its 95% confidence interval (CI). Evidence grades were classified as convincing, probable, limited suggestive, limited no conclusion, or substantial effect on risk unlikely (indicating the likelihood of causality). Note: † Indicates exposure–cancer pairs for which sufficient data for meta-analyses were only available after the combination of sedentary time definitions. These analyses were not possible when restricting to specific, non-mixed definitions of sedentary time. ‡ Includes one study on liver cancer mortality.

The positive associations between occupational sitting and breast and colon cancers, television watching and colorectal cancer, and sedentary time (mixed definitions) and colorectal, lung, ovarian, and premenopausal and postmenopausal breast cancers were graded as limited suggestive evidence. For colorectal cancer, the magnitude of the associations was attenuated compared to the analyses for colon cancer, likely driven by the null results for rectal cancer, resulting in lower evidence grades. Evidence for postmenopausal (smaller magnitude of association) and premenopausal (fewer cases, wider CIs) breast cancer was considered weaker compared to overall breast cancer but was not heterogenous by menopausal status. The association for ovarian cancer was based on a small number of cases, while that for lung cancer was smaller in magnitude with substantial observed heterogeneity. In general, limited suggestive gradings reflected positive associations that were supported by a combination of smaller number of studies or cases, more modest or less precise estimates, greater between-study heterogeneity, or weaker consistency across related exposure definitions than for the associations graded as probable.

Compared with the WCRF/AICR Third Expert Report, the accumulated evidence allowed updated evidence conclusions for some exposure–cancer pairs. The positive association between sitting time and endometrial cancer was previously graded as limited suggestive, based on a categorical highest versus lowest meta-analysis of three studies investigating total sitting, recreational sitting, or television watching time. The accumulated research findings allowed us to conduct a linear dose-response meta-analysis specifically for television watching time, where we observed no association. Furthermore, there was no association for sedentary time (mixed definitions) based on five cohorts, resulting in an updated grading of limited no conclusion.

Conversely, the Panel concluded that a substantial effect on risk is unlikely for the associations between sedentary time (mixed definitions) and kidney, oesophageal, and prostate cancers, and television watching time and premenopausal and postmenopausal breast cancers, based on summary estimates very close to the null and an evidence base considered sufficiently consistent for these associations at the time of the review. While for other exposure-cancer pairs we observed null associations of similar magnitudes, those were graded as limited no conclusion, reflecting a combination of a small number of studies or cases, more imprecise estimates, substantial between-study heterogeneity, or had an evidence base which was considered not sufficient to draw conclusions.

## Synopsis and discussion

We comprehensively reviewed 62 publications from 27 observational cohort studies, reporting results for 162,902 cancer incidence cases across 19 major anatomical sites and for 5,985 deaths across five major anatomical sites. There was evidence for probable causal positive associations between sedentary time (mixed definitions) and breast and colon cancer risk, and television watching time and colon cancer risk. The evidence for other cancer sites was generally weaker and more heterogeneous. In general, there was no evidence of deviations from linearity or any threshold effects identified.

Descriptive synthesis of isolated associations that were not included in meta-analyses indicated mostly null associations across major cancer sites, with few exceptions. Taken together, these findings build on the evidence considered in the WCRF/AICR Third Expert Report by strengthening support for specific exposure-cancer associations while also updating conclusions for others as more recent data have accumulated. The stronger evidence for breast and colon cancers reflects the greater consistency of findings across the specific exposure definitions and the larger evidence base available for these cancer sites.

Biological plausibility for these associations is supported by evidence linking sedentary behaviour with metabolic and hormonal pathways relevant to cancer development, although the underlying mechanisms remain incompletely understood.^110^ While sedentary behaviour is conceptually distinct from physical activity, the two exposures are closely related and may influence cancer through overlapping pathways, such as insulin resistance and the dysregulation of sex hormones, adipokines, and related biomarkers.^111^ Sedentary time has been shown to predict insulin resistance independently of physical activity,^112^ and interventions targeting sedentary behaviour, with or without physical activity, yield small but statistically significant reductions in insulin levels.^113,114^ Observational data from the UK Biobank suggest that sedentary time may impact the levels of sex hormones in a manner opposite to physical activity, potentially contributing to tumorigenesis.^115^ Additionally, sedentary behaviour correlates with elevated levels of adipokines and related biomarkers, though these associations are likely mediated by adiposity.^116^

Exposure misclassification was a potential limitation for all but one study included in the meta-analyses. Except for one accelerometer-based study,^88^ all studies utilised self-reported measures without an objective assessment or a validated tool. Self-reported tools correlate poorly with objective measures,^117–119^ often underestimating sedentary duration levels, particularly when single-item questions are used (e.g., “how may hours do you spend sitting in total?”).^120^ If the exposure misclassification is non-differential, and there is no additional misclassification of covariates, then misclassification have resulted in attenuated observed associations, which may further explain the modest magnitude of associations observed in our meta-analyses, as the true effects may be larger in magnitude. In addition, most studies included in the meta-analyses assessed sedentary behaviour at one point in time further misclassifying long-term exposure levels and reducing the precision of the estimated associations. To date, prospective studies examining objectively measured sedentary behaviour in relation to cancer incidence remain scarce, although relevant evidence is currently emerging in the literature.^88,121–123^ For example, the Women’s Health Accelerometry Collaboration reported a positive association between accelerometer-measured sitting time and overall cancer incidence among postmenopausal women, but the results for individual cancer sites were largely null, but those analyses were based on a small number of cancer cases.^121^ A UK Biobank study examining device-based sedentary behaviour and combined cancer sites reported no direct association, but a decreased risk of 13 physical-activity-related cancers in models substituting sedentary behaviour with physical activity.^122^ Additional evidence from ongoing cohorts linking cancer registries with accelerometer data, such as the Cancer Prevention Study-3 Accelerometry Substudy,^123^ may help clarify in the future associations with specific cancer sites.

Potential confounding bias was also a concern, with about half of the studies rated as critical risk due to inadequate adjustment for important confounders. However, subgroup analyses did not identify heterogeneity between studies at critical compared to studies at moderate/serious risk of bias or for studies adjusting and not adjusting for physical activity. We hypothesised that adiposity-related measures are potential mediators between sedentary behaviour and cancer risk, but 83% of studies included in the meta-analyses utilised them as confounders, despite evidence supporting a positive link of sedentariness to adiposity,^22,23^ which could explain the summary associations of modest magnitude observed in our meta-analyses. In studies providing both adiposity-adjusted and unadjusted models, the adjusted results were in general attenuated slightly towards the null.

Despite the inherent limitations of the field,^15^ this review also has several strengths. We provided an updated and comprehensive evaluation of the current evidence on sedentary behaviour and cancer risk, incorporating a substantial body of accumulated evidence from cohort and MR studies that has emerged since the Third Expert Report. The review was conducted following rigorous CUP Global protocols, and the quality of evidence was graded by a Panel of experts in the field. Additional strengths included the ability to conduct refined analyses by specific sedentary behaviour definitions and domains, and the use of both linear and non-linear dose-response meta-analyses, allowing for a more detailed assessment of the shape of the associations and whether there was evidence of thresholds. Taken together, this evidence will form part of the Fourth Expert Report, which will be developed by WCRF International in the coming years.

Given the global rise in sedentary time over recent decades,^1–4^ reducing sedentary behaviour could play a critical role in lowering the population-level risk of chronic diseases, including cancer. Importantly, the determinants of sedentary time, such as environmental, social, and individual level factors, are distinct from those influencing engagement in physical activity.^124^ Daily environments and personal habits play a particularly critical role in shaping sedentary lifestyles.^125,126^ As most studies in this review assessed sedentary behaviours in earlier periods, they may not fully capture the higher exposure levels seen today. If the observed associations are causal, current increases in sedentary behaviour could result in an even greater cancer burden in future populations. Thus, innovative interventions targeting sitting and low-intensity activities that utilise those environmental and habitual cues may have synergistic effects when combined with strategies designed to promote higher physical activity levels.^127^

In summary, this SLR indicated evidence for probable causal positive associations between sedentary time and risk of breast and colon cancers, and television watching time and risk of colon cancer, and limited suggestive evidence for positive associations for several other exposure–cancer pairs, including lung and ovarian cancers. Future research should focus on device-based assessments across different sedentary behaviour domains and dimensions, combined physical activity-sedentary behaviour patterns or exposure substitution models, inclusion of diverse populations and investigation of biological mechanisms to further clarify the impact of sedentary behaviour on cancer risk and strengthen the evidence base for more accurate public health guidance.

## Research in context

### Evidence before this study

At the time of the publication of the World Cancer Research Fund/American Institute for Cancer Research (WCRF/AICR) Third Expert Report in 2018, epidemiological evidence linking sedentary behaviour with cancer risk was limited and inconsistent. Evidence for a positive association with endometrial cancer was graded as limited suggestive, whereas no conclusion could be drawn for other investigated cancer sites. Since then, additional evidence from observational cohort studies has emerged, allowing more comprehensive investigation, such as associations across specific sedentary behaviour domains, particularly in occupational and recreational settings. Therefore, sedentary behaviour was identified as a priority topic for an updated systematic literature review (SLR) within the World Cancer Research Fund International’s Global Cancer Update Programme (CUP Global).

### Added value of this study

We conducted a SLR including dose–response meta-analyses of observational cohort studies and MR analyses examining sedentary behaviour and cancer risk. We reviewed 62 publications from 27 cohort studies including 162,902 incident cancer cases across 19 major anatomical sites and 15 MR studies. We evaluated associations across different domains and types of sedentary behaviour, including total and occupational sitting time, television watching time, and additional analyses of sedentary time using mixed definitions. We observed evidence for a probable causal positive association between sedentary time (mixed definitions) and breast and colon cancer risk, and television watching time and colon cancer risk. Limited suggestive evidence supported the associations of prolonged sedentary time (mixed definitions) with higher risk of colorectal, lung, ovarian, and premenopausal and postmenopausal breast cancers, as well as for occupational sitting time and breast and colon cancers, and television watching time and colorectal cancer.

### Implications of all the available evidence

The available evidence suggests that prolonged sedentary behaviour is probably a cause of breast and colon cancer while limited suggestive evidence supports positive associations for several other exposure-cancer pairs, including lung and ovarian cancers. While the magnitude of the observed associations is modest, given the increasingly high prevalence of sedentary behaviour at the population level, even these modest associations may result in a substantial cancer burden in the future. Still, evidence remains limited for many cancer sites and important uncertainties exist, particularly regarding the role of different sedentary behaviour domains, and the potential influence of exposure measurement error and residual confounding. Future research should prioritise objective measurements of sedentary behaviour and analyses integrating sedentary behaviour and physical activity in large prospective cohorts to better clarify these associations. Taken together, these findings support public health recommendations encouraging reductions in sedentary time alongside increasing physical activity as part of cancer prevention strategies.

### Panel: Grading the evidence

An Expert Panel convened by WCRF International interpreted and graded the quality of the evidence following pre-defined CUP Global evidence grading criteria (**Supplementary Table 5**). Evidence grading was based on aspects related to the quantity, consistency, magnitude and precision of the summary estimates, presence of a dose–response relationship, study design and risk of bias, generalisability, and mechanistic plausibility of the results from observational studies, as well as results from Mendelian Randomisation (MR) studies, where applicable. Evidence grades were classified as convincing, probable, limited suggestive, limited no conclusion, or substantial effect on risk unlikely (indicating the likelihood of causality).

Based on evaluation by the CUP Global Expert Panel, there was evidence for a probable causal positive association between sedentary time (mixed definitions) and breast and colon cancers, and between television watching time and colon cancer, supported by large number of prospective studies and cancer cases, consistent positive findings, and low between-study heterogeneity. Limited suggestive evidence supported the positive associations between occupational sitting time and breast and colon cancers, television watching time and colorectal cancer, sedentary time (mixed definitions) and colorectal, lung, ovarian, and premenopausal and postmenopausal breast cancers. An online interactive tool to help explore the evidence can be found at: https://teacup.cc.ic.ac.uk/sedentary-behaviour-cancer.html. Evidence presented in this systematic literature review will form part of the Fourth Expert Report which will be developed by WCRF International in the coming years.

## Acknowledgements

We thank Teresa Norat for leading the WCRF/AICR Continuous Update Project (CUP) as principal investigator from 2007 to 2020 and for developing the original review protocols. We thank the current and past WCRF International CUP/CUP Global Imperial team members for their contribution to the literature search, study selection, and/or data extraction; Lam Teng and past CUP/CUP Global Imperial database managers for implementing and updating the CUP Global database; and project manager: Eduardo Seleiro for organising the references, proofreading the manuscript and coordinating the work of CUP Global at Imperial College London.

## Data availability statement

Only publicly available data were used in our study. Data sources and handling of these data are described in the materials and methods section. Data are extracted into the CUP Global database and further details are available from the corresponding author upon request.

## Ethics statement

Ethical approval was not required, since this work was based solely on the analysis of published data without collection of new data or participant involvement.

