## Supplementary Table 1 for "Sedentary behaviour and cancer risk: a World Cancer Research Fund International Global Cancer Update Programme (CUP Global) systematic literature review and meta-analysis"

### Sedentary behaviour and cancer risk

#### Supplementary materials

#### Table of tables

Supplementary Figure 43: Linear dose-response meta-analysis of the association between occupational sitting time and colorectal cancer incidence, by physical activity adjustment. 185

Supplementary Figure 44: Linear dose-response meta-analysis of the association between occupational sitting time and colorectal cancer incidence, by risk of bias due to confounding (Domain 1). 186

Supplementary Figure 45: Linear dose-response meta-analysis of the association between occupational sitting time and colorectal cancer incidence, by risk of bias due to selective reporting (Domain 7). 187

Supplementary Figure 46: Linear dose-response meta-analysis of the association between total sitting time and colorectal cancer incidence, leave-one-out sensitivity analysis. 188

Supplementary Figure 47: Linear dose-response meta-analysis of the association between total sitting time and lung cancer incidence, leave-one-out sensitivity analysis. 189

Supplementary Figure 48: Linear dose-response meta-analysis of the association between total sitting time and breast cancer incidence, leave-one-out sensitivity analysis. 190

Supplementary Figure 49: Linear dose-response meta-analysis of the association between occupational sitting time and colorectal cancer incidence, leave-one-out sensitivity analysis. 191

Supplementary Figure 50: Linear dose-response meta-analysis of the association between occupational sitting time and pancreatic cancer incidence, leave-one-out sensitivity analysis. 192

Supplementary Figure 51: Linear dose-response meta-analysis of the association between occupational sitting time and breast cancer incidence, leave-one-out sensitivity analysis. 193

Supplementary Figure 52: Linear dose-response meta-analysis of the association between television watching time and oesophageal cancer incidence. 195

Supplementary Figure 53: Linear dose-response meta-analysis of the association between television watching time and stomach cancer incidence. 196

Supplementary Figure 54: Linear dose-response meta-analysis of the association between television watching time and colorectal cancer incidence. 197

Supplementary Figure 55: Linear dose-response meta-analysis of the association between television watching time and lung cancer incidence. 198

Supplementary Figure 66: Linear dose-response meta-analysis of the association between television watching time and colorectal cancer incidence, by physical activity adjustment. 210

Supplementary Figure 77: Linear dose-response meta-analysis of the association between television watching time and endometrial cancer incidence, by adjustment for adiposity... 221

Supplementary Figure 78: Linear dose-response meta-analysis of the association between television watching time and ovarian cancer incidence, by physical activity adjustment. ... 222

Supplementary Figure 90: Linear dose-response meta-analysis of the association between television watching time and breast cancer incidence, leave-one-out sensitivity analysis.. 234

Supplementary Figure 91: Linear dose-response meta-analysis of the association between television watching time and ovarian cancer incidence, leave-one-out sensitivity analysis.235

Supplementary Figure 105: Linear dose-response meta-analysis of the association between mixed definitions of sedentary time and breast cancer incidence by menopausal status. .. 249

#### Supplementary Text 1: Search strategy to find potential eligible studies.

##### A. PubMed

**a. Searching for studies on cancer**

| | |
|---|---|
| 1 | “Neoplasms”[MeSH Terms] |
| 2 | Neoplasm*[Title/Abstract] OR cancer*[Title/Abstract] OR carcinoma*[Title/Abstract] OR tumour*[Title/Abstract] OR tumor*[Title/Abstract] OR malignancy[Title/Abstract] OR malignant[Title/Abstract] OR adenocarcinoma[Title/Abstract] |
| 3 | leukemia[Title/Abstract] OR lymphoma[Title/Abstract] OR myeloma[Title/Abstract] |
| 4 | melanoma[Title/Abstract] |
| 5 | glioma[Title/Abstract] |
| 6 | #1 OR #2 OR #3 OR #4 OR #5 |

**b. Searching for studies on sedentary behaviour**

| | |
|---|---|
| 7 | “Sedentary Behavior”[MeSH Terms] OR “Screen Time”[MeSH Terms] OR “Leisure Activities”[MeSH Terms] OR “Inactivity”[Text Word] OR “sedentary*”[Text Word] OR “leisure activit*”[Text Word] OR “occupation* activity”[Text Word] OR “occupation* physical activity”[Text Word]) OR “sitting time*”[Text Word] OR “sitting hour*”[Text Word] OR “Screen Time”[Text Word] OR “screen use”[Text Word] OR “screen watching”[Text Word] OR “computer use”[Text Word] OR “internet use”[Text Word] OR (“television*”[Text Word] AND (“watching”[Text Word] OR “viewing”[Text Word])) OR (“sitting”[Text Word] AND (“occupation*”[Text Word] OR “work”[Text Word] OR “job”[Text Word])) |

**c. Combining the searches**

| | |
|---|---|
| 8 | #6 AND #7 |

**d. Limiting to human studies**

| | |
|---|---|
| 9 | animal [MeSH Terms] NOT human [MeSH Terms] |
| 10 | #8 NOT #9 |

##### B. For sedentary behaviour in Embase (Ovid)

**a. Searching for studies on cancer**

| | |
|---|---|
| 1 | exp neoplasm/ |
| 2 | (Cancer* or carcinoma* or tumour* or tumor* or malignancy or malignant or adenocarcinoma or leukemia or lymphoma or myeloma or melanoma or glioma).ab,ti. |
| 3 | 1 or 2 |

**b. Searching for studies on sedentary behaviour**

| | |
|---|---|
| 4 | exp sedentary lifestyle/ or exp sedentary time/ or exp screen time/ or exp television viewing/ or exp physical inactivity/ or inactivity.mp. |
| 5 | (Inactivity or 14edentary* or “leisure activities” or “occupation* activity” or “occupation* physical activity” or “sitting time*” or “sitting hour*” or “screen time” or “screen use” or “screen watching” or “computer use” or “internet use”).tw. |
| 6 | television.tw. |
| 7 | (watching or viewing).tw. |
| 8 | sitting.tw. |
| 9 | (occupation* or work or job).tw. |
| 10 | 6 and 7 |
| 11 | 8 and 9 |
| 12 | 4 or 5 or 10 or 11 |

**c. Combining the searches**

| | |
|---|---|
| 13 | 3 and 12 |

**d. Limiting to human studies**

| | |
|---|---|
| 14 | exp animal/ not exp human/ |
| 15 | 13 not 14 |

**e. Limiting to certain publication types**

| | |
|---|---|
| 16 | limit 15 to (article or article in press or conference paper or "conference review" or editorial or erratum or "review" or tombstone) |

#### Supplementary Text 2: Detailed information for study selection, data extraction and data analysis.

##### Search strategy and study selection criteria

We conducted comprehensive searches in PubMed and Embase until 01 September 2024, using pre-specified search algorithms (**Supplementary Text 1**). We also checked the reference lists of all relevant original publications of cohorts and related reviews for potential missed studies. Eligible studies were 1) randomised controlled trials, observational cohort studies, or pooled analyses of such studies, or Mendelian randomisation (MR) studies; 2) studies involving individuals who were not diagnosed with cancer (excluding non-melanoma skin cancer) at the time of the intervention or exposure evaluation; 3) studies assessing reported or measured sedentary behaviour (but not physical inactivity that captures the lack of meeting the physical activity guideline levels) as an exposure; 4) measured the incidence or mortality of primary cancer at any anatomical site during adulthood (a non-exhaustive list of cancer outcomes considered eligible for the present SLR is provided in **Supplementary Table 2**); and 5) provided multivariable adjusted estimates expressed as hazard ratios, risk ratios, relative risks, or odds ratios. For publications that reported on the same exposure-outcome associations from the same cohort, we selected those with sufficient information for dose-response meta-analyses, otherwise, we selected those with the highest number of cancer cases. Pooled analyses were selected over the individual articles published by their component studies. If available, we used study-specific estimates included in the pooled analyses**.**

##### Definition of exposures

Included studies used diverse definitions and/or assessments of sedentary behaviour. We primarily focused on studies assessing and comparing the levels of each sedentary behaviour. Studies that contrasted sedentary behaviour with other non-sedentary behaviours (e.g., various levels of physical activity) were considered out of the scope of this review, as they may not accurately reflect associations that could be attributed to sedentary behaviours[128,129]. Type of work was not used as proxy for sedentariness, as it is unlikely to provide a clear distinction on the level of sedentariness.

To include comparable exposures in a meta-analysis and avoid the combination of definitions that may reflect incompatible behaviours within the same meta-analysis, we categorised the sedentary behaviour based on its domains (total, occupational, recreational, transportational, and/or other) and dimensions (duration, frequency). Whenever possible, we further categorised specific types of sedentary behaviours, such as sitting, computer screening, and television watching time.

Additional meta-analyses were conducted pooling all related evidence across all exposure definitions to reflect overall sedentary behaviour time. Where multiple exposure definitions were available within a study, a pre-specified hierarchy was applied to prioritise inclusion in the mixed-definition additional analyses, based on definitions considered most informative for population-level interpretation. Total sedentary or sitting time was prioritised where available. Across the few studies that reported on multiple non-overlapping domains, when total measures were unavailable, the recreational sedentary behaviour domain (including focused definitions such as recreational screening time) was prioritised because it occurs across both working and non-working adults, contributes substantially to overall sedentary time at the population level, and represents modifiable behaviours that are relevant to population-level public health guidance. Occupational domain was subsequently selected for mixed-definition analyses, as it is more restricted to specific employment contexts and therefore may contribute to overall sedentary exposure in more context-specific way. Transportational sitting was given lower priority in the mixed-definition analyses, as it typically represents a smaller component of total sedentary time and is more constrained by contextual factors. Order of selection of exposure definitions for the mixed-definition additional analyses:

- Sedentary time, total
- Sedentary time, non-occupational (includes recreational and transportational)

- Sitting time, total
- Sitting time, recreational
  - Screen time, recreational
  - Screen time, recreational TV
  - Screen time, recreational, computers
- Sitting time, non-recreational (includes occupational and transportational)
- Sitting time, occupational
- Sitting time, transportational
- Sitting time, other definitions

**List of sedentary behaviour definitions in included studies**

| **Sedentary behaviour category** | **Study definitions** |
|---|---|
| Sedentary behaviour overall | |
| Sedentary time (total) | ➢ Time spent in sedentary activity (e.g., driving or watching television).<br>➢ Any sedentary behaviour that does not include sleeping, hard labour, walking, and standing. |
| Sedentary time (non-occupational) | ➢ Sum of driving, watching TV, computer using (not including during work) as part of healthy lifestyle score.<br>➢ Sum of time watching TV, using computer during leisure time, and driving for transportation. |
| Sitting time | |
| Sitting time (total) | ➢ Sitting at work, mealtimes, watching TV or seeing movies, sitting in a car and other sitting activities (e.g., reading, playing cards, sewing).<br>➢ Any sitting beside sleeping.<br>➢ Time spent at work, at home, while doing course work, and during leisure time. This may include time spent sitting at a desk, visiting friends, reading, or sitting, or lying down to watch television'<br>➢ Total sitting. |
| Sitting time (recreational) | ➢ Sitting outside of work (e.g., watching TV, reading etc.) |
| Sitting time (occupational) | ➢ Sitting time completing tasks at work.<br>➢ Sitting time at work.<br>➢ Sitting time at longest held job.<br>➢ Sitting time at work (including work-related computer/cell-phone use). |
| Sitting time (transportational) | ➢ Sitting time while driving.<br>➢ Sitting in a car or bus. |
| Sitting time (other)* | ➢ Occupational sitting time in women, alongside home duties.<br>➢ Other time spent sitting at home (e.g., reading, during meals, time at a desk, talking on the phone, reading, playing games, sewing)<br>➢ Sitting time excluding when watching TV. |
| Screen time | |
| TV watching time | ➢ Time spent sitting while watching TV.<br>➢ Time spent watching TV.<br>➢ Time spent watching TV or videos.<br>➢ Time spent in front of the television.<br>➢ Sitting while watching television or movies.<br>➢ Television viewing time. |
| TV watching frequency | ➢ How often watch television. |

| | |
|---|---|
| Screen time (recreational) | ➢ Using a computer and watching TV. |
| Screen time (recreational, computers) | ➢ Time spent using computers. |

* These exposure definitions were always reported along with at least one of the other main definitions.

#### Evidence synthesis

For inclusion in the meta-analyses, a model was considered multivariable if it adjusted for at least two variables in addition to the exposure of interest. Models adjusted for age or sex only were not considered multivariable. We aimed to include the estimated from multivariable maximally adjusted models in the meta-analyses. Literature evidence suggests that sedentary behaviour may be associated with higher adiposity[22,23], thus we considered adiposity as a potential mediator of the association between sedentary behaviour and cancer risk.

When possible, we opted to use the maximally-adjusted association estimates that were not adjusted for adiposity measures in our analyses. Since the handling of adiposity measures varied across studies, we adopted a structured approach to model selection and sensitivity analyses to address potential mediator adjustment bias. Primary studies were categorised based on how they handled adiposity measures in their reported multivariable models: (1) studies could report a single multivariable-adjusted model that excluded adiposity measures. These estimates were considered the most appropriate for inclusion in the primary meta-analysis, as these models avoided adjusting for mediators and captured the total effect of the exposure; (2) studies could report only a multivariable-adjusted model that included adiposity measures. These results were also included in the meta-analysis when no alternative multivariable estimates were available. Although adjusting for mediators (e.g., BMI) may attenuate the observed associations, these studies were retained to avoid omission of relevant studies; and (3) for studies presenting both types of models, we prioritised the mediator-unadjusted estimates (i.e., models without adiposity adjustment) as the most appropriate for inclusion in the primary meta-analysis.

Additionally, we compared the adiposity-unadjusted and adiposity-adjusted estimates, focusing on the magnitude of the estimates and the statistical significance of the associations. This approach aimed to evaluate the potential bias introduced by adjusting for mediator variables. In cases where large qualitative differences were observed (e.g., changes in statistical significance between the two models), sensitivity meta-analyses were performed using the adiposity-adjusted estimates.

The measures of association, including hazard ratio, risk ratio, and odds ratio, derived from the individual studies were aggregated and referred to collectively as the summary relative risk. These measures were roughly equivalent in research examining the current topic, based on the assumption that cancer cases are infrequent, and the absolute risks of cancer are minimal.

Studies of both sexes combined or of single sex only were meta-analysed together. When a study did not report an overall result but only reported results by sub-populations (e.g. males and females, smokers and non-smokers), a fixed effect model was used to estimate an overall result before pooling with the other studies in the meta-analysis. Similarly, studies reporting results only by cancer subtype (e.g., colon and rectal) were first pooled before meta-analysing with the other studies. In this case, the Hamling's method that accounts for the common denominators used in calculating the risk estimates for the outcomes was used [24]. When results from multiple studies were reported in the same publication (e.g. in a pooled analysis), the results of each component study were included separately if they were provided. The pooled result was not used directly, if possible, to maintain the independence of observations included and to allow exploration of heterogeneity across study results.

#### Statistical methods for meta-analysis

Separate meta-analyses were conducted for incidence (primary) and mortality (secondary) outcomes. For cancers with high mortality rate, including pancreas, liver, lung, and gallbladder cancers, we combined incidence and mortality outcomes in an additional analysis.

Both inverse variance fixed effect and DerSimonian-Laird random effects model[27] were used for categorical and linear dose-response meta-analyses. When important differences such as influence from small studies on the summary RRs were identified, both models were reported, otherwise, only the random effects summary estimates were reported. Linear dose-response meta-analyses were conducted when there were at least two independent studies reporting results on the same exposure-outcome association to allow expressing the results of each study in the same increment unit for a given exposure. To be included in a meta-analysis, studies must have reported the following data:

- For studies investigating cancer risk associated with a continuous exposure variable: relative risk and its measure of variance (standard errors (SE), 95% confidence intervals (CIs) or P-value) per unit increment in exposure that can be readily converted or re-scaled if needed. In cases where the units of measurement differed between results the units were converted, where possible, such that all results used the same measurement.
- For studies investigating cancer risk associated with a categorical comparison of exposure variable: relative risk and its measure of variance (as above), number of cases, person-years or number of non-cases, exposure categories (cut-offs, mean or median) for at least three exposure level comparisons. Standard approaches were used to derive missing data (e.g., number of participants or cases across categories) for meta-analysis [130,131].

The slope of the association estimated from a categorical analysis in a study was derived using generalised least-squares for trend estimation. When the assigned point of exposure of the reference group (relative risk = 1) was not zero, all the assigned exposures were rescaled as such[132,133].

When linear dose-response meta-analysis was not possible or when more than a third of the included studies did not have sufficient information for inclusion in a linear dose-response meta-analysis, a supporting categorical high versus low analysis was conducted. When a meta-analysis was not possible or for individual studies that could not be meta-analysed, the studies were descriptively reviewed. This approach allows a comprehensive summation of study results while minimising the number of categorical meta-analysis that may pool results for different exposure comparisons. For descriptive synthesis, the results for each exposure-outcome association reported in the individual studies were presented in supplementary tables and reviewed briefly in the text.

Non-linear dose-response meta-analyses were assessed by one-stage mixed effects models using restricted cubic splines with three knots placed at $10^{th}$, $50^{th}$, and $90^{th}$ percentiles of the overall exposure distribution[28]. The likelihood ratio test was used to assess deviations from linearity[29]. Two hours/day was selected as increment unit for the linear meta-analyses and as referent point of exposure for the non-linear meta-analyses. An increment unit of 2 hours/day in sedentary time was used, consistent with the previous CUP SLRs.

The proportion of total variability in effect estimates attributable to heterogeneity between studies was assessed by the $I^2$ metric[30]. Potential sources of heterogeneity were explored by pre-specified subgroup analyses when at least two primary studies reported results for the same subgroup. Pre-specified subgroups included participant's characteristics (sex, race/ethnicity, BMI status, physical activity level, and smoking status), study characteristics (geographical location, publication year, study size, length of follow-up, and risk of bias domains), and exposure assessment.

Leave-one-out sensitivity analyses were conducted to assess the impact of omitting individual studies on the summary estimate[31]. In case where primary studies reported both adiposity adjusted and adiposity unadjusted estimates that resulted in differences in the inference (change in statistical significance) we conducted a sensitivity analysis by replacing the adiposity-unadjusted estimate with the adiposity-adjusted one, to assess its impact on the summary estimate. The low number of studies per meta-analysis (less than 10) precluded the investigation of small study effects by the Egger's regression test[32] or by visual inspection of the funnel plots.

#### Mendelian randomisation (MR) studies

We included MR studies using genetic variants derived from genome-wide association studies (GWAS) as instrumental variables to assess associations of genetically predicted sedentary behaviour and cancer risk. We extracted the following information: the study design (one- or two-sample MR), exposure and outcome GWAS data sources, population ancestries and sample sizes, information on genetic instruments including number of single nucleotide polymorphisms (SNPs) associated with sedentary traits utilised in the analyses and variance explained ($R^2$) by the instrument, primary analysis (e.g., fixed or random effects inverse-variance weighted [IVW] method), sensitivity analyses assessed (e.g., MR-Egger, weighted median, MR-PRESSO), and any multivariable MR analyses conducted and their results. When multiple analyses on overlapping exposure and outcome populations from different publications were identified, the analysis utilising the largest outcome sample size (reflecting the largest power) was considered as the primary MR analysis, while the rest were also kept and checked for consistency of results against the main analysis. We further assessed the concordance and robustness of the main MR results against the sensitivity analyses.

### Supplementary Text 3: Detailed information for study results.

#### Sedentary time

Two studies (four publications)[83,88,91,92] with 4,656 cases of cancer incidence at three anatomical sites reported on total and non-occupational sedentary time **(Supplementary Tables 7, 8)**. Meta-analysis was not conducted. Results for the categorical highest versus lowest comparisons are presented in forest plots in **Supplementary Figures 8-20**.

In J-MICC, total sedentary time was positively associated with breast cancer in categorical analysis (MV, PA and BMI adjusted $RR_{\geq 13 \text{ vs } <7 \text{ hours/day}}$:1.34; 95%CI: 1.02-1.75; 554 cases), and there was indication of association in linear analyses but with 95%CI crossing the null (MV, PA and BMI adjusted $RR_{\text{per 1 hour/day}}$: 1.02; 95%CI: 1.00-1.04)[91]. Similar associations were observed for pre- and postmenopausal breast cancer **(Supplementary Table 11)**. When the association of leisure-time physical activity was examined in combination with sedentary time, sedentary time was shown to outweigh the protective association of physical activity[91] **(Supplementary Table 12)**.

In UK Biobank, objectively measured sedentary time was indicative of an association with non-melanoma skin cancer but the 95%CI crossed the null (MV, PA and BMI not adjusted $RR_{\text{Per 20 mins/day}}$: 0.99; 95%CI: 0.97-1.00; 1,734 cases), while in a substitution model where 20 minutes/day of sedentary time were replaced with equivalent time of light or moderate-to-vigorous physical activity, no association was observed[88]. Also in UK Biobank, self-reported non-occupational sedentary time was not linearly associated with melanoma[83]. In addition, low versus high levels of non-occupational sedentary time were inversely associated with pancreatic cancer risk in males (MV, PA and BMI not adjusted $RR_{<3 \text{ vs } \geq 5 \text{ hours/day}}$:0.70; 95%CI: 0.53-0.92) but not in females (MV, PA and BMI not adjusted $RR_{<3 \text{ vs } \geq 5 \text{ hours/day}}$:0.78; 95%CI: 0.60-1.02)[92] **(Supplementary Table 11; Supplementary Figure 12)**.

#### Sitting time

Twenty-one studies (41 publications)[33,34,36,38-46,48,49,51-53,55-63,65,66,70-73,75-77,84-86,89,90,94] with 119,737 cases of cancer incidence at 19 anatomical sites, and 15,899 cases of cancer mortality at two anatomical sites reported on sitting time **(Supplementary Tables 7, 9)**. Meta-analyses were possible for total and occupational sitting time.

##### *Meta-analyses of sitting time and cancer incidence*

Meta-analyses were conducted for total sitting time and cancer incidence at 4 anatomical sites with 2 to 6 studies each, and for occupational sitting time and cancer incidence at 5 anatomical sites with 2 to 5 studies each **(Supplementary Figures 21-45)**.

Total sitting time was linearly associated with higher risk of breast cancer ($RR_{per\ 2\ hours/day}$: 1.04; 95%CI: 1.01-1.07; $I^2$: 0%; 4 studies[46,55,60,85]; 3,658 cases) **(Supplementary Figure 23)**. Total sitting time was not linearly associated with colorectal ($RR_{per\ 2\ hours/day}$: 1.01; 95%CI: 0.98-1.03; $I^2$: 34.6%; 3 studies[61,66,73]; 5,561 cases) **(Supplementary Figure 21)**, lung ($RR_{per\ 2\ hours/day}$: 1.02; 95%CI: 0.97-1.07; $I^2$: 27.8%; 3 studies[48,59,72]; 2,084 cases) **(Supplementary Figure 22)**, and ovarian ($RR_{per\ 2\ hours/day}$: 1.03; 95%CI: 0.98-1.09; $I^2$: 0%; 3 studies[49,84]; 1,347 cases) cancer **(Supplementary Figure 24)**. A high versus low analysis was conducted for prostate cancer with an RR of 1.08 (95%CI: 0.87-1.34; $I^2$: 84.9%; 2 studies[53,70]; 14,640 cases) **(Supplementary Figure 25)**.

Occupational sitting time was positively linearly associated with the risk of ovarian ($RR_{per\ 2\ hours/day}$: 1.25; 95%CI: 1.12-1.40; $I^2$: 3.7%; 3 studies in two publications[77,84]; 908 cases) **(Supplementary Figure 29)** and breast cancer ($RR_{per\ 2\ hours/day}$: 1.05; 95%CI: 1.01-1.09; $I^2$: 0%; 4 studies[42,46,60,77]; 3,334 cases) **(Supplementary Figure 28)**. No association was observed with colorectal ($RR_{per\ 2\ hours/day}$: 1.03; 95%CI: 0.99-1.07; $I^2$: 24.4%; 5 studies in four publications[45,58,76,77]; 4,352 cases) **(Supplementary Figure 26)**, pancreatic ($RR_{per\ 2\ hours/day}$: 1.01; 95%CI: 0.91-1.12; $I^2$: 62.1%; 4 studies in three publications[43,65,77]; 580 cases) **(Supplementary Figure 27)**, and prostate cancer risk ($RR_{per\ 2\ hours/day}$: 0.97; 95%CI: 0.93-1.03; $I^2$: 0%; 2 studies[34,77]; 1,648 cases) **(Supplementary Figure 30)**.

##### *Subgroup and sensitivity analyses*

The results of a categorical high versus low analysis for breast cancer, with two additional studies not included in the dose-response analysis[70,86], supported the linear analysis findings **(Supplementary Figure 31)**. For breast cancer, no heterogeneity was observed in subgroup analyses of total sitting time by menopause status (pre- and postmenopausal), hormone receptor status (ER+/PR+ and ER-PR-), physical activity adjustment (adjusted and not adjusted), and risk of bias due to confounding (critical and moderate/serious) and for occupational sitting time by menopause status **(Supplementary Table 13; Supplementary Figures 32-37)**.

For colorectal cancer, the results of a categorical high versus low analysis, with one additional study not included in the dose-response analysis[70], supported the linear analysis findings **(Supplementary Figure 38)**. For colorectal cancer anatomical subsites (colon and rectal), no subgroup heterogeneity was observed for total sitting time **(Supplementary Figure 39)**, however there was an indication of subgroup heterogeneity for occupational sitting time ($P_{subgroup-heterogeneity}$: 0.021) **(Supplementary Figure 40)**. Specifically, the association was more profound in colon cancer ($RR_{per\ 2\ hours/day}$: 1.08; 95%CI: 1.02-1.15; $I^2$: 28.2%; 2 studies) compared to no observed association in rectal cancer ($RR_{per\ 2\ hours/day}$: 0.97; 0.91-1.04; $I^2$: 0%; 2 studies) **(Supplementary Table 13; Supplementary Figure 40)**. No heterogeneity was observed in subgroup analyses of occupational sitting time by sex (male and female), region (Southeast/eastern Asia and North America), physical activity adjustment (adjusted and not adjusted), and risk of bias due to confounding and selective reporting (critical and moderate/serious) **(Supplementary Table 13; Supplementary Figures 41-45)**.

In general, we did not observe large influences of individual studies on the summary estimates in the leave-one-out sensitivity analysis **(Supplementary Figures 46-51)**, although the number of included studies in the meta-analyses was generally low.

##### *Descriptive synthesis of sitting time and cancer incidence*

Results for the categorical highest versus lowest comparisons are presented in forest plots in **Supplementary Figures 8-20**.

Four studies (8 publications[39-41,49,52,53,70,72,75]) investigating total sitting time with cancer incidence at 8 major anatomical sites could not be included in the meta-analyses. No associations were observed across the 16 exposure-cancer/cancer subsite associations, except for a higher risk for endometrial cancer in NIH-AARP (MV, PA adjusted and BMI not adjusted $RR_{\geq 9\ vs\ \leq 2.99\ hours/day}$:1.45; 95%CI: 1.10-1.92; $P_{trend}$ <0.01; 888 cases)[40] **(Supplementary Table 11; Supplementary Figures 8-20)**.

Nine publications[40,49,53,60,61,63,66,73,84] reported results on combined, joint, or interaction analyses of sedentary behaviour with physical activity. Interactions were observed between total sitting time and strenuous/vigorous recreational physical activity in relation to endometrial [40] and breast cancer risk[60,63], but were not observed for total sitting time and walking or total physical activity with breast cancer. For endometrial cancer, the positive association with sitting time was stronger among less active compared to more active individuals. In contrast, for breast cancer, the positive association with sitting time was stronger among more active compared to less active individuals. No interactions were observed with colorectal cancer in the three studies reporting results[61,66,73]. No interactions were observed on ovarian[49,84], or prostate cancers risk[53]**(Supplementary Table 12)**.

Five publications from one study, the CPS II[44,51,56,57,89], investigated the association of recreational sitting time and 30,071 incidence/mortality cancer cases across 18 anatomical sites or subsites, reporting results on 38 exposure-outcomes pairs (overall and sex-specific). Of these, associations were observed only with endometrial (MV, PA adjusted and BMI not adjusted $RR_{\geq 6\ vs\ \leq 2\ hours/day}$:1.34; 95%CI: 1.08-1.67; 387 cases)[56], ovarian (MV, PA not adjusted and BMI adjusted $RR_{\geq 6\ vs\ \leq 2.9\ hours/day}$: 1.44; 95%CI: 1.12-1.85; $P_{trend}$ = 0.006; 638 cases) and serous ovarian cancer (MV, PA not adjusted and with BMI adjusted $RR_{\geq 6\ vs\ \leq 2.9\ hours/day}$: 1.52; 95%CI: 1.06-2.16; $P_{trend}$ = 0.01; 313 cases)[57], and non-Hodgkin lymphoma in females (MV, PA and BMI adjusted $RR_{\geq 6\ vs\ \leq 2\ hours/day}$: 1.26; 95%CI: 1.01-1.59; $P_{trend}$ = 0.011; 863 cases)[44] **(Supplementary Table 11; Supplementary Figures 8-20)**. No interaction or heterogeneity was observed between recreational siting time and physical activity in the two studies reporting results)[44,56] **(Supplementary Table 12)**.

One publication from NHS II found no association between non-recreational sitting time and colorectal cancer incidence in young adults (diagnosed before 50 years old)[71] **(Supplementary Table 11; Supplementary Figure 8)**.

One study on occupational sitting time and colon and rectal cancer incidence from the 1960 Swedish census, which lacked sufficient data for inclusion in the linear meta-analyses, reported similar results to the main meta-analysis[94]. Results for different colon anatomical subsites were reported in two studies, indicating generally positive associations with occupational sitting time[45,94]. One and two studies on premenopausal[60] and postmenopausal breast cancer[33,60], respectively, found no association with occupational sitting time, contrasting with our main meta-analysis results. In the JPHC study, sex-specific associations of occupational sitting time with cancer incidence at seven anatomical sites (that were not assessed in other studies and not possible to meta-analyse) were mostly null, except for a positive association with female lung cancer (MV, PA and BMI adjusted $RR_{\geq 7\ vs\ 1\text{-}<3\ hours/day}$: 2.80; 95%CI: 1.33-5.90; $P_{trend}$ = 0.013; 81 cases)[77] **(Supplementary Table 11; Supplementary Figures 8-20)**. No interaction or heterogeneity was observed between occupational siting time and physical activity in the three publications reporting results)[42,77,84] **(Supplementary Table 12)**.

Three studies investigated transportational sitting time, one study each from HPFS in colorectal cancer[58], UK Biobank in pancreatic cancer[90], and SCCS in breast cancer[46], reporting no association **(Supplementary Table 11; Supplementary Figures 10, 12, 14)**.

Five studies (five publications[46,62,71,77,84]) investigated various other siting times with cancer incidence at 11 major anatomical sites across 17 main exposure-anatomical cancer site/subsite pairs, with no association observed **(Supplementary Table 11; Supplementary Figures 8-20)**. No heterogeneity was observed by physical activity level in one study reporting results[84] **(Supplementary Table 12)**.

##### *Descriptive synthesis of sitting time and cancer mortality*

Two studies investigated total sitting time, one study each from NIH-AARP on prostate cancer mortality[53] and one from WHI on lung cancer mortality[59], reporting no associations **(Supplementary Figures 13, 17)**.

#### Screen time

Fourteen studies (33 publications)[35-39,41,46-50,52-55,58,60,62,64,67-69,71,73,74,78-82,84,87,93] with 60,171 cases of cancer incidence at 16 anatomical sites and 5,171 cases of cancer mortality at five anatomical sites reported on screen time and its different types(one study in two publications on recreational screen time[68,93]; one study in one publication on recreational computer screen time[67]; 13 studies in 30 publications on TV watching time[35-39,41,46-50,52-55,58,60,62,64,67,69,71,73,74,78,79,81,82,84,87]; one study in one publication on TV watching frequency[80]) **(Supplementary Tables 7, 10)**. Meta-analyses were only possible for TV watching time.

##### *Meta-analyses of TV watching time and cancer incidence*

Meta-analyses were conducted for TV watching time and cancer incidence at 11 major anatomical sites with 2 to 6 studies each **(Supplementary Table 7; Supplementary Figures 52-95)**. The categorical highest versus lowest comparisons are presented in forest plots without a meta-analysis in **Supplementary Figures 8-20**.

In linear dose-response meta-analyses, TV watching time was positively linearly associated with colorectal ($RR_{per\ 2\ hours/day}$: 1.03; 95%CI: 1.01-1.06; $I^2$=0%; 6 studies in five publications; 11,619 cases) **(Supplementary Figure 54)** and lung cancer incidence ($RR_{per\ 2\ hours/day}$: 1.05; 95%CI: 1.01-1.10; $I^2$=0%; 3 studies[47,48,78]; 3,079 cases) **(Supplementary Figure 55)**. Non-linear meta-analyses were possible for, colorectal, colon, and breast cancers, without evidence of non-linearity **(Supplementary Figures 61-63)**.

There was an indication of a positive linear association with stomach cancer but with the 95%CI crossing the null ($RR_{per\ 2\ hours/day}$: 1.09; 95%CI: 1.00-1.18; $I^2$=0%; 2 studies[52,78]; 888 cases), while no association was observed with breast (RR: 1.03; 95%CI: 0.98-1.08; $I^2$=27.8%; 6 studies [46,55,60,74,78,82], $P_{non-linearity}$ = 0.794; 9,562 cases), endometrial ($RR_{per\ 2\ hours/day}$: 1.05; 95%CI: 0.89-1.25; $I^2$=82%; 3 studies[38,78,79]; 1,600 cases), oesophageal (RR: 0.92; 95%CI: 0.76-1.13; $I^2$=84%; 2 studies[52,78]; 1,046 cases), ovarian (RR: 1.00; 95%CI: 0.94-1.07; $I^2$=0%; 5 studies in four publications[49,64,78,84]; 1,984 cases), prostate (RR: 0.99; 95%CI: 0.98-1.01; $I^2$=0%; 2 studies[53,78]; 19,730 cases) or kidney cancer (RR: 1.01; 95%CI: 0.95-1.07; $I^2$=0%; 2 studies[41,78]; 1,985 cases). A categorical high versus low analysis was possible for bladder cancer with a summary RR of 1.15 (95%CI: 0.90-1.46; $I^2$=0%; 2 studies[37,78]; 2,386 cases) **(Supplementary Figure 64)**.

##### *Subgroup and sensitivity analyses*

Subgroup analyses were possible by menopause status (premenopausal and postmenopausal) for breast cancer, anatomical subsite (colon and rectal) for colorectal cancer, sex (male and female) for colorectal, colon, and rectal cancers, region (Europe and North America) for breast cancer, physical activity adjustment (adjusted and not adjusted) for breast and ovarian cancers, adiposity adjustment (adjusted and not adjusted for adiposity across all studies and restricted to studies with both models) for colorectal cancer and endometrial cancer, BMI category (BMI<30 kg/m$^2$ and BMI>30 kg/m$^2$) for prostate cancer, and risk of bias due to confounding (critical and moderate/serious) for colorectal, breast, and lung cancers. There was no evidence of heterogeneity in most subgroup analyses (all $P_{subgroup-heterogeneity}$ > 0.158) **(Supplementary Table 14; Supplementary Figures 65-79)**, except for endometrial cancer incidence by adjustment for adiposity ($P_{subgroup-heterogeneity}$: 0.001), but this was based on 2 and 1 study in each subgroup. Specifically, the association was more profound in one study not adjusted for adiposity ($RR_{per\ 2\ hours/day}$: 1.16; 95%CI: 1.07-1.26) compared to no observed association in studies adjusting for adiposity ($RR_{per\ 2\ hours/day}$: 0.95; 0.87-1.04; $I^2$: 0%; 2 studies) **(Supplementary Table 14; Supplementary Figure 101)**.

For colorectal cancer, when the adiposity unadjusted study results were replaced with the adiposity-adjusted study results, the summary estimate was similar ($RR_{per\ 2\ hours/day}$: 1.03; 95%CI: 1.00-1.06; $I^2$=8%) **(Supplementary Figure 80)**, while for endometrial cancer the association was attenuated ($RR_{per\ 2\ hours/day}$: 1.00; 95%CI: 0.92-1.01; $I^2$=40.4%) **(Supplementary Figure 81)**. The results of a categorical high versus low analysis with the addition of one extra study each for endometrial[35] and for lung cancer[87] not included in the dose-response analyses, supported the linear dose-response findings **(Supplementary Figures 82-83)**, without evidence for subgroup heterogeneity by adjustment for adiposity for endometrial cancer and by risk of bias due to confounding for lung cancer **(Supplementary Figures 84-85)**. No evidence of non-linearity was observed for colorectal and colon cancers in the sensitivity analyses using the adiposity-adjusted results **(Supplementary Figures 86-87)**.

In general, we did not observe large influences of individual studies on the summary estimates in the leave-one-out sensitivity analyses **(Supplementary Figures 88-92)**, apart from endometrial cancer incidence, where the exclusion of the UK Biobank study[78] resulted in a positive association ($RR_{per\ 2\ hours/day}$: 1.16; 95%CI: 1.07-1.25; $I^2$=0%) **(Supplementary Figure 92)**.

##### *Descriptive synthesis of TV watching time and cancer incidence*

One publication from NHS II[71] observed a positive association between TV watching time and colorectal and rectal cancer incidence in young adults (diagnosed before 50 years old). Two publications from NIH-AARP[49,53] and UK Biobank[67,78] each provided results on single exposure-cancer associations that could only be descriptively synthesised, reporting no associations for epithelial ovarian[49], advanced prostate[53], [39]proximal and distal colon[67], hepatobiliary tract[78], pancreatic[78], brain[78], thyroid[78], and non-Hodgkin lymphoma cancers[78] incidence **(Supplementary Table 11; Supplementary Figures 8-20)**. Substitution models in UK Biobank showed that replacing 1 hour/day of TV watching with 1 hour of moderate-intensity physical activity or walking reduced the risk of oropharyngeal, lung, breast, and colorectal cancers [78].

Nine publications reporting results[53,58,62,67,71,73,78,82,84], did not find evidence of interaction or combined effects for most cancer sites, with the exception of some inconsistent evidence of interaction on colorectal cancer. Out of the six publications reporting results[58,62,67,71,73,78], only two reported some evidence of interaction for colorectal[62] or rectal cancer[78], suggesting that the inverse association of physical activity and cancer risk was more profound among individuals with higher compared to lower time spent watching television. **(Supplementary Table 12)**.

##### *Meta-analyses of TV watching time and cancer mortality*

Meta-analyses were possible for TV watching time and colorectal, prostate, and breast cancer mortality with two studies each **(Supplementary Figures 93-95)**. No association was observed with colorectal (RR: 1.07; 95%CI: 0.96-1.19; $I^2$=57.5%; 2 studies; 1,438 cases[50,81]), and prostate cancer mortality (RR: 1.10; 95%CI: 0.98-1.22; $I^2$=38.7%; 2 studies; 993 cases[50,53]). A categorical meta-analysis was possible for breast cancer mortality, indicating no association with high versus low levels of TV watching time (RR: 1.17; 95%CI: 0.85-1.61; $I^2$=0%; 2 studies; 775 cases[50,69]).

##### *Descriptive synthesis of TV watching time and cancer mortality*

One study each reported on TV watching time and colon, rectal[81], and lung[50] cancer mortality. In JACC, TV watching time was positively linearly associated with risk of colon cancer (MV, PA and BMI adjusted $RR_{per\ 1\ hour/day}$:1.07; 95%CI: 1.02-1.13; 531 cases), but not for rectal cancer (MV, PA and BMI adjusted $RR_{per\ 1\ hour/day}$:1.03; 95%CI: 0.94-1.13; 218 cases)[81]. In MEC, TV watching time was not associated with lung cancer mortality[50] **(Supplementary Table 11; Supplementary Figures 8, 13)**.

Two publications reported interaction or combined results with physical activity[54,81]. No interaction was observed on colorectal[81] or liver cancer mortality[54] **(Supplementary Table 12)**.

##### *Descriptive synthesis of other types of screen time and cancer incidence*

Three publications from UK Biobank investigated general recreational screen time (watching TV and using a computer, excluding at work)[68,93] and recreational computer screen time[67] with cancer incidence. Since the distinct associations on TV watching time from UK Biobank have already included in the TV watching time syntheses, these exposures are assessed separately. A positive association was observed between recreational screen time and lung cancer incidence (MV, PA and BMI adjusted $RR_{>4 \text{ vs } <3 \text{ hours/day}}$:1.26; 95%CI: 1.13-1.40; 2,765 cases[93]), and there was an indication of a positive association with colorectal cancer but with the 95%CI crossing the null (MV, PA and BMI adjusted $RR_{>4 \text{ vs } <3 \text{ hours/day}}$:1.08; 95%CI: 1.00-1.17; 4,560 cases[93]). No associations were reported for breast[93], prostate[93], oesophageal[68], and gastric[68] cancers, or between recreational computer screen time and colorectal, colon (total, distal, proximal), or rectal cancers incidence[67]. One study from ARIC investigated TV watching frequency (how often) with colorectal, lung, postmenopausal breast, and prostate cancers, reporting no associations[80] **(Supplementary Table 11; Supplementary Figures 8-20)**. In one study reporting results, no interaction was observed between total physical activity and recreational screen time on prostate, breast, colorectal, and lung cancer risks[93] **(Supplementary Table 12)**.

#### Mixed definitions of sedentary time

Additional meta-analyses on mixed definitions of sitting and sedentary time were largely consistent with the main analyses on total and occupational sitting time and television watching time for colorectal, colon, rectal, pancreatic, lung, breast (total, pre-menopausal, post-menopausal), ovarian, endometrial, oesophageal, kidney, stomach, and prostate cancers incidence **(Supplementary Figures 96-111)**. Positive or marginally positive associations were observed between mixed definitions sedentary time and colorectal cancer ($RR_{\text{per 2 hours/day}}$: 1.02; 95%CI: 1.00-1.04; $I^2$=46.5%; 11 studies; 18,136 cases[45,56,58,61,62,66,73,76,77,93]) **(Supplementary Figure 98)**, colon ($RR_{\text{per 2 hours/day}}$: 1.05; 95%CI: 1.03-1.07; $I^2$=0%; 9 studies; 8,834 cases[36,45,58,61,62,66,77,78]) **(Supplementary Figure 99)**, lung ($RR_{\text{per 2 hours/day}}$: 1.04; 95%CI: 1.00-1.09; $I^2$=69.3%; 7 studies; 7,631 cases[47,48,56,59,72,77,93]) **(Supplementary Figure 102)**, breast ($RR_{\text{per 2 hours/day}}$: 1.03; 95%CI: 1.02-1.05; $I^2$=9.8%; 11 studies; 17,433 cases[42,46,55,56,60,74,77,82,85,91,93]) **(Supplementary Figure 104)**, and pre and post-menopausal breast ($RR_{\text{per 2 hours/day}}$: 1.03; 95%CI: 0.99-1.08; $I^2$=20.1%; 7 studies; 2,870 cases and $RR_{\text{per 2 hours/day}}$: 1.02; 95%CI: 1.01-1.04; $I^2$=7.4%; 11 studies; 17,554 cases, respectively[33,39,51,55,60,63,74,78,82,85,91]) **(Supplementary Figure 105)**, and ovarian cancers ($RR_{\text{per 2 hours/day}}$: 1.06; 95%CI: 1.01-1.10; $I^2$=0%; 7 studies; 2,646 cases[49,57,64,77,78,84]) **(Supplementary Figure 107)**. Furthermore, new meta-analyses using the mixed definition of sedentary time were possible for liver, advanced prostate, skin (melanoma), and bladder cancer incidence, for which, meta-analyses on specific definitions of sitting or television watching time were not possible. All new associations were null. In general, we observed little evidence of influential studies in the leave-one-out sensitivity analysis **(Supplementary Figures 112-121)**.

#### Supplementary Text 4: Risk of bias of included studies.

The risk of bias assessment showed considerable variation across domains **(Supplementary Figure 122)**. In bias due to confounding, 43% of studies were classified as having a serious risk and 46% as critical, while only 11% had a moderate risk. Studies with critical risk did not adjust for all critically important confounding variables **(Supplementary Table 4)**. Studies were assessed as having serious risk mainly because key confounders such as physical activity, dietary factors, and comorbidities were often self-reported without further validation. In addition, 83% studies adjusted for adiposity, which we considered as potential mediator in the association between sedentary behaviour and cancer risk, but we did not consider this adjustment in the risk of bias assessment. For analyses specifically examining TV watching time, we assessed whether studies adjusted for education and dietary habits. Most associations were adjusted for at least one of these factors, and only four associations pertaining to lung[47], ovarian[84], and endometrial cancer incidence (main and sensitivity analyses)[38] were not adjusted for either education or any aspect of diet.

Regarding bias in the selection of participants, 48% of studies were at low risk, 34% had a moderate risk and 18% a serious/critical risk. Bias in the classification of exposures was rated as low/moderate in 57% of studies and serious in 41%, with a small proportion (2%) at critical risk. Specifically, exposure assessment in all studies included in meta-analyses relied on self-reported measures, and validity or reliability statistics were not presented in about half of them.

Bias due to departures from intended exposures was the most severe, with 95% of studies at critical risk, 4% at serious risk, and only 2% at moderate risk. These studies were rated as such because changes in exposure status may have occurred after baseline exposure assessment. Most included studies assessed exposure only at baseline, except for one study with time-varying analyses[60] and two with cumulative average data[58,61]. None of these studies compared the results with models utilising only the baseline data.

Regarding bias due to missing data, 56% of studies had a low or moderate risk, 29% had a serious risk, 2% had a critical risk, and 14% lacked sufficient information. Specifically, studies were rated as serious or critical risk because authors excluded substantial proportions (>10%) of participants due to missing data on exposure status and other variables (besides outcome data and exposure status), while these studies did not use appropriate statistical methods (e.g., multiple imputation technique) to account for missing data.

Bias in the measurement of outcomes was not a concern, as 98% of studies were rated as low risk with 2% lacking sufficient information. Bias in the selection of reported results was moderate in 88% of studies, while 11% were classified as serious/critical, because not all findings for pre-specified outcomes and analyses were reported, or only statistically significant results were shown without mentioning the statistical significance of non-reported findings. Overall, confounding and exposure measurement error posed the greatest risks.

#### Supplementary Text 5: Mendelian randomisation studies.

##### *Study characteristics of MR studies*

Fifteen publications were identified[95-109] **(Supplementary Tables 15-17)** reporting results on 191 MR analyses of sedentary time with cancer, using summary or individual-level information from large GWAS studies or consortia. Six publications investigated genetically predicted measures of sedentary behaviour[96-98,101,105,134] across 19 MR analyses and 11 publications investigated self-reported sedentary behaviour[97,99,100,102-109] across 172 MR analyses. All studies comprised individuals of European descent. There was considerable overlap of data sources across studies for the exposure, with most studies utilising data from UK Biobank[95-107,109] and from a GWAS meta-analysis of 51 studies[105,107,108]. There was also some overlap for the outcomes, specifically for breast[98,101,103,104,107], lung[97,99,100,106,107], prostate[95,103,104,107], and ovarian[98,103,107] cancers.

##### *Results of MR studies*

The studies reported little evidence of an association with genetically predicted objectively measured sedentary behaviour. The genetic instruments included 2-6 SNPs and only explained 0.08%-0.14% of variance in sedentary behaviour. There was a lack of associations with genetically predicted self-reported sedentary behaviour at work (4 SNPs, 0.04% variance), except for an inverse association with breast cancer[107], which was robust in the MR sensitivity analyses, but discordant to the average positive association estimated from the observational studies investigating occupational sitting time and breast cancer risk. The associations with genetically predicted self-reported driving time (3-5 SNPs, 0.03%-0.08% variance) were generally null, except for a positive association with follicular lymphoma[109], and an inverse association with oesophageal cancer[107] that was not consistent with another overlapping study with a smaller number of cases[103]. For genetically predicted self-reported time of computer use or playing computer games (18-83 SNPs, 0.23%-0.89% variance), the associations were also largely null across major cancers, but an inverse association with colorectal and lung cancers was reported[107]. Other overlapping publications reported no associations[99,100,104]. These genetic instruments included small numbers of SNPs, capturing low genetic variations, leading to reduced statistical power to detect a potential association with cancer.

The studies used two sets of genetic instruments to represent time spent in watching television (56-209 SNPs, 0.46%-2% variance). The results are largely supportive of the findings in the observational studies. Positive associations were reported for genetically predicted self-reported leisure screen time and TV watching time with lung cancer[107]. The associations were robust in the sensitivity analyses and were consistently reported in overlapping publications[97,99,100,106]. Multivariable MR (MVMR) analyses showed that the association between TV watching time and lung cancer risk persisted after the adjustment for education[100], BMI[99,100], or triglyceride and total cholesterol[99], but was partially mediated by smoking (mediated proportion 51.8%)[99,100] and emotional well-being (mediated proportion 38.0%)[106]. In stratified analysis by smoking, the significant association between TV watching time and lung cancer persisted among ever smokers but not in never smokers[107]. A positive association was observed between genetically predicted TV watching time and colorectal cancer[107], but not with leisure screen time [107,108]. The positive association was robust in the sensitivity analyses and was consistent with another overlapping study[104]. MVMR analyses showed that the association attenuated after adjusting for years of education and smoking[104].

Genetically predicted self-reported leisure screen time and TV watching time were positively associated with breast cancer[107]. The association with TV watching was consistently reported[103,104]. Adjustment for years of education [103,104] and smoking[104] attenuated the association in MVMR analyses. For endometrial cancer, positive associations were observed with leisure screen time [107], and TV watching time[103], although not consistently with the other overlapping publication[107]. Sensitivity analyses showed that the results were robust for TV watching time, but in MVMR analyses, the association attenuated after adjustment for educational attainment and also for BMI and type 2 diabetes[103]. For ovarian cancer, a positive association was shown with leisure screen time that was robust in sensitivity analyses[107], but no association was reported with television watching time[103,107]. Observational studies investigating these associations, on average, found non-significant results.

Genetically predicted leisure screen time and TV watching time were not associated with oesophageal cancer[107]. The results were consistent with one overlapping publication[103] but not with another showing a positive association with leisure screen time, which was however attenuated after mutual adjustment for moderate-to-vigorous intensity physical activity in MVMR analysis[108]. For prostate cancer, one study[107] reported no association with genetically predicted leisure screen time but an inverse association with TV watching time that was not observed in two other overlapping publications[103,104]. Observational studies, on average, showed no association.

No association was reported for genetically predicted leisure screen time and pancreatic cancer [108], but a positive association for TV watching time was observed[105]. Excluding IVs associated with potential confounders, including history of pancreatitis, diabetes, BMI, alcohol and tobacco, showed similar results to the main analysis. Sensitivity analyses indicated robust findings. The results were not robust in the smaller component study[105] and not consistent with the overlapping publication of fewer cases[103]. Genetically predicted TV watching time was modestly positively associated with cervical cancer (OR per 1 SD 1.003, 95% CI: 1.001-1.005)[103], and was positively associated with Hodgkin lymphoma (OR per 1 SD 2.187, 95% CI: 1.023-4.674) and Diffuse large B-cell lymphoma (OR per 1 SD 4.048, 95% CI: 1.688-9.708)[109]. The results for cervical cancer and Hodgkin lymphoma were robust in sensitivity analyses, but showed heterogeneity (MR-PRESSO P<0.001) that could be due to outlying SNPs in the genetic instrument of TV watching for Diffuse large B-cell lymphoma[109].

No observational studies were identified investigating the same associations, except for TV watching time and pancreatic cancer and non-Hodgkin lymphoma, showing a null association[78].

No association was reported in the MR studies investigating oral cavity and pharyngeal cancer[103], stomach[108], liver/hepatobiliary tract[103,108], melanoma[103], basal cell carcinoma[102], kidney[107], bladder or urinary organs[103], glioma[103], or thyroid[103] cancers.

#### Supplementary Table 1: PRISMA checklist

| Section and Topic | Item # | Checklist item | Location where item is reported |
|---|---|---|---|
| TITLE | | | |
| Title | 1 | Identify the report as a systematic review. | Title page |
| ABSTRACT | | | |
| Abstract | 2 | See the PRISMA 2020 for Abstracts checklist. | NA |
| INTRODUCTION | | | |
| Rationale | 3 | Describe the rationale for the review in the context of existing knowledge. | Pages 5, 6 |
| Objectives | 4 | Provide an explicit statement of the objective(s) or question(s) the review addresses. | Page 6 |
| METHODS | | | |
| Eligibility criteria | 5 | Specify the inclusion and exclusion criteria for the review and how studies were grouped for the syntheses. | Page 7, Supplementary Text 2 |
| Information sources | 6 | Specify all databases, registers, websites, organisations, reference lists and other sources searched or consulted to identify studies. Specify the date when each source was last searched or consulted. | Page 7, Supplementary Text 2 |
| Search strategy | 7 | Present the full search strategies for all databases, registers and websites, including any filters and limits used. | Page 7, Supplementary Text 2 |
| Selection process | 8 | Specify the methods used to decide whether a study met the inclusion criteria of the review, including how many reviewers screened each record and each report retrieved, whether they worked independently, and if applicable, details of automation tools used in the process. | Page 7, Supplementary Text 2, Supplementary Table 2 |

| Section and Topic | Item # | Checklist item | Location where item is reported |
| --- | --- | --- | --- |
| Data collection process | 9 | Specify the methods used to collect data from reports, including how many reviewers collected data from each report, whether they worked independently, any processes for obtaining or confirming data from study investigators, and if applicable, details of automation tools used in the process. | Page 7, Supplementary Text 2 |
| Data items | 10a | List and define all outcomes for which data were sought. Specify whether all results that were compatible with each outcome domain in each study were sought (e.g. for all measures, time points, analyses), and if not, the methods used to decide which results to collect. | Page 7, Supplementary Text 2 |
| | 10b | List and define all other variables for which data were sought (e.g. participant and intervention characteristics, funding sources). Describe any assumptions made about any missing or unclear information. | Page 7, Supplementary Text 2, Supplementary Tables 2, 4 |
| Study risk of bias assessment | 11 | Specify the methods used to assess risk of bias in the included studies, including details of the tool(s) used, how many reviewers assessed each study and whether they worked independently, and if applicable, details of automation tools used in the process. | Page 8, Supplementary Text 2, Supplementary Tables 3, 4 |
| Effect measures | 12 | Specify for each outcome the effect measure(s) (e.g. risk ratio, mean difference) used in the synthesis or presentation of results. | Page 8, Supplementary Text 2 |
| Synthesis methods | 13a | Describe the processes used to decide which studies were eligible for each synthesis (e.g. tabulating the study intervention characteristics and comparing against the planned groups for each synthesis (item #5)). | Pages 8, 9, Supplementary Text 2 |
| | 13b | Describe any methods required to prepare the data for presentation or synthesis, such as handling of missing summary statistics, or data conversions. | Pages 8, 9, Supplementary Text 2 |
| | 13c | Describe any methods used to tabulate or visually display results of individual studies and syntheses. | Pages 8, 9, Supplementary Text 2 |
| | 13d | Describe any methods used to synthesize results and provide a rationale for the choice(s). If meta-analysis was performed, describe the model(s), method(s) to identify the presence and extent of statistical heterogeneity, and software package(s) used. | Pages 8, 9, Supplementary Text 2 |

| Section and Topic | Item # | Checklist item | Location where item is reported |
|---|---|---|---|
| | 13e | Describe any methods used to explore possible causes of heterogeneity among study results (e.g. subgroup analysis, meta-regression). | Pages 8, 9, Supplementary Text 2 |
| | 13f | Describe any sensitivity analyses conducted to assess robustness of the synthesized results. | Pages 8, 9, Supplementary Text 2 |
| Reporting bias assessment | 14 | Describe any methods used to assess risk of bias due to missing results in a synthesis (arising from reporting biases). | Pages 8, 9, Supplementary Text 2 |
| Certainty assessment | 15 | Describe any methods used to assess certainty (or confidence) in the body of evidence for an outcome. | Page 9, Supplementary Table 5 |
| RESULTS | | | |
| Study selection | 16a | Describe the results of the search and selection process, from the number of records identified in the search to the number of studies included in the review, ideally using a flow diagram. | Page 10, Supplementary Text 3, Figure 1 |
| | 16b | Cite studies that might appear to meet the inclusion criteria, but which were excluded, and explain why they were excluded. | Page 10, Supplementary Text 3, Supplementary Tables 6, 7 |
| Study characteristics | 17 | Cite each included study and present its characteristics. | Pages 10, 11, Supplementary Text 3, Supplementary Tables 8-10, 15-17 |
| Risk of bias in studies | 18 | Present assessments of risk of bias for each included study. | Page 13, Supplementary Text 4, Supplementary Figure 122 |

| Section and Topic | Item # | Checklist item | Location where item is reported |
|---|---|---|---|
| Results of individual studies | 19 | For all outcomes, present, for each study: (a) summary statistics for each group (where appropriate) and (b) an effect estimate and its precision (e.g. confidence/credible interval), ideally using structured tables or plots. | Supplementary Tables 11, 12, 15-18 Supplementary Figures 8-20 |
| Results of syntheses | 20a | For each synthesis, briefly summarise the characteristics and risk of bias among contributing studies. | Pages 10-13, Supplementary Text 3-5 |
| | 20b | Present results of all statistical syntheses conducted. If meta-analysis was done, present for each the summary estimate and its precision (e.g. confidence/credible interval) and measures of statistical heterogeneity. If comparing groups, describe the direction of the effect. | Pages 10-13, Figure 2, Supplementary Text 3-5, Supplementary Figures 1-7 |
| | 20c | Present results of all investigations of possible causes of heterogeneity among study results. | Pages 10-13, Supplementary Text 3-5, Supplementary Figures 21-121 |
| | 20d | Present results of all sensitivity analyses conducted to assess the robustness of the synthesized results. | Pages 10-13, Supplementary Text 3-5, Supplementary Tables 14,15, Supplementary Figures 21-121 |
| Reporting biases | 21 | Present assessments of risk of bias due to missing results (arising from reporting biases) for each synthesis assessed. | Pages 10-13, Supplementary Text 3-5, Supplementary Figures 21-121 |
| Certainty of evidence | 22 | Present assessments of certainty (or confidence) in the body of evidence for each outcome assessed. | Pages 14, 15, Figure 1 |

| Section and Topic | Item # | Checklist item | Location where item is reported |
|---|---|---|---|
| DISCUSSION | | | |
| Discussion | 23a | Provide a general interpretation of the results in the context of other evidence. | Pages 15-19 |
| | 23b | Discuss any limitations of the evidence included in the review. | Pages 15-19 |
| | 23c | Discuss any limitations of the review processes used. | Pages 15-19 |
| | 23d | Discuss implications of the results for practice, policy, and future research. | Pages 18-19 |
| OTHER INFORMATION | | | |
| Registration and protocol | 24a | Provide registration information for the review, including register name and registration number, or state that the review was not registered. | Page 6 |
| | 24b | Indicate where the review protocol can be accessed, or state that a protocol was not prepared. | Page 6 |
| | 24c | Describe and explain any amendments to information provided at registration or in the protocol. | NA |
| Support | 25 | Describe sources of financial or non-financial support for the review, and the role of the funders or sponsors in the review. | Page 9, Page 23 |
| Competing interests | 26 | Declare any competing interests of review authors. | Page 23 |
| Availability of data, code and other materials | 27 | Report which of the following are publicly available and where they can be found: template data collection forms; data extracted from included studies; data used for all analyses; analytic code; any other materials used in the review. | Page 24 |

#### Supplementary Table 2: Eligibility criteria for cancer outcomes commonly investigated in the epidemiological studies [a]

| Cancer sites (incidence or mortality) | Inclusion criteria | Exclusion criteria |
|---|---|---|
| Bladder cancer | Bladder cancer; transitional cell carcinoma (urothelial carcinoma), squamous cell carcinoma, adenocarcinoma; histologically defined carcinoma in situ; and imprecise anatomical definitions of cancer sites that include bladder cancer, such as urological tract cancer, provided that they are of invasive carcinoma. | Pre-malignant bladder cancer other than histologically defined carcinoma in situ. |
| Brain and other central nervous system cancers | Brain cancer; glioma, glioblastoma; grade III meningioma; spinal cord cancer; brain combined with meninges and spinal cord cancers. | Benign brain tumours; benign spinal cord tumours; metastatic spinal cancer. |
| Breast cancer | Invasive breast cancer; premenopausal or postmenopausal breast cancer; any molecular, genetic, or histopathological subtypes. | In situ breast cancer |
| Cervical cancer | Invasive or microinvasive cervical cancer; squamous cell carcinoma (SCC), epidermoid carcinoma (large cell or small cell, keratinizing or nonkeratinizing), or adenocarcinoma; cervical intraepithelial neoplasia III (CIN 3)/carcinoma in situ (CIS). | Pre-invasive neoplastic lesions (cervical dysplasia, cervical intraepithelial neoplasia (CIN I or II), squamous intraepithelial lesion (SIL, different grades). |
| Colorectal cancer | Colorectal cancer; (distal, proximal) colon cancer, rectal cancer; any molecular, genetic, or histopathological subtypes; Lynch syndrome colorectal cancer, early onset colorectal cancer. | Pre-malignant colorectal conditions, e.g. colorectal adenomas or polyps |
| Endometrial cancer | Endometrial cancer; type I endometrial cancer (endometrioid adenocarcinoma), type II endometrial cancer (serous, clear cell, carcinosarcoma and undifferentiated/dedifferentiated cancers) | ---- |
| Gallbladder cancer | Gallbladder cancer; imprecise anatomical definitions of cancer sites that include gallbladder cancer, such as biliary tract cancer. | ---- |
| Kidney cancer | Kidney cancer; (clear cell, non-clear cell) renal cell carcinoma (RCC), renal parenchyma cancer; renal pelvis cancer, transitional cell carcinoma (TCC) (urothelial carcinoma). | Wilm’s tumour (more common in children) or cancer related to schistosomiasis infection. |
| Liver cancer | Liver cancer; hepatocellular carcinoma (HCC). | ---- |
| Lung cancer | Lung cancer; non-small cell lung cancer (NSCLC), adenocarcinoma, squamous cell carcinoma, large cell carcinoma; small cell lung cancer (SCLC). | ---- |

| Cancer sites (incidence or mortality) | Inclusion criteria | Exclusion criteria |
|---|---|---|
| Lymphoid and hematopoietic cancers | Leukaemia; acute lymphoblastic/lymphocytic leukaemia (ALL), acute myeloid leukaemia (AML), chronic lymphocytic leukaemia (CLL)[b], chronic myeloid leukaemia (CML), chronic myelomonocytic leukaemia (CMML).<br><br>Lymphoma; non-Hodgkin lymphoma, Hodgkin lymphoma, diffuse large B-cell lymphoma, follicular lymphoma, small lymphocytic lymphoma (SLL)[b]<br><br>Multiple myeloma. | ---- |
| Mouth, pharyngeal, and laryngeal cancer | Cancer of the mouth, pharynx or larynx as separate outcomes, as combinations of these cancers, or for all these cancers combined; head and neck cancer; upper aerodigestive cancer; and other cancer groups that explicitly include mouth, pharynx, or larynx cancers. | ---- |
| Nasopharyngeal cancer | Nasopharyngeal cancer. | Cancer combinations that include nasopharyngeal cancer. |
| Oesophageal cancer | Oesophageal cancer; gastro-oesophageal cancer; squamous cell carcinoma, adenocarcinoma. | Upper aerodigestive cancer that includes oesophageal cancer that is evaluated under mouth, pharyngeal, and laryngeal cancers. |
| Ovarian cancer | Ovarian cancer; epithelial ovarian carcinoma; serous carcinomas. | ---- |
| Pancreatic cancer | Pancreatic cancer. | ---- |
| Prostate cancer | Prostate cancer; advanced/aggressive prostate cancer (see (a)-(h) below), early localised/low grade prostate cancer;<br>imprecise anatomical definitions of cancer sites that include prostate cancer, such as urogenital cancer.<br><br>(a) stage 3-4 on the AJCC 1992 classification<br>(b) advanced cancer<br>(c) advanced or metastatic cancer<br>(d) metastatic cancer | Pre-malignant prostate conditions, e.g. high-grade prostate intra epithelial neoplasia. |

| Cancer sites (incidence or mortality) | Inclusion criteria | Exclusion criteria |
|---|---|---|
| | (e) stage C or D on the Whitemore/Jewertt scale<br>(f) fatal cancer<br>(g) high stage or grade<br>(h) Gleason grade >= 7 | |
| Skin cancer | Skin cancer, including basal cell carcinoma (BCC), basal cell epithelioma; squamous cell carcinoma (SCC) of skin, squamous cell epithelioma; (invasive or in situ) melanoma, cutaneous melanoma; non-melanoma skin cancer (NMSC). | Skin cancer in patients with AIDS (e.g. Kaposi's sarcoma and AIDS). |
| Small intestine cancer | Small intestine cancer. | Small intestine polyps. |
| Stomach cancer | Gastric (stomach) cancer; cardia or non-cardia gastric cancers. | Anatomical definitions of cancer sites that include gastric cancer, e.g. gastrointestinal cancer; gastro-oesophageal cancer that is evaluated under oesophageal cancer. |
| Thyroid cancer | Thyroid cancer; differentiated thyroid cancer (papillary, follicular, and oncocytic carcinoma). | ---- |

[a] All listed cancers were reviewed in the TER, apart from brain and other central nervous system cancers, lymphoid and hematopoietic cancers, small intestine cancer, and thyroid cancer. The list is not meant to be exhaustive. Any additional cancers or cancer subtypes identified in eligible studies during the literature search will be included in the present SLRs.

[b] CLL is the same disease as SLL.

#### Supplementary Table 3: Risk of Bias for Nutrition Observational Studies (RoB-NObs) tool for evaluation of cancer incidence studies (modified version dated 29/01/2025).

Assess for each exposure-outcome association for publications included in the meta-analyses or narrative reviews.

Some sections can be assessed for the overall publication (1, 2, 5, 7) but exposure and outcome-related domains should be assessed separately (3, 4, 6). We consider lumping together similar exposures (e.g., dietary exposures) or outcomes (e.g., mortality outcomes).

The levels of judgement are low, moderate, serious, critical, and no information for each of the seven domains. There is no overall risk of bias.

Abbreviations:
Y: Yes
PY: Probably yes
N: No
PN: Probably no
NI: No information

##### Bias due to confounding.

| Question | Issues/ Remarks | Prompting questions/ indicators |
|---|---|---|
| 1.1 Is there potential for confounding of the effect of exposure in this study?<br>**If N or PN**: skip all remaining questions (1.2 to 1.8) and go to Bias due to confounding: Risk of bias judgement; the study can be considered to be at low risk of bias due to confounding and no further signalling questions need be considered.<br>**If Y or PY**: answer question 1.2 to determine whether there is a need to assess time-varying confounding. | Issues:<br>- Confounding cannot be ruled out in observational studies. | This question should be always answered as **PY** in observational studies of the areas we work on. |
| 1.2. **If Y or PY to 1.1**: Was the analysis based on splitting follow-up time according to exposure received?<br>**If N or PN:** skip 1.3 and answer questions 1.4 to 1.6, which relate to baseline confounding. If Y or PY, go to question 1.3. | Issues:<br>- Exposures may change across time and are subject to time-varying confounding.<br>Remarks:<br>- This question asks whether a time-varying exposure analysis was conducted (which | Consider whether the study has evaluated the exposure at multiple time points. This question should be always answered as either **Y** or **N.**<br><br>**-Y** The study has evaluated the exposure at multiple time points and the investigators have |

| Question | Issues/ Remarks | Prompting questions/ indicators |
|---|---|---|
| | also requires to adjust for time-varying confounders). | performed time-varying exposure/ confounder analyses.<br>**-N** The study has not evaluated the exposure at multiple time points OR the investigators have not performed time-varying exposure/ confounder analyses. |
| 1.3. **If Y or PY to 1.2**: Were exposure discontinuations or switches likely to be related to factors that are prognostic for the outcome?<br>**If N or PN:** answer questions 1.4 to 1.6, which relate to baseline confounding only. Do not answer 1.7 and 1.8, which relate to both baseline and time-varying confounding.<br>**If Y or PY**: skip questions 1.4 to 1.6, and answer questions 1.7 and 1.8, which relate to both baseline confounding and time-varying confounding. | Issues:<br>- How to tell whether the changes are related to prognostic factors for the outcome?<br>Remarks:<br>- If exposure switches are unrelated to the outcome, for example when the outcome is an unexpected harm, then time-varying confounding will not be present and only control for baseline confounding is required.<br>- This is unlikely on the areas and outcomes we work on. | This question should be always answered as **PY** in observational studies of the areas we work on. |
| Questions relating to baseline confounding | | |
| 1.4. **If N or PN to 1.2 or 1.3**: Did the authors use an appropriate analysis method that adjusted for all the critically important confounding variables at baseline?<br>Go to 1.5 | Issues:<br>- Which method of adjustments can be considered as adequate?<br>Remarks:<br>- Any method to adjust for measured confounders should be acceptable e.g., stratification, regression, matching, restriction, standardization, and inverse probability weighting, multivariable analysis.<br>- Consider the association as "adjusted" when the required factors (as they appear in the Prompting questions/ indicators) were used in matching between those with or without a study outcome or stratification in analysis. | **-N** Not adjusted at least for all the required confounding variables.<br>**-PY** Adjusted at least for all the required confounders.<br>**-Y** Adjusted at least for all the required plus the desired confounding variables.<br>Please consult the external list of important confounding variables below. |

| Question | Issues/ Remarks | Prompting questions/ indicators |
|---|---|---|
| | - Consider the association as “adjusted” when the required factors (as they appear in the Prompting questions/ indicators) were tested but not included in the final statistical model due to lack of statistical significance or small change in risk estimate.<br>E.g., age was tested univariably and was found to be not significant or the estimates did not change by more than 10%. | |
| 1.5. Were confounders that were adjusted for measured validly and reliably by the variables available in this study?<br>Go to 1.6 | Issues:<br>- Residual confounding is possible if covariates are not measured validly and reliably.<br>Remarks:<br>- For some topics, a list of valid and reliable measures of confounding domains will be specified in the review protocol but for others such a list may not be available.<br>- Study authors may cite references to support the use of a particular measure.<br>- If authors control for confounding variables with no indication of their validity or reliability pay attention to the subjectivity of the measure.<br>- Subjective measures (e.g. based on self-report) may have lower validity and reliability than objective measures such as lab findings. | -**Y** Adiposity (any index or measure) was measured. Dietary factors and physical activity were self-reported but based on a validated questionnaire, or their assessment was validated against another assessment method (within the same study or externally in another study reported in the paper). Comorbidities (if relevant) were ascertained by a physician, medical records, registries.<br>-**PY** Adiposity (any index or measure) was self-reported. Dietary factors and physical activity were self-reported but based on a validated questionnaire, or their assessment was validated against another assessment method (within the same study or externally in another study reported in the paper). Comorbidities (if relevant) were self-reported but further ascertained by a physician, medical records, registries.<br>-**N/ PN** Adiposity (any index or measure) was self-reported. Dietary factors and physical activity were self-reported but were not based on a validated questionnaire, or their assessment was not validated against another assessment method. Comorbidities (if relevant) are self-reported and not further ascertained.<br>-**NI** Information was not reported in detail/ separately for each critical confounder |

| Question | Issues/ Remarks | Prompting questions/ indicators |
|---|---|---|
| | | When two or more different confounders lead to different judgement then go with the worst-case scenario. |
| 1.6. Did the authors avoid adjusting for post-exposure variables?<br><br>Skip to Bias due to confounding: Risk of bias judgement. | Issues:<br>- Controlling for post-exposure variables that are affected by the exposure is not appropriate.<br>- Controlling for mediating variables estimates the direct effect of the exposure and may introduce bias.<br>- Controlling for common effects of the exposure and outcome introduces bias.<br>Remarks:<br>- Confounders should be measured at or before (ideally) the time that exposure was assessed and not after (a few days/ weeks is ok but definitively not longer). | This question should be always answered as either **PY** or **PN**.<br><br>**-PY** The study has not adjusted for post-exposure variables affected by the exposure, mediators, colliders (common effects of the exposure and outcome) AND the adjustment variables should be measured at or before the time of exposure assessment.<br>**-PN** All other cases. |
| Questions relating to baseline and time-varying confounding | | |
| 1.7. **If Y or PY to 1.3**: Did the authors use an appropriate analysis method that adjusted for all the critically important confounding variables, including baseline and time-varying confounding?<br>**If N or PN to 1.7**: skip to Bias due to confounding: Risk of bias judgement.<br>**If Y or PY to 1.7**, answer question 1.8. | Issues:<br>- Exposures and confounders may change across time.<br>Remarks:<br>- Any method to adjust for baseline and time-varying confounding should be considered appropriate (e.g., regular regression, methods based on inverse probability weighting, marginal structural models). | Please consult the external list of important confounding variables.<br>Time-fixed confounders<br>**-PY** Age, sex (if relevant), race (if relevant), reproductive factors (if relevant), comorbidities (if relevant).<br>Time-varying confounders<br>**-Y** Adiposity, smoking, dietary factors, physical activity, comorbidities (if relevant).<br>**-PN** All other cases. |
| 1.8 **If Y or PY to 1.3 and Y or PY to 1.7**: Were confounders that were adjusted for measured validly and reliably by the variables available in this study? | | **-Y** Adiposity (any index or measure) was measured. Dietary factors and physical activity were self-reported but based on a validated questionnaire, or their assessment was validated against another assessment method (within the same study or externally in another study reported in the paper). |

| Question | Issues/ Remarks | Prompting questions/ indicators |
|---|---|---|
| Skip to Bias due to confounding: Risk of bias judgement. | | Comorbidities (if relevant) were ascertained by a physician, medical records, registries.<br>**-PY** Adiposity (any index or measure) was self-reported. Dietary factors and physical activity were self-reported but based on a validated questionnaire, or their assessment was validated against another assessment method (within the same study or externally in another study reported in the paper). Comorbidities (if relevant) were self-reported but further ascertained by a physician, medical records, registries.<br>**-N/ PN** Adiposity (any index or measure) was self-reported. Dietary factors and physical activity were self-reported but were not based on a validated questionnaire, or their assessment was not validated against another assessment method. Comorbidities (if relevant) are self-reported and not further ascertained.<br>**-NI** Information was not reported in detail/ separately for each critical confounder<br>When two or more different confounders lead to different judgement then go with the worst-case scenario. |
| RoB judgement | | |
| Low risk of bias<br>The study is comparable to a well-performed randomized trial with regard to this domain. | No confounding expected. Observational studies should never have low RoB in this domain. | |
| Moderate risk of bias<br>The study is sound for an observational study with regard to this domain but cannot be considered comparable to a well-performed randomized trial. | (i) Confounding expected, all known important confounding domains appropriately measured and controlled for;<br>and<br>(ii) Reliability and validity of measurement of important domains were sufficient, such that we do not expect serious residual confounding. | |
| Serious risk of bias<br>The study has some important problems. | (i) At least one key confounder was not appropriately measured, or not controlled for;<br>or | |

| Question | Issues/ Remarks | Prompting questions/ indicators |
|---|---|---|
| | (ii) Reliability or validity of measurement of a key confounder **could not be evaluated or** was low enough that we expect serious residual confounding. | |
| Critical risk of bias<br>The study is too problematic to provide any useful evidence. | (i) Confounding is inherently not controllable;<br>or<br>(ii) The use of negative controls strongly suggests unmeasured confounding. | |

#### Bias in selection of participants into the study.

| Question | Issues/ Remarks | Prompting questions/ indicators |
|---|---|---|
| 2.1. Was selection of participants into the study or into the analysis based on participant characteristics observed after the start of the exposure assessment?<br>If **N or PN:** go to 2.4 (skip 2.2 and 2.3).<br>If **Y or PY**: go to 2.2 and 2.3. | Issues:<br>- Systematic difference in risk factors and risk of the outcome between study participants and non-participants. Can arise from the initial self-selection of study participants into the study. | Expect to answer this question almost always as **Y/ PY**.<br><br>**-N/ PN** Study population was a random sample of the target population or was based on a national registry.<br>**-Y/ PY** If participant contact or consent is required, selection bias may occur. |
| 2.2. **If Y or PY to 2.1**: Were the post-exposure variables that influenced selection of participants (into the study or analysis) associated with exposure?<br>Go to 2.3 | Issues:<br>- Post-exposure variables may influence study selection which can lead to unrepresentative exposure groups.<br>Remarks:<br>- The focus of this question should only be on the exposure and not on other variables.<br>- Consider whether an analysis (by exposure status) comparing participants and non-participants has been performed.<br>- Consider whether the characteristics (age, sex [when appropriate], BMI or other exposures e.g., diet, physical activity [when appropriate]) between the participants and non-participants at baseline (at the time of exposure) are comparable. | **-Y** An analysis has been performed and there is a difference (10% or more) in the exposure value/ rate between the groups.<br>**-PY** An analysis has not been performed.<br>**-PY** An analysis has been performed and there is a difference (5-10%) in the exposure value/ rate between the groups.<br>**-PN** An analysis has been performed and there is a difference (<5%) in the exposure value/ rate between the groups.<br>**-N** An analysis has been performed and there is no difference in the exposure value/ rate between the groups OR there is a statement in the manuscript that potential selection of participants was not associated with exposure. |

| Question | Issues/ Remarks | Prompting questions/ indicators |
|---|---|---|
| | - Keep in mind that sometimes studies compare baseline characteristics of included vs. excluded individuals based on a certain exclusion criterion, but we need to evaluate here whether they have also performed something similar for individuals that never participated [volunteered] in the study. | |
| 2.3. **If Y or PY to 2.1**: Were the post-exposure variables that influenced selection of participants (into the study or analysis) associated with the outcome?<br>Go to 2.4 | Issues:<br>- Post-exposure variables may influence study selection leading to unrepresentative outcome ascertainment.<br>- In cohort studies (of cancer incidence), any self-selection of participants should not be associated with the outcome that occurs years later. | *Expect to answer this question almost always as **N/PN** when cohort studies are evaluated.* |
| 2.4. Do start of follow-up and start of exposure assessment coincide for most participants?<br>If **N or PN**: answer 2.5.<br>If **Y or PY**: go to Bias in selection of participants into the study: Risk of bias judgement. | Remarks:<br>- The time of follow-up start should coincide with the time of exposure assessment. | **-Y** Follow-up for outcome assessment started at the time of exposure assessment (or within 3 months maximum).<br>**-PN** The study has mentioned that the start of follow-up for outcome assessment and the time of exposure assessment did not coincide (the exposure was assessed after the start of follow-up) but there was no information on the timing between the two measurements.<br>**-N** The start of follow-up for outcome assessment only started after the time of exposure assessment (more than 3 months).<br>**-NI** The study has not mentioned anything about the start of follow-up for outcome assessment and the time of exposure assessment. |
| 2.5 **If Y or PY to 2.2 and 2.3, or N or PN to 2.4**: Were adjustment techniques that were likely to correct for the presence of selection biases used? | Remarks:<br>- It is in principle possible to correct for selection biases, for example by using inverse | **-Y/ PY** Such an analysis was performed.<br>**-N/ PN** Such an analysis was not performed. |

| **Question** | **Issues/ Remarks** | **Prompting questions/ indicators** |
|---|---|---|
| Go to Bias in selection of participants into the study: Risk of bias judgement. | probability weights to create a pseudo-population in which the selection bias has been removed, or by modelling the distributions of the missing participants or follow up times and outcome events and including them using missing data methodology.<br>- Check whether any adjustment techniques (e.g., inverse probability weighting, g-formula, marginal structural models) were used for accounting for selection bias either as a main analysis or a sensitivity analysis. | |
| RoB judgement | | |
| Low risk of bias<br>The study is comparable to a well-performed randomized trial with regard to this domain. | (i) All participants who would have been eligible for inclusion, were included in the study;<br>and<br>(ii) For each participant, start of follow up and start of exposure coincided. | |
| Moderate risk of bias<br>The study is sound for an observational study with regard to this domain but cannot be considered comparable to a well-performed randomized trial. | (i) All participants who would have been eligible for inclusion, were included in the study;<br>and<br>There was no information on whether start of follow up and start of exposure coincided for all participants.<br>or<br>(ii) Selection into the study may have been related to exposure and outcome;<br>and<br>The authors used appropriate methods to adjust for the selection bias;<br>or<br>(iii) Start of follow up and start of exposure do not coincide for all participants;<br>and<br>(a) the proportion of participants for which this was the case was too low to induce important bias;<br>or<br>(b) the authors used appropriate methods to adjust for the selection bias;<br>or<br>(c) the review authors are confident that the rate (hazard) ratio for the effect of exposure remains constant over time. | |

| Question | Issues/ Remarks | Prompting questions/ indicators |
|---|---|---|
| Serious risk of bias<br>The study has some important problems. | (i) Selection into the study was related (but not very strongly) to exposure and outcome;<br>and<br>This could not be adjusted for in analyses;<br>or<br>(ii) Start of follow up and start of exposure do not coincide;<br>and<br>A potentially important amount of follow-up time is missing from analyses;<br>and<br>The rate ratio is not constant over time | |
| Critical risk of bias<br>The study is too problematic to provide any useful evidence. | (i) Selection into the study was very strongly related to exposure and outcome;<br>and<br>This could not be adjusted for in analyses;<br>or<br>(ii) A substantial amount of follow-up time is likely to be missing from analyses;<br>and<br>The rate ratio is not constant over time. | |

#### Bias in classification of exposures.

| Question | Issues/ Remarks | Prompting questions/ indicators |
|---|---|---|
| 3.1. Is the exposure that was assessed clearly defined? | Issues:<br>- Ambiguity in the definition may lead to bias in the classification of participants.<br>Remarks:<br>- Example where exposure may not be clearly defined may be e.g., a study has an unclear definition of dietary and supplement intake of vitamins and ends up with a mixture of info for the different participants in the study. | Expect to answer this question most often as **Y/ PY**.<br><br>-**Y/ PY** The study provided details for the definition of the exposure.<br>-**N/ PN** The study did not provide sufficient details for the definition of the exposure.<br>E.g., The study aim is to assess vitamin D, but did not define what is considered as vitamin D. |
| 3.2. Does the exposure that was assessed represent the exposure of interest? | | Expect to answer this question most often as **Y/ PY**.<br><br>-**Y/ PY** The assessed exposure directly represents the exposure of interest. |

| Question | Issues/ Remarks | Prompting questions/ indicators |
|---|---|---|
| | | **-N/ PN** The assessed exposure does not directly represent the exposure of interest.<br>E.g., The study aim is to assess total physical activity, but data are only on recreational physical activity. |
| 3.3. Were the methods used to assess the exposure clearly described? | | Expect to answer this question most often as **Y/ PY**.<br><br>**-Y/ PY** The study provided details on the assessment methods of the exposure of interest.<br>**-N/ PN** The study did not provide sufficient details on the assessment methods of the exposure of interest. |
| 3.4. Were the methods used to measure the exposure valid and/ or reliable? | | **-Y** Assessment was based on any objective measure of assessing the exposure (measured adiposity, measured physical activity, measured sedentary behaviour, diet biomarkers).<br>**-Y** Exposure was self-reported and assessment was validated against another exposure assessment method, in whole or subset of study populations AND replicated with the same exposure method at a different time point.<br>**-PY** Assessment was based on self-reported weight/ height/ BMI.<br>**-PY** Exposure was self-reported, and assessment was validated against another exposure assessment method, in whole or subset of study populations OR replicated with the same exposure method at a different time point.<br>**-PY** Exposure was self-reported, and assessment was validated against another exposure assessment method AND replicated with the same exposure method at a different time point, but these were performed in another study cohort (also check discussion section).<br>**-PN** Exposure was self-reported, and assessment was validated against another exposure assessment method OR replicated with the same exposure method at a different time point, but these were |

| Question | Issues/ Remarks | Prompting questions/ indicators |
|---|---|---|
| | | performed in another study cohort (also check discussion section).<br>**-PN** Assessment was based on retrospective assessment or chart review of exposures.<br>**-N** Exposure was self-reported and assessment was not validated against another exposure assessment method OR not replicated with the same exposure method at a different time point.<br>**-NI** No data on validation AND replication of the self-reported measurement. |
| 3.5. Were the same methods used to assess the exposure status for all participants/ groups? | | **-PY/ Y** Assessment method and timeframe were the same regardless of exposure status.<br>**-PN/ N** Assessment method and timeframe differed in terms of exposure status. |
| 3.6. Were the methods used to define exposure status for participants/ groups clearly described? | Remarks:<br>- A study may use different definitions of exposure, an exposure may be measured based on a score, or a continuously measured exposure may be used as categories. The definition of the variable used for the exposure should be clearly defined. | **-Y/ PY** The study provided details on how the exposure status was defined.<br>**-N/ PN** The study did not provide sufficient details on how the exposure status was defined. |
| 3.7. Were the methods used to define exposure status for participants/ groups likely to result in minimal random or systematic exposure misclassification? | Remarks:<br>- Misclassification exists in self-reported diet, adiposity, and physical activity assessments.<br>- The question is whether this misclassification is different in those with high vs. low levels of exposure, which might be the case.<br>- It is expected to see different statistics evaluating validity/ reliability (correlation coefficient, sensitivity, specificity, kappa, etc.).<br>- Please use common sense (or check for established thresholds for the specific exposure) about what an acceptable level for the specific statistic is, but as guidance: a correlation | **-Y** Exposure assessment is based on objective measures (measured adiposity, measured physical activity, diet biomarkers) or based on self-reported anthropometric factors AND a validity/ reliability statistic is presented and is at an acceptable level.<br>**-PY** Exposure assessment is based on objective measures (measured adiposity, measured physical activity, diet biomarkers) or based on self-reported anthropometric factors AND a validity/ reliability statistic is not presented.<br>**-PY** Exposure assessment is based on any self-reported measure (except for anthropometric factors) AND a validity/ reliability statistic is presented and is at an acceptable level |

| Question | Issues/ Remarks | Prompting questions/ indicators |
|---|---|---|
| | coefficient of 0.4 or larger can be considered acceptable for nutritional exposures but not acceptable for BMI for which we would expect a correlation coefficient of 0.7 or larger. | **-PN** Exposure assessment is based on any self-reported measure (except for anthropometric factors) AND a validity/ reliability statistic is not presented.<br>**-N** A validity/ reliability statistic is presented, either for objective or the self-reported measures and is not at an acceptable level. |
| 3.8. Could classification of exposure status been affected by the presence of the outcome, knowledge of the outcome or risk of the outcome?<br>**If Y or PY**, there may be serious risk of bias. Go to Bias in classification of exposures: Risk of bias judgement. | Issues:<br>- There is the possibility that the exposure of interest may be influenced by undiagnosed disease, thus resulting in misclassification. | **-Y/ PY** The study included participants for which assessment of the exposure was performed at least 1 or 2 years before diagnosis and there was no mention about performing some type of lag analysis (e.g., the study excluded events that happen within 1 or 2 years of exposure assessment in a sensitivity analysis).<br>**-N/ PN** All other cases. |
| RoB judgement | | |
| Low risk of bias<br>The study is comparable to a well-performed randomized trial with regard to this domain. | (i) The exposure and the methods used to assess the exposure were well defined and represent the exposure of interest;<br>and<br>(ii) Methods were valid, reliable, the same across groups, and likely to result in minimal random or systematic exposure misclassification;<br>and<br>(iii) Exposure status was not affected by the presence of the outcome, knowledge of the outcome or risk of the outcome. | |
| Moderate risk of bias<br>The study is sound for an observational study with regard to this domain but cannot be considered comparable to a well-performed randomized trial. | (i) The exposure and the methods used to assess the exposure are defined and represent the exposure of interest;<br>and<br>(ii) Methods were valid, reliable, the same across groups, and likely to result in minimal random or systematic exposure misclassification;<br>or<br>Exposure status was not affected by the presence of the outcome, knowledge of the outcome or risk of the outcome. | |
| Serious risk of bias<br>The study has some important problems. | (i) Exposure status or the methods used to assess the exposure are not well defined or do not represent the exposure of interest;<br>and | |

| Question | Issues/ Remarks | Prompting questions/ indicators |
|---|---|---|
| | (ii) Methods were not valid and reliable, the same across groups, or were likely to result in some degree of random or systematic exposure misclassification;<br>or<br>Exposure status was affected by the presence of the outcome, knowledge of the outcome or risk of the outcome. | |
| Critical risk of bias<br>The study is too problematic to provide any useful evidence. | (i) Exposure status and the methods used to assess the exposure are not well defined or do not represent the exposure of interest;<br>and<br>(ii) Methods were not valid and reliable, were not the same across groups, and were likely to result in substantial random or systematic exposure misclassification;<br>and<br>(iii) Exposure status was affected by the presence of the outcome, knowledge of the outcome or risk of the outcome. | |

#### Bias due to departures from intended exposures.

| Question | Issues/ Remarks | Prompting questions/ indicators |
|---|---|---|
| 4.1. Is there concern that changes in exposure status occurred among participants that were unbalanced across groups and likely to impact the outcome? | Issues:<br>- Changes of exposure status may happen after baseline exposure assessment | This question should be always answered as **PY** in observational studies of the areas we work on. |
| 4.2. Were any critical co-exposures that occurred unbalanced between exposure groups and likely to impact the outcome? | | This question should be always answered as **PY** in observational studies of the areas we work on. |
| 4.3. **If Y or PY to 4.1, or 4.2**: Were adjustment techniques that are likely to correct for these issues (i.e., changes in exposure status and/ or unbalanced co-exposures) used?<br><br>Go to Bias due to departures from intended exposures: Risk of bias judgement. | Remarks:<br>- Check whether there were repeated exposure assessments and whether these were used in time-varying models.<br>- Check whether there was any type of sensitivity analysis. | **-Y** The study has measured exposure and important confounders more than once and performed a time-varying exposure (and confounder) analysis.<br>**-PY** The study has measured exposure more than once in a subset of the cohort and either performed a time-varying exposure (and confounder) analysis in that subset or estimated a correlation |

| Question | Issues/ Remarks | Prompting questions/ indicators |
| --- | --- | --- |
| | E.g., the study calculated an intraclass correlation coefficient (ICC) of exposure assessment with time and the ICC is high. | between the several exposure (and confounder) measures and the correlation was high.<br>**-PN** The study has measured only exposure more than once and either performed a time-varying exposure but not confounder analysis in the total sample or in a subset or estimated a correlation between the exposure measures and the correlation was adequate.<br>**-N** The study has not performed a time-varying exposure analysis. |
| RoB judgement | | |
| Low risk of bias<br>The study is comparable to a well-performed randomized trial with regard to this domain. | There were no changes in the exposure status that were likely to impact the outcome, and any important co-exposures were balanced across exposure groups. ***No study should have low RoB in this domain.*** | |
| Moderate risk of bias<br>The study is sound for an observational study with regard to this domain but cannot be considered comparable to a well-performed randomized trial. | (i) There were changes in exposures status or important coexposures were not balanced across groups; and<br>(ii) the impact on the outcome is expected to be slight or measurement and/or adjustment techniques were used to correct for the issues. | |
| Serious risk of bias<br>The study has some important problems. | (i) There were changes in exposure status or important co-exposures were not balanced across groups that were likely to impact the outcome;<br>and<br>(ii) No or inappropriate measurement and/or adjustment techniques were used to correct for the issues. | |
| Critical risk of bias<br>The study is too problematic to provide any useful evidence. | (i) There were substantial changes in exposures status, or important co-exposures were not balanced across groups, that were likely to impact the outcome;<br>and<br>(ii) No or inappropriate measurement and/or adjustment techniques were used to correct for the issues. | |

#### Bias due to missing data.

| Question | Issues/ Remarks | Prompting questions/ indicators |
| --- | --- | --- |
| 5.1. Were there missing outcome data? | Issues: | **-N/PN** The participants were completely followed-up and assessed for outcomes or the percent of missingness was <10%. |

| Question | Issues/ Remarks | Prompting questions/ indicators |
|---|---|---|
| | - Missing outcome measurements due to loss to follow-up or any missed outcome assessment may lead to bias in the results. | -**N/PN** No mention of missingness but outcomes were ascertained via record linkages.<br>-**Y/PY** ≥10% of the participants missing an outcome.<br>**-NI** There was no information about missing outcome data. |
| 5.2. Were participants excluded due to missing data on exposure status? | Issues:<br>- Differential inclusion of participants in the analysis may lead to bias in the results. | -**N/PN** The participants were completely assessed for exposures or the percent of missingness was <10%.<br>-**Y/PY** ≥10% of the participants missing the exposure.<br>**-Y/PY** Study mentions that only participants with complete data on exposure were included.<br>**-NI** There was no information about missing data on exposure status. |
| 5.3. Were participants excluded due to missing data on other variables (besides outcome data and exposure status) needed for the analysis? | Issues:<br>- Missing covariates needed in the analysis may lead to incomplete adjustment or residual confounding.<br>Remarks:<br>- Please consult the external list of important confounding variables. | -**N/PN** The participants were completely assessed for important confounders or the percent of missingness (cumulatively) was <10%.<br>-**Y/PY** ≥10% of the participants (cumulatively) missing important confounders.<br>**-Y/PY** Study mentions that only participants with complete data on confounders were included.<br>**-NI** There was no information about missing data on confounders. |
| 5.4. **If Y or PY to 5.1, 5.2 or 5.3**: Are the proportion of participants and reasons for missing data similar across exposure groups? | Issues:<br>- Differential rates of missing data between the exposure groups may lead to bias in the results.<br>Remarks:<br>- To answer, the study has to provide the information, such as, in a sub-analysis comparing the proportion (and reason) of participants with missing outcome (or confounder) data across exposure groups. | **-Y** The proportions of missing outcome (or confounder) data were found to be the same or similar (<10%) across exposure groups.<br>**-Y** The proportions of missing data were dissimilar (≥10% difference) BUT reasons for missingness did not differ across exposure groups.<br>**-PY** The proportions of missing data were slightly different (10-<15%) AND reasons for missingness were either not presented or differ across exposure groups.<br>**-PN** The proportions of missing data were substantially different (15-<20%) AND reasons for |

| Question | Issues/ Remarks | Prompting questions/ indicators |
|---|---|---|
| | | missingness were either not presented or differ across exposure groups.<br>**-N** The proportions of missing data were critically different (≥20%) AND reasons for missingness were either not presented or differ across exposure groups.<br>-**NI** No sub-analysis performed. Studies with missing exposure data are highly unlikely to have conducted such a sensitivity analysis. |
| 5.5. **If Y or PY to 5.1, 5.2 or 5.3**: Were appropriate statistical methods used to account for missing data?<br><br>Go to Bias due to missing data: Risk of bias judgement. | Remarks:<br>- The use of appropriate statistical methods would remove/minimise any risk of bias. | -**Y/ PY** The study performed a multiple imputation analysis, inverse probability weighting, g-formula, marginal structural model (or any other equivalent method) for missing data.<br>**-PN** The study performed a simple imputation analysis for missing data.<br>**-PN** The study used a missing-indicator for categorical variables.<br>**-PN** The study performed a complete case analysis and a sensitivity analysis for missing data using a simple imputation method and found no materially different results.<br>-**N** The study did not account for missing data using any statistical methods.<br>-**N** The study had only performed a complete case analysis. |
| RoB judgement | | |
| Low risk of bias<br>The study is comparable to a well-performed randomized trial with regard to this domain. | (i) Data were reasonably complete;<br>or<br>(ii) Proportions of and reasons for missing participants were similar across exposure groups;<br>or<br>(iii) The analysis addressed missing data and is likely to have removed any risk of bias. | |
| Moderate risk of bias<br>The study is sound for an observational study with regard to this domain but cannot be considered comparable to a well-performed randomized trial. | (i) Proportions of and reasons for missing participants differ slightly across exposure groups;<br>and<br>(ii) The analysis is unlikely to have removed the risk of bias arising from the missing data. | |

| Question | Issues/ Remarks | Prompting questions/ indicators |
|---|---|---|
| Serious risk of bias<br>The study has some important problems. | (i) Proportions of missing participants differ substantially across exposures;<br>or<br>Reasons for missingness differ substantially across exposures;<br>and<br>(ii) The analysis is unlikely to have removed the risk of bias arising from the missing data;<br>or<br>Missing data were addressed inappropriately in the analysis;<br>or<br>The nature of the missing data means that the risk of bias cannot be removed through appropriate analysis. | |
| Critical risk of bias<br>The study is too problematic to provide any useful evidence. | (i) (Unusual) There were critical differences between exposures in participants with missing data;<br>and<br>(ii) Missing data were not, or could not, be addressed through appropriate analysis. | |
| **No information** on which to base a judgement about risk of bias for this domain. | There is too little information to make a judgement (for example if only an abstract is available for the study). | |

#### Bias in measurement of outcomes.

| Question | Issues/ Remarks | Prompting questions/ indicators |
|---|---|---|
| 6.1. Could the outcome measure have been influenced by knowledge of the exposure received? | Remarks:<br>- Outcome measures such as cancer incidence and mortality involve negligible assessor judgment. Risk of bias due to measurement of these outcomes would be expected to be low. | **-N** Mortality (all-cause mortality, cause-specific mortality) or any primary cancer outcomes were ascertained by record linkages to death registries, cancer registries, or hospital records.<br>**-N** Mortality outcomes were reported by their proxies and verified by record linkages to death registries, cancer registries, or hospital records.<br>**-N** Incidence outcomes were based on biopsy/ measures of laboratory testing.<br>**-PN** Mortality outcomes were reported by their proxies, and it has not been verified or verification not reported. |

| Question | Issues/ Remarks | Prompting questions/ indicators |
|---|---|---|
| | | **-PN** Outcomes were self-reported by the participants or their proxies, with an objective verification.<br>**-PY** Outcomes were self-reported by the participants or their proxies, without any objective verification. |
| 6.2. Were outcome assessors aware of the exposure received by study participants? | Remarks:<br>- "Outcome assessors" here is interpreted as "the researchers who processed the data".<br>- For self-reported measures, "outcome assessors" are "the participants" who reported the data. | **-N** Outcomes were ascertained by record linkages to death registries, cancer registries, hospital records, or laboratory measurements.<br>**-N** Outcomes were ascertained by assessors "blinded" to the exposure status of the participants.<br>**-PN** Outcomes were ascertained by independent assessors without "blinding" or "blinding" not reported.<br>**-PY** Any outcomes self-reported by the participants or their proxies, without mention of any further ascertainment.<br>**-NI** No information was reposted about outcome ascertainment. |
| 6.3. Were the methods of outcome assessment the same across exposure groups? | Issues:<br>Outcome assessment methods (detection methods and threshold, time point, definition, and measurements) should be compatible across exposure groups.<br>Remarks:<br>- Studies normally only describe one outcome assessment method that was applied to all participants, any difference would likely be described explicitly. | **-Y/ PY** Same outcome detection methods, thresholds, time point (ended on the date of event or end of study follow-up, whichever came first), same definition, same measurements, across exposure groups.<br>**-PY** It was not explicitly reported whether outcome detection methods were the same across exposure groups.<br>**-PN** The authors have explicitly described some differences of outcome ascertainment e.g., definition, detection methods, threshold, and measurement across exposure groups.<br>**-N** The authors have explicitly described very different outcome assessment, e.g., definition, detection methods, threshold, and measurement across exposure groups. |

| Question | Issues/ Remarks | Prompting questions/ indicators |
|---|---|---|
| 6.4. Were any systematic errors during measurement of the outcome related to exposure received?<br><br>Go to Bias in measurement of outcomes: Risk of bias judgement. | Issues:<br>- This question refers to differential misclassification of outcomes.<br>- Systematic errors in measuring the outcome, if present, could cause bias if they are related to exposure (or to a confounder of the exposure-outcome association).<br>Remarks:<br>Example: Outcome was detected by health screening/ examination and there was differential attendance related to the lifestyle of the participants. | **-PY** The outcome was self-reported by the participants.<br>**-PY** The authors have explicitly described different outcome assessment methods across exposure groups.<br>**-N/ PN** All other instances. |
| RoB judgement | | |
| Low risk of bias<br>The study is comparable to a well-performed randomized trial with regard to this domain. | (i) The methods of outcome assessment were comparable across exposure groups;<br>and<br>(ii) The outcome measure was unlikely to be influenced by knowledge of the exposure received by study participants (i.e. is objective) or the outcome assessors were unaware of the exposure received by study participants ***or there was no information provided about outcome ascertainment***;<br>and<br>(iii) Any error in measuring the outcome is unrelated to exposure status. | |
| Moderate risk of bias<br>The study is sound for an observational study with regard to this domain but cannot be considered comparable to a well-performed randomized trial. | (i) The methods of outcome assessment were comparable across exposure groups;<br>and<br>(ii) The outcome measure is only minimally influenced by knowledge of the exposure received by study participants ***or there was no information provided about outcome ascertainment***;<br>and<br>(iii) Any error in measuring the outcome is only minimally related to exposure status. | |
| Serious risk of bias<br>The study has some important problems. | (i) The methods of outcome assessment were not comparable across exposure groups;<br>or<br>(ii) The outcome measure was subjective (i.e. vulnerable to influence by knowledge of the exposure received by study participants);<br>and<br>The outcome was assessed by assessors aware of the exposure received by study participants; | |

| Question | Issues/ Remarks | Prompting questions/ indicators |
|---|---|---|
| | or<br>(iii) Error in measuring the outcome was related to exposure status. | |
| Critical risk of bias<br>The study is too problematic to provide any useful evidence. | The methods of outcome assessment were so different that they cannot reasonably be compared across exposure groups. | |

#### Bias in selection of reported result.

| Question | Issues/ Remarks | Prompting questions/ indicators |
|---|---|---|
| 7.1. Is the reported effect estimate likely to be selected on the basis of the results from multiple outcome measurements within the outcome domain? | Issues:<br>- It is possible to generate multiple effect estimates for different measurements of the same or related outcome/s.<br>- Studies may selectively report a subset of results for the pre-specified outcomes depending on their clinical importance (but without having stated this in the methods/protocol) or statistical significance, magnitude, or direction of the findings, which may threaten the validity of the review.<br>Remarks:<br>- Research protocol and statistical analysis plan is often unclear in observational studies. This question has to be determined from the study aim and methods and their consistency against the results reported. | -**N** All findings for the pre-specified outcomes (primary or secondary) as stated in the methods or the available protocol were reported fully in the results.<br>-**PN** Some of the findings for the pre-specified outcomes (primary or secondary) as stated in the methods or the available protocol were reported briefly as null (data not shown or with a P-value only) because of not achieving statistical significance.<br>-**Y/ PY** Not all findings for the pre-specified outcomes were reported or only statistically significant findings were reported, without mentioning of the statistical significance of the non-reported findings. |
| 7.2. Is the reported effect estimate likely to be selected on the basis of the results from multiple analyses of the exposure-outcome relationship? | Issues:<br>- Studies may analyse an exposure-outcome association in a number of pre-specified manners. Application of such methods generates multiple effect estimates. | -**N** All findings for the pre-specified analyses (different adjustments, different transformation of variables, different analysis techniques) as stated in the methods or the available protocol were reported fully in the results.<br>-**PN** Some of the findings for the pre-specified analyses (different adjustments, different |

| Question | Issues/ Remarks | Prompting questions/ indicators |
|---|---|---|
| | - If the analyst does not pre-specify the methods to be applied, and multiple estimates are generated but only one or a subset is reported (e.g., depending on their nature or significance of the findings), there is a risk of selective reporting on the basis of results.<br>Remarks:<br>Examples include unadjusted and adjusted models; use of final value vs change from baseline vs analysis of covariance; different transformations of variables; a continuously scaled outcome converted to categorical data with different cut-points; different sets of covariates used for adjustment; and different analytic strategies for dealing with missing data. | transformation of variables, different analysis techniques) as stated in the methods or the available protocol were reported briefly/ partially as null (data not shown or with a P-value only) because of not achieving statistical significance.<br>-**Y/ PY** Not all findings for the pre-specified analyses (different adjustments, different transformation of variables, different analysis techniques) were reported or only statistically significant findings were reported, without mentioning of the statistical significance of the non-reported findings. |
| 7.3. Is the reported effect estimate likely to be selected on the basis of the results from different subgroups?<br><br>Go to Bias in selection of reported result: Risk of bias judgement. | Issues:<br>- Particularly with large cohorts often available from routine data sources, it is possible to generate multiple effect estimates for different subgroups or simply to omit varying proportions of the original cohort that is not part of the design. If multiple estimates are generated but only one or a subset is reported, there is a risk of selective reporting on the basis of results. | -**N** All findings for the pre-specified subgroup analyses (exposure and cancer subtypes, population subgroups) as stated in the methods or the available protocol were reported fully in the results.<br>-**PN** Some of the findings for the pre-specified subgroup analyses (exposure and cancer subtypes, population subgroups) as stated in the methods or the available protocol were reported briefly/ partially as null (data not shown or with a P-value only) because of not achieving statistical significance.<br>-**Y/ PY** Not all findings for the pre-specified subgroup analyses (exposure and cancer subtypes, population subgroups) were reported or only statistically significant findings were reported, without mentioning of the statistical significance of the non-reported findings. |
| RoB judgement | | |
| Low risk of bias | There is clear evidence (usually through examination of a pre-registered protocol or statistical analysis plan) that all reported results correspond to all intended outcomes, analyses and sub-cohorts. | |

| Question | Issues/ Remarks | Prompting questions/ indicators |
|---|---|---|
| The study is comparable to a well-performed randomized trial with regard to this domain. | | |
| Moderate risk of bias<br>The study is sound for an observational study with regard to this domain but cannot be considered comparable to a well-performed randomized trial. | (i) The outcome measurements and analyses are consistent with an a priori plan; or are clearly defined and both internally and externally consistent;<br>and<br>(ii) There is no indication of selection of the reported analysis from among multiple analyses;<br>and<br>(iii) There is no indication of selection of the cohort or subgroups for analysis and reporting on the basis of the results. | |
| Serious risk of bias<br>The study has some important problems. | (i) Outcomes are defined in different ways in the methods and results sections, or in different publications of the study;<br>or<br>(ii) There is a high risk of selective reporting from among multiple analyses;<br>or<br>(iii) The cohort or subgroup is selected from a larger study for analysis and appears to be reported on the basis of the results. | |
| Critical risk of bias<br>The study is too problematic to provide any useful evidence. | (i) There is evidence or strong suspicion of selective reporting of results;<br>and<br>(ii) The unreported results are likely to be substantially different from the reported results. | |

**Supplementary Table 4: List of important confounding variables.**

| Cancer | Confounder* | | Mediator variable(s)** |
|---|---|---|---|
| Bladder | Required | Age, sex, smoking, physical activity | Body fatness |
| | Desirable | --- | |
| Brain and spinal cord | Required | Age, sex, physical activity | --- |
| | Desirable | --- | |
| Colorectal | Required | Age, sex, physical activity | Body fatness |
| | Desirable | Smoking, alcohol, physical activity, red/processed meat | |
| Gallbladder/bile ducts | Required | Age, sex, smoking, physical activity | Body fatness |
| | Desirable | Gallstones/gallbladder disease/cholecystitis | |
| Kidney | Required | Age, sex, alcohol, physical activity | Body fatness |
| | Desirable | --- | |
| Liver | Required | Age, sex, alcohol, physical activity | Body fatness |
| | Desirable | Smoking, race, one of: *HBV, HCV, liver cirrhosis* | |
| Lung | Required | Age, sex, smoking, physical activity | --- |
| | Desirable | Race, environmental tobacco smoke | |
| Lymphoid and hematopoietic | Required | Age, sex, physical activity | --- |
| | Desirable | --- | |
| Mouth, pharynx, and larynx | Required | Age, sex, smoking, alcohol, physical activity | Body fatness |
| | Desirable | Race, environmental tobacco smoke | |
| Nasopharynx | Required | Age, sex, smoking, physical activity | Body fatness |
| | Desirable | --- | |

| Cancer | Confounder* | | Mediator variable(s)** |
|---|---|---|---|
| Oesophageal adenocarcinoma | Required<br>Desirable | Age, sex, physical activity<br>Smoking, alcohol, race | Body fatness |
| Oesophageal squamous cell carcinoma | Required<br>Desirable | Age, sex, smoking, alcohol, physical activity<br>Race | Body fatness |
| Pancreas | Required<br>Desirable | Age, sex, smoking, physical activity<br>Pancreatitis, alcohol | Body fatness |
| Melanoma | Required<br>Desirable | Age, sex, physical activity, at least one of the following: UV light -sun exposure/ indoor tanning, sunscreen use, moles, fair skin, red/blond hair, blue/green eyes, freckles<br>Race | --- |
| Basal cell skin cancer | Required<br>Desirable | Age, sex, physical activity, at least one of the following: UV light - sun exposure/ indoor tanning, sunscreen use, radiation therapy, immune-suppressing drugs<br>Race | --- |
| Small intestine | Required<br>Desirable | Age, sex, physical activity<br>Inflammatory Bowel Disease | --- |
| Stomach | Required<br>Desirable | Age, sex, smoking, alcohol, physical activity<br>Salt/preserved foods, race, Helicobacter Pylori | Body fatness |
| Thyroid | Required<br>Desirable | Age, sex, physical activity<br>One of: Benign thyroid nodules, goiter, thyroiditis | --- |
| Sex-specific cancers | | | |
| Breast - premenopausal | Required | Age, physical activity, reproductive factor(s) | Body fatness |

| Cancer | Confounder* | | Mediator variable(s)** |
|---|---|---|---|
| | Desirable | Alcohol, smoking, HRT/ oral contraceptive use | |
| Breast - postmenopausal | Required<br>Desirable | Age, physical activity, reproductive factor(s)<br>Alcohol, smoking, HRT | Body fatness |
| Cervix | Required<br>Desirable | Age, HPV, smoking, physical activity | Body fatness |
| Endometrium | Required<br>Desirable | Age, race, physical activity<br>HRT/ oral contraceptive use | Body fatness |
| Ovary | Required<br>Desirable | Age, physical activity, reproductive factor(s)<br>HRT/ oral contraceptive use | Body fatness |
| Prostate | Required<br>Desirable | Age, race, physical activity<br>Smoking | --- |

***Smoking:** For cancers not related very strongly to smoking such as prostate, breast or colorectal, status is ok. For smoking-related cancers, adjustments should also include duration and intensity.

**** Mediator variable(s)** should be considered as an intermediate factor(s) between sedentary behaviour and cancer and should not be adjusted for when investigating these associations.

**Reproductive factor(s):** At least one of the following: age at menarche, age at menopause, parity, lactation.

#### Supplementary Table 5: WCRF grading criteria for evidence on diet, nutrition, physical activity and cancer incidence.

| Evidence grades | GRADING CRITERIA FOR EVIDENCE ON DIET, NUTRITION, PHYSICAL ACTIVITY AND CANCER INCIDENCE |
|---|---|
| **Convincing** | A combination of all the following points<br><br>• **Volume and type of evidence**: Convincing epidemiological evidence from several well-conducted, independent cohort studies and, when available, supporting evidence from pertinent RCTs<br>• **Consistency of evidence**: This includes absence of unexplained heterogeneity between studies<br>• **Quality of evidence**: This excludes with confidence the possibility that the observed association results from systematic error, including confounding, measurement error, and selection bias<br>• **Level of statistical significance and precision**: When available, summary risk estimates from meta-analyses with sufficiently narrow 95%CI, clearly excluding no association |
| **Probable** | A combination of all the following points<br><br>• **Volume and type of evidence**: Sufficient epidemiological evidence from at least two well-conducted, independent cohort studies<br>• **Consistency of evidence**: This includes absence of unexplained heterogeneity between studies<br>• **Quality of evidence**: This excludes with confidence the possibility that the observed association results from systematic error, including confounding, measurement error, and selection bias<br>• **Level of statistical significance and precision**: When available, summary risk estimates from meta-analyses indicating an association with 95%CI excluding the null |
| **Limited suggestive** | A combination of both of the following points<br><br>• **Volume and type of evidence**: Evidence from at least two independent cohort studies<br>• **Consistency of evidence**: Some unexplained heterogeneity may be present<br>• **Quality of Evidence:** Not able to rule out the possibility that the observed association may primarily result from systematic error, including confounding and selection bias<br>• **Level of statistical significance and precision:** When available, summary risk estimates from meta-analyses suggesting an association, but with 95% CI that may include the null |
| **Limited – no conclusion** | Any of the following reasons – or a combination of them:<br><br>• **Volume and type of evidence**: Too few studies available. Note there may be instances where the Panel considers an individual study warrants a *Limited-no conclusion* judgement<br>• **Consistency of evidence**: Inconsistency of direction of effect across different studies<br>• **Quality of the evidence**: Poor quality of studies (for example, lack of adjustment for known confounders)<br>• **Level of statistical significance and precision:** Too imprecise risk estimate(s) |
| **Substantial effect on risk unlikely** | A combination of all the following points<br><br>• **Volume and type of evidence**: Convincing epidemiological evidence from several well-conducted, independent cohort studies of an absence of effect and, when available, supporting evidence of no effect from pertinent RCTs<br>• **Consistency of evidence**: This includes absence of unexplained heterogeneity between studies |

| | |
|---|---|
| | • **Quality of evidence**: This excludes with confidence the possibility that the observed association results from systematic error, including confounding, measurement error, and selection bias<br>• **Level of statistical significance and precision**: When available, summary risk estimates from meta-analyses close to 1.0, with sufficiently narrow 95%CI clearly indicating that an association is unlikely |

Note that there are exposures for which there is insufficient data to make a judgement (typically when only a single cohort study, of moderate quality, is available for a specific exposure-outcome pair of interest). These do not qualify for any of the evidence grades above. These include:

1- Total sedentary or sitting time: These definitions were not graded separately, as they are always encompassed within the mixed-definition analyses of overall sedentary behaviour, following the pre-specified hierarchy used for exposure selection for the mixed definition analyses.

2- Cancer subsites or subtypes (e.g., ER+ or ER- breast cancer, advanced prostate cancer): The numbers of studies for these cancer subtypes/subsites are often too low to inform evidence grading, unlike for colon or rectum cancers and cancers among pre- or postmenopausal women. For the latter, overall cancers (colorectal cancer, breast cancer) are also graded, utilising all available evidence.

3- Cancer mortality: The evidence for cancer mortality was not graded and considered as supportive evidence.

**Upgrading factors**

- Presence of a **plausible gradient (‘dose-response’)** in the association. Such a gradient need not be linear or even in the same direction across the different levels of exposure, so long as this can be explained plausibly.
- **Strong and plausible mechanistic evidence**, either from human studies or relevant animal models, that typical human exposures can lead to relevant cancer outcomes.
- A particularly **large summary effect size** (an odds ratio or relative risk of 2.0 or more - depending on the unit of exposure - with a 95%CI excluding the null), after appropriate control for confounders.
- Evidence from well-conducted, pertinent (in terms of exposure, dosage, and duration of intervention and follow-up period) **randomised controlled trials in humans supporting** the epidemiological evidence.
- Additional consideration regarding **Mendelian randomisation (MR) studies:**
  There is currently no formal method for assessing the quality of MR studies in a rapidly evolving field or for determining how well these studies apply to specific nutritional exposures. Therefore, MR studies are not formally listed as an upgrading factor. However, all MR studies that appear potentially relevant will be presented to the Panel, who will decide, case by case, whether to incorporate that evidence into its judgment and, if so, how.

**Downgrading factors**

- Evidence from well-conducted, pertinent (in terms of exposure, dosage, and duration of intervention and follow-up period) **randomised controlled trials in humans unsupportive** of the epidemiological evidence
- Evidence of **publication bias**

#### Supplementary Table 6: Publications excluded and reasons of exclusion from the potentially eligible studies after further assessment.

| Reason of exclusion | Number of studies | Reference (PMID) |
|---|---|---|
| **78** publications excluded after further inspection | | |
| Definition of exposure/Comparison not of interest | **67** | Lund Nilsen 2000 (10755415)[135]; Krishnadasan 2008 (18064535)[136]; Cerhan 2005 (16215871)[137]; Thune 1994 (7827242)[138]; Thune 1996 (8624277)[139]; Thune 1997 (9113929)[140]; Veierod 1997 (9398038)[141]; Hartman 1998 (9486459)[142]; Nilsen 2000 (10977109)[143]; Colbert 2001 (11303597)[144]; Colbert 2002 (11920649)[145]; Moradi 2002 (12115590)[146]; Stolzenberg-Solomon 2002 (12146846)[147]; Berrington de Gonzalez 2006 (16702364)[148]; Furberg 2003 (12640672)[149]; Mahabir 2004 (14696127)[150]; Gerhardsson 1988 (3272134)[151]; Severson 1989 (2763997)[152]; Veierod 1997 (9178814)[153]; Lund Haheim 2006 (16952929)[154]; Batty 2009 (19190162)[155]; Huerta 2010 (20052611)[156]; van Veldhoven 2011 (21159506)[157]; Vena 1987 (3799522)[158]; Garabrant 1984 (6731427)[159]; Fraser 1993 (8431530)[160]; Thune 1997 (8985091)[161]; Moradi 1998 (9610723)[162]; Bergstrom 1999 (10471525)[163]; Moradi 1999 (10530613)[164]; Putnam 2000 (10964002)[165]; Bergstrom 2001 (11279620)[166]; Weiderpass 2001 (11385641)[167]; Norman 2002 (11857014)[168]; Isaksson 2002 (11920604)[169]; Rintala b 2002 (12109554)[170]; Weiderpass 2003 (12661188)[171]; Charles 2003 (12697572)[172]; Robsahm 2005 (15677890)[173]; Bak 2005 (15800940)[174]; Norat 2005 (15956652)[175]; Washio 2005 (16127235)[176]; Steindorf 2006 (16894558)[177]; Larsson 2006 (16914307)[178]; Johnsen 2006 (17160429)[179]; Friedenreich 2006 (17164362)[180]; Lahmann 2007 (17179488)[181]; Friedenreich 2007 (17357139)[182]; Wilson 2008 (18067176)[183]; Moradi 2008 (18414190)[184]; Lahmann 2009 (19124520)[185]; Johnsen 2009 (19415749)[186]; Orsini 2009 (19861965)[187]; Rundle 2010 (20050820)[188]; Lahmann 2011 (22165995)[189]; Steindorf 2013 (22903273)[190]; Steindorf 2012 (23074288)[191]; Grotta 2015 (25557943)[192]; Hrafnkelsdottir 2015 (26152935)[193]; Ekenga 2015 (26450605)[194]; Sormunen 2016 (27420632)[195]; Masala 2017 (27832394)[196]; Johnsson 2017 (27919198)[197]; Pahwa 2017 (28651179)[198]; Sari 2020 (32664915)[199]; Eshak 2022 (35048204)[200]; An 2024 (37817564)[201] |

| Reason of exclusion | Number of studies | Reference (PMID) |
|---|---|---|
| Superseded publications | **6** | Patel 2006 (16495470)[202]; Patel 2008 (18651569)[203]; Rosenberg 2014 (25103823)[204]; Gaudet 2021 (33398477)[205]; Sasamoto 2021 (33653814)[206]; Jenniskens 2022 (35546360)[207] |
| Standardised incidence ratios | **3** | Hsing 1994 (8167260)[208]; Chow 1993 (8449643)[209]; Zheng 1993 (8490910)[210] |
| Only unadjusted results | **1** | Saberi Hosnijeh 2021 (33128820)[211] |
| Examined total cancer incidence | **1** | Robsahm 2016 (27227704))[212] |

#### Supplementary Table 7: List of publications included in the systematic literature review, and studies included and not included in the meta-analyses of specific definitions of sedentary behaviour (cancers are ordered by ICD-10 codes).

##### Publications included in or excluded from the systematic literature review and meta-analyses of sedentary time and risk of cancer.

| Cancer | Outcome | Publications identified | Publications included in various meta-analyses | Publications included in the descriptive synthesis only | Superseded |
|---|---|---|---|---|---|
| **Sedentary time, total** | | | | | |
| Skin, non-melanoma | Incidence | 1 publication: Watts 2023[88] | N/A | 1 publication: Watts 2023[88] | N/A |
| Breast | Incidence | 1 publication: Tomida 2024[91] | N/A | 1 publication: Tomida 2024[91] | N/A |
| Breast, postmenopausal | Incidence | 1 publication: Tomida 2024[91] | N/A | 1 publication: Tomida 2024[91] | N/A |
| Breast, premenopausal | Incidence | 1 publication: Tomida 2024[91] | N/A | 1 publication: Tomida 2024[91] | N/A |
| **Sedentary time, non-occupational** | | | | | |
| Pancreas | Incidence | 1 publication: Zeng 2023[92] | N/A | 1 publication: Zeng 2023[92] | N/A |
| Melanoma | Incidence | 1 publication: Weber 2021[83] | N/A | 1 publication: Weber 2021[83] | N/A |

##### Publications included in or excluded from the systematic literature review and meta-analyses of sitting time and risk of cancer.

| Cancer | Outcome | Publications identified | Publications included in various meta-analyses | Publications included in the descriptive synthesis only | Superseded |
|---|---|---|---|---|---|
| **Sitting time, total** | | | | | |
| Oesophageal, adenocarcinoma | Incidence | 1 publication: Cook 2013[52] | N/A | 1 publication: Cook 2013[52] | N/A |
| Oesophageal, squamous cell carcinoma | Incidence | 1 publication: Cook 2013[52] | N/A | 1 publication: Cook 2013[52] | N/A |
| Gastric, cardia | Incidence | 1 publication: Cook 2013[52] | N/A | 1 publication: Cook 2013[52] | N/A |
| Gastric, non-cardia | Incidence | 1 publication: Cook 2013[52] | N/A | 1 publication: Cook 2013[52] | N/A |
| Colorectal | Incidence | 4 publications: Gorczyca 2018[61], Rangul 2018[70], Park 2019[73], Nunez 2018[66] | 3 publications: Gorczyca 2018[61], Park 2019[73], Nunez 2018[66]<br><br>1 extra publication: in high vs low Rangul 2018[70] | N/A | N/A |
| Colon | Incidence | 3 publications: Howard 2008[36], Gorczyca 2018[61], Nunez 2018[66] | 3 publications: Howard 2008[36], Gorczyca 2018[61], Nunez 2018[66] | N/A | N/A |
| Rectal | Incidence | 2 publications: Gorczyca 2018[61], Nunez 2018[66] | 2 publications: Gorczyca 2018[61], Nunez 2018[66] | N/A | N/A |
| Lung | Incidence | 4 publications: Lam 2013[48], Wang 2016[59], Rangul 2018[70], Jiang 2019[72] | 3 publications: Lam 2013[48], Wang 2016[59], Jiang 2019[72] | 1 publication: Rangul 2018[70] (Only categorical analysis by sex) | N/A |
| Lung | Mortality | 1 publication: Wang 2016[59] | N/A | 1 publication: Wang 2016[59] | N/A |
| Lung, non-small cell | Incidence | 1 publication: Jiang 2019[72] | N/A | 1 publication: Jiang 2019[72] | N/A |
| Lung, small cell | Incidence | 1 publication: Jiang 2019[72] | N/A | 1 publication: Jiang 2019[72] | N/A |

| | | | | | |
|---|---|---|---|---|---|
| Breast | Incidence | 7 publications: Cohen 2013[46], Catsburg 2014[55], Nomura 2016[60], Rangul 2018[70], Stahr 2019[85], Wang 2022[86], Rosenberg 2014[204] | 4 publications: Cohen 2013[46], Catsburg 2014[55], Nomura 2016[60], Stahr 2019[85]<br>2 extra publications: in high vs low Rangul 2018[70], Wang 2022[86] | N/A | 1 publication: Rosenberg 2014[204] |
| Breast, postmenopausal | Incidence | 6 publications: George 2010[39], Cohen 2013[46], Catsburg 2014[55], Nomura 2016[60], Nomura 2017[63], Stahr 2019[85] | 5 publications: George 2010[39], Catsburg 2014[55], Nomura 2016[60], Nomura 2017[63], Stahr 2019[85] | 1 publication: Cohen 2013[46] (Only categorical analysis) | N/A |
| Breast, premenopausal | Incidence | 3 publications: Catsburg 2014[55], Nomura 2016[60], Stahr 2019[85] | 3 publications: Catsburg 2014[55], Nomura 2016[60], Stahr 2019[85] | N/A | N/A |
| Endometrial | Incidence | 2 publications: Gierach 2009[38], Moore 2010[40] | N/A | 1 publication: Moore 2010[40] | 1 publication: Gierach 2009[38] |
| Ovary | Incidence | 3 publications: Buras 2022[84], Xiao 2013[49], Sasamoto 2021[206] | 2 publications: Buras 2022[84], Xiao 2013[49] | N/A | 1 publications: Sasamoto 2021[206] |
| Ovary, epithelial | Incidence | 1 publication: Xiao 2013[49] | N/A | 1 publication: Xiao 2013[49] | N/A |
| Prostate | Incidence | 2 publications: Lynch 2014b[53], Rangul 2018[70] | 2 publications: in high vs low Lynch 2014b[53], Rangul 2018[70] | N/A | N/A |
| Prostate | Mortality | 1 publication: Lynch 2014b[53] | N/A | 1 publication: Lynch 2014b[53] | N/A |
| Prostate, advanced | Incidence | 1 publication: Lynch 2014b[53] | N/A | 1 publication: Lynch 2014b[53] | N/A |
| Kidney | Incidence | 1 publication: George 2011[41] | N/A | 1 publication: George 2011[41] | N/A |
| Acute myeloid leukaemia | Incidence | 1 publication: Rees-Punia 2019[75] | N/A | 1 publication: Rees-Punia 2019[75] | N/A |
| Myeloid leukaemia | Incidence | 1 publication: Rees-Punia 2019[75] | N/A | 1 publication: Rees-Punia 2019[75] | N/A |
| **Sitting time, recreational** | | | | | |
| Mouth | Incidence | 1 publication: Patel 2015[56] | N/A | 1 publication: Patel 2015[56] | N/A |
| Oesophageal | Incidence | 1 publication: Patel 2015[56] | N/A | 1 publication: Patel 2015[56] | N/A |
| Gastric | Incidence | 1 publication: Patel 2015[56] | N/A | 1 publication: Patel 2015[56] | N/A |
| Colorectal | Incidence | 1 publication: Patel 2015[56] | N/A | 1 publication: Patel 2015[56] | N/A |
| Liver | Incidence | 1 publication: Patel 2015[56] | N/A | 1 publication: Patel 2015[56] | N/A |
| Gallbladder | Incidence | 1 publication: Patel 2015[56] | N/A | 1 publication: Patel 2015[56] | N/A |
| Pancreas | Incidence | 1 publication: Patel 2015[56] | N/A | 1 publication: Patel 2015[56] | N/A |
| Lung | Incidence | 1 publication: Patel 2015[56] | N/A | 1 publication: Patel 2015[56] | N/A |
| Melanoma | Incidence | 1 publication: Patel 2015[56] | N/A | 1 publication: Patel 2015[56] | N/A |
| Breast | Incidence | 1 publication: Patel 2015[56] | N/A | 1 publication: Patel 2015[56] | N/A |
| Breast, postmenopausal | Incidence | 1 publication: Hildebrand 2013[51] | N/A | 1 publication: Hildebrand 2013[51] | N/A |
| Endometrial | Incidence | 2 publications: Patel 2015[56], Patel 2008[203] | N/A | 1 publication: Patel 2015[56] | 1 publication: Patel 2008[203] |
| Ovary | Incidence | 3 publications: Hildebrand 2015[57], Patel 2006[202], Patel 2015[56] | N/A | 1 publication: Hildebrand 2015[57] | 2 publications: Patel 2006[202], Patel 2015[56] |
| Ovary, non-serous | Incidence | 1 publication: Hildebrand 2015[57] | N/A | 1 publication: Hildebrand 2015[57] | N/A |
| Ovary, serous | Incidence | 1 publication: Hildebrand 2015[57] | N/A | 1 publication: Hildebrand 2015[57] | N/A |
| Prostate | Incidence | 1 publication: Patel 2015[56] | N/A | 1 publication: Patel 2015[56] | N/A |

| | | | | | |
|---|---|---|---|---|---|
| Prostate, advanced | Incidence | 1 publication: Patel 2015[56] | N/A | 1 publication: Patel 2015[56] | N/A |
| Kidney | Incidence | 1 publication: Patel 2015[56] | N/A | 1 publication: Patel 2015[56] | N/A |
| Bladder | Incidence | 2 publications: Chantaprasopsuk 2023[89], Patel 2015[56] | N/A | 1 publication: Chantaprasopsuk 2023[89] | 1 publication: Patel 2015[56] |
| Follicular lymphoma | Incidence | 1 publication: Teras 2012[44] | N/A | 1 publication: Teras 2012[44] | N/A |
| Non-Hodgkin Lymphoma | Incidence | 2 publications: Teras 2012[44], Patel 2015[56] | N/A | 1 publication: Teras 2012[44] | 1 publication: Patel 2015[56] |
| Diffuse large B cell lymphoma | Incidence | 1 publication: Teras 2012[44] | N/A | 1 publication: Teras 2012[44] | N/A |
| Multiple Myeloma | Incidence | 2 publications: Patel 2015[56], Teras 2012[44] | N/A | 1 publication: Teras 2012[44] | 1 publication: Teras 2012[44] |
| Leukaemia | Incidence | 1 publication: Teras 2012[44] | N/A | 1 publication: Teras 2012[44] | N/A |
| **Sitting time, non-recreational** | | | | | |
| Colorectal | Incidence | 1 publication: Nguyen 2018[71] | N/A | 1 publication: Nguyen 2018[71] | N/A |
| **Sitting time, occupational** | | | | | |
| Oesophageal | Incidence | 1 publication: Ihira 2020[77] | N/A | 1 publication: Ihira 2020[77] | N/A |
| Gastric | Incidence | 1 publication: Ihira 2020[77] | N/A | 1 publication: Ihira 2020[77] | N/A |
| Colorectal | Incidence | 4 publications: Keum 2016[58], Guo 2020[76], Ihira 2020[77], Simons 2013[45] | 4 publications: Keum 2016[58], Guo 2020[76], Ihira 2020[77], Simons 2013[45] | N/A | N/A |
| Colon | Incidence | 4 publications: Simons 2013[45], Ihira 2020[77], Gerhardsson 1986[94], Jenniskens 2022[207] | 2 publications: Simons 2013[45], Ihira 2020[77], | 1 publication: Gerhardsson 1986[94] (Only categorical analysis) | 1 publication: Jenniskens 2022[207] |
| Rectal | Incidence | 4 publications: Simons 2013[45], Ihira 2020[77], Gerhardsson 1986[94], Jenniskens 2022[207] | 2 publications: Simons 2013[45], Ihira 2020[77] | 1 publication: Gerhardsson 1986[94] (Only categorical analysis) | 1 publication: Jenniskens 2022[207] |
| Liver | Incidence | 1 publication: Ihira 2020[77] | N/A | 1 publication: Ihira 2020[77] | N/A |
| Pancreas | Incidence | 3 publications: Heinen 2011[43], Wu 2018[65], Ihira 2020[77] | 3 publications: Heinen 2011[43], Wu 2018[65], Ihira 2020[77] | N/A | N/A |
| Lung | Incidence | 1 publication: Ihira 2020[77] | N/A | 1 publication: Ihira 2020[77] | N/A |
| Breast | Incidence | 5 publications: Cohen 2013[46], Nomura 2016[60], Ihira 2020[77], Rosenberg 2014[204], Pronk 2011[42] | 4 publications: Cohen 2013[46], Nomura 2016[60], Ihira 2020[77], Pronk 2011[42] | N/A | 1 publication: Rosenberg 2014[204] |
| Breast, postmenopausal | Incidence | 2 publications: Dirx 2001[33], Nomura 2016[60] | 2 publications: Dirx 2001[33], Nomura 2016[60] | N/A | N/A |
| Breast, premenopausal | Incidence | 1 publication: Nomura 2016[60] | N/A | 1 publication: Nomura 2016[60] | N/A |
| Endometrial | Incidence | 1 publication: Ihira 2020[77] | N/A | 1 publication: Ihira 2020[77] | N/A |
| Ovary | Incidence | 2 publications: Ihira 2020[77], Buras 2022[84] | 2 publications: Ihira 2020[77], Buras 2022[84] | N/A | N/A |
| Prostate | Incidence | 2 publications: Zeegers 2005[34], Ihira 2020[77] | 2 publications: Zeegers 2005[34], Ihira 2020[77] | N/A | N/A |
| Kidney | Incidence | 1 publication: Ihira 2020[77] | N/A | 1 publication: Ihira 2020[77] | N/A |
| Bladder | Incidence | 1 publication: Ihira 2020[77] | N/A | 1 publication: Ihira 2020[77] | N/A |
| **Sitting time, transportational** | | | | | |
| Colorectal | Incidence | 1 publication: Keum 2016[58] | N/A | 1 publication: Keum 2016[58] | N/A |

| | | | | | |
|---|---|---|---|---|---|
| Pancreas | Incidence | 1 publication: Peduzzi 2024[90] | N/A | 1 publication: Peduzzi 2024[90] | N/A |
| Breast | Incidence | 1 publication: Cohen 2013[46] | N/A | 1 publication: Cohen 2013[46] | N/A |
| **Sitting time, other** | | | | | |
| Oesophageal | Incidence | 1 publication: Ihira 2020[77] | N/A | 1 publication: Ihira 2020[77] | N/A |
| Gastric | Incidence | 1 publication: Ihira 2020[77] | N/A | 1 publication: Ihira 2020[77] | N/A |
| Colorectal | Incidence | 3 publications: Eaglehouse 2017[62], Nguyen 2018[71], Ihira 2020[77] | N/A | 2 publications: Eaglehouse 2017[62], Ihira 2020[77] (different exposure definitions)<br>1 publication: Nguyen 2018[71] (early onset) | N/A |
| Colon | Incidence | 1 publication: Ihira 2020[77] | N/A | 1 publication: Ihira 2020[77], | N/A |
| Rectal | Incidence | 1 publication: Ihira 2020[77] | N/A | 1 publication: Ihira 2020[77], | N/A |
| Liver | Incidence | 1 publication: Ihira 2020[77] | N/A | 1 publication: Ihira 2020[77], | N/A |
| Pancreas | Incidence | 1 publication: Ihira 2020[77] | N/A | 1 publication: Ihira 2020[77], | N/A |
| Lung | Incidence | 1 publication: Ihira 2020[77] | N/A | 1 publication: Ihira 2020[77], | N/A |
| Breast | Incidence | 2 publications: Cohen 2013[46], Ihira 2020[77] | N/A | 2 publications: Cohen 2013[46], Ihira 2020[77] (different exposure definitions) | N/A |
| Endometrial | Incidence | 1 publication: Ihira 2020[77] | N/A | 1 publication: Ihira 2020[77] | N/A |
| Ovary | Incidence | 2 publications: Ihira 2020[77], Buras 2022[84] | N/A | 2 publications: Ihira 2020[77], Buras 2022[84] (different exposure definitions) | N/A |
| Kidney | Incidence | 1 publication: Ihira 2020[77] | N/A | 1 publication: Ihira 2020[77] | N/A |
| Bladder | Incidence | 1 publication: Ihira 2020[77] | N/A | 1 publication: Ihira 2020[77] | N/A |

#### Publications included in or excluded from the systematic literature review and meta-analyses of screen time and risk of cancer.

| Cancer | Outcome | Publications identified | Publications included in various meta-analyses | Publications included in the descriptive synthesis only | Superseded |
|---|---|---|---|---|---|
| **Screen time, recreational** | | | | | |
| Oesophageal | Incidence | 1 publication: Kunzmann 2018[68] | N/A | 1 publication: Kunzmann 2018[68] | N/A |
| Gastric | Incidence | 1 publication: Kunzmann 2018[68] | N/A | 1 publication: Kunzmann 2018[68] | N/A |
| Gastric, adeno | Incidence | 1 publication: Kunzmann 2018[68] | N/A | 1 publication: Kunzmann 2018[68] | N/A |
| Gastric, cardia | Incidence | 1 publication: Kunzmann 2018[68] | N/A | 1 publication: Kunzmann 2018[68] | N/A |
| Gastric, non-cardia | Incidence | 1 publication: Kunzmann 2018[68] | N/A | 1 publication: Kunzmann 2018[68] | N/A |
| Colorectal | Incidence | 1 publication: Zhang 2024[93] | N/A | 1 publication: Zhang 2024[93] | N/A |
| Lung | Incidence | 1 publication: Zhang 2024[93] | N/A | 1 publication: Zhang 2024[93] | N/A |
| Breast | Incidence | 1 publication: Zhang 2024[93] | N/A | 1 publication: Zhang 2024[93] | N/A |
| Prostate | Incidence | 1 publication: Zhang 2024[93] | N/A | 1 publication: Zhang 2024[93] | N/A |
| **Screen time, computers, recreational** | | | | | |
| Colon | Incidence | 1 publication: Morris 2018[67] | N/A | 1 publication: Morris 2018[67] | N/A |

| | | | | | |
|---|---|---|---|---|---|
| Colorectal | Incidence | 1 publication: Morris 2018[67] | N/A | 1 publication: Morris 2018[67] | N/A |
| Rectal | Incidence | 1 publication: Morris 2018[67] | N/A | 1 publication: Morris 2018[67] | N/A |
| **Television watching time** | | | | | |
| Oesophageal | Incidence | 2 publications: Hunter 2020[78]; Cook 2013[52] | 2 publications: Hunter 2020[78]; Cook 2013[52] | N/A | N/A |
| Gastric | Incidence | 2 publications: Hunter 2020[78]; Cook 2013[52] | 2 publications: Hunter 2020[78]; Cook 2013[52] | N/A | N/A |
| Colorectal | Incidence | 6 publications: Keum 2016[58];Eaglehouse 2017[62];Morris 2018[67];Nguyen 2018[71];Park 2019[73];Hunter 2020[78] | 5 publications: Keum 2016[58];Eaglehouse 2017[62];Nguyen 2018[71];Park 2019[73];Hunter 2020[78]<br><br>1 publication: Morris 2018[67] (only for sex-specific analyses) | N/A | 1 publication: Morris 2018[67] (only for both sexes) |
| Colorectal | Mortality | 3 publications: Kim 2013[50];Li 2021[81];Morris 2018[67] | 2 publications: Kim 2013[50];Li 2021[81] | N/A | 1 publication: Morris 2018[67] |
| Colon | Incidence | 6 publications: Howard 2008[36];Keum 2016[58];Eaglehouse 2017[62];Morris 2018[67];Nguyen 2018[71];Hunter 2020[78] | 4 publications: Howard 2008[36];Keum 2016[58];Eaglehouse 2017[62];Hunter 2020[78]<br><br>1 publication: Morris 2018[67] (only for sex-specific analyses) | 1 publication: Nguyen 2018[71] (early onset) | 1 publication: Morris 2018[67] (only for both sexes) |
| Colon | Mortality | 1 publication: Li 2021[81] | N/A | 1 publication: Li 2021[81] | N/A |
| Rectal | Incidence | 5 publications: Keum 2016[58];Eaglehouse 2017[62];Morris 2018[67];Nguyen 2018[71];Hunter 2020[78] | 3 publications: Keum 2016[58];Eaglehouse 2017[62];Hunter 2020[78]<br><br>1 publication: Morris 2018[67] (only for sex-specific analyses) | 1 publication: Nguyen 2018[71] (early onset) | 1 publication: Morris 2018[67] (only for both sexes) |
| Rectal | Mortality | 1 publication: Li 2021[81] | N/A | 1 publication: Li 2021[81] | N/A |
| Hepatobiliary tract | Incidence | 1 publication: Hunter 2020[78] | N/A | 1 publication: Hunter 2020[78] | |
| Liver | Mortality | 1 publication: Ukawa 2014[54] | N/A | 1 publication: Ukawa 2014[54] | N/A |
| Pancreas | Incidence | 1 publication: Hunter 2020[78] | N/A | 1 publication: Hunter 2020[78] | N/A |
| Lung | Incidence | 4 publications: Ukawa 2013[47];Lam 2013[48];Hunter 2020[78];Bethea 2022[87] | 3 publications: Ukawa 2013[47];Lam 2013[48];Hunter 2020[78]<br><br>1 extra publication: in high vs low Bethea 2022[87] | N/A | N/A |
| Lung | Mortality | 1 publication: Kim 2013[50] | N/A | 1 publication: Kim 2013[50] | N/A |
| Breast | Incidence | 8 publications: Cohen 2013[46];Catsburg 2014[55];Nomura 2016[60];Cifu 2018[69];Cao 2019[74];Hunter 2020[78];Sanchez-Bayona 2021[82]; Rosenberg 2014[204] | 6 publications: Cohen 2013[46];Catsburg 2014[55];Nomura 2016[60];Cao 2019[74];Hunter 2020[78];Sanchez-Bayona 2021[82] | 1 publication: Cifu 2018[69] (Only categorical analysis) | 1 publication: Rosenberg 2014[204] |

| | | | | | |
|---|---|---|---|---|---|
| Breast | Mortality | 2 publications: Kim 2013[50];Cifu 2018[69] | 2 publications: in high vs low Kim 2013[50];Cifu 2018[69] | N/A | N/A |
| Breast, postmenopausal | Incidence | 6 publications: George 2010[39]; Catsburg 2014[55];Nomura 2016[60];Cao 2019[74]; Hunter 2020[78];Sanchez-Bayona 2021[82] | 6 publications: George 2010[39]; Catsburg 2014[55];Nomura 2016[60];Cao 2019[74]; Hunter 2020[78];Sanchez-Bayona 2021[82] | N/A | N/A |
| Breast, premenopausal | Incidence | 5 publications: Catsburg 2014[55];Nomura 2016[60];Cao 2019[74]; Hunter 2020[78];Sanchez-Bayona 2021[82] | 5 publications: Catsburg 2014[55];Nomura 2016[60];Cao 2019[74];Hunter 2020[78] ;Sanchez-Bayona 2021[82] | N/A | N/A |
| Endometrial | Incidence | 4 publications: Friberg 2006[35];Gierach 2009[38];Miyata 2021[79];Hunter 2020[78] | 3 publications: Gierach 2009[38];Miyata 2021[79];Hunter 2020[78]<br>1 extra publication: in high vs low Friberg 2006[35] | N/A | N/A |
| Ovary | Incidence | 4 publications: Xiao 2013[49];Ukawa 2018[64];Hunter 2020[78];Buras 2022[84] | 4 publications: Xiao 2013[49];Ukawa 2018[64];Hunter 2020[78];Buras 2022[84] | N/A | N/A |
| Ovary, epithelial | Incidence | 1 publication: Xiao 2013[49] | N/A | 1 publication: Xiao 2013[49] | N/A |
| Prostate | Incidence | 2 publications: Lynch 2014b[53];Hunter 2020[78] | 2 publications: Lynch 2014b[53];Hunter 2020[78] | N/A | N/A |
| Prostate | Mortality | 2 publications: Kim 2013[50];Lynch 2014b[53] | 2 publications: Kim 2013[50];Lynch 2014b[53] | N/A | N/A |
| Prostate, advanced | Incidence | 1 publication: Lynch 2014b[53] | N/A | 1 publication: Lynch 2014b[53] | N/A |
| Kidney | Incidence | 2 publications: George 2011[41];Hunter 2020[78] | 2 publications: George 2011[41];Hunter 2020[78] | N/A | N/A |
| Bladder | Incidence | 2 publications: Koebnick 2008[37];Hunter 2020[78] | 2 publications: in high vs low Koebnick 2008[37];Hunter 2020[78] | N/A | N/A |
| Brain | Incidence | 1 publication: Hunter 2020[78] | N/A | 1 publication: Hunter 2020[78] | N/A |
| Thyroid | Incidence | 1 publication: Hunter 2020[78] | N/A | 1 publication: Hunter 2020[78] | N/A |
| Non-Hodgkin Lymphoma | Incidence | 1 publication: Hunter 2020[78] | N/A | 1 publication: Hunter 2020[78] | N/A |
| **Television watching frequency** | | | | | |
| Colorectal | Incidence | 1 publication: Cuthbertson 2020[80] | N/A | 1 publication: Cuthbertson 2020[80] | N/A |
| Lung | Incidence | 1 publication: Cuthbertson 2020[80] | N/A | 1 publication: Cuthbertson 2020[80] | N/A |
| Breast, postmenopausal | Incidence | 1 publication: Cuthbertson 2020[80] | N/A | 1 publication: Cuthbertson 2020[80] | N/A |
| Prostate | Incidence | 1 publication: Cuthbertson 2020[80] | N/A | 1 publication: Cuthbertson 2020[80] | N/A |

### Supplementary Table 8: Descriptive table of the included observational studies of general sedentary time and cancer risk.

| Author, Year Country PMID | Study name, type Age, sex, ethnicity, characteristics of participants | Cases/ Study size Median follow-up (years) | Case ascertainment | Exposure Assessment, detail | Outcome Sub-population | Comparison | RR (95%CI) Ptrend | Adjustments | | | | | | | | | | | |
|---|---|---|---|---|---|---|---|---|---|---|---|---|---|---|---|---|---|---|---|
| | | | | | | | | Age | Sex | Smo | Alc | Obe | Phy | Diet | Med | Mens | Repr | Oth | Skin |
| **Pancreatic cancer** | | | | | | | | | | | | | | | | | | | |
| Zeng 2023 UK (38066552)[92] | UK Biobank Prospective Cohort Mean age:57 years M/F | 1,129 / 340,631 13.05 years | National cancer registries, hospital inpatient records | Self-reported in a touchscreen questionnaire<br><br>**Sedentary time:** Sum of driving, watching TV, and computer using, as part of the healthy lifestyle score | Incidence, Pancreas cancer<br><br>Men | Favourable (<3 h/d) vs Unfavourable (≥5 h/d) | 0.70 (0.53 - 0.92) | ✓ | ✓ | | | | | | | | | ✓ | |
| | | | | | Women | | 0.78 (0.60 - 1.02) | ✓ | ✓ | | | | | | | | | ✓ | |
| **Skin cancer** | | | | | | | | | | | | | | | | | | | |
| Weber 2021 UK (34059803)[83] | UK Biobank Prospective Cohort Range:38-73 years M/F | 1,239 / 350,512 7 years | National Health Service, central registers | Self-reported in a touchscreen questionnaire<br><br>**Total sedentary behaviour**: sum of time spent watching TV, using a computer during leisure time and driving for transportation | Incidence, melanoma | Per 3 hours/ day | 1.04 (0.97 - 1.12) Ptrend:0.306 | ✓ | ✓ | ✓ | ✓ | | ✓ | ✓ | | | | ✓ | ✓ |
| | | 372 | | | Low UV | | 1.09 (0.96 - 1.24) Ptrend:0.178 | ✓ | ✓ | ✓ | ✓ | | ✓ | ✓ | | | | ✓ | ✓ |
| | | 846 | | | High UV | | 1.00 (0.92 - 1.10) Ptrend:0.935 | ✓ | ✓ | ✓ | ✓ | | ✓ | ✓ | | | | ✓ | ✓ |
| | | 680 | | | Men | | 1.01 (0.92 - 1.11) Ptrend:0.755 | ✓ | ✓ | ✓ | ✓ | | ✓ | ✓ | | | | ✓ | ✓ |
| | | 559 | | | Women | | 1.07 (0.97 - 1.19) Ptrend:0.161 | ✓ | ✓ | ✓ | ✓ | | ✓ | ✓ | | | | ✓ | ✓ |
| | | 507 | | | chronically UV exposed | | 0.99 (0.88 - 1.11) Ptrend:0.178 | ✓ | ✓ | ✓ | ✓ | | ✓ | ✓ | | | | ✓ | ✓ |
| | | 712 | | | intermittently UV exposed | | 1.07 (0.97 - 1.18) Ptrend:0.164 | ✓ | ✓ | ✓ | ✓ | | ✓ | ✓ | | | | ✓ | ✓ |
| | | 232 | | | Men and low UV sensitivity | | 1.07 (0.92 - 1.25) Ptrend:0.403 | ✓ | ✓ | ✓ | ✓ | | ✓ | ✓ | | | | ✓ | ✓ |
| | | 434 | | | Men and high UV sensitivity | | 0.98 (0.87 - 1.10) Ptrend:0.747 | ✓ | ✓ | ✓ | ✓ | | ✓ | ✓ | | | | ✓ | ✓ |
| | | 140 | | | Women and low UV sensitivity | | 1.11 (0.92 - 1.35) Ptrend:0.264 | ✓ | ✓ | ✓ | ✓ | | ✓ | ✓ | | | | ✓ | ✓ |

| Author, Year Country PMID | Study name, type Age, sex, ethnicity, characteristics of participants | Cases/ Study size Median follow-up (years) | Case ascertainment | Exposure Assessment, detail | Outcome Sub-population | Comparison | RR (95%CI) Ptrend | Adjustments | | | | | | | | | | | |
|---|---|---|---|---|---|---|---|---|---|---|---|---|---|---|---|---|---|---|---|
| | | | | | | | | Age | Sex | Smo | Alc | Obe | Phy | Diet | Med | Mens | Repr | Oth | Skin |
| | | 412 | | | Women and high UV sensitivity | | 1.04 (0.93 - 1.17) Ptrend:0.500 | ✓ | ✓ | ✓ | ✓ | | ✓ | ✓ | | | | ✓ | ✓ |
| **Breast cancer** | | | | | | | | | | | | | | | | | | | |
| Tomida 2024 Japan (38041484)[91] | Japan Multi-Institutional Collaborative Cohort Study (J-MICC) Prospective Cohort Range:35-69 years F | 554 / 36,023 9.41 years | cancer registries confirmed by medical records | , Self-administered lifestyle questionnaire **Daily sedentary time**: 24 hours minus time spent on sleeping, hard labour, walking, and standing | Incidence, Breast cancer | >13 vs <7 hours/day | 1.34 (1.02 - 1.75) Ptrend:0.046 | ✓ | | ✓ | ✓ | ✓ | ✓ | | | ✓ | ✓ | ✓ | |
| | | | | | | Per 1 hour | 1.02 (1.00 - 1.04) Ptrend:0.102 | ✓ | | ✓ | ✓ | ✓ | ✓ | | | ✓ | ✓ | ✓ | |
| | | 174 | | , | Incidence, Breast cancer, premenopausal | >13 vs <7 hours/day | 1.64 (1.03 - 2.60) Ptrend:0.037 | ✓ | | ✓ | ✓ | ✓ | ✓ | | | ✓ | ✓ | ✓ | |
| | | | | | | Per 1 hour | 1.03 (1.00 - 1.08) Ptrend:0.080 | ✓ | | ✓ | ✓ | ✓ | ✓ | | | ✓ | ✓ | ✓ | |
| | | 276 | | | Incidence, Breast cancer, postmenopausal | >13 vs <7 hours/day | 1.22 (0.87 - 1.70) Ptrend:0.251 | ✓ | | ✓ | ✓ | ✓ | ✓ | | | ✓ | ✓ | ✓ | |
| | | | | | | Per 1 hour | 1.01 (0.98 - 1.04) Ptrend:0.080 | ✓ | | ✓ | ✓ | ✓ | ✓ | | | ✓ | ✓ | ✓ | |
| | | 31 | | | BMI <18.5 | >13 vs <7 hours/day | 1.40 (0.50 - 3.93) Ptrend:0.629 | ✓ | | ✓ | ✓ | | ✓ | | | ✓ | ✓ | ✓ | |
| | | 401 | | | BMI 18.5-25 kg/m2 | | 1.23 (0.90 - 1.69) Ptrend:0.159 | ✓ | | ✓ | ✓ | | ✓ | | | ✓ | ✓ | ✓ | |
| | | 122 | | | BMI >25 | | 1.78 (0.96 - 3.32) Ptrend:0.165 | ✓ | | ✓ | ✓ | | ✓ | | | ✓ | ✓ | ✓ | |
| | | 93 | | | Smokers | | 1.21 (0.67 - 2.20) Ptrend:0.465 | ✓ | | | ✓ | ✓ | ✓ | | | ✓ | ✓ | ✓ | |
| | | 460 | | | Non smokers | | 1.36 (1.01 - 1.85) Ptrend:0.076 | ✓ | | ✓ | ✓ | ✓ | ✓ | | | ✓ | ✓ | ✓ | |

**Adjustment groups: Age**: age; **Alc**: alcohol; **Diet**: dietary factors and energy intake; **Med**: medical factors (e.g. cancer-related infections, diseases in the breast, digestive tract, gallbladder, liver, or pancreas, cancer screening, precancerous conditions, or pre-existing skin cancer); **Mens**: menstrual characteristics (e.g. age at menarche, menopausal status/duration, age at menopause, or type of menopause); **Obe**: obesity (adiposity measures); **Oth**: others (e.g. family history/genetic factor, race/ethnicity, ancestry, socioeconomic factors, or marriage); **Phy**: physical activity; **Repr**: reproductive or hormonal factors (e.g. parity, age at first/last birth, postmenopausal hormone use, or oral contraceptive use); **Sex**: sex; **Skin**: skin-related factors (e.g. UV light/sun exposure, sunburns, time spent outdoors, sunscreen use, indoor tanning, moles, fair skin, red/blond hair, blue/green eyes, or freckles); **Smo**: smoking, cancer-related

#### Supplementary Table 9: Descriptive table of the included observational studies of sitting time and cancer risk.

| Author, Year Country PMID | Study name, type Age, sex, ethnicity, characteristics of participants | Cases/ Study size Median follow-up (years) | Case ascertainment | Exposure Assessment, detail | Outcome Sub-population | Compariso n | RR (95%CI) Ptrend | Adjustments Age | Sex | Smo | Alc | Obe | Phy | Diet | Med | Mens | Repr | Oth | Skin |
|---|---|---|---|---|---|---|---|---|---|---|---|---|---|---|---|---|---|---|---|
| **Head and Neck cancer** | | | | | | | | | | | | | | | | | | | |
| Patel 2015 USA (26126627)[56] | CPS II Nutrition Cohort<br>Prospective Cohort<br>Age:63 years<br>M/F<br>Mostly white | 274 / 146,722<br>17 years | Cancer registry and death certificates and medical records | Self-reported questionnaire, **Sitting**:<br>Leisure-time spent sitting | Incidence / Mortality, Head and Neck cancer, Men | >6 vs <3 hours/day | 1.22 (0.88 - 1.69) | ✓ | | ✓ | ✓ | | ✓ | ✓ | | | | ✓ | |
| | | 97 | | | Women | | 1.46 (0.84 - 2.54) | ✓ | | ✓ | ✓ | | ✓ | ✓ | | ✓ | ✓ | ✓ | |
| | | 274 | | | Men | | 1.22 (0.88 - 1.69) | ✓ | | ✓ | ✓ | ✓ | ✓ | ✓ | | | | ✓ | |
| | | 97 | | | Women | | 1.49 (0.86 - 2.61) | ✓ | | ✓ | ✓ | ✓ | ✓ | ✓ | | ✓ | ✓ | ✓ | |
| **Oesophageal cancer** | | | | | | | | | | | | | | | | | | | |
| Cook 2013 USA (24367697)[52] | NIH- AARP Diet and Health Study<br>Prospective Cohort<br>Range:50-71 years<br>M/F<br>Retired | 377 / 303,033 | Postal Service, Social Security Administration Death Master File, National Death Index | Self-reported questionnaire, **Sitting**:<br>Typical number of hours per day siting during the last 12 months | Incidence, oesophageal adenocarcinoma | >9 vs <3 hours/day | 0.69 (0.41 - 1.15) | ✓ | ✓ | ✓ | ✓ | | | ✓ | | | | ✓ | |
| | | 128 / 303,033 | | | Incidence, oesophageal squamous cell carcinoma | | 0.86 (0.39 - 1.88) | ✓ | ✓ | ✓ | ✓ | | | ✓ | | | | ✓ | |
| Patel 2015 USA (26126627)[56] | CPS II Nutrition Cohort<br>Prospective Cohort<br>Age:63 years<br>M/F<br>Mostly white | 270 / 146,722<br>17 years | Cancer registry and death certificates and medical records | Self-reported questionnaire, **Sitting**:<br>Leisure-time spent sitting | Incidence / Mortality, Oesophageal cancer, Men | >6 vs <3 hours/day | 1.05 (0.75 - 1.48) | ✓ | | ✓ | ✓ | | ✓ | ✓ | | | | ✓ | |
| | | 45 | | | Women | | 1.14 (0.48 - 2.74) | ✓ | | ✓ | ✓ | | ✓ | ✓ | | ✓ | ✓ | ✓ | |
| | | 270 | | | Men | | 1.04 (0.74 - 1.46) | ✓ | | ✓ | ✓ | ✓ | ✓ | ✓ | | | | ✓ | |
| | | 45 | | | Women | | 1.13 (0.47 - 2.72) | ✓ | | ✓ | ✓ | ✓ | ✓ | ✓ | | ✓ | ✓ | ✓ | |
| Ihira 2020 Japan (31925977)[77] | Japan Public Health Center-based Prospective Study<br>Prospective Cohort<br>M/F<br>Japanese | 100 / 33,307<br>10.2 years | | Self-reported questionnaire, **Sedentary**:<br>Daily occupational sitting time | Incidence, oesophageal cancer, Men | ≥7 vs 1-<3 h/d hour/day | 1.05 (0.58 - 1.87)<br>Ptrend:0.924 | ✓ | | ✓ | ✓ | ✓ | ✓ | ✓ | | | | | |
| | | 8 | | | Women | <1 h/d vs 1-<3 h/d hour/day | 1.64 (0.25 - 10.75) | ✓ | | ✓ | ✓ | ✓ | ✓ | ✓ | | | | | |
| | | 13 | | Daily occupational sitting time including home duties | Women | | 1.68 (0.25 - 10.75) | ✓ | | ✓ | ✓ | ✓ | ✓ | ✓ | | | | | |
| **Stomach cancer** | | | | | | | | | | | | | | | | | | | |

| Author, Year Country PMID | Study name, type Age, sex, ethnicity, characteristics of participants | Cases/ Study size Median follow-up (years) | Case ascertainment | Exposure Assessment, detail | Outcome Sub-population | Comparison | RR (95%CI) Ptrend | Adjustments | | | | | | | | | | | |
|---|---|---|---|---|---|---|---|---|---|---|---|---|---|---|---|---|---|---|---|
| | | | | | | | | Age | Sex | Smo | Alc | Obe | Phy | Diet | Med | Mens | Repr | Oth | Skin |
| Cook 2013 USA (24367697)[52] | NIH- AARP Diet and Health Study<br>Prospective Cohort<br>Range:50-71 years<br>M/F<br>Retired | 255 / 303,033 | Postal Service, Social Security Administration Death Master File, National Death Index | Self-reported questionnaire, **Sitting**: Typical number of hours per day siting during the last 12 months | Incidence, Gastric cardia adenocarcinoma | >9 vs <3 hours/day | 1.00 (0.62 - 1.63) | ✓ | ✓ | ✓ | ✓ | | | ✓ | | | | ✓ | |
| | | 277 | | | Incidence, Gastric noncardia adenocarcinoma | | 0.81 (0.45 - 1.45) | ✓ | ✓ | ✓ | ✓ | | | ✓ | | | | ✓ | |
| Patel 2015 USA (26126627)[56] | CPS II Nutrition Cohort<br>Prospective Cohort<br>Age:63 years<br>M/F<br>Mostly white | 226 / 146,722<br>17 years | Cancer registry and death certificates and medical records | Self-reported questionnaire, **Sitting**: Leisure-time spent sitting | Incidence / Mortality, Stomach cancer, Men | >6 vs <3 hours/day | 1.04 (0.70 - 1.55) | ✓ | | ✓ | ✓ | | ✓ | ✓ | | | | ✓ | |
| | | 80 | | | Women | | 1.02 (0.54 - 1.96) | ✓ | | ✓ | ✓ | | ✓ | ✓ | | ✓ | ✓ | ✓ | |
| | | 226 | | | Men | | 1.05 (0.71 - 1.55) | ✓ | | ✓ | ✓ | ✓ | ✓ | ✓ | | | | ✓ | |
| | | 80 | | | Women | | 1.06 (0.55 - 2.03) | ✓ | | ✓ | ✓ | ✓ | ✓ | ✓ | | ✓ | ✓ | ✓ | |
| Ihira 2020 Japan (31925977)[77] | Japan Public Health Center-based Prospective Study<br>Prospective Cohort<br>M/F<br>Japanese | 552 / 33,307<br>10.2 years | | Self-reported questionnaire, **Sedentary**: Daily occupational sitting time | Incidence, Stomach cancer, Men | ≥7 vs 1-<3 h/d hour/day | 1.08 (0.82 - 1.41) Ptrend:0.568 | ✓ | | ✓ | ✓ | ✓ | ✓ | ✓ | | | | | |
| | | 127 | | | Women | | 1.03 (0.59 - 1.81) Ptrend:0.696 | ✓ | | ✓ | ✓ | ✓ | ✓ | ✓ | | | | | |
| | | 216 | | Daily occupational sitting time including home duties | Women | | 1.14 (0.73 - 1.75) Ptrend:0.533 | ✓ | | ✓ | ✓ | ✓ | ✓ | ✓ | | | | | |
| **Colorectal cancer** | | | | | | | | | | | | | | | | | | | |
| Park 2019 USA (30910780)[73] | Multiethnic Cohort<br>Prospective Cohort<br>Range:45-75 years<br>M/F<br>Multi-ethnic | 2,341 / 172,502<br>16.8 years | Cancer registry and national death index | Self-reported questionnaire, **Sitting**: | Incidence, colorectal cancer, Men | >11 vs <5 hrs/day | 0.96 (0.84 - 1.10) Ptrend:0.95 | ✓ | | ✓ | ✓ | ✓ | | ✓ | ✓ | | | ✓ | |
| | | 1,955 | | | Women | | 0.92 (0.80 - 1.07) Ptrend:0.39 | ✓ | | ✓ | ✓ | ✓ | | ✓ | ✓ | | ✓ | ✓ | |
| Nunez 2018 Australia (29510753)[66] | Prospective Cohort<br>Age:62 years<br>M/F | 765 / 226,584<br>2.7 years | Cancer registry, hospital admission and death certificate | Self-administered Questionnaire, **Sitting time**: | Incidence, colon cancer | > 8 - vs 0 - 2.99 hrs/day | 1.02 (0.79 - 1.32) | ✓ | ✓ | ✓ | ✓ | | ✓ | ✓ | | | | ✓ | |
| | | NA | | | Incidence, rectal cancer | | 0.90 (0.61 - 1.32) | ✓ | ✓ | ✓ | ✓ | | ✓ | ✓ | | | | ✓ | |
| Rangul 2018 Norway (30352079)[70] | The Nord-Trøndelag Health Study (HUNT)<br>Prospective Cohort<br>M/F | 388 / 37,810<br>16 years | Cancer Registry of Norway | Self-reported questionnaire, **Sitting time**: | Incidence, colorectal cancer, Men | <8 vs >8 hours/day | 1.14 (0.91 - 1.43) Ptrend:0.26 | ✓ | | ✓ | ✓ | ✓ | | | | | | ✓ | |
| | | 255 | | | Women | | 1.02 (0.75 - 1.39) Ptrend:0.91 | ✓ | | ✓ | ✓ | ✓ | | | | | | ✓ | |

| Author, Year Country PMID | Study name, type Age, sex, ethnicity, characteristics of participants | Cases/ Study size Median follow-up (years) | Case ascertainment | Exposure Assessment, detail | Outcome Sub-population | Comparison | RR (95%CI) Ptrend | Adjustments | | | | | | | | | | | |
|---|---|---|---|---|---|---|---|---|---|---|---|---|---|---|---|---|---|---|---|
| | | | | | | | | Age | Sex | Smo | Alc | Obe | Phy | Diet | Med | Mens | Repr | Oth | Skin |
| Gorczyca 2017 USA (28538039)[61] | Women's Health Initiative (WHI) Observational Study Prospective Cohort Range:50-79 years F Mostly white | 1,145 / 74,870 13.4 years | self-report + pathology reports | Self-reported questionnaire, **Sitting time**: | Incidence, colorectal cancer | ≥10 vs ≤5 h/day | 1.15 (0.95 - 1.40) Ptrend:0.16 | ✓ | | ✓ | ✓ | | ✓ | ✓ | ✓ | | ✓ | ✓ | |
| | | 1,145 | | | Incidence, colorectal cancer | | 1.12 (0.92 - 1.35) Ptrend:0.29 | ✓ | | ✓ | ✓ | ✓ | ✓ | ✓ | ✓ | | ✓ | ✓ | |
| | | 964 | | | Colon cancer | | 1.14 (0.92 - 1.40) Ptrend:0.25 | ✓ | | ✓ | ✓ | ✓ | ✓ | ✓ | ✓ | | ✓ | ✓ | |
| | | 964 | | | Colon cancer | | 1.17 (0.95 - 1.45) Ptrend:0.14 | ✓ | | ✓ | ✓ | | ✓ | ✓ | ✓ | | ✓ | ✓ | |
| | | 202 | | | Rectal cancer | | 0.94 (0.59 - 1.48) Ptrend:0.74 | ✓ | | ✓ | ✓ | ✓ | ✓ | ✓ | ✓ | | ✓ | ✓ | |
| | | 202 | | | Rectal cancer | | 0.97 (0.61 - 1.53) Ptrend:0.85 | ✓ | | ✓ | ✓ | | ✓ | ✓ | ✓ | | ✓ | ✓ | |
| | | NA | | | colorectal cancer, normal BMI | | 0.86 (0.61 - 1.21) Ptrend:0.43 | | | ✓ | ✓ | | ✓ | ✓ | ✓ | | ✓ | ✓ | |
| | | NA | | | colorectal cancer, Overweight | | 1.34 (0.96 - 1.87) Ptrend:0.09 | | | ✓ | ✓ | | ✓ | ✓ | ✓ | | ✓ | ✓ | |
| | | NA | | | colorectal cancer, Obese | | 1.22 (0.86 - 1.73) Ptrend:0.30 | | | ✓ | ✓ | | ✓ | ✓ | ✓ | | ✓ | ✓ | |
| Howard 2008 USA (18437512)[36] | NIH- AARP Diet and Health Study Prospective Cohort Age:62 years M/F Multi-ethnic | 1,264 / 488,720 6.9 years | Cancer registry | Self-reported questionnaire, **Sitting time**: Time spent sitting, over past 12 months, from 2nd questionnaire | Incidence / Mortality, colon cancer, Men | 9 - vs 0 - 2.9 hours/day | 1.24 (0.98 - 1.57) Ptrend:0.050 | ✓ | | ✓ | ✓ | | ✓ | ✓ | | | | ✓ | |
| | | 667 | | | Women | | 1.24 (0.90 - 1.70) Ptrend:0.361 | ✓ | | ✓ | ✓ | | ✓ | ✓ | | | ✓ | ✓ | |
| | | 1,264 | | | Men | | 1.22 (0.96 - 1.55) Ptrend:0.073 | ✓ | | ✓ | ✓ | ✓ | ✓ | ✓ | | | | ✓ | |
| | | 667 | | | Women | | 1.23 (0.89 - 1.70) Ptrend:0.382 | ✓ | | ✓ | ✓ | ✓ | ✓ | ✓ | | | ✓ | ✓ | |
| Patel 2015 USA (26126627)[56] | CPS II Nutrition Cohort Prospective Cohort Age:63 years M/F Mostly white | 1,447 / 146,722 17 years | Cancer registry and death certificates and medical records | Self-reported questionnaire, **Sitting**: Leisure-time spent sitting | Incidence / Mortality, colorectal cancer, Men | >6 vs <3 hours/day | 1.03 (0.89 - 1.19) | ✓ | | ✓ | ✓ | | ✓ | ✓ | ✓ | | | ✓ | |
| | | 1,447 | | | Men | | 1.01 (0.87 - 1.18) | ✓ | | ✓ | ✓ | ✓ | ✓ | ✓ | ✓ | | | ✓ | |
| | | 1,199 | | | Women | | 0.97 (0.81 - 1.16) | ✓ | | ✓ | ✓ | | ✓ | ✓ | ✓ | ✓ | ✓ | ✓ | |
| | | 1,199 | | | Women | | 0.95 (0.79 - 1.14) | ✓ | | ✓ | ✓ | ✓ | ✓ | ✓ | ✓ | ✓ | ✓ | ✓ | |
| Nguyen 2018 USA (30740587)[71] | NHS II Prospective Cohort Age:45 years F nurses | 118 / 89,278 13.9 years | Self report verified by medical record | Self-reported questionnaire, **Sitting work**: sitting at work or away from home or driving | Incidence, colorectal cancer, diagnosed before 50y | >14 vs <7 hours/week | 1.07 (0.70 - 1.65) Ptrend:0.46 | | | ✓ | ✓ | | ✓ | ✓ | ✓ | ✓ | ✓ | ✓ | |
| | | 118 | | | | | 1.05 (0.68 - 1.62) Ptrend:0.52 | | | ✓ | ✓ | ✓ | ✓ | ✓ | ✓ | ✓ | ✓ | ✓ | |

| Author, Year Country PMID | Study name, type Age, sex, ethnicity, characteristics of participants | Cases/ Study size Median follow-up (years) | Case ascertainment | Exposure Assessment, detail | Outcome Sub-population | Comparison | RR (95%CI) Ptrend | Adjustments Age | Sex | Smo | Alc | Obe | Phy | Diet | Med | Mens | Repr | Oth | Skin |
|---|---|---|---|---|---|---|---|---|---|---|---|---|---|---|---|---|---|---|---|
| Guo 2020 China (31773920)[76] | Kailuan cohort<br>Prospective Cohort<br>Age:59 years<br>M | 353 / 92,923<br>10 years | self report verified by medical records or by linkage with state cancer registries | Interview,<br>**Sitting time**:<br>occupation sitting time occupation | Incidence, colorectal cancer | <4 vs >8 hours/day | 2.99 (0.96 - 9.37) | ✓ | | | ✓ | ✓ | | | | | | | |
| Ihira 2020 Japan (31925977)[77] | Japan Public Health Center-based Prospective Study<br>Prospective Cohort<br>M/F<br>Japanese | 552 | | Self-reported questionnaire,<br>**Sedentary**:<br>Daily occupational sitting time | Incidence, colorectal cancer, Men | ≥7 vs 1-<3 hour/day | 1.17 (0.91 - 1.52)<br>Ptrend:0.319 | ✓ | | ✓ | ✓ | ✓ | ✓ | ✓ | | | | | |
| | | 212 | | | Women | | 1.03 (0.59 - 1.81)<br>Ptrend:0.696 | ✓ | | ✓ | ✓ | ✓ | ✓ | ✓ | | | | | |
| | | 362 | | | colon cancer, Men | | 1.19 (0.87 - 1.65)<br>Ptrend:0.356 | ✓ | | ✓ | ✓ | ✓ | ✓ | ✓ | | | | | |
| | | 159 | | | Women | | 0.91 (0.52 - 1.60)<br>Ptrend:0.967 | ✓ | | ✓ | ✓ | ✓ | ✓ | ✓ | | | | | |
| | | 193 | | | rectal cancer, Men | | 1.13 (0.73 - 1.75)<br>Ptrend:0.697 | ✓ | | ✓ | ✓ | ✓ | ✓ | ✓ | | | | | |
| | | 53 | | | Women | | 1.00 (0.37 - 2.69)<br>Ptrend:0.821 | ✓ | | ✓ | ✓ | ✓ | ✓ | ✓ | | | | | |
| Keum 2016 USA (26649988)[58] | Nurses Health Study (NHS) + Health Professionals Follow-up Study (HPFS)<br>Prospective Cohort<br>Range:30-75 years<br>M/F<br>Health professionals | 774 / 106,521<br>22 years | Self report verified by medical record | Self-reported questionnaire,<br>**Sitting work**:<br>sitting time at work | Incidence, colorectal cancer, Men | >21 vs 0-6.9 hours/week | 1.08 (0.84 - 1.37)<br>Ptrend:0.90 | ✓ | | ✓ | ✓ | | ✓ | ✓ | ✓ | | | ✓ | |
| | | 732 | | | Men | Per 7 hours week | 1.00 (0.94 - 1.06) | ✓ | | ✓ | ✓ | | ✓ | ✓ | ✓ | | | ✓ | |
| | | 1,065 | | | Women | >21 vs 0-6.9 hours/week | 1.14 (0.94 - 1.38)<br>Ptrend:0.48 | ✓ | | ✓ | ✓ | | ✓ | ✓ | ✓ | ✓ | ✓ | ✓ | |
| | | 1,065 | | | Women | Per 7 hours week | 1.01 (0.98 - 1.05) | ✓ | | ✓ | ✓ | | ✓ | ✓ | ✓ | ✓ | ✓ | ✓ | |
| | | 774 | | | Men | >21 vs 0-6.9 hours/week | 1.08 (0.85 - 1.38)<br>Ptrend:0.91 | ✓ | | ✓ | ✓ | ✓ | ✓ | ✓ | ✓ | | | ✓ | |
| | | 732 | | | Men | Per 7 hours week | 1.00 (0.94 - 1.06) | ✓ | | ✓ | ✓ | ✓ | ✓ | ✓ | ✓ | | | ✓ | |
| | | 1,065 | | | Women | >21 vs 0-6.9 hours/week | 1.13 (0.93 - 1.37)<br>Ptrend:0.53 | ✓ | | ✓ | ✓ | ✓ | ✓ | ✓ | ✓ | ✓ | ✓ | ✓ | |
| | | 1,065 | | | Women | Per 7 hours week | 1.01 (0.97 - 1.05) | ✓ | | ✓ | ✓ | ✓ | ✓ | ✓ | ✓ | ✓ | ✓ | ✓ | |
| Simons 2013 Netherlands (23420352)[45] | The Netherlands Cohort study<br>Prospective Cohort<br>Range:55-69 years<br>M/Fs | 1,033 / 4,416<br>16 years | Cancer registry | Self-reported questionnaire,<br>**Sitting time**:<br>Sitting time at longest held job, hours/day | Incidence, colon cancer, Men | <2 vs >6-8 hours/day | 0.72 (0.58 - 0.89)<br>Ptrend:0.003 | ✓ | | ✓ | ✓ | ✓ | | ✓ | | | | ✓ | |
| | | 456 | | | Proximal cancer, Men | | 0.84 (0.63 - 1.12)<br>Ptrend:0.25 | ✓ | | ✓ | ✓ | ✓ | | ✓ | | | | ✓ | |
| | | 535 | | | Distal colon cancer, Men | | 0.63 (0.48 - 0.83)<br>Ptrend:0.001 | ✓ | | ✓ | ✓ | ✓ | | ✓ | | | | ✓ | |

| Author, Year Country PMID | Study name, type Age, sex, ethnicity, characteristics of participants | Cases/ Study size Median follow-up (years) | Case ascertainment | Exposure Assessment, detail | Outcome Sub-population | Comparison | RR (95%CI) Ptrend | Adjustments: Age | Sex | Smo | Alc | Obe | Phy | Diet | Med | Mens | Repr | Oth | Skin |
|---|---|---|---|---|---|---|---|---|---|---|---|---|---|---|---|---|---|---|---|
| | | 402 | | | Rectal cancer, Men | | 1.10 (0.82 - 1.47) Ptrend:0.49 | ✓ | | ✓ | ✓ | ✓ | | ✓ | | | | ✓ | |
| Gerhardsson 1986 Sweden (3962961)[94] | Sweden 1960 census study Prospective Cohort Range:20-64 years M working men | 7,115 / 1,055,715 19 years | population | Self-reported questionnaire, **Sitting time**: 95% confidence interval=90% confidence interval. Sedentary is sitting 50% of the time of more | Incidence, colon cancer | Sedentary job (sitting >50% of time) vs physically active job ((sitting <50% of time) | 1.30 (1.20 - 1.50) | ✓ | | | | | | | | | | ✓ | |
| | | 1,775 / 1,055,715 19 years | | | Incidence, Cecum ascending colon | | 1.50 (1.20 - 1.80) | ✓ | | | | | | | | | | ✓ | |
| | | 774 | | | Transverse and flexures, | | 1.60 (1.20 - 2.10) | ✓ | | | | | | | | | | ✓ | |
| | | 330 | | | Descending colon cancer | | 1.30 (0.90 - 1.90) | ✓ | | | | | | | | | | ✓ | |
| | | 2,221 | | | Sigmoid cancer | | 1.20 (1.00 - 1.40) | ✓ | | | | | | | | | | ✓ | |
| | | 4,533 / 1,055,715 19 years | | | rectal cancer | | 1.10 (1.00 - 1.20) | ✓ | | | | | | | | | | ✓ | |
| Keum 2016 USA (26649988)[58] | Nurses Health Study (NHS) + Health Professionals Follow-up Study (HPFS) Prospective Cohort Range:30-75 years M/F Health professionals | 771 / 106,521 22 years | Self report verified by medical record | Self-reported questionnaire, **Sitting time**: sitting time driving | Incidence, colorectal cancer, Men | >14 vs 0-3.4 hours/week | 1.04 (0.69 - 1.58) Ptrend:0.65 | ✓ | | ✓ | ✓ | | ✓ | ✓ | ✓ | | | ✓ | |
| | | 771 | | | Men | Per 7 hours week | 0.97 (0.84 - 1.11) | ✓ | | ✓ | ✓ | | ✓ | ✓ | ✓ | | | ✓ | |
| | | 771 | | | Men | >14 vs 0-3.4 hours/week | 1.01 (0.66 - 1.52) Ptrend:0.46 | ✓ | | ✓ | ✓ | ✓ | ✓ | ✓ | ✓ | | | ✓ | |
| | | 771 | | | Men | Per 7 hours week | 0.95 (0.82 - 1.09) | ✓ | | ✓ | ✓ | ✓ | ✓ | ✓ | ✓ | | | ✓ | |
| Ihira 2020 Japan (31925977)[77] | Japan Public Health Center-based Prospective Study Prospective Cohort M/F Japanese | 353 / 33,307 10.2 years | | Self-reported questionnaire, **Sedentary**: Daily occupational sitting time in women including home duties | Incidence, colorectal cancer, Women | ≥7 vs 1-<3 hour/day | 1.02 (0.70 - 1.50) Ptrend:0.695 | ✓ | | ✓ | ✓ | ✓ | ✓ | ✓ | | | | | |
| | | 261 | | | Colon cancer, Women | | 0.96 (0.60 - 1.51) Ptrend:0.948 | ✓ | | ✓ | ✓ | ✓ | ✓ | ✓ | | | | | |
| | | 92 | | | Rectal cancer, Women | | 1.22 (0.60 - 2.48) Ptrend:0.537 | ✓ | | ✓ | ✓ | ✓ | ✓ | ✓ | | | | | |
| Nguyen 2018 USA (30740587)[71] | NHS II Prospective Cohort Age:45 years F nurses | 118 / 89,278 13.9 years | Self report verified by medical record | Self-reported questionnaire, **Sitting time**: other measures of sitting | Incidence, colorectal cancer, diagnosed before 50y | >14 vs <7 hours/week | 1.36 (0.85 - 2.18) Ptrend:0.21 | | | ✓ | ✓ | | ✓ | ✓ | ✓ | ✓ | ✓ | ✓ | |
| | | 118 | | | | | 1.34 (0.84 - 2.15) Ptrend:0.23 | | | ✓ | ✓ | ✓ | ✓ | ✓ | ✓ | ✓ | ✓ | ✓ | |

| Author, Year Country PMID | Study name, type Age, sex, ethnicity, characteristics of participants | Cases/ Study size Median follow-up (years) | Case ascertainment | Exposure Assessment, detail | Outcome Sub-population | Comparison | RR (95%CI) Ptrend | Adjustments | | | | | | | | | | | |
|---|---|---|---|---|---|---|---|---|---|---|---|---|---|---|---|---|---|---|---|
| | | | | | | | | Age | Sex | Smo | Alc | Obe | Phy | Diet | Med | Mens | Repr | Oth | Skin |
| Eaglehouse 2017 Singapore (28542077)[62] | Singapore Chinese Health study Prospective Cohort Age:56.4 years M/F Chinese | 1,981 / 61,308 16.8 years | Cancer registry | Self-reported questionnaire, **Sitting time**: Excluding when watching TV | Incidence, colorectal cancer | ≥3 vs 0 hours/day | 0.99 (0.84 - 1.17) Ptrend:0.99 | ✓ | ✓ | ✓ | ✓ | ✓ | | ✓ | | | | ✓ | |
| **Liver cancer** | | | | | | | | | | | | | | | | | | | |
| Patel 2015 USA (26126627)[56] | CPS II Nutrition Cohort Prospective Cohort Age:63 years M/F Mostly white | 171 / 146,722 17 years | Cancer registry and death certificates and medical records | Self-reported questionnaire, **Sitting**: Leisure-time spent sitting | Incidence / Mortality, liver cancer, Men | >6 vs <3 hours/day | 0.84 (0.55 - 1.29) | ✓ | | ✓ | ✓ | | ✓ | ✓ | | | | ✓ | |
| | | 79 | | | Women | | 0.77 (0.37 - 1.60) | ✓ | | ✓ | ✓ | | ✓ | ✓ | | ✓ | ✓ | ✓ | |
| | | 171 | | | Men | | 0.83 (0.54 - 1.28) | ✓ | | ✓ | ✓ | ✓ | ✓ | ✓ | | | | ✓ | |
| | | 79 | | | Women | | 0.73 (0.35 - 1.53) | ✓ | | ✓ | ✓ | ✓ | ✓ | ✓ | | ✓ | ✓ | ✓ | |
| Ihira 2020 Japan (31925977)[77] | Japan Public Health Center-based Prospective Study Prospective Cohort M/F Japanese | 134 | | Self-reported questionnaire, **Sedentary**: Daily occupational sitting time | Incidence, liver cancer, Men | ≥7 vs 1-<3 hour/day | 1.54 (0.92 - 2.58) Ptrend:0.159 | ✓ | | ✓ | ✓ | ✓ | ✓ | ✓ | | | | | |
| | | 21 | | | Women | | 0.30 (0.04 - 2.20) Ptrend:0.109 | ✓ | | ✓ | ✓ | ✓ | ✓ | ✓ | | | | | |
| | | 40 | | Daily occupational sitting time in women including home duties | Women | | 0.62 (0.14 - 2.82) Ptrend:0.685 | ✓ | | ✓ | ✓ | ✓ | ✓ | ✓ | | | | | |
| **Gallbladder cancer** | | | | | | | | | | | | | | | | | | | |
| Patel 2015 USA (26126627)[56] | CPS II Nutrition Cohort Prospective Cohort Age:63 years M/F Mostly white | 33 / 146,722 17 years | Cancer registry and death certificates and medical records | Self-reported questionnaire, **Sitting**: Leisure-time spent sitting | Incidence / Mortality, Gallbladder cancer, Men | >6 vs <3 hours/day | 2.14 (0.89 - 5.16) | ✓ | | ✓ | ✓ | | ✓ | ✓ | | | | ✓ | |
| | | 57 | | | Women | | 1.52 (0.70 - 3.32) | ✓ | | ✓ | ✓ | | ✓ | ✓ | | ✓ | ✓ | ✓ | |
| | | 33 | | | Men | | 2.11 (0.87 - 5.09) | ✓ | | ✓ | ✓ | ✓ | ✓ | ✓ | | | | ✓ | |
| | | 57 | | | Women | | 1.43 (0.65 - 3.14) | ✓ | | ✓ | ✓ | ✓ | ✓ | ✓ | | ✓ | ✓ | ✓ | |
| **Pancreatic cancer** | | | | | | | | | | | | | | | | | | | |
| Patel 2015 USA (26126627)[56] | | 425 / 146,722 17 years | | | Incidence / Mortality, Pancreas cancer, Men | >6 vs <3 hours/day | 1.17 (0.89 - 1.53) | ✓ | | ✓ | ✓ | | ✓ | ✓ | ✓ | | | ✓ | |
| | | 350 | | | Women | | 1.04 (0.75 - 1.44) | ✓ | | ✓ | ✓ | | ✓ | ✓ | ✓ | ✓ | ✓ | ✓ | |
| | | 425 | | | Men | | 1.14 (0.87 - 1.49) | ✓ | | ✓ | ✓ | ✓ | ✓ | ✓ | ✓ | | | ✓ | |
| | | 4,165 | | | Women | | 1.02 (0.73 - 1.41) | ✓ | | ✓ | ✓ | ✓ | ✓ | ✓ | ✓ | ✓ | ✓ | ✓ | |
| Ihira 2020 Japan (31925977)[77] | Japan Public Health Center-based Prospective Study Prospective Cohort | 86 / 33,307 10.2 years | | Self-reported questionnaire, **Sedentary**: | Incidence, Pancreas cancer, Men | ≥7 vs 1-<3 hour/day | 2.25 (1.17 - 4.34) Ptrend:0.021 | ✓ | | ✓ | ✓ | ✓ | ✓ | ✓ | | | | | |

| Author, Year Country PMID | Study name, type Age, sex, ethnicity, characteristics of participants | Cases/ Study size Median follow-up (years) | Case ascertainment | Exposure Assessment, detail | Outcome Sub-population | Comparison | RR (95%CI) Ptrend | Adjustments: Age | Sex | Smo | Alc | Obe | Phy | Diet | Med | Mens | Repr | Oth | Skin |
|---|---|---|---|---|---|---|---|---|---|---|---|---|---|---|---|---|---|---|---|
| | M/F<br>Japanese | | | Daily occupational sitting time | | | | | | | | | | | | | | | |
| | | 39 | | | Women | ≥7 vs 1-<3 hour/day | 0.90 (0.26 - 3.07)<br>Ptrend:0.858 | ✓ | | ✓ | ✓ | ✓ | ✓ | ✓ | | | | | |
| | | 62 | | | BMI at baseline <25 in men | ≥3 vs <3 hour/day | 1.83 (1.07 - 3.13) | ✓ | | ✓ | ✓ | | ✓ | ✓ | | | | | |
| | | 24 | | | BMI at baseline >=25 in men | | 1.53 (0.65 - 3.61) | ✓ | | ✓ | ✓ | | ✓ | ✓ | | | | | |
| | | 49 | | | , Short MVPA time (<30min/wk) in leisure time in men at baseline | | 1.74 (0.96 - 3.15) | ✓ | | ✓ | ✓ | ✓ | ✓ | ✓ | | | | | |
| | | 37 | | | Long MVPA time (≥30min/wk) in leisure time in men at baseline | | 1.80 (0.89 - 3.66) | ✓ | | ✓ | ✓ | ✓ | ✓ | ✓ | | | | | |
| Wu 2018 China (29475964)[65] | Shanghai Women's Health Study (SWHS) and the Shanghai Men's Health Study (SMHS)<br>Pooled Analysis Cohorts<br>Range:40-74 years<br>M/F | 157 / 132,488<br>16.1 years | Cancer registry and annual in-person re-interviews of surviving cohort members | Interview,<br>**Occupational physical activity**:<br>Sitting time | Incidence, Pancreas cancer, Men | low vs high | 1.22 (0.75 - 1.99)<br>Ptrend:0.48 | | | ✓ | ✓ | ✓ | | | ✓ | | | ✓ | |
| | | 222 | | | Women | low vs high | 1.06 (0.69 - 1.62)<br>Ptrend:0.85 | | | ✓ | ✓ | ✓ | | | ✓ | | | ✓ | |
| Heinen 2011 Netherlands (21955648)[43] | The Netherlands Cohort study<br>Prospective Cohort<br>Range:55-69 years<br>M/F | 162 / 120,852<br>13.3 years | Cancer registry | Self-reported questionnaire,<br>**Sitting time**:<br>longest held job | Incidence, Pancreatic cancer | <2 vs > 6 h/d | 1.25 (0.81 - 1.93)<br>Ptrend:0.28 | ✓ | | ✓ | | ✓ | | ✓ | | | | | |
| Peduzzi 2024 UK (37985251)[90] | UK Biobank<br>Nested Case Control<br>Range:37-73 years<br>M/F<br>White | / 303,461 | cancer registries | Touchscreen questionnaire,<br>**Sedentary**:<br>time spent driving | Incidence, Pancreatic cancer | Per 1 hour day | 1.05 (0.93 - 1.19) | ✓ | ✓ | | | | | | | | | | |
| Ihira 2020 Japan (31925977)[77] | Japan Public Health Center-based Prospective Study<br>Prospective Cohort<br>M/F<br>Japanese | 67 / 33,307<br>10.2 years | | Self-reported questionnaire,<br>**Sedentary**:<br>Daily occupational sitting time in women including home duties | Incidence, Pancreas cancer, Women | ≥7 vs 1-<3 hour/day | 0.67 (0.24 - 1.82)<br>Ptrend:0.626 | ✓ | | ✓ | ✓ | ✓ | ✓ | ✓ | | | | | |
| **Lung cancer** | | | | | | | | | | | | | | | | | | | |
| Jiang 2019 Norway (30859092)[72] | HUNT (North-Trondelag Health Study), Norway<br>Prospective Cohort<br>M/F | 549 / 45,810<br>18.3 years | Cancer registry | Self-reported questionnaire,<br>**Sitting time**: | Incidence, Lung cancer | >8 vs 0-4 hours/day | 1.05 (0.66 - 1.29) | ✓ | ✓ | ✓ | ✓ | ✓ | ✓ | | | | | ✓ | |
| | | 76 | | | Small cell carcinoma | | 1.18 (0.69 - 2.03) | ✓ | ✓ | ✓ | ✓ | ✓ | ✓ | | | | | ✓ | |
| | | 333 | | | Non-small cell (SCC, Adenocarcinoma, Large cell) | | 0.97 (0.74 - 1.26) | ✓ | ✓ | ✓ | ✓ | ✓ | ✓ | | | | | ✓ | |

| Author, Year Country PMID | Study name, type Age, sex, ethnicity, characteristics of participants | Cases/ Study size Median follow-up (years) | Case ascertainment | Exposure Assessment, detail | Outcome Sub-population | Comparison | RR (95%CI) Ptrend | Adjustments | | | | | | | | | | | |
|---|---|---|---|---|---|---|---|---|---|---|---|---|---|---|---|---|---|---|---|
| | | | | | | | | Age | Sex | Smo | Alc | Obe | Phy | Diet | Med | Mens | Repr | Oth | Skin |
| Rangul 2018 Norway (30352079)[70] | The Nord-Trøndelag Health Study (HUNT) Prospective Cohort M/F | 242 / 37,810 16 years | Cancer Registry of Norway | Self-reported questionnaire, **Sitting time**: | Incidence, Lung cancer, Men | < 8 vs > 8 hours/day | 1.14 (0.86 - 1.54) Ptrend:0.36 | ✓ | | ✓ | ✓ | ✓ | | | | | | ✓ | |
| | | 149 | | | Women | | 0.85 (0.55 - 1.31) Ptrend:0.46 | ✓ | | ✓ | ✓ | ✓ | | | | | | ✓ | |
| Wang 2016 USA (27439221)[59] | Womens Health Initiative (WHI) Prospective Cohort F Postmenopausal | 1,329 / 129,401 11.8 years | Medical and pathology records | Interview, **Sedentary**: sitting time | Incidence, Lung cancer | ≥10 hrs/d vs ≤5 hrs/day sitting time | 1.10 (0.95 - 1.28) | ✓ | | ✓ | ✓ | ✓ | | ✓ | ✓ | | ✓ | ✓ | |
| | | 814 / 129,401 11.8 years | Medical and pathology records | | Mortality, Lung cancer | | 1.16 (0.96 - 1.40) | ✓ | | ✓ | ✓ | ✓ | | ✓ | ✓ | | ✓ | ✓ | |
| Lam 2013 USA (23940620)[48] | NIH- AARP Diet and Health Study Prospective Cohort Age:61.8 years M/F never smokers | 206 / 158,415 9.96 years | Postal Service, Social Security Administration Death Master File, National Death Index | Self-reported questionnaire, **Sitting**: | Incidence, Lung cancer, never smokers | 5+ vs <3 hours/day | 1.28 (0.96 - 1.72) Ptrend:0.23 | ✓ | | | ✓ | | ✓ | ✓ | | | | ✓ | |
| Patel 2015 USA (26126627)[56] | CPS II Nutrition Cohort Prospective Cohort Age:63 years M/F Mostly white | 1,903 / 146,722 17 years | Cancer registry and death certificates and medical records | Self-reported questionnaire, **Sitting**: Leisure-time spent sitting | Incidence / Mortality, Lung cancer, Men | <3 vs >6 hours/day | 1.01 (0.89 - 1.15) | ✓ | | ✓ | ✓ | | ✓ | ✓ | | | | ✓ | |
| | | 1,118 | | | Women | | 0.96 (0.80 - 1.14) | ✓ | | ✓ | ✓ | | ✓ | ✓ | | ✓ | ✓ | ✓ | |
| | | 1,903 | | | Men | | 1.01 (0.89 - 1.15) | ✓ | | ✓ | ✓ | ✓ | ✓ | ✓ | | | | ✓ | |
| | | 1,118 | | | Women | | 0.98 (0.82 - 1.17) | ✓ | | ✓ | ✓ | ✓ | ✓ | ✓ | | ✓ | ✓ | ✓ | |
| Ihira 2020 Japan (31925977)[77] | Japan Public Health Center-based Prospective Study Prospective Cohort M/F Japanese | 367 / 33,307 10.2 years | | Self-reported questionnaire, **Sedentary**: Daily occupational sitting time | Incidence, Lung cancer, Men | ≥7 vs 1-<3 hour/day | 1.07 (0.75 - 1.51) Ptrend:0.685 | ✓ | | ✓ | ✓ | ✓ | ✓ | ✓ | | | | | |
| | | 81 | | | Women | | 2.80 (1.33 - 5.90) Ptrend:0.013 | ✓ | | ✓ | ✓ | ✓ | ✓ | ✓ | | | | | |
| | | 128 | | Daily occupational sitting time in women including home duties | Women | | 1.63 (0.89 - 2.99) Ptrend:0.139 | ✓ | | ✓ | ✓ | ✓ | ✓ | ✓ | | | | | |
| **Skin cancer** | | | | | | | | | | | | | | | | | | | |
| Watts 2023 UK (36795414)[88] | UK Biobank Prospective Cohort Age:61.5 years M/F | 1,734 / 81,717 6.8 years | cancer registries | Touchscreen questionnaire, **Sedentary**: | Incidence, non-melanoma | Per 20 min day | 0.99 (0.97 - 1.00) | ✓ | ✓ | ✓ | ✓ | | | | | ✓ | ✓ | ✓ | |
| Patel 2015 USA (26126627)[56] | CPS II Nutrition Cohort Prospective Cohort Age:63 years M/F Mostly white | 1,154 / 146,722 17 years | Cancer registry and death certificates and medical records | Self-reported questionnaire, **Sitting**: Leisure-time spent sitting | Incidence / Mortality, melanoma, Men | >6 vs <3 hours/day | 1.05 (0.89 - 1.24) | ✓ | | ✓ | ✓ | | ✓ | ✓ | | | | ✓ | |
| | | 769 | | | Women | | 0.99 (0.78 - 1.24) | ✓ | | ✓ | ✓ | | ✓ | ✓ | | ✓ | ✓ | ✓ | |

| Author, Year Country PMID | Study name, type Age, sex, ethnicity, characteristics of participants | Cases/ Study size Median follow-up (years) | Case ascertainment | Exposure Assessment, detail | Outcome Sub-population | Comparison | RR (95%CI) Ptrend | Adjustments | | | | | | | | | | | |
|---|---|---|---|---|---|---|---|---|---|---|---|---|---|---|---|---|---|---|---|
| | | | | | | | | Age | Sex | Smo | Alc | Obe | Phy | Diet | Med | Mens | Repr | Oth | Skin |
| | | 1,154 | | | Men | | 1.05 (0.88 - 1.24) | ✓ | | ✓ | ✓ | ✓ | ✓ | ✓ | | | | ✓ | |
| | | 769 | | | Women | | 0.99 (0.79 - 1.25) | ✓ | | ✓ | ✓ | ✓ | ✓ | ✓ | | ✓ | ✓ | ✓ | |
| **Breast cancer** | | | | | | | | | | | | | | | | | | | |
| Wang 2022 Taiwan (35159055)[86] | National Health Interview Survey (NHIS) Prospective Cohort Age:49.74 years F Taiwanese | 112 / 5,879 13.25 years | National Health Insurance Research Database and death registries and a panel of disease experts verifies diagnosis | Interview, **Sitting time**: | Incidence, Breast cancer | 3 - vs 0 - 2.9 hours/day | 1.89 (1.08 - 3.32) Ptrend:0.026 | ✓ | | | ✓ | ✓ | | ✓ | | | ✓ | | |
| Stahr 2019 USA (35117114)[85] | Arkansas Rural Community Health (ARCH) Study cohort, Prospective Cohort Range:18-100 years F | 72 / 21,665 11 years | medical records and cancer registries | Self-reported questionnaire, **Sitting**: | Incidence, Breast cancer, premenopausal | >49h/w vs <28h/w | 1.13 (0.64 - 2.01) Ptrend:0.65 | | | | | ✓ | | | | | ✓ | ✓ | |
| | | 179 | | | postmenopausal | >42h/w vs <28h/w | 1.11 (0.78 - 1.57) Ptrend:0.59 | | | | | ✓ | | | | | ✓ | ✓ | |
| Rangul 2018 Norway (30352079)[70] | The Nord-Trøndelag Health Study (HUNT) Prospective Cohort F | 513 / 19,039 16 years | Cancer Registry of Norway | Self-reported questionnaire, **Sitting time**: | Incidence, Breast cancer | High vs Low | 1.04 (0.85 - 1.27) Ptrend:0.74 | ✓ | | ✓ | ✓ | ✓ | | | | | | ✓ | |
| Nomura 2017 USA (28975422)[63] | The Women's Health Initiative Observational Study Prospective Cohort Range:50-79 years F Multi-ethnic | 4,115 / 70,233 19 years | annual health status questionnaire or by cause of death reports verified by physicians | Self-completed questionnaire, **Sitting time**: Including at work, driving or riding in car/bus, watching TV, and sitting while talking | Incidence, Breast cancer, postmenopausal | ≥10 vs ≤5 hours/day | 1.00 (0.92 - 1.09) | ✓ | | ✓ | ✓ | ✓ | ✓ | ✓ | ✓ | | ✓ | ✓ | |
| Nomura 2017 USA (28975422)[63] | | 4,115 / 70,233 19 years | | | Incidence, Breast cancer, postmenopausal | Per 1 hour day | 1.00 (0.98 - 1.02) Ptrend:0.72 | ✓ | | ✓ | ✓ | ✓ | ✓ | ✓ | ✓ | | ✓ | ✓ | |
| | | 3,333 | | | Hormone receptor positive, postmenopausal | ≥10 vs ≤5 hours/day | 1.01 (0.92 - 1.11) | ✓ | | ✓ | ✓ | ✓ | ✓ | ✓ | ✓ | | ✓ | ✓ | |
| | | 3,333 | | | Hormone receptor positive, postmenopausal | Per 1 hour day | 1.00 (0.98 - 1.02) Ptrend:0.91 | ✓ | | ✓ | ✓ | ✓ | ✓ | ✓ | ✓ | | ✓ | ✓ | |
| | | 510 | | | Hormone receptor negative, postmenopausal | ≥10 vs ≤5 hours/day | 1.03 (0.81 - 1.30) | ✓ | | ✓ | ✓ | ✓ | ✓ | ✓ | ✓ | | ✓ | ✓ | |
| | | 510 | | | Hormone receptor negative, postmenopausal | Per 1 hour day | 1.00 (0.94 - 1.06) Ptrend:0.94 | ✓ | | ✓ | ✓ | ✓ | ✓ | ✓ | ✓ | | ✓ | ✓ | |
| | | 299 | | | Triple negative, postmenopausal | ≥10 vs ≤5 hours/day | 0.86 (0.63 - 1.18) | ✓ | | ✓ | ✓ | ✓ | ✓ | ✓ | ✓ | | ✓ | ✓ | |
| | | 299 | | | Triple negative, postmenopausal | Per 1 hour day | 0.96 (0.89 - 1.04) Ptrend:0.3 | ✓ | | ✓ | ✓ | ✓ | ✓ | ✓ | ✓ | | ✓ | ✓ | |

| Author, Year Country PMID | Study name, type Age, sex, ethnicity, characteristics of participants | Cases/ Study size Median follow-up (years) | Case ascertainment | Exposure Assessment, detail | Outcome Sub-population | Comparison | RR (95%CI) Ptrend | Adjustments Age | Sex | Smo | Alc | Obe | Phy | Diet | Med | Mens | Repr | Oth | Skin |
|---|---|---|---|---|---|---|---|---|---|---|---|---|---|---|---|---|---|---|---|
| | | 1,679 | | | BMI <25 kg/m2 | ≥10 vs ≤5 hours/day | 1.07 (0.94 - 1.22) | ✓ | | ✓ | ✓ | | ✓ | ✓ | ✓ | | ✓ | ✓ | |
| | | 1,679 | | | Breast cancer, BMI <25 kg/m2 | Per 1 hour day | 1.02 (0.99 - 1.05) Ptrend:0.31 | ✓ | | ✓ | ✓ | | ✓ | ✓ | ✓ | | ✓ | ✓ | |
| | | 1,415 | | | BMI 25-29.9 | ≥10 vs ≤5 hours/day | 0.88 (0.76 - 1.02) | ✓ | | ✓ | ✓ | | ✓ | ✓ | ✓ | | ✓ | ✓ | |
| | | 1,415 | | | BMI 25-29.9 | Per 1 hour day | 0.97 (0.93 - 1.00) Ptrend:0.31 | ✓ | | ✓ | ✓ | | ✓ | ✓ | ✓ | | ✓ | ✓ | |
| | | 1,021 | | | BMI 30+ | ≥10 vs ≤5 hours/day | 1.09 (0.92 - 1.29) | ✓ | | ✓ | ✓ | | ✓ | ✓ | ✓ | | ✓ | ✓ | |
| | | 1,021 | | | BMI 30+ | Per 1 hour day | 1.01 (0.97 - 1.05) Ptrend:0.31 | ✓ | | ✓ | ✓ | | ✓ | ✓ | ✓ | | ✓ | ✓ | |
| | | 488 | | | No recreational physical activity | ≥10 vs ≤5 hours/day | 1.03 (0.81 - 1.31) | ✓ | | ✓ | ✓ | ✓ | | ✓ | ✓ | | ✓ | ✓ | |
| | | 488 | | | No recreational physical activity | Per 1 hour day | 0.99 (0.94 - 1.04) Ptrend:0.009 | ✓ | | ✓ | ✓ | ✓ | | ✓ | ✓ | | ✓ | ✓ | |
| | | 1,544 | | | Low recreational activity | ≥10 vs ≤5 hours/day | 0.92 (0.80 - 1.05) | ✓ | | ✓ | ✓ | ✓ | | ✓ | ✓ | | ✓ | ✓ | |
| | | 1,544 | | | Low recreational activity | Per 1 hour day | 0.98 (0.95 - 1.01) Ptrend:0.009 | ✓ | | ✓ | ✓ | ✓ | | ✓ | ✓ | | ✓ | ✓ | |
| | | 807 | | | Moderate physical activity | ≥10 vs ≤5 hours/day | 0.82 (0.67 - 0.99) | ✓ | | ✓ | ✓ | ✓ | | ✓ | ✓ | | ✓ | ✓ | |
| | | 807 | | | Moderate physical activity | Per 1 hour day | 0.95 (0.91 - 0.99) Ptrend:0.009 | ✓ | | ✓ | ✓ | ✓ | | ✓ | ✓ | | ✓ | ✓ | |
| | | 1,276 | | | High physical activity | ≥10 vs ≤5 hours/day | 1.24 (1.07 - 1.44) | ✓ | | ✓ | ✓ | ✓ | | ✓ | ✓ | | ✓ | ✓ | |
| | | 1,276 | | | Incidence, Breast cancer, High physical activity | Per 1 hour day | 1.05 (1.01 - 1.09) Ptrend:0.009 | ✓ | | ✓ | ✓ | ✓ | | ✓ | ✓ | | ✓ | ✓ | |
| | | 3,675 | | | Non-Hispanic white | ≥10 vs ≤5 hours/day | 1.01 (0.93 - 1.11) | ✓ | | ✓ | ✓ | ✓ | ✓ | ✓ | ✓ | | ✓ | ✓ | |
| | | 3,675 | | | Non-Hispanic white | Per 1 hour day | 1.00 (0.98 - 1.02) Ptrend:0.93 | ✓ | | ✓ | ✓ | ✓ | ✓ | ✓ | ✓ | | ✓ | ✓ | |
| | | 208 | | | Non-Hispanic black | ≥10 vs ≤5 hours/day | 0.91 (0.62 - 1.31) | ✓ | | ✓ | ✓ | ✓ | ✓ | ✓ | ✓ | | ✓ | ✓ | |
| | | 208 | | | Non-Hispanic black | Per 1 hour day | 0.99 (0.91 - 1.07) Ptrend:0.8 | ✓ | | ✓ | ✓ | ✓ | ✓ | ✓ | ✓ | | ✓ | ✓ | |
| | | 92 | | | Hispanic | ≥10 vs ≤5 hours/day | 0.82 (0.47 - 1.43) | ✓ | | ✓ | ✓ | ✓ | ✓ | ✓ | ✓ | | ✓ | ✓ | |
| | | 92 | | | Hispanic | Per 1 hour day | 0.95 (0.84 - 1.08) Ptrend:0.6 | ✓ | | ✓ | ✓ | ✓ | ✓ | ✓ | ✓ | | ✓ | ✓ | |
| | | 93 | | | Asians | ≥10 vs ≤5 hours/day | 0.60 (0.33 - 1.07) | ✓ | | ✓ | ✓ | ✓ | ✓ | ✓ | ✓ | | ✓ | ✓ | |
| | | 93 | | | Asians | Per 1 hour day | 0.86 (0.75 - 0.98) Ptrend:0.03 | ✓ | | ✓ | ✓ | ✓ | ✓ | ✓ | ✓ | | ✓ | ✓ | |
| Nomura 2016 USA (27632430)[60] | Black Women's Health Study, 1995 Prospective Cohort Range:21-69 years F African American | 1,984 / 46,734 15.4 years | self-report, linkage to cancer registries, medical and pathology records | Self-completed questionnaire, **Sitting time**: Total sitting time, at baseline 1995 | Incidence, Breast cancer | ≥10 vs <5 hours/day | 1.27 (1.06 - 1.53) Ptrend:0.01 | ✓ | | ✓ | | ✓ | ✓ | ✓ | ✓ | ✓ | ✓ | ✓ | |
| | | 752 | | | Premenopausal | | 1.08 (0.80 - 1.46) Ptrend:0.43 | ✓ | | ✓ | | ✓ | ✓ | ✓ | ✓ | ✓ | ✓ | ✓ | |

| Author, Year Country PMID | Study name, type Age, sex, ethnicity, characteristics of participants | Cases/ Study size Median follow-up (years) | Case ascertainment | Exposure Assessment, detail | Outcome Sub-population | Comparison | RR (95%CI) Ptrend | Adjustments | | | | | | | | | | | |
|---|---|---|---|---|---|---|---|---|---|---|---|---|---|---|---|---|---|---|---|
| | | | | | | | | Age | Sex | Smo | Alc | Obe | Phy | Diet | Med | Mens | Repr | Oth | Skin |
| | | 965 | | | Postmenopausal | | 1.36 (1.06 - 1.76) Ptrend:0.04 | ✓ | | ✓ | | ✓ | ✓ | ✓ | ✓ | ✓ | ✓ | ✓ | |
| | | 1,008 | | | ER+/PR+ | | 1.03 (0.79 - 1.34) Ptrend:0.87 | ✓ | | ✓ | | ✓ | ✓ | ✓ | ✓ | ✓ | ✓ | ✓ | |
| | | 433 | | | ER- & PR- | | 1.56 (1.08 - 2.25) Ptrend:0.16 | ✓ | | ✓ | | ✓ | ✓ | ✓ | ✓ | ✓ | ✓ | ✓ | |
| | | 173 | | | ER- & PR- & HER-2- | | 1.86 (1.10 - 3.15) Ptrend:0.36 | ✓ | | ✓ | | ✓ | ✓ | ✓ | ✓ | ✓ | ✓ | ✓ | |
| Catsburg 2014 Canada (24781974)[55] | Canadian Study of Diet, Lifestyle and Health<br>Case Cohort<br>F<br>Mostly white alumnae | 1,040 / 4,417<br>15 years | Cancer registry | Self-reported questionnaire, **Sitting**:<br>Time spent sitting | Incidence, Invasive breast cancer | >54 vs <12.5 hours/week | 0.98 (0.76 - 1.25) Ptrend:0.63 | | | | ✓ | ✓ | | | | ✓ | ✓ | ✓ | |
| | | 538 | | | Premenopausal | | 0.99 (0.68 - 1.43) Ptrend:0.82 | | | | ✓ | ✓ | | | | ✓ | ✓ | ✓ | |
| | | 502 | | | Postmenopausal | | 0.98 (0.69 - 1.39) Ptrend:0.54 | | | | ✓ | ✓ | | | | ✓ | ✓ | ✓ | |
| Cohen 2013 USA (23576427)[46] | Southern Community Cohort Study<br>Nested Case Control<br>Age:55 years<br>White, black, other | 455 / 2,730<br>9 years | Cancer registry | Self-reported questionnaire, **Sitting time**:<br>Total sedentary behaviour, in a car or bus, at work, watching television or seeing movies, using a computer at home, and for other reasons | Incidence, Invasive breast cancer | ≥12 vs 0 - 5.4 hours/day | 1.41 (1.01 - 1.95) Ptrend:0.60 | ✓ | | ✓ | | ✓ | ✓ | | | ✓ | ✓ | ✓ | |
| | | 260 | | | ER+/PR+ | | 2.10 (1.32 - 3.34) Ptrend:0.05 | ✓ | | ✓ | | ✓ | | | | ✓ | ✓ | ✓ | |
| | | 117 | | | ER-/PR- | | 0.80 (0.43 - 1.51) Ptrend:0.45 | ✓ | | ✓ | | ✓ | | | | ✓ | ✓ | ✓ | |
| | | 371 | | | Postmenopausal | | 1.55 (1.08 - 2.23) | ✓ | | ✓ | | ✓ | | | | ✓ | ✓ | ✓ | |
| | | 131 | | | White | | 1.94 (1.01 - 3.70) Ptrend:0.10 | ✓ | | ✓ | | ✓ | ✓ | | | ✓ | ✓ | ✓ | |
| | | 311 | | | Black | | 1.23 (0.82 - 1.83) Ptrend:0.58 | ✓ | | ✓ | | ✓ | ✓ | | | ✓ | ✓ | ✓ | |
| | | 82 | | | ER+/PR+, White | | 2.86 (1.15 - 7.11) Ptrend:0.04 | ✓ | | ✓ | | ✓ | | | | ✓ | ✓ | ✓ | |
| | | 171 | | | ER+/PR+, Black | | 1.94 (1.09 - 3.44) Ptrend:0.31 | ✓ | | ✓ | | ✓ | | | | ✓ | ✓ | ✓ | |
| | | 28 | | | ER-/PR-, White | | 1.83 (0.37 - 9.06) Ptrend:0.34 | ✓ | | ✓ | | ✓ | | | | ✓ | ✓ | ✓ | |
| | | 88 | | | ER-/PR-, Black | | 0.73 (0.34 - 1.56) Ptrend:0.27 | ✓ | | ✓ | | ✓ | | | | ✓ | ✓ | ✓ | |
| | | NA | | | Postmenopausal, White | | 2.68 (1.30 - 5.53) | ✓ | | ✓ | | ✓ | | | | ✓ | ✓ | ✓ | |
| | | NA | | | Postmenopausal Black | | 1.24 (0.80 - 1.93) | ✓ | | ✓ | | ✓ | | | | ✓ | ✓ | ✓ | |
| Pronk 2011 China (21934685)[42] | Shanghai Women's Health Study<br>Prospective Cohort<br>Range:40-70 years<br>F | 717 / 73,049<br>9 years | Cancer registry | Self-reported questionnaire, **Sitting time, occupational** | Incidence, Breast cancer | | 0.81 (0.65 - 1.01) Ptrend:0.04 | ✓ | | | | | | | | | ✓ | ✓ | |

| Author, Year Country PMID | Study name, type Age, sex, ethnicity, characteristics of participants | Cases/ Study size Median follow-up (years) | Case ascertainment | Exposure Assessment, detail | Outcome Sub-population | Comparison | RR (95%CI) Ptrend | Adjustments Age | Sex | Smo | Alc | Obe | Phy | Diet | Med | Mens | Repr | Oth | Skin |
|---|---|---|---|---|---|---|---|---|---|---|---|---|---|---|---|---|---|---|---|
| George 2010 USA (20864719)[39] | NIH- AARP Diet and Health Study Prospective Cohort Range:50-71 years M/F Postmenopausal | 2,866 / 97,039 7 years | Cancer registry | Self-administrated questionnaire, **Sitting**: sitting H/day | Incidence, Invasive breast cancer, postmenopausal | ≥9 vs <3 hours/day | 1.12 (0.95 - 1.31) | ✓ | | | ✓ | | ✓ | ✓ | ✓ | | ✓ | ✓ | |
| | | 2,866 | | | | | 1.08 (0.92 - 1.27) | ✓ | | | ✓ | ✓ | ✓ | ✓ | ✓ | | ✓ | ✓ | |
| Hildebrand 2013 USA (24097200)[51] | CPS II Nutrition Cohort Prospective Cohort Age:62.7 years F Mostly white Postmenopausal | 4,681 / 73,615 14.2 years | Self-report verified by medical record | Questionnaire (general), **Sitting**: Leisure-time sitting, time spent watching TV, reading, | Incidence / Mortality, Breast cancer, Postmenopausal | ≥6 vs <3 hours/day | 1.05 (0.96 - 1.16) | ✓ | | ✓ | ✓ | ✓ | ✓ | | ✓ | ✓ | ✓ | ✓ | |
| Ihira 2020 Japan (31925977)[77] | Japan Public Health Center-based Prospective Study Prospective Cohort M/F Japanese | 174 / 33,307 10.2 years | | Self-reported questionnaire, **Sedentary**: Daily occupational sitting time | Incidence, Breast cancer | ≥7 vs 1-<3 hour/day | 1.11 (0.69 - 1.81) Ptrend:0.618 | ✓ | | ✓ | ✓ | ✓ | ✓ | ✓ | | ✓ | | | |
| Nomura 2016 USA (27632430)[60] | Black Women's Health Study, 1995 Prospective Cohort Range:21-69 years F African American | 1,984 / 46,734 15.4 years | self-report, linkage to cancer registries, medical and pathology records | Self-completed questionnaire, **Sitting work**: Sitting at work, at baseline 1995 | Incidence, Breast cancer | ≥5 vs <1 hours/day | 1.15 (1.01 - 1.30) Ptrend:0.03 | ✓ | | ✓ | | ✓ | ✓ | ✓ | ✓ | ✓ | ✓ | ✓ | |
| | | 752 | | | Premenopausal | | 1.14 (0.91 - 1.42) Ptrend:0.26 | ✓ | | ✓ | | ✓ | ✓ | ✓ | ✓ | ✓ | ✓ | ✓ | |
| | | 965 | | | Postmenopausal | | 1.14 (0.96 - 1.35) Ptrend:0.18 | ✓ | | ✓ | | ✓ | ✓ | ✓ | ✓ | ✓ | ✓ | ✓ | |
| | | 1,008 | | | ER+/PR+ | | 1.11 (0.93 - 1.33) Ptrend:0.64 | ✓ | | ✓ | | ✓ | ✓ | ✓ | ✓ | ✓ | ✓ | ✓ | |
| ~~Nomura 2016 USA (27632430)[60]~~ | | 433 / 46,734 15.4 years | | | ER- & PR- | | 1.02 (0.79 - 1.32) Ptrend:0.64 | ✓ | | ✓ | | ✓ | ✓ | ✓ | ✓ | ✓ | ✓ | ✓ | |
| | | 173 | | | ER- & PR- & HER-2- | | 0.96 (0.63 - 1.44) Ptrend:0.72 | ✓ | | ✓ | | ✓ | ✓ | ✓ | ✓ | ✓ | ✓ | ✓ | |
| Cohen 2013 USA (23576427)[46] | Southern Community Cohort Study Nested Case Control Age:55 years White, black, other | 459 / 2,730 9 years | Cancer registry | Self-reported questionnaire, **Sitting time**: Sitting at work | Incidence, Invasive breast cancer | >3 vs 0 hours/day | 1.13 (0.82 - 1.56) Ptrend:0.63 | ✓ | | ✓ | | ✓ | ✓ | | | ✓ | ✓ | ✓ | |
| | | 132 | | | White | | 1.64 (0.90 - 2.99) Ptrend:0.10 | ✓ | | ✓ | | ✓ | ✓ | | | ✓ | ✓ | ✓ | |
| | | 314 | | | Black | | 0.96 (0.65 - 1.44) Ptrend:0.54 | ✓ | | ✓ | | ✓ | ✓ | | | ✓ | ✓ | ✓ | |
| Dirx, 2001 Netherlands (11745243)[33] | The Netherlands Cohort study Case Cohort Range:55-69 years F Not specified Postmenopausal | 755 / 62,573 7.3 years | | Self-reported questionnaire, **Sitting-time index (during working hours)**: life-long | Incidence, Breast cancer, postmenopausal | <2 vs 6-8 hours/day | 1.21 (0.94 - 1.56) Ptrend:0.54 | ✓ | | | ✓ | ✓ | | ✓ | ✓ | ✓ | ✓ | ✓ | |
| Cohen 2013 USA (23576427)[46] | Southern Community Cohort Study Nested Case Control Age:55 years White, black, other | 457 / 2,730 9 years | Cancer registry | Self-reported questionnaire, **Sitting time**: Sitting in a car or bus | Incidence, Invasive breast cancer | >2 vs 0 - 0.32 hours/day | 1.05 (0.72 - 1.53) Ptrend:0.20 | ✓ | | ✓ | | ✓ | ✓ | | | ✓ | ✓ | ✓ | |

| Author, Year Country PMID | Study name, type Age, sex, ethnicity, characteristics of participants | Cases/ Study size Median follow-up (years) | Case ascertainment | Exposure Assessment, detail | Outcome Sub-population | Comparison | RR (95%CI) Ptrend | Adjustments | | | | | | | | | | | |
|---|---|---|---|---|---|---|---|---|---|---|---|---|---|---|---|---|---|---|---|
| | | | | | | | | Age | Sex | Smo | Alc | Obe | Phy | Diet | Med | Mens | Repr | Oth | Skin |
| | | 131 | | | Incidence, Invasive breast cancer, White | | 1.26 (0.56 - 2.85) Ptrend:0.20 | ✓ | | ✓ | | ✓ | ✓ | | | ✓ | ✓ | ✓ | |
| | | 313 | | | Incidence, Invasive breast cancer, Black | | 1.06 (0.68 - 1.65) Ptrend:0.25 | ✓ | | ✓ | | ✓ | ✓ | | | ✓ | ✓ | ✓ | |
| Ihira 2020 Japan (31925977)[77] | Japan Public Health Center-based Prospective Study Prospective Cohort M/F Japanese | 255 / 33,307 10.2 years | | Self-reported questionnaire, **Sedentary**: Daily occupational sitting time including home duties | Incidence, Breast cancer, Women | ≥7 vs 1-<3 hour/day | 1.21 (0.80 - 1.81) Ptrend:0.425 | ✓ | | ✓ | ✓ | ✓ | ✓ | ✓ | | ✓ | | | |
| Cohen 2013 USA (23576427)[46] | Southern Community Cohort Study Nested Case Control Age:55 years White, black, other | 457 / 2,730 9 years | Cancer registry | Self-reported questionnaire, **Sitting time**: Other sitting includes using computer at home, sitting at meals, talking on the phone, reading, playing games, or sewing | Incidence, Invasive breast cancer | >4 vs 0 - 0.9 hours/day | 1.28 (0.91 - 1.80) Ptrend:0.52 | ✓ | | ✓ | | ✓ | ✓ | | | ✓ | ✓ | ✓ | |
| | | 132 | | | White | >4 vs 0 - 0.9 hours/day | 1.56 (0.77 - 3.14) Ptrend:0.11 | ✓ | | ✓ | | ✓ | ✓ | | | ✓ | ✓ | ✓ | |
| | | 312 | | | Black | | 1.13 (0.75 - 1.71) Ptrend:0.63 | ✓ | | ✓ | | ✓ | ✓ | | | ✓ | ✓ | ✓ | |
| **Endometrial cancer** | | | | | | | | | | | | | | | | | | | |
| Moore 2010 USA (20877336)[40] | NIH- AARP Diet and Health Study Prospective Cohort F Retired | 888 / 69,648 | linkage of the cohort with database to state cancer registries | Self-reported questionnaire, **time spent sitting per day** | Incidence, Endometrial Cancer | <9 vs>3 h/d | 1.45 (1.10 - 1.92) Ptrend:<0.01 | ✓ | | ✓ | | | ✓ | | | ✓ | ✓ | ✓ | |
| | | 888 | | | | | 1.15 (0.87 - 1.53) Ptrend:<0.01 | ✓ | | ✓ | | ✓ | ✓ | | | ✓ | ✓ | ✓ | |
| Gierach 2009 USA (19123463)[38] | NIH- AARP Diet and Health Study Prospective Cohort Range:50-71 years F Retired | 649 / 109,621 776170 person-years | linkage to eight state cancer registries | Self-reported questionnaire, **Sitting time**: hours spent sitting during typical 24 hours period in past 12 months11 | Incidence | 7+ vs <3 hours | 1.56 (1.22 - 1.99) Ptrend:<0.0001 | ✓ | | ✓ | | | | | | ✓ | ✓ | ✓ | |
| | | 649 | | | Incidence | | 1.23 (0.96 - 1.57) Ptrend:0.04 | ✓ | | ✓ | | ✓ | ✓ | | | ✓ | ✓ | ✓ | |
| Patel 2015 USA (26126627)[56] | CPS II Nutrition Cohort Prospective Cohort Age:63 years F Mostly white | 387 / 77,462 15.8 years | Cancer registry and death certificates and medical records | Self-reported questionnaire, **Sitting**: Leisure-time spent sitting | Incidence / Mortality, Endometrial Cancer | >6 vs >3 hours/day | 1.34 (1.08 - 1.67) | ✓ | | ✓ | ✓ | | ✓ | ✓ | | ✓ | ✓ | ✓ | |
| | | 387 | | | | | 1.21 (0.97 - 1.50) | ✓ | | ✓ | ✓ | ✓ | ✓ | ✓ | | ✓ | ✓ | ✓ | |
| Ihira 2020 Japan (31925977)[77] | Japan Public Health Center-based Prospective Study Prospective Cohort M/F Japanese | 50 / 33,307 10.2 years | | Self-reported questionnaire, **Sedentary**: Daily occupational sitting time | Incidence, Endometrial Cancer | ≥7 vs 1-<3 hour/day | 0.49 (0.15 - 1.52) Ptrend:0.314 | ✓ | | ✓ | ✓ | ✓ | ✓ | ✓ | | ✓ | | | |
| | | 62 | | Daily occupational sitting time including home duties | | | 0.96 (0.40 - 2.30) Ptrend:0.813 | ✓ | | ✓ | ✓ | ✓ | ✓ | ✓ | | ✓ | | | |

| Author, Year Country PMID | Study name, type Age, sex, ethnicity, characteristics of participants | Cases/ Study size Median follow-up (years) | Case ascertainment | Exposure Assessment, detail | Outcome Sub-population | Comparison | RR (95%CI) Ptrend | Adjustments: Age | Sex | Smo | Alc | Obe | Phy | Diet | Med | Mens | Repr | Oth | Skin |
|---|---|---|---|---|---|---|---|---|---|---|---|---|---|---|---|---|---|---|---|
| **Ovarian cancer** | | | | | | | | | | | | | | | | | | | |
| Buras 2022 USA<br>(35094053)[84] | NHS and NHS II<br>Prospective Cohort<br>Age:47 years<br>F<br>nurses | 884 / 171,085<br>17.2 | self-report, cancer registry, death report | Self-reported questionnaire, **Sitting time**:<br>total sedentary behaviour | Incidence, Ovarian cancer | >40 vs <20 hours/week | 1.09 (0.92 - 1.30)<br>Ptrend:0.47 | ✓ | | | | ✓ | ✓ | | | ✓ | ✓ | ✓ | |
| Xiao 2013 USA<br>(23966580)[49] | NIH- AARP Diet and Health Study<br>Prospective Cohort<br>Range:50-71 years<br>F<br>Retired | 463 / 153,123<br>12 years | cancer registry, death master file, national death index plus, postal service database | Self-reported questionnaire, **Sitting time**:<br>Overall sitting | Incidence, Epithelial ovarian cancer | 7+ vs <3 hours/day | 1.06 (0.81 - 1.39) | ✓ | | ✓ | | | | | | ✓ | ✓ | ✓ | |
| Hildebrand 2015 USA<br>(26335264)[57] | CPS-II Nutrition Cohort<br>Prospective Cohort<br>Age:62.2 years<br>F<br>Not reported<br>Postmenopausal | 638 / 63,972<br>19 years | medical record | Self-reported questionnaire, **Sedentary**:<br>Sitting (hours/day) | Cancer occurrence, Total ovarian cancer | >/6 vs <3 hours/day | 1.44 (1.12 - 1.85)<br>Ptrend:0.006 | ✓ | | ✓ | | ✓ | | | | | ✓ | ✓ | |
| | | 313 | | | Serous Ovarian cancer | | 1.52 (1.06 - 2.16)<br>Ptrend:0.01 | ✓ | | ✓ | | ✓ | | | | | ✓ | ✓ | |
| | | 112 | | | Nonserous ovarian cancer | | 1.08 (0.57 - 2.04)<br>Ptrend:0.83 | ✓ | | ✓ | | ✓ | | | | | ✓ | ✓ | |
| Buras 2022 USA<br>(35094053)[84] | NHS and NHS II<br>Prospective Cohort<br>Age:47 years<br>F<br>nurses | 884 / 171,085<br>17.2 | self-report, cancer registry, death report | Self-reported questionnaire, **Sitting work**: | Incidence, Ovarian cancer | >20 vs <5 hours/week | 1.40 (1.14 - 1.71)<br>Ptrend:<0.01 | ✓ | | | | ✓ | ✓ | | | ✓ | ✓ | ✓ | |
| Ihira 2020 Japan<br>(31925977)[77] | Japan Public Health Center-based Prospective Study<br>Prospective Cohort<br>F<br>Japanese | 24 / 33,307<br>10.2 years | | Self-reported questionnaire, **Sedentary**:<br>Daily occupational sitting time | Incidence, Ovarian cancer | ≥7 vs 1-<3 hour/day | 1.51 (0.48 - 4.72)<br>Ptrend:0.618 | ✓ | | ✓ | ✓ | ✓ | ✓ | ✓ | | ✓ | | | |
| Buras 2022 USA<br>(35094053)[84] | NHS and NHS II<br>Prospective Cohort<br>Age:47 years<br>F<br>nurses | 884 / 171,085<br>17.2 | self-report, cancer registry, death report | Self-reported questionnaire, **Sitting time**:<br>reading, eating meals, using a computer/cell phone, or sitting at a desk | Incidence, Ovarian cancer | >20 vs <5 hours/week | 0.84 (0.66 - 1.08)<br>Ptrend:0.04 | ✓ | | | | ✓ | ✓ | | | ✓ | ✓ | ✓ | |
| Ihira 2020 Japan<br>(31925977)[77] | Japan Public Health Center-based Prospective Study<br>Prospective Cohort<br>F<br>Japanese | 39 / 33,307<br>10.2 years | | Self-reported questionnaire, **Sedentary**:<br>Daily occupational sitting time including home duties | Incidence, Ovarian cancer | ≥7 vs 1-<3 hour/day | 1.37 (0.49 - 3.85)<br>Ptrend:0.800 | ✓ | | ✓ | ✓ | ✓ | ✓ | ✓ | | ✓ | | | |
| **Prostate cancer** | | | | | | | | | | | | | | | | | | | |
| Rangul 2018 Norway<br>(30352079)[70] | The Nord-Trøndelag Health Study (HUNT)<br>Prospective Cohort<br>M | 889 / 18,771<br>16 years | Cancer Registry of Norway | Self-reported questionnaire, **Sitting time**: | Incidence, prostate cancer | High vs Low | 1.22 (1.05 - 1.42)<br>Ptrend:0.01 | ✓ | | ✓ | ✓ | ✓ | | | | | | ✓ | |
| Lynch 2014 USA<br>(24526287)[53] | NIH- AARP Diet and Health Study<br>Prospective Cohort<br>Range:50-71 years | 13,751 / 170,481<br>8.5 years | Cancer registry and national death index | Self-administered Questionnaire, | Incidence, prostate cancer | ≥9 vs <3 hours/day | 0.98 (0.91 - 1.05)<br>Ptrend:0.09 | ✓ | | ✓ | ✓ | ✓ | ✓ | ✓ | ✓ | | | ✓ | |

| Author, Year Country PMID | Study name, type Age, sex, ethnicity, characteristics of participants | Cases/ Study size Median follow-up (years) | Case ascertainment | Exposure Assessment, detail | Outcome Sub-population | Comparison | RR (95%CI) Ptrend | Adjustments: Age | Sex | Smo | Alc | Obe | Phy | Diet | Med | Mens | Repr | Oth | Skin |
|---|---|---|---|---|---|---|---|---|---|---|---|---|---|---|---|---|---|---|---|
| | M<br>White, black, other | | | **Sitting time**: | | | | | | | | | | | | | | | |
| | | 4,435 | | | Incidence, prostate cancer, BMI 18.5-24.9kg/m2 | | 0.97 (0.85 - 1.10)<br>Ptrend:0.13 | ✓ | | ✓ | ✓ | ✓ | ✓ | ✓ | ✓ | | | ✓ | |
| | | 6,895 | | | BMI 25.0 to 29.9 kg/m^2 | | 0.99 (0.89 - 1.10)<br>Ptrend:0.30 | ✓ | | ✓ | ✓ | ✓ | ✓ | ✓ | ✓ | | | ✓ | |
| | | 2,366 | | | BMI >=30 kg/m^2 | | 0.99 (0.89 - 1.10)<br>Ptrend:0.79 | ✓ | | ✓ | ✓ | ✓ | ✓ | ✓ | ✓ | | | ✓ | |
| | | 1,365 | | | advanced prostate cancer | ≥7 vs <3 hours/day | 0.91 (0.77 - 1.08)<br>Ptrend:0.16 | ✓ | | ✓ | ✓ | ✓ | ✓ | ✓ | ✓ | | | ✓ | |
| | | 420 | | | BMI 18.5-24.9kg/m2 | | 0.86 (0.64 - 1.14)<br>Ptrend:0.08 | ✓ | | ✓ | ✓ | ✓ | ✓ | ✓ | ✓ | | | ✓ | |
| | | 673 | | | BMI 25.0 to 29.9 kg/m^2 | | 0.87 (0.69 - 1.10)<br>Ptrend:0.14 | ✓ | | ✓ | ✓ | ✓ | ✓ | ✓ | ✓ | | | ✓ | |
| | | 262 | | | BMI >=30 kg/m^2 | | 1.24 (0.82 - 1.85)<br>Ptrend:0.18 | ✓ | | ✓ | ✓ | ✓ | ✓ | ✓ | ✓ | | | ✓ | |
| | | 669 | | | Mortality, Prostate cancer | | 1.07 (0.84 - 1.35)<br>Ptrend:0.98 | ✓ | | ✓ | ✓ | ✓ | ✓ | ✓ | ✓ | | | ✓ | |
| | | 181 | | | BMI 18.5-24.9kg/m2 | | 0.90 (0.57 - 1.42)<br>Ptrend:0.23 | ✓ | | ✓ | ✓ | ✓ | ✓ | ✓ | ✓ | | | ✓ | |
| | | 335 | | | BMI 25.0 to 29.9 kg/m^2 | | 1.07 (0.77 - 1.49)<br>Ptrend:0.91 | ✓ | | ✓ | ✓ | ✓ | ✓ | ✓ | ✓ | | | ✓ | |
| | | 151 | | | BMI >=30 kg/m^2 | | 1.34 (0.79 - 2.26)<br>Ptrend:0.18 | ✓ | | ✓ | ✓ | ✓ | ✓ | ✓ | ✓ | | | ✓ | |
| Patel 2015 USA<br>(26126627)[56] | CPS II Nutrition Cohort<br>Prospective Cohort<br>Age:63 years<br>M<br>Mostly white | 8,276 /<br>69,260<br>13.2 years | Cancer registry and death certificates and medical records | Self-reported questionnaire,<br>**Sitting**:<br>Leisure-time spent sitting | Incidence / Mortality, prostate cancer | >6 vs <3 hours/day | 0.96 (0.90 - 1.02) | ✓ | | ✓ | ✓ | | ✓ | ✓ | ✓ | | | ✓ | |
| | | 1,705 | | | Advance prostate cancer | | 0.96 (0.85 - 1.09) | ✓ | | ✓ | ✓ | | ✓ | ✓ | ✓ | | | ✓ | |
| | | 8,276 | | | Prostate cancer | | 0.97 (0.91 - 1.03) | ✓ | | ✓ | ✓ | ✓ | ✓ | ✓ | ✓ | | | ✓ | |
| | | 1,705 | | | Advance prostate cancer | | 0.96 (0.85 - 1.09) | ✓ | | ✓ | ✓ | ✓ | ✓ | ✓ | ✓ | | | ✓ | |
| Ihira 2020 Japan<br>(31925977)[77] | Japan Public Health Center-based Prospective Study<br>Prospective Cohort<br>M<br>Japanese | 521 / 20,300<br>10.2 years | | Self-reported questionnaire,<br>**Sedentary**:<br>Daily occupational sitting time | Incidence, prostate cancer | ≥7 vs 1-<3 hour/day | 1.10 (0.84 - 1.45)<br>Ptrend:0.543 | ✓ | | ✓ | ✓ | ✓ | ✓ | ✓ | | | | | |
| Zeegers, 2005 Netherlands<br>(15941961)[34] | The Netherlands Cohort study<br>Nested Case Control<br>Range:55-69 years<br>M<br>not stated | 1,127 /<br>58,279<br>9.3 years | municipal population registries | Self-reported questionnaire,<br>**Sitting time**:<br>Longest held job | Incidence, prostate cancer | | 1.16 (0.91 - 1.47)<br>Ptrend:0.22 | ✓ | | | ✓ | ✓ | | ✓ | | | | ✓ | |
| **Kidney cancer** | | | | | | | | | | | | | | | | | | | |
| George 2011 USA<br>(21737302)[41] | National Institute of Health (NIH)-AARP(formerly the American Association for Retired Persons) Diet and Health Study | 1,206 /<br>289,512<br>10 years | Cancer registry | Self-administrated questionnaire,<br>**Sitting**:<br>Total sitting | Incidence, renal cell carcinoma, Men and women | ≥9 vs <3 hours/day | 1.11 (0.87 - 1.41)<br>Ptrend:0.765 | ✓ | ✓ | ✓ | ✓ | | ✓ | ✓ | | | | ✓ | |

| Author, Year Country PMID | Study name, type Age, sex, ethnicity, characteristics of participants | Cases/ Study size Median follow-up (years) | Case ascertainment | Exposure Assessment, detail | Outcome Sub-population | Comparison | RR (95%CI) Ptrend | Age | Sex | Smo | Alc | Obe | Phy | Diet | Med | Mens | Repr | Oth | Skin |
|---|---|---|---|---|---|---|---|---|---|---|---|---|---|---|---|---|---|---|---|
| | Prospective Cohort<br>Age:63 years<br>M/F | | | | | | | | | | | | | | | | | | |
| | | 909 | | | Men | | 1.08 (0.82 - 1.42)<br>Ptrend:0.524 | ✓ | | ✓ | ✓ | | ✓ | ✓ | | | | ✓ | |
| | | 297 | | | Women | | 1.20 (0.72 - 1.99)<br>Ptrend:0.572 | ✓ | | ✓ | ✓ | | ✓ | ✓ | | | ✓ | ✓ | |
| Patel 2015 USA (26126627)[56] | CPS II Nutrition Cohort<br>Prospective Cohort<br>Age:63 years<br>M/F<br>Mostly white | 379 / 146,722<br>17 years | Cancer registry and death certificates and medical records | Self-reported questionnaire, **Sitting**:<br>Leisure-time spent sitting | Incidence / Mortality, kidney cancer, Men | >6 vs <3 hours/day | 1.11 (0.82 - 1.52) | ✓ | | ✓ | ✓ | | ✓ | ✓ | | | | ✓ | |
| | | 186 | | | Women | | 1.01 (0.65 - 1.57) | ✓ | | ✓ | ✓ | | ✓ | ✓ | | ✓ | ✓ | ✓ | |
| | | 379 | | | Men | | 1.09 (0.80 - 1.48) | ✓ | | ✓ | ✓ | ✓ | ✓ | ✓ | | | | ✓ | |
| | | 186 | | | Women | | 0.97 (0.62 - 1.51) | ✓ | | ✓ | ✓ | ✓ | ✓ | ✓ | | ✓ | ✓ | ✓ | |
| Ihira 2020 Japan (31925977)[77] | Japan Public Health Center-based Prospective Study<br>Prospective Cohort<br>M/F<br>Japanese | 57 / 33,307<br>10.2 years | | Self-reported questionnaire, **Sedentary**:<br>Daily occupational sitting time | Incidence, kidney cancer, Men | ≥7 vs 1-<3 hour/day | 1.04 (0.44 - 2.47)<br>Ptrend:0.731 | ✓ | | ✓ | ✓ | ✓ | ✓ | ✓ | | | | | |
| | | 12 | | | Women | 5-<7 vs 1-<3 hour/day | 4.15 (0.66 - 26.11) | ✓ | | ✓ | ✓ | ✓ | ✓ | ✓ | | | | | |
| | | 19 | | Daily occupational sitting time including home duties | Women | | 2.89 (0.7-11.92) | ✓ | | ✓ | ✓ | ✓ | ✓ | ✓ | | | | | |
| **Bladder cancer** | | | | | | | | | | | | | | | | | | | |
| Chantaprasopsuk 2023 USA (37202564)[89] | CPS II Nutrition Cohort<br>Prospective Cohort<br>Age:63 years<br>M/F | 2,215 / 146,027 | self-report, linkage to cancer registries, medical and pathology records | Self-reported questionnaire, **Sitting time**: | Incidence, bladder cancer | >6 vs 0-3 hours/day | 1.09 (0.96 - 1.24) | ✓ | ✓ | ✓ | | | ✓ | ✓ | | | | ✓ | |
| | | 2,215 | | | | | 1.09 (0.95 - 1.24) | ✓ | ✓ | ✓ | | ✓ | ✓ | ✓ | | | | ✓ | |
| | | 1,103 | | | Non-invasive bladder cancer | | 1.01 (0.82 - 1.20) | ✓ | ✓ | ✓ | | | ✓ | ✓ | | | | ✓ | |
| | | 1,103 | | | | | 0.99 (0.82 - 1.20) | ✓ | ✓ | ✓ | | ✓ | ✓ | ✓ | | | | ✓ | |
| | | 1,034 | | | Invasive bladder cancer | | 1.23 (1.02 - 1.47) | ✓ | ✓ | ✓ | | | ✓ | ✓ | | | | ✓ | |
| | | 1,034 | | | | | 1.22 (1.02 - 1.47) | ✓ | ✓ | ✓ | | ✓ | ✓ | ✓ | | | | ✓ | |
| | | 1,764 | | | Bladder cancer, Men | | 1.07 (0.92 - 1.23) | ✓ | | ✓ | | ✓ | ✓ | ✓ | | | | ✓ | |

| Author, Year Country PMID | Study name, type Age, sex, ethnicity, characteristics of participants | Cases/ Study size Median follow-up (years) | Case ascertainment | Exposure Assessment, detail | Outcome Sub-population | Comparison | RR (95%CI) Ptrend | Adjustments | | | | | | | | | | | |
|---|---|---|---|---|---|---|---|---|---|---|---|---|---|---|---|---|---|---|---|
| | | | | | | | | Age | Sex | Smo | Alc | Obe | Phy | Diet | Med | Mens | Repr | Oth | Skin |
| | | 451 | | | Women | | 1.20 (0.88 - 1.63) | ✓ | | ✓ | | ✓ | ✓ | ✓ | | | | ✓ | |
| Ihira 2020 Japan (31925977)[77] | Japan Public Health Center-based Prospective Study Prospective Cohort M/F Japanese | 93 / 33,307 10.2 years | | Self-reported questionnaire, **Sedentary**: Daily occupational sitting time | Incidence, bladder cancer, Men | ≥7 vs 1-<3 hour/day | 0.56 (0.27 - 1.17) Ptrend:0.188 | ✓ | | ✓ | ✓ | ✓ | ✓ | ✓ | | | | | |
| | | 15 | | | Women | 5-<7 vs 1-<3 hour/day | 1.42 (0.33 - 6.13) Ptrend:0.927 | ✓ | | ✓ | ✓ | ✓ | ✓ | ✓ | | | | | |
| | | 19 | | Daily occupational sitting time in women including home duties | Women | ≥7 vs 1-<3 hour/day | 0.73 (0.14 - 3.87) | ✓ | | ✓ | ✓ | ✓ | ✓ | ✓ | | | | | |
| **Haematological cancers** | | | | | | | | | | | | | | | | | | | |
| Rees-Punia 2019 USA (31196856)[75] | CPS II Nutrition Cohort Prospective Cohort Age:69.2 years M/F | 155 / 109,030 10 years | Cancer registry and national death index | Self-reported questionnaire, **Sitting time**: | Incidence, Acute myeloid leukaemia (AML) | >6 vs <3 hours/day | 1.01 (0.60 - 1.64) | ✓ | ✓ | ✓ | ✓ | ✓ | | | | | | ✓ | |
| | | 255 / 109,030 10 years | | | Incidence, Myeloid leukaemia | | 1.28 (0.89 - 1.85) | ✓ | ✓ | ✓ | ✓ | ✓ | | | | | | ✓ | |
| Teras 2012 USA (22275172)[44] | American Cancer Society Cancer Prevention Study II Nutrition Cohort Prospective Cohort Age:62.9 years | 143 / 146,850 | medical records and cancer registries | Questionnaire, **Sedentary**: sitting (hr/day) | Incidence, Follicular lymphoma, Men | 6+ vs <3 hours/day | 0.76 (0.45 - 1.31) Ptrend:0.19 | ✓ | | ✓ | ✓ | ✓ | ✓ | | | | | ✓ | |
| | | 137 / 146,850 | | | Women | | 1.43 (0.81 - 2.53) Ptrend:0.11 | ✓ | | ✓ | ✓ | ✓ | ✓ | | | | | ✓ | |
| | | 1,138 | | | Incidence, Non-Hodgkin Lymphoma, Men | | 0.95 (0.79 - 1.15) Ptrend:0.67 | ✓ | | ✓ | ✓ | ✓ | ✓ | | | | | ✓ | |
| | | 863 | | | Women | | 1.26 (1.01 - 1.59) Ptrend:0.011 | ✓ | | ✓ | ✓ | ✓ | ✓ | | | | | ✓ | |
| | | 271 | | | Incidence, Leukaemia (CLL +SLL), Men | | 0.75 (0.50 - 1.13) Ptrend:0.30 | ✓ | | ✓ | ✓ | ✓ | ✓ | | | | | ✓ | |
| | | 197 | | | Incidence, Diffuse large-Bcell lymphoma, Women | 6+h/d vs <3hr/d hours/day | 0.78 (0.45 - 1.37) Ptrend:0.94 | ✓ | | ✓ | ✓ | ✓ | ✓ | | | | | ✓ | |
| Patel 2015 USA (26126627)[56] | CPS II Nutrition Cohort Prospective Cohort Age:63 years M Mostly white | 243 / 69,260 13.2 years | Cancer registry and death certificates and medical records | Self-reported questionnaire, **Sitting**: Leisure-time spent sitting | Incidence / Mortality, multiple Myeloma, Men | >6 vs <3hours/day | 1.00 (0.68 - 1.45) | ✓ | | ✓ | ✓ | ✓ | ✓ | ✓ | | | | ✓ | |
| | | 171 | | | Women | | 1.65 (1.07 - 2.54) | ✓ | | ✓ | ✓ | ✓ | ✓ | ✓ | | ✓ | ✓ | ✓ | |
| | | 243 | | | Men | | 1.02 (0.70 - 1.48) | ✓ | | ✓ | ✓ | | ✓ | ✓ | | | | ✓ | |
| | | 171 | | | Women | | 1.64 (1.06 - 2.53) | ✓ | | ✓ | ✓ | | ✓ | ✓ | | ✓ | ✓ | ✓ | |

| Author, Year Country PMID | Study name, type Age, sex, ethnicity, characteristics of participants | Cases/ Study size Median follow-up (years) | Case ascertainment | Exposure Assessment, detail | Outcome Sub-population | Compariso n | RR (95%CI) Ptrend | Adjustments Age | Sex | Smo | Alc | Obe | Phy | Diet | Med | Mens | Repr | Oth | Skin |
|---|---|---|---|---|---|---|---|---|---|---|---|---|---|---|---|---|---|---|---|
| Teras 2012 USA (22275172)[44] | American Cancer Society Cancer Prevention Study II Nutrition Cohort Prospective Cohort Age:62.9 years | 238 / 146,850 | medical records and cancer registries | Questionnaire, **Sedentary**: sitting (hr/day) | Incidence, Diffuse large-Bcell lymphoma, Men | 6+ vs <3 hours/day | 1.11 (0.75 - 1.65) Ptrend:0.57 | ✓ | | ✓ | ✓ | ✓ | ✓ | | | | | ✓ | |
| | | 211 | | | Incidence, Leukaemia (CLL +SLL), Women | | 1.16 (0.74 - 1.82) Ptrend:0.75 | ✓ | | ✓ | ✓ | ✓ | ✓ | | | | | ✓ | |

**Adjustment groups: Age**: age; **Alc**: alcohol; **Diet**: dietary factors and energy intake; **Med**: medical factors (e.g. cancer-related infections, diseases in the breast, digestive tract, gallbladder, liver, or pancreas, cancer screening, precancerous conditions, or pre-existing skin cancer); **Mens**: menstrual characteristics (e.g. age at menarche, menopausal status/duration, age at menopause, or type of menopause); **Obe**: obesity (adiposity measures); **Oth**: others (e.g. family history/genetic factor, race/ethnicity, ancestry, socioeconomic factors, or marriage); **Phy**: physical activity; **Repr**: reproductive or hormonal factors (e.g. parity, age at first/last birth, postmenopausal hormone use, or oral contraceptive use); **Sex**: sex; **Skin**: skin-related factors (e.g. UV light/sun exposure, sunburns, time spent outdoors, sunscreen use, indoor tanning, moles, fair skin, red/blond hair, blue/green eyes, or freckles); **Smo**: smoking, cancer-related

### Supplementary Table 10: Descriptive table of the included observational studies of screen time and cancer risk.

| Author, Year Country (PMID) | Study name, type Age, sex, ethnicity, characteristics of participants | Cases/ Study size Median follow-up (years) | Case ascertainment | Exposure Assessment, detail | Outcome Sub-population | Comparison | RR (95%CI) Ptrend | Adjustments: Age | Sex | Smo | Alc | Obe | Phy | Diet | Med | Mens | Repr | Oth | Skin |
|---|---|---|---|---|---|---|---|---|---|---|---|---|---|---|---|---|---|---|---|
| **Oesophageal cancer** | | | | | | | | | | | | | | | | | | | |
| Kunzmann 2018 UK (30288276)[68] | UK Biobank Prospective Cohort Age:56.5 years M/F | 294 / 359,033 8 years | cancer registries | Self-report questionnaire , **Sedentary**: screen time | Incidence, Oesophageal cancer | 4 - 16 vs 0 - 3 hours/day | 1.14 (0.87 - 1.49) Ptrend:0.23 | ✓ | ✓ | ✓ | ✓ | | | | | | | ✓ | |
| | | 218 | | | Adenocarcinoma | | 1.31 (0.95 - 1.79) Ptrend:0.08 | ✓ | ✓ | ✓ | ✓ | | | | | | | ✓ | |
| | | 57 | | | Squamous cell carcinoma | | 0.68 (0.37 - 1.26) Ptrend:0.41 | ✓ | ✓ | ✓ | ✓ | | | | | | | ✓ | |
| Hunter 2020 UK (32746843)[78] | UK Biobank Prospective Cohort Range:40-69 years M/F | 541 / 28,992 7.6 years | cancer registries | Self- reported questionnaire, **Television watching** | Incidence, Oesophageal cancer | >5 vs 1-3 hours/day | 1.02 (0.73 - 1.42) | ✓ | ✓ | ✓ | ✓ | ✓ | | | | | | ✓ | |
| | | 541 | | | | Per 1 hour day | 1.02 (0.97 - 1.08) Ptrend:0.34 | ✓ | ✓ | ✓ | ✓ | ✓ | | | | ✓ | ✓ | ✓ | |
| | | 161 | | | Obese | | 1.04 (0.96 - 1.13) | ✓ | ✓ | ✓ | ✓ | ✓ | | | | ✓ | ✓ | ✓ | |
| | | 341 | | | Non obese | | 1.01 (0.95 - 1.08) | ✓ | ✓ | ✓ | ✓ | ✓ | | | | ✓ | ✓ | ✓ | |
| | | 168 | | | High physical activity | | 1.02 (0.93 - 1.13) | ✓ | ✓ | ✓ | ✓ | ✓ | | | | ✓ | ✓ | ✓ | |
| | | 142 | | | Moderate physical activity | | 1.01 (0.92 - 1.12) | ✓ | ✓ | ✓ | ✓ | ✓ | | | | ✓ | ✓ | ✓ | |
| | | 76 | | | Low physical activity | | 1.03 (0.92 - 1.15) | ✓ | ✓ | ✓ | ✓ | ✓ | | | | ✓ | ✓ | ✓ | |
| | | 154 | | | Never smokers | | 1.04 (0.94 - 1.15) Ptrend:0.34 | ✓ | ✓ | | ✓ | ✓ | | | | ✓ | ✓ | ✓ | |
| | | 112 | | | Former heavy smokers | | 1.01 (0.91 - 1.12) | ✓ | ✓ | | ✓ | ✓ | | | | ✓ | ✓ | ✓ | |
| | | 139 | | | Former light smokers | | 1.02 (0.92 - 1.14) | ✓ | ✓ | | ✓ | ✓ | | | | ✓ | ✓ | ✓ | |
| | | 59 | | | Current heavy smokers | | 1.07 (0.95 - 1.19) | ✓ | ✓ | | ✓ | ✓ | | | | ✓ | ✓ | ✓ | |
| | | 38 | | | Current light smokers | | 0.93 (0.77 - 1.13) | ✓ | ✓ | | ✓ | ✓ | | | | ✓ | ✓ | ✓ | |
| Cook 2013 USA (24367697)[52] | NIH- AARP Diet and Health Study Prospective Cohort Range:50-71 years M/F Retired | 377 / 303,033 | Postal Service, Social Security Administration Death Master File, National Death Index | Self-reported questionnaire, **Television watching**: Typical number of hours per day watching TV during the last 12 months | Incidence, oesophageal adenocarcinoma | >7 vs <1 hours/day | 0.55 (0.29 - 1.02) | ✓ | ✓ | ✓ | ✓ | | | ✓ | | | | ✓ | |
| | | 128 | | | Oesophageal squamocellular carcinoma | | 0.78 (0.26 - 2.32) | ✓ | ✓ | ✓ | ✓ | | | ✓ | | | | ✓ | |
| **Stomach cancer** | | | | | | | | | | | | | | | | | | | |
| Kunzmann 2018 UK (30288276)[68] | UK Biobank Prospective Cohort Age:56.5 years M/F | 216 / 359,033 8 years | cancer registries | Self-reported questionnaire, **Sedentary**: screen time | Incidence, Gastric cancer | 4 - 16 vs 0 - 3 hours/day | 1.13 (0.83 - 1.55) Ptrend:0.42 | ✓ | ✓ | ✓ | ✓ | | | | | | | ✓ | |

| Author, Year Country (PMID) | Study name, type Age, sex, ethnicity, characteristics of participants | Cases/ Study size Median follow-up (years) | Case ascertainment | Exposure Assessment, detail | Outcome Sub-population | Comparison | RR (95%CI) Ptrend | Adjustments: Age | Sex | Smo | Alc | Obe | Phy | Diet | Med | Mens | Repr | Oth | Skin |
|---|---|---|---|---|---|---|---|---|---|---|---|---|---|---|---|---|---|---|---|
| Hunter 2020 UK (32746843)[78] | UK Biobank Prospective Cohort Range:40-69 years M/F | 183 | cancer registries | Self- reported questionnaire, **Television watching** | Incidence, Gastric Adenocarcinoma | >5 vs 1-3 hours/day | 1.11 (0.79 - 1.57) Ptrend:0.51 | ✓ | ✓ | ✓ | ✓ | | | | | | | ✓ | |
| | | 89 | | | Cardia adenocarcinoma | | 0.95 (0.58 - 1.57) Ptrend:0.87 | ✓ | ✓ | ✓ | ✓ | | | | | | | ✓ | |
| | | 128 | | | Noncardia adenocarcinoma | | 1.29 (0.86 - 1.93) Ptrend:0.21 | ✓ | ✓ | ✓ | ✓ | | | | | | | ✓ | |
| | | 356 / 28,992 7.6 years | | | Incidence, Stomach cancer | >5 vs 1-3 hours/day | 1.03 (0.69 - 1.53) | ✓ | ✓ | ✓ | ✓ | ✓ | | | | | | ✓ | |
| | | 356 | | | Incidence, Stomach cancer | Per 1 hour day | 1.06 (1.00 - 1.13) Ptrend:0.045 | ✓ | ✓ | ✓ | ✓ | ✓ | | | | | | ✓ | |
| | | 103 | | | Obese | | 0.95 (0.86 - 1.06) | ✓ | ✓ | ✓ | ✓ | ✓ | | | | | | ✓ | |
| | | 229 | | | Non obese | | 1.12 (1.05 - 1.20) | ✓ | ✓ | ✓ | ✓ | ✓ | | | | | | ✓ | |
| | | 104 | | | High physical activity | | 1.11 (0.99 - 1.28) | ✓ | ✓ | ✓ | ✓ | ✓ | | | | | | ✓ | |
| | | 108 | | | Moderate physical activity | | 0.99 (0.88 - 1.11) | ✓ | ✓ | ✓ | ✓ | ✓ | | | | | | ✓ | |
| | | 53 | | | Low physical activity | | 1.09 (0.98 - 1.23) | ✓ | ✓ | ✓ | ✓ | ✓ | | | | | | ✓ | |
| | | 123 | | | Never smokers | | 1.19 (1.09 - 1.30) Ptrend:0.045 | ✓ | ✓ | | ✓ | ✓ | | | | | | ✓ | |
| | | 72 | | | Former heavy smokers | | 1.01 (0.89 - 1.15) | ✓ | ✓ | | ✓ | ✓ | | | | | | ✓ | |
| | | 78 | | | Former light smokers | | 1.04 (0.91 - 1.19) | ✓ | ✓ | | ✓ | ✓ | | | | | | ✓ | |
| | | 36 | | | Current heavy smokers | | 0.97 (0.83 - 1.14) | ✓ | ✓ | | ✓ | ✓ | | | | | | ✓ | |
| | | 23 | | | Current light smokers | | 0.96 (0.76 - 1.22) | ✓ | ✓ | | ✓ | ✓ | | | | | | ✓ | |
| Cook 2013 USA (24367697)[52] | NIH- AARP Diet and Health Study Prospective Cohort Range:50-71 years M/F Retired | 255 / 303,033 | Postal Service, Social Security Administration Death Master File, National Death Index | Self-reported questionnaire, **Television watching**: Typical number of hours per day watching TV during the last 12 months | Cardia adenocarcinoma | >7 vs <1 hours/day | 1.37 (0.61 - 3.09) | ✓ | ✓ | ✓ | ✓ | | | ✓ | | | | ✓ | |
| | | 277 | | | Noncardia adenocarcinoma | | 0.93 (0.42 - 2.09) | ✓ | ✓ | ✓ | ✓ | | | ✓ | | | | ✓ | |
| **Colorectal cancer** | | | | | | | | | | | | | | | | | | | |
| Zhang 2024 UK (38974470)[93] | UK Biobank Prospective Cohort Age:55.94 years M/F Mostly white | 4,560 / 360,271 12.52 years | Cancer registry | Self-reported questionnaire, **Screen time, recreational** | Incidence, colorectal cancer | high vs low | 1.08 (1.00 - 1.17) | ✓ | ✓ | ✓ | ✓ | ✓ | ✓ | | ✓ | | | ✓ | |
| Morris 2018 UK (29520109)[67] | UK Biobank Prospective Cohort Range:40-69 years M/F | 2,377 / 430,584 5.6 years | national cancer registers | Self-administered Questionnaire, **Other sedentary activity**: time spent on computers hours/day | Colorectal cancer | >4 vs 0 h/day | 0.94 (0.68 - 1.29) | ✓ | ✓ | ✓ | ✓ | | | ✓ | | | ✓ | ✓ | |

| Author, Year Country (PMID) | Study name, type Age, sex, ethnicity, characteristics of participants | Cases/ Study size Median follow-up (years) | Case ascertainment | Exposure Assessment, detail | Outcome Sub-population | Comparison | RR (95%CI) Ptrend | Adjustments Age | Sex | Smo | Alc | Obe | Phy | Diet | Med | Mens | Repr | Oth | Skin |
|---|---|---|---|---|---|---|---|---|---|---|---|---|---|---|---|---|---|---|---|
| | | 1,383 | | | Men | | 1.04 (0.86 - 1.29) | ✓ | ✓ | ✓ | ✓ | | | ✓ | | | ✓ | ✓ | |
| | | 994 | | | Women | | 1.03 (0.73 - 1.44) | ✓ | ✓ | ✓ | ✓ | | | ✓ | | | ✓ | ✓ | |
| | | 1,546 | | | Colon cancer | | 0.86 (0.56 - 1.30) | ✓ | ✓ | ✓ | ✓ | | | ✓ | | | ✓ | ✓ | |
| | | 829 | | | Men | | 1.05 (0.80 - 1.37) | ✓ | ✓ | ✓ | ✓ | | | ✓ | | | ✓ | ✓ | |
| | | 717 | | | Women | | 1.01 (0.68 - 1.52) | ✓ | ✓ | ✓ | ✓ | | | ✓ | | | ✓ | ✓ | |
| | | 778 | | | Proximal colon cancer | | 0.84 (0.46 - 1.53) | ✓ | ✓ | ✓ | ✓ | | | ✓ | | | ✓ | ✓ | |
| | | 411 | | | Men | | 0.95 (0.64 - 1.42) | ✓ | ✓ | ✓ | ✓ | | | ✓ | | | ✓ | ✓ | |
| | | 367 | | | Women | | 0.84 (0.45 - 1.59) | ✓ | ✓ | ✓ | ✓ | | | ✓ | | | ✓ | ✓ | |
| | | 690 | | | Distal colon cancer | | 1.00 (0.56 - 1.79) | ✓ | ✓ | ✓ | ✓ | | | ✓ | | | ✓ | ✓ | |
| | | 380 | | | Men | | 1.18 (0.81 - 1.74) | ✓ | ✓ | ✓ | ✓ | | | ✓ | | | ✓ | ✓ | |
| | | 310 | | | Women | | 1.31 (0.76 - 2.26) | ✓ | ✓ | ✓ | ✓ | | | ✓ | | | ✓ | ✓ | |
| | | 817 | | | Rectal cancer | | 1.10 (0.68 - 1.79) | ✓ | ✓ | ✓ | ✓ | | | ✓ | | | ✓ | ✓ | |
| | | 547 | | | Men | | 1.03 (0.72 - 1.45) | ✓ | ✓ | ✓ | ✓ | | | ✓ | | | ✓ | ✓ | |
| | | 270 | | | Women | | 1.19 (0.66 - 2.13) | ✓ | ✓ | ✓ | ✓ | | | ✓ | | | ✓ | ✓ | |
| | | 2,377 | | | Colorectal cancer | | 0.91 (0.67 - 1.25) | ✓ | ✓ | ✓ | ✓ | ✓ | | ✓ | | | ✓ | ✓ | |
| | | 1,383 | | | Men | | 1.01 (0.82 - 1.25) | ✓ | ✓ | ✓ | ✓ | ✓ | | ✓ | | | ✓ | ✓ | |
| | | 994 | | | Women | | 1.01 (0.72 - 1.41) | ✓ | ✓ | ✓ | ✓ | ✓ | | ✓ | | | ✓ | ✓ | |
| | | 1,546 | | | Colon cancer | | 0.83 (0.55 - 1.26) | ✓ | ✓ | ✓ | ✓ | ✓ | | ✓ | | | ✓ | ✓ | |
| | | 829 | | | Men | | 1.00 (0.76 - 1.32) | ✓ | ✓ | ✓ | ✓ | ✓ | | ✓ | | | ✓ | ✓ | |
| | | 717 | | | Women | | 0.99 (0.66 - 1.49) | ✓ | ✓ | ✓ | ✓ | ✓ | | ✓ | | | ✓ | ✓ | |
| | | 778 | | | Proximal colon cancer | | 0.81 (0.44 - 1.47) | ✓ | ✓ | ✓ | ✓ | ✓ | | ✓ | | | ✓ | ✓ | |
| | | 411 | | | Men | | 0.91 (0.61 - 1.35) | ✓ | ✓ | ✓ | ✓ | ✓ | | ✓ | | | ✓ | ✓ | |
| | | 367 | | | Women | | 0.81 (0.43 - 1.54) | ✓ | ✓ | ✓ | ✓ | ✓ | | ✓ | | | ✓ | ✓ | |
| | | 690 | | | Distal colon cancer | | 0.98 (0.55 - 1.75) | ✓ | ✓ | ✓ | ✓ | ✓ | | ✓ | | | ✓ | ✓ | |
| | | 380 | | | Men | | 1.15 (0.78 - 1.69) | ✓ | ✓ | ✓ | ✓ | ✓ | | ✓ | | | ✓ | ✓ | |
| | | 310 | | | Women | | 1.30 (0.75 - 2.24) | ✓ | ✓ | ✓ | ✓ | ✓ | | ✓ | | | ✓ | ✓ | |
| | | 817 | | | Rectal cancer | | 1.08 (0.66 - 1.76) | ✓ | ✓ | ✓ | ✓ | ✓ | | ✓ | | | ✓ | ✓ | |
| | | 547 | | | Men | | 1.00 (0.71 - 1.42) | ✓ | ✓ | ✓ | ✓ | ✓ | | ✓ | | | ✓ | ✓ | |
| | | 270 | | | Women | | 1.17 (0.65 - 2.11) | ✓ | ✓ | ✓ | ✓ | ✓ | | ✓ | | | ✓ | ✓ | |
| Cuthbertson 2020 USA (32978174)[80] | ARIC Study Prospective Cohort Age:68.8 years | 361 / 14,508 23.6 years | cancer registry and medical records | Self-reported questionnaire, **Television watching frequency** | Incidence, colorectal cancer | seldom/never vs often/very often | 0.96 (0.70 - 1.31) | ✓ | ✓ | ✓ | ✓ | | | | | | | ✓ | |

| Author, Year Country (PMID) | Study name, type Age, sex, ethnicity, characteristics of participants | Cases/ Study size Median follow-up (years) | Case ascertainment | Exposure Assessment, detail | Outcome Sub-population | Comparison | RR (95%CI) Ptrend | Adjustments | | | | | | | | | | | |
|---|---|---|---|---|---|---|---|---|---|---|---|---|---|---|---|---|---|---|---|
| | | | | | | | | Age | Sex | Smo | Alc | Obe | Phy | Diet | Med | Mens | Repr | Oth | Skin |
| Hunter 2020 UK (32746843)[78] | M/F<br>Multi-ethnic<br>UK Biobank<br>Prospective Cohort<br>Range:40-69 years<br>M/F | 3,358 / 28,992<br>7.6 years | cancer registries | Self-reported questionnaire, **Television watching** | Incidence, colorectal cancer | >5 vs 1-3 hours/day | 1.03 (0.88 - 1.20) | ✓ | ✓ | ✓ | ✓ | ✓ | | ✓ | | | ✓ | ✓ | |
| | | 3,358 | | | | Per 1 hour day | 1.02 (1.00 - 1.04) Ptrend:0.13 | ✓ | ✓ | ✓ | ✓ | ✓ | | ✓ | | | ✓ | ✓ | |
| | | 2,155 | | | Colon cancer | >5 vs 1-3 hours/day | 1.17 (0.98 - 1.41) | ✓ | ✓ | ✓ | ✓ | ✓ | | ✓ | | | ✓ | ✓ | |
| | | 2,155 | | | | Per 1 hour day | 1.04 (1.01 - 1.06) Ptrend:0.02 | ✓ | ✓ | ✓ | ✓ | ✓ | | ✓ | | | ✓ | ✓ | |
| | | 1,127 | | | Rectal cancer | >5 vs 1-3 hours/day | 0.82 (0.62 - 1.10) | ✓ | ✓ | ✓ | ✓ | ✓ | | ✓ | | | ✓ | ✓ | |
| | | 1,127 | | | Rectal cancer | Per 1 hour day | 0.99 (0.95 - 1.03) Ptrend:0.67 | ✓ | ✓ | ✓ | ✓ | ✓ | | ✓ | | | ✓ | ✓ | |
| | | 835 | | | Incidence, colorectal cancer obese | | 1.03 (0.99 - 1.07) | ✓ | ✓ | ✓ | ✓ | ✓ | | ✓ | | | ✓ | ✓ | |
| | | 2,228 | | | Non obese | | 1.01 (0.99 - 1.04) | ✓ | ✓ | ✓ | ✓ | ✓ | | ✓ | | | ✓ | ✓ | |
| | | 572 | | | Incidence, colon cancer, obese | | 1.04 (0.99 - 1.09) | ✓ | ✓ | ✓ | ✓ | ✓ | | ✓ | | | ✓ | ✓ | |
| | | 1,400 | | | Non obese | | 1.03 (1.00 - 1.07) | ✓ | ✓ | ✓ | ✓ | ✓ | | ✓ | | | ✓ | ✓ | |
| | | 256 | | | Incidence, rectal cancer, obese | | 1.01 (0.94 - 1.09) | ✓ | ✓ | ✓ | ✓ | ✓ | | ✓ | | | ✓ | ✓ | |
| | | 770 | | | Non obese | | 0.98 (0.94 - 1.03) | ✓ | ✓ | ✓ | ✓ | ✓ | | ✓ | | | ✓ | ✓ | |
| | | 969 | | | High physical activity | | 1.03 (0.99 - 1.08) | ✓ | ✓ | ✓ | ✓ | ✓ | | ✓ | | | ✓ | ✓ | |
| | | 1,008 | | | Moderate physical activity | | 1.04 (1.00 - 1.09) | ✓ | ✓ | ✓ | ✓ | ✓ | | ✓ | | | ✓ | ✓ | |
| | | 429 | | | Low physical activity | | 0.98 (0.93 - 1.04) | ✓ | ✓ | ✓ | ✓ | ✓ | | ✓ | | | ✓ | ✓ | |
| | | 583 | | | Incidence, colon cancer, high physical activity | | 1.01 (0.95 - 1.07) | ✓ | ✓ | ✓ | ✓ | ✓ | | ✓ | | | ✓ | ✓ | |
| | | 658 | | | Moderate physical activity | | 1.06 (1.01 - 1.11) | ✓ | ✓ | ✓ | ✓ | ✓ | | ✓ | | | ✓ | ✓ | |
| | | 290 | | | Low physical activity | | 1.01 (0.95 - 1.08) | ✓ | ✓ | ✓ | ✓ | ✓ | | ✓ | | | ✓ | ✓ | |
| | | 363 | | | Incidence, rectal cancer, high physical activity | | 1.09 (1.01 - 1.17) | ✓ | ✓ | ✓ | ✓ | ✓ | | ✓ | | | ✓ | ✓ | |
| | | 327 | | | moderate physical activity | | 1.00 (0.93 - 1.08) | ✓ | ✓ | ✓ | ✓ | ✓ | | ✓ | | | ✓ | ✓ | |
| | | 131 | | | low physical activity | | 0.96 (0.86 - 1.06) | ✓ | ✓ | ✓ | ✓ | ✓ | | ✓ | | | ✓ | ✓ | |
| | | 1,407 | | | Incidence, colorectal cancer, never smokers | | 1.02 (0.98 - 1.06) | ✓ | ✓ | | ✓ | ✓ | | ✓ | | | ✓ | ✓ | |
| | | 480 | | | Former heavy smokers | | 0.97 (0.92 - 1.03) | ✓ | ✓ | | ✓ | ✓ | | ✓ | | | ✓ | ✓ | |
| | | 864 | | | Former light smokers | | 1.03 (0.99 - 1.08) | ✓ | ✓ | | ✓ | ✓ | | ✓ | | | ✓ | ✓ | |

| Author, Year Country (PMID) | Study name, type Age, sex, ethnicity, characteristics of participants | Cases/ Study size Median follow-up (years) | Case ascertainment | Exposure Assessment, detail | Outcome Sub-population | Comparison | RR (95%CI) Ptrend | Adjustments | | | | | | | | | | | |
|---|---|---|---|---|---|---|---|---|---|---|---|---|---|---|---|---|---|---|---|
| | | | | | | | | Age | Sex | Smo | Alc | Obe | Phy | Diet | Med | Mens | Repr | Oth | Skin |
| | | 157 | | | Current heavy smokers | | 1.06 (0.99 - 1.14) | ✓ | ✓ | | ✓ | ✓ | | ✓ | | | ✓ | ✓ | |
| | | 155 | | | Current light smokers | | 1.04 (0.95 - 1.14) | ✓ | ✓ | | ✓ | ✓ | | ✓ | | | ✓ | ✓ | |
| | | 939 | | | Never smokers | | 1.03 (0.99 - 1.08) | ✓ | ✓ | | ✓ | ✓ | | ✓ | | | ✓ | ✓ | |
| | | 295 | | | Former heavy smokers | | 0.97 (0.90 - 1.04) | ✓ | ✓ | | ✓ | ✓ | | ✓ | | | ✓ | ✓ | |
| | | 550 | | | Incidence, colon cancer, Former light smokers | | 1.05 (1.00 - 1.11) | ✓ | ✓ | | ✓ | ✓ | | ✓ | | | ✓ | ✓ | |
| | | 88 | | | Current heavy smokers | | 1.10 (1.01 - 1.21) | ✓ | ✓ | | ✓ | ✓ | | ✓ | | | ✓ | ✓ | |
| | | 100 | | | Current light smokers | | 1.10 (0.99 - 1.21) | ✓ | ✓ | | ✓ | ✓ | | ✓ | | | ✓ | ✓ | |
| | | 441 | | | Incidence, rectal cancer, never smokers | | 1.01 (0.94 - 1.07) | ✓ | ✓ | | ✓ | ✓ | | ✓ | | | ✓ | ✓ | |
| | | 178 | | | Former heavy smokers | | 0.98 (0.89 - 1.07) | ✓ | ✓ | | ✓ | ✓ | | ✓ | | | ✓ | ✓ | |
| | | 301 | | | Former light smokers | | 1.00 (0.92 - 1.08) | ✓ | ✓ | | ✓ | ✓ | | ✓ | | | ✓ | ✓ | |
| | | 55 | | | Current heavy smokers | | 1.00 (0.88 - 1.14) | ✓ | ✓ | | ✓ | ✓ | | ✓ | | | ✓ | ✓ | |
| | | 51 | | | Current light smokers | | 0.91 (0.77 - 1.09) | ✓ | ✓ | | ✓ | ✓ | | ✓ | | | ✓ | ✓ | |
| Park 2019 USA (30910780)[73] | Multiethnic Cohort Prospective Cohort Range:45-75 years M/F Multi-ethnic | 2,192 / 172,502 16.8 years | Cancer registry and national death index | Self-reported questionnaire, **Sitting while watching tv** | Incidence, colorectal cancer, Men | | Ptrend:0.72 | ✓ | | ✓ | ✓ | ✓ | | ✓ | ✓ | | | ✓ | |
| | | 1,891 | | | Women | | Ptrend:0.36 | ✓ | | ✓ | ✓ | ✓ | | ✓ | ✓ | | ✓ | ✓ | |
| Morris 2018 UK (29520109)[67] | UK Biobank Prospective Cohort Range:40-69 years M/F | 1,382 / 430,584 5.6 years | national cancer registers | Self-administered Questionnaire, **Television watching**: television watching time h/day | Colorectal cancer, Men | 4-5 vs <1 h/day | 1.35 (1.13 - 1.61) | ✓ | ✓ | ✓ | ✓ | | | ✓ | | | ✓ | ✓ | |
| | | 999 | | | Women | | 1.11 (0.91 - 1.35) | ✓ | ✓ | ✓ | ✓ | | | ✓ | | | ✓ | ✓ | |
| | | 830 | | | Colon cancer Men | | 1.45 (1.15 - 1.83) | ✓ | ✓ | ✓ | ✓ | | | ✓ | | | ✓ | ✓ | |
| | | 721 | | | Women | | 1.09 (0.87 - 1.37) | ✓ | ✓ | ✓ | ✓ | | | ✓ | | | ✓ | ✓ | |
| | | 780 | | | Proximal colon cancer | >5 vs <1 h/d | 1.29 (0.93 - 1.80) | ✓ | ✓ | ✓ | ✓ | | | ✓ | | | ✓ | ✓ | |
| | | 411 | | | Men | 4-5 vs <1 h/day | 1.45 (1.05 - 1.99) | ✓ | ✓ | ✓ | ✓ | | | ✓ | | | ✓ | ✓ | |
| | | 369 | | | Women | | 1.14 (0.83 - 1.57) | ✓ | ✓ | ✓ | ✓ | | | ✓ | | | ✓ | ✓ | |
| | | 693 | | | Distal colon cancer | >5 vs <1 h/d | 1.33 (0.92 - 1.92) | ✓ | ✓ | ✓ | ✓ | | | ✓ | | | ✓ | ✓ | |
| | | 381 | | | Men | 4-5 vs <1 h/day | 1.45 (1.02 - 2.06) | ✓ | ✓ | ✓ | ✓ | | | ✓ | | | ✓ | ✓ | |
| | | 312 | | | Women | | 1.09 (0.76 - 1.56) | ✓ | ✓ | ✓ | ✓ | | | ✓ | | | ✓ | ✓ | |
| | | 816 | | | Rectal cancer | >5 vs <1 h/d | 1.13 (0.79 - 1.61) | ✓ | ✓ | ✓ | ✓ | | | ✓ | | | ✓ | ✓ | |

| Author, Year Country (PMID) | Study name, type Age, sex, ethnicity, characteristics of participants | Cases/ Study size Median follow-up (years) | Case ascertainment | Exposure Assessment, detail | Outcome Sub-population | Comparison | RR (95%CI) Ptrend | Adjustments Age | Sex | Smo | Alc | Obe | Phy | Diet | Med | Mens | Repr | Oth | Skin |
|---|---|---|---|---|---|---|---|---|---|---|---|---|---|---|---|---|---|---|---|
| | | 545 | | | Men | 4-5 vs <1 h/day | 1.24 (0.95 - 1.63) | ✓ | ✓ | ✓ | ✓ | | | ✓ | | | ✓ | ✓ | |
| | | 271 | | | Women | | 1.15 (0.77 - 1.73) | ✓ | ✓ | ✓ | ✓ | | | ✓ | | | ✓ | ✓ | |
| | | 1,382 | | | Colorectal cancer, Men | | 1.28 (1.07 - 1.53) | ✓ | ✓ | ✓ | ✓ | ✓ | | ✓ | | | ✓ | ✓ | |
| | | 999 | | | Women | | 1.08 (0.88 - 1.32) | ✓ | ✓ | ✓ | ✓ | ✓ | | ✓ | | | ✓ | ✓ | |
| | | 830 | | | Colon cancer, Men | | 1.37 (1.08 - 1.72) | ✓ | ✓ | ✓ | ✓ | ✓ | | ✓ | | | ✓ | ✓ | |
| | | 721 | | | Women | | 1.05 (0.84 - 1.33) | ✓ | ✓ | ✓ | ✓ | ✓ | | ✓ | | | ✓ | ✓ | |
| | | 780 | | | Proximal colon cancer | >5 vs <1 h/d | 1.18 (0.85 - 1.65) | ✓ | ✓ | ✓ | ✓ | ✓ | | ✓ | | | ✓ | ✓ | |
| | | 411 | | | Men | 4-5 vs <1 h/day | 1.35 (0.98 - 1.86) | ✓ | ✓ | ✓ | ✓ | ✓ | | ✓ | | | ✓ | ✓ | |
| | | 369 | | | Women | | 1.08 (0.78 - 1.48) | ✓ | ✓ | ✓ | ✓ | ✓ | | ✓ | | | ✓ | ✓ | |
| | | 693 | | | Distal colon cancer | | 1.27 (0.88 - 1.83) | ✓ | ✓ | ✓ | ✓ | ✓ | | ✓ | | | ✓ | ✓ | |
| | | 381 | | | Men | >5 vs <1 h/d | 1.39 (0.98 - 1.97) | ✓ | ✓ | ✓ | ✓ | ✓ | | ✓ | | | ✓ | ✓ | |
| | | 312 | | | Women | 4-5 vs <1 h/day | 1.07 (0.74 - 1.53) | ✓ | ✓ | ✓ | ✓ | ✓ | | ✓ | | | ✓ | ✓ | |
| | | 816 | | | Rectal cancer | >5 vs <1 h/d | 1.09 (0.76 - 1.55) | ✓ | ✓ | ✓ | ✓ | ✓ | | ✓ | | | ✓ | ✓ | |
| | | 545 | | | Men | | 1.20 (0.91 - 1.58) | ✓ | ✓ | ✓ | ✓ | ✓ | | ✓ | | | ✓ | ✓ | |
| | | 271 | | | Women | | 1.14 (0.76 - 1.71) | ✓ | ✓ | ✓ | ✓ | ✓ | | ✓ | | | ✓ | ✓ | |
| Nguyen 2018 USA (30740587)[71] | NHS II Prospective Cohort Age:45 years F nurses | 165 / 89,278 13.9 years | Cancer registry | Self-reported questionnaire, **Sitting while watching tv** | Incidence, colorectal cancer (diagnosed after 50 years old) | >14 vs <7 hours/week | 1.20 (0.79 – 1.84) Ptrend:0.29 | | | ✓ | ✓ | | ✓ | ✓ | ✓ | ✓ | ✓ | ✓ | |
| | | 165 | | | Incidence, colorectal cancer (diagnosed after 50 years old) | | 1.20 (0.80 – 1.89) Ptrend:0.24 | | | ✓ | ✓ | ✓ | ✓ | ✓ | ✓ | ✓ | ✓ | ✓ | |
| | | 118 | | | Incidence, colorectal cancer (diagnosed before 50 years old) | | 1.77 (1.12 - 2.78) Ptrend:0.02 | | | ✓ | ✓ | | ✓ | ✓ | ✓ | ✓ | ✓ | ✓ | |
| | | 118 | | | Incidence, colorectal cancer (diagnosed before 50 years old) | | 1.69 (1.07 - 2.67) Ptrend:0.03 | | | ✓ | ✓ | ✓ | ✓ | ✓ | ✓ | ✓ | ✓ | ✓ | |
| | | 82 | | | Colon cancer (diagnosed before 50 years old) | | 1.47 (0.85 - 2.54) Ptrend:0.22 | | | ✓ | ✓ | ✓ | ✓ | ✓ | ✓ | ✓ | ✓ | ✓ | |
| | | 36 | | , | Rectal cancer (diagnosed before 50 years old) | | 2.44 (1.03 - 5.78) Ptrend:0.04 | | | ✓ | ✓ | ✓ | ✓ | ✓ | ✓ | ✓ | ✓ | ✓ | |
| Eaglehouse 2017 Singapore (28542077)[62] | Singapore Chinese Health study Prospective Cohort Age:56.4 years M/F Chinese | 1,981 / 61,308 16.8 years | Cancer registry | Self-reported questionnaire, **Sitting while watching tv** | Incidence, colorectal cancer | >3 vs 0 - 0.9 hours/day | 1.08 (0.95 - 1.23) Ptrend:0.28 | ✓ | ✓ | ✓ | ✓ | ✓ | | ✓ | | | | ✓ | |
| | | 1,249 | | | Colon cancer | | 1.11 (0.94 - 1.32) Ptrend:0.29 | ✓ | ✓ | ✓ | ✓ | ✓ | | ✓ | | | | ✓ | |

| Author, Year Country (PMID) | Study name, type Age, sex, ethnicity, characteristics of participants | Cases/ Study size Median follow-up (years) | Case ascertainment | Exposure Assessment, detail | Outcome Sub-population | Comparison | RR (95%CI) Ptrend | Adjustments: Age | Sex | Smo | Alc | Obe | Phy | Diet | Med | Mens | Repr | Oth | Skin |
|---|---|---|---|---|---|---|---|---|---|---|---|---|---|---|---|---|---|---|---|
| | | 732 | | | Rectal cancer | | 1.03 (0.84 - 1.27)<br>Ptrend:0.64 | ✓ | ✓ | ✓ | ✓ | ✓ | | ✓ | | | | ✓ | |
| | | 171 | | | Colorectal cancer, BMI >=27.5 | ≥3 vs =<2 hours/day | 0.91 (0.66 - 1.23) | ✓ | ✓ | ✓ | ✓ | ✓ | | ✓ | | | | ✓ | |
| | | 124 | | | BMI 18.5-27.4 | | 1.05 (0.95 - 1.15) | ✓ | ✓ | ✓ | ✓ | ✓ | | ✓ | | | | ✓ | |
| | | 124 | | | BMI <18.5 | | 1.12 (0.78 - 1.61) | ✓ | ✓ | ✓ | ✓ | ✓ | | ✓ | | | | ✓ | |
| Keum 2016 USA (26649988)[58] | Nurses Health Study (NHS) + Health Professionals Follow-up Study (HPFS)<br>Prospective Cohort<br>Range:30-75 years<br>M/F<br>Health professionals | 913 / 106,521<br>22 years | Self-report verified by medical record | Self-reported questionnaire, **Sitting while watching tv** | Incidence, colorectal cancer, Men | >21 vs 0-6.9 hours/week | 1.06 (0.84 - 1.34)<br>Ptrend:0.93 | ✓ | | ✓ | ✓ | | ✓ | ✓ | ✓ | | | ✓ | |
| | | 913 | | | | Per 7 hours week | 1.00 (0.94 - 1.07) | ✓ | | ✓ | ✓ | | ✓ | ✓ | ✓ | | | ✓ | |
| | | 1,119 | | | Women | >21 vs 0-6.9 hours/week | 1.21 (1.02 - 1.43)<br>Ptrend:0.01 | ✓ | | ✓ | ✓ | | ✓ | ✓ | ✓ | ✓ | ✓ | ✓ | |
| | | 1,119 | | | | Per 7 hours week | 1.04 (1.01 - 1.08) | ✓ | | ✓ | ✓ | | ✓ | ✓ | ✓ | ✓ | ✓ | ✓ | |
| | | 596 | | | Incidence, colon cancer, Men | >21 vs 0-6.9 hours/week | 1.13 (0.85 - 1.50)<br>Ptrend:0.42 | ✓ | | ✓ | ✓ | | ✓ | ✓ | ✓ | | | ✓ | |
| | | 596 | | | | Per 7 hours week | 1.03 (0.96 - 1.11) | ✓ | | ✓ | ✓ | | ✓ | ✓ | ✓ | | | ✓ | |
| | | 939 | | | Women | >21 vs 0-6.9 hours/week | 1.19 (0.98 - 1.45)<br>Ptrend:0.02 | ✓ | | ✓ | ✓ | | ✓ | ✓ | ✓ | ✓ | ✓ | ✓ | |
| | | 884 | | | | Per 7 hours week | 1.05 (1.01 - 1.08) | ✓ | | ✓ | ✓ | | ✓ | ✓ | ✓ | ✓ | ✓ | ✓ | |
| | | 210 | | | Incidence, rectal cancer, Men | >21 vs 0-6.9 hours/week | 1.21 (0.76 - 1.93)<br>Ptrend:0.63 | ✓ | | ✓ | ✓ | | ✓ | ✓ | ✓ | | | ✓ | |
| | | 210 | | | | Per 7 hours week | 1.03 (0.91 - 1.17) | ✓ | | ✓ | ✓ | | ✓ | ✓ | ✓ | | | ✓ | |
| | | 215 | | | Women | >21 vs 0-6.9 hours/week | 1.32 (0.90 - 1.95)<br>Ptrend:0.38 | ✓ | | ✓ | ✓ | | ✓ | ✓ | ✓ | ✓ | ✓ | ✓ | |
| | | 215 | | | | Per 7 hours week | 1.04 (0.96 - 1.12) | ✓ | | ✓ | ✓ | | ✓ | ✓ | ✓ | ✓ | ✓ | ✓ | |
| | | 913 | | | Incidence, colorectal cancer, Men | >21 vs 0-6.9 hours/week | 1.02 (0.81 - 1.29)<br>Ptrend:0.79 | ✓ | | ✓ | ✓ | ✓ | ✓ | ✓ | ✓ | | | ✓ | |
| | | 913 | | | | Per 7 hours week | 0.99 (0.93 - 1.06) | ✓ | | ✓ | ✓ | ✓ | ✓ | ✓ | ✓ | | | ✓ | |
| | | 1,119 | | | Women | >21 vs 0-6.9 hours/week | 1.18 (1.00 - 1.41)<br>Ptrend:0.02 | ✓ | | ✓ | ✓ | ✓ | ✓ | ✓ | ✓ | ✓ | ✓ | ✓ | |
| | | 1,119 | | | | Per 7 hours week | 1.04 (1.01 - 1.08) | ✓ | | ✓ | ✓ | ✓ | ✓ | ✓ | ✓ | ✓ | ✓ | ✓ | |
| | | 596 | | | Incidence, colon cancer, Men | >21 vs 0-6.9 hours/week | 1.09 (0.82 - 1.45)<br>Ptrend:0.6 | ✓ | | ✓ | ✓ | ✓ | ✓ | ✓ | ✓ | | | ✓ | |
| | | 596 | | | | Per 7 hours week | 1.03 (0.96 - 1.11) | ✓ | | ✓ | ✓ | | ✓ | ✓ | ✓ | | | ✓ | |
| | | 939 | | | Incidence, colon cancer, Women | >21 vs 0-6.9 hours/week | 1.18 (0.97 - 1.43)<br>Ptrend:0.03 | ✓ | | ✓ | ✓ | ✓ | ✓ | ✓ | ✓ | ✓ | ✓ | ✓ | |
| | | 884 | | | | Per 7 hours week | 1.04 (1.00 - 1.08) | ✓ | | ✓ | ✓ | ✓ | ✓ | ✓ | ✓ | ✓ | ✓ | ✓ | |

| Author, Year Country (PMID) | Study name, type Age, sex, ethnicity, characteristics of participants | Cases/ Study size Median follow-up (years) | Case ascertainment | Exposure Assessment, detail | Outcome Sub-population | Comparison | RR (95%CI) Ptrend | Adjustments | | | | | | | | | | | |
|---|---|---|---|---|---|---|---|---|---|---|---|---|---|---|---|---|---|---|---|
| | | | | | | | | Age | Sex | Smo | Alc | Obe | Phy | Diet | Med | Mens | Repr | Oth | Skin |
| | | 210 | | | Incidence, rectal cancer, Men | >21 vs 0-6.9 hours/week | 1.17 (0.73 - 1.87) Ptrend:0.75 | ✓ | | ✓ | ✓ | ✓ | ✓ | ✓ | ✓ | | | ✓ | |
| | | 210 | | | | Per 7 hours week | 1.02 (0.90 - 1.16) | ✓ | | ✓ | ✓ | | ✓ | ✓ | ✓ | | | ✓ | |
| | | 215 | | | Women | >21 vs 0-6.9 hours/week | 1.29 (0.87 - 1.90) Ptrend:0.46 | ✓ | | ✓ | ✓ | ✓ | ✓ | ✓ | ✓ | | | ✓ | |
| | | 215 | | | | Per 7 hours week | 1.02 (0.95 - 1.11) | ✓ | | ✓ | ✓ | ✓ | ✓ | ✓ | ✓ | ✓ | ✓ | ✓ | |
| Howard 2008 USA (18437512)[36] | NIH- AARP Diet and Health Study Prospective Cohort Age:62 years M/F Multi-ethnic | 1,261 / 488,720 6.9 years | Cancer registry | Self-reported questionnaire, **Television watching**: Time spent watching TV or videos, over past 12 months, from 2nd questionnaire | Incidence / Mortality, colon cancer, Men | 9 - vs 0 - 2.9 hours/day | 1.61 (1.14 - 2.27) Ptrend:<0.001 | ✓ | | ✓ | ✓ | | ✓ | ✓ | | | | ✓ | |
| | | 669 | | | Women | | 1.45 (0.99 - 2.12) Ptrend:0.174 | ✓ | | ✓ | ✓ | | ✓ | ✓ | | | ✓ | ✓ | |
| | | 1,261 | | | Incidence / Mortality, colon cancer, Men | | 1.56 (1.11 - 2.20) Ptrend:0.002 | ✓ | | ✓ | ✓ | ✓ | ✓ | ✓ | | | | ✓ | |
| | | 669 | | | Women | | 1.45 (0.99 - 2.13) Ptrend:0.180 | ✓ | | ✓ | ✓ | ✓ | ✓ | ✓ | | | ✓ | ✓ | |
| Li 2021 Japan (33138348)[81] | JACC Japan Prospective Cohort Range:40-79 years M/F | 749 / 90,834 19.1 years | Follow up survey/cancer registry/vital statistics registry | Self-reported questionnaire, **Television watching** | Mortality, colorectal cancer | >4.5 vs <1.5 hrs/day | 1.33 (1.02 - 1.73) Ptrend:0.038 | ✓ | ✓ | ✓ | ✓ | ✓ | ✓ | ✓ | | | | ✓ | |
| | | 749 | | | | Per 1 hour | | ✓ | ✓ | ✓ | ✓ | ✓ | ✓ | ✓ | | | | ✓ | |
| | | 531 | | | Mortality, colon cancer | >4.5 vs <1.5 hrs/day | 1.43 (1.04 - 1.96) Ptrend:0.049 | ✓ | ✓ | ✓ | ✓ | ✓ | ✓ | ✓ | | | | ✓ | |
| | | 531 | | | | Per 1 hour | 1.07 (1.02 - 1.13) | ✓ | ✓ | ✓ | ✓ | ✓ | ✓ | ✓ | | | | ✓ | |
| | | 218 | | | Mortality, rectal cancer | >4.5 vs <1.5 hrs/day | 1.13 (0.69 - 1.85) Ptrend:0.450 | ✓ | ✓ | ✓ | ✓ | ✓ | ✓ | ✓ | | | | ✓ | |
| | | 218 | | | | Per 1 hour | | ✓ | ✓ | ✓ | ✓ | ✓ | ✓ | ✓ | | | | ✓ | |
| | | 150 | | | Mortality, colorectal cancer, BMI >=25 | >4.5 vs <1.5 hrs/day | 1.16 (0.63 - 2.12) Ptrend:0.751 | ✓ | ✓ | ✓ | ✓ | ✓ | ✓ | ✓ | | | | ✓ | |
| | | 150 | | | | Per 1 hour | | ✓ | ✓ | ✓ | ✓ | ✓ | ✓ | ✓ | | | | ✓ | |
| | | 544 | | | BMI <25 | >4.5 vs <1.5 hrs/day | 1.39 (1.01 - 1.90) Ptrend:0.034 | ✓ | ✓ | ✓ | ✓ | ✓ | ✓ | ✓ | | | | ✓ | |
| | | 544 | | | | Per 1 hour | | ✓ | ✓ | ✓ | ✓ | ✓ | ✓ | ✓ | | | | ✓ | |
| | | 108 | | | Mortality, colon cancer, BMI >=25 | >4.5 vs <1.5 hrs/day | 1.62 (0.76 - 3.47) Ptrend:0.425 | ✓ | ✓ | ✓ | ✓ | ✓ | ✓ | ✓ | | | | ✓ | |
| | | 108 | | | | Per 1 hour | | ✓ | ✓ | ✓ | ✓ | ✓ | ✓ | ✓ | | | | ✓ | |
| | | 381 | | | BMI<25 | >4.5 vs <1.5 hrs/day | 1.39 (0.96 - 2.03) Ptrend:0.090 | ✓ | ✓ | ✓ | ✓ | ✓ | ✓ | ✓ | | | | ✓ | |
| | | 381 | | | | Per 1 hour | | ✓ | ✓ | ✓ | ✓ | ✓ | ✓ | ✓ | | | | ✓ | |
| | | 42 | | | Mortality, rectal cancer, BMI >=25 | >4.5 vs <1.5 hrs/day | 0.56 (0.19 - 1.65) Ptrend:0.520 | ✓ | ✓ | ✓ | ✓ | ✓ | ✓ | ✓ | | | | ✓ | |
| | | 42 | | | | Per 1 hour | 0.95 (0.78 - 1.17) | ✓ | ✓ | ✓ | ✓ | ✓ | ✓ | ✓ | | | | ✓ | |

| Author, Year Country (PMID) | Study name, type Age, sex, ethnicity, characteristics of participants | Cases/ Study size Median follow-up (years) | Case ascertainment | Exposure Assessment, detail | Outcome Sub-population | Comparison | RR (95%CI) Ptrend | Adjustments Age | Sex | Smo | Alc | Obe | Phy | Diet | Med | Mens | Repr | Oth | Skin |
|---|---|---|---|---|---|---|---|---|---|---|---|---|---|---|---|---|---|---|---|
| | | 163 | | | BMI <25 | >4.5 vs <1.5 hrs/day | 1.40 (0.78 - 2.49) Ptrend:0.206 | ✓ | ✓ | ✓ | ✓ | ✓ | ✓ | ✓ | | | | ✓ | |
| | | 163 | | | | Per 1 hour | | ✓ | ✓ | ✓ | ✓ | ✓ | ✓ | ✓ | | | | ✓ | |
| | | 365 | | | Mortality, colorectal cancer, High recreational activity | >4.5 vs <1.5 hrs/day | 1.53 (1.04 - 2.24) Ptrend:0.045 | ✓ | ✓ | ✓ | ✓ | ✓ | ✓ | ✓ | | | | ✓ | |
| | | 365 | | | | Per 1 hour | | ✓ | ✓ | ✓ | ✓ | ✓ | ✓ | ✓ | | | | ✓ | |
| | | 362 | | | Mortality, colorectal cancer, low leisure time PA | >4.5 vs <1.5 hrs/day | 1.10 (0.75 - 1.61) Ptrend:0.515 | ✓ | ✓ | ✓ | ✓ | ✓ | ✓ | ✓ | | | | ✓ | |
| | | 362 | | | | Per 1 hour | | ✓ | ✓ | ✓ | ✓ | ✓ | ✓ | ✓ | | | | ✓ | |
| | | 258 | | | Mortality, colon cancer, High recreational activity | >4.5 vs <1.5 hrs/day | 1.72 (1.08 - 2.76) Ptrend:0.059 | ✓ | ✓ | ✓ | ✓ | ✓ | ✓ | ✓ | | | | ✓ | |
| | | 258 | | | | Per 1 hour | | ✓ | ✓ | ✓ | ✓ | ✓ | ✓ | ✓ | | | | ✓ | |
| | | 257 | | | Mortality, colon cancer, low leisure time PA | >4.5 vs <1.5 hrs/day | 1.13 (0.72 - 1.77) Ptrend:0.575 | ✓ | ✓ | ✓ | ✓ | ✓ | ✓ | ✓ | | | | ✓ | |
| | | 257 | | | | Per 1 hour | | ✓ | ✓ | ✓ | ✓ | ✓ | ✓ | ✓ | | | | ✓ | |
| | | 107 | | | Mortality, rectal cancer, High recreational activity | >4.5 vs <1.5 hrs/day | 1.20 (0.60 - 2.39) Ptrend:0.456 | ✓ | ✓ | ✓ | ✓ | ✓ | ✓ | ✓ | | | | ✓ | |
| | | 107 | | | | Per 1 hour | 1.03 (0.90 - 1.18) | ✓ | ✓ | ✓ | ✓ | ✓ | ✓ | ✓ | | | | ✓ | |
| | | 105 | | | Mortality, rectal cancer, low leisure time PA | >4.5 vs <1.5 hours/day | 1.02 (0.49 - 2.11) Ptrend:0.727 | ✓ | ✓ | ✓ | ✓ | ✓ | ✓ | ✓ | | | | ✓ | |
| | | 105 | | | | Per 1 hour | | ✓ | ✓ | ✓ | ✓ | ✓ | ✓ | ✓ | | | | ✓ | |
| Kim 2013 USA (24062293)[50] | Multiethnic Cohort Study (MEC) Prospective Cohort M/F Multi-ethnic | / 134,596 13.7 years | National Death Index | Self-reported questionnaire **Other sedentary activity**: Daily sitting watching TV (hours) | Mortality, colorectal cancer, Men | ≥5 h vs <1 hours /day | 1.08 (0.68 - 1.73) Ptrend:0.66 | ✓ | | ✓ | ✓ | | ✓ | ✓ | | | | ✓ | |
| | | NA | | | Women | | 1.05 (0.65 - 1.69) Ptrend:0.67 | ✓ | | ✓ | ✓ | | ✓ | ✓ | | | | ✓ | |
| **Hepatobiliary tract** | | | | | | | | | | | | | | | | | | | |
| Hunter 2020 UK (32746843)[78] | UK Biobank Prospective Cohort Range:40-69 years M/F | 456 / 28,992 7.6 years | cancer registries | Self-reported questionnaire, **Television watching**: | Incidence, hepatobiliary tract cancer | >5 vs 1-3 hours/day | 1.26 (0.90 - 1.77) | ✓ | ✓ | ✓ | ✓ | ✓ | | | | | | ✓ | |
| | | 456 | | | | Per 1 hour day | 1.01 (0.96 - 1.07) Ptrend:0.62 | ✓ | ✓ | ✓ | ✓ | ✓ | | | | | | ✓ | |
| | | 166 | | | Obese | | 1.00 (0.92 - 1.09) | ✓ | ✓ | ✓ | ✓ | | | | | | | ✓ | |
| | | 262 | | | Non obese | | 1.02 (0.94 - 1.10) | ✓ | ✓ | ✓ | ✓ | | | | | | | ✓ | |
| | | 119 | | | High physical activity | | 1.06 (0.94 - 1.19) | ✓ | ✓ | ✓ | ✓ | ✓ | | | | | | ✓ | |

| Author, Year Country (PMID) | Study name, type Age, sex, ethnicity, characteristics of participants | Cases/ Study size Median follow-up (years) | Case ascertainment | Exposure Assessment, detail | Outcome Sub-population | Comparison | RR (95%CI) Ptrend | Adjustments Age | Sex | Smo | Alc | Obe | Phy | Diet | Med | Mens | Repr | Oth | Skin |
|---|---|---|---|---|---|---|---|---|---|---|---|---|---|---|---|---|---|---|---|
| | | 138 | | | Moderate physical activity | | 1.01 (0.91 - 1.12) | ✓ | ✓ | ✓ | ✓ | ✓ | | | | | | ✓ | |
| | | 74 | | | Low physical activity | | 0.97 (0.86 - 1.10) | ✓ | ✓ | ✓ | ✓ | ✓ | | | | | | ✓ | |
| | | 164 | | | Never smokers | | 1.03 (0.93 - 1.14) | ✓ | ✓ | | ✓ | ✓ | | | | | | ✓ | |
| | | 78 | | | Former heavy smokers | | 0.95 (0.83 - 1.08) | ✓ | ✓ | | ✓ | ✓ | | | | | | ✓ | |
| | | 128 | | | Former light smokers | | 0.99 (0.88 - 1.11) | ✓ | ✓ | | ✓ | ✓ | | | | | | ✓ | |
| | | 33 | | | Current heavy smokers | | 1.03 (0.89 - 1.20) | ✓ | ✓ | | ✓ | ✓ | | | | | | ✓ | |
| | | 25 | | | Current light smokers | | 1.13 (0.97 - 1.31) | ✓ | ✓ | | ✓ | ✓ | | | | | | ✓ | |
| **Liver cancer** | | | | | | | | | | | | | | | | | | | |
| Ukawa 2014 Japan (24728669)[54] | Japan Collaborative Cohort Study Prospective Cohort Age:56.8 years M/F Japanese | 267 / 69752 | Death certificate | Self-administrated-Questionnaire, **Sitting while watching tv**: TV viewing time | Mortality, liver cancer | ≥4 vs <2 hours/day | 1.20 (0.82 - 1.77) Ptrend:0.27 | ✓ | ✓ | ✓ | ✓ | ✓ | | ✓ | ✓ | | | ✓ | |
| | | 163 | | | Men | | 1.23 (0.76 - 2.02) Ptrend:0.64 | ✓ | ✓ | ✓ | ✓ | ✓ | | ✓ | ✓ | | | ✓ | |
| | | 104 | | | Women | | 1.13 (0.62 - 2.13) Ptrend:0.34 | ✓ | ✓ | ✓ | ✓ | ✓ | | ✓ | ✓ | | | ✓ | |
| **Pancreatic cancer** | | | | | | | | | | | | | | | | | | | |
| Hunter 2020 UK (32746843)[78] | UK Biobank Prospective Cohort Range:40-69 years M/F | 615 / 28,992 7.6 years | cancer registries | Self-reported questionnaire, **Television watching** | Incidence, Pancreatic cancer | >5 vs 1-3 hours/day | 1.03 (0.74 - 1.44) | ✓ | ✓ | ✓ | ✓ | ✓ | | | | | | ✓ | |
| | | 615 | | | | Per 1 hour day | 1.03 (0.98 - 1.08) Ptrend:0.20 | ✓ | ✓ | ✓ | ✓ | ✓ | | | | | | ✓ | |
| | | 177 | | | Obese | | 1.02 (0.93 - 1.10) | ✓ | ✓ | ✓ | ✓ | | | | | | | ✓ | |
| | | 401 | | | Non obese | | 1.04 (0.98 - 1.11) | ✓ | ✓ | ✓ | ✓ | | | | | | | ✓ | |
| | | 187 | | | High physical activity | | 1.00 (0.91 - 1.10) | ✓ | ✓ | ✓ | ✓ | ✓ | | | | | | ✓ | |
| | | 190 | | | Moderate physical activity | | 1.08 (1.00 - 1.18) | ✓ | ✓ | ✓ | ✓ | ✓ | | | | | | ✓ | |
| | | 90 | | | Low physical activity | | 0.99 (0.89 - 1.11) | ✓ | ✓ | ✓ | ✓ | ✓ | | | | | | ✓ | |
| | | 262 | | | Never smokers | | 0.99 (0.92 - 1.08) | ✓ | ✓ | | ✓ | ✓ | | | | | | ✓ | |
| | | 80 | | | Former heavy smokers | | 1.12 (1.01 - 1.25) | ✓ | ✓ | | ✓ | ✓ | | | | | | ✓ | |
| | | 139 | | | Former light smokers | | 1.07 (0.96 - 1.19) | ✓ | ✓ | | ✓ | ✓ | | | | | | ✓ | |
| | | 60 | | | Current heavy smokers | | 0.96 (0.85 - 1.10) | ✓ | ✓ | | ✓ | ✓ | | | | | | ✓ | |
| | | 37 | | | Current light smokers | | 1.08 (0.92 - 1.25) | ✓ | ✓ | | ✓ | ✓ | | | | | | ✓ | |
| **Lung cancer** | | | | | | | | | | | | | | | | | | | |
| Zhang 2024 UK (38974470)[93] | UK Biobank Prospective Cohort | 8,644 / 360,271 12.52 years | Cancer registry | Self-reported questionnaire, **Screen time, recreational** | Incidence, Lung cancer | high vs low | 1.26 (1.13 - 1.40) | ✓ | ✓ | ✓ | ✓ | ✓ | ✓ | | | | | ✓ | |

| Author, Year Country (PMID) | Study name, type Age, sex, ethnicity, characteristics of participants | Cases/ Study size Median follow-up (years) | Case ascertainment | Exposure Assessment, detail | Outcome Sub-population | Comparison | RR (95%CI) Ptrend | Adjustments | | | | | | | | | | | |
|---|---|---|---|---|---|---|---|---|---|---|---|---|---|---|---|---|---|---|---|
| | | | | | | | | Age | Sex | Smo | Alc | Obe | Phy | Diet | Med | Mens | Repr | Oth | Skin |
| Bethea 2022 USA (35325667)[87] | Age:55.94 years M/F Mostly white Black Women Health Study Prospective Cohort Age:42.3 years Black Black women | 387 / 38,432 22 years | post office, relatives, linkage to National Death Index | Self-reported questionnaire, **Sitting while watching tv** | Incidence, Lung cancer | ≥1 vs <1h/week | 1.27 (0.72 - 2.21) | ✓ | | ✓ | ✓ | ✓ | | ✓ | | ✓ | | ✓ | |
| | | 51 | | | Never smokers | | 1.99 (0.58 - 8.29) | ✓ | | ✓ | ✓ | ✓ | | ✓ | | ✓ | | ✓ | |
| | | 172 | | | Former smokers | | 1.75 (0.65 - 4.76) | ✓ | | ✓ | ✓ | ✓ | | ✓ | | ✓ | | ✓ | |
| | | 162 | | | Current smoker | | 0.72 (0.33 - 1.55) | ✓ | | ✓ | ✓ | ✓ | | ✓ | | ✓ | | ✓ | |
| | | 187 | | | >=20 pack years smoking | | 0.84 (0.37 - 1.92) | ✓ | | ✓ | ✓ | ✓ | | ✓ | | ✓ | | ✓ | |
| | | 147 | | | < 20 pack years smoking | | 1.62 (0.66 - 3.97) | ✓ | | ✓ | ✓ | ✓ | | ✓ | | ✓ | | ✓ | |
| Cuthbertson 2020 USA (32978174)[80] | ARIC Study Prospective Cohort Age:69.7 years M/F Multi-ethnic | 560 / 14,508 23.6 years | cancer registry and medical records | Self-reported questionnaire, **TV watching frequency** | Incidence, Lung cancer | seldom/never vs often/very often | 1.09 (0.85 - 1.40) | ✓ | ✓ | ✓ | ✓ | | | | | | | ✓ | |
| Hunter 2020 UK (32746843)[78] | UK Biobank Prospective Cohort Range:40-69 years M/F | 2,076 / 28,992 7.6 years | cancer registries | Self-reported questionnaire, **Television watching**: | Incidence, Lung cancer | <5 vs 1-3 hours/day | 1.09 (0.94 - 1.27) | ✓ | ✓ | ✓ | ✓ | ✓ | | | | | | ✓ | |
| | | 2,076 | | | | Per 1 hour day | 1.02 (1.00 - 1.05) Ptrend:0.09 | ✓ | ✓ | ✓ | ✓ | ✓ | | | | | | ✓ | |
| | | 480 | | | Obese | | 1.01 (0.96 - 1.06) | ✓ | ✓ | ✓ | ✓ | | | | | | | ✓ | |
| | | 1,377 | | | Non obese | | 1.03 (1.00 - 1.06) | ✓ | ✓ | ✓ | ✓ | | | | | | | ✓ | |
| | | 516 | | | High physical activity | | 1.03 (0.98 - 1.09) | ✓ | ✓ | ✓ | ✓ | ✓ | | | | | | ✓ | |
| | | 555 | | | Moderate physical activity | | 1.05 (1.00 - 1.10) | ✓ | ✓ | ✓ | ✓ | ✓ | | | | | | ✓ | |
| | | 284 | | | Low physical activity | | 1.03 (0.97 - 1.08) | ✓ | ✓ | ✓ | ✓ | ✓ | | | | | | ✓ | |
| | | 256 | | | Never smokers | | 1.03 (0.95 - 1.12) | ✓ | ✓ | | ✓ | ✓ | | | | | | ✓ | |
| | | 553 | | | Former heavy smokers | | 1.02 (0.97 - 1.07) | ✓ | ✓ | | ✓ | ✓ | | | | | | ✓ | |
| | | 294 | | | Former light smokers | | 1.01 (0.94 - 1.09) | ✓ | ✓ | | ✓ | ✓ | | | | | | ✓ | |
| | | 35 | | | Current heavy smokers | | 1.01 (0.97 - 1.05) | ✓ | ✓ | | ✓ | ✓ | | | | | | ✓ | |
| | | 47 | | | Current light smokers | | 1.08 (1.01 - 1.16) | ✓ | ✓ | | ✓ | ✓ | | | | | | ✓ | |
| Ukawa 2013 Japan (23686441)[47] | Japan Collaborative Cohort study for Evaluation of Cancer (JACC) Prospective Cohort Age:57.6 years M/F | 598 / 54,258 15.6 years | Cancer registry | Self-administrated questionnaire, **Television watching** | Incidence, Lung cancer, Men | ≥4 vs <2 hours/day | 1.36 (1.04 - 1.79) Ptrend:0.06 | ✓ | | ✓ | | | | | | | | | |
| | | 200 | | | Women | | 1.01 (0.66 - 1.59) Ptrend:0.37 | ✓ | | ✓ | | | | | | | | | |

| Author, Year Country (PMID) | Study name, type Age, sex, ethnicity, characteristics of participants | Cases/ Study size Median follow-up (years) | Case ascertainment | Exposure Assessment, detail | Outcome Sub-population | Comparison | RR (95%CI) Ptrend | Adjustments Age | Sex | Smo | Alc | Obe | Phy | Diet | Med | Mens | Repr | Oth | Skin |
|---|---|---|---|---|---|---|---|---|---|---|---|---|---|---|---|---|---|---|---|
| | | 598 | | | Men | | 1.36 (1.04 - 1.80) Ptrend:0.06 | ✓ | | ✓ | ✓ | ✓ | | ✓ | | | | ✓ | |
| | | 200 | | | Women | | 1.03 (0.67 - 1.62) Ptrend:0.40 | ✓ | | ✓ | ✓ | ✓ | | ✓ | | | | ✓ | |
| Lam 2013 USA (23940620)[48] | NIH- AARP Diet and Health Study Prospective Cohort Age:61.8 years M/F never smokers | 205 / 158,415 9.96 years | Postal Service, Social Security Administration Death Master File, National Death Index | Self-reported questionnaire, **Television watching**: | Incidence, Lung cancer, never smokers | 5+ vs <3 hours/day | 1.06 (0.77 - 1.46) Ptrend:0.53 | ✓ | | | ✓ | | ✓ | ✓ | | | | ✓ | |
| Kim 2013 USA (24062293)[50] | Multiethnic Cohort Study (MEC) Prospective Cohort M/F Multi-ethnic | / 134,596 13.7 years | National Death Index | Self-administrated questionnaire, **Other sedentary activity**: Daily sitting watching TV (hours) | Mortality, Lung cancer, Men | >5 vs <1 h/day | 1.14 (0.87 - 1.51) Ptrend:0.76 | ✓ | | ✓ | ✓ | | ✓ | ✓ | | | | ✓ | |
| | | NA | | | Women | | 1.09 (0.81 - 1.49) Ptrend:0.14 | ✓ | | ✓ | ✓ | | ✓ | ✓ | | | | ✓ | |
| **Breast cancer** | | | | | | | | | | | | | | | | | | | |
| Zhang 2024 UK (38974470)[93] | UK Biobank Prospective Cohort Age:55.94 years F Mostly white | 7,817 / 360,271 12.52 years | Cancer registry | Self-reported questionnaire, **Screen time, recreational** | Incidence, Breast cancer | high vs low | 1.01 (0.96 - 1.08) | ✓ | ✓ | ✓ | ✓ | ✓ | ✓ | | | ✓ | ✓ | ✓ | |
| Sanchez-Bayona 2021 Spain (33798533)[82] | The SUN Project Prospective Cohort Age:34.7 years F University students | 101 / 10,812 11.8 years | self-reported/death certificate/ medical records | Self-reported questionnaire, **Television watching**: | Incidence, Breast cancer | >2 vs <1 hours/day | 1.67 (1.03 - 2.72) Ptrend:0.02 | | | | ✓ | ✓ | ✓ | ✓ | | ✓ | | ✓ | |
| | | 57 | | | Premenopausal | | 2.22 (1.15 - 4.28) Ptrend:0.01 | | | | ✓ | ✓ | ✓ | ✓ | | ✓ | | ✓ | |
| | | 34 | | | Postmenopausal | | 1.07 (0.45 - 2.60) Ptrend:0.87 | | | | ✓ | ✓ | ✓ | ✓ | | ✓ | ✓ | ✓ | |
| Cuthbertson 2020 USA (32978174)[80] | ARIC Study Prospective Cohort Age:66.7 years F Multi-ethnic Postmenopausal | 564 / 14,508 23.6 years | cancer registry and medical records | Self-reported questionnaire, **TV watching frequency** | Incidence, Postmenopausal breast cancer | seldom/never vs often/very often | 1.03 (0.81 - 1.30) | ✓ | | ✓ | ✓ | | | | | | | ✓ | |
| Hunter 2020 UK (32746843)[78] | UK Biobank Prospective Cohort Range:40-69 years F | 5,702 / 253,188 7.6 years | cancer registries | Self-reported questionnaire, **Television watching** | Incidence, Breast cancer | >5 vs 1-3 hours/day | 1.01 (0.87 - 1.16) | ✓ | ✓ | ✓ | ✓ | ✓ | | | | ✓ | ✓ | ✓ | |
| | | | | | | Per 1 hour day | 1.01 (0.99 - 1.03) Ptrend:0.59 | ✓ | ✓ | ✓ | ✓ | ✓ | | | | | | ✓ | |
| | | 1107 | | | Incidence, Breast cancer, premenopausal | Per 1 hour day | 1.01 (0.97 - 1.06) | ✓ | ✓ | ✓ | ✓ | ✓ | | | | | | ✓ | |
| | | 3104 | | | Incidence, Breast cancer, postmenopausal | Per 1 hour day | 1.003 (0.98 - 1.03) | ✓ | ✓ | ✓ | ✓ | ✓ | | | | | | ✓ | |
| | | 1,092 | | | Incidence, Breast cancer, Obese | | 0.97 (0.94 - 1.01) | ✓ | ✓ | ✓ | ✓ | | | | | | | ✓ | |

| Author, Year Country (PMID) | Study name, type Age, sex, ethnicity, characteristics of participants | Cases/ Study size Median follow-up (years) | Case ascertainment | Exposure Assessment, detail | Outcome Sub-population | Comparison | RR (95%CI) Ptrend | Adjustments: Age | Sex | Smo | Alc | Obe | Phy | Diet | Med | Mens | Repr | Oth | Skin |
|---|---|---|---|---|---|---|---|---|---|---|---|---|---|---|---|---|---|---|---|
| | | 3,365 | | | Non obese | | 1.02 (0.99 - 1.04) | ✓ | ✓ | ✓ | ✓ | | | | | | | ✓ | |
| | | 1,259 | | | High physical activity | | 1.02 (0.97 - 1.06) | ✓ | ✓ | ✓ | ✓ | ✓ | | | | | | ✓ | |
| | | 1,570 | | | Moderate physical activity | | 1.00 (0.96 - 1.00) | ✓ | ✓ | ✓ | ✓ | ✓ | | | | | | ✓ | |
| | | 625 | | | low physical activity | | 0.99 (0.94 - 1.04) | ✓ | ✓ | ✓ | ✓ | ✓ | | | | | | ✓ | |
| | | 2,604 | | | never smokers | | 1.01 (0.98 - 1.04) | ✓ | ✓ | | ✓ | ✓ | | | | | | ✓ | |
| | | 302 | | | Former heavy smokers | | 1.04 (0.97 - 1.12) | ✓ | ✓ | | ✓ | ✓ | | | | | | ✓ | |
| | | 1,153 | | | Former light smokers | | 1.01 (0.97 - 1.05) | ✓ | ✓ | | ✓ | ✓ | | | | | | ✓ | |
| | | 181 | | | Current heavy smokers | | 0.97 (0.89 - 1.05) | ✓ | ✓ | | ✓ | ✓ | | | | | | ✓ | |
| | | 217 | | | Current light smokers | | 0.96 (0.88 - 1.05) | ✓ | ✓ | | ✓ | ✓ | | | | | | ✓ | |
| Cao 2019 Japan (30913861)[74] | JAAC<br>Prospective Cohort<br>Range:40-79 years<br>F<br>Premenopausal+ postmenopausal | 247 / 33,276<br>16.8 years | Cancer registry and hospital records | Self-administrated questionnaire, **Sitting while watching tv**: Hours of TV viewing | Incidence, Breast cancer | ≥4.5 vs <1.5 hours/day | 1.45 (0.91 - 2.32)<br>Ptrend:0.028 | ✓ | | ✓ | ✓ | ✓ | ✓ | | | ✓ | ✓ | ✓ | |
| | | 170 | | | premenopausal | | 1.34 (0.76 - 2.36)<br>Ptrend:0.493 | ✓ | | ✓ | ✓ | ✓ | ✓ | | | ✓ | ✓ | ✓ | |
| | | 77 | | | postmenopausal | | 2.37 (0.92 - 6.10)<br>Ptrend:0.009 | ✓ | | ✓ | ✓ | ✓ | ✓ | | | ✓ | ✓ | ✓ | |
| | | 129 | | | BMI >=23 | | 1.26 (0.65 - 2.44)<br>Ptrend:0.076 | ✓ | | ✓ | ✓ | ✓ | ✓ | | | ✓ | ✓ | ✓ | |
| | | 118 | | | BMI <23 | | 1.24 (0.57 - 2.72)<br>Ptrend:0.105 | ✓ | | ✓ | ✓ | ✓ | ✓ | | | ✓ | ✓ | ✓ | |
| Cifu 2018 USA (30309689)[69] | NIH- AARP Diet and Health Study<br>Prospective Cohort<br>Age:62 years<br>F<br>Mostly white | 7,088 / 106,126<br>12.5 years | self-report, cancer registry, death report | Self-reported questionnaire **Sitting while watching tv**: Daily television time, component of lifestyle adherence to ACS guidelines | Incidence, Breast cancer | <1 vs >7 hour/day | 1.02 (0.89 - 1.17)<br>Ptrend:0.879 | ✓ | | ✓ | ✓ | ✓ | ✓ | ✓ | | | ✓ | ✓ | |
| Nomura 2016 USA (27632430)[60] | Black Women's Health Study, 1995<br>Prospective Cohort<br>Range:21-69 years<br>F<br>African American | 1,984 / 46,734<br>15.4 years | self-report, linkage to cancer registries, medical and pathology records | Self-reported questionnaire, **Sitting while watching tv**: Watching TV, at baseline 1995 | Incidence, Breast cancer | ≥5 vs <1 hours/day | 1.04 (0.87 - 1.24)<br>Ptrend:0.53 | ✓ | | ✓ | | ✓ | ✓ | ✓ | ✓ | ✓ | ✓ | ✓ | |
| | | 752 | | | premenopausal | | 0.87 (0.65 - 1.16)<br>Ptrend:0.41 | ✓ | | ✓ | | ✓ | ✓ | ✓ | ✓ | ✓ | ✓ | ✓ | |
| | | 965 | | | postmenopausal | | 1.01 (0.79 - 1.30)<br>Ptrend:0.65 | ✓ | | ✓ | | ✓ | ✓ | ✓ | ✓ | ✓ | ✓ | ✓ | |
| | | 1,008 | | | ER+/PR+ | | 0.92 (0.71 - 1.19)<br>Ptrend:0.18 | ✓ | | ✓ | | ✓ | ✓ | ✓ | ✓ | ✓ | ✓ | ✓ | |
| | | 433 | | | ER- & PR- | | 1.40 (0.95 - 2.05)<br>Ptrend:0.09 | ✓ | | ✓ | | ✓ | ✓ | ✓ | ✓ | ✓ | ✓ | ✓ | |
| | | 173 | | | ER- & PR- & HER-2- | | 1.67 (0.92 - 3.01)<br>Ptrend:0.14 | ✓ | | ✓ | | ✓ | ✓ | ✓ | ✓ | ✓ | ✓ | ✓ | |

| Author, Year Country (PMID) | Study name, type Age, sex, ethnicity, characteristics of participants | Cases/ Study size Median follow-up (years) | Case ascertainment | Exposure Assessment, detail | Outcome Sub-population | Comparison | RR (95%CI) Ptrend | Adjustments | | | | | | | | | | | |
|---|---|---|---|---|---|---|---|---|---|---|---|---|---|---|---|---|---|---|---|
| | | | | | | | | Age | Sex | Smo | Alc | Obe | Phy | Diet | Med | Mens | Repr | Oth | Skin |
| Catsburg 2014 Canada (24781974)[55] | Canadian Study of Diet, Lifestyle and Health Case Cohort F Mostly white alumnae | 1,069 / 4,417 15 years | Cancer registry | Self-reported questionnaire, **Television watching**: Time spent in front of the television | Incidence, Invasive breast cancer | ≥21 vs ≤1 hours/week | 1.17 (0.86 - 1.59) Ptrend:0.62 | | | | ✓ | ✓ | | | | ✓ | ✓ | ✓ | |
| | | 543 | | | premenopausal | | 1.08 (0.65 - 1.79) Ptrend:0.74 | | | | ✓ | ✓ | | | | ✓ | ✓ | ✓ | |
| | | 526 | | | postmenopausal | | 1.20 (0.81 - 1.80) Ptrend:0.27 | | | | ✓ | ✓ | | | | ✓ | ✓ | ✓ | |
| Cohen 2013 USA (23576427)[46] | Southern Community Cohort Study Nested Case Control Age:55 years White, black, other | 459 / 2,730 9 years | Cancer registry | Self-reported questionnaire, **Sitting while watching tv**: Watching television or movies | Incidence, Invasive breast cancer | ≥5 vs 0 - 1.9 hours/day | 0.97 (0.70 - 1.35) Ptrend:0.31 | ✓ | | ✓ | | ✓ | ✓ | | | ✓ | ✓ | ✓ | |
| | | 132 | | | White | | 1.13 (0.59 - 2.19) Ptrend:0.45 | ✓ | | ✓ | | ✓ | ✓ | | | ✓ | ✓ | ✓ | |
| | | 314 | | | Black | | 0.95 (0.64 - 1.39) Ptrend:0.45 | ✓ | | ✓ | | ✓ | ✓ | | | ✓ | ✓ | ✓ | |
| Cifu 2018 USA (30309689)[69] | NIH- AARP Diet and Health Study Prospective Cohort Age:62 years F Mostly white | 462 / 106,126 12.5 years | self-report, cancer registry, death report | Self-administrated questionnaire, **Sitting while watching tv**: Daily television time, component of lifestyle adherence to ACS guidelines | Mortality, Breast cancer | < 1 vs > 7 hour/day | 0.85 (0.52 - 1.26) Ptrend:0.286 | ✓ | | ✓ | ✓ | ✓ | ✓ | ✓ | | | ✓ | ✓ | |
| Kim 2013 USA (24062293)[50] | Multiethnic Cohort Study (MEC) Prospective Cohort M/F Multi-ethnic | / 134,596 13.7 years | National Death Index | Self-administrated questionnaire, **Other sedentary activity**: Daily sitting watching TV (hours) | Mortality, Breast cancer | ≥5 vs <1 hour /day | 1.16 (0.73 - 1.84) Ptrend:0.36 | ✓ | | ✓ | ✓ | | ✓ | ✓ | | | | ✓ | |
| **Endometrial cancer** | | | | | | | | | | | | | | | | | | | |
| Miyata 2021 Japan (32963209)[79] | JACC Japan Prospective Cohort Range:40-79 years F | 79 / 33,801 14.8 years | Cancer registry/death certificates/medical records | Self-administrated questionnaire, **Sitting while watching tv**: | Incidence, Endometrial Cancer | ≥4 vs 1-<2 hours/day | 1.05 (0.51 - 2.15) Ptrend:0.635 | ✓ | | ✓ | ✓ | ✓ | ✓ | | | ✓ | ✓ | | |
| Hunter 2020 UK (32746843)[78] | UK Biobank Prospective Cohort Range:40-69 years F | 872 / 28,992 7.6 years | cancer registries | Self-reported questionnaire, **Television watching** | Incidence, Uterus corpus cancer | >5 vs 1-3 hours/day | 0.61 (0.42 - 0.88) | ✓ | ✓ | ✓ | ✓ | ✓ | | | | ✓ | ✓ | ✓ | |
| | | 872 | | | | Per 1 hour day | 0.97 (0.93 - 1.02) Ptrend:0.24 | ✓ | ✓ | ✓ | ✓ | ✓ | | | | ✓ | ✓ | ✓ | |
| | | 342 | | | obese | | 0.94 (0.88 - 1.00) | ✓ | ✓ | ✓ | ✓ | | | | | ✓ | ✓ | ✓ | |
| | | 406 | | | non obese | | 1.02 (0.95 - 1.09) | ✓ | ✓ | ✓ | ✓ | | | | | ✓ | ✓ | ✓ | |
| | | 197 | | | High physical activity | | 0.98 (0.89 - 1.09) | ✓ | ✓ | ✓ | ✓ | ✓ | | | | ✓ | ✓ | ✓ | |
| | | 258 | | | Moderate physical activity | | 1.00 (0.92 - 1.09) | ✓ | ✓ | ✓ | ✓ | ✓ | | | | ✓ | ✓ | ✓ | |
| | | 115 | | | Low physical activity | | 0.92 (0.82 - 1.03) | ✓ | ✓ | ✓ | ✓ | ✓ | | | | ✓ | ✓ | ✓ | |
| | | 495 | | | Never smokers | | 1.00 (0.95 - 1.06) Ptrend:0.24 | ✓ | ✓ | | ✓ | ✓ | | | | ✓ | ✓ | ✓ | |

| Author, Year Country (PMID) | Study name, type Age, sex, ethnicity, characteristics of participants | Cases/ Study size Median follow-up (years) | Case ascertainment | Exposure Assessment, detail | Outcome Sub-population | Comparison | RR (95%CI) Ptrend | Adjustments | | | | | | | | | | | |
|---|---|---|---|---|---|---|---|---|---|---|---|---|---|---|---|---|---|---|---|
| | | | | | | | | Age | Sex | Smo | Alc | Obe | Phy | Diet | Med | Mens | Repr | Oth | Skin |
| | | 51 | | | Former heavy smokers | | 0.88 (0.74 - 1.04) | ✓ | ✓ | | ✓ | ✓ | | | | ✓ | ✓ | ✓ | |
| | | 170 | | | Former light smokers | | 0.90 (0.81 - 1.01) | ✓ | ✓ | | ✓ | ✓ | | | | ✓ | ✓ | ✓ | |
| | | 15 | | | Current heavy smokers | | 0.89 (0.66 - 1.21) | ✓ | ✓ | | ✓ | ✓ | | | | ✓ | ✓ | ✓ | |
| | | 17 | | | Current light smokers | | 1.08 (0.81 - 1.44) | ✓ | ✓ | | ✓ | ✓ | | | | ✓ | ✓ | ✓ | |
| Gierach 2009 USA (19123463)[38] | NIH- AARP Diet and Health Study Prospective Cohort Range:50-71 years F Retired | 649 / 109,621 776170 person-years | linkage to eight state cancer registries | Self-reported questionnaire, **Television watching**: hours spending watching TV/video during typical 24-hour period in past 12 months | Incidence | 7+ vs <3 hours | 1.66 (1.20 - 2.28) Ptrend:0.0003 | ✓ | | ✓ | | | | | | ✓ | ✓ | ✓ | |
| | | 649 | | | | | 1.21 (0.87 - 1.67) Ptrend:0.26 | ✓ | | ✓ | | ✓ | | | | ✓ | ✓ | ✓ | |
| Friberg 2006 Sweden (17057024)[35] | Swedish Mammography Cohort (SMC) Prospective Cohort Range:50-83 years F | 199 / 33,723 7.25 years | Linkage with the National Swedish Cancer Register | Self-reported questionnaire, **Recreational activity**: leisure time inactivity (watching tv, sitting) | Incidence, Endometrial Cancer | high vs low | 1.80 (1.14 - 2.83) | ✓ | | | | | ✓ | ✓ | | | ✓ | ✓ | |
| | | 199 | | | | | 1.66 (1.05 - 2.61) | ✓ | | | | ✓ | ✓ | ✓ | | | ✓ | ✓ | |

#### Ovarian cancer

| Author, Year Country (PMID) | Study name, type Age, sex, ethnicity, characteristics of participants | Cases/ Study size Median follow-up (years) | Case ascertainment | Exposure Assessment, detail | Outcome Sub-population | Comparison | RR (95%CI) Ptrend | Age | Sex | Smo | Alc | Obe | Phy | Diet | Med | Mens | Repr | Oth | Skin |
|---|---|---|---|---|---|---|---|---|---|---|---|---|---|---|---|---|---|---|---|
| Buras 2022 USA (35094053)[84] | NHS and NHS II Prospective Cohort Age:47 years F nurses | 884 / 171,085 17.2 | self-report, cancer registry, death report | Self-reported questionnaire, **Television watching**: | Incidence, Ovarian cancer | >20 vs <5 hours/week | 0.96 (0.77 - 1.20) | ✓ | | | | ✓ | ✓ | | | ✓ | ✓ | ✓ | |
| | | 884 | | | average physical activity >= 15 MET/h-week | | 1.18 (0.84-1.64) Ptrend:0.38 | ✓ | | | | ✓ | ✓ | | | ✓ | ✓ | ✓ | |
| | | 884 | | | average physical activity < 15 MET/h-week | | 0.86 (0.62-1.19) Ptrend:0.14 | ✓ | | | | ✓ | ✓ | | | ✓ | ✓ | ✓ | |
| Hunter 2020 UK (32746843)[78] | UK Biobank Prospective Cohort Range:40-69 years F | 578 / 28,992 7.6 years | cancer registries | Self-reported questionnaire, **Television watching** | Incidence, Ovarian cancer | >5 vs 1-3 hours/day | 0.93 (0.63 - 1.38) | ✓ | ✓ | ✓ | ✓ | ✓ | | | | ✓ | ✓ | ✓ | |
| | | 578 | | | | Per 1 hour day | 1.02 (0.96 - 1.08) Ptrend:0.53 | ✓ | ✓ | ✓ | ✓ | ✓ | | | | ✓ | ✓ | ✓ | |
| | | 139 | | | Obese | | 0.96 (0.86 - 1.07) | ✓ | ✓ | ✓ | ✓ | | | | | ✓ | ✓ | ✓ | |
| | | 407 | | | Non obese | | 1.04 (0.97 - 1.11) | ✓ | ✓ | ✓ | ✓ | | | | | ✓ | ✓ | ✓ | |
| | | 170 | | | High physical activity | | 0.97 (0.87 - 1.08) | ✓ | ✓ | ✓ | ✓ | ✓ | | | | ✓ | ✓ | ✓ | |
| | | 171 | | | Moderate physical activity | | 1.05 (0.95 - 1.16) | ✓ | ✓ | ✓ | ✓ | ✓ | | | | ✓ | ✓ | ✓ | |
| | | 64 | | | Low physical activity | | 1.00 (0.86 - 1.17) | ✓ | ✓ | ✓ | ✓ | ✓ | | | | ✓ | ✓ | ✓ | |
| | | 316 | | | Never smokers | | 1.00 (0.93 - 1.08) Ptrend:0.53 | ✓ | ✓ | | ✓ | ✓ | | | | ✓ | ✓ | ✓ | |

| Author, Year Country (PMID) | Study name, type Age, sex, ethnicity, characteristics of participants | Cases/ Study size Median follow-up (years) | Case ascertainment | Exposure Assessment, detail | Outcome Sub-population | Comparison | RR (95%CI) Ptrend | Adjustments: Age | Sex | Smo | Alc | Obe | Phy | Diet | Med | Mens | Repr | Oth | Skin |
|---|---|---|---|---|---|---|---|---|---|---|---|---|---|---|---|---|---|---|---|
| | | 49 | | | Former heavy smokers | | 0.88 (0.73 - 1.05) | ✓ | ✓ | | ✓ | ✓ | | | | ✓ | ✓ | ✓ | |
| | | 138 | | | Former light smokers | | 1.06 (0.94 - 1.18) | ✓ | ✓ | | ✓ | ✓ | | | | ✓ | ✓ | ✓ | |
| | | 19 | | | Current heavy smokers | | 0.96 (0.75 - 1.23) | ✓ | ✓ | | ✓ | ✓ | | | | ✓ | ✓ | ✓ | |
| | | 24 | | | Current light smokers | | 1.23 (1.08 - 1.42) | ✓ | ✓ | | ✓ | ✓ | | | | ✓ | ✓ | ✓ | |
| Ukawa 2018 Japan (29340890)[64] | Japan Collaborative Cohort Study Prospective Cohort Range:40-79 years F | 59 / 34,758 19.4 years | hospital and cancer registry | Self-administrated questionnaire, **Television watching**: average TV viewing time to represent sedentary behaviour | Incidence, Ovarian cancer | ≥5 vs <2 hours/day | 1.81 (0.75 - 4.39) | ✓ | | ✓ | ✓ | ✓ | ✓ | | | ✓ | ✓ | ✓ | |
| Xiao 2013 USA (23966580)[49] | NIH- AARP Diet and Health Study Prospective Cohort Range:50-71 years F Retired | 463 / 153,123 12 years | cancer registry, death master file, national death index plus, postal service database | Self-reported questionnaire, **Television watching**: | Incidence, Epithelial ovarian cancer | 7+ vs <3 hours/day | 1.02 (0.67 - 1.55) | ✓ | | ✓ | | | | | | ✓ | ✓ | ✓ | |
| **Prostate cancer** | | | | | | | | | | | | | | | | | | | |
| Zhang 2024 UK (38974470)[93] | UK Biobank Prospective Cohort Age:55.94 years M Mostly white | 8,644 / 360,271 12.52 years | Cancer registry | Self-reported questionnaire, **Screen time, recreational** | Incidence, prostate cancer | high vs low | 1.01 (0.95 - 1.07) | ✓ | ✓ | ✓ | ✓ | ✓ | ✓ | | | | | ✓ | |
| Cuthbertson 2020 USA (32978174)[80] | ARIC Study Prospective Cohort Age:68.6 years M Multi-ethnic | 813 / 14,508 23.6 years | cancer registry and medical records | Self-reported questionnaire, **TV watching frequency** | Incidence, prostate cancer | seldom/never vs often/very often | 1.20 (0.99 - 1.47) | ✓ | | ✓ | ✓ | | | | | | | ✓ | |
| Hunter 2020 UK (32746843)[78] | UK Biobank Prospective Cohort Range:40-69 years M | 5,979 / 28,992 7.6 years | cancer registries | Self-reported questionnaire, **Television watching** | Incidence, prostate cancer | >5 vs 1-3 hours/day | 0.95 (0.84 - 1.07) | ✓ | ✓ | ✓ | ✓ | ✓ | | | | | | ✓ | |
| | | 5,979 | | | | Per 1 hour day | 0.99 (0.97 - 1.01) | ✓ | ✓ | ✓ | ✓ | ✓ | | | | | | ✓ | |
| | | 1,290 | | | Obese | | 0.99 (0.96 - 1.03) | ✓ | ✓ | ✓ | ✓ | | | | | | | ✓ | |
| | | 4,276 | | | Non obese | | 0.99 (0.97 - 1.01) | ✓ | ✓ | ✓ | ✓ | | | | | | | ✓ | |
| | | 1,998 | | | High physical activity | | 0.99 (0.96 - 1.02) | ✓ | ✓ | ✓ | ✓ | ✓ | | | | | | ✓ | |
| | | 1,890 | | | Moderate physical activity | | 0.98 (0.95 - 1.01) | ✓ | ✓ | ✓ | ✓ | ✓ | | | | | | ✓ | |
| | | 741 | | | Low physical activity | | 1.01 (0.97 - 1.06) | ✓ | ✓ | ✓ | ✓ | ✓ | | | | | | ✓ | |
| | | 2,659 | | | Never smokers | | 0.99 (0.96 - 1.01) | ✓ | ✓ | | ✓ | ✓ | | | | | | ✓ | |
| | | 710 | | | Former heavy smokers | | 1.01 (0.97 - 1.06) | ✓ | ✓ | | ✓ | ✓ | | | | | | ✓ | |
| | | 1,697 | | | Former light smokers | | 0.99 (0.96 - 1.03) | ✓ | ✓ | | ✓ | ✓ | | | | | | ✓ | |
| | | 222 | | | Current heavy smokers | | 0.88 (0.82 - 0.95) | ✓ | ✓ | | ✓ | ✓ | | | | | | ✓ | |

| Author, Year Country (PMID) | Study name, type Age, sex, ethnicity, characteristics of participants | Cases/ Study size Median follow-up (years) | Case ascertainment | Exposure Assessment, detail | Outcome Sub-population | Comparison | RR (95%CI) Ptrend | Adjustments | | | | | | | | | | | |
|---|---|---|---|---|---|---|---|---|---|---|---|---|---|---|---|---|---|---|---|
| | | | | | | | | Age | Sex | Smo | Alc | Obe | Phy | Diet | Med | Mens | Repr | Oth | Skin |
| | | 276 | | | Current light smokers | | 1.01 (0.95 - 1.09) | ✓ | ✓ | | ✓ | ✓ | | | | | | ✓ | |
| Lynch 2014 USA (24526287)[53] | NIH- AARP Diet and Health Study Prospective Cohort Range:50-71 years M White, black, other | 13,751 / 170,481 8.5 years | Cancer registry and national death index | Self-administered Questionnaire, **Sitting while watching tv**: by BMI category | Incidence, prostate cancer | ≥7 vs <1 hours/day | 1.03 (0.92 - 1.15) Ptrend:0.53 | ✓ | | ✓ | ✓ | ✓ | ✓ | ✓ | ✓ | | | ✓ | |
| | | 1,365 | | | Advanced prostate cancer | ≥5 vs <3 hours/day | 0.93 (0.79 - 1.09) Ptrend:0.49 | ✓ | | ✓ | ✓ | ✓ | ✓ | ✓ | ✓ | | | ✓ | |
| | | 2,366 | | | prostate cancer BMI >=30 kg/m^2 | ≥7 vs <1 hours/day | 1.28 (0.98 - 1.69) Ptrend:0.11 | ✓ | | ✓ | ✓ | ✓ | ✓ | ✓ | ✓ | | | ✓ | |
| | | NA | | | BMI 25.0 to 29.9 kg/m^2 | | 1.04 (0.89 - 1.22) Ptrend:0.98 | ✓ | | ✓ | ✓ | ✓ | ✓ | ✓ | ✓ | | | ✓ | |
| | | 4,435 | | | BMI 18.5-24.9kg/m2 | | 0.82 (0.66 - 1.01) Ptrend:0.04 | ✓ | | ✓ | ✓ | ✓ | ✓ | ✓ | ✓ | | | ✓ | |
| | | 262 | | | advanced prostate cancer, BMI >=30 kg/m^2 | ≥5 vs <3 hours/day | 1.22 (0.86 - 1.73) Ptrend:0.22 | ✓ | | ✓ | ✓ | ✓ | ✓ | ✓ | ✓ | | | ✓ | |
| | | 673 | | | BMI 25.0 to 29.9 kg/m^2 | | 0.88 (0.70 - 1.11) Ptrend:0.27 | ✓ | | ✓ | ✓ | ✓ | ✓ | ✓ | ✓ | | | ✓ | |
| | | 420 | | | BMI 18.5-24.9kg/m2 | | 0.86 (0.63 - 1.17) Ptrend:0.44 | ✓ | | ✓ | ✓ | ✓ | ✓ | ✓ | ✓ | | | ✓ | |
| | | 669 | | | Mortality, Prostate cancer | | 1.07 (0.85 - 1.33) Ptrend:0.15 | ✓ | | ✓ | ✓ | ✓ | ✓ | ✓ | ✓ | | | ✓ | |
| | | 151 | | | BMI >=30 kg/m^2 | | 1.52 (0.97 - 2.40) Ptrend:0.03 | ✓ | | ✓ | ✓ | ✓ | ✓ | ✓ | ✓ | | | ✓ | |
| | | 335 | | | BMI 25.0 to 29.9 kg/m^2 | | 0.99 (0.72 - 1.38) Ptrend:0.59 | ✓ | | ✓ | ✓ | ✓ | ✓ | ✓ | ✓ | | | ✓ | |
| | | 181 | | | BMI 18.5-24.9kg/m2 | | 0.83 (0.53 - 1.30) Ptrend:0.96 | ✓ | | ✓ | ✓ | ✓ | ✓ | ✓ | ✓ | | | ✓ | |
| Kim 2013 USA (24062293)[50] | Multiethnic Cohort Study (MEC) Prospective Cohort M Multi-ethnic | / 134,596 13.7 years | National Death Index | Self-administrated questionnaire, **Other sedentary activity**: Daily sitting watching TV (hours) | Mortality, prostate cancer | ≥5 vs <1 hour/day | 1.39 (0.90 - 2.15) Ptrend:0.01 | ✓ | | ✓ | ✓ | | ✓ | ✓ | | | | ✓ | |
| **Kidney cancer** | | | | | | | | | | | | | | | | | | | |
| George 2011 USA (21737302)[41] | National Institute of Health (NIH)-AARP(formerly the American Association for Retired Persons) Diet and Health Study Prospective Cohort Age:63 years M/F | 1,206 / 289,512 10 years | Cancer registry | Self-administrated questionnaire, **Sitting while watching tv** | Incidence, renal cell carcinoma, Men and women | ≥7 vs <1 hours/day | 0.96 (0.66 - 1.38) Ptrend:0.707 | ✓ | | ✓ | ✓ | | ✓ | ✓ | | | | ✓ | |
| | | 909 | | | Men | | 0.97 (0.61 - 1.53) Ptrend:0.743 | ✓ | | ✓ | ✓ | | ✓ | ✓ | | | | ✓ | |
| | | 297 | | | Women | | 0.96 (0.51 - 1.82) Ptrend:0.700 | ✓ | | ✓ | ✓ | | ✓ | ✓ | | | ✓ | ✓ | |
| Hunter 2020 UK (32746843)[78] | UK Biobank Prospective Cohort Range:40-69 years M/F | 779 / 28,992 7.6 years | cancer registries | Self-reported questionnaire, **Television watching** | Incidence, kidney cancer | >5 vs 1-3 hours/day | 1.08 (0.82 - 1.42) | ✓ | ✓ | ✓ | ✓ | ✓ | | | | | | ✓ | |

| Author, Year Country (PMID) | Study name, type Age, sex, ethnicity, characteristics of participants | Cases/ Study size Median follow-up (years) | Case ascertainment | Exposure Assessment, detail | Outcome Sub-population | Comparison | RR (95%CI) Ptrend | Adjustments | | | | | | | | | | | |
|---|---|---|---|---|---|---|---|---|---|---|---|---|---|---|---|---|---|---|---|
| | | | | | | | | Age | Sex | Smo | Alc | Obe | Phy | Diet | Med | Mens | Repr | Oth | Skin |
| | | 779 | | | | Per 1 hour day | 1.00 (0.95 - 1.04) Ptrend:0.86 | ✓ | ✓ | ✓ | ✓ | ✓ | | | | | | ✓ | |
| | | 273 | | | Obese | | 0.98 (0.92 - 1.06) | ✓ | ✓ | ✓ | ✓ | | | | | | | ✓ | |
| | | 465 | | | Non obese | | 1.00 (0.95 - 1.07) | ✓ | ✓ | ✓ | ✓ | | | | | | | ✓ | |
| | | 207 | | | High physical activity | | 1.04 (0.95 - 1.14) | ✓ | ✓ | ✓ | ✓ | ✓ | | | | | | ✓ | |
| | | 248 | | | Moderate physical activity | | 1.01 (0.93 - 1.10) | ✓ | ✓ | ✓ | ✓ | ✓ | | | | | | ✓ | |
| | | 104 | | | Low physical activity | | 0.89 (0.79 - 1.01) | ✓ | ✓ | ✓ | ✓ | ✓ | | | | | | ✓ | |
| | | 315 | | | Never smokers | | 1.01 (0.94 - 1.08) | ✓ | ✓ | | ✓ | ✓ | | | | | | ✓ | |
| | | 115 | | | Former heavy smokers | | 0.97 (0.87 - 1.08) | ✓ | ✓ | | ✓ | ✓ | | | | | | ✓ | |
| | | 206 | | | Former light smokers | | 0.98 (0.89 - 1.07) | ✓ | ✓ | | ✓ | ✓ | | | | | | ✓ | |
| | | 60 | | | Current heavy smokers | | 1.00 (0.88 - 1.14) | ✓ | ✓ | | ✓ | ✓ | | | | | | ✓ | |
| | | 42 | | | Current light smokers | | 1.05 (0.89 - 1.23) | ✓ | ✓ | | ✓ | ✓ | | | | | | ✓ | |
| **Bladder cancer** | | | | | | | | | | | | | | | | | | | |
| Hunter 2020 UK (32746843)[78] | UK Biobank Prospective Cohort Range:40-69 years M/F | 677 / 28,992 7.6 years | cancer registries | Self-reported questionnaire, **Television watching** | Incidence, bladder cancer | >5 vs 1-3 hours/day | 1.29 (0.97 - 1.73) | ✓ | ✓ | ✓ | ✓ | ✓ | | | | | | ✓ | |
| | | 677 | | | | Per 1 hour day | 1.04 (0.99 - 1.09) Ptrend:0.13 | ✓ | ✓ | ✓ | ✓ | ✓ | | | | | | ✓ | |
| | | 204 | | | Obese | | 1.05 (0.97 - 1.13) | ✓ | ✓ | ✓ | ✓ | | | | | | | ✓ | |
| | | 424 | | | Non obese | | 1.03 (0.97 - 1.09) | ✓ | ✓ | ✓ | ✓ | | | | | | | ✓ | |
| | | 213 | | | High physical activity | | 1.01 (0.93 - 1.11) | ✓ | ✓ | ✓ | ✓ | ✓ | | | | | | ✓ | |
| | | 207 | | | Moderate physical activity | | 1.05 (0.97 - 1.14) | ✓ | ✓ | ✓ | ✓ | ✓ | | | | | | ✓ | |
| | | 82 | | | Low physical activity | | 1.08 (0.98 - 1.20) | ✓ | ✓ | ✓ | ✓ | ✓ | | | | | | ✓ | |
| | | 187 | | | Never smokers | | 1.07 (0.98 - 1.18) | ✓ | ✓ | | ✓ | ✓ | | | | | | ✓ | |
| | | 141 | | | Former heavy smokers | | 1.01 (0.92 - 1.12) | ✓ | ✓ | | ✓ | ✓ | | | | | | ✓ | |
| | | 182 | | | Former light smokers | | 1.08 (0.99 - 1.18) | ✓ | ✓ | | ✓ | ✓ | | | | | | ✓ | |
| | | 68 | | | Current heavy smokers | | 0.99 (0.88 - 1.11) | ✓ | ✓ | | ✓ | ✓ | | | | | | ✓ | |
| | | 50 | | | Current light smokers | | 0.99 (0.84 - 1.17) | ✓ | ✓ | | ✓ | ✓ | | | | | | ✓ | |
| Koebnick 2008 USA (18483344)[37] | NIH- AARP Diet and Health Study Prospective Cohort Range:50-71 years M/F Mixed | 1719 / 471,760 3404642 person-years | linkages with national cancer and death registries | Self-reported questionnaire, **Television watching**: Watching television or videos<br><br>not adjusted for BMI | Incidence, bladder cancer | ≥9 vs <3 hours/day | 0.99 (0.66 - 1.51) | ✓ | ✓ | ✓ | ✓ | | | ✓ | | | ✓ | ✓ | |

| Author, Year Country (PMID) | Study name, type Age, sex, ethnicity, characteristics of participants | Cases/ Study size Median follow-up (years) | Case ascertainment | Exposure Assessment, detail | Outcome Sub-population | Comparison | RR (95%CI) Ptrend | Adjustments | | | | | | | | | | | |
|---|---|---|---|---|---|---|---|---|---|---|---|---|---|---|---|---|---|---|---|
| | | | | | | | | Age | Sex | Smo | Alc | Obe | Phy | Diet | Med | Mens | Repr | Oth | Skin |
| | | NA | | Adjusted for BMI as well. | | | 0.98 (0.65 - 1.49) | ✓ | ✓ | ✓ | ✓ | ✓ | | ✓ | | | ✓ | ✓ | |
| **Brain cancer** | | | | | | | | | | | | | | | | | | | |
| Hunter 2020 UK (32746843)[78] | UK Biobank Prospective Cohort Range:40-69 years M/F | 463 / 28,992 7.6 years | cancer registries | Self-reported questionnaire **Television watching**: | Incidence, Brain cancer | >5 vs 1-3 hours/day | 0.96 (0.63 - 1.46) | ✓ | ✓ | ✓ | ✓ | ✓ | | | | | | ✓ | |
| | | 463 | | | | Per 1 hour day | 1.04 (0.98 - 1.10) Ptrend:0.20 | ✓ | ✓ | ✓ | ✓ | ✓ | | | | | | ✓ | |
| | | 110 | | | Obese | | 1.03 (0.92 - 1.14) | ✓ | ✓ | ✓ | ✓ | | | | | | | ✓ | |
| | | 324 | | | Non obese | | 1.05 (0.98 - 1.13) | ✓ | ✓ | ✓ | ✓ | | | | | | | ✓ | |
| | | 137 | | | High physical activity | | 1.01 (0.90 - 1.14) | ✓ | ✓ | ✓ | ✓ | ✓ | | | | | | ✓ | |
| | | 145 | | | Moderate physical activity | | 0.94 (0.84 - 1.06) | ✓ | ✓ | ✓ | ✓ | ✓ | | | | | | ✓ | |
| | | 63 | | | Low physical activity | | 1.14 (1.03 - 1.26) | ✓ | ✓ | ✓ | ✓ | ✓ | | | | | | ✓ | |
| | | 212 | | | Never smokers | | 1.00 (0.91 - 1.09) | ✓ | ✓ | | ✓ | ✓ | | | | | | ✓ | |
| | | 38 | | | Former heavy smokers | | 1.18 (1.02 - 1.36) | ✓ | ✓ | | ✓ | ✓ | | | | | | ✓ | |
| | | 141 | | | Former light smokers | | 1.04 (0.93 - 1.16) | ✓ | ✓ | | ✓ | ✓ | | | | | | ✓ | |
| | | 23 | | | Current heavy smokers | | 0.97 (0.80 - 1.19) | ✓ | ✓ | | ✓ | ✓ | | | | | | ✓ | |
| | | 20 | | | Current light smokers | | 1.13 (0.94 - 1.35) | ✓ | ✓ | | ✓ | ✓ | | | | | | ✓ | |
| **Thyroid cancer** | | | | | | | | | | | | | | | | | | | |
| Hunter 2020 UK (32746843)[78] | UK Biobank Prospective Cohort Range:40-69 years M/F | 242 / 28,992 7.6 years | cancer registries | Self-reported questionnaire **Television watching**: | Incidence, Thyroid cancer | >5 vs 1-3 hours/day | 1.14 (0.63 - 2.06) | ✓ | ✓ | ✓ | ✓ | ✓ | | | | | | ✓ | |
| | | 242 | | | | Per 1 hour day | 1.00 (0.92 - 1.03) Ptrend:0.89 | ✓ | ✓ | ✓ | ✓ | ✓ | | | | | | ✓ | |
| | | 68 | | | Obese | | 0.95 (0.81 - 1.11) | ✓ | ✓ | ✓ | ✓ | | | | | | | ✓ | |
| | | 157 | | | Non obese | | 1.03 (0.93 - 1.15) | ✓ | ✓ | ✓ | ✓ | | | | | | | ✓ | |
| | | 74 | | | High physical activity | | 1.12 (0.97 - 1.29) | ✓ | ✓ | ✓ | ✓ | | | | | | | ✓ | |
| | | 73 | | | Moderate physical activity | | 0.92 (0.78 - 1.10) | ✓ | ✓ | ✓ | ✓ | | | | | | | ✓ | |
| | | 34 | | | Low physical activity | | 1.05 (0.87 - 1.26) | ✓ | ✓ | ✓ | ✓ | | | | | | | ✓ | |
| | | 130 | | | Never smokers | | 1.02 (0.90 - 1.14) | ✓ | ✓ | | ✓ | ✓ | | | | | | ✓ | |
| | | 20 | | | Former heavy smokers | | 1.06 (0.82 - 1.38) | ✓ | ✓ | | ✓ | ✓ | | | | | | ✓ | |
| | | 57 | | | Former light smokers | | 0.91 (0.75 - 1.10) | ✓ | ✓ | | ✓ | ✓ | | | | | | ✓ | |
| | | 7 | | | Current heavy smokers | | 1.04 (0.72 - 1.53) | ✓ | ✓ | | ✓ | ✓ | | | | | | ✓ | |

| Author, Year Country (PMID) | Study name, type Age, sex, ethnicity, characteristics of participants | Cases/ Study size Median follow-up (years) | Case ascertainment | Exposure Assessment, detail | Outcome Sub-population | Comparison | RR (95%CI) Ptrend | Adjustments | | | | | | | | | | | |
|---|---|---|---|---|---|---|---|---|---|---|---|---|---|---|---|---|---|---|---|
| | | | | | | | | Age | Sex | Smo | Alc | Obe | Phy | Diet | Med | Mens | Repr | Oth | Skin |
| | | 11 | | | Current light smokers | | 1.04 (0.77 - 1.41) | ✓ | ✓ | | ✓ | ✓ | | | | | | ✓ | |
| **Haematological cancer** | | | | | | | | | | | | | | | | | | | |
| Hunter 2020 UK (32746843)[78] | UK Biobank Prospective Cohort Range:40-69 years M/F | 1,193 / 28,992 7.6 years | cancer registries | Self-reported questionnaire **Television watching**: | Incidence, Non-Hodgkin's lymphoma (NHL) | >5 vs 1-3 hours/day | 0.85 (0.65 - 1.11) | ✓ | ✓ | ✓ | ✓ | ✓ | | | | | | ✓ | |
| | | 1,193 | | | | Per 1 hour day | 1.01 (0.98 - 1.05) Ptrend:0.48 | ✓ | ✓ | ✓ | ✓ | ✓ | | | | | | ✓ | |
| | | 289 | | | Obese | | 1.00 (0.94 - 1.07) | ✓ | ✓ | ✓ | ✓ | | | | | | | ✓ | |
| | | 841 | | | Non obese | | 1.02 (0.97 - 1.06) | ✓ | ✓ | ✓ | ✓ | | | | | | | ✓ | |
| | | 362 | | | High physical activity | | 1.05 (0.98 - 1.12) | ✓ | ✓ | ✓ | ✓ | ✓ | | | | | | ✓ | |
| | | 350 | | | Moderate physical activity | | 1.02 (0.95 - 1.09) | ✓ | ✓ | ✓ | ✓ | ✓ | | | | | | ✓ | |
| | | 152 | | | Low physical activity | | 0.97 (0.89 - 1.07) | ✓ | ✓ | ✓ | ✓ | ✓ | | | | | | ✓ | |
| | | 584 | | | Never smokers | | 0.99 (0.94 - 1.05) | ✓ | ✓ | | ✓ | ✓ | | | | | | ✓ | |
| | | 134 | | | Former heavy smokers | | 0.99 (0.89 - 1.09) | ✓ | ✓ | | ✓ | ✓ | | | | | | ✓ | |
| | | 290 | | | Former light smokers | | 1.12 (1.04 - 1.05) | ✓ | ✓ | | ✓ | ✓ | | | | | | ✓ | |
| | | 70 | | | Current heavy smokers | | 0.91 (0.81 - 1.03) | ✓ | ✓ | | ✓ | ✓ | | | | | | ✓ | |
| | | 52 | | | Current light smokers | | 0.98 (0.83 - 1.15) | ✓ | ✓ | | ✓ | ✓ | | | | | | ✓ | |

**Adjustment groups: Age**: age; **Alc**: alcohol; **Diet**: dietary factors and energy intake; **Med**: medical factors (e.g. cancer-related infections, diseases in the breast, digestive tract, gallbladder, liver, or pancreas, cancer screening, precancerous conditions, or pre-existing skin cancer); **Mens**: menstrual characteristics (e.g. age at menarche, menopausal status/duration, age at menopause, or type of menopause); **Obe**: obesity (adiposity measures); **Oth**: others (e.g. family history/genetic factor, race/ethnicity, ancestry, socioeconomic factors, or marriage); **Phy**: physical activity; **Repr**: reproductive or hormonal factors (e.g. parity, age at first/last birth, postmenopausal hormone use, or oral contraceptive use); **Sex**: sex; **Skin**: skin-related factors (e.g. UV light/sun exposure, sunburns, time spent outdoors, sunscreen use, indoor tanning, moles, fair skin, red/blond hair, blue/green eyes, or freckles); **Smo**: smoking, cancer-related

#### Supplementary Table 11: List of associations between sedentary behaviour and cancer incidence or mortality outcomes with only one available study (for specific definitions of sedentary behaviour).

| Author Year | Cancer site | Outcome type* | Study name | Sex | Cases / cohort size | RR (95%CI) | p-value for trend/ linear analysis | Comparison |
|---|---|---|---|---|---|---|---|---|
| **Sedentary time, total** | | | | | | | | |
| Tomida 2024 (38041484)[91] | Breast | Inc | J-MICC | F | 554 / 36023 | 1.34 (1.02 - 1.75) | 0.046 | >13 vs <7 hours/day |
| Tomida 2024 (38041484)[91] | Breast, postmenopausal | Inc | J-MICC | F | 276 / 36023 | 1.22 (0.87 - 1.70) | 0.251 | >13 vs <7 hours/day |
| Tomida 2024 (38041484)[91] | Breast, postmenopausal | Inc | J-MICC | F | 276 / 36023 | 1.01 (0.98 - 1.04) | 0.08 | Per 1 hours/day |
| Tomida 2024 (38041484)[91] | Breast, premenopausal | Inc | J-MICC | F | 174 / 36023 | 1.64 (1.03 - 2.60) | 0.037 | >13 vs <7 hours/day |
| Tomida 2024 (38041484)[91] | Breast, premenopausal | Inc | J-MICC | F | 174 / 36023 | 1.03 (1.00 - 1.08) | 0.08 | Per 1 hours/day |
| Watts 2023 (36795414)[88] | Skin, non-melanoma | Inc | UK Biobank | M/F | 1734 / 81717 | 0.99 (0.97 - 1.00) | N/A | Per 20 min/day |
| **Sedentary time, non-occupational** | | | | | | | | |
| Zeng 2023 (38066552)[92] | Pancreas | Inc | UK Biobank | M | N/A / 340631 | 0.70 (0.53 - 0.92) | N/A | <3 vs ≥5 hours/day |
| Zeng 2023 (38066552)[92] | Pancreas | Inc | UK Biobank | F | N/A / 340631 | 0.78 (0.60 - 1.02) | N/A | <3 vs ≥5 hours/day |
| Weber 2021 (34059803)[83] | Melanoma | Inc | UK Biobank | M/F | 1239 / 350512 | 1.04 (0.97 - 1.12) | 0.306 | Per 3 hours/day |
| **Sitting time, total** | | | | | | | | |
| Cook 2013 (24367697)[52] | Oesophageal, adenocarcinoma | Inc | NIH-AARP | M/F | 377 / 493802 | 0.69 (0.41 - 1.15) | 0.84 | ≥9 vs ≤3 hours/day |
| Cook 2013 (24367697)[52] | Oesophageal, SCC | Inc | NIH-AARP | M/F | 128 / 493802 | 0.86 (0.39 - 1.88) | 0.82 | ≥9 vs ≤3 hours/day |
| Cook 2013 (24367697)[52] | Gastric, cardia | Inc | NIH-AARP | M/F | 255 / 303033 | 1.00 (0.62 - 1.63) | 0.63 | ≥9 vs ≤3 hours/day |
| Cook 2013 (24367697)[52] | Gastric, non-cardia | Inc | NIH-AARP | M/F | 277 / 303033 | 0.81 (0.45 - 1.45) | 0.49 | ≥9 vs ≤3 hours/day |
| Wang 2016 (27439221)[59] | Lung | Mort | WHI | F | 814 / 129401 | 1.16 (0.96 - 1.40) | N/A | ≥10 vs ≤5 hours/day |
| Rangul 2018 Norway (30352079)[70] | Lung | Inc | HUNT | M | 242 / 37810 | 1.14 (0.86 - 1.54) | N/A | >8 vs 0-4 hours/day |
| Rangul 2018 Norway (30352079)[70] | Lung | Inc | HUNT | F | 149 / 37810 | 0.85 (0.55 - 1.31) | N/A | >8 vs 0-4 hours/day |
| Jiang 2019 (30859092)[72] | Lung, non-small cell | Inc | HUNT | M/F | 333 / 45810 | 0.97 (0.74 - 1.26) | N/A | >8 vs 0-4 hours/day |
| Jiang 2019 (30859092)[72] | Lung, small cell | Inc | HUNT | M/F | 76 / 45810 | 1.18 (0.69 - 2.03) | N/A | >8 vs 0-4 hours/day |
| Moore, 2010 (20877336)[40] | Endometrial | Inc | NIH-AARP | F | 888 / 109621 | 1.45 (1.10 - 1.92) | <0.0001 | ≥9 vs ≤2.99 hours/day |
| Xiao 2013 (23966580)[49] | Ovary, epithelial | Inc | NIH-AARP | F | 463 / 153123 | 1.06 (0.81 - 1.39) | N/A | ≥7 vs ≤2.9 hours/day |
| Lynch 2014b (24526287)[53] | Prostate, advanced | Inc | NIH-AARP | M | 1365 / 170481 | 0.91 (0.77 - 1.08) | 0.16 | ≥7 vs <3 hours/day |
| Lynch 2014b (24526287)[53] | Prostate | Mort | NIH-AARP | M | 669 / 170481 | 1.07 (0.84 - 1.35) | 0.98 | ≥7 vs <3 hours/day |
| George 2011 (21737302)[41] | Kidney | Inc | NIH-AARP | M/F | 1206 / 289512 | 1.11 (0.87 - 1.41) | 0.765 | ≥9 vs ≤2.9 hours/day |
| Rees-Punia 2019 (31196856)[75] | Acute myeloid leukaemia | Inc / Mort | CPS II | M/F | 155 / 109030 | 1.01 (0.60 - 1.64) | N/A | >6 vs <3 hours/day |
| Rees-Punia 2019 (31196856)[75] | Myeloid leukaemia | Inc / Mort | CPS II | M/F | 255 / 109030 | 1.28 (0.89 - 1.85) | N/A | >6 vs <3 hours/day |
| **Sitting time, recreational** | | | | | | | | |
| Patel 2015 (26126627)[56] | Mouth | Inc / Mort | CPS II | M | 274 / 146722 | 1.22 (0.88 - 1.69) | N/A | ≥6 vs ≤2 hours/day |
| Patel 2015 (26126627)[56] | Mouth | Inc / Mort | CPS II | F | 97 / 146722 | 1.46 (0.84 - 2.54) | N/A | ≥6 vs ≤2 hours/day |
| Patel 2015 (26126627)[56] | Oesophageal | Inc / Mort | CPS II | M | 270 / 146722 | 1.05 (0.75 - 1.48) | N/A | ≥6 vs ≤2 hours/day |
| Patel 2015 (26126627)[56] | Oesophageal | Inc / Mort | CPS II | F | 45 / 146722 | 1.14 (0.48 - 2.74) | N/A | ≥6 vs ≤2 hours/day |

| Author Year | Cancer site | Outcome type* | Study name | Sex | Cases / cohort size | RR (95%CI) | p-value for trend/ linear analysis | Comparison |
|---|---|---|---|---|---|---|---|---|
| Patel 2015 (26126627)[56] | Stomach | Inc / Mort | CPS II | M | 226 / 146722 | 1.04 (0.70 - 1.55) | N/A | ≥6 vs ≤2 hours/day |
| Patel 2015 (26126627)[56] | Stomach | Inc / Mort | CPS II | F | 80 / 146722 | 1.02 (0.54 - 1.96) | N/A | ≥6 vs ≤2 hours/day |
| Patel 2015 (26126627)[56] | Colorectal | Inc / Mort | CPS II | M | 1447 / 146722 | 1.03 (0.89 - 1.19) | N/A | ≥6 vs ≤2 hours/day |
| Patel 2015 (26126627)[56] | Colorectal | Inc / Mort | CPS II | F | 1199 / 146722 | 0.97 (0.81 - 1.16) | N/A | ≥6 vs ≤2 hours/day |
| Patel 2015 (26126627)[56] | Liver | Inc / Mort | CPS II | M | 171 / 146722 | 0.84 (0.55 - 1.29) | N/A | ≥6 vs ≤2 hours/day |
| Patel 2015 (26126627)[56] | Liver | Inc / Mort | CPS II | F | 79 / 146722 | 0.77 (0.37 - 1.60) | N/A | ≥6 vs ≤2 hours/day |
| Patel 2015 (26126627)[56] | Gallbladder | Inc / Mort | CPS II | M | 33 / 146722 | 2.14 (0.89 - 5.16) | N/A | ≥6 vs ≤2 hours/day |
| Patel 2015 (26126627)[56] | Gallbladder | Inc / Mort | CPS II | F | 57 / 146722 | 1.52 (0.70 - 3.32) | N/A | ≥6 vs ≤2 hours/day |
| Patel 2015 (26126627)[56] | Pancreas | Inc / Mort | CPS II | M | 425 / 146722 | 1.17 (0.89 - 1.53) | N/A | ≥6 vs ≤2 hours/day |
| Patel 2015 (26126627)[56] | Pancreas | Inc / Mort | CPS II | F | 350 / 146722 | 1.04 (0.75 - 1.44) | N/A | ≥6 vs ≤2 hours/day |
| Patel 2015 (26126627)[56] | Lung | Inc / Mort | CPS II | M | 1903 / 146722 | 1.01 (0.89 - 1.15) | N/A | ≥6 vs ≤2 hours/day |
| Patel 2015 (26126627)[56] | Lung | Inc / Mort | CPS II | F | 1118 / 146722 | 0.96 (0.80 - 1.14) | N/A | ≥6 vs ≤2 hours/day |
| Patel 2015 (26126627)[56] | Melanoma | Inc / Mort | CPS II | M | 1154 / 146722 | 1.05 (0.89 - 1.24) | N/A | ≥6 vs ≤2 hours/day |
| Patel 2015 (26126627)[56] | Melanoma | Inc / Mort | CPS II | F | 769 / 146722 | 0.99 (0.78 - 1.24) | N/A | ≥6 vs ≤2 hours/day |
| Hildebrand 2013 (24097200)[51] | Breast, postmenopausal | Inc / Mort | CPS II | F | 4681 / 73615 | 1.05 (0.96 - 1.16) | N/A | ≥6 vs ≤2.9 hours/day |
| Patel 2015 (26126627)[56] | Endometrial | Inc / Mort | CPS II | F | 387 / 77462 | 1.34 (1.08 - 1.67) | N/A | ≥6 vs ≤2 hours/day |
| Hildebrand 2015 (26335264)[57] | Ovary | Inc / Mort | CPS II | F | 638 / 63972 | 1.44 (1.12 - 1.85) | 0.006 | ≥6 vs ≤2.9 hours/day |
| Hildebrand 2015 (26335264)[57] | Ovary, nonserous | Inc / Mort | CPS II | F | 112 / 63972 | 1.08 (0.57 - 2.04) | 0.83 | ≥6 vs ≤2.9 hours/day |
| Hildebrand 2015 (26335264)[57] | Ovary, serous | Inc / Mort | CPS II | F | 313 / 63972 | 1.52 (1.06 - 2.16) | 0.01 | ≥6 vs ≤2.9 hours/day |
| Patel 2015 (26126627)[56] | Prostate | Inc / Mort | CPS II | M | 8276 / 69260 | 0.96 (0.90 - 1.02) | N/A | ≥6 vs ≤2 hours/day |
| Patel 2015 (26126627)[56] | Prostate, advanced | Inc / Mort | CPS II | M | 1705 / 69260 | 0.96 (0.85 - 1.09) | N/A | ≥6 vs ≤2 hours/day |
| Patel 2015 (26126627)[56] | Kidney | Inc / Mort | CPS II | M | 379 / 146722 | 1.11 (0.82 - 1.52) | N/A | ≥6 vs ≤2 hours/day |
| Patel 2015 (26126627)[56] | Kidney | Inc / Mort | CPS II | F | 186 / 146722 | 0.97 (0.62 - 1.51) | N/A | ≥6 vs ≤2 hours/day |
| Chantaprasopsuk 2023 (37202564)[89] | Bladder | Inc / Mort | CPS II | M/F | 2215 / 146027 | 1.09 (0.96 - 1.24) | N/A | >6 vs 0-3 hours/day |
| Teras 2012 (22275172)[44] | Follicular lymphoma | Inc / Mort | CPS II | F | 137 / 146850 | 1.43 (0.81 - 2.53) | 0.11 | ≥6 vs ≤2.9 hours/day |
| Teras 2012 (22275172)[44] | Follicular lymphoma | Inc / Mort | CPS II | M | 143 / 146850 | 0.76 (0.45 - 1.31) | 0.19 | ≥6 vs ≤2.9 hours/day |
| Teras 2012 (22275172)[44] | Non-Hodgkin Lymphoma | Inc / Mort | CPS II | F | 863 / 146850 | 1.26 (1.01 - 1.59) | 0.011 | ≥6 vs ≤2 hours/day |
| Teras 2012 (22275172)[44] | Non-Hodgkin Lymphoma | Inc / Mort | CPS II | M | 1138 / 146850 | 0.95 (0.79 - 1.15) | 0.67 | ≥6 vs ≤2 hours/day |
| Teras 2012 (22275172)[44] | Diffuse large-Bcell lymphoma | Inc / Mort | CPS II | F | 197 / 146850 | 0.78 (0.45 - 1.37) | 0.94 | ≥6 vs ≤2.9 hours/day |
| Teras 2012 (22275172)[44] | Diffuse large-Bcell lymphoma | Inc / Mort | CPS II | M | 271 / 146850 | 0.75 (0.50 - 1.13) | 0.3 | ≥6 vs ≤2.9 hours/day |
| Patel 2015 (26126627)[56] | Multiple Myeloma | Inc / Mort | CPS II | M | 243 / 69260 | 1.02 (0.70 - 1.48) | N/A | ≥6 vs ≤2 hours/day |
| Patel 2015 (26126627)[56] | Multiple Myeloma | Inc / Mort | CPS II | F | 171 / 69260 | 1.64 (1.06 - 2.53) | N/A | ≥6 vs ≤2 hours/day |
| Teras 2012 (22275172)[44] | Leukaemia (CLL +SLL) | Inc / Mort | CPS II | F | 211 / 146850 | 1.16 (0.74 - 1.82) | 0.75 | ≥6 vs ≤2.9 hours/day |
| Teras 2012 (22275172)[44] | Leukaemia (CLL +SLL) | Inc / Mort | CPS II | M | 238 / 146850 | 1.11 (0.75 - 1.65) | 0.57 | ≥6 vs ≤2.9 hours/day |
| **Sitting time, non-recreational** | | | | | | | | |
| Nguyen 2018 (30740587)[71] | Colorectal (early onset) | Inc | NHS II | F | 118 / 89278 | 1.07 (0.70 - 1.65) | 0.46 | >14 vs <7 hours/week |
| **Sitting time, occupational** | | | | | | | | |

| Author Year | Cancer site | Outcome type* | Study name | Sex | Cases / cohort size | RR (95%CI) | p-value for trend/ linear analysis | Comparison |
|---|---|---|---|---|---|---|---|---|
| Ihira 2020 (31925977)[77] | Oesophageal | Inc | JPHC | M | 100 / 33307 | 1.05 (0.58 - 1.87) | 0.924 | ≥7 vs 1-<3 hours/day |
| Ihira 2020 (31925977)[77] | Oesophageal | Inc | JPHC | F | 8 / 33307 | 2.38 (0.35 - 16.45) | N/A | 3-5 vs 1-<3 hours/day |
| Ihira 2020 (31925977)[77] | Stomach | Inc | JPHC | M | 552 / 33307 | 1.08 (0.82 - 1.41) | 0.568 | ≥7 vs 1-<3 hours/day |
| Ihira 2020 (31925977)[77] | Stomach | Inc | JPHC | F | 127 / 33307 | 1.03 (0.59 - 1.81) | 0.696 | ≥7 vs 1-<3 hours/day |
| Gerhardsson 1986 (3962961)[94] | Colon | Inc | Sweden 1960 census | M | N/A / 9633 | 1.30 (1.20 - 1.50) | N/A | sitting >50% vs <50% of time |
| Gerhardsson 1986 (3962961)[94] | Rectal | Inc | Sweden 1960 census | M | N/A / 9633 | 1.10 (1.00 - 1.20) | N/A | sitting >50% vs <50% of time |
| Gerhardsson 1986 (3962961)[94] | Colon, ascending | Inc | Sweden 1960 census | M | N/A / 9633 | 1.50 (1.20 - 1.80) | N/A | sitting >50% vs <50% of time |
| Gerhardsson 1986 (3962961)[94] | Colon, transverse | Inc | Sweden 1960 census | M | N/A / 9633 | 1.60 (1.20 - 2.10) | N/A | sitting >50% vs <50% of time |
| Gerhardsson 1986 (3962961)[94] | Colon, descending | Inc | Sweden 1960 census | M | N/A / 9633 | 1.30 (0.90 - 1.90) | N/A | sitting >50% vs <50% of time |
| Gerhardsson 1986 (3962961)[94] | Colon, sigmoid | Inc | Sweden 1960 census | M | N/A / 9633 | 1.20 (1.00 - 1.40) | N/A | sitting >50% vs <50% of time |
| Simons 2013 (23420352)[45] | Colon, proximal | Inc | Sweden 1960 census | M/F | 456 / 4416 | 0.84 (0.63 - 1.12) | 0.25 | <2 vs >6-8 hours/day |
| Simons 2013 (23420352)[45] | Colon, distal | Inc | Sweden 1960 census | M/F | 535 / 4416 | 0.63 (0.48 - 0.83) | 0.001 | <2 vs >6-8 hours/day |
| Ihira 2020 (31925977)[77] | Liver | Inc | JPHC | M | 134 / 33307 | 1.54 (0.92 - 2.58) | 0.159 | ≥7 vs 1-<3 hours/day |
| Ihira 2020 (31925977)[77] | Liver | Inc | JPHC | F | 21 / 33307 | 0.30 (0.04 - 2.20) | 0.109 | ≥7 vs 1-<3 hours/day |
| Ihira 2020 (31925977)[77] | Lung | Inc | JPHC | M | 367 / 33307 | 1.07 (0.75 - 1.51) | 0.685 | ≥7 vs 1-<3 hours/day |
| Ihira 2020 (31925977)[77] | Lung | Inc | JPHC | F | 81 / 33307 | 2.80 (1.33 - 5.90) | 0.013 | ≥7 vs 1-<3 hours/day |
| Nomura 2016 (27632430)[60] | Breast, premenopausal | Inc | BWHS | F | 752 / 46734 | 1.14 (0.91 - 1.42) | 0.26 | ≥5 vs ≤0.9 hours/day |
| Ihira 2020 (31925977)[77] | Endometrial | Inc | JPHC | F | 50 / 33307 | 0.49 (0.15 - 1.52) | 0.314 | ≥7 vs 1-<3 hours/day |
| Ihira 2020 (31925977)[77] | Kidney | Inc | JPHC | M | 57 / 33307 | 1.04 (0.44 - 2.47) | 0.731 | ≥7 vs 1-<3 hours/day |
| Ihira 2020 (31925977)[77] | Kidney | Inc | JPHC | F | 12 / 33307 | 4.15 (0.66 - 26.11) | N/A | 5-7 vs 1-<3 hours/day |
| Ihira 2020 (31925977)[77] | Bladder | Inc | JPHC | M | 93 / 33307 | 0.56 (0.27 - 1.17) | 0.188 | ≥7 vs 1-<3 hours/day |
| Ihira 2020 (31925977)[77] | Bladder | Inc | JPHC | F | 15 / 33307 | 0.79 (0.14 - 4.27) | 0.927 | ≥7 vs 1-<3 hours/day |
| **Sitting time, transportational** | | | | | | | | |
| Keum 2016 (26649988)[58] | Colorectal | Inc | HPFS | M | 771 / 106521 | 1.04 (0.69 - 1.58) | 0.65 | >14 vs 0-3.4 hours/week |
| Keum 2016 (26649988)[58] | Colorectal | Inc | HPFS | M | 771 / 106521 | 0.97 (0.84 - 1.11) | N/A | Per 7 hours/day |
| Peduzzi 2024 (37985251)[90] | Pancreas | Inc | UK Biobank | M/F | N/A / 303461 | 1.05 (0.93 - 1.19) | N/A | Per 1 hours/day |
| Cohen 2013 (23576427)[46] | Breast | Inc | SCCS | F | 457 / 2730 | 1.05 (0.72 - 1.53) | 0.2 | ≥2 vs ≤0.32 hours/day |
| **Sitting time, other** | | | | | | | | |
| Ihira 2020 (31925977)[77] | Oesophageal | Inc | JPHC | F | 13 / 33307 | 0.76 (0.07 - 7.87) | N/A | ≥7 vs 1-<3 hours/day |
| Ihira 2020 (31925977)[77] | Stomach | Inc | JPHC | F | 216 / 33307 | 1.14 (0.73 - 1.75) | 0.533 | ≥7 vs 1-<3 hours/day |
| Eaglehouse 2017 (28542077)[62] | Colorectal | Inc | SCHS | M/F | 1981 / 61308 | 0.99 (0.84 - 1.17) | 0.99 | ≥3 vs 0 hours/day |
| Nguyen 2018 (30740587)[71] | Colorectal (early onset) | Inc | NHS II | F | 118 / 89278 | 1.34 (0.84 - 2.15) | 0.23 | >14 vs <7 hours/week |
| Ihira 2020 (31925977)[77] | Colorectal | Inc | JPHC | M/F | 353 / 33307 | 1.02 (0.70 - 1.50) | 0.695 | ≥7 vs 1-<3 hours/day |
| Ihira 2020 (31925977)[77] | Colon | Inc | JPHC | F | 261 / 33307 | 0.96 (0.60 - 1.51) | 0.948 | ≥7 vs 1-<3 hours/day |
| Ihira 2020 (31925977)[77] | Rectal | Inc | JPHC | F | 92 / 33307 | 1.22 (0.60 - 2.48) | 0.537 | ≥7 vs 1-<3 hours/day |
| Ihira 2020 (31925977)[77] | Liver | Inc | JPHC | F | 40 / 33307 | 0.62 (0.14 - 2.82) | 0.685 | ≥7 vs 1-<3 hours/day |
| Ihira 2020 (31925977)[77] | Pancreas | Inc | JPHC | F | 67 / 33307 | 0.67 (0.24 - 1.82) | 0.626 | ≥7 vs 1-<3 hours/day |

| Author Year | Cancer site | Outcome type* | Study name | Sex | Cases / cohort size | RR (95%CI) | p-value for trend/ linear analysis | Comparison |
|---|---|---|---|---|---|---|---|---|
| Ihira 2020 (31925977)[77] | Lung | Inc | JPHC | F | 128 / 33307 | 1.63 (0.89 - 2.99) | 0.139 | ≥7 vs 1-<3 hours/day |
| Cohen 2013 (23576427)[46] | Breast | Inc | SCCS | F | 457 / 2730 | 1.28 (0.91 - 1.80) | 0.52 | ≥4 vs 0 hours/day |
| Ihira 2020 (31925977)[77] | Breast | Inc | JPHC | F | 255 / 33307 | 1.21 (0.80 - 1.81) | 0.425 | ≥7 vs 1-<3 hours/day |
| Ihira 2020 (31925977)[77] | Endometrial | Inc | JPHC | F | 62 / 33307 | 0.96 (0.40 - 2.30) | 0.813 | ≥7 vs 1-<3 hours/day |
| Ihira 2020 (31925977)[77] | Ovary | Inc | JPHC | F | 39 / 33307 | 1.37 (0.49 - 3.85) | 0.8 | ≥7 vs 1-<3 hours/day |
| Buras 2022 (35094053)[84] | Ovary | Inc | NHS, NHS II | F | 884 / 171085 | 0.84 (0.66 - 1.08) | 0.04 | >20 vs <5 hours/week |
| Ihira 2020 (31925977)[77] | Kidney | Inc | JPHC | F | 19 / 33307 | 2.89 (0.70 - 11.92) | N/A | 5-7 vs 1-<3 hours/day |
| Ihira 2020 (31925977)[77] | Bladder | Inc | JPHC | F | 19 / 33307 | 0.73 (0.14 - 3.87) | N/A | ≥7 vs 1-<3 hours/day |
| **Screen time, recreational** | | | | | | | | |
| Kunzmann 2018 (30288276)[68] | Oesophageal | Inc | UK Biobank | M/F | 294 / 359033 | 1.14 (0.87 - 1.49) | 0.23 | 4-16 vs 0-3 hours/day |
| Kunzmann 2018 (30288276)[68] | Gastric | Inc | UK Biobank | M/F | 216 / 359033 | 1.13 (0.83 - 1.55) | 0.42 | 4-16 vs 0-3 hours/day |
| Kunzmann 2018 (30288276)[68] | Gastric, adeno | Inc | UK Biobank | M/F | 183 / 359033 | 1.11 (0.79 - 1.57) | 0.51 | 4-16 vs 0-3 hours/day |
| Kunzmann 2018 (30288276)[68] | Gastric, cardia | Inc | UK Biobank | M/F | 89 / 359033 | 0.95 (0.58 - 1.57) | 0.87 | 4-16 vs 0-3 hours/day |
| Kunzmann 2018 (30288276)[68] | Gastric, non-cardia | Inc | UK Biobank | M/F | 128 / 359033 | 1.29 (0.86 - 1.93) | 0.21 | 4-16 vs 0-3 hours/day |
| Zhang 2024 (38974470)[93] | Colorectal | Inc | UK Biobank | M/F | 4560 / 360271 | 1.08 (1.00 - 1.17) | N/A | >4 vs <3 hours/day |
| Zhang 2024 (38974470)[93] | Lung | Inc | UK Biobank | M/F | 2765 / 360271 | 1.26 (1.13 - 1.40) | N/A | >4 vs <3 hours/day |
| Zhang 2024 (38974470)[93] | Breast | Inc | UK Biobank | F | 7817 / 360271 | 1.01 (0.96 - 1.08) | N/A | >4 vs <3 hours/day |
| Zhang 2024 (38974470)[93] | Prostate | Inc | UK Biobank | M | 8644 / 360271 | 1.01 (0.95 - 1.07) | N/A | >4 vs <3 hours/day |
| **Screen time, computers, recreational** | | | | | | | | |
| Morris 2018 (29520109)[67] | Colorectal | Inc | UK Biobank | M/F | 2377 / 430584 | 0.94 (0.68 - 1.29) | 0.75 | ≥4 vs 0 hours/day |
| Morris 2018 (29520109)[67] | Colon | Inc | UK Biobank | M/F | 1546 / 430584 | 0.86 (0.56 - 1.30) | 0.81 | ≥4 vs 0 hours/day |
| Morris 2018 (29520109)[67] | Colon, proximal | Inc | UK Biobank | M/F | 778 / 430584 | 0.84 (0.46 - 1.53) | 0.79 | ≥4 vs 0 hours/day |
| Morris 2018 (29520109)[67] | Colon, distal | Inc | UK Biobank | M/F | 690 / 430584 | 1.00 (0.56 - 1.79) | 0.62 | ≥4 vs 0 hours/day |
| Morris 2018 (29520109)[67] | Rectal | Inc | UK Biobank | M/F | 817 / 430584 | 1.10 (0.68 - 1.79) | 0.98 | ≥4 vs 0 hours/day |
| **Television watching time** | | | | | | | | |
| Nguyen 2018 (30740587)[71] | Colorectal (early onset) | Inc | NHS II | F | 118 / 89278 | 1.77 (1.12 - 2.78) | 0.02 | >14 vs <7 hours/week |
| Li 2021 (33138348)[81] | Colon | Mort | JACC | M/F | 531 / 90834 | 1.43 (1.04 - 1.96) | 0.049 | >4.5 vs <1.5 hours/day |
| Li 2021 (33138348)[81] | Colon | Mort | JACC | M/F | 531 / 90834 | 1.07 (1.02 - 1.13) | N/A | Per 1 hours/day |
| Morris 2018 (29520109)[67] | Colon, proximal | Inc | UK Biobank | M/F | 780 / 430584 | 1.29 (0.93 - 1.80) | 0.009 | ≥4 vs 0 hours/day |
| Morris 2018 (29520109)[67] | Colon, distal | Inc | UK Biobank | M/F | 693 / 430584 | 1.33 (0.92 - 1.92) | 0.28 | ≥4 vs 0 hours/day |
| Nguyen 2018 (30740587)[71] | Colon (early onset) | Inc | NHS II | F | 82 / 89278 | 1.47 (0.85 - 2.54) | 0.22 | >14 vs <7 hours/week |
| Li 2021 (33138348)[81] | Rectal | Mort | JACC | M/F | 218 / 90834 | 1.13 (0.69 - 1.85) | 0.45 | >4.5 vs <1.5 hours/day |
| Li 2021 (33138348)[81] | Rectal | Mort | JACC | M/F | 218 / 90834 | 1.03 (0.94 - 1.13) | N/A | Per 1 hours/day |
| Nguyen 2018 (30740587)[71] | Rectal (early onset) | Inc | NHS II | F | 36 / 89278 | 2.44 (1.03 - 5.78) | 0.04 | >14 vs <7 hours/week |
| Hunter 2020 (32746843)[78] | Hepatobiliary tract | Inc | UK Biobank | M/F | 456 / 28992 | 1.26 (0.90 - 1.77) | N/A | >5 vs 1-<3 hours/day |
| Hunter 2020 (32746843)[78] | Hepatobiliary tract | Inc | UK Biobank | M/F | 456 / 28992 | 1.01 (0.96 - 1.07) | 0.62 | Per 1 hours/day |
| Hunter 2020 (32746843)[78] | Pancreas | Inc | UK Biobank | M/F | 615 / 28992 | 1.03 (0.74 - 1.44) | N/A | >5 vs 1-<3 hours/day |

| Author Year | Cancer site | Outcome type* | Study name | Sex | Cases / cohort size | RR (95%CI) | p-value for trend/ linear analysis | Comparison |
|---|---|---|---|---|---|---|---|---|
| Hunter 2020 (32746843)[78] | Pancreas | Inc | UK Biobank | M/F | 615 / 28992 | 1.03 (0.98 - 1.08) | 0.2 | Per 1 hours/day |
| Kim 2013 (24062293)[50] | Lung | Mort | MEC | M | 1010 / 134596 | 1.14 (0.87 - 1.51) | 0.76 | ≥5 vs <1 hours/day |
| Kim 2013 (24062293)[50] | Lung | Mort | MEC | F | 688 / 134596 | 1.09 (0.81 - 1.49) | 0.14 | ≥5 vs <1 hours/day |
| Xiao 2013 (23966580)[49] | Ovary, epithelial | Inc | NIH-AARP | F | 463 / 153123 | 1.02 (0.67 - 1.55) | N/A | ≥7 vs ≤2.9 hours/day |
| Lynch 2014b (24526287)[53] | Prostate, advanced | Inc | NIH-AARP | M | 1365 / 170481 | 0.93 (0.79 - 1.09) | 0.49 | ≥5 vs <3 hours/day |
| Hunter 2020 (32746843)[78] | Brain | Inc | UK Biobank | M/F | 463 / 28992 | 0.96 (0.63 - 1.46) | N/A | >5 vs 1-<3 hours/day |
| Hunter 2020 (32746843)[78] | Brain | Inc | UK Biobank | M/F | 463 / 28992 | 1.04 (0.98 - 1.10) | 0.2 | Per 1 hours/day |
| Hunter 2020 (32746843)[78] | Thyroid | Inc | UK Biobank | M/F | 242 / 28992 | 1.14 (0.63 - 2.06) | N/A | >5 vs 1-<3 hours/day |
| Hunter 2020 (32746843)[78] | Thyroid | Inc | UK Biobank | M/F | 242 / 28992 | 1.00 (0.92 - 1.03) | 0.89 | Per 1 hours/day |
| Hunter 2020 (32746843)[78] | Non-Hodgkin Lymphoma | Inc | UK Biobank | M/F | 1193 / 28992 | 0.85 (0.65 - 1.11) | N/A | >5 vs 1-<3 hours/day |
| Hunter 2020 (32746843)[78] | Non-Hodgkin Lymphoma | Inc | UK Biobank | M/F | 1193 / 28992 | 1.01 (0.98 - 1.05) | 0.48 | Per 1 hours/day |
| **Television watching frequency** | | | | | | | | |
| Cuthbertson 2020 (32978174)[80] | Colorectal | Inc | ARIC | M/F | 361 / 14508 | 0.96 (0.70 - 1.31) | N/A | Seldom/never vs often/very often |
| Cuthbertson 2020 (32978174)[80] | Lung | Inc | ARIC | M/F | 560 / 14508 | 1.09 (0.85 - 1.40) | N/A | Seldom/never vs often/very often |
| Cuthbertson 2020 (32978174)[80] | Breast, postmenopausal | Inc | ARIC | F | 564 / 14508 | 1.03 (0.81 - 1.30) | N/A | Seldom/never vs often/very often |
| Cuthbertson 2020 (32978174)[80] | Prostate | Inc | ARIC | M | 813 / 14508 | 1.20 (0.99 - 1.47) | N/A | Seldom/never vs often/very often |

**Abbreviations:** CI: confidence interval; CLL: chronic lymphocytic leukaemia; F: female; M: male; N/A: not applicable; RR: relative risk; SLL: small lymphocytic lymphoma; SCC: squamous cell carcinoma.

**Study abbreviations:** ARIC: Atherosclerosis risk in communities study; BWHS: Black women's health study; CPS II: Cancer Prevention Study II; HPFS: Health professionals follow-up study; HUNT: HUNT - The Trøndelag health study; J-MICC: Japan multi-institutional collaborative cohort study; JACC: Japan collaborative cohort study; JPHC: Japan public health center-based prospective study; MEC: Multiethnic cohort study; NHS: Nurses' health study; NHS II: Nurses' health study II; NIH-AARP: National institute of health - American association for retired persons diet and health study; NLCS: The Netherlands cohort study; SCCS :Southern community cohort study; SCHS :Singapore Chinese health study; Sweden 1960 census: Sweden 1960 census study; UK Biobank: UK Biobank; WHI: Women’s health initiative study.

* In outcome type, the abbreviation “Inc / Mort” denotes that the study included both incident cancers and cancer deaths in the definitions for cancer cases.

#### Supplementary Table 12: Descriptive results of combined, joint, or interaction analyses between sedentary behaviour and physical activity on cancer risk or mortality.

| Author, year (reference) | Description of results |
| --- | --- |
| **Total sedentary time** | |
| Tomida 2024 (38041484)[91] | • When the effects of leisure-time physical activity were examined in combination with the sedentary time, sedentary time was shown to outweigh the protective effects of physical activity and be a risk factor for breast cancer. |
| **Total sitting time** | |
| Buras 2022 (35094053)[84] | • No heterogeneity was observed by physical activity level for the association between total sitting time (p heterogeneity = 0.50) and ovarian cancer risk. |
| Park 2019 (30910780)[73] | • In combined analyses of physical activity and sitting time on colorectal cancer risk, an inverse association with moderate/vigorous activity was seen in males with total sitting time longer than 6 hours per day, but not in those with 6 or less hours (p heterogeneity = 0.06).<br>• No heterogeneity was observed for total sitting in females (p heterogeneity = 0.89). |
| Nunez 2018 (29510753)[66] | • No interaction was observed between moderate or vigorous physical activity and prolonged sitting on colon or rectal cancer risk (all p interaction > 0.1). |
| Gorczyca 2018 (28538039)[61] | • No interaction was observed between total sitting time and physical activity on colorectal cancer risk (p interaction = 0.62). |
| Nomura 2017 (28975422)[63] | • An interaction was observed between total sitting time and moderate and strenuous recreational physical activity on breast cancer risk (p interaction = 0.009):<br>o High recreational activity: $HR_{\geq 10 \text{ vs } \leq 5 \text{ hours/day total sitting time}}$: 1.24 (95%CI: 1.07-1.44)<br>o Low recreational activity: $HR_{\geq 10 \text{ vs } \leq 5 \text{ hours/day total sitting time}}$: 0.92 (95%CI: 0.80-1.05)<br>• An interaction was observed between total sitting time and total recreational physical activity on breast cancer risk (p interaction = 0.002).<br>o >20 MET hours/week recreational activity: $HR_{\geq 10 \text{ vs } \leq 5 \text{ hours/day total sitting time}}$: 1.32 (95%CI: 1.12-1.56)<br>o >0-10 MET hours/week recreational activity: $HR_{\geq 10 \text{ vs } \leq 5 \text{ hours/day total sitting time}}$: 0.95 (95%CI: 0.83-1.09)<br>• No interaction was observed between total sitting time and walking (p interaction = 0.97) or combined physical activity (p interaction = 0.32) on breast cancer risk. |
| Nomura 2016 (27632430)[60] | • An interaction was observed between total sitting time and vigorous activity (p = 0.04), but few females who reported 10 or more hours of sitting time also reported regular vigorous exercise.<br>• No interaction was observed between sitting time and combined physical activity level (p interaction = 0.27) or time spent walking for exercise (p interaction = 0.43) on breast cancer risk.<br>• In the joint effects analysis, no clear pattern was observed across levels of total physical activity, vigorous exercise or walking for exercise and total sitting time on breast cancer risk. |
| Lynch 2014b (24526287)[53] | • No interaction was observed between total sitting time and moderate-to-vigorous intensity physical activity on prostate cancer risk (results not shown). |
| Xiao 2013 (23966580)[49] | • No interaction was observed between total sitting time and moderate-to-vigorous physical activity on epithelial ovarian cancer (p interaction = 0.66). |
| Moore 2010 (20877336)[40] | • In a joint effects analysis, females who were inactive (vigorous physical activity fewer than three times per week) and who sat for 9 or more hours/day had twice the risk of endometrial cancer of active females who sat fewer than 3 hours/day [RR:2.14 (95%CI: 1.48-3.10)] vs [RR: 1.19 (95%CI: 0.69-2.05)]. |
| **Occupational sitting time** | |

| Author, year (reference) | Description of results |
|---|---|
| Buras 2022 (35094053)[84] | • No heterogeneity was observed by physical activity level for the association between sitting at work (p heterogeneity = 0.91) and ovarian cancer risk. |
| Ihira 2020 (31925977)[77] | • No interaction was observed between occupational siting time (<3 hours/day or ≥3 hours/day) and leisure time moderate-to-vigorous physical activity (<30 minutes/week or ≥30 minutes/week) on pancreatic cancer risk (p interaction = 0.965). |
| Pronk 2011 (21934685)[42] | • No interaction was observed in a joint effects analysis between occupational sitting time and exercise participation (below or above the recommended level of 8 MET hours/week/year) on breast cancer risk. |
| **Recreational sitting time** | |
| Patel 2015 (26126627)[56] | • Exercise activity did not modify associations of sitting time with total or site-specific cancer risk (data not shown). |
| Teras 2012 (22275172)[44] | • No interaction was observed between physical activity and sitting time on the risk of non-Hodgkin lymphoid neoplasms. |
| **Other sitting time** | |
| Buras 2022 (35094053)[84] | • No heterogeneity was observed by physical activity level for the association between other sitting at home (p heterogeneity = 0.15) and ovarian cancer risk. |
| **TV watching time** | |
| Buras 2022 (35094053)[84] | • No heterogeneity was observed by physical activity level for the association between watching TV or movies at home (p heterogeneity = 0.26) and ovarian cancer risk. |
| Sanchez-Bayona 2021 (33798533)[82] | • No interaction was observed between the levels of leisure-time physical activity and TV watching on breast cancer risk (p interaction = 0.98). |
| Li 2021 (33138348)[81] | • No interaction was observed between the levels of leisure-time physical activity and TV watching on colorectal, colon, and rectal cancer mortality (all p interaction > 0.1). |
| Hunter 2020 (32746843)[78] | • An interaction was observed between physical activity level and TV viewing time only for rectum cancers amongst males (p interaction = 0.004).<br>• No interactions were observed for other cancers either in males or in females (all p interaction > 0.01). |
| Park 2019 (30910780)[73] | • In combined analyses of moderate/vigorous activity and TV watching time on colorectal cancer risk, no heterogeneity was observed for both males (p heterogeneity = 0.52) and females (p heterogeneity = 0.52). |
| Morris 2018 (29520109)[67] | • No interaction was observed between total physical activity and subgroups of TV watching time on colorectal cancer risk (p interaction = 0.24). |
| Nguyen 2018 (30740587)[71] | • No heterogeneity between TV viewing time and young-onset colorectal cancer according to physical activity strata (median <15 vs ≥15 METs-hours/week) (p interaction = 0.83) |
| Eaglehouse 2017 (28542077)[62] | • An interaction was observed between moderate-intensity physical activity and TV watching time on colorectal cancer risk (p interaction = 0.042). A suggestive inverse association between moderate-intensity physical activity and colorectal cancer risk was observed in those who sat for at least 3 hours/day watching TV, but not in those who sat up to 2 hours/day watching TV.<br>• No interaction was observed between TV watching time and other physical activity (i.e. strenuous–vigorous intensity (p interaction = 0.51) and overall physical activity (p interaction = 0.1) levels on colorectal cancer risk. |
| Keum 2016 (26649988)[58] | • In a joint association of TV watching time and physical activity on colorectal cancer risk, females who were highly sedentary (>21 hours/week of sitting watching TV) and physically less active (below median MET-hours/week) had an ~41% elevated risk (95%CI: 1.03-1.92) compared with females who were less sedentary (<14 hours/week of sitting watching TV) and physically more active (above median MET-hours/week). No pattern was observed in the joint analysis among males.<br>• No interaction was observed between TV watching time and physical activity on colorectal cancer risk in males (p interaction = 0.55) or females (p interaction = 0.23). |

| Author, year (reference) | Description of results |
|---|---|
| Lynch 2014b (24526287)[53] | • No interaction was observed between TV viewing time and moderate-to-vigorous intensity physical activity on prostate cancer risk (results not shown). |
| Ukawa 2014 (24728669)[54] | • Compared with participants in the shorter walking category (<0.5 hours/day) and longest TV watching category (>4 hours/day) those in the longer walking category (>0.5 hours/day) and middle and shortest TV watching categories (2 to 4 and <2 hours/day) had a significantly reduced risk of liver cancer mortality (HR: 0.58; 95%CI: 0.39-0.89 and HR 0.58; 95%CI: 0.35-0.98, respectively). ***Estimates were comparable between shorter and longer walking time with largely overlapping 95%Cis providing little evidence of interaction.*** |
| **Recreational screen time** | |
| Zhang 2024 (38974470)[93] | • No interaction was observed between total physical activity and recreational screen time on prostate, breast, colorectal, and lung cancer risks (all p interaction > 0.46). |

**Abbreviations:** CI: confidence interval; HR: hazard ratio; MET: metabolic equivalent of task; RR: risk ratio.

#### Supplementary Table 13: Subgroup analyses of sitting time and cancer risk.

| | Studies | RR (95%CI) | $I^2$ (%) | $P_{heterogeneity}$ between subgroups |
|---|---|---|---|---|
| **Total sitting time** | | | | |
| **Breast cancer, main analysis, per 2 hours/day** | **4** | **1.04 (1.01 - 1.07)** | **0** | |
| **Subgroup** | | | | |
| ER+/PR+ | 2 | 1.05 (0.95 - 1.17) | 83 | 0.585 |
| ER-PR- | 2 | 1.01 (0.91 - 1.13) | 65 | |
| Premenopausal | 3 | 1.02 (0.98 - 1.07) | 0 | 0.879 |
| Postmenopausal | 5 | 1.03 (1.00 - 1.05) | 0 | |
| Not adjusted for physical activity | 2 | 1.02 (0.97 - 1.07) | 0 | 0.417 |
| Adjusted for physical activity | 2 | 1.05 (1.02 - 1.08) | 0 | |
| Critical risk of bias in confounding | 2 | 1.02 (0.97 - 1.07) | 0 | 0.417 |
| Moderate/serious risk of bias in confounding | 2 | 1.05 (1.02 - 1.08) | 0 | |
| **Breast cancer, main analysis, high vs low** | **6** | **1.17 (1.02 - 1.35)** | **38.7** | |
| **Subgroup** | | | | |
| Critical risk of bias in confounding | 3 | 1.17 (0.87 - 1.56) | 54.6 | 0.148 |
| Moderate/serious risk of bias in confounding | 3 | 1.20 (1.02 - 1.42) | 38.1 | |
| **Colorectal cancer, main analysis, per 2 hours/day** | **3** | **1.01 (0.98 - 1.03)** | **34.6** | |
| **Subgroup** | | | | |
| Colon | 3 | 1.03 (1.01 - 1.06) | 0 | 0.203 |
| Rectal | 2 | 0.99 (0.93 - 1.05) | 0 | |
| **Occupational sitting time** | | | | |
| **Breast cancer, main analysis, per 2 hours/day** | **4** | **1.05 (1.01 - 1.09)** | **0** | |
| **Subgroup** | | | | |
| Premenopausal | 1 | 1.05 (0.97 - 1.13) | - | 0.451 |
| Postmenopausal | 2 | 1.00 (0.90 - 1.10) | 75.5 | |
| **Colorectal cancer, main analysis, per 2 hours/day** | **5** | **1.03 (0.99 - 1.07)** | **27.0** | |
| **Subgroup** | | | | |
| Colon | 2 | 1.08 (1.02 - 1.15) | 28.2 | **0.021** |
| Rectal | 2 | 0.97 (0.91 - 1.04) | 0 | |
| Male | 4 | 1.03 (0.97 - 1.09) | 42.6 | 0.745 |
| Female | 2 | 1.01 (0.96 - 1.07) | 0 | |
| Asia | 2 | 0.92 (0.70 - 1.21) | 75.4 | 0.778 |

| | | | | |
|---|---|---|---|---|
| North America | 2 | 1.02 (0.96 - 1.08) | 0 | |
| Not adjusted for physical activity | 2 | 0.93 (0.69 - 1.26) | 79.1 | 0.543 |
| Adjusted for physical activity | 3 | 1.02 (0.98 - 1.07) | 0 | |
| Critical risk of bias in confounding | 2 | 0.93 (0.69 - 1.26) | 79.1 | 0.543 |
| Moderate/serious risk of bias in confounding | 3 | 1.02 (0.98 - 1.07) | 0 | |
| Critical risk of bias in selective reporting | 2 | 1.02 (0.96 - 1.08) | 0 | 0.864 |
| Moderate/serious risk of bias in selective reporting | 3 | 1.02 (0.95 - 1.10) | 58.8 | |

**Abbreviations:** CI: confidence interval; RR: relative risk.

#### Supplementary Table 14: Subgroup analyses of television watching time and cancer risk.

| | Studies | RR (95%CI) | $I^2$ (%) | $P_{heterogeneity}$ between subgroups |
|---|---|---|---|---|
| **Breast cancer, main analysis, per 2 hours/day** | **6** | **1.03 (0.98 - 1.08)** | **27.8** | |
| **Subgroup** | | | | |
| Europe | 2 | 1.27 (0.76 - 2.14) | 79.5 | 0.412 |
| North America | 3 | 1.02 (0.97 - 1.07) | 0 | |
| Premenopausal | 5 | 1.01 (0.91 - 1.13) | 49.9 | 0.983 |
| Postmenopausal | 6 | 1.02 (0.99 - 1.04) | 0 | |
| Not adjusted for physical activity | 2 | 1.02 (0.98 - 1.06) | 0 | 0.518 |
| Adjusted for physical activity | 4 | 1.06 (0.95 - 1.20) | 56.2 | |
| Critical risk of bias in confounding | 4 | 1.06 (0.95 - 1.20) | 56.2 | 0.518 |
| Moderate/serious risk of bias in confounding | 2 | 1.02 (0.98 - 1.06) | 0 | |
| **Colorectal cancer, main analysis, per 2 hours/day** | **6** | **1.03 (1.01 - 1.06)** | **0** | |
| **Subgroup** | | | | |
| Colon | 5 | 1.08 (1.01 - 1.11) | 0 | 0.063 |
| Rectal | 4 | 1.01 (0.95 - 1.08) | 0 | |
| Male | 3 | 1.04 (0.94 - 1.15) | 76.1 | 0.892 |
| Female | 4 | 1.05 (1.01 - 1.09) | 0 | |
| Not adjusted for BMI | 3 | 1.07 (1.01 - 1.13) | 0 | 0.297 |
| Adjusted for BMI | 6 | 1.03 (1.00 - 1.06) | 8 | |
| Not adjusted for BMI - both models | 3 | 1.07 (1.01 - 1.13) | 0 | 0.855 |
| Adjusted for BMI - both models | 3 | 1.06 (0.97 - 1.15) | 22.5 | |
| Not adjusted for physical activity | 3 | 1.02 (1.00 - 1.05) | 0 | 0.201 |
| Adjusted for physical activity | 3 | 1.07 (1.01 - 1.13) | 0 | |
| Critical risk of bias in confounding | 2 | 1.02 (0.99 - 1.06) | 22.7 | 0.245 |
| Moderate/serious risk of bias in confounding | 4 | 1.06 (1.01 - 1.11) | 0 | |
| **Colon cancer, main analysis, per 2 hours/day** | **6** | **1.04 (1.01 - 1.06)** | **69.6** | |
| **Subgroup** | | | | |
| Male | 3 | 1.11 (1.06 - 1.17) | 0 | 0.158 |
| Female | 4 | 1.06 (1.01 - 1.12) | 0 | |
| **Rectal cancer, main analysis, per 2 hours/day** | **5** | **1.01 (0.99 - 1.03)** | **16.8** | |
| **Subgroup** | | | | |
| Male | 2 | 1.01 (0.98 - 1.23) | 0 | 0.676 |
| Female | 2 | 1.06 (0.94 - 1.20) | 0 | |

| | | | | |
|---|---|---|---|---|
| **Lung cancer, main analysis, high vs low** | **4** | **1.26 (1.15 - 1.38)** | 0 | |
| **Subgroup** | | | | |
| Critical risk of bias in confounding | 2 | 1.28 (1.15 - 1.42) | 0 | 0.463 |
| Moderate/serious risk of bias in confounding | 2 | 1.18 (0.98 - 1.43) | 0 | |
| **Ovarian cancer, main analysis, per 2 hours/day** | **5** | **1.00 (0.94 - 1.07)** | **0** | |
| **Subgroup** | | | | |
| Not adjusted for physical activity | 3 | 1.05 (0.87 - 1.28) | 32.2 | 0.583 |
| Adjusted for physical activity | 2 | 0.99 (0.91 - 1.08) | 8.3 | |
| **Endometrial cancer, main analysis, per 2 hours/day** | **3** | **1.05 (0.98 - 1.25)** | **82.1** | |
| **Subgroup** | | | | |
| Not adjusted for BMI | 1 | 1.16 (1.07 - 1.26) | - | 0.001 |
| Adjusted for BMI | 2 | 0.95 (0.87 - 1.04) | 0 | |
| **Endometrial cancer, main analysis, high vs low** | **4** | **1.30 (0.69 – 2.46)** | **86.3** | |
| **Subgroup** | | | | |
| Not adjusted for BMI | 2 | 1.71 (1.31 - 2.22) | 0 | 0.362 |
| Adjusted for BMI | 2 | 0.96 (0.29 - 3.21) | 72 | |
| **Prostate cancer, main analysis, per 2 hours/day** | **2** | **0.99 (0.98 - 1.01)** | **0** | |
| **Subgroup** | | | | |
| BMI<30 kg/m$^2$ | 2 | 0.99 (0.97 - 1.02) | 0 | 0.476 |
| BMI>30 kg/m$^2$ | 2 | 1.01 (0.96 - 1.07) | 43 | |

**Abbreviations:** BMI: body mass index; CI: confidence interval; RR: relative risk.

#### Supplementary Table 15: Descriptive table of the included Mendelian randomisation analyses of sedentary behaviour and cancer risk.

| Author, Year, PMID | MR design | Exposure population source | Exposure population ancestry, sample size | No. IVs (SNPs) | Cancer population source | Cancer population ancestry | Cancer cases/ controls | Outcome, comparison | Main results OR (95% CI) | Sensitivity analyses |
|---|---|---|---|---|---|---|---|---|---|---|
| Zhang 2021 (33510439)[96] | Two-sample; summary-level data | Measured sedentary time/total period of accelerometer with 30s intervals<br>UK Biobank GWAS | European<br>91105 individuals | 6 | GWAS meta-analysis of 15 datasets (NSCCG, the SCOT study, SOCCS/GS, SOCCS/LBC, UK Biobank, UK1, Scotland1, VQ58, CCFR1, CCFR2, COIN, Finnish GWAS, CORSA, DACHS and Croatia) | European | 31197/ 61770 | Colorectal cancer<br>Per 1 SD | Random-effects IVW<br>0.80 (0.57-1.12) p=0.19 | Leave-one-out, MR-Egger, MR-PRESSO, MR-Robust, median-based estimate, mode-based estimate |
| Gentiluomo 2024 (38425433)[105] | Two-sample; summary-level data | Measured sedentary time/total period of accelerometer with 30s intervals<br>UK Biobank GWAS | European<br>91105 individuals | 4 | PanScan I-III-PanC4 | European | 8769/ 7055 | Pancreatic cancer<br>Per 1 SD | Random-effects IVW<br>0.70 (0.31-1.61) p=0.405 | Leave-one-out analysis, weighted median, MR-Egger, Lasso, MR-PRESSO, contamination mixture |
| Shen 2021 (34076349)[97] | Two-sample; summary-level data | Measured sedentary time/total period of accelerometer with 30s intervals<br>UK Biobank GWAS | European<br>408815 individuals | 4 | International Lung Cancer Consortium GWAS meta-analysis | European | 11348/ 15861 | Lung cancer<br>Per 1 SD | IVW<br>0.95 (0.45-2.00) p=0.893 | Leave-one-out analysis, weighted median, MR-Egger, MR-PRESSO |
| | | | | | | | | Lung adenocarcinoma | IVW<br>0.52 (0.17-1.63) p=0.261 | |
| | | | | | | | | Lung squamous cell carcinoma | IVW<br>1.02 (0.32-3.23) p=0.969 | |
| Dixon-Suen 2022 (36328784)[101] | Two-sample; individual-level data | Measured sedentary time/total period of accelerometer with 30s intervals | European<br>91105 individuals | 6 | BCAC | European | 69838/ 54452 | Breast cancer, invasive | IVW<br>1.20 (0.93-1.55) p≥0.05 | Leave-one-out analysis, weighted median, MR-Egger, MR-PRESSO |

| Author, Year, PMID | MR design | Exposure population source | Exposure population ancestry, sample size | No. IVs (SNPs) | Cancer population source | Cancer population ancestry | Cancer cases/ controls | Outcome, comparison | Main results OR (95% CI) | Sensitivity analyses |
|---|---|---|---|---|---|---|---|---|---|---|
| | | UK Biobank GWAS | | | | | | Per 7% time spent in sedentary | | |
| | | | | | | | 23999/ 54452 | Pre/perimenopausal breast cancer<br>Per 7% time spent in sedentary | IVW<br>1.22 (0.78-1.90) p≥0.05 | |
| | | | | | | | 45839/ 54452 | Postmenopausal breast cancer | IVW<br>1.21 (0.89-1.65) p≥0.05 | |
| | | | | | | | 46528/ 54452 | ER+ breast cancer | IVW<br>1.19 (0.90-1.57) p≥0.05 | |
| | | | | | | | 11246/ 54452 | ER- breast cancer | IVW<br>1.43 (0.90-2.26) p≥0.05 | |
| | | | | | | | 34891/ 54452 | PR+ breast cancer | IVW<br>1.19 (0.87-1.63) p≥0.05 | |
| | | | | | | | 16432/ 54452 | PR- breast cancer | IVW<br>1.40 (0.94-2.09) p≥0.05 | |
| | | | | | | | 6945/ 54452 | HER2+ breast cancer | IVW<br>1.17 (0.67-2.06) p≥0.05 | |
| | | | | | | | 33214/ 54452 | HER2- breast cancer | IVW<br>1.27 (0.93-1.74) p≥0.05 | |

| Author, Year, PMID | MR design | Exposure population source | Exposure population ancestry, sample size | No. IVs (SNPs) | Cancer population source | Cancer population ancestry | Cancer cases/ controls | Outcome, comparison | Main results OR (95% CI) | Sensitivity analyses |
|---|---|---|---|---|---|---|---|---|---|---|
| Si 2021 (34671129)[98] | Two-sample; summary-level data | Measured sedentary time/total period of accelerometer with 30s intervals<br>UK Biobank GWAS | European<br>91105 individuals | 4 | BCAC, DRIVE, iCOGs, GWAS meta-analyses | European | 122977/ 105974 | Breast cancer<br>Per 1 SD | IVW<br>1.23 (0.97-1.55) p=0.088 | Weighted median, MR-Egger, MVMR |
| Si 2021 (34671129)[98] | Two-sample; summary-level data | Measured sedentary time/total period of accelerometer with 30s intervals<br>UK Biobank GWAS | European<br>91105 individuals | 4 | OCAC | European | 25509/ 40941 | Ovarian cancer<br>Per 1 SD | IVW<br>0.86 (0.52-1.42) p=0.563 | Weighted median, MR-Egger, MVMR |
| Kazmi 2020 (31802111)[95] | Two-sample; summary-level data | Measured sedentary time/total period of accelerometer with 30s intervals<br>UK Biobank GWAS | European<br>91105 individuals | 2 | PRACTICAL, GAME-ON/ELLIPSE | European | 79148/ 61106 | Prostate cancer<br>Per 1 SD | Fixed effect IVW<br>0.95 (0.63-1.42) p=0.79 | Weighted median, MR-Egger, weighted mode, MR-PRESSO |
| | | | | | | | 15167/ 58308 | Advanced prostate cancer | Fixed effect IVW<br>0.59 (0.15-2.41) p=0.46 | |

#### Supplementary Table 16: Descriptive table of the included Mendelian randomisation analyses of sitting time, sedentary at work and driving and cancer risk.

| First author, Year, PMID | MR design | Exposure population source | Exposure population ancestry, sample size | No. IVs (SNPs) | Cancer population source | Cancer population ancestry | Number of cases/controls | Outcome, comparison | Main results OR (95% CI) | Sensitivity analysis |
|---|---|---|---|---|---|---|---|---|---|---|
| Chen 2023 (37148539)[103] | Two-sample; summary-level data | Driving time UK Biobank | European 408815 | 5 | INHANCE | European | 2497/2928 | Oral cavity and pharyngeal cancer Per 1 SD | Fixed effect IVW 1.28 (0.05-32.69) p>0.05 | Weighted median, MR-Egger, MR-PRESSO, Weighted mode |
| Went 2024 (38527997)[107] | Two-sample; summary-level data | Sedentary behaviour at work 51 studies | European 372605 | 4 | GWAS meta-analysis | European | 16790/32476 | Oesophageal cancer Per 1 SD | Random-effects IVW 0.66 (0.41-1.07) p=0,093 | Leave-one-out analysis, Weighted median, MR-Egger, MR-PRESSO, MR Steiger, Weighted mode |
| | | Driving time UK Biobank MRC-IEU ukb-b-3793 | European 310555 | 6 | | | | | Random-effects IVW 0.20 (0.05-0.77) p=0,654 | |
| Chen 2023 (37148539)[103] | Two-sample; summary-level data | Driving time UK Biobank | European 408815 | 5 | Finnish Biobank N/A | European | 232/218560 | Oesophageal cancer Per 1 SD | Fixed effect IVW 34.63 (0.11-1.11x10^4) p>0.05 | Weighted median, MR-Egger, MR-PRESSO, Weighted mode |
| Went 2024 (38527997)[107] | Two-sample; summary-level data | Sedentary behaviour at work 51 studies | European 372605 | 4 | GWAS meta-analysis | European | 74673/86854 | Colorectal cancer Per 1 SD | Random-effects IVW 0.88 (0.64-1.21) p=0,437 | Leave-one-out analysis, Weighted median, MR-Egger, MR-PRESSO, MR Steiger, Weighted mode |
| | | Driving time UK Biobank MRC-IEU ukb-b-3793 | European 310555 | 6 | | | | | Random-effects IVW 0.69 (0.26-1.84) p=0,463 | |
| Chen 2023 (37148539)[103] | Two-sample; summary-level data | Driving time UK Biobank | European 408815 | 5 | UK Biobank N/A | European | 2437/358757 | Colon cancer Per 1 SD | Random-effects IVW 1.01 (1.00-1.03) p>0.05 | Weighted median, MR-Egger, MR-PRESSO, Weighted mode |
| | | | | | Finnish Biobank N/A | European | 304/218488 | Liver and intrahepatic bile duct cancer Per 1 SD | Random-effects IVW 3.48 (0.01-2207.12) p=>0.05 | |
| Gentiluomo 2024 (38425433)[105] | Two-sample; summary-level data | Sedentary behaviour at work 51 studies | European 372605 | 4 | PanScan I-III-PanC4 | European | 8769/7055 | Pancreatic cancer Per 1 SD | Random-effects IVW 0.71 (0.38-1.32) p=0,281 | Leave-one-out analysis, weighted median, MR-Egger, Lasso, MR-PRESSO, |

| First author, Year, PMID | MR design | Exposure population source | Exposure population ancestry, sample size | No. IVs (SNPs) | Cancer population source | Cancer population ancestry | Number of cases/controls | Outcome, comparison | Main results OR (95% CI) | Sensitivity analysis |
|---|---|---|---|---|---|---|---|---|---|---|
| | | | | | | | | | | contamination mixture |
| | | Driving time UK Biobank | European<br>408815 | 4 | | | | | Random-effects IVW 0.45 (0.09-2.21) p=0,323 | |
| Chen 2023 (37148539)[103] | Two-sample; summary-level data | Driving time UK Biobank | European 408815 | 3 | PanScan1 | European | 1896/1939 | Pancreatic cancer<br>Per 1 SD | Fixed effect IVW 5.59 (0.09-335.42) p>0.05 | Weighted median, MR-Egger, MR-PRESSO, Weighted mode |
| Went 2024 (38527997)[107] | Two-sample; summary-level data | Sedentary behaviour at work 51 studies | European 372605 | 4 | LC3, ILCCO meta-analysis | European | 29266/56450 | Lung cancer<br>Per 1 SD | Random-effects IVW 0.91 (0.56-1.48) p=0,694 | Leave-one-out analysis, Weighted median, MR-Egger, MR-PRESSO, MR Steiger, Weighted mode |
| | | Driving time UK Biobank MRC-IEU ukb-b-3793 | European 310555 | 6 | | | | | Random-effects IVW 1.09 (0.40-2.96) p=0,860 | |
| Peng 2021 (34899835)[100] | Two-sample; summary-level data | Driving time UK Biobank | European<br>408815 | 4 | ILCCO meta-analysis | European | 11348/15861 | Lung cancer<br>Per 1 h/d | IVW 1.02 (0.29-3.62) p>0.05 | Weighted median, MR-Egger, MR-PRESSO, Weighted mode |
| Gao 2021 (34777480)[99] | Two-sample; summary-level data | Driving time UK Biobank | European<br>408815 | 3 | ILCCO meta-analysis | European | 11348/15861 | Lung cancer<br>Per 1.5 h/d | Random-effects IVW (P het<0.1) or fixed effect IVW 0.77 (0.19-3.08) p=0,707 | Leave-one-out analysis, weighted median, MR-Egger, MR-PRESSO, MR-APSS, constrained max likelihood, model averaging (cML-MA) |
| | | | | | | | 3442/14894 | Lung cancer, adenocarcinoma | Random-effects IVW (P het<0.1) or fixed effect IVW 1.13 (0.13-9.87) p=0,912 | max likelihood, model averaging (cML-MA) |
| | | | | | | | 3275/ 15038 | Lung cancer, squamous cell carcinoma | Random-effects IVW (P het<0.1) or fixed effect IVW 0.86 (0.02-39.12) p=0,938 | |
| Chen 2023 (37148539)[103] | Two-sample; | Driving time UK Biobank | European 408815 | 5 | UK Biobank N/A | European | 2677 /334482 | Melanoma Per 1 SD | Fixed effect IVW 1.00 (0.98-1.03) p>0.05 | Weighted median, MR-Egger, MR- |

| First author, Year, PMID | MR design | Exposure population source | Exposure population ancestry, sample size | No. IVs (SNPs) | Cancer population source | Cancer population ancestry | Number of cases/controls | Outcome, comparison | Main results OR (95% CI) | Sensitivity analysis |
|---|---|---|---|---|---|---|---|---|---|---|
| | summary-level data | | | | | | | | | PRESSO, Weighted mode |
| Li 2023 (36815291)[102] | Two-sample; summary-level data | Playing computer games UK Biobank MRC-IEU ukb-b-4779 | European 462433 | 46 | UK Biobank | European | 17416/375455 | Skin cancer, basal cell carcinoma<br><br>Per 1 SD | IVW 0.66 (0.33-1.35) p=0,250 | Weighted median, MR-Egger, simple mode, weighted mode |
| Went 2024 (38527997)[107] | Two-sample; summary-level data | Sedentary behaviour at work 51 studies | European 372605 | 4 | BCAC | European | 133384/113789 | Breast cancer<br><br>Per 1 SD | Random-effects IVW 0.76 (0.62-0.93) p=0,006 | Leave-one-out analysis, Weighted median, MR-Egger, MR-PRESSO, MR Steiger, Weighted mode |
| | | Driving time UK Biobank MRC-IEU ukb-b-3793 | European 310555 | 5 | | | | | Random-effects IVW 1.01 (0.62-1.65) p=0,975 | |
| Chen 2023 (37148539)[103] | Two-sample; summary-level data | Driving time UK Biobank | European 408815 | 4 | BCAC, DRIVE | European | 122977/105974 | Breast cancer<br><br>Per 1 SD | Fixed effect IVW 0.70 (0.32-1.52) p>0.05 | Weighted median, MR-Egger, MR-PRESSO, Weighted mode |
| | | | | | | | 69501 /105974 | Breast cancer, ER+ | Fixed effect IVW 0.59 (0.26-1.30) p>0.05 | |
| | | | | | | | 21468 /105974 | Breast cancer, ER- | Fixed effect IVW 0.90 (0.26-3.07) p>0.05 | |
| | | | | 5 | UK Biobank | N/A | 1889/461044 | Cervical cancer | Fixed effect IVW 0.99 (0.99-1.00) p>0.05 | |
| Went 2024 (38527997)[107] | Two-sample; summary-level data | Sedentary behaviour at work 51 studies | European 372605 | 4 | ECAC, E2C2, UKB | European | 8758/46126 | Endometrial cancer<br><br>Per 1 SD | Random-effects IVW 0.88 (0.54-1.43) p=0,602 | Leave-one-out analysis, Weighted median, MR-Egger, MR-PRESSO, MR Steiger, Weighted mode |
| | | Driving time UK Biobank MRC-IEU ukb-b-3793 | | 6 | | | | | Random-effects IVW 1.96 (0.51-7.59) p=0,331 | |
| Chen 2023 (37148539)[103] | Two-sample; summary-level data | Driving time UK Biobank | European 408815 | 5 | ECAC, E2C2, UKB | European | 12906/108979 | Endometrial cancer<br><br>Per 1 SD | Fixed effect IVW 0.65 (0.17-2.52) p>0.05 | Weighted median, MR-Egger, MR-PRESSO, Weighted mode |

| First author, Year, PMID | MR design | Exposure population source | Exposure population ancestry, sample size | No. IVs (SNPs) | Cancer population source | Cancer population ancestry | Number of cases/controls | Outcome, comparison | Main results OR (95% CI) | Sensitivity analysis |
|---|---|---|---|---|---|---|---|---|---|---|
| | | | | | | | 1230 /35447 | Endometrial cancer, non-endometrioid Per 1 SD | Fixed effect IVW 0.67 (0.04-11.77) p>0.05 | |
| Tan 2024 (38911370)[109] | Two-sample; summary-level data | Driving time UK Biobank | European 408815 | 5 | ECAC, E2C2, UKB | European | 8758 /46126 | Endometrial cancer, endometrioid Per 1 SD | Fixed effect IVW 0.78 (0.24-2.52) p>0.05 | Weighted median, MR-Egger, MR-PRESSO, Weighted mode |
| Went 2024 (38527997)[107] | Two-sample; summary-level data | Sedentary behaviour at work 51 studies | European 372605 | 4 | OCAC, CIMBA | European | 26293 /68502 | Ovarian cancer Per 1 SD | Random-effects IVW 0.92 (0.64-1.32) p=0,654 | Leave-one-out analysis, Weighted median, MR-Egger, MR-PRESSO, MR Steiger, Weighted mode |
| | | Driving time UK Biobank MRC-IEU ukb-b-3793 | European 310555 | 6 | | | | | Random-effects IVW 2.28 (0.84-6.20) p=0,108 | |
| Chen2023 (37148539)[103] | Two-sample; summary-level data | Driving time UK Biobank | European 408815 | 4 | OCAC, CIMBA | European | 25509 /40941 | Ovarian cancer Per 1 SD | Fixed effect IVW 0.71 (0.14-3.74) p>0.05 | Weighted median, MR-Egger, MR-PRESSO, Weighted mode |
| | | | | | | | 2966/40941 | Ovarian cancer, low-grade and low malignant potential serous | Fixed effect IVW 0.45 (0.03-6.22) p>0.05 | |
| | | | | | | | 13037 /40941 | Ovarian cancer, high-grade serous | Fixed effect IVW 0.90 (0.17-4.73) p>0.05 | |
| | | | | | | | 2566/40941 | Ovarian cancer, mucinous | Fixed effect IVW 0.52 (0.06-4.49) p>0.05 | |
| | | | | | | | 2810 /40941 | Ovarian cancer, endometrioid | Fixed effect IVW 1.13 (0.02-84.29) p>0.05 | |
| | | | | | | | 1366 /40941 | Ovarian cancer, clear cell | Fixed effect IVW 1.86 (0.02-171.04) p>0.05 | |
| Went 2024 (38527997)[107] | Two-sample; summary-level data | Sedentary behaviour at work 51 studies | European 372605 | 3 | PRACTICAL | European | 79194/ 61112 | Prostate cancer Per 1 SD | Fixed effect IVW 0.59 (0.21-1.68) p=0,324 | Leave-one-out analysis, Weighted median, MR-Egger, MR-PRESSO, MR |

| First author, Year, PMID | MR design | Exposure population source | Exposure population ancestry, sample size | No. IVs (SNPs) | Cancer population source | Cancer population ancestry | Number of cases/controls | Outcome, comparison | Main results OR (95% CI) | Sensitivity analysis |
|---|---|---|---|---|---|---|---|---|---|---|
| | | Driving time UK Biobank MRC-IEU ukb-b-3793 | European 310555 | 3 | | | | | Random-effects IVW 1.09 (0.46-2.57) p=0,845 | Steiger, Weighted mode |
| Chen 2023 (37148539)[103] | Two-sample; summary-level data | Driving time UK Biobank | European 408815 | 5 | PRACTICAL | European | 79148 /61106 | Prostate cancer Per 1 SD | Fixed effect IVW 1.40 (0.72-2.72) p>0.05 | Weighted median, MR-Egger, MR-PRESSO, Weighted mode |
| Went 2024 (38527997)[107] | Two-sample; summary-level data | Sedentary behaviour at work 51 studies | European 372605 | 4 | GWAS meta-analysis | European | 10784 /20406 | Kidney cancer Per 1 SD | Random-effects IVW 0.78 (0.47-1.28) p=0,319 | Leave-one-out analysis, Weighted median, MR-Egger, MR-PRESSO, MR Steiger, Weighted mode |
| | | Driving time UK Biobank MRC-IEU ukb-b-3793 | European 310555 | 6 | | | | | Random-effects IVW 0.77 (0.17-3.42) p=0,733 | |
| Chen 2023 (37148539)[103] | Two-sample; summary-level data | Driving time UK Biobank | European 408815 | 5 | Finnish Biobank N/A | European | 2168 /216624 | Cancer of urinary organs Per 1 SD | Fixed effect IVW 1.51 (0.22-10.34) p>0.05 | Weighted median, MR-Egger, MR-PRESSO, Weighted mode |
| | | | | 3 | UK Biobank N/A | | 1101/461832 | Bladder cancer Per 1 SD | Fixed effect IVW 1.01 (1.00-1.02) p>0.05 | |
| | | | | 3 | Italian GWAS | | 649/431 | Thyroid cancer Per 1 SD | Fixed effect IVW 0.01 (0.00-12.16) p>0.05 | |
| Tan 2024 (38911370)[109] | Two-sample; summary-level data | Driving time UK Biobank | European 408815 | 4 | FinnGen | European | 1181/324650 | Follicular lymphoma Per 1 SD | IVW 31.799 (1.848-547.031) p=0,017 | Leave-one-out analysis, weighted median, MR-Egger, MR-PRESSO, weighted mode, simple mode |

#### Supplementary Table 17: Descriptive table of the included Mendelian randomisation analyses of screen time (computer use, gaming, television watching) and cancer risk.

| First author, Year, PMID | MR design | Exposure population source | Exposure population ancestry, sample size | No. IVs (SNPs) | Cancer population source | Cancer population ancestry | Number of cases/controls | Outcome, comparison | Main results OR (95% CI) | Sensitivity analysis |
|---|---|---|---|---|---|---|---|---|---|---|
| Chen 2023 (37148539)[103] | Two-sample; summary-level data | Leisure computer use time UK Biobank | European 408815 | 46 | INHANCE | European | 2497/ 2928 | Oral cavity and pharyngeal cancer Per 1 SD | Fixed effect IVW 0.54 (0.21-1.35) p>0.05 | Weighted median, MR-Egger, MR-PRESSO, Weighted mode |
| | | Television watching time UK Biobank | | 170 | | | | | Fixed effect IVW 1.62 (0.98 - 2.67) p>0.05 | |
| Went 2024 (38527997)[107] | Two-sample; summary-level data | Leisure computer use time UK Biobank MRC-IEU ukb-b-4522 | European 360895 | 72 | GWAS meta-analysis | European | 16790/ 32476 | Oesophageal cancer Per 1 SD | Random-effects IVW 0.68 (0.45-1.01) p=0,057 | Leave-one-out analysis, Weighted median, MR-Egger, MR-PRESSO, MR Steiger, Weighted mode |
| Chen 2023 (37148539)[103] | Two-sample; summary-level data | Leisure computer use time UK Biobank | European 408815 | 48 | Finnish Biobank | European | 232/ 218560 | Oesophageal cancer Per 1 SD | Fixed effect IVW 2.36 (0.34-16.27) p>0.05 | Weighted median, MR-Egger, MR-PRESSO, Weighted mode |
| Chen 2024b (38583262)[108] | Two-sample; summary-level data | Leisure screen time 51 studies | European 526725 | 132 | UK Biobank, FinnGen | European | 1223/ 652955 | Oesophageal cancer Per 1 SD | Random-effects IVW 1.36 (1.00-1.84) p=0,049 | Weighted median, MR-Egger, MR-PRESSO |
| Went 2024 (38527997)[107] | Two-sample; summary-level data | Leisure screen time 51 studies | European 526725 | 73 | GWAS meta-analysis | European | 16790/ 32476 | Oesophageal cancer Per 1 SD | Random-effects IVW 1.03 (0.84-1.26) p=0,800 | Leave-one-out analysis, Weighted median, MR-Egger, MR-PRESSO, MR Steiger, Weighted mode |
| | | Television watching time UK Biobank MRC-IEU ukb-b-5192 | European 437887 | 97 | | | | | Random-effects IVW 1.15 (0.78-1.70) p=0,481 | |
| Chen 2023 (37148539)[103] | Two-sample; summary-level data | Television watching time UK Biobank | European 408815 | 152 | Finnish Biobank | European | 232/ 218560 | Oesophageal cancer Per 1 SD | Fixed effect IVW 1.69 (0.60-4.74) p>0.05 | Weighted median, MR-Egger, MR-PRESSO, Weighted mode |
| Chen 2024b (38583262)[108] | Two-sample; summary-level data | Leisure screen time 51 studies | European 526725 | 132 | UK Biobank, FinnGen | European | 1781/ 652955 | Stomach cancer Per 1 SD | Random-effects IVW 1.06 (0.83-1.34) p=0,64 | Weighted median, MR-Egger, MR-PRESSO |
| Went 2024 (38527997)[107] | Two-sample; summary-level data | Leisure computer use time UK Biobank MRC-IEU ukb-b-4522 | European 360895 | 82 | GWAS meta-analysis | | 74673/ 86854 | Colorectal cancer Per 1 SD | Random-effects IVW 0.74 (0.61-0.91) p=0,004 | Leave-one-out analysis, Weighted median, MR-Egger, MR-PRESSO, MR Steiger, Weighted mode |

| First author, Year, PMID | MR design | Exposure population source | Exposure population ancestry, sample size | No. IVs (SNPs) | Cancer population source | Cancer population ancestry | Number of cases/controls | Outcome, comparison | Main results OR (95% CI) | Sensitivity analysis |
|---|---|---|---|---|---|---|---|---|---|---|
| Papadimitriou 2024 (38155458)[104] | Two-sample; summary-level data | Leisure computer use time UK Biobank | European 408815 | 52 | CORECT, CCFR, GECCO | European | 52775/ 45940 | Colorectal cancer Per 1.2 h/d | Fixed effect IVW 0.90 (0.70-1.13) p=0,33 | Weighted median, MR-Egger, simple mode, MR-PRESSO |
| | | | | | | | N/A/ N/A | Colorectal cancer, males | Fixed effect IVW 0.79 (0.61-1.04) p=0,1 | |
| | | | | | | | | Colorectal cancer, females | Fixed effect IVW 1.02 (0.74-1.40) p=0,89 | |
| | | | | | | | | Colon cancer | Fixed effect IVW 0.90 (0.72-1.14) p=0,42 | |
| Chen 2023 (37148539)[103] | Two-sample; summary-level data | Leisure computer use time UK Biobank | European 408815 | 49 | UK Biobank | European | 2437/ 358757 | Colon cancer Per 1 SD | Fixed effect IVW 1.000 (0.995-1.004) p>0.05 | Weighted median, MR-Egger, MR-PRESSO, Weighted mode |
| Papadimitriou 2024 (38155458)[104] | Two-sample; summary-level data | Leisure computer use time UK Biobank | European 408815 | 52 | CORECT, CCFR, GECCO | European | N/A/ N/A | Rectal cancer Per 1.2 h/d | Fixed effect IVW 0.66 (0.49-0.89) p=0,006 | Weighted median, MR-Egger, simple mode, MR-PRESSO |
| Chen 2024b (38583262)[108] | Two-sample; summary-level data | Leisure screen time 51 studies | European 526725 | 132 | UK Biobank, FinnGen | European | 10020/ 642339 | Colorectal cancer Per 1 SD | Random-effects IVW 0.98 (0.87-1.09) p=0,685 | Weighted median, MR-Egger, MR-PRESSO |
| Went 2024 (38527997)[107] | Two-sample; summary-level data | Leisure screen time 51 studies | European 526725 | 63 | GWAS meta-analysis | European | 74673/ 86854 | Colorectal cancer Per 1 SD | Random-effects IVW 1.02 (0.91-1.14) p=0,729 | Leave-one-out analysis, Weighted median, MR-Egger, MR-PRESSO, MR Steiger, Weighted mode |
| | | Television watching time UK Biobank MRC-IEU ukb-b-5192 | European 437887 | 109 | | | | | Random-effects IVW 1.38 (1.13-1.68) p=0,002 | |
| Papadimitriou 2024 (38155458)[104] | Two-sample; summary-level data | Television watching time UK Biobank | European 408815 | 209 | CORECT, CCFR, GECCO | European | 52775/ 45940 | Colorectal cancer Per 1.5 h/d | Fixed effect IVW 1.32 (1.16-1.49) p=2,0E-05 | Weighted median, MR-Egger, simple mode, MR-PRESSO |
| | | | | | | | N/A/ N/A | Colorectal cancer, males | Fixed effect IVW 1.45 (1.23-1.67) p=5,0E-06 | |
| | | | | | | | N/A/ N/A | Colorectal cancer, females | Fixed effect IVW 1.25 (1.06-1.46) p=0,007 | |
| | | | | | | | N/A/ N/A | Colon cancer | Fixed effect IVW 1.36 (1.19-1.57) p=2,0E-05 | |
| Chen 2023 (37148539)[103] | Two-sample; | Television watching time UK Biobank | European 408815 | 187 | UK Biobank | European | 2437/ 358757 | Colon cancer Per 1 SD | Fixed effect IVW 1.001 (0.998-1.003) p>0.05 | Weighted median, MR-Egger, MR-PRESSO, Weighted mode |

| First author, Year, PMID | MR design | Exposure population source | Exposure population ancestry, sample size | No. IVs (SNPs) | Cancer population source | Cancer population ancestry | Number of cases/controls | Outcome, comparison | Main results OR (95% CI) | Sensitivity analysis |
|---|---|---|---|---|---|---|---|---|---|---|
| | summary-level data | | | | | | | | | |
| Papadimitriou 2024 (38155458)[104] | Two-sample; summary-level data | Television watching time UK Biobank | European 408815 | 209 | CORECT, CCFR, GECCO | European | N/A/ N/A | Rectal cancer Per 1.5 h/d | Fixed effect IVW 1.60 (1.32-1.93) p=2,0E-06 | Weighted median, MR-Egger, simple mode, MR-PRESSO |
| Chen 2023 (37148539)[103] | Two-sample; summary-level data | Leisure computer use time UK Biobank | European 408815 | 48 | Finnish Biobank | European | 304/ 218488 | Liver and intrahepatic bile duct cancer Per 1 SD | Fixed effect IVW 0.71 (0.11-4.80) p>0.05 | Weighted median, MR-Egger, MR-PRESSO, Weighted mode |
| Chen 2024b (38583262)[108] | Two-sample; summary-level data | Leisure screen time 51 studies | European 526725 | 132 | UK Biobank, FinnGen | European | 844/ 652955 | Liver cancer Per 1 SD | Random-effects IVW 1.00 (0.67-1.48) p=0,985 | Weighted median, MR-Egger, MR-PRESSO |
| Chen 2023 (37148539)[103] | Two-sample; summary-level data | Television watching time UK Biobank | European 408815 | 176 | Finnish Biobank | European | 304/ 218488 | Liver and intrahepatic bile duct cancer Per 1 SD | Fixed effect IVW 1.41 (0.58-3.40) p>0.05 | Weighted median, MR-Egger, MR-PRESSO, Weighted mode |
| Gentiluomo 2024 (38425433)[105] | Two-sample; summary-level data | Leisure computer use time UK Biobank | European 408815 | 44 | PanScan I-III-PanC4 | European | 8769/ 7055 | Pancreatic cancer Per 1 SD | Random-effects IVW 0.66 (0.41-1.08) p=0,1 | Leave-one-out analysis, weighted median, MR-Egger, Lasso, MR-PRESSO, contamination mixture |
| Chen 2023 (37148539)[103] | Two-sample; summary-level data | Leisure computer use time UK Biobank | European 408815 | 23 | PanScan1 | European | 1896/ 1939 | Pancreatic cancer Per 1 SD | Fixed effect IVW 0.51 (0.08-3.40) p>0.05 | Weighted median, MR-Egger, MR-PRESSO, Weighted mode |
| Chen 2024b (38583262)[108] | Two-sample; summary-level data | Leisure screen time 51 studies | European 526725 | 132 | UK Biobank, FinnGen | European | 1838/ 652955 | Pancreatic cancer Per 1 SD | Random-effects IVW 1.09 (0.85-1.38) p=0,498 | Weighted median, MR-Egger, MR-PRESSO |
| Gentiluomo 2024 (38425433)[105] | Two-sample; summary-level data | Television watching time UK Biobank | European 408815 | 177 | PanScan I-III-PanC4 | European | 8769/ 7055 | Pancreatic cancer Per 1 h/d | Random-effects IVW 1.32 (1.07-1.63) p=0,01 | Leave-one-out analysis, weighted median, MR-Egger, Lasso, MR-PRESSO, contamination mixture |
| | | Television watching time UK Biobank | | 182 | FinnGen | | 1249/ 259583 | | Random-effects IVW 1.42 (0.98-2.05) p=0,061 | |
| Chen 2023 (37148539)[103] | Two-sample; summary-level data | Television watching time UK Biobank | European 408815 | 86 | PanScan1 | European | 1896/ 1939 | Pancreatic cancer Per 1 SD | Fixed effect IVW 0.73 (0.36-1.50) p>0.05 | Weighted median, MR-Egger, MR-PRESSO, Weighted mode |
| Went 2024 (38527997)[107] | Two-sample; | Leisure computer use time UK Biobank | European 360895 | 77 | LC3, ILCCO meta-analysis | European | 29266/ 56450 | Lung cancer Per 1 SD | Random-effects IVW 0.68 (0.50-0.92) p=0,013 | Leave-one-out analysis, Weighted median, MR- |

| First author, Year, PMID | MR design | Exposure population source | Exposure population ancestry, sample size | No. IVs (SNPs) | Cancer population source | Cancer population ancestry | Number of cases/controls | Outcome, comparison | Main results OR (95% CI) | Sensitivity analysis |
|---|---|---|---|---|---|---|---|---|---|---|
| | summary-level data | MRC-IEU ukb-b-4522 | | | | | | | | Egger, MR-PRESSO, MR Steiger, Weighted mode |
| Peng 2021 (34899835)[100] | Two-sample; summary-level data | Leisure computer use time UK Biobank | European 408815 | 37 | ILCCO meta-analysis | European | 11348/ 15861 | Lung cancer Per 1.2 h/d | IVW 0.81 (0.47-1.40) p=0,461 | |
| Gao 2021 (34777480)[99] | Two-sample; summary-level data | Leisure computer use time UK Biobank | European 408815 | 22 | ILCCO meta-analysis | European | 11348/ 15861 | Lung cancer Per 1.5 h/d | Random-effects IVW (P het<0.1) or fixed effect IVW 0.81 (0.48-1.37) p=0,431 | Leave-one-out analysis, weighted median, MR-Egger, MR-PRESSO, MR-APSS, constrained max likelihood, model averaging (cML-MA) |
| | | | | | | | 3442/ 14894 | Lung cancer, adenocarcinoma | Random-effects IVW (P het<0.1) or fixed effect IVW 0.70 (0.31-1.57) p=0,386 | |
| | | | | | | | 3275/ 15038 | Lung cancer, squamous cell carcinoma | Random-effects IVW (P het<0.1) or fixed effect IVW 0.76 (0.34-1.72) p=0,511 | |
| Went 2024 (38527997)[107] | Two-sample; summary-level data | Leisure screen time 51 studies | European 526725 | 63 | LC3, ILCCO meta-analysis | European | 29266/ 56450 | Lung cancer Per 1 SD | Random-effects IVW 1.31 (1.09-1.59) p=0,004 | Leave-one-out analysis, Weighted median, MR-Egger, MR-PRESSO, MR Steiger, Weighted mode |
| | | Television watching time UK Biobank MRC-IEU ukb-b-5192 | | 105 | | | 29266/ 56450 | Lung cancer | Random-effects IVW 2.22 (1.66-2.96) p=7,4E-08 | |
| | | Television watching time UK Biobank MRC-IEU ukb-b-5192 | | 103 | | | N/A/ N/A | Lung cancer, ever smoked | Random-effects IVW 2.32 (1.70-3.16) p=1,0E-07 | |
| | | | | | | | | Lung cancer, never smoked | Random-effects IVW 1.48 (0.73-3.01) p=0,28 | |
| Chen 2024a (38481241)[106] | Two-sample; summary-level data | Television watching time UK Biobank MRC-IEU ukb-b-5192 | European 437887 | 100 | UK Biobank MRC-IEU ukb-b-4954 | European | 2671/ 372016 | Lung cancer Per 1 SD | IVW 1.009 (1.005-1.014) p=2,9E-05 | Weighted median, MR-Egger |
| Peng 2021 (34899835)[100] | Two-sample; summary-level data | Television watching time UK Biobank | European 408815 | 98 | ILCCO meta-analysis | European | 11348/ 15861 | Lung cancer Per 1.5 h/d | IVW 1.90 (1.44-2.50) p<0.001 | Weighted median, MR-Egger, MR-PRESSO,Weighted mode |

| First author, Year, PMID | MR design | Exposure population source | Exposure population ancestry, sample size | No. IVs (SNPs) | Cancer population source | Cancer population ancestry | Number of cases/controls | Outcome, comparison | Main results OR (95% CI) | Sensitivity analysis |
|---|---|---|---|---|---|---|---|---|---|---|
| Gao 2021 (34777480)[99] | Two-sample; summary-level data | Television watching time UK Biobank | European 408815 | 92 | ILCCO meta-analysis | European | 11348/ 15861 | Lung cancer Per 1.5 h/d | Random-effects IVW (P het<0.1) or fixed effect IVW 1.89 (1.41-2.54) p=2,3E-05 | Leave-one-out analysis, weighted median, MR-Egger, MR-PRESSO, MR-APSS, constrained max likelihood, model averaging (cML-MA) |
| Shen 2021 (34076349)[97] | Two-sample; summary-level data | Television watching time UK Biobank MRC-IEU ukb-b-5192 | European 408815 | 108 | ILCCO meta-analysis | | 11348/ 15861 | Lung cancer Per 1 SD | IVW 1.96 (1.32-2.89) p=7,6E-04 | Leave-one-out analysis, weighted median, MR-Egger, MR-PRESSO |
| Gao 2021 (34777480)[99] | Two-sample; summary-level data | Television watching time UK Biobank | European 408815 | 92 | ILCCO meta-analysis | European | 3442/ 14894 | Lung cancer, adenocarcinoma Per 1.5 h/d | Random-effects IVW (P het<0.1) or fixed effect IVW 1.40 (0.94-2.09) p=0,100 | Leave-one-out analysis, weighted median, MR-Egger, MR-PRESSO, MR-APSS, constrained max likelihood, model averaging (cML-MA) |
| Shen 2021 (34076349)[97] | Two-sample; summary-level data | Television watching time UK Biobank MRC-IEU ukb-b-5192 | European 408815 | 108 | ILCCO meta-analysis | European | N/A/ N/A | Lung cancer, adenocarcinoma Per 1 SD | IVW 1.81 (1.02-3.21) p=0,04 | Leave-one-out analysis, weighted median, MR-Egger, MR-PRESSO |
| Gao 2021 (34777480)[99] | Two-sample; summary-level data | Television watching time UK Biobank | European 408815 | 92 | ILCCO meta-analysis | European | 3275/ 15038 | Lung cancer, squamous cell carcinoma Per 1.5 h/d | Random-effects IVW (P het<0.1) or fixed effect IVW 2.37 (1.58-3.55) p=3,2E-05 | Leave-one-out analysis, weighted median, MR-Egger, MR-PRESSO, MR-APSS, constrained max likelihood, model averaging (cML-MA) |
| Shen 2021 (34076349)[97] | Two-sample; summary-level data | Television watching time UK Biobank MRC-IEU ukb-b-5192 | European 408815 | 108 | ILCCO meta-analysis | European | N/A/ N/A | Lung cancer, squamous cell carcinoma Per 1 SD | IVW 3.03 (1.71-5.35) p<0.001 | Leave-one-out analysis, weighted median, MR-Egger, MR-PRESSO |
| Chen 2023 (37148539)[103] | Two-sample; summary-level data | Leisure computer use time UK Biobank | European 408815 | 48 | UK Biobank | European | 2677/ 334482 | Melanoma Per 1 SD | Fixed effect IVW 1.00 (1.00-1.01) p>0.05<br><br>Fixed effect IVW 0.997 (0.995-1.000) p>0.05 | Weighted median, MR-Egger, MR-PRESSO, Weighted mode |
| Li 2023 (36815291)[102] | Two-sample; summary-level data | Playing computer games UK Biobank MRC-IEU ukb-b-4779 | European 462433 | 46 | UK Biobank | European | 17416/ 375455 | Skin cancer, basal cell carcinoma Per 1 SD | IVW 0.66 (0.33-1.35) p=0,25 | Weighted median, MR-Egger, simple mode, weighted mode |
| Li 2023 (36815291)[102] | Two-sample; summary-level data | Television watching time UK Biobank MRC-IEU ukb-b-5192 | European 437887 | 108 | UK Biobank | European | 17416/ 375455 | Skin cancer, basal cell carcinoma Per 1 SD | IVW 0.84 (0.60-1.17) p=0,3 | Weighted median, MR-Egger, simple mode, weighted mode |

| First author, Year, PMID | MR design | Exposure population source | Exposure population ancestry, sample size | No. IVs (SNPs) | Cancer population source | Cancer population ancestry | Number of cases/controls | Outcome, comparison | Main results OR (95% CI) | Sensitivity analysis |
|---|---|---|---|---|---|---|---|---|---|---|
| Went 2024 (38527997)[107] | Two-sample; summary-level data | Leisure computer use time UK Biobank MRC-IEU ukb-b-4522 | European 360895 | 67 | BCAC | European | 133384/ 113789 | Breast cancer Per 1 SD | Random-effects IVW 1.04 (0.85-1.27) p=0,703 | Leave-one-out analysis, Weighted median, MR-Egger, MR-PRESSO, MR Steiger, Weighted mode |
| Papadimitriou 2024 (38155458)[104] | Two-sample; summary-level data | Leisure computer use time UK Biobank | European 408815 | 52 | BCAC | European | 133384/ 113789 | Breast cancer Per 1.2 h/d | Fixed effect IVW 1.01 (0.84-1.23) p=0,89 | Weighted median, MR-Egger, simple mode, MR-PRESSO |
| Chen 2023 (37148539)[103] | Two-sample; summary-level data | Leisure computer use time UK Biobank | European 408815 | 47 | BCAC, DRIVE | European | 122977/ 105974 | Breast cancer Per 1 SD | Fixed effect IVW 0.94 (0.75-1.17) p>0.05 | Weighted median, MR-Egger, MR-PRESSO, Weighted mode |
| | | | | | | | 69501/ 105974 | Breast cancer, ER+ | Fixed effect IVW 1.00 (0.77-1.30) p>0.05 | |
| | | | | | | | 21468/ 105974 | Breast cancer, ER- | Fixed effect IVW 0.72 (0.55-0.94) p<=0.05 | |
| Papadimitriou 2024 (38155458)[104] | Two-sample; summary-level data | Leisure computer use time UK Biobank | European 408815 | 52 | BCAC | European | N/A/ N/A | Breast cancer, Luminal A Per 1.2 h/d | Fixed effect IVW 1.06 (0.84-1.34) p=0,62 | Weighted median, MR-Egger, simple mode, MR-PRESSO |
| | | | | | | | | Breast cancer, Luminal B | Fixed effect IVW 0.89 (0.58-1.36) p=0,58 | |
| | | | | | | | | Breast cancer, Luminal B HER2 negative | Fixed effect IVW 1.03 (0.76-1.40) p=0,84 | |
| | | | | | | | | Breast cancer, HER2 enriched | Fixed effect IVW 0.67 (0.40-1.13) p=0,13 | |
| | | | | | | | | Breast cancer, Triple-negative | Fixed effect IVW 0.68 (0.50-0.93) p=0,02 | |
| Went 2024 (38527997)[107] | Two-sample; summary-level data | Leisure screen time 51 studies | European 526725 | 67 | BCAC | European | 133384/ 113789 | Breast cancer Per 1 SD | Random-effects IVW 1.17 (1.06-1.31) p=0,003 | Leave-one-out analysis, Weighted median, MR-Egger, MR-PRESSO, MR Steiger, Weighted mode |
| | | Television watching time UK Biobank MRC-IEU ukb-b-5192 | European 437887 | 105 | | | | | Random-effects IVW 1.21 (1.01-1.46) p=0,043 | |
| Papadimitriou 2024 (38155458)[104] | Two-sample; | Television watching time UK Biobank | European 408815 | 209 | BCAC | European | 133384/ 113789 | Breast cancer Per 1.5 h/d | Fixed effect IVW 1.15 (1.05-1.26) p=0,002 | Weighted median, MR-Egger, simple mode, MR-PRESSO |

| First author, Year, PMID | MR design | Exposure population source | Exposure population ancestry, sample size | No. IVs (SNPs) | Cancer population source | Cancer population ancestry | Number of cases/controls | Outcome, comparison | Main results OR (95% CI) | Sensitivity analysis |
|---|---|---|---|---|---|---|---|---|---|---|
| | summary-level data | | | | | | | | | |
| Chen 2023 (37148539)[103] | Two-sample; summary-level data | Television watching time UK Biobank | European 408815 | 178 | BCAC, DRIVE | European | 122977/ 105974 | Breast cancer Per 1 SD | Fixed effect IVW 1.21 (1.09-1.33) p<0.001 | Weighted median, MR-Egger, MR-PRESSO, Weighted mode |
| | | | | | | | 69501/ 105974 | Breast cancer, ER+ | Fixed effect IVW 1.21 (1.07-1.36) p=0,002 | |
| | | | | | | | 21468/ 105974 | Breast cancer, ER- | Fixed effect IVW 1.32 (1.12-1.54) p<0.001 | |
| Papadimitriou 2024 (38155458)[104] | Two-sample; summary-level data | Television watching time UK Biobank | European 408815 | 209 | BCAC | European | N/A/ N/A | Breast cancer, Luminal A Per 1.5 h/d | Fixed effect IVW 1.20 (1.06-1.35) p=0,002 | Weighted median, MR-Egger, simple mode, MR-PRESSO |
| | | | | | | | N/A/ N/A | Breast cancer, Luminal B | Fixed effect IVW 1.14 (0.94-1.38) p=0,19 | |
| | | | | | | | N/A/ N/A | Breast cancer, Luminal B HER2 negative | Fixed effect IVW 1.14 (0.96-1.36) p=0,13 | |
| | | | | | | | N/A/ N/A | Breast cancer, HER2 enriched | Fixed effect IVW 1.21 (0.91-1.60) p=0,19 | |
| | | | | | | | N/A/ N/A | Breast cancer, Triple-negative | Fixed effect IVW 1.16 (0.99-1.35) p=0,06 | |
| Chen 2023 (37148539)[103] | Two-sample; summary-level data | Leisure computer use time UK Biobank | European 408815 | 41 | UK Biobank | European | 1889/ 461044 | Cervical cancer Per 1 SD | Fixed effect IVW 1.00 (1.00-1.01) p=0,208 | Weighted median, MR-Egger, MR-PRESSO, Weighted mode |
| | | Television watching time UK Biobank | | 132 | | | | | Fixed effect IVW 1.003 (1.001-1.005) p<0.05 | |
| Went 2024 (38527997)[107] | Two-sample; summary-level data | Leisure computer use time UK Biobank MRC-IEU ukb-b-4522 | European 360895 | 83 | ECAC, E2C2, UKB | European | 8758/ 46126 | Endometrial cancer Per 1 SD | Random-effects IVW 0.88 (0.61-1.28) p=0,505 | Leave-one-out analysis, Weighted median, MR-Egger, MR-PRESSO, MR Steiger, Weighted mode |
| Chen 2023 (37148539)[103] | Two-sample; summary-level data | Leisure computer use time UK Biobank | European 408815 | 49 | ECAC, E2C2, UKB | European | 12906/ 108979 | Endometrial cancer Per 1 SD | Fixed effect IVW 0.89 (0.63-1.27) p>0.05 | Weighted median, MR-Egger, MR-PRESSO, Weighted mode |
| | | | | | | | 1230/ 35447 | Non-endometrioid | Fixed effect IVW 2.99 (1.19-7.49) p=0,019 | |
| | | | | | | | 8758/ 46126 | Endometrioid | Fixed effect IVW 0.80 (0.52-1.23) p>0.05 | |
| Went 2024 (38527997)[107] | Two-sample; | Leisure screen time 51 studies | European 526725 | 78 | ECAC, E2C2, UKB | European | 8758/ 46126 | Endometrial cancer Per 1 SD | Random-effects IVW 1.33 (1.11-1.59) p=0,002 | Leave-one-out analysis, Weighted median, MR- |

| First author, Year, PMID | MR design | Exposure population source | Exposure population ancestry, sample size | No. IVs (SNPs) | Cancer population source | Cancer population ancestry | Number of cases/controls | Outcome, comparison | Main results OR (95% CI) | Sensitivity analysis |
|---|---|---|---|---|---|---|---|---|---|---|
| | summary-level data | | | | | | | | | Egger, MR-PRESSO, MR Steiger, Weighted mode |
| | | Television watching time UK Biobank MRC-IEU ukb-b-5192 | European 437887 | 111 | | | | | Random-effects IVW 1.41 (0.96-2.08) p=0,082 | |
| Chen 2023 (37148539)[103] | Two-sample; summary-level data | Television watching time UK Biobank | European 408815 | 188 | ECAC, E2C2, UKB | European | 12906/ 108979 | Endometrial cancer Per 1 SD | Fixed effect IVW 1.28 (1.06-1.55) p=0,01 | Weighted median, MR-Egger, MR-PRESSO, Weighted mode |
| | | | | | | | 1230/ 35447 | Non-endometrioid | Fixed effect IVW 1.15 (0.72-1.86) p=0,56 | |
| | | | | | | | 8758/ 46126 | Endometrioid | Fixed effect IVW 1.28 (1.02-1.60) p=0,031 | |
| Went 2024 (38527997)[107] | Two-sample; summary-level data | Leisure computer use time UK Biobank MRC-IEU ukb-b-4522 | European 360895 | 82 | OCAC, CIMBA | European | 26293/ 68502 | Ovarian cancer Per 1 SD | Random-effects IVW 0.89 (0.66-1.20) p=0,703 | Leave-one-out analysis, Weighted median, MR-Egger, MR-PRESSO, MR Steiger, Weighted mode |
| Chen 2023 (37148539)[103] | Two-sample; summary-level data | Leisure computer use time UK Biobank | European 408815 | 47 | OCAC, CIMBA | European | 25509/ 40941 | Ovarian cancer Per 1 SD | Fixed effect IVW 0.97 (0.72-1.30) p>0.05 | Weighted median, MR-Egger, MR-PRESSO, Weighted mode |
| | | | | | | | 2966/ 40941 | Ovarian cancer, low-grade and low malignant potential serous | Fixed effect IVW 0.96 (0.51-1.81) p>0.05 | |
| | | | | | | | 13037/ 40941 | Ovarian cancer, high-grade serous | Fixed effect IVW 0.99 (0.70-1.40) p>0.05 | |
| | | | | | | | 2566/ 40941 | Ovarian cancer, mucinous | Fixed effect IVW 1.03 (0.55-1.94) p>0.05 | |
| | | | | | | | 2810/ 40941 | Ovarian cancer, endometrioid | Fixed effect IVW 0.57 (0.31-1.04) p>0.05 | |
| | | | | | | | 1366/ 40941 | Ovarian cancer, clear cell | Fixed effect IVW 0.61 (0.25-1.47) p>0.05 | |
| Went 2024 (38527997)[107] | Two-sample; summary-level data | Leisure screen time 51 studies | European 526725 | 69 | OCAC, CIMBA | European | 26293/ 68502 | Ovarian cancer Per 1 SD | Random-effects IVW 1.21 (1.02-1.43) p=0,026 | Leave-one-out analysis, Weighted median, MR-Egger, MR-PRESSO, MR Steiger, Weighted mode |

| First author, Year, PMID | MR design | Exposure population source | Exposure population ancestry, sample size | No. IVs (SNPs) | Cancer population source | Cancer population ancestry | Number of cases/controls | Outcome, comparison | Main results OR (95% CI) | Sensitivity analysis |
|---|---|---|---|---|---|---|---|---|---|---|
| | | Television watching time UK Biobank MRC-IEU ukb-b-5192 | European 437887 | 108 | | | | | Random-effects IVW 1.21 (0.91-1.61) p=0,186 | |
| Chen 2023 (37148539)[103] | Two-sample; summary-level data | Television watching time UK Biobank | European 408815 | 178 | OCAC, CIMBA | European | 25509/ 40941 | Ovarian cancer Per 1 SD | Fixed effect IVW 1.02 (0.87-1.20) p=0,78 | Weighted median, MR-Egger, MR-PRESSO, Weighted mode |
| | | | | | | | 2966/ 40941 | Ovarian cancer, low-grade and low malignant potential serous | Fixed effect IVW 1.49 (1.07-2.08) p=0,02 | |
| | | | | | | | 13037/ 40941 | Ovarian cancer, high-grade serous | Fixed effect IVW 1.02 (0.85-1.23) p=0,81 | |
| | | | | | | | 2566/ 40941 | Ovarian cancer, mucinous | Fixed effect IVW 1.19 (0.86-1.65) p=0,3 | |
| | | | | | | | 2810/ 40941 | Ovarian cancer, endometrioid | Fixed effect IVW 0.89 (0.64-1.24) p=0,49 | |
| | | | | | | | 1366/ 40941 | Ovarian cancer, clear cell | Fixed effect IVW 0.88 (0.57-1.37) p=0,57 | |
| Went 2024 (38527997)[107] | Two-sample; summary-level data | Leisure computer use time UK Biobank MRC-IEU ukb-b-4522 | European 360895 | 65 | PRACTICAL | European | 79194/ 61112 | Prostate cancer Per 1 SD | Random-effects IVW 1.02 (0.81-1.29) p=0,842 | Leave-one-out analysis, Weighted median, MR-Egger, MR-PRESSO, MR Steiger, Weighted mode |
| Papadimitriou 2024 (38155458)[104] | Two-sample; summary-level data | Leisure computer use time UK Biobank | European 408815 | 52 | PRACTICAL, GAME-ON/ELLIPSE | European | 79148/ 61106 | Prostate cancer Per 1.2 h/d | Fixed effect IVW 1.08 (0.89-1.34) p=0,42 | Weighted median, MR-Egger, simple mode, MR-PRESSO |
| Chen 2023 (37148539)[103] | Two-sample; summary-level data | Leisure computer use time UK Biobank | European 408815 | 46 | PRACTICAL | European | 79148/ 61106 | Prostate cancer Per 1 SD | Fixed effect IVW 1.07 (0.86-1.33) p>0.05 | Weighted median, MR-Egger, MR-PRESSO, Weighted mode |
| Papadimitriou 2024 (38155458)[104] | Two-sample; summary-level data | Leisure computer use time UK Biobank | European 408815 | 52 | PRACTICAL, GAME-ON/ELLIPSE | European | 15167/ 58308 | Prostate cancer, advanced Per 1.2 h/d | Fixed effect IVW 0.91 (0.69-1.22) p=0,54 | Weighted median, MR-Egger, simple mode, MR-PRESSO |
| Went 2024 (38527997)[107] | Two-sample; summary-level data | Leisure screen time 51 studies | European 526725 | 56 | PRACTICAL | European | 79194/ 61112 | Prostate cancer Per 1 SD | Random-effects IVW 0.97 (0.87-1.07) p=0,543 | Leave-one-out analysis, Weighted median, MR-Egger, MR-PRESSO, MR Steiger, Weighted mode |

| First author, Year, PMID | MR design | Exposure population source | Exposure population ancestry, sample size | No. IVs (SNPs) | Cancer population source | Cancer population ancestry | Number of cases/controls | Outcome, comparison | Main results OR (95% CI) | Sensitivity analysis |
|---|---|---|---|---|---|---|---|---|---|---|
| | | Television watching time UK Biobank MRC-IEU ukb-b-5192 | European 437887 | 82 | | | | | Random-effects IVW 0.79 (0.63-0.99) p=0,040 | |
| Papadimitriou 2024 (38155458)[104] | Two-sample; summary-level data | Television watching time UK Biobank | European 408815 | 209 | PRACTICAL, GAME-ON/ELLIPSE | European | 79148/ 61106 | Prostate cancer Per 1.5 h/d | Fixed effect IVW 0.94 (0.84-1.06) p=0,34 | Weighted median, MR-Egger, simple mode, MR-PRESSO |
| Chen 2023 (37148539)[103] | Two-sample; summary-level data | Television watching time UK Biobank | European 408815 | 176 | PRACTICAL | European | 79148/ 61106 | Prostate cancer Per 1 SD | Fixed effect IVW 0.95 (0.85-1.07) p>0.05 | Weighted median, MR-Egger, MR-PRESSO, Weighted mode |
| Papadimitriou 2024 (38155458)[104] | Two-sample; summary-level data | Television watching time UK Biobank | European 408815 | 209 | PRACTICAL, GAME-ON/ELLIPSE | European | 15167/ 58308 | Prostate cancer, advanced Per 1.5 h/d | Fixed effect IVW 0.95 (0.81-1.13) p=0,59 | Weighted median, MR-Egger, simple mode, MR-PRESSO |
| Chen 2023 (37148539)[103] | Two-sample; summary-level data | Leisure computer use time UK Biobank | European 408815 | 48 | Finnish Biobank | European | 2168/ 216624 | Cancer of urinary organs Per 1 SD | Fixed effect IVW 0.86 (0.42-1.74) p>0.05 | Weighted median, MR-Egger, MR-PRESSO, Weighted mode |
| | | Television watching time UK Biobank | | 176 | | | | Cancer of urinary organs Per 1 SD | Fixed effect IVW 1.10 (0.77-1.58) p>0.05 | |
| Went 2024 (38527997)[107] | Two-sample; summary-level data | Leisure computer use time UK Biobank MRC-IEU ukb-b-4522 | European 360895 | 82 | GWAS meta-analysis | European | 10784/ 20406 | Kidney cancer Per 1 SD | Random-effects IVW 0.96 (0.63-1.46) p=0,853 | Leave-one-out analysis, Weighted median, MR-Egger, MR-PRESSO, MR Steiger, Weighted mode |
| | | Leisure screen time 51 studies | European 526725 | 64 | | | | | Random-effects IVW 1.16 (0.93-1.44) p=0,191 | |
| | | Television watching time UK Biobank MRC-IEU ukb-b-5192 | European 437887 | 108 | | | | | Random-effects IVW 1.34 (0.92-1.96) p=0,124 | |
| Chen 2023 (37148539)[103] | Two-sample; summary-level data | Leisure computer use time UK Biobank | European 408815 | 24 | UK Biobank | European | 1101/ 461832 | Bladder cancer Per 1 SD | Fixed effect IVW 1.001 (0.998-1.004) p>0.05 | Weighted median, MR-Egger, MR-PRESSO, Weighted mode |
| | | Television watching time UK Biobank | | 87 | UK Biobank | | 1101/ 461832 | Bladder cancer Per 1 SD | Fixed effect IVW 0.999 (0.998-1.001) p>0.05 | |
| | | Leisure computer use time UK Biobank | | 18 | GliomaScan | | 1856/ 4955 | Glioma Per 1 SD | Fixed effect IVW 3.23 (0.70-14.91) p>0.05 | |
| | | Television watching time UK Biobank | | 67 | GliomaScan | | 1856/ 4955 | Glioma Per 1 SD | Fixed effect IVW 0.70 (0.31-1.57) p>0.05 | |

| First author, Year, PMID | MR design | Exposure population source | Exposure population ancestry, sample size | No. IVs (SNPs) | Cancer population source | Cancer population ancestry | Number of cases/controls | Outcome, comparison | Main results OR (95% CI) | Sensitivity analysis |
|---|---|---|---|---|---|---|---|---|---|---|
| | | Leisure computer use time UK Biobank | | 31 | Italian GWAS | | 649/ 431 | Thyroid cancer Per 1 SD | Fixed effect IVW 0.1 (0.01-0.86) p>0.05 | |
| | | Television watching time UK Biobank | | 105 | Italian GWAS | | 649/ 431 | Thyroid cancer Per 1 SD | Fixed effect IVW 1.79 (0.53-6.09) p>0.05 | |
| Tan 2024 (38911370)[109] | Two-sample; summary-level data | Television watching time UK Biobank | European 408815 | 72 | FinnGen | European | 846/ 324650 | Hodgkin lymphoma Per 1 SD | IVW 2.187 (1.023-4.674) p=0,043 | Leave-one-out analysis, weighted median, MR-Egger, MR-PRESSO, weighted mode, simple mode |
| | | Leisure computer use time UK Biobank | | 20 | | | 1050/ 314193 | Diffuse large B-cell lymphoma Per 1 SD | IVW 0.244 (0.065-0.915) p=0,036 | |
| | | Television watching time UK Biobank | | 72 | | | 1050/ 314193 | Diffuse large B-cell lymphoma Per 1 SD | IVW 4.048 (1.688-9.708) p=0,002 | |

#### Summary forestplots

##### Supplementary Figure 1: Summary forest plot of the linear dose-response associations of sedentary time and cancer incidence and mortality.

**Sedentary time per 2 hours/day and cancer risk**

| Exposure, cancer | Studies | Cases | RR (95% CI) | $I^2$(%) |
|---|---|---|---|---|
| **Sitting time, total** | | | | |
| Breast | 4 | 3,658 | 1.04 (1.01 - 1.07)* | 0 |
| Colon | 3 | 2,396 | 1.03 (1.01 - 1.06)* | 0 |
| Ovarian | 3 | 1,347 | 1.03 (0.98 - 1.09) | 0 |
| Breast, Postmenopausal | 5 | 8,627 | 1.03 (1.00 - 1.05)* | 0 |
| Breast, Premenopausal | 3 | 1,362 | 1.02 (0.98 - 1.07) | 0 |
| Lung | 3 | 2,084 | 1.02 (0.97 - 1.07) | 28 |
| Colorectal | 3 | 5,561 | 1.01 (0.98 - 1.03) | 35 |
| Rectal | 2 | 535 | 0.99 (0.93 - 1.05) | 0 |
| **Sitting time, occupational** | | | | |
| Ovarian | 3 | 908 | 1.25 (1.12 - 1.40)* | 4 |
| Colon | 2 | 1,554 | 1.08 (1.02 - 1.15)* | 28 |
| Breast | 4 | 3,334 | 1.05 (1.01 - 1.09)* | 0 |
| Colorectal | 5 | 4,352 | 1.03 (0.99 - 1.07) | 24 |
| Pancreatic | 4 | 580 | 1.01 (0.91 - 1.12) | 62 |
| Breast, Postmenopausal | 2 | 1,720 | 1.00 (0.90 - 1.10) | 76 |
| Prostate | 2 | 1,648 | 0.97 (0.93 - 1.03) | 0 |
| Rectal | 2 | 648 | 0.97 (0.91 - 1.04) | 0 |
| **Television watching time** | | | | |
| Prostate, mortality | 2 | 993 | 1.10 (0.98 - 1.22) | 39 |
| Stomach | 2 | 888 | 1.08 (1.00 - 1.18) | 0 |
| Colon | 5 | 5,553 | 1.08 (1.05 - 1.11)* | 0 |
| Colorectal, mortality | 2 | 1,438 | 1.07 (0.96 - 1.19) | 57 |
| Endometrial | 3 | 1,600 | 1.05 (0.89 - 1.25) | 82 |
| Lung | 3 | 3,079 | 1.05 (1.01 - 1.10)* | 0 |
| Colorectal | 6 | 11,619 | 1.03 (1.01 - 1.06)* | 0 |
| Breast | 6 | 9,562 | 1.03 (0.98 - 1.08) | 28 |
| Breast, Postmenopausal | 6 | 7,572 | 1.02 (0.99 - 1.04) | 0 |
| Rectal | 4 | 2,284 | 1.01 (0.95 - 1.08) | 0 |
| Breast, Premenopausal | 5 | 2,629 | 1.01 (0.91 - 1.13) | 50 |
| Kidney | 2 | 1,985 | 1.01 (0.95 - 1.07) | 0 |
| Ovarian | 5 | 1,984 | 1.00 (0.94 - 1.07) | 0 |
| Prostate | 2 | 19,730 | 0.99 (0.98 - 1.01) | 0 |
| Oesophageal | 2 | 1,046 | 0.92 (0.76 - 1.13) | 84 |
| **Sedentary time (mixed definitions)** | | | | |
| Ovarian | 7 | 2,646 | 1.06 (1.01 - 1.10)* | 0 |
| Colon | 9 | 8,834 | 1.05 (1.03 - 1.07)* | 0 |
| Endometrial | 5 | 2,037 | 1.04 (0.95 - 1.15) | 74 |
| Lung | 7 | 7,631 | 1.04 (1.00 - 1.09)* | 69 |
| Liver † ‡ | 3 | 459 | 1.04 (0.91 - 1.19) | 65 |
| Breast | 11 | 17,433 | 1.03 (1.02 - 1.05)* | 10 |
| Breast, Premenopausal | 7 | 2,870 | 1.03 (0.99 - 1.08) | 20 |
| Bladder † | 3 | 3,000 | 1.03 (0.97 - 1.09) | 28 |
| Colorectal | 11 | 18,136 | 1.02 (1.00 - 1.04) | 47 |
| Melanoma † | 2 | 3,162 | 1.02 (0.99 - 1.05) | 0 |
| Breast, Postmenopausal | 11 | 17,554 | 1.02 (1.00 - 1.04)* | 7 |
| Pancreatic | 6 | 1,005 | 1.01 (0.96 - 1.07) | 41 |
| Stomach | 4 | 1,241 | 1.01 (0.95 - 1.07) | 54 |
| Kidney | 4 | 2,376 | 1.01 (0.97 - 1.05) | 0 |
| Oesophageal | 4 | 1,424 | 1.00 (0.96 - 1.05) | 0 |
| Rectal | 8 | 3,467 | 0.99 (0.96 - 1.03) | 0 |
| Prostate | 5 | 32,319 | 0.99 (0.98 - 1.00) | 0 |
| Prostate, advanced † | 2 | 3,070 | 0.98 (0.95 - 1.01) | 0 |

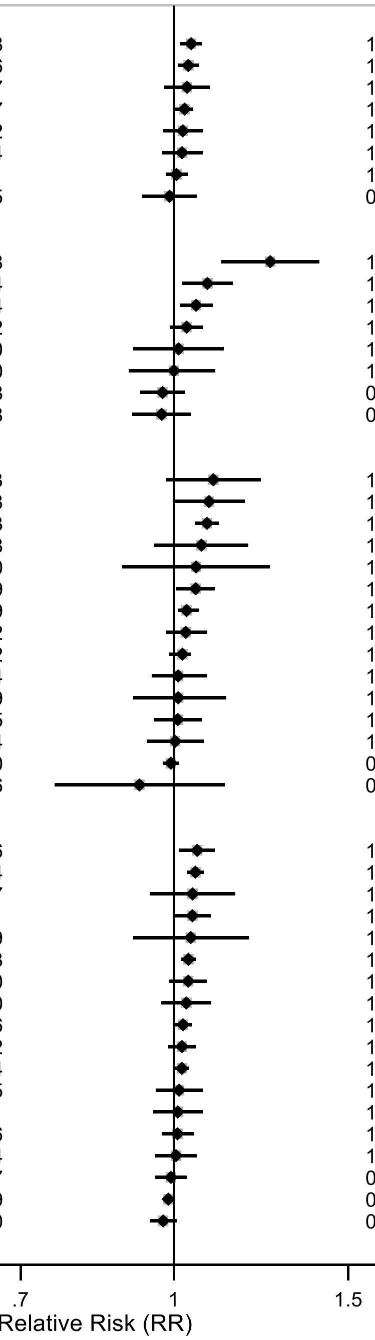

* $p < 0.05$

Forest plot shows the summary results across the linear dose-response (per 2 hours/day of sedentary time) inverse variance DerSimonian-Laird random-effects model meta-analyses. Each square and the horizontal line across the square represent the relative risk (RR) estimate of the corresponding meta-analysis and its 95% confidence interval (CI).

Note: † Indicates exposure–cancer pairs for which sufficient data for meta-analyses were only available after the combination of sedentary time definitions. These analyses were not possible when restricting to specific, non-mixed definitions of sitting or sedentary time. ‡ Includes one study on liver cancer mortality.

Supplementary Materials

#### Supplementary Figure 2: Summary forest plot of the categorical high versus low associations of sedentary time and cancer incidence and mortality.

**High versus low levels of sedentary time and cancer risk**

| Exposure, cancer | Studies | Cases | RR (95% CI) | $I^2$(%) |
|---|---|---|---|---|
| **Sitting time, total** | | | | |
| Breast | 6 | 4,283 | 1.17 (1.02 - 1.35)* | 38.7 |
| Prostate | 2 | 14,640 | 1.08 (0.87 - 1.34) | 84.9 |
| Colorectal | 4 | 3,688 | 1.02 (0.91 - 1.14) | 33.8 |
| **Television watching time** | | | | |
| Endometrial | 4 | 1,799 | 1.30 (0.69 - 2.46) | 86.3 |
| Lung | 4 | 3,466 | 1.26 (1.15 - 1.38)* | 0 |
| Breast, mortality | 2 | 775 | 1.16 (0.85 - 1.61) | 0 |
| Bladder | 2 | 2,386 | 1.15 (0.90 - 1.46) | 0 |
| **Sedentary time (mixed definitions)** | | | | |
| Bladder | 4 | 4,719 | 1.09 (0.97 - 1.22) | 0 |

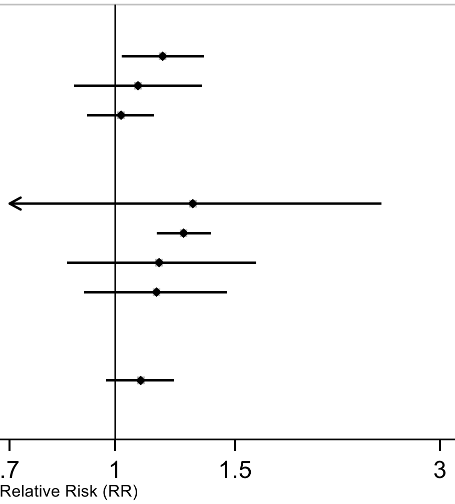

* $p < 0.05$

Forest plot shows the summary results across the highest versus the lowest category inverse variance DerSimonian-Laird random-effects model meta-analyses. Each square and the horizontal line across the square represent the relative risk (RR) estimate of the corresponding meta-analysis and its 95% confidence interval (CI).

Note: A categorical high versus low meta-analysis is conducted when a linear dose-response meta-analysis was not possible (total sitting time and prostate cancer risk and television watching time and bladder cancer risk and breast cancer mortality) or a third or more of the studies could not be included in the linear dose-response meta-analysis (total sitting time and breast and colorectal cancer risk and television watching time and endometrial and lung cancer risk). † Indicates exposure–cancer pairs for which sufficient data for meta-analyses were only available after the combination of sedentary time definitions. These analyses were not possible when restricting to specific, non-mixed definitions of sitting or sedentary time.

#### Supplementary Figure 3: Summary forest plot of the associations of sedentary time and colorectal cancer incidence and mortality.

**Sedentary time and colorectal cancer risk**

| Exposure | Contrast | Studies | Cases | RR (95% CI) | $I^2$(%) |
|---|---|---|---|---|---|
| **Sitting time, total** | | | | | |
| Colon | 2 h/day | 3 | 2,396 | 1.03 (1.01 - 1.06)* | 0 |
| Colorectal | High vs low | 4 | 3,688 | 1.02 (0.91 - 1.14) | 34 |
| Colorectal | 2 h/day | 3 | 5,561 | 1.01 (0.98 - 1.03) | 35 |
| Rectal | 2 h/day | 2 | 535 | 0.99 (0.93 - 1.05) | 0 |
| **Sitting time, occupational** | | | | | |
| Colon | 2 h/day | 2 | 1,554 | 1.08 (1.02 - 1.15)* | 28 |
| Colorectal | 2 h/day | 5 | 4,352 | 1.03 (0.99 - 1.07) | 24 |
| Rectal | 2 h/day | 2 | 648 | 0.97 (0.91 - 1.04) | 0 |
| **Television watching time** | | | | | |
| Colon | 2 h/day | 5 | 5,553 | 1.08 (1.05 - 1.11)* | 0 |
| Colorectal, mortality | 2 h/day | 2 | 1,438 | 1.07 (0.96 - 1.19) | 57 |
| Colorectal | 2 h/day | 6 | 11,619 | 1.03 (1.01 - 1.06)* | 0 |
| Rectal | 2 h/day | 4 | 2,284 | 1.01 (0.95 - 1.08) | 0 |
| **Sedentary time (mixed definitions)** | | | | | |
| Colon | 2 h/day | 9 | 8,834 | 1.05 (1.03 - 1.07)* | 0 |
| Colorectal | 2 h/day | 11 | 18,136 | 1.02 (1.00 - 1.04) | 47 |
| Rectal | 2 h/day | 8 | 3,467 | 0.99 (0.96 - 1.03) | 0 |

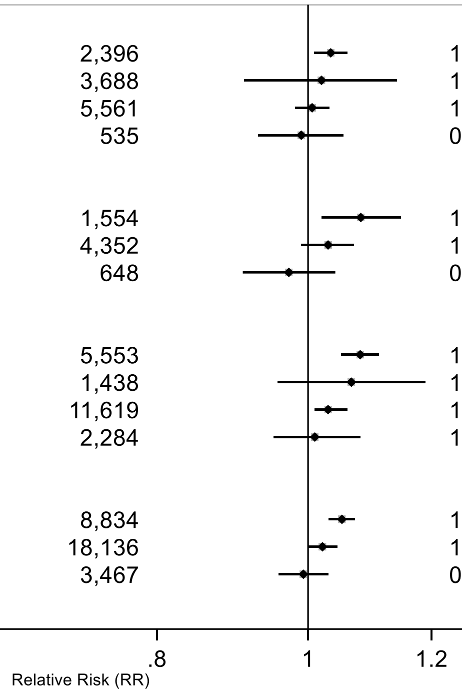

* p < 0.05

Figure legend: Forest plot shows the summary results across the linear dose-response (per 2 hours/day of sedentary time) or categorical high versus low inverse variance DerSimonian-Laird random-effects model meta-analyses. Each square and the horizontal line across the square represent the relative risk (RR) estimate of the corresponding meta-analysis and its 95% confidence interval (CI). Note: A categorical high versus low meta-analysis is conducted when a linear dose-response meta-analysis was not possible or a third or more of the studies could not be included in the linear dose-response meta-analysis.

#### Supplementary Figure 4: Summary forest plot of the associations of sedentary time and breast cancer incidence and mortality.

**Sedentary time and breast cancer risk**

| Exposure | Contrast | Studies | Cases | RR (95% CI) | $I^2$(%) |
|---|---|---|---|---|---|
| **Sitting time, total** | | | | | |
| Breast | High vs low | 6 | 4,283 | 1.17 (1.02 - 1.35)* | 39 |
| Breast | 2 h/day | 4 | 3,658 | 1.04 (1.01 - 1.07)* | 0 |
| Breast, Postmenopausal | 2 h/day | 5 | 8,627 | 1.03 (1.00 - 1.05)* | 0 |
| Breast, Premenopausal | 2 h/day | 3 | 1,362 | 1.02 (0.98 - 1.07) | 0 |
| **Sitting time, occupational** | | | | | |
| Breast | 2 h/day | 4 | 3,334 | 1.05 (1.01 - 1.09)* | 0 |
| Breast, Postmenopausal | 2 h/day | 2 | 1,720 | 1.00 (0.90 - 1.10) | 76 |
| **Television watching time** | | | | | |
| Breast, mortality | High vs low | 2 | 775 | 1.16 (0.85 - 1.61) | 0 |
| Breast | 2 h/day | 6 | 9,562 | 1.03 (0.98 - 1.08) | 28 |
| Breast, Postmenopausal | 2 h/day | 6 | 7,572 | 1.02 (0.99 - 1.04) | 0 |
| Breast, Premenopausal | 2 h/day | 5 | 2,629 | 1.01 (0.91 - 1.13) | 50 |
| **Sedentary time (mixed definitions)** | | | | | |
| Breast | 2 h/day | 11 | 17,433 | 1.03 (1.02 - 1.05)* | 10 |
| Breast, Premenopausal | 2 h/day | 7 | 2,870 | 1.03 (0.99 - 1.08) | 20 |
| Breast, Postmenopausal | 2 h/day | 11 | 17,554 | 1.02 (1.00 - 1.04)* | 7 |

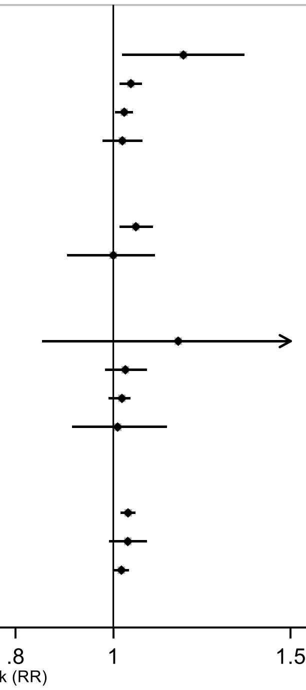

* p < 0.05

Figure legend: Forest plot shows the summary results across the linear dose-response (per 2 hours/day of sedentary time) or categorical high versus low inverse variance DerSimonian-Laird random-effects model meta-analyses. Each square and the horizontal line across the square represent the relative risk (RR) estimate of the corresponding meta-analysis and its 95% confidence interval (CI).

Note: A categorical high versus low meta-analysis is conducted when a linear dose-response meta-analysis was not possible or a third or more of the studies could not be included in the linear dose-response meta-analysis.

#### Supplementary Figure 5: Summary forest plot of the associations of sedentary time and ovarian cancer incidence and mortality.

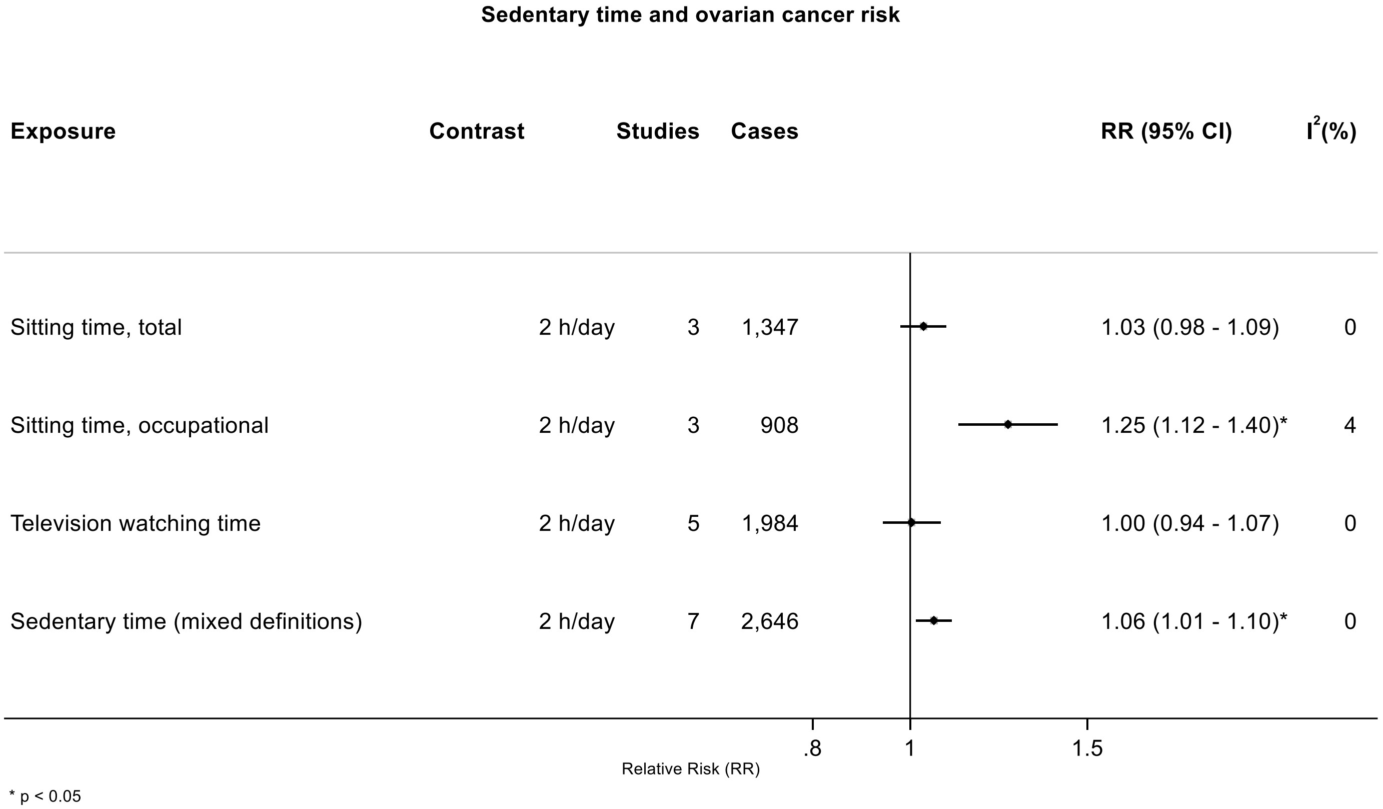

Figure legend: Forest plot shows the summary results across the linear dose-response (per 2 hours/day of sedentary time) or categorical high versus low inverse variance DerSimonian-Laird random-effects model meta-analyses. Each square and the horizontal line across the square represent the relative risk (RR) estimate of the corresponding meta-analysis and its 95% confidence interval (CI).

Note: A categorical high versus low meta-analysis is conducted when a linear dose-response meta-analysis was not possible or a third or more of the studies could not be included in the linear dose-response meta-analysis.

#### Supplementary Figure 6: Summary forest plot of the associations of sedentary time and prostate cancer incidence and mortality.

**Sedentary time and prostate cancer risk**

| Exposure | Contrast | Studies | Cases | RR (95% CI) | $I^2$(%) |
|---|---|---|---|---|---|
| **Sitting time, total** | | | | | |
| Prostate | High vs low | 2 | 14,640 | 1.08 (0.87 - 1.34) | 85 |
| **Sitting time, occupational** | | | | | |
| Prostate | 2 h/day | 2 | 1,648 | 0.97 (0.93 - 1.03) | 0 |
| **Television watching time** | | | | | |
| Prostate, mortality | 2 h/day | 2 | 993 | 1.10 (0.98 - 1.22) | 39 |
| Prostate | 2 h/day | 2 | 19,730 | 0.99 (0.98 - 1.01) | 0 |
| **Sedentary time (mixed definitions)** | | | | | |
| Prostate | 2 h/day | 5 | 32,319 | 0.99 (0.98 - 1.00) | 0 |
| Prostate, advanced † | 2 h/day | 2 | 3,070 | 0.98 (0.95 - 1.01) | 0 |

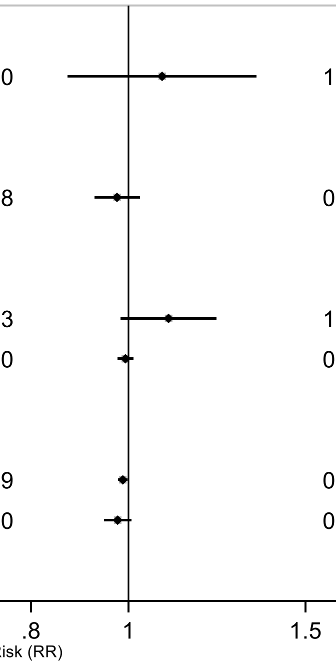

* p < 0.05

Figure legend: Forest plot shows the summary results across the linear dose-response (per 2 hours/day of sedentary time) or categorical high versus low inverse variance DerSimonian-Laird random-effects model meta-analyses. Each square and the horizontal line across the square represent the relative risk (RR) estimate of the corresponding meta-analysis and its 95% confidence interval (CI).

Note: A categorical high versus low meta-analysis is conducted when a linear dose-response meta-analysis was not possible or a third or more of the studies could not be included in the linear dose-response meta-analysis. † Indicates exposure–cancer pairs for which sufficient data for meta-analyses were only available after the combination of sedentary time definitions. These analyses were not possible when restricting to specific, non-mixed definitions of sitting or sedentary time.

#### Supplementary Figure 7: Summary forest plot of the associations of sedentary time and cancer incidence and mortality at other anatomical sites.

**Sedentary time and cancer risk at other anatomical sites**

| Exposure | Contrast | Studies | Cases | RR (95% CI) | $I^2$(%) |
|---|---|---|---|---|---|
| **Bladder** | | | | | |
| Television watching time | 2 h/day | 2 | 2,386 | 1.15 (0.90 - 1.46) | 0 |
| Sedentary time (mixed definitions) † | High vs low | 4 | 4,719 | 1.09 (0.97 - 1.22) | 0 |
| Sedentary time (mixed definitions) † | 2 h/day | 3 | 3,000 | 1.03 (0.97 - 1.09) | 28 |
| **Endometrial** | | | | | |
| Television watching time | High vs low | 4 | 1,799 | 1.30 (0.69 - 2.46) | 86 |
| Television watching time | 2 h/day | 3 | 1,600 | 1.05 (0.89 - 1.25) | 82 |
| Sedentary time (mixed definitions) | 2 h/day | 5 | 2,037 | 1.04 (0.95 - 1.15) | 74 |
| **Kidney** | | | | | |
| Television watching time | 2 h/day | 2 | 1,985 | 1.01 (0.95 - 1.07) | 0 |
| Sedentary time (mixed definitions) | 2 h/day | 4 | 2,376 | 1.01 (0.97 - 1.05) | 0 |
| **Liver** | | | | | |
| Sedentary time (mixed definitions) † ‡ | 2 h/day | 3 | 459 | 1.04 (0.91 - 1.19) | 65 |
| **Lung** | | | | | |
| Sitting time, total | 2 h/day | 3 | 2,084 | 1.02 (0.97 - 1.07) | 28 |
| Television watching time | High vs low | 4 | 3,466 | 1.26 (1.15 - 1.38)* | 0 |
| Television watching time | 2 h/day | 3 | 3,079 | 1.05 (1.01 - 1.10)* | 0 |
| Sedentary time (mixed definitions) | 2 h/day | 7 | 7,631 | 1.04 (1.00 - 1.09)* | 69 |
| **Melanoma** | | | | | |
| Sedentary time (mixed definitions) † | 2 h/day | 2 | 3,162 | 1.02 (0.99 - 1.05) | 0 |
| **Oesophageal** | | | | | |
| Television watching time | 2 h/day | 2 | 1,046 | 0.92 (0.76 - 1.13) | 84 |
| Sedentary time (mixed definitions) | 2 h/day | 4 | 1,424 | 1.00 (0.96 - 1.05) | 0 |
| **Pancreatic** | | | | | |
| Sitting time, occupational | 2 h/day | 4 | 580 | 1.01 (0.91 - 1.12) | 62 |
| Sedentary time (mixed definitions) | 2 h/day | 6 | 1,005 | 1.01 (0.96 - 1.07) | 41 |
| **Stomach** | | | | | |
| Television watching time | 2 h/day | 2 | 888 | 1.08 (1.00 - 1.18) | 0 |
| Sedentary time (mixed definitions) | 2 h/day | 4 | 1,241 | 1.01 (0.95 - 1.07) | 54 |

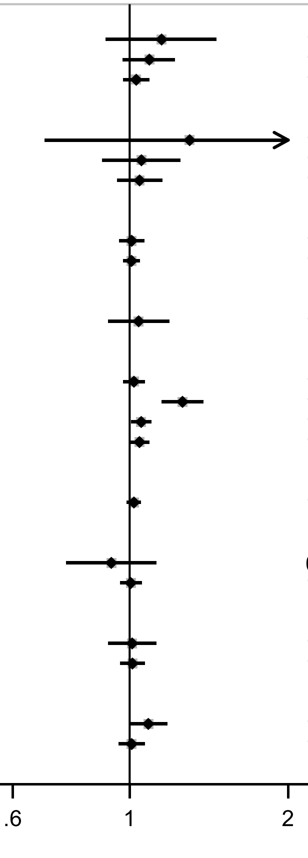

* $p < 0.05$

Figure legend: Forest plot shows the summary results across the linear dose-response (per 2 hours/day of sedentary time) or categorical high versus low inverse variance DerSimonian-Laird random-effects model meta-analyses. Each square and the horizontal line across the square represent the relative risk (RR) estimate of the corresponding meta-analysis and its 95% confidence interval (CI).

Note: A categorical high versus low meta-analysis is conducted when a linear dose-response meta-analysis was not possible or a third or more of the studies could not be included in the linear dose-response meta-analysis. † Indicates exposure–cancer pairs for which sufficient data for meta-analyses were only available after the combination of sedentary time definitions. These analyses were not possible when restricting to specific, non-mixed definitions of sitting or sedentary time. ‡ Includes one study on liver cancer mortality.

#### High versus lowest results plots

##### Supplementary Figure 8: Highest versus the lowest category results from individual studies on sedentary time and oesophageal cancer.

**Sedentary time, highest vs lowest category and oesophageal cancer incidence**

| Author | Study | Outcome | Outcome Type | RR (95% CI) | Comparison |
|---|---|---|---|---|---|
| **Sitting time, total** | | | | | |
| Cook, 2013 | NIH-AARP | Oesophageal, adeno | Incidence | 0.69 (0.41, 1.15) | ≥9 vs ≤3 hours day |
| Cook, 2013 | NIH-AARP | Oesophageal, SCC | Incidence | 0.86 (0.39, 1.88) | ≥9 vs ≤3 hours day |
| **Sitting time, recreational** | | | | | |
| Patel, 2015 | CPS II | Oesophageal | Incidence / Mortality | 1.06 (0.77, 1.46) | ≥6 vs ≤2 hours day |
| **Sitting time, occupational** | | | | | |
| Ihira, 2020 | JPHC | Oesophageal | Incidence | 1.17 (0.68, 2.03) | ≥7 vs <1 hours day |
| **Sitting time, other** | | | | | |
| Ihira, 2020 | JPHC | Oesophageal | Incidence | 0.45 (0.06, 3.53) | ≥7 vs <1 hours day |
| **Screen time, recreational** | | | | | |
| Kunzmann, 2018 | UK Biobank | Oesophageal | Incidence | 1.14 (0.87, 1.49) | 4-16 vs 0-3 hours day |
| Kunzmann, 2018 | UK Biobank | Oesophageal, adeno | Incidence | 1.31 (0.95, 1.79) | 4-16 vs 0-3 hours day |
| Kunzmann, 2018 | UK Biobank | Oesophageal, SCC | Incidence | 0.68 (0.37, 1.26) | 4-16 vs 0-3 hours day |
| **Television watching time** | | | | | |
| Hunter, 2020 | UK Biobank | Oesophageal | Incidence | 1.19 (0.83, 1.69) | >5 vs <1 hours day |
| Cook, 2013 | NIH-AARP | Oesophageal, adeno | Incidence | 0.55 (0.29, 1.02) | >7 vs <1 hours day |
| Cook, 2013 | NIH-AARP | Oesophageal, SCC | Incidence | 0.78 (0.26, 2.32) | >7 vs <1 hours day |

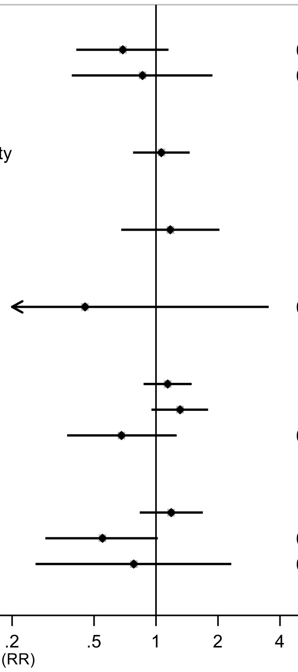

Figure legend: Forest plot shows the highest versus the lowest category results from the individual studies. Each square and the horizontal line across the square represent the relative risk (RR) estimate of the individual study and its 95% confidence interval (CI).

Study abbreviations: NIH-AARP, National Institute of Health + American Association of Retired Persons Diet and Health Study, CPS II, Cancer Prevention Study; JPHC, Japan Public Health Center-Based Prospective Study

Cook 2013 (24367697)[52]; Patel 2015 (26126627)[56]; Ihira 2020 (31925977)[77]; Kunzmann 2018 (30288276)[68]; Hunter 2020 (32746843)[78]

#### Supplementary Figure 9: Highest versus the lowest category results from individual studies on sedentary time and stomach cancer.

**Sedentary time, highest vs lowest category and stomach cancer incidence**

| Author | Study | Outcome | Outcome Type | RR (95% CI) | Comparison |
|---|---|---|---|---|---|
| **Sitting time, total** | | | | | |
| Cook, 2013 | NIH-AARP | Gastric, cardia | Incidence | 1.00 (0.62, 1.63) | ≥9 vs ≤3 hours day |
| Cook, 2013 | NIH-AARP | Gastric, noncardia | Incidence | 0.81 (0.45, 1.45) | ≥9 vs ≤3 hours day |
| **Sitting time, recreational** | | | | | |
| Patel, 2015 | CPS II | Stomach | Incidence / Mortality | 1.03 (0.74, 1.45) | ≥6 vs ≤2 hours day |
| **Sitting time, occupational** | | | | | |
| Ihira, 2020 | JPHC | Stomach | Incidence | 1.10 (0.87, 1.41) | ≥7 vs <1 hours day |
| **Sitting time, other** | | | | | |
| Ihira, 2020 | JPHC | Stomach | Incidence | 1.52 (0.39, 5.92) | ≥7 vs <1 hours day |
| **Screen time, recreational** | | | | | |
| Kunzmann, 2018 | UK Biobank | Stomach | Incidence | 1.13 (0.83, 1.55) | 4-16 vs 0-3 hours day |
| Kunzmann, 2018 | UK Biobank | Gastric, adeno | Incidence | 1.11 (0.79, 1.57) | 4-16 vs 0-3 hours day |
| Kunzmann, 2018 | UK Biobank | Gastric, cardia | Incidence | 0.95 (0.58, 1.57) | 4-16 vs 0-3 hours day |
| Kunzmann, 2018 | UK Biobank | Gastric, noncardia | Incidence | 1.29 (0.86, 1.93) | 4-16 vs 0-3 hours day |
| **Television watching time** | | | | | |
| Hunter, 2020 | UK Biobank | Stomach | Incidence | 1.56 (0.90, 2.71) | >5 vs <1 hours day |
| Cook, 2013 | NIH-AARP | Gastric, cardia | Incidence | 1.37 (0.61, 3.09) | ≥7 vs ≤1 hours day |
| Cook, 2013 | NIH-AARP | Gastric, noncardia | Incidence | 0.93 (0.42, 2.09) | ≥7 vs ≤1 hours day |

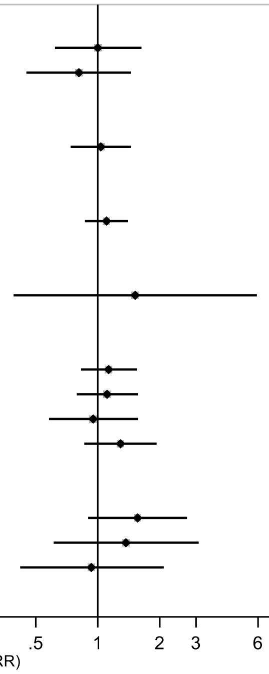

Figure legend: Forest plot shows the highest versus the lowest category results from the individual studies. Each square and the horizontal line across the square represent the relative risk (RR) estimate of the individual study and its 95% confidence interval (CI).

Study abbreviations: NIH-AARP, National Institute of Health + American Association of Retired Persons Diet and Health Study, CPS II, Cancer Prevention Study; JPHC, Japan Public Health Center-Based Prospective Study

Cook 2013 (24367697)[52]; Patel 2015 (26126627)[56]; Ihira 2020 (31925977)[77]; Kunzmann 2018 (30288276)[68]; Hunter 2020 (32746843)[78]

#### Supplementary Figure 10: Highest versus the lowest category results from individual studies on sedentary time and colorectal cancer.

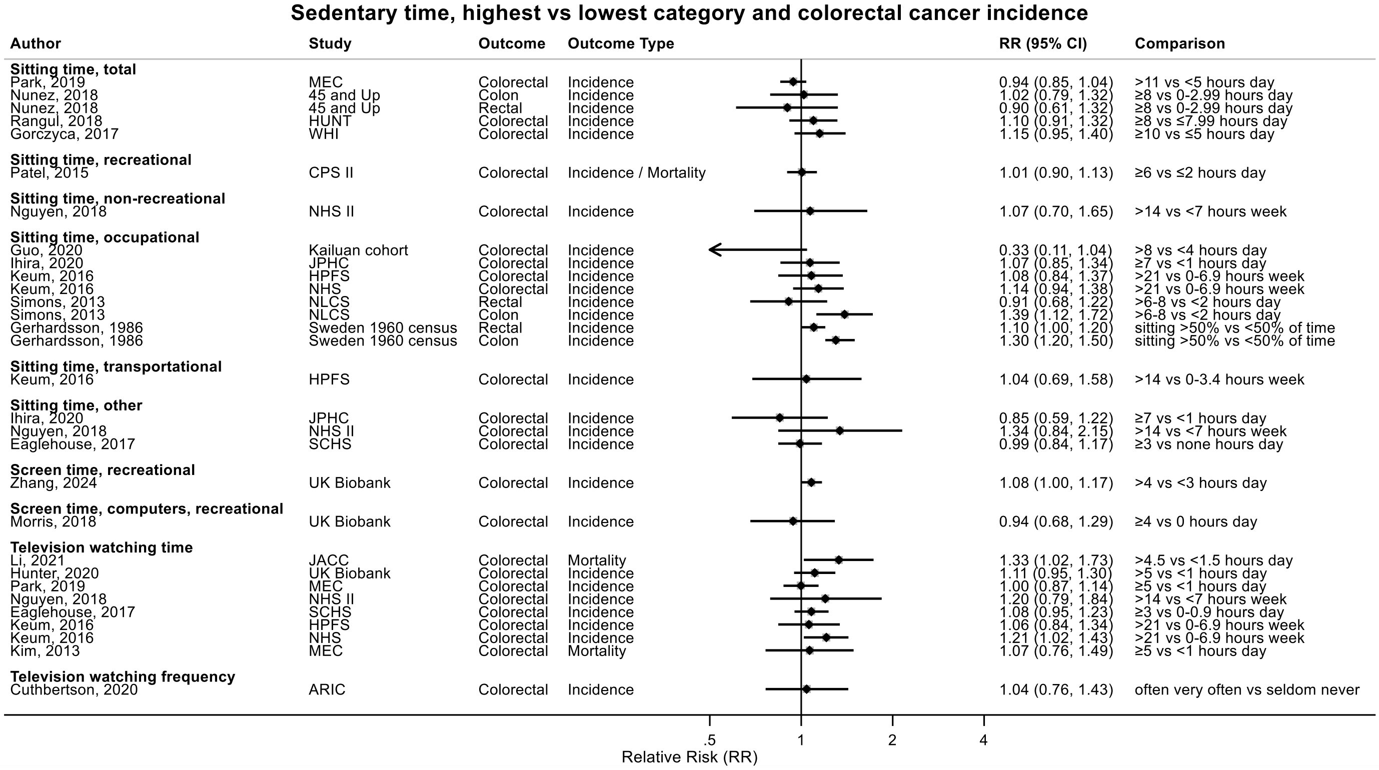

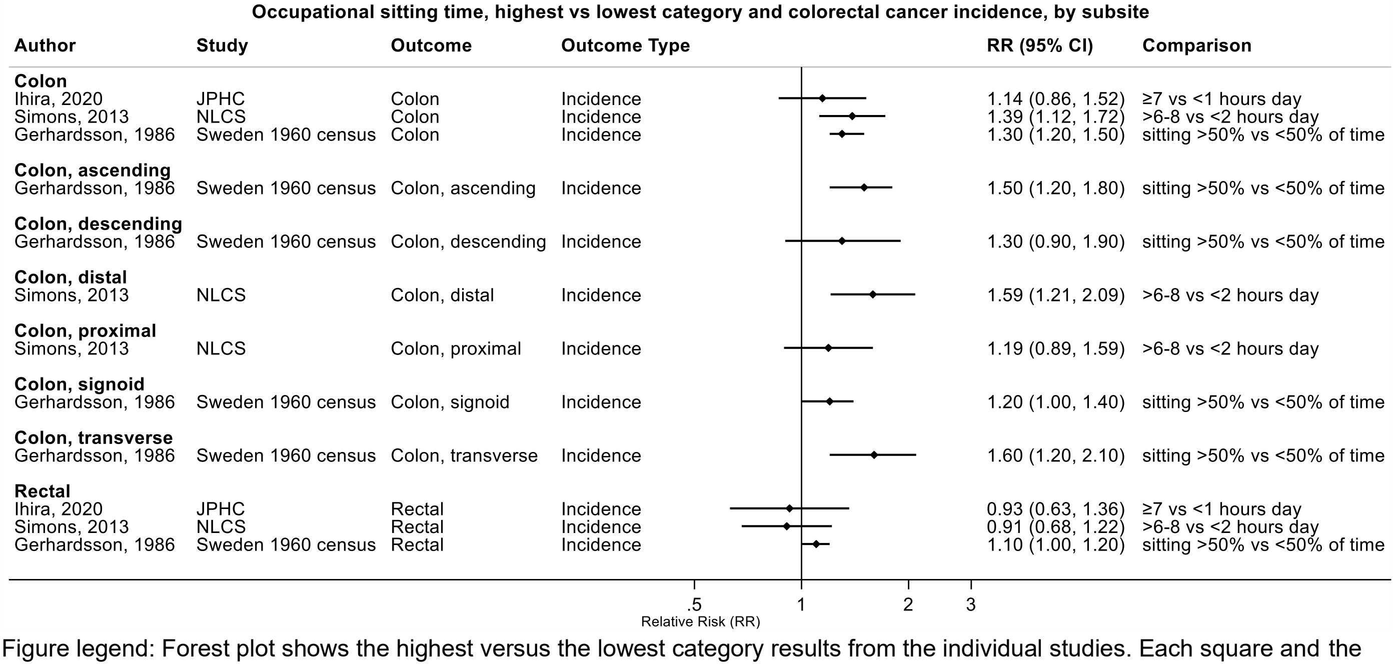

Figure legend: Forest plot shows the highest versus the lowest category results from the individual studies. Each square and the horizontal line across the square represent the relative risk (RR) estimate of the individual study and its 95% confidence interval (CI).

Study abbreviations: CPS II, Cancer Prevention Study; JPHC, Japan Public Health Center-Based Prospective Study; MEC, Multiethnic Cohort Study; NHS, Nurses' Health Study, HPFS, Health Professional Follow-up Study; SCHS, Singapore Chinese Health study; NLCS, Netherlands Cohort Study on Diet and Cancer; HUNT, Trøndelag Health Study, WHI, Women Health Initiative; JACC, Japan Collaborative Cohort study; ARIC, Atherosclerosis Risk in Communities Study

Park 2019 (30910780)[73]; Nunez 2018 (29510753)[66]; Rangul 2018 (30352079)[70]; Gorczyca 2018 (28538039)[61]; Patel 2015 (26126627)[56]; Guo 2020 (31773920)[76]; Ihira 2020 (31925977)[77]; Keum 2016 (26649988)[58]; Simons 2013 (23420352)[45]; Gerhardsson 1986 (3962961)[94]; Eaglehouse 2017 (28542077)[62]; Zhang 2024 (38974470)[93]; Morris 2018 (29520109)[67]; Li 2021 (33138348)[81]; Hunter 2020 (32746843)[78]; Kim 2013 (24062293)[50]; Cuthbertson 2020 (32978174)[80]

#### Supplementary Figure 11: Highest versus the lowest category results from individual studies on sedentary time and liver/hepatobiliary tract cancer.

**Sedentary time, highest vs lowest category and liver cancer incidence**

| Author | Study | Outcome | Outcome Type | RR (95% CI) | Comparison |
|---|---|---|---|---|---|
| **Sitting time, recreational** | | | | | |
| Patel, 2015 | CPS II | Liver | Incidence / Mortality | 0.82 (0.57, 1.19) | ≥6 vs ≤2 hours day |
| **Sitting time, occupational** | | | | | |
| Ihira, 2020 | JPHC | Liver | Incidence | 1.67 (1.00, 2.80) | ≥7 vs <1 hours day |
| **Sitting time, other** | | | | | |
| Ihira, 2020 | JPHC | Liver | Incidence | 0.64 (0.15, 2.78) | ≥7 vs <1 hours day |
| **Television watching time** | | | | | |
| Hunter, 2020 | UK Biobank | Hepatobiliary tract | Incidence | 1.17 (0.83, 1.64) | >5 vs <1 hours day |
| Ukawa, 2014 | JACC | Liver | Mortality | 1.20 (0.82, 1.77) | ≥4 vs ≤1.9 hours day |

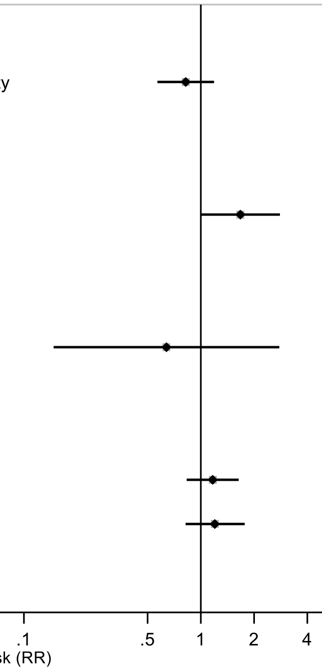

Figure legend: Forest plot shows the highest versus the lowest category results from the individual studies. Each square and the horizontal line across the square represent the relative risk (RR) estimate of the individual study and its 95% confidence interval (CI).

Study abbreviations: CPS II, Cancer Prevention Study; JPHC, Japan Public Health Center-Based Prospective Study; JACC, Japan Collaborative Cohort study

Patel 2015 (26126627)[56]; Ihira 2020 (31925977)[77]; Hunter 2020 (32746843)[78]; Ukawa 2014 (24728669)[54]

#### Supplementary Figure 12: Highest versus the lowest category results from individual studies on sedentary time and pancreatic cancer.

**Sedentary time, highest vs lowest category and pancreatic cancer incidence**

| Author | Study | Outcome | Outcome Type | RR (95% CI) | Comparison |
|---|---|---|---|---|---|
| **Sedentary time, non-occupational** | | | | | |
| Zeng, 2023 | UK Biobank | Pancreas | Incidence | 1.35 (1.12, 1.64) | ≥5 vs <3 hours day |
| **Sitting time, recreational** | | | | | |
| Patel, 2015 | CPS II | Pancreas | Incidence / Mortality | 1.12 (0.91, 1.37) | ≥6 vs ≤2 hours day |
| **Sitting time, occupational** | | | | | |
| Ihira, 2020 | JPHC | Pancreas | Incidence | 1.52 (0.88, 2.61) | ≥7 vs <1 hours day |
| Wu, 2018 | SWHS | Pancreas | Incidence | 0.94 (0.62, 1.45) | ≥6 vs ≤2 hours day |
| Wu, 2018 | SMHS | Pancreas | Incidence | 0.82 (0.50, 1.34) | ≥6 vs ≤2 hours day |
| Heinen, 2011 | NLCS | Pancreas | Incidence | 0.80 (0.52, 1.23) | ≥6 vs ≤2 hours day |
| **Sitting time, other** | | | | | |
| Ihira, 2020 | JPHC | Pancreas | Incidence | 0.68 (0.25, 1.83) | ≥7 vs <1 hours day |
| **Television watching time** | | | | | |
| Hunter, 2020 | UK Biobank | Pancreas | Incidence | 1.03 (0.74, 1.45) | >5 vs <1 hours day |

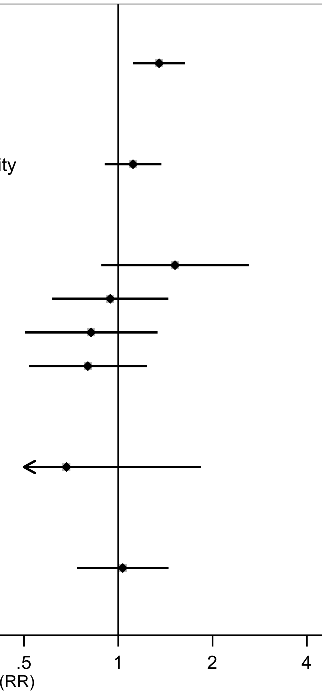

Figure legend: Forest plot shows the highest versus the lowest category results from the individual studies. Each square and the horizontal line across the square represent the relative risk (RR) estimate of the individual study and its 95% confidence interval (CI).

Study abbreviations: CPS II, Cancer Prevention Study; JPHC, Japan Public Health Center-Based Prospective Study; JACC, Japan Collaborative Cohort study; SWHS, Shanghai Women’s Health Study; SMHS, Shanghai Men’s Health Study;  NLCS, Netherlands Cohort Study on Diet and Cancer

Zeng 2023 (38066552)[92]; Patel 2015 (26126627)[56]; Ihira 2020 (31925977)[77]; Wu 2018 (29475964)[65]; Heinen 2011 (21955648)[43]; Hunter 2020 (32746843)[78]

#### Supplementary Figure 13: Highest versus the lowest category results from individual studies on sedentary time and lung cancer.

**Sedentary time, highest vs lowest category and lung cancer incidence**

| Author | Study | Outcome | Outcome Type | RR (95% CI) | Comparison |
|---|---|---|---|---|---|
| **Sitting time, total** | | | | | |
| Jiang, 2019 | HUNT | Lung | Incidence | 1.05 (0.66, 1.29) | >8 vs 0-4 hours day |
| Jiang, 2019 | HUNT | Lung, non-small cell | Incidence | 0.97 (0.74, 1.26) | >8 vs 0-4 hours day |
| Jiang, 2019 | HUNT | Lung, small cell | Incidence | 1.18 (0.69, 2.03) | >8 vs 0-4 hours day |
| Wang, 2016 | WHI | Lung | Mortality | 1.16 (0.96, 1.40) | ≥10 vs ≤5 hours day |
| Wang, 2016 | WHI | Lung | Incidence | 1.10 (0.95, 1.28) | ≥10 vs ≤5 hours day |
| Lam, 2013 | NIH-AARP | Lung | Incidence | 1.28 (0.96, 1.72) | ≥5 vs <3 hours day |
| **Sitting time, recreational** | | | | | |
| Patel, 2015 | CPS II | Lung | Incidence / Mortality | 0.99 (0.89, 1.10) | ≥6 vs ≤2 hours day |
| **Sitting time, occupational** | | | | | |
| Ihira, 2020 | JPHC | Lung | Incidence | 1.03 (0.77, 1.38) | ≥7 vs <1 hours day |
| **Sitting time, other** | | | | | |
| Ihira, 2020 | JPHC | Lung | Incidence | 1.11 (0.64, 1.91) | ≥7 vs <1 hours day |
| **Screen time, recreational** | | | | | |
| Zhang, 2024 | UK Biobank | Lung | Incidence | 1.26 (1.13, 1.40) | >4 vs <3 hours day |
| **Television watching time** | | | | | |
| Bethea, 2022 | BWHS | Lung | Incidence | 1.27 (0.72, 2.21) | ≥1 vs <1 hours week |
| Hunter, 2020 | UK Biobank | Lung | Incidence | 1.28 (1.15, 1.43) | >5 vs <1 hours day |
| Kim, 2013 | MEC | Lung | Mortality | 1.12 (0.91, 1.37) | ≥5 vs <1 hours day |
| Lam, 2013 | NIH-AARP | Lung | Incidence | 1.06 (0.77, 1.46) | ≥5 vs <3 hours day |
| Ukawa, 2013 | JACC | Lung | Incidence | 1.25 (0.99, 1.58) | ≥4 vs <2 hours day |
| **Television watching frequency** | | | | | |
| Cuthbertson, 2020 | ARIC | Lung | Incidence | 0.92 (0.71, 1.18) | often very often vs seldom never |

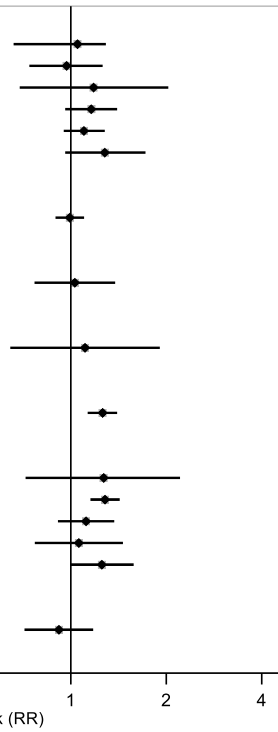

Figure legend: Forest plot shows the highest versus the lowest category results from the individual studies. Each square and the horizontal line across the square represent the relative risk (RR) estimate of the individual study and its 95% confidence interval (CI).

Study abbreviations: HUNT, Trøndelag Health Study; WHI, Women's Health Initiative; NIH-AARP, National Institute of Health + American Association of Retired Persons Diet and Health Study, CPS II, Cancer Prevention Study; JPHC, Japan Public Health Center-Based Prospective Study; BWHS, Black Women Health Study; MEC, Multiethnic Cohort Study; JACC, Japan Collaborative Cohort study; WHI, Women Health Initiative; ARIC, Atherosclerosis Risk in Communities Study

Jiang 2019 (30859092)[72]; Wang 2016 (27439221)[59]; Lam 2013 (23940620)[48]; Patel 2015 (26126627)[56]; Ihira 2020 (31925977)[77]; Zhang 2024 (38974470)[93]; Bethea 2022 (35325667)[87]; Hunter 2020 (32746843)[78]; Kim 2013 (24062293)[50]; Ukawa 2013 (23686441)[47]; Cuthbertson 2020 (32978174)[80]

Supplementary Materials

#### Supplementary Figure 14: Highest versus the lowest category results from individual studies on sedentary time and breast cancer.

**Sedentary time, highest vs lowest category and breast cancer incidence**

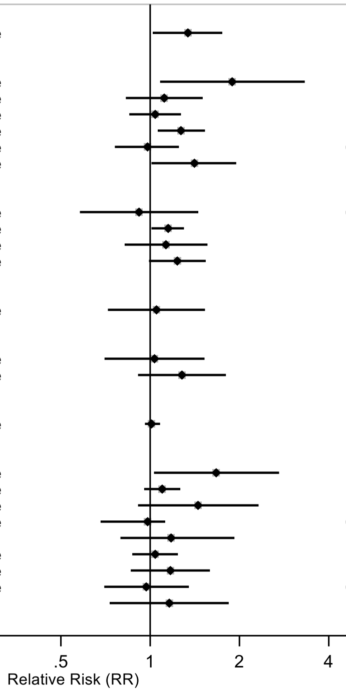

| Author | Study | Outcome | Outcome Type | RR (95% CI) | Comparison |
|---|---|---|---|---|---|
| **Sedentary time, total** | | | | | |
| Tomida, 2024 | J-MICC | Breast | Incidence | 1.34 (1.02, 1.75) | >13 vs <7 hours day |
| **Sitting time, total** | | | | | |
| Wang, 2022 | NHIS | Breast | Incidence | 1.89 (1.08, 3.32) | ≥3 vs 0-2.9 hours day |
| Stahr, 2019 | ARCH | Breast | Incidence | 1.12 (0.83, 1.50) | ≥42 vs ≤28 hours week |
| Rangul, 2018 | HUNT | Breast | Incidence | 1.04 (0.85, 1.27) | ≥8 vs ≤7.99 hours day |
| Nomura, 2016 | BWHS | Breast | Incidence | 1.27 (1.06, 1.53) | ≥10 vs ≤4.9 hours day |
| Catsburg, 2014 | CSDLH | Breast | Incidence | 0.98 (0.76, 1.25) | ≥54.1 vs ≤12.4 hours week |
| Cohen, 2013 | SCCS | Breast | Incidence | 1.41 (1.01, 1.95) | ≥12 vs ≤5.4 hours day |
| **Sitting time, occupational** | | | | | |
| Ihira, 2020 | JPHC | Breast | Incidence | 0.92 (0.58, 1.45) | ≥7 vs <1 hours day |
| Nomura, 2016 | BWHS | Breast | Incidence | 1.15 (1.01, 1.30) | ≥5 vs ≤0.9 hours day |
| Cohen, 2013 | SCCS | Breast | Incidence | 1.13 (0.82, 1.56) | ≥3 vs 0 hours day |
| Pronk, 2011 | SWHS | Breast | Incidence | 1.23 (0.99, 1.54) | ≥4 vs ≤1.2 hours day |
| **Sitting time, transportational** | | | | | |
| Cohen, 2013 | SCCS | Breast | Incidence | 1.05 (0.72, 1.53) | ≥2 vs ≤0.32 hours day |
| **Sitting time, other** | | | | | |
| Ihira, 2020 | JPHC | Breast | Incidence | 1.03 (0.70, 1.52) | ≥7 vs <1 hours day |
| Cohen, 2013 | SCCS | Breast | Incidence | 1.28 (0.91, 1.80) | ≥4 vs ≤0.9 hours day |
| **Screen time, recreational** | | | | | |
| Zhang, 2024 | UK Biobank | Breast | Incidence | 1.01 (0.96, 1.08) | >4 vs <3 hours day |
| **Television watching time** | | | | | |
| Sanchez-Bayona, 2021 | SUN Project | Breast | Incidence | 1.67 (1.03, 2.72) | >2h vs <1h hours day |
| Hunter, 2020 | UK Biobank | Breast | Incidence | 1.10 (0.95, 1.26) | >5 vs <1 hours day |
| Cao, 2019 | JACC | Breast | Incidence | 1.45 (0.91, 2.32) | ≥4.5 vs <1.5 hours day |
| Cifu, 2018 | NIH-AARP | Breast | Incidence | 0.98 (0.68, 1.12) | ≥7 vs 0-0.9 hours day |
| Cifu, 2018 | NIH-AARP | Breast | Mortality | 1.18 (0.79, 1.92) | ≥7 vs 0-0.9 hours day |
| Nomura, 2016 | BWHS | Breast | Incidence | 1.04 (0.87, 1.24) | ≥5 vs ≤0.9 hours day |
| Catsburg, 2014 | CSDLH | Breast | Incidence | 1.17 (0.86, 1.59) | ≥21 vs ≤1 hours week |
| Cohen, 2013 | SCCS | Breast | Incidence | 0.97 (0.70, 1.35) | ≥5 vs ≤1.9 hours day |
| Kim, 2013 | MEC | Breast | Mortality | 1.16 (0.73, 1.84) | ≥5 vs <1 hours day |

Figure legend: Forest plot shows the highest versus the lowest category results from the individual studies. Each square and the horizontal line across the square represent the relative risk (RR) estimate of the individual study and its 95% confidence interval (CI).

Study abbreviations: J-MICC, Japan Multi-institutional Collaborative Cohort; NHIS, National Health Interview Survey; ARCH, Arkansas Rural Community Health Study; HUNT, Trøndelag Health Study; NIH-AARP, National Institute of Health + American Association of Retired Persons Diet and Health Study, JPHC, Japan Public Health Center-Based Prospective Study; BWHS, Black Women Health Study; MEC, Multiethnic Cohort Study; JACC, Japan Collaborative Cohort study; WHI, Women's Health Initiative; CSDLH, Canadian Study of Diet, Lifestyle and Health; SCCS, Southern Community Cohort Study; SWHS, Shanghai Women's Health Study; NIH-AARP, National Institute of Health + American Association of Retired Persons Diet and Health Study

Tomida 2024 (38041484)[91]; Wang 2022a (35159055)[86]; Stahr 2019 (35117114)[85]; Rangul 2018 (30352079)[70]; Nomura 2016 (27632430)[60]; Catsburg 2014 (24781974)[55]; Cohen 2013 (23576427)[46]; Pronk 2011 (21934685)[42]; Ihira 2020 (31925977)[77]; Zhang 2024 (38974470)[93]; Sanchez-Bayona 2021 (33798533)[82]; Hunter 2020 (32746843)[78]; Cao 2019 (30913861)[74]; Cifu 2018 (30309689)[69]; Kim 2013 (24062293)[50]

### Supplementary Figure 15: Highest versus the lowest category results from individual studies on sedentary time and endometrial cancer.

**Sedentary time, highest vs lowest category and endometrial cancer incidence**

| Author | Study | Outcome | Outcome Type | RR (95% CI) | Comparison |
|---|---|---|---|---|---|
| **Sitting time, total** | | | | | |
| Moore, 2010 | NIH-AARP | Endometrial | Incidence | 1.45 (1.10, 1.92) | ≥9 vs ≤2.99 hours day |
| **Sitting time, recreational** | | | | | |
| Patel, 2015 | CPS II | Endometrial | Incidence / Mortality | 1.34 (1.08, 1.67) | ≥6 vs ≤2 hours day |
| **Sitting time, occupational** | | | | | |
| Ihira, 2020 | JPHC | Endometrial | Incidence | 0.36 (0.12, 1.08) | ≥7 vs <1 hours day |
| **Sitting time, other** | | | | | |
| Ihira, 2020 | JPHC | Endometrial | Incidence | 0.70 (0.31, 1.56) | ≥7 vs <1 hours day |
| **Television watching time** | | | | | |
| Miyata, 2021 | JACC | Endometrial | Incidence | 2.10 (0.60, 7.40) | ≥4 vs <1 hours day |
| Hunter, 2020 | UK Biobank | Endometrial | Incidence | 0.59 (0.41, 0.85) | >5 vs <1 hours day |
| Gierach, 2009 | NIH-AARP | Endometrial | Incidence | 1.66 (1.20, 2.28) | ≥7 vs <3 hours day |
| Friberg, 2006 | SMC | Endometrial | Incidence | 1.80 (1.14, 2.83) | ≥5 vs <5 hours day |

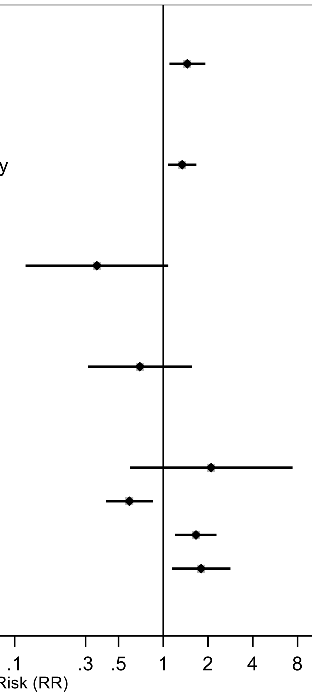

Figure legend: Forest plot shows the highest versus the lowest category results from the individual studies. Each square and the horizontal line across the square represent the relative risk (RR) estimate of the individual study and its 95% confidence interval (CI).

Study abbreviations: CPS II, Cancer Prevention Study; JPHC, Japan Public Health Center-Based Prospective Study; JACC, Japan Collaborative Cohort study; NIH-AARP, National Institute of Health + American Association of Retired Persons Diet and Health Study; SMC, Swedish Mammography Cohort

Moore 2010 (20877336)[40]; Patel 2015 (26126627)[56]; Ihira 2020 (31925977)[77]; Miyata 2021 (32963209)[79]; Hunter 2020 (32746843)[78]; Gierach 2009 (19123463)[38]; Friberg 2006 (17057024)[35]

Adjustment for physical activity and adiposity in models:

Adjusted for physical activity: Miyata, 2021; Ihira, 2020; Patel, 2015; Moore, 2010; Friberg, 2006

Not adjusted for physical activity: Hunter, 2020; Gierach, 2009

Adjusted for adiposity: Miyata, 2021; Hunter, 2020; Ihira, 2020

Not adjusted for adiposity: Patel, 2015; Moore, 2010; Gierach, 2009; Friberg, 2006

#### Supplementary Figure 16: Highest versus the lowest category results from individual studies on sedentary time and ovarian cancer.

**Sedentary time, highest vs lowest category and ovarian cancer incidence**

| Author | Study | Outcome | Outcome Type | RR (95% CI) | Comparison |
|---|---|---|---|---|---|
| **Sitting time, total** | | | | | |
| Buras, 2022 | NHS, NHS II | Ovary | Incidence | 1.09 (0.92, 1.30) | >40 vs <20 hours week |
| Xiao, 2013 | NIH-AARP | Ovary, epithelial | Incidence | 1.06 (0.81, 1.39) | ≥7 vs ≤2.9 hours day |
| **Sitting time, recreational** | | | | | |
| Hildebrand, 2015 | CPS II | Ovary | Incidence / Mortality | 1.44 (1.12, 1.85) | ≥6 vs ≤2.9 hours day |
| Hildebrand, 2015 | CPS II | Ovary, nonserous | Incidence / Mortality | 1.08 (0.57, 2.04) | ≥6 vs ≤2.9 hours day |
| Hildebrand, 2015 | CPS II | Ovary, serous | Incidence / Mortality | 1.52 (1.06, 2.16) | ≥6 vs ≤2.9 hours day |
| **Sitting time, occupational** | | | | | |
| Buras, 2022 | NHS, NHS II | Ovary | Incidence | 1.40 (1.14, 1.71) | >20 vs <5 hours week |
| Ihira, 2020 | JPHC | Ovary | Incidence | 1.36 (0.49, 3.80) | ≥7 vs <1 hours day |
| **Sitting time, other** | | | | | |
| Buras, 2022 | NHS, NHS II | Ovary | Incidence | 0.84 (0.66, 1.08) | >20 vs <5 hours week |
| Ihira, 2020 | JPHC | Ovary | Incidence | 1.05 (0.40, 2.75) | ≥7 vs <1 hours day |
| **Television watching time** | | | | | |
| Buras, 2022 | NHS, NHS II | Ovary | Incidence | 0.96 (0.77, 1.20) | >20 vs <5 hours week |
| Hunter, 2020 | UK Biobank | Ovary | Incidence | 1.03 (0.70, 1.52) | >5 vs <1 hours day |
| Ukawa, 2018 | JACC | Ovary | Incidence | 1.81 (0.75, 4.39) | ≥5 vs ≤1.9 hours day |
| Xiao, 2013 | NIH-AARP | Ovary, epithelial | Incidence | 1.02 (0.67, 1.55) | ≥7 vs ≤2.9 hours day |

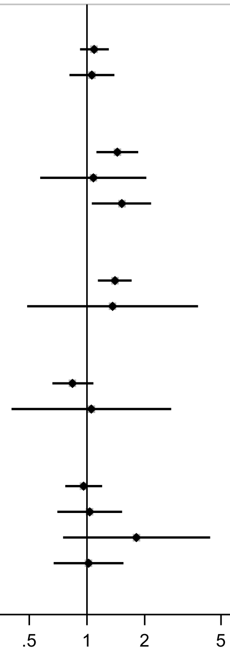

Figure legend: Forest plot shows the highest versus the lowest category results from the individual studies. Each square and the horizontal line across the square represent the relative risk (RR) estimate of the individual study and its 95% confidence interval (CI).

Study abbreviations: CPS II, Cancer Prevention Study; JPHC, Japan Public Health Center-Based Prospective Study; JACC, Japan Collaborative Cohort study; NIH-AARP, National Institute of Health + American Association of Retired Persons Diet and Health Study; NHS, Nurses' Health Study

Buras 2022 (35094053)[84]; Xiao 2013 (23966580)[49]; Hildebrand 2015 (26335264)[57]; Ihira 2020 (31925977)[77]; Hunter 2020 (32746843)[78]; Ukawa 2018 (29340890)[64]

#### Supplementary Figure 17: Highest versus the lowest category results from individual studies on sedentary time and prostate cancer.

**Sedentary time, highest vs lowest category and prostate cancer incidence**

| Author | Study | Outcome | Outcome Type | RR (95% CI) | Comparison |
|---|---|---|---|---|---|
| **Sitting time, total** | | | | | |
| Rangul, 2018 | HUNT | Prostate | Incidence | 1.22 (1.05, 1.42) | ≥8 vs ≤7.99 hours day |
| Lynch, 2014 | NIH-AARP | Prostate | Incidence | 0.98 (0.91, 1.05) | ≥9 vs <3 hours day |
| Lynch, 2014 | NIH-AARP | Prostate | Mortality | 1.07 (0.84, 1.35) | ≥7 vs <3 hours day |
| Lynch, 2014 | NIH-AARP | Prostate, advanced | Incidence | 0.91 (0.77, 1.08) | ≥7 vs <3 hours day |
| **Sitting time, recreational** | | | | | |
| Patel, 2015 | CPS II | Prostate | Incidence / Mortality | 0.96 (0.90, 1.02) | ≥6 vs ≤2 hours day |
| Patel, 2015 | CPS II | Prostate, advanced | Incidence / Mortality | 0.96 (0.85, 1.09) | ≥6 vs ≤2 hours day |
| **Sitting time, occupational** | | | | | |
| Ihira, 2020 | JPHC | Prostate | Incidence | 1.01 (0.77, 1.32) | ≥7 vs <1 hours day |
| Zeegers, 2005 | NLCS | Prostate | Incidence | 0.86 (0.68, 1.10) | 6-8 vs ≤1.9 hours day |
| **Screen time, recreational** | | | | | |
| Zhang, 2024 | UK Biobank | Prostate | Incidence | 1.01 (0.95, 1.07) | >4 vs <3 hours day |
| **Television watching time** | | | | | |
| Hunter, 2020 | UK Biobank | Prostate | Incidence | 0.91 (0.81, 1.03) | >5 vs <1 hours day |
| Lynch, 2014 | NIH-AARP | Prostate | Incidence | 1.03 (0.92, 1.15) | ≥7 vs <1 hours day |
| Lynch, 2014 | NIH-AARP | Prostate | Mortality | 1.07 (0.85, 1.33) | ≥5 vs <3 hours day |
| Lynch, 2014 | NIH-AARP | Prostate, advanced | Incidence | 0.93 (0.79, 1.09) | ≥5 vs <3 hours day |
| Kim, 2013 | MEC | Prostate | Mortality | 1.39 (0.90, 2.15) | ≥5 vs <1 hours day |
| **Television watching frequency** | | | | | |
| Cuthbertson, 2020 | ARIC | Prostate | Incidence | 0.83 (0.68, 1.01) | often very often vs seldom never |

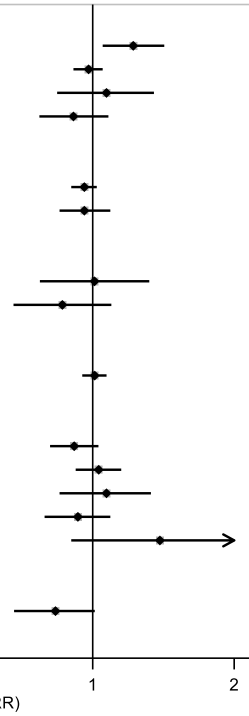

Figure legend: Forest plot shows the highest versus the lowest category results from the individual studies. Each square and the horizontal line across the square represent the relative risk (RR) estimate of the individual study and its 95% confidence interval (CI).

Study abbreviations: HUNT, Trøndelag Health Study; NIH-AARP, National Institute of Health + American Association of Retired Persons Diet and Health Study; CPS II, Cancer Prevention Study; JPHC, Japan Public Health Center-Based Prospective Study; NLCS, Netherlands Cohort Study on Diet and Cancer; MEC, Multiethnic Cohort Study; ARIC, Atherosclerosis Risk in Communities Study

Rangul 2018 (30352079)[70]; Lynch 2014b (24526287)[53]; Patel 2015 (26126627)[56]; Ihira 2020 (31925977)[77]; Zeegers 2005 (15941961)[34]; Zhang 2024 (38974470)[93]; Hunter 2020 (32746843)[78]; Kim 2013 (24062293)[50]; Cuthbertson 2020 (32978174)[80]

**Supplementary Figure 18: Highest versus the lowest category results from individual studies on sedentary time and kidney cancer.**

**Sedentary time, highest vs lowest category and kidney cancer incidence**

| Author | Study | Outcome | Outcome Type | RR (95% CI) | Comparison |
|---|---|---|---|---|---|
| **Sitting time, total** | | | | | |
| George, 2011 | NIH-AARP | Kidney | Incidence | 1.11 (0.87, 1.41) | ≥9 vs ≤2.9 hours day |
| **Sitting time, recreational** | | | | | |
| Patel, 2015 | CPS II | Kidney | Incidence / Mortality | 1.08 (0.84, 1.39) | ≥6 vs ≤2 hours day |
| **Sitting time, occupational** | | | | | |
| Ihira, 2020 | JPHC | Kidney | Incidence | 1.03 (0.50, 2.08) | 5-6 vs <1 hours day |
| **Sitting time, other** | | | | | |
| Ihira, 2020 | JPHC | Kidney | Incidence | 1.84 (0.58, 5.87) | ≥7 vs <1 hours day |
| **Television watching time** | | | | | |
| Hunter, 2020 | UK Biobank | Kidney | Incidence | 1.17 (0.89, 1.56) | >5 vs <1 hours day |
| George, 2011 | NIH-AARP | Kidney | Incidence | 0.96 (0.66, 1.38) | ≥7 vs ≤0.9 hours day |

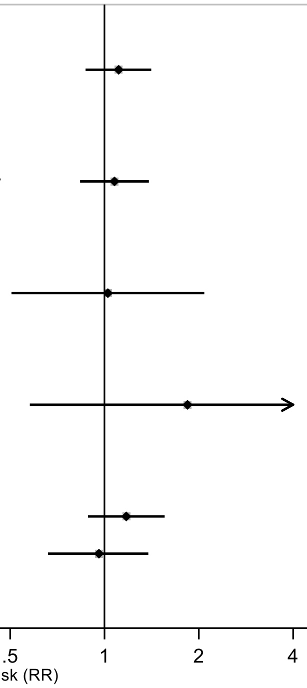

Figure legend: Forest plot shows the highest versus the lowest category results from the individual studies. Each square and the horizontal line across the square represent the relative risk (RR) estimate of the individual study and its 95% confidence interval (CI).

Study abbreviations: NIH-AARP, National Institute of Health + American Association of Retired Persons Diet and Health Study; CPS II, Cancer Prevention Study; JPHC, Japan Public Health Center-Based Prospective Study

George 2011 (21737302)[41]; Patel 2015 (26126627)[56]; Ihira 2020 (31925977)[77]; Hunter 2020 (32746843)[78]

#### Supplementary Figure 19: Highest versus the lowest category results from individual studies on sedentary time and bladder cancer.

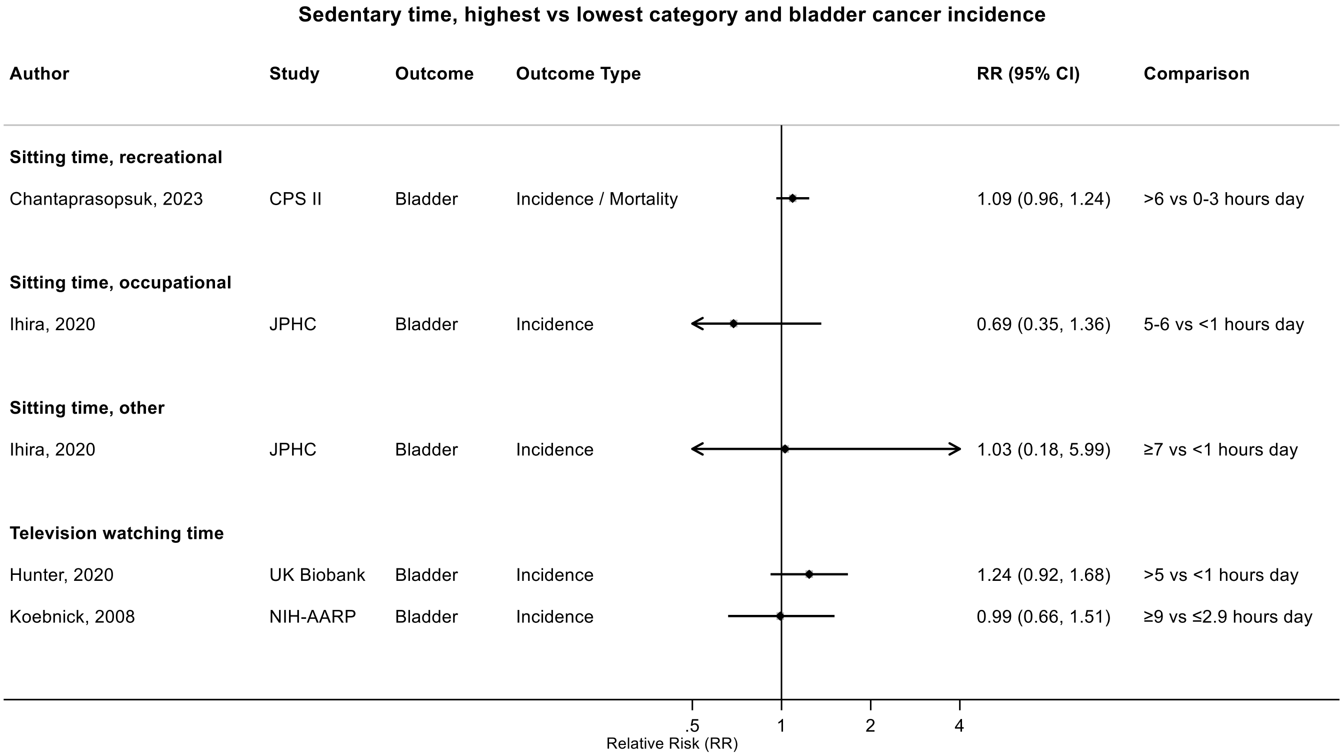

Figure legend: Forest plot shows the highest versus the lowest category results from the individual studies. Each square and the horizontal line across the square represent the relative risk (RR) estimate of the individual study and its 95% confidence interval (CI).

Study abbreviations: NIH-AARP, National Institute of Health + American Association of Retired Persons Diet and Health Study; CPS II, Cancer Prevention Study; JPHC, Japan Public Health Center-Based Prospective Study

Chantaprasopsuk 2023 (37202564)[89]; Ihira 2020 (31925977)[77]; Hunter 2020 (32746843)[78]; Koebnick 2008 (18483344)[37]

### Supplementary Figure 20: Highest versus the lowest category results from individual studies on sedentary time and cancer at other anatomical sites.

**Sedentary time, highest vs lowest category and cancer incidence at other anatomical sites**

| Author | Study | Outcome | Outcome Type | RR (95% CI) | Comparison |
|---|---|---|---|---|---|
| **Sitting time, total** | | | | | |
| Rees-Punia, 2019 | CPS II | Myeloid leukaemia | Incidence / Mortality | 1.28 (0.89, 1.85) | >6 vs <3 hours day |
| Rees-Punia, 2019 | CPS II | Acute myeloid leukaemia | Incidence / Mortality | 1.01 (0.60, 1.64) | >6 vs <3 hours day |
| **Sitting time, recreational** | | | | | |
| Patel, 2015 | CPS II | Gallbladder | Incidence / Mortality | 1.77 (0.99, 3.16) | ≥6 vs ≤2 hours day |
| Patel, 2015 | CPS II | Mouth | Incidence / Mortality | 1.28 (0.96, 1.69) | ≥6 vs ≤2 hours day |
| Patel, 2015 | CPS II | Melanoma | Incidence / Mortality | 1.03 (0.90, 1.18) | ≥6 vs ≤2 hours day |
| Patel, 2015 | CPS II | Multiple Myeloma | Incidence / Mortality | 1.25 (0.94, 1.66) | ≥6 vs ≤2 hours day |
| Teras, 2012 | CPS II | Diffuse large-Bcell lymphoma | Incidence / Mortality | 0.76 (0.55, 1.06) | ≥6 vs ≤2.9 hours day |
| Teras, 2012 | CPS II | Leukaemia (CLL +SLL) | Incidence / Mortality | 1.13 (0.84, 1.52) | ≥6 vs ≤2.9 hours day |
| Teras, 2012 | CPS II | Non-Hodgkin Lymphoma | Incidence / Mortality | 1.07 (0.92, 1.23) | ≥6 vs ≤2 hours day |
| Teras, 2012 | CPS II | Follicular lymphoma | Incidence / Mortality | 1.02 (0.69, 1.51) | ≥6 vs ≤2.9 hours day |
| **Television watching time** | | | | | |
| Hunter, 2020 | UK Biobank | Non-Hodgkin Lymphoma | Incidence | 0.96 (0.73, 1.25) | >5 vs <1 hours day |
| Hunter, 2020 | UK Biobank | Brain | Incidence | 1.10 (0.73, 1.67) | >5 vs <1 hours day |

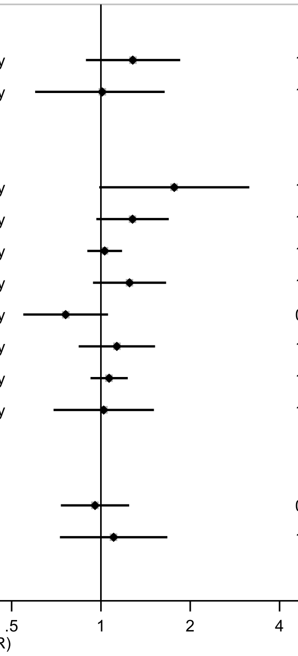

Figure legend: Forest plot shows the highest versus the lowest category results from the individual studies. Each square and the horizontal line across the square represent the relative risk (RR) estimate of the individual study and its 95% confidence interval (CI).

Study abbreviations: CPS II, Cancer Prevention Study

Rees-Punia 2019 (31196856)[75]; Patel 2015 (26126627)[56]; Teras 2012 (22275172)[44]; Hunter 2020 (32746843)[78]

#### Specific definitions of sitting time

##### Supplementary Figure 21: Linear dose-response meta-analysis of the association between total sitting time and colorectal cancer incidence.

**Total sitting time, per 2 hours/day and colorectal cancer incidence**

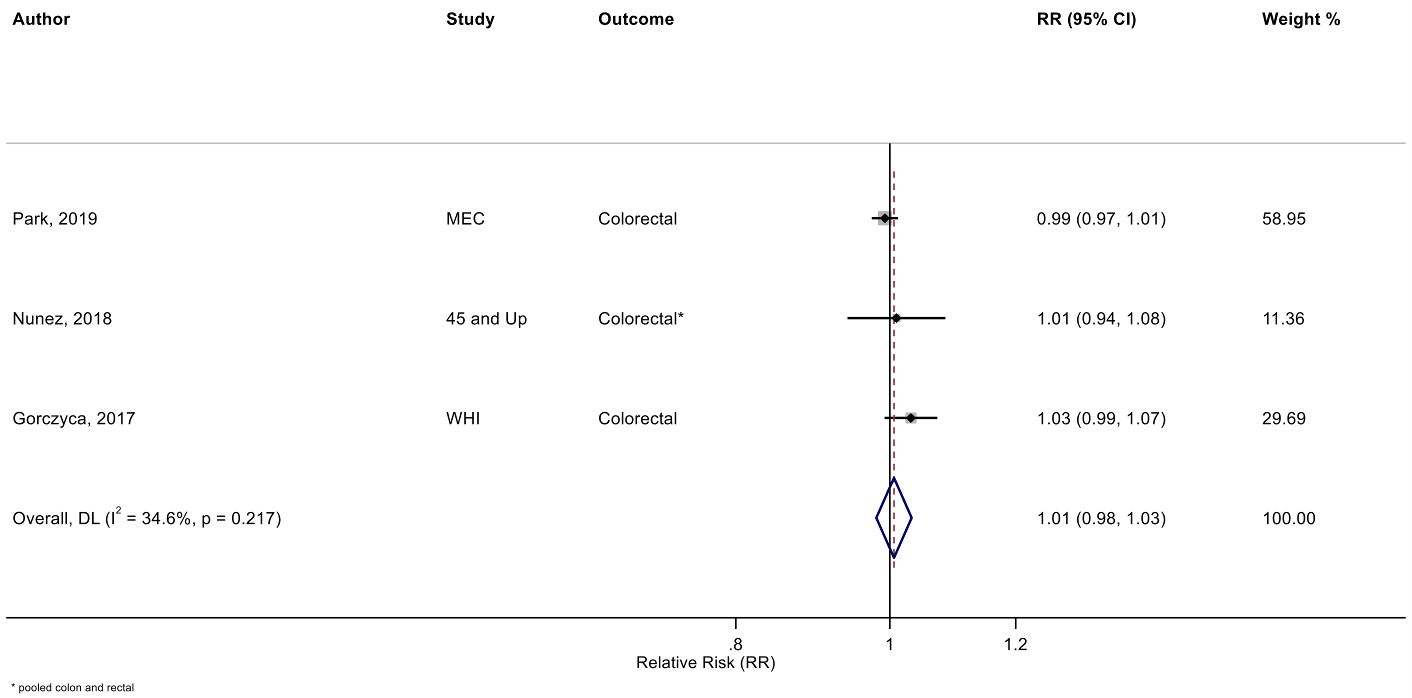

Figure legend: Forest plot shows the linear dose-response results (per 2 hours/day of sedentary time) from the inverse variance DerSimonian-Laird random-effects model. The diamond represents the summary relative risk (RR) estimate and the diamond's width the 95% confidence interval (CI). Each square and the horizontal line across the square represent the RR estimate of the individual study and its 95%CI.

Study abbreviations: MEC, Multiethnic Cohort Study; WHI, Women’s Health Initiative

Park 2019 (30910780)[73]; Nunez 2018 (29510753)[66]; Gorczyca 2018 (28538039)[61]

#### Supplementary Figure 22: Linear dose-response meta-analysis of the association between total sitting time and lung cancer incidence.

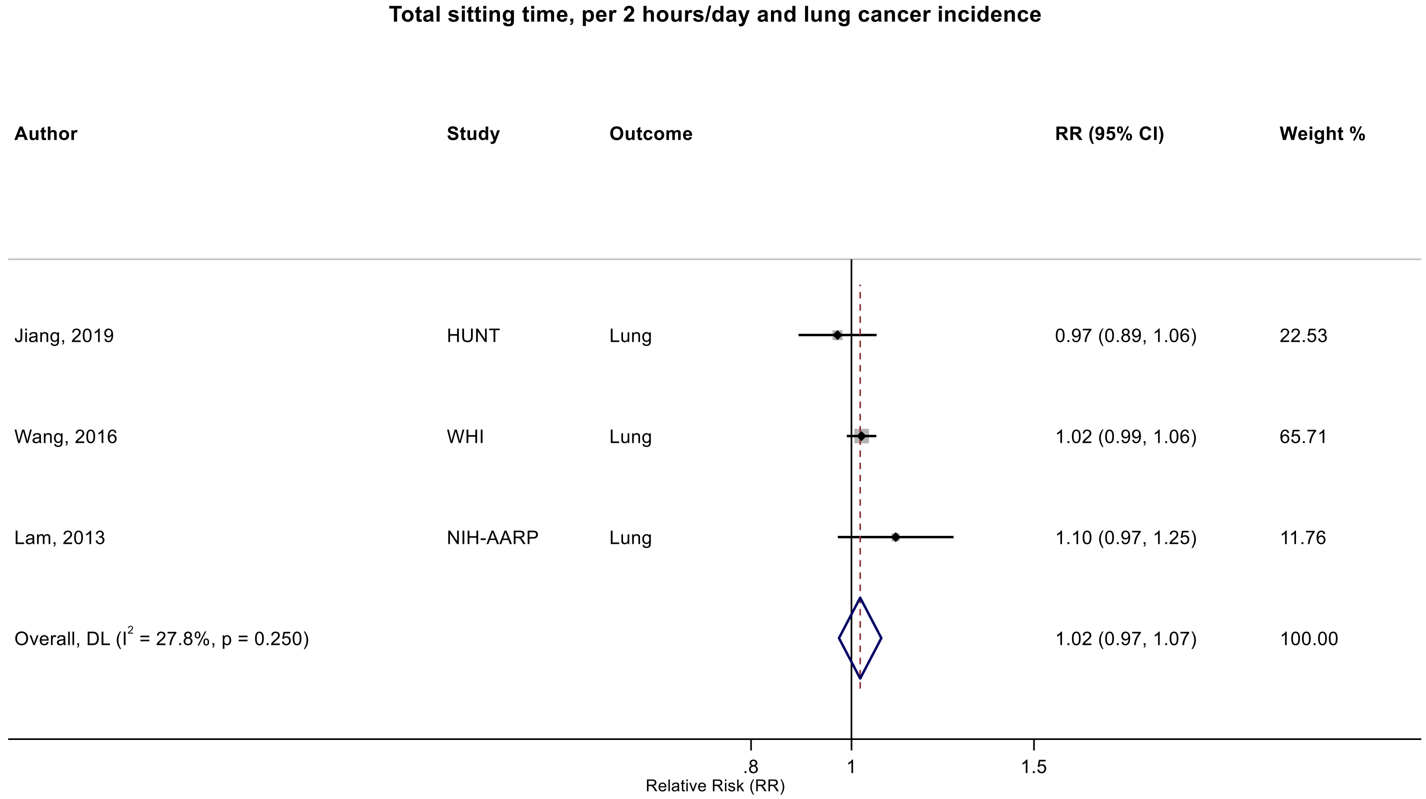

Figure legend: Forest plot shows the linear dose-response results (per 2 hours/day of sedentary time) from the inverse variance DerSimonian-Laird random-effects model. The diamond represents the summary relative risk (RR) estimate and the diamond's width the 95% confidence interval (CI). Each square and the horizontal line across the square represent the RR estimate of the individual study and its 95%CI.

Study abbreviations: HUNT, Trøndelag Health Study; WHI, Women’s Health Initiative; NIH-AARP, National Institute of Health + American Association of Retired Persons Diet and Health Study

Jiang 2019 (30859092)[72]; Wang 2022 (35159055)[86]; Lam 2013 (23940620)[48]

#### Supplementary Figure 23: Linear dose-response meta-analysis of the association between total sitting time and breast cancer incidence.

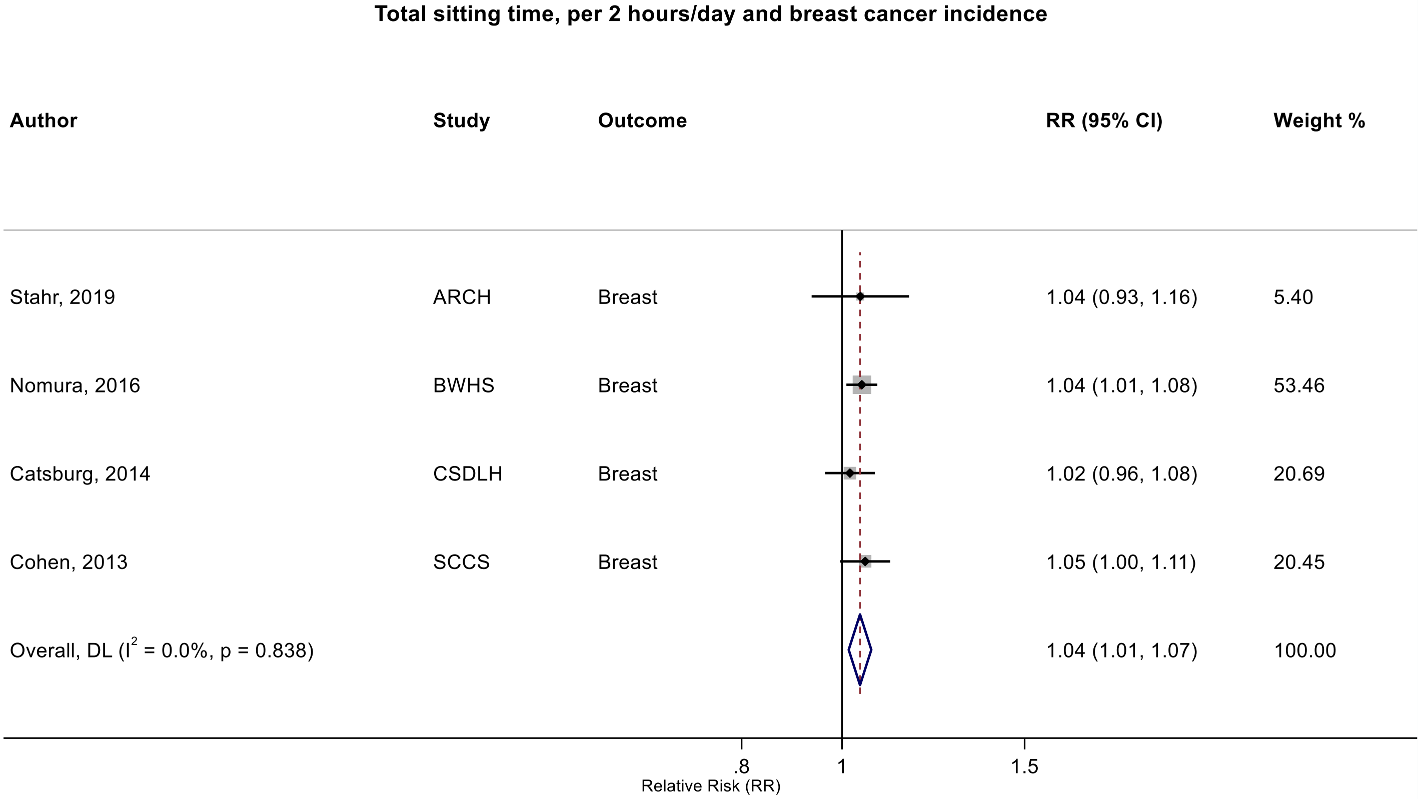

Figure legend: Forest plot shows the linear dose-response results (per 2 hours/day of sedentary time) from the inverse variance DerSimonian-Laird random-effects model. The diamond represents the summary relative risk (RR) estimate and the diamond's width the 95% confidence interval (CI). Each square and the horizontal line across the square represent the RR estimate of the individual study and its 95%CI.

Study abbreviations: ARCH, Arkansas Rural Community Health Study; BWHS, Black Women's Health Study; CSDLH, Canadian Study of Diet, Lifestyle and Health; SCCS, Southern Community Cohort Study

Stahr 2019 (35117114)[85]; Nomura 2016 (27632430)[60]; Catsburg 2014 (24781974)[55]; Cohen 2013 (23576427)[46]

#### Supplementary Figure 24: Linear dose-response meta-analysis of the association between total sitting time and ovarian cancer incidence.

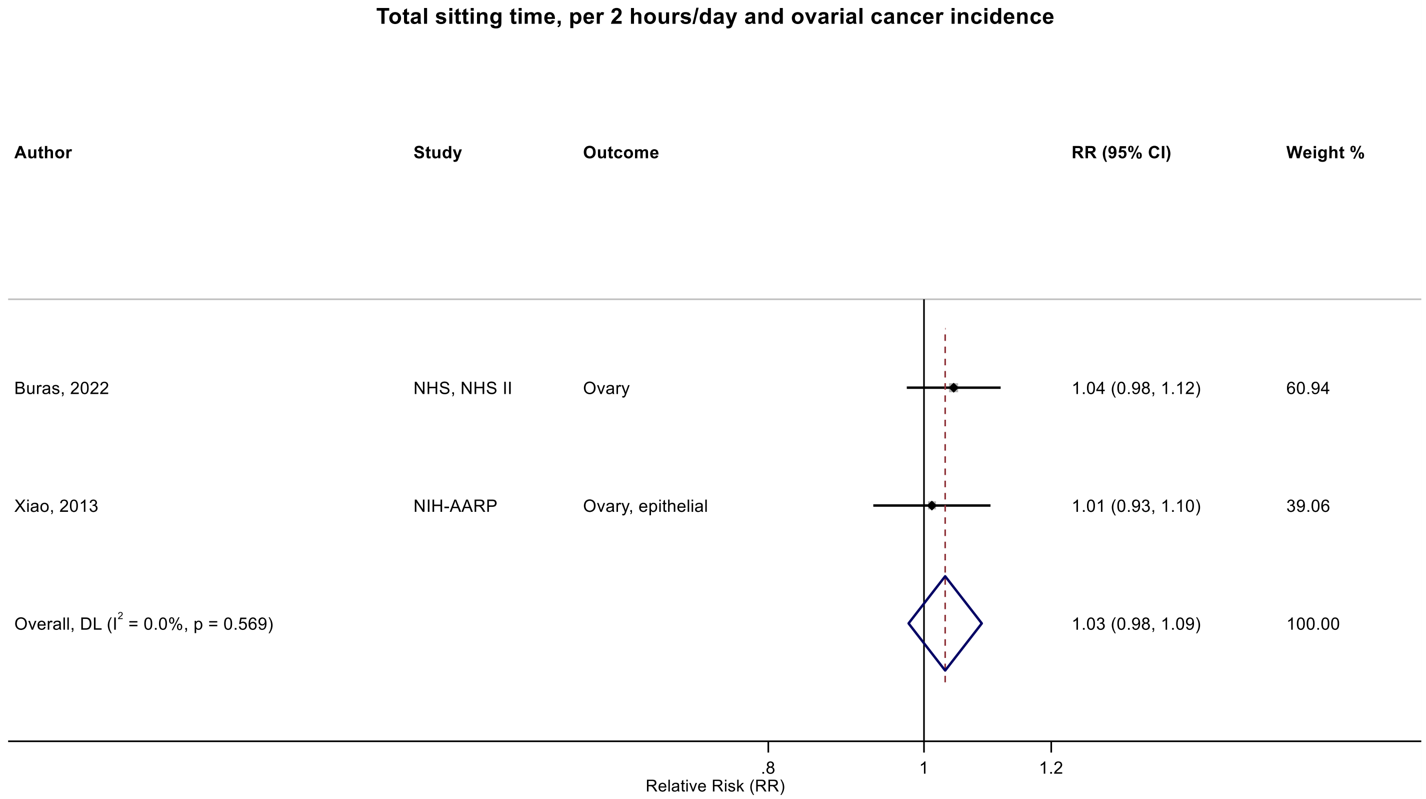

Figure legend: Forest plot shows the linear dose-response results (per 2 hours/day of sedentary time) from the inverse variance DerSimonian-Laird random-effects model. The diamond represents the summary relative risk (RR) estimate and the diamond's width the 95% confidence interval (CI). Each square and the horizontal line across the square represent the RR estimate of the individual study and its 95%CI.

Study abbreviations: NHS, Nurse's Health Study; NIH-AARP, National Institute of Health + American Association of Retired Persons Diet and Health Study

Buras 2022 (35094053)[84]; Xiao 2013 (23966580)[49]

#### Supplementary Figure 25: Categorical highest versus the lowest meta-analysis of the association between total sitting time and prostate cancer incidence.

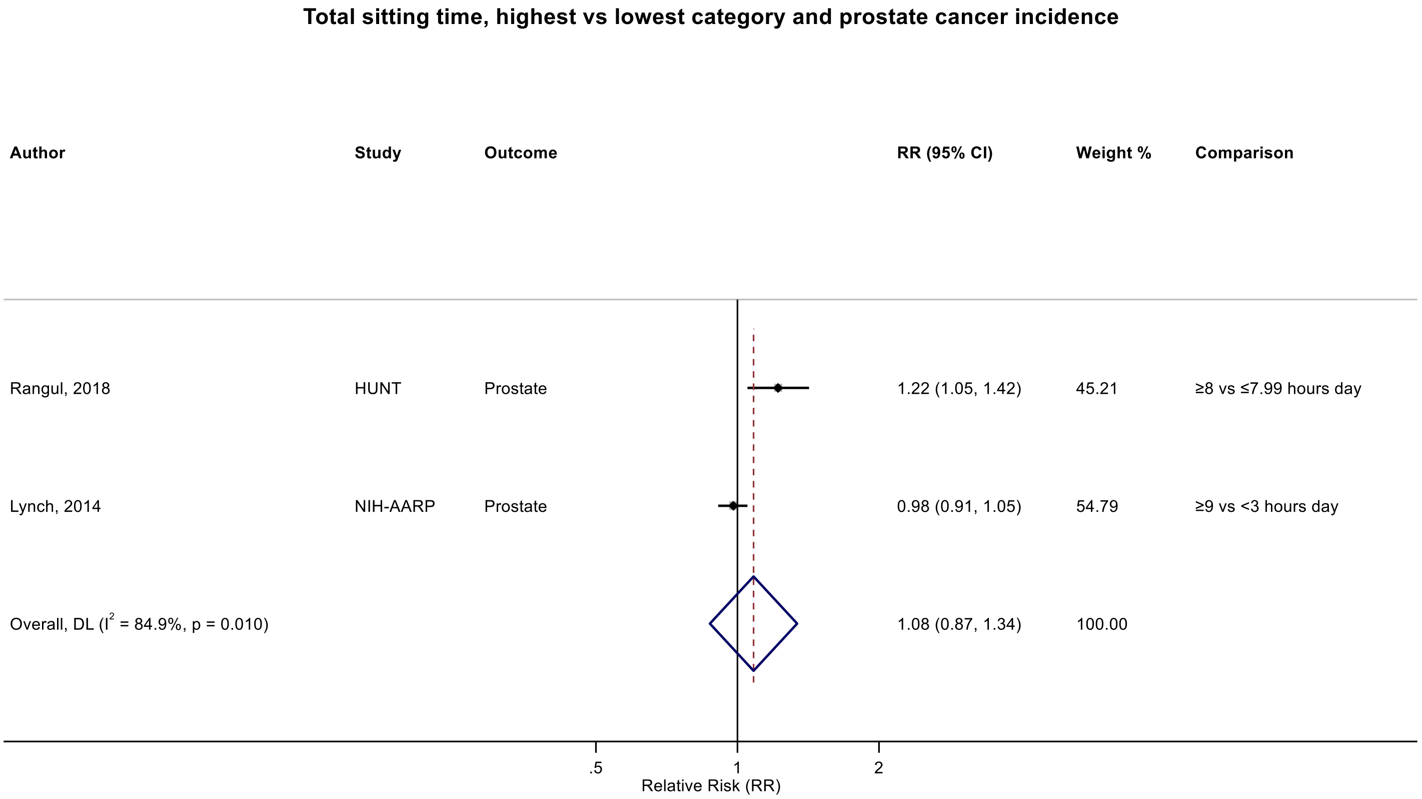

Figure legend: Forest plot shows the categorical comparison results from the individual studies for the associations with the largest contrast, as reported in the respective publication. Each square and the horizontal line across the square represent the relative risk (RR) estimate of the individual study and its 95% confidence interval (CI).

Study abbreviations: HUNT, Trøndelag Health Study; NIH-AARP, National Institute of Health + American Association of Retired Persons Diet and Health Study

Rangul 2018 (30352079)[70]; Lynch 2014 (24526287)[53]

#### Supplementary Figure 26: Linear dose-response meta-analysis of the association between occupational sitting time and colorectal cancer incidence.

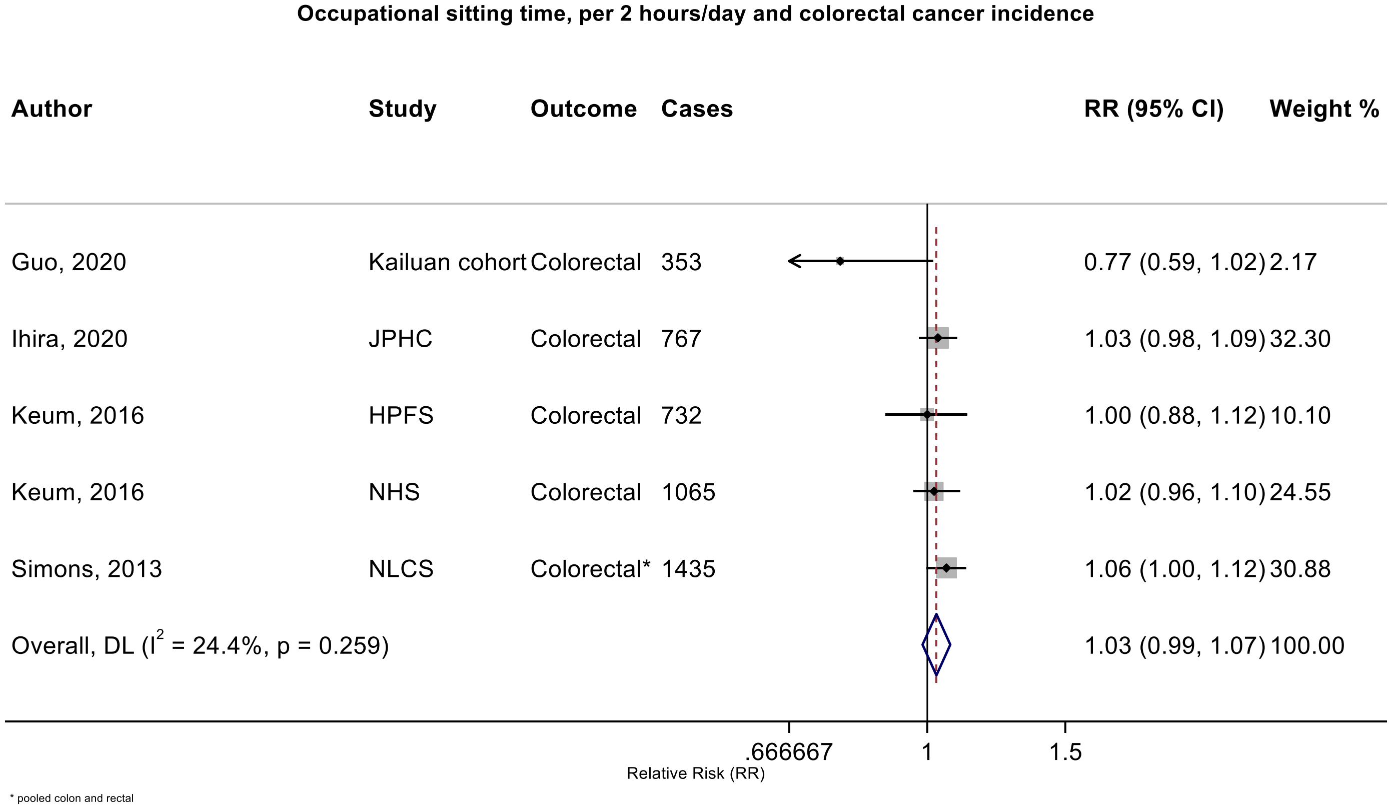

Figure legend: Forest plot shows the linear dose-response results (per 2 hours/day of sedentary time) from the inverse variance DerSimonian-Laird random-effects model. The diamond represents the summary relative risk (RR) estimate and the diamond's width the 95% confidence interval (CI). Each square and the horizontal line across the square represent the RR estimate of the individual study and its 95%CI.

Study abbreviations: JPHC, Japan Public Health Center-Based Prospective Study; HPFS, Health Professionals Follow-up Study; NHS, Nurses' Health Study; NLCS, Netherlands Cohort Study on Diet and Cancer

Guo 2020 (31773920)[76]; Ihira 2020 (31925977)[77]; Keum 2016 (26649988)[58]; Simons 2013 (23420352)[45]

#### Supplementary Figure 27: Linear dose-response meta-analysis of the association between occupational sitting time and pancreatic cancer incidence.

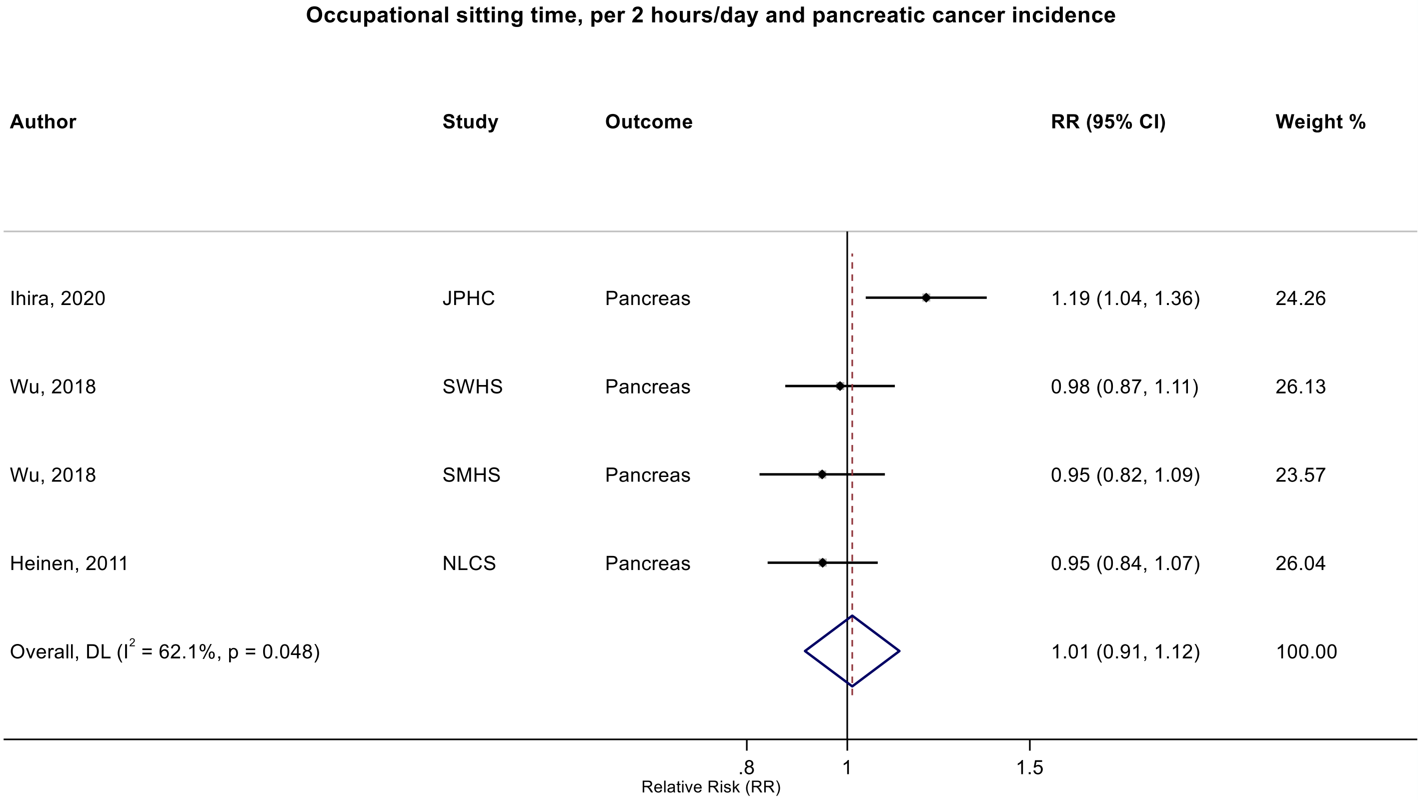

Figure legend: Forest plot shows the linear dose-response results (per 2 hours/day of sedentary time) from the inverse variance DerSimonian-Laird random-effects model. The diamond represents the summary relative risk (RR) estimate and the diamond's width the 95% confidence interval (CI). Each square and the horizontal line across the square represent the RR estimate of the individual study and its 95%CI.

Study abbreviations: JPHC, Japan Public Health Center-Based Prospective Study; SWHS, Shanghai Women's Health Study; SMHS, Shanghai Men's Health Study, NLCS, Netherlands Cohort Study on Diet and Cancer

Ihira 2020 (31925977)[77]; Wu 2018 (29475964)[65]; Heinen 2011 (21955648)[43]

#### Supplementary Figure 28: Linear dose-response meta-analysis of the association between occupational sitting time and breast cancer incidence.

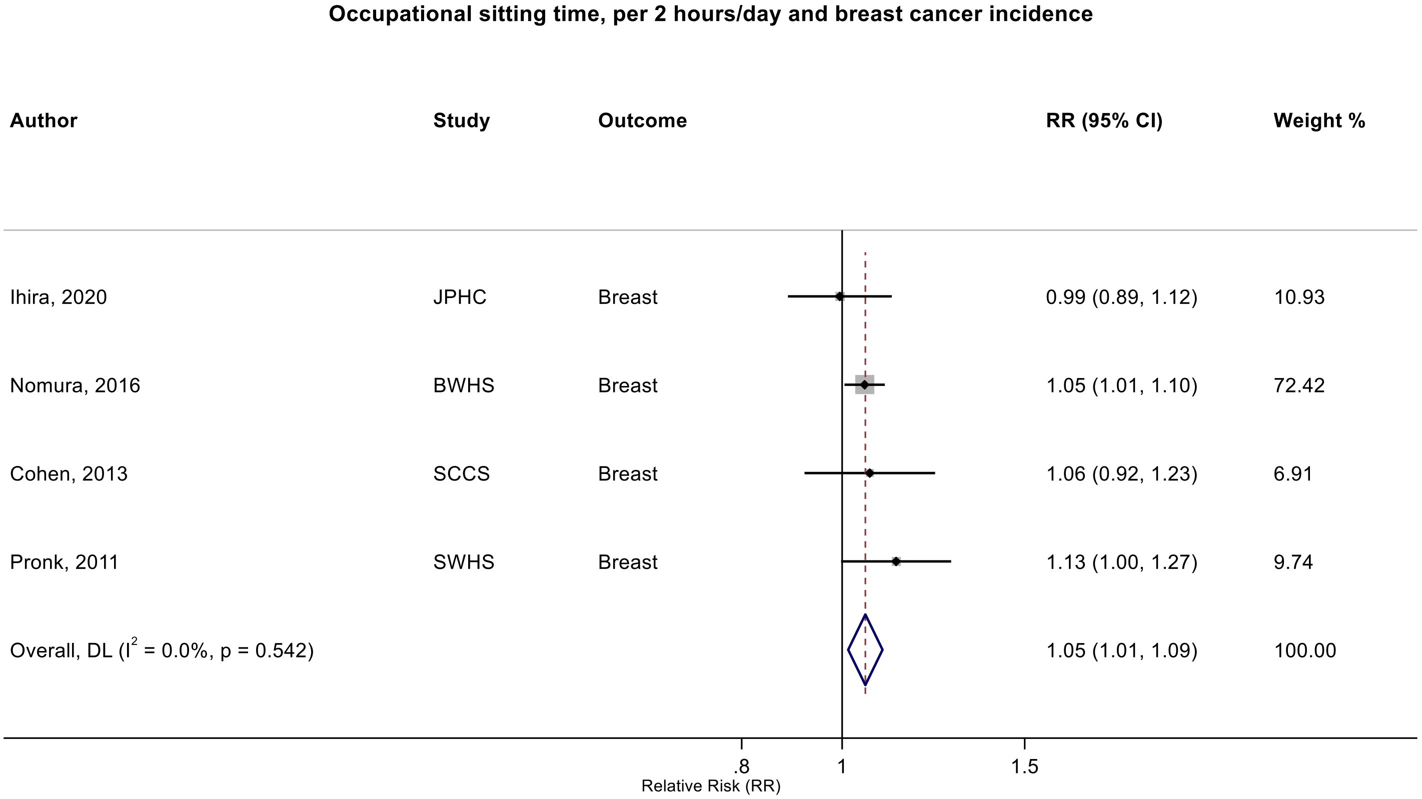

Figure legend: Forest plot shows the linear dose-response results (per 2 hours/day of sedentary time) from the inverse variance DerSimonian-Laird random-effects model. The diamond represents the summary relative risk (RR) estimate and the diamond's width the 95% confidence interval (CI). Each square and the horizontal line across the square represent the RR estimate of the individual study and its 95%CI.

Study abbreviations: JPHC, Japan Public Health Center-Based Prospective Study; BWHS, Black Women's Health Study; SCCS, Southern Community Cohort Study; SWHS, Shanghai Women’s Health Study

Ihira 2020 (31925977)[77]; Nomura 2016 (27632430)[60]; Cohen 2013 (23576427)[46]; Pronk 2011 (21934685)[42]

**Supplementary Figure 29: Linear dose-response meta-analysis of the association between occupational sitting time and ovarian cancer incidence.**

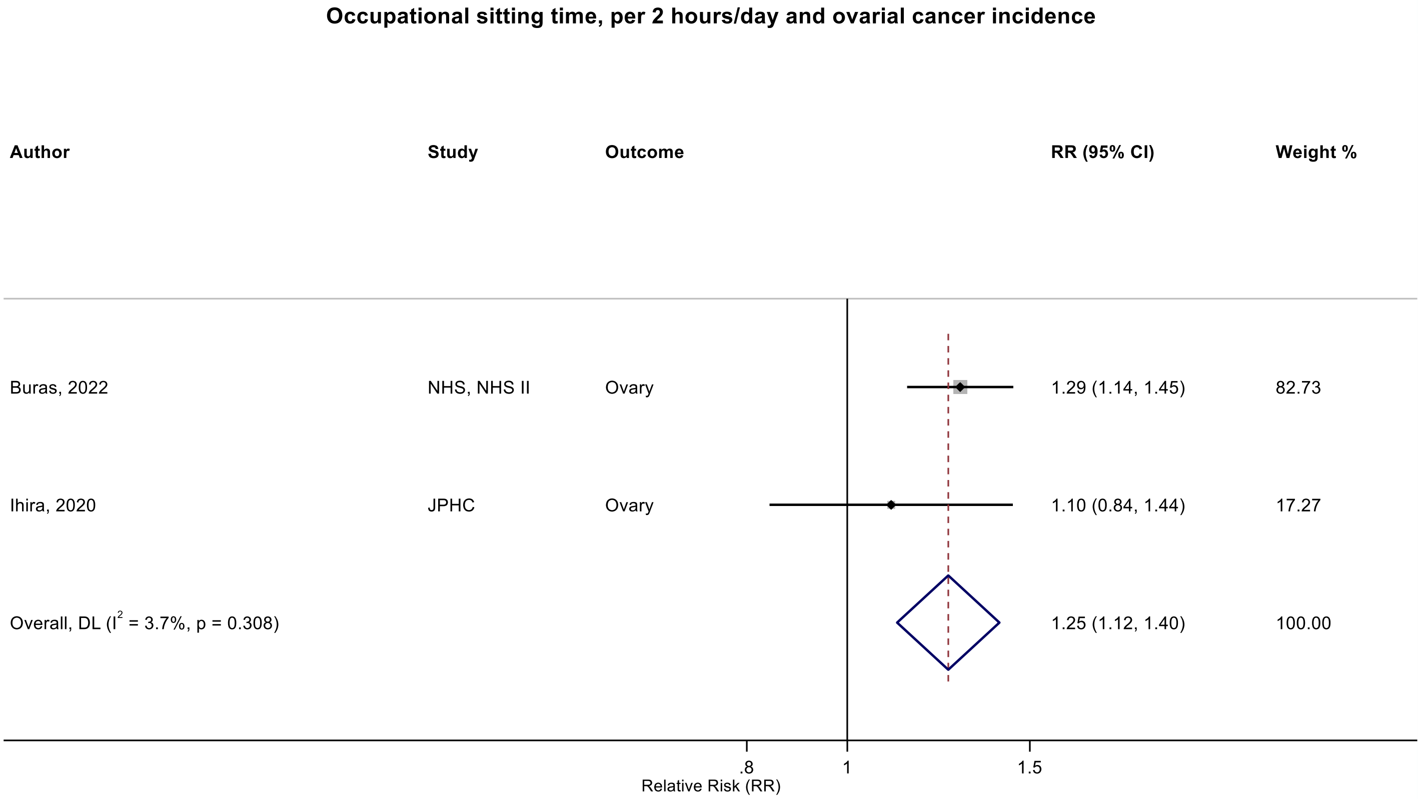

Figure legend: Forest plot shows the linear dose-response results (per 2 hours/day of sedentary time) from the inverse variance DerSimonian-Laird random-effects model. The diamond represents the summary relative risk (RR) estimate and the diamond's width the 95% confidence interval (CI). Each square and the horizontal line across the square represent the RR estimate of the individual study and its 95%CI.

Study abbreviations: NHS, Nurses' Health Study; JPHC, Japan Public Health Center-Based Prospective Study

Buras 2022 (35094053)[84]; Ihira 2020 (31925977)[77]

**Supplementary Figure 30: Linear dose-response meta-analysis of the association between occupational sitting time and prostate cancer incidence.**

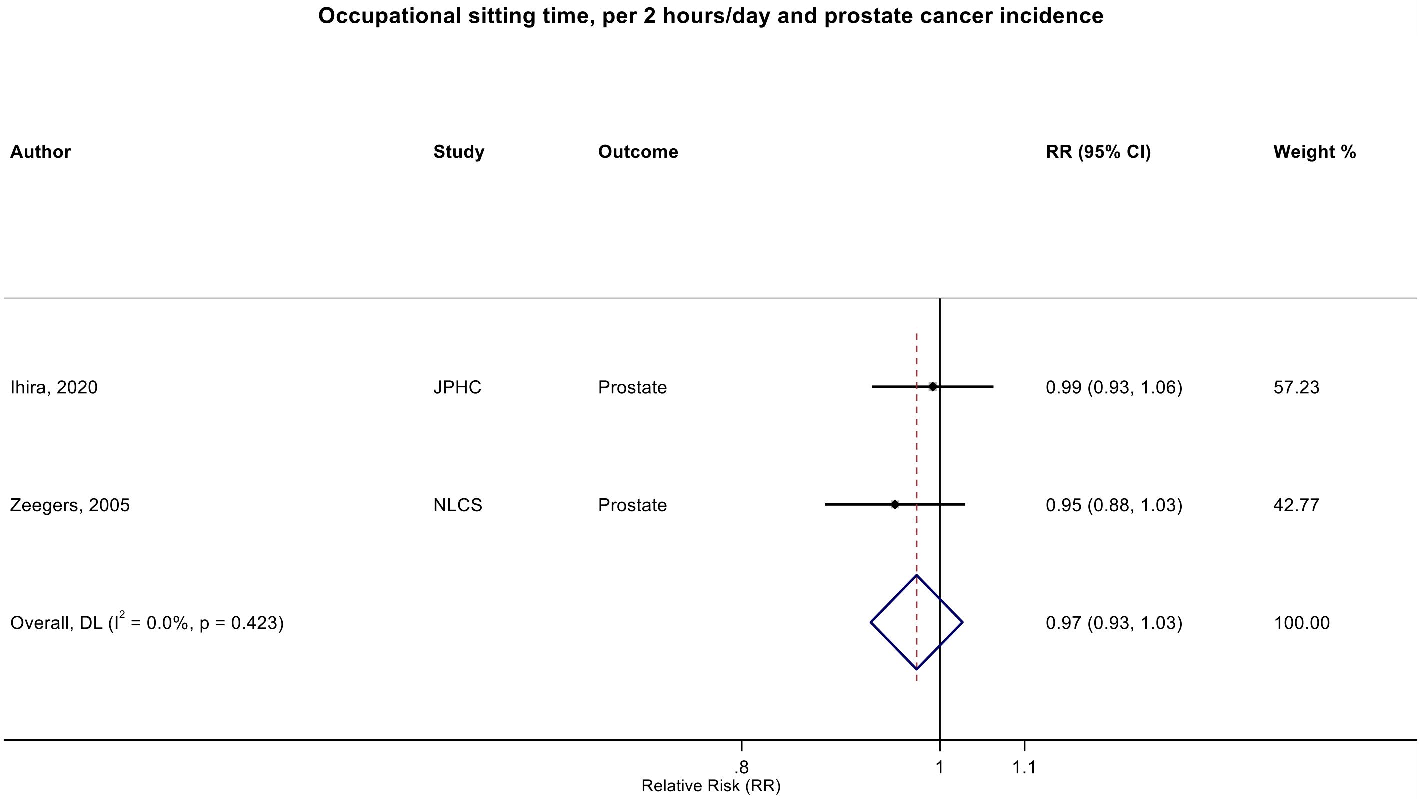

Figure legend: Forest plot shows the linear dose-response results (per 2 hours/day of sedentary time) from the inverse variance DerSimonian-Laird random-effects model. The diamond represents the summary relative risk (RR) estimate and the diamond's width the 95% confidence interval (CI). Each square and the horizontal line across the square represent the RR estimate of the individual study and its 95%CI.

Study abbreviations: JPHC, Japan Public Health Center-Based Prospective Study; NLCS, Netherlands Cohort Study on Diet and Cancer

Zeegers 2005 (15941961)[34]; Ihira 2020 (31925977)[77]

#### Supplementary Figure 31: Categorical highest versus the lowest meta-analysis of the association between total sitting time and breast cancer incidence.

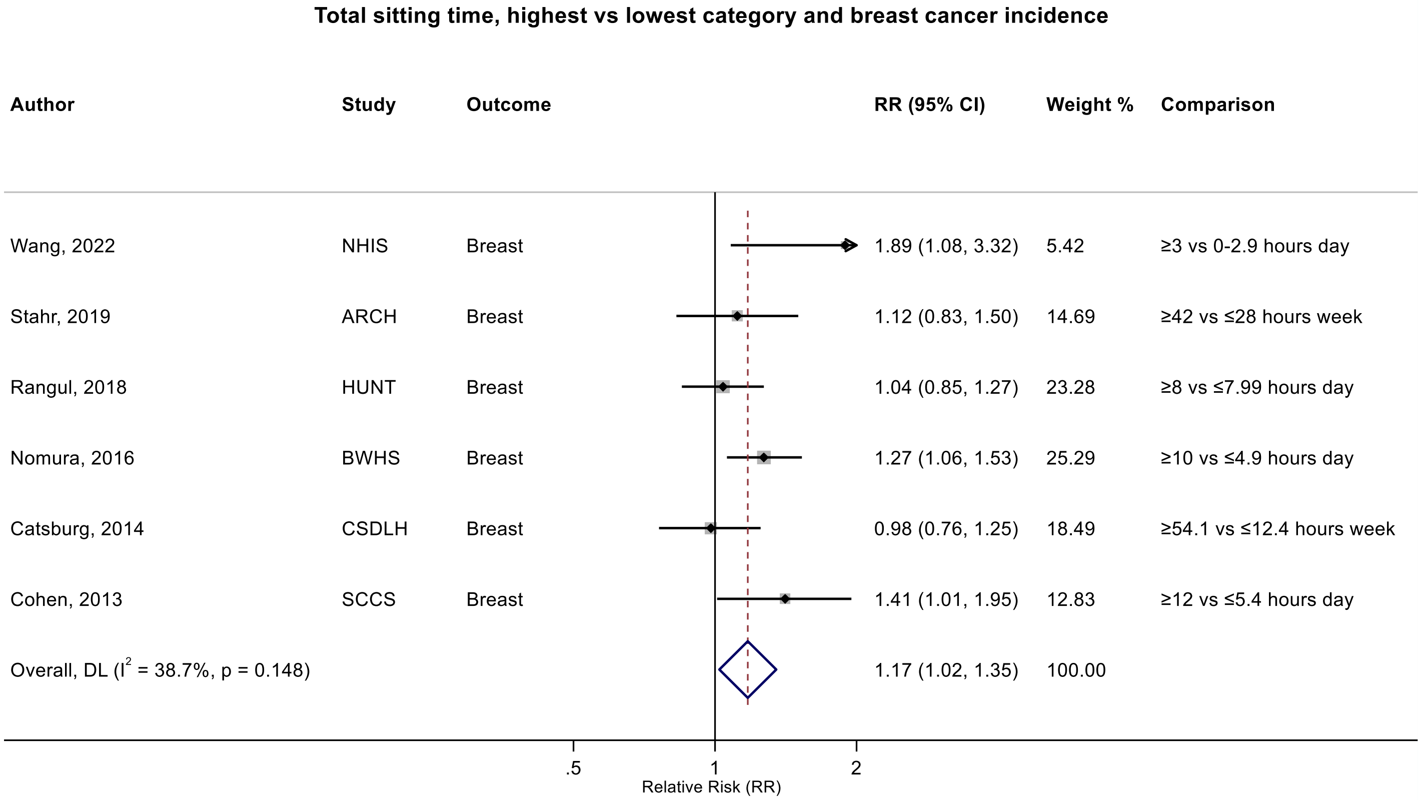

Figure legend: Forest plot shows the categorical comparison results from the individual studies for the associations with the largest contrast, as reported in the respective publication. Each square and the horizontal line across the square represent the relative risk (RR) estimate of the individual study and its 95% confidence interval (CI).

Study abbreviations: NHIS, National Health Interview Survey; ARCH, Arkansas Rural Community Health Study; HUNT, Trøndelag Health Study; BWHS, Black Women's Health Study; CSDLH, Canadian Study of Diet, Lifestyle and Health; SCCS, Southern Community Cohort Study

Wang 2022 (35159055)[86]; Stahr 2019 (35117114)[85]; Rangul 2018 (30352079)[70]; Nomura 2016 (27632430)[60]; Catsburg 2014 (24781974)[55]; Cohen 2013 (23576427)[46]

**Supplementary Figure 32: Linear dose-response meta-analysis of the association between total sitting time and breast cancer incidence, by menopause status.**

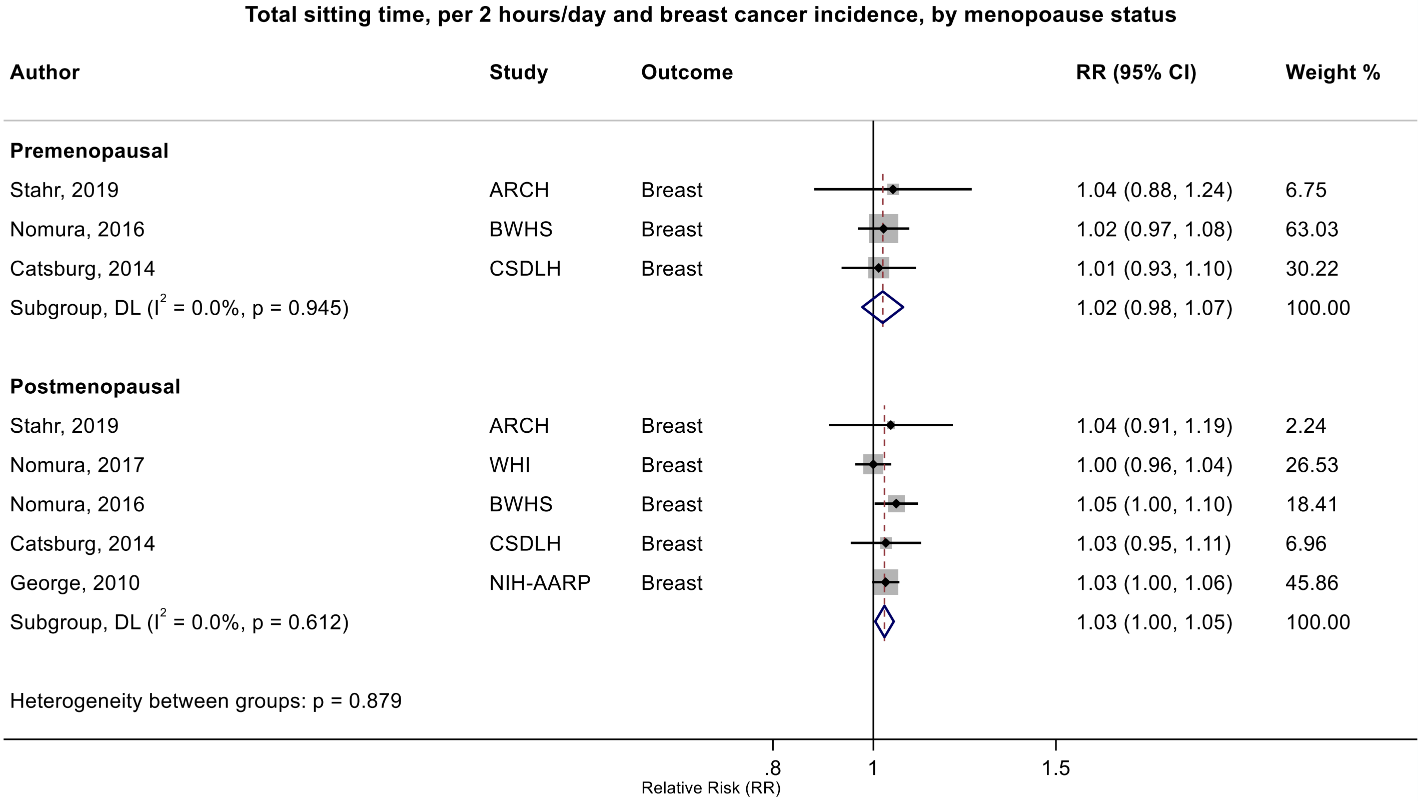

Figure legend: Forest plot shows the linear dose-response results (per 2 hours/day of sedentary time) from the inverse variance DerSimonian-Laird random-effects model. The diamond represents the summary relative risk (RR) estimate and the diamond's width the 95% confidence interval (CI). Each square and the horizontal line across the square represent the RR estimate of the individual study and its 95%CI.

Study abbreviations: ARCH, Arkansas Rural Community Health Study; BWHS, Black Women's Health Study; CSDLH, Canadian Study of Diet, Lifestyle and Health; ; NIH-AARP, National Institute of Health + American Association of Retired Persons Diet and Health Study; SCCS, Southern Community Cohort Study; WHI, Women’s Health Initiative

Stahr 2019 (35117114)[85]; Nomura 2016 (27632430)[60]; Nomura 2017 (28975422)[63]; Catsburg 2014 (24781974)[55] ; George 2010 (20864719)[39]

#### Supplementary Figure 33: Linear dose-response meta-analysis of the association between total sitting time and breast cancer incidence, by hormone receptor status.

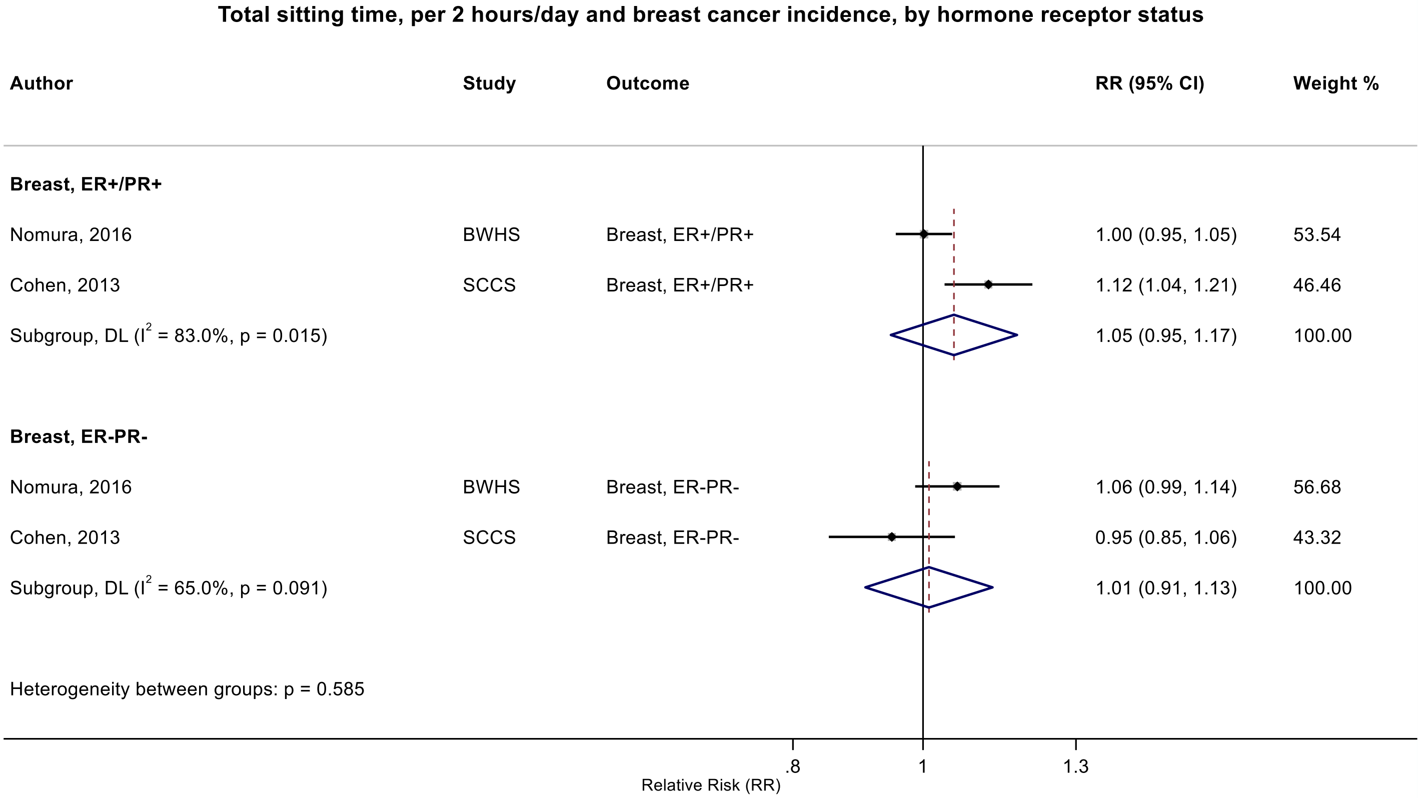

Figure legend: Forest plot shows the linear dose-response results (per 2 hours/day of sedentary time) from the inverse variance DerSimonian-Laird random-effects model. The diamond represents the summary relative risk (RR) estimate and the diamond's width the 95% confidence interval (CI). Each square and the horizontal line across the square represent the RR estimate of the individual study and its 95%CI.

Study abbreviations: BWHS, Black Women's Health Study; SCCS, Southern Community Cohort Study

Nomura 2016 (27632430)[60]; Cohen 2013 (23576427)[46]

#### Supplementary Figure 34: Linear dose-response meta-analysis of the association between total sitting time and breast cancer incidence, by physical activity adjustment.

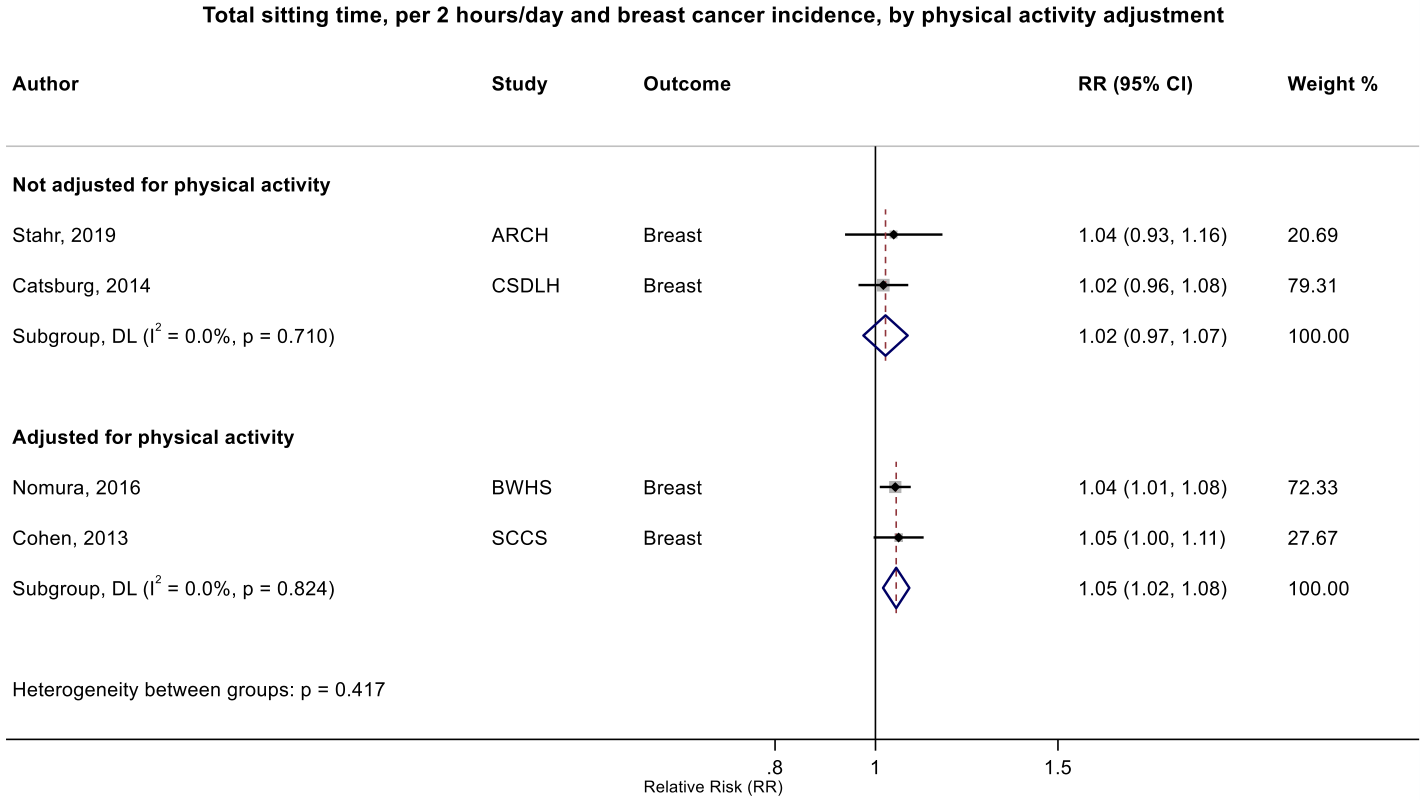

Figure legend: Forest plot shows the linear dose-response results (per 2 hours/day of sedentary time) from the inverse variance DerSimonian-Laird random-effects model. The diamond represents the summary relative risk (RR) estimate and the diamond's width the 95% confidence interval (CI). Each square and the horizontal line across the square represent the RR estimate of the individual study and its 95%CI.

Study abbreviations: ARCH, Arkansas Rural Community Health Study; BWHS, Black Women's Health Study; CSDLH, Canadian Study of Diet, Lifestyle and Health; SCCS, Southern Community Cohort Study

Stahr 2019 (35117114)[85]; Catsburg 2014 (24781974)[55]; Nomura 2016 (27632430)[60]; Cohen 2013 (23576427)[46]

**Supplementary Figure 35: Linear dose-response meta-analysis of the association between total sitting time and breast cancer incidence, by risk of bias due to confounding (Domain 1).**

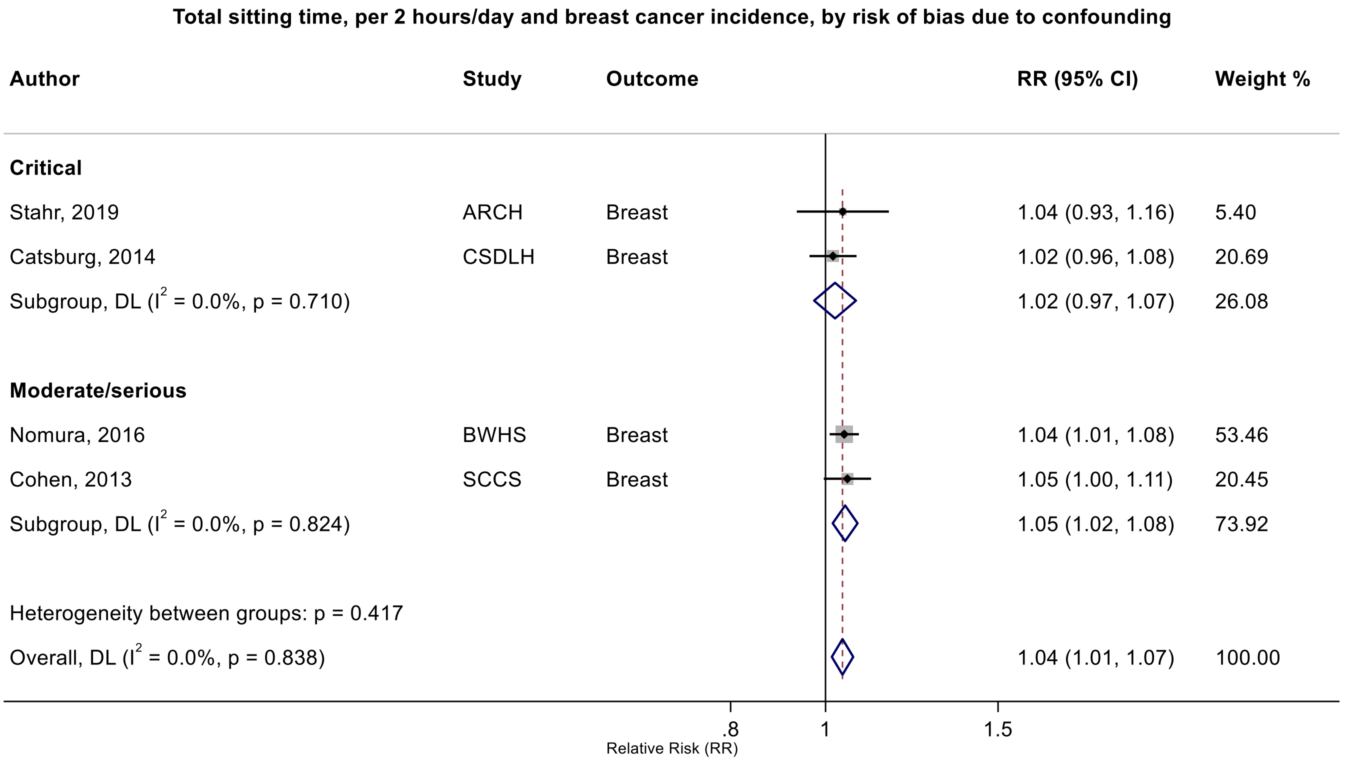

Figure legend: Forest plot shows the linear dose-response results (per 2 hours/day of sedentary time) from the inverse variance DerSimonian-Laird random-effects model. The diamond represents the summary relative risk (RR) estimate and the diamond's width the 95% confidence interval (CI). Each square and the horizontal line across the square represent the RR estimate of the individual study and its 95%CI.

Study abbreviations: ARCH, Arkansas Rural Community Health Study; BWHS, Black Women's Health Study; CSDLH, Canadian Study of Diet, Lifestyle and Health; SCCS, Southern Community Cohort Study

Stahr 2019 (35117114)[85]; Catsburg 2014 (24781974)[55]; Nomura 2016 (27632430)[60]; Cohen 2013 (23576427)[46]

#### Supplementary Figure 36: Categorical highest versus the lowest meta-analysis of the association between total sitting time and breast cancer incidence, by risk of bias due to confounding (Domain 1).

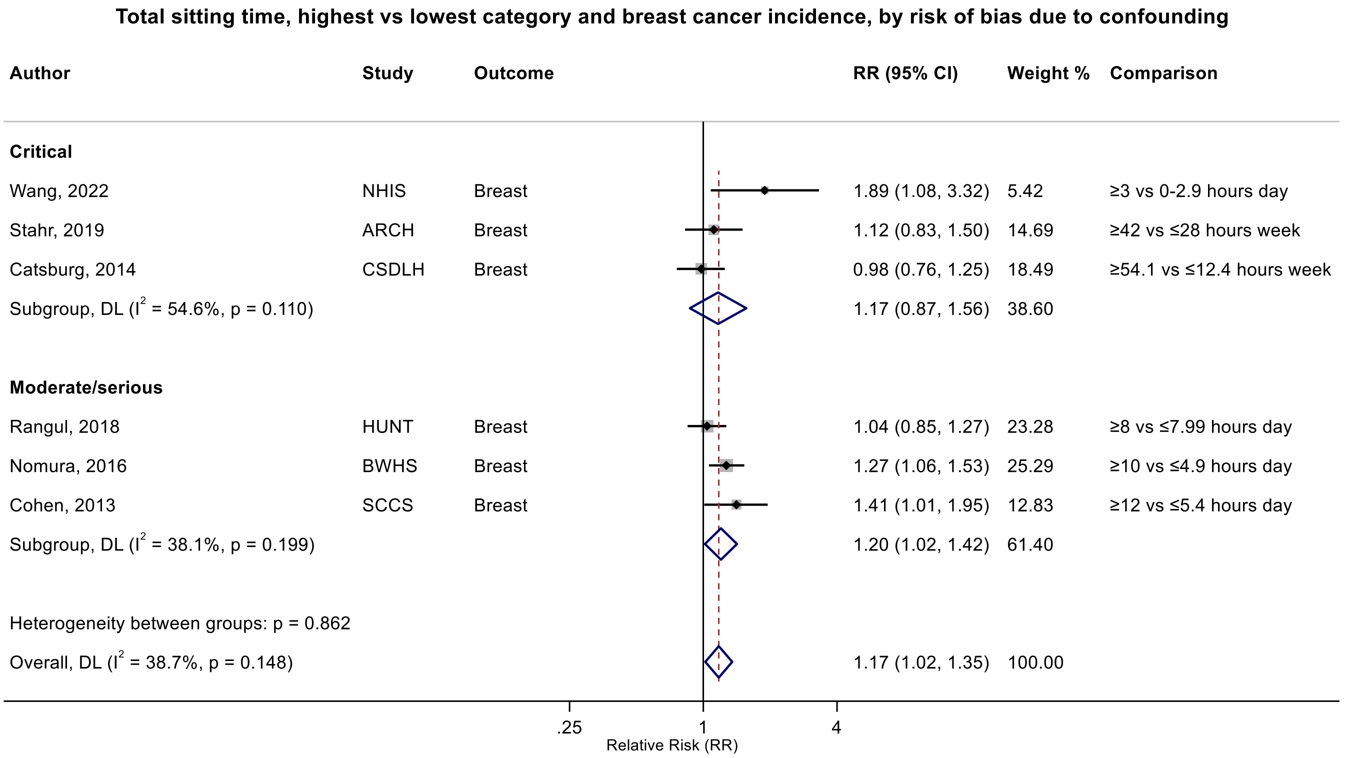

Figure legend: Forest plot shows the categorical comparison results from the individual studies for the associations with the largest contrast, as reported in the respective publication. Each square and the horizontal line across the square represent the relative risk (RR) estimate of the individual study and its 95% confidence interval (CI).

Study abbreviations: NHIS, National Health Interview Survey; ARCH, Arkansas Rural Community Health Study; HUNT, Trøndelag Health Study; BWHS, Black Women's Health Study; CSDLH, Canadian Study of Diet, Lifestyle and Health; SCCS, Southern Community Cohort Study

Wang 2022 (35159055)[86]; Stahr 2019 (35117114)[85]; Rangul 2018 (30352079)[70]; Nomura 2016 (27632430)[60]; Catsburg 2014 (24781974)[55]; Cohen 2013 (23576427)[46]

**Supplementary Figure 37: Linear dose-response meta-analysis of the association between occupational sitting time and breast cancer incidence, by menopausal status.**

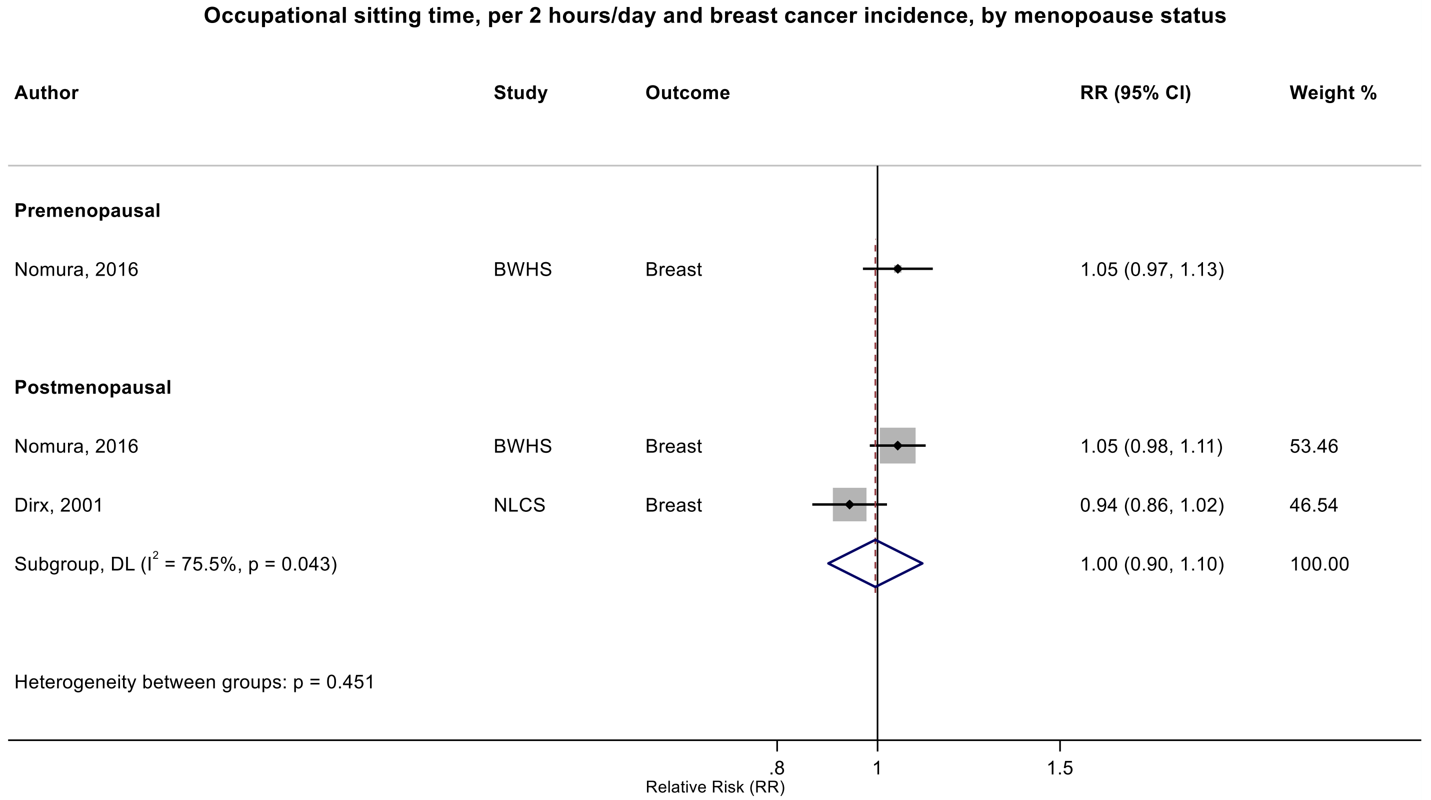

Figure legend: Forest plot shows the linear dose-response results (per 2 hours/day of sedentary time) from the inverse variance DerSimonian-Laird random-effects model. The diamond represents the summary relative risk (RR) estimate and the diamond's width the 95% confidence interval (CI). Each square and the horizontal line across the square represent the RR estimate of the individual study and its 95%CI.

Study abbreviations: BWHS, Black Women's Health Study, NLCS, Netherlands Cohort Study on Diet and Cancer

Nomura 2016 (27632430)[60]; Dirx 2001 (11745243)[33]

#### Supplementary Figure 38: Categorical highest versus the lowest meta-analysis of the association between total sitting time and colorectal cancer incidence.

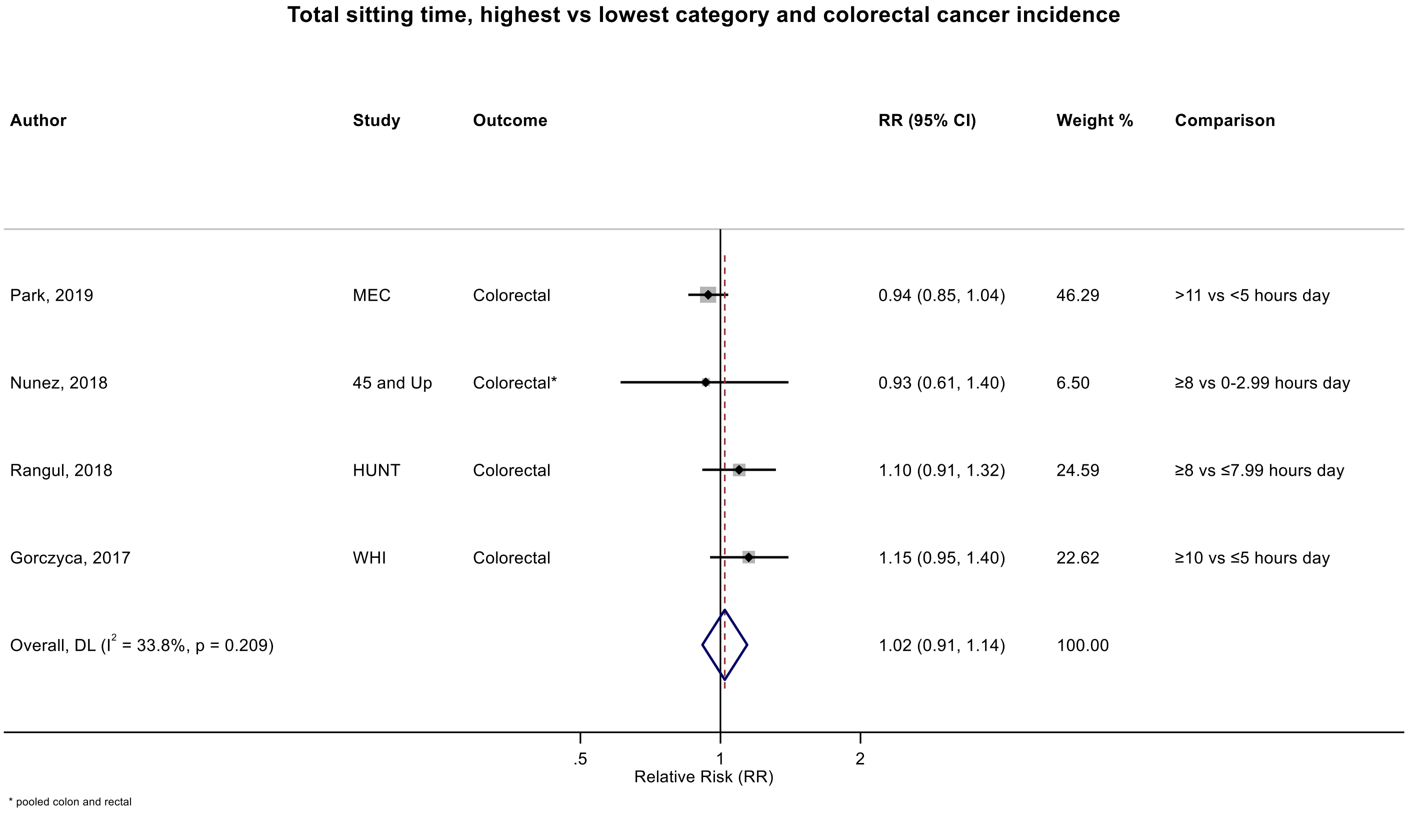

Figure legend: Forest plot shows the categorical comparison results from the individual studies for the associations with the largest contrast, as reported in the respective publication. Each square and the horizontal line across the square represent the relative risk (RR) estimate of the individual study and its 95% confidence interval (CI).

Study abbreviations: MEC, Multiethnic Cohort Study; HUNT, Trøndelag Health Study; WHI, Women's Health Initiative

Park 2019 (30910780)[73]; Nunez 2018 (29510753)[66]; Rangul 2018 (30352079)[70]; Gorczyca 2018 (28538039)[61]

#### Supplementary Figure 39: Linear dose-response meta-analysis of the association between total sitting time and colorectal cancer incidence, by anatomical subsite.

**Total sitting time, per 2 hours/day and colorectal cancer incidence, by anatomical subsite**

| Author | Study | Outcome | RR (95% CI) | Weight % |
|---|---|---|---|---|
| **Colon** | | | | |
| Nunez, 2018 | 45 and Up | Colon | 1.02 (0.96, 1.08) | 18.23 |
| Gorczyca, 2017 | WHI | Colon | 1.03 (0.99, 1.08) | 35.71 |
| Howard, 2008 | NIH-AARP | Colon | 1.04 (1.00, 1.08) | 46.06 |
| Subgroup, DL ($I^2$ = 0.0%, p = 0.785) | | | 1.03 (1.01, 1.06) | 100.00 |
| **Rectal** | | | | |
| Nunez, 2018 | 45 and Up | Rectal | 0.98 (0.90, 1.07) | 51.35 |
| Gorczyca, 2017 | WHI | Rectal | 1.00 (0.92, 1.10) | 48.65 |
| Subgroup, DL ($I^2$ = 0.0%, p = 0.705) | | | 0.99 (0.93, 1.05) | 100.00 |
| Heterogeneity between groups: p = 0.203 | | | | |

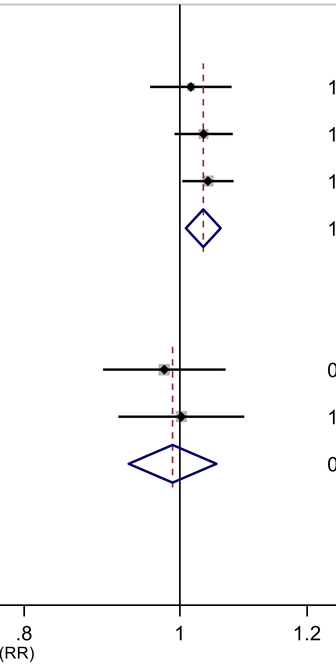

Figure legend: Forest plot shows the linear dose-response results (per 2 hours/day of sedentary time) from the inverse variance DerSimonian-Laird random-effects model. The diamond represents the summary relative risk (RR) estimate and the diamond's width the 95% confidence interval (CI). Each square and the horizontal line across the square represent the RR estimate of the individual study and its 95%CI.

Study abbreviations: WHI, Women's Health Initiative; NIH-AARP, National Institute of Health + American Association of Retired Persons Diet and Health Study

Nunez 2018 (29510753)[66]; Gorczyca 2018 (28538039)[61]; Howard 2008 (18437512)[36]

#### Supplementary Figure 40: Linear dose-response meta-analysis of the association between occupational sitting time and colorectal cancer incidence, by anatomical subsite.

**Occupational sitting time, per 2 hours/day and colorectal cancer incidence, by anatomical subsite**

| Author | Study | Outcome | RR (95% CI) | Weight % |
|---|---|---|---|---|
| **Colon** | | | | |
| Ihira, 2020 | JPHC | Colon | 1.05 (0.98, 1.13) | 50.49 |
| Simons, 2013 | NLCS | Colon | 1.11 (1.04, 1.20) | 49.51 |
| Subgroup, DL ($I^2$ = 28.2%, p = 0.238) | | | 1.08 (1.02, 1.15) | 100.00 |
| **Rectal** | | | | |
| Ihira, 2020 | JPHC | Rectal | 0.99 (0.90, 1.09) | 48.82 |
| Simons, 2013 | NLCS | Rectal | 0.95 (0.87, 1.05) | 51.18 |
| Subgroup, DL ($I^2$ = 0.0%, p = 0.569) | | | 0.97 (0.91, 1.04) | 100.00 |
| Heterogeneity between groups: p = 0.021 | | | | |

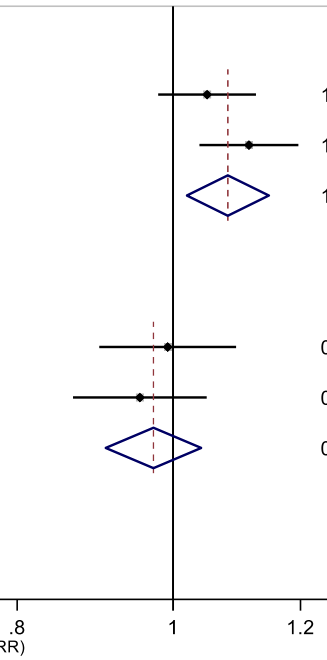

Figure legend: Forest plot shows the linear dose-response results (per 2 hours/day of sedentary time) from the inverse variance DerSimonian-Laird random-effects model. The diamond represents the summary relative risk (RR) estimate and the diamond's width the 95% confidence interval (CI). Each square and the horizontal line across the square represent the RR estimate of the individual study and its 95%CI.

Study abbreviations: JPHC, Japan Public Health Center-Based Prospective Study; NLCS, Netherlands Cohort Study on Diet and Cancer

Ihira 2020 (31925977)[77]; Simons 2013 (23420352)[45]

Supplementary Materials

#### Supplementary Figure 41: Linear dose-response meta-analysis of the association between occupational sitting time and colorectal cancer incidence, by sex.

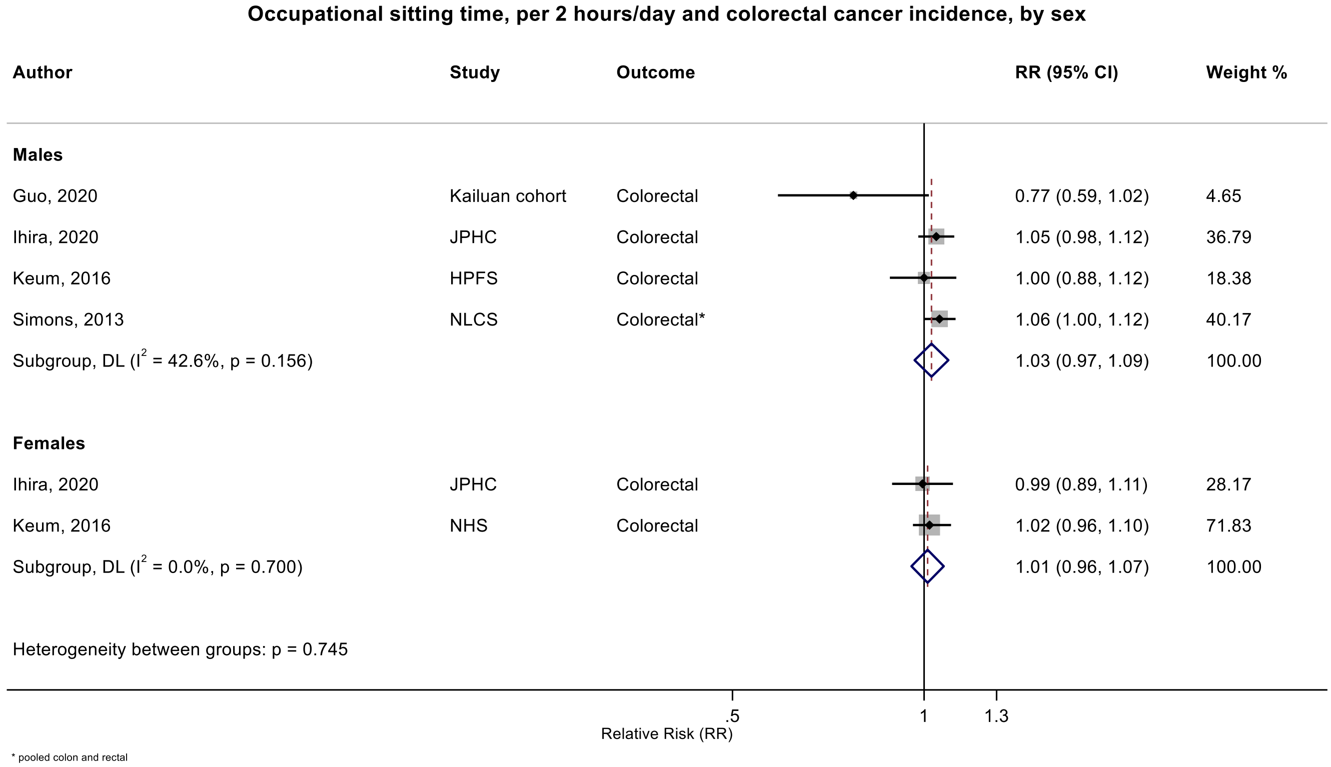

Figure legend: Forest plot shows the linear dose-response results (per 2 hours/day of sedentary time) from the inverse variance DerSimonian-Laird random-effects model. The diamond represents the summary relative risk (RR) estimate and the diamond's width the 95% confidence interval (CI). Each square and the horizontal line across the square represent the RR estimate of the individual study and its 95%CI.

Study abbreviations: JPHC, Japan Public Health Center-Based Prospective Study; NLCS, Netherlands Cohort Study on Diet and Cancer; HPFS, Health Professionals Follow-up Study; NHS, Nurses' Health Study

Guo 2020 (31773920)[76]; Ihira 2020 (31925977)[77]; Keum 2016 (26649988)[58]; Simons 2013 (23420352)[45]

#### Supplementary Figure 42: Linear dose-response meta-analysis of the association between occupational sitting time and colorectal cancer incidence, by region.

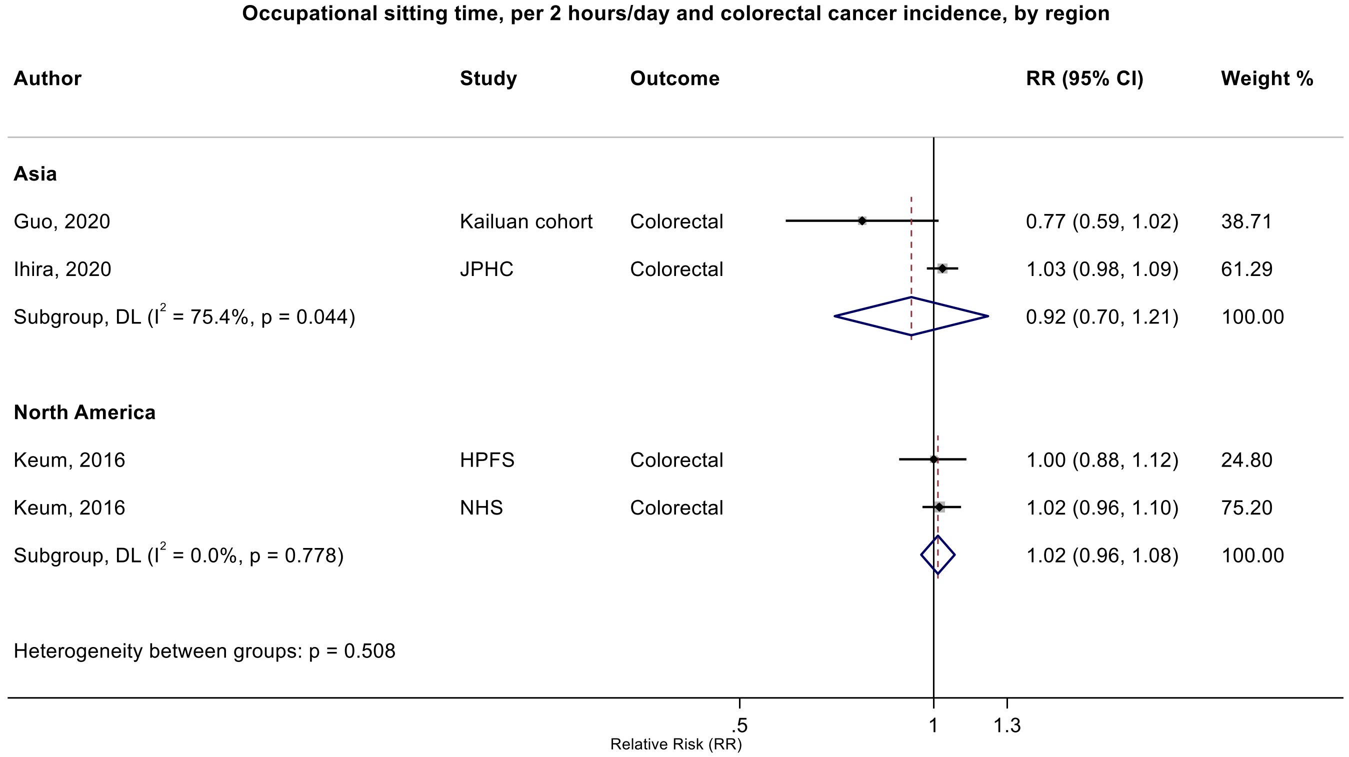

Figure legend: Forest plot shows the linear dose-response results (per 2 hours/day of sedentary time) from the inverse variance DerSimonian-Laird random-effects model. The diamond represents the summary relative risk (RR) estimate and the diamond's width the 95% confidence interval (CI). Each square and the horizontal line across the square represent the RR estimate of the individual study and its 95%CI.

Study abbreviations: JPHC, Japan Public Health Center-Based Prospective Study; HPFS, Health Professionals Follow-up Study; NHS, Nurses' Health Study

Guo 2020 (31773920)[76]; Ihira 2020 (31925977)[77]; Keum 2016 (26649988)[58]

#### Supplementary Figure 43: Linear dose-response meta-analysis of the association between occupational sitting time and colorectal cancer incidence, by physical activity adjustment.

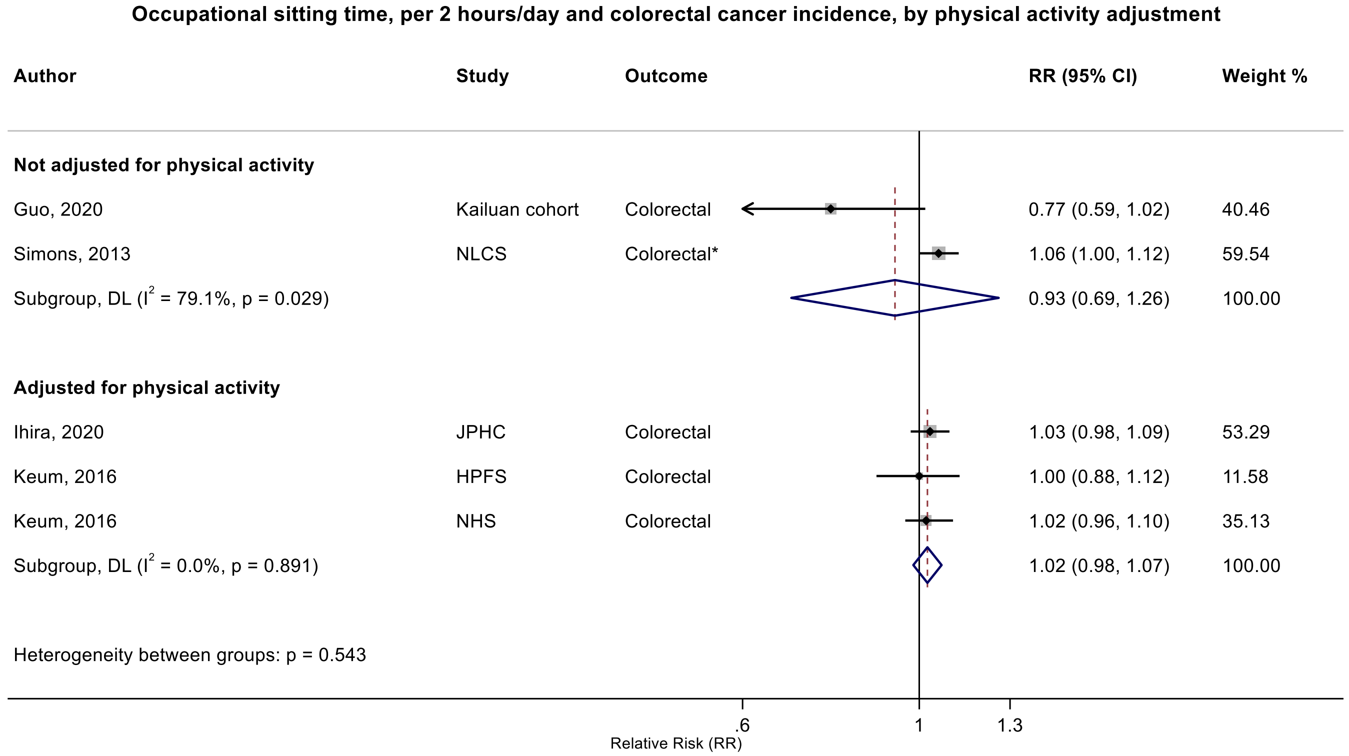

Figure legend: Forest plot shows the linear dose-response results (per 2 hours/day of sedentary time) from the inverse variance DerSimonian-Laird random-effects model. The diamond represents the summary relative risk (RR) estimate and the diamond's width the 95% confidence interval (CI). Each square and the horizontal line across the square represent the RR estimate of the individual study and its 95%CI.

Study abbreviations: JPHC, Japan Public Health Center-Based Prospective Study; NLCS, Netherlands Cohort Study on Diet and Cancer; HPFS, Health Professionals Follow-up Study; NHS, Nurses' Health Study

Guo 2020 (31773920)[76]; Simons 2013 (23420352)[45]; Ihira 2020 (31925977)[77]; Keum 2016 (26649988)[58]

**Supplementary Figure 44: Linear dose-response meta-analysis of the association between occupational sitting time and colorectal cancer incidence, by risk of bias due to confounding (Domain 1).**

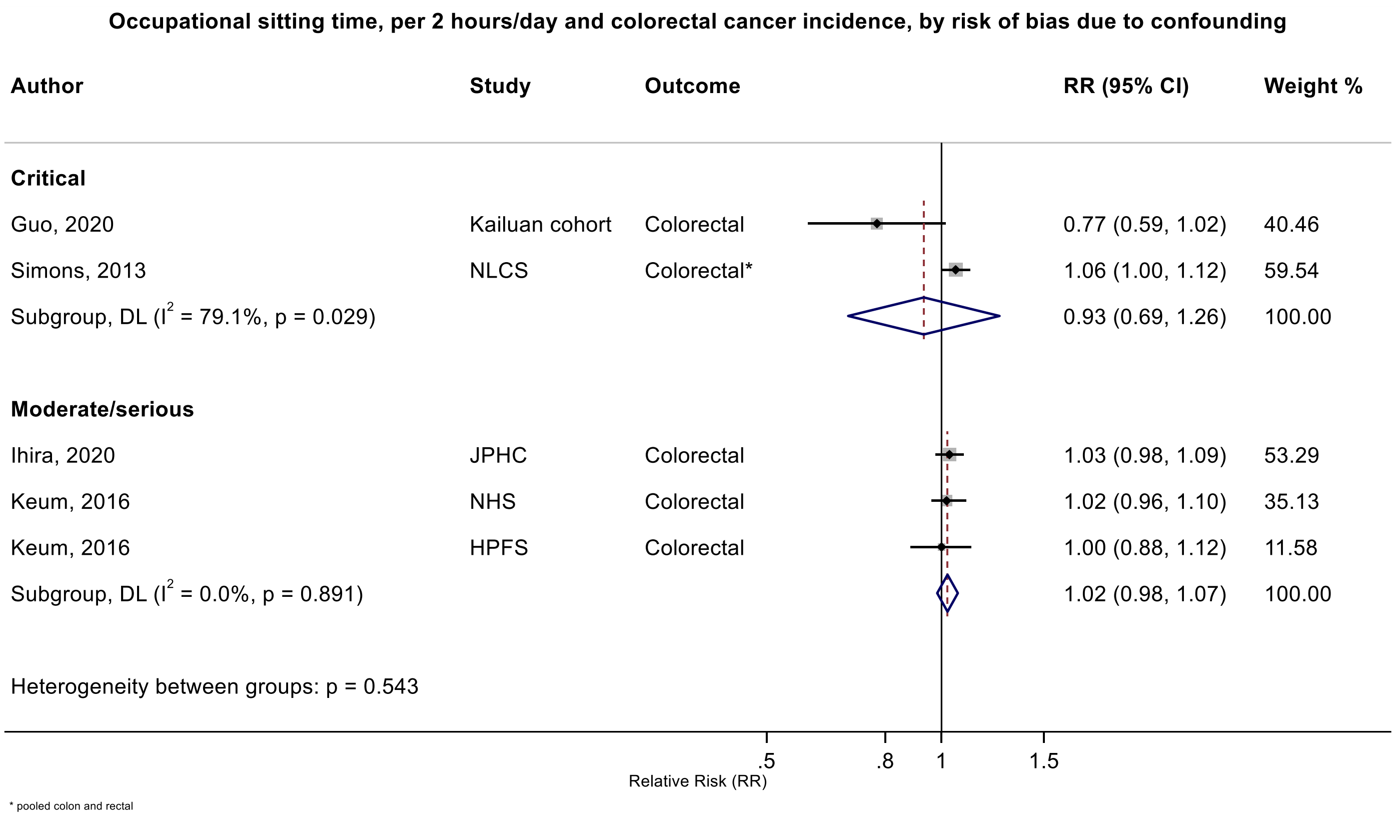

Figure legend: Forest plot shows the linear dose-response results (per 2 hours/day of sedentary time) from the inverse variance DerSimonian-Laird random-effects model. The diamond represents the summary relative risk (RR) estimate and the diamond's width the 95% confidence interval (CI). Each square and the horizontal line across the square represent the RR estimate of the individual study and its 95%CI.

Study abbreviations: JPHC, Japan Public Health Center-Based Prospective Study; NLCS, Netherlands Cohort Study on Diet and Cancer; HPFS, Health Professionals Follow-up Study; NHS, Nurses' Health Study

Guo 2020 (31773920)[76]; Simons 2013 (23420352)[45]; Ihira 2020 (31925977)[77]; Keum 2016 (26649988)[58]

**Supplementary Figure 45: Linear dose-response meta-analysis of the association between occupational sitting time and colorectal cancer incidence, by risk of bias due to selective reporting (Domain 7).**

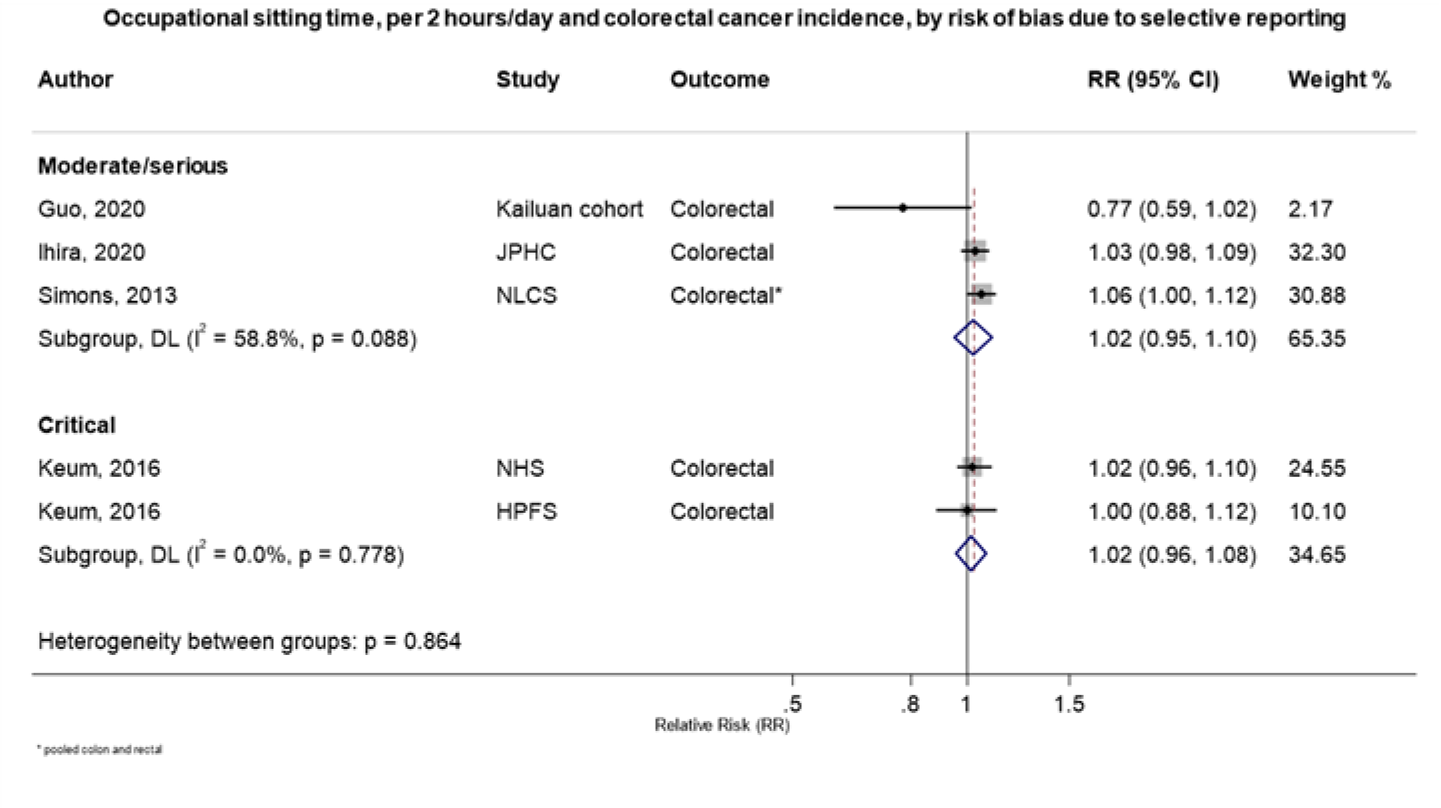

Figure legend: Forest plot shows the linear dose-response results (per 2 hours/day of sedentary time) from the inverse variance DerSimonian-Laird random-effects model. The diamond represents the summary relative risk (RR) estimate and the diamond's width the 95% confidence interval (CI). Each square and the horizontal line across the square represent the RR estimate of the individual study and its 95%CI.

Study abbreviations: JPHC, Japan Public Health Center-Based Prospective Study; NLCS, Netherlands Cohort Study on Diet and Cancer; HPFS, Health Professionals Follow-up Study; NHS, Nurses' Health Study

Guo 2020 (31773920)[76]; Simons 2013 (23420352)[45]; Ihira 2020 (31925977)[77]; Keum 2016 (26649988)[58]

**Supplementary Figure 46: Linear dose-response meta-analysis of the association between total sitting time and colorectal cancer incidence, leave-one-out sensitivity analysis.**

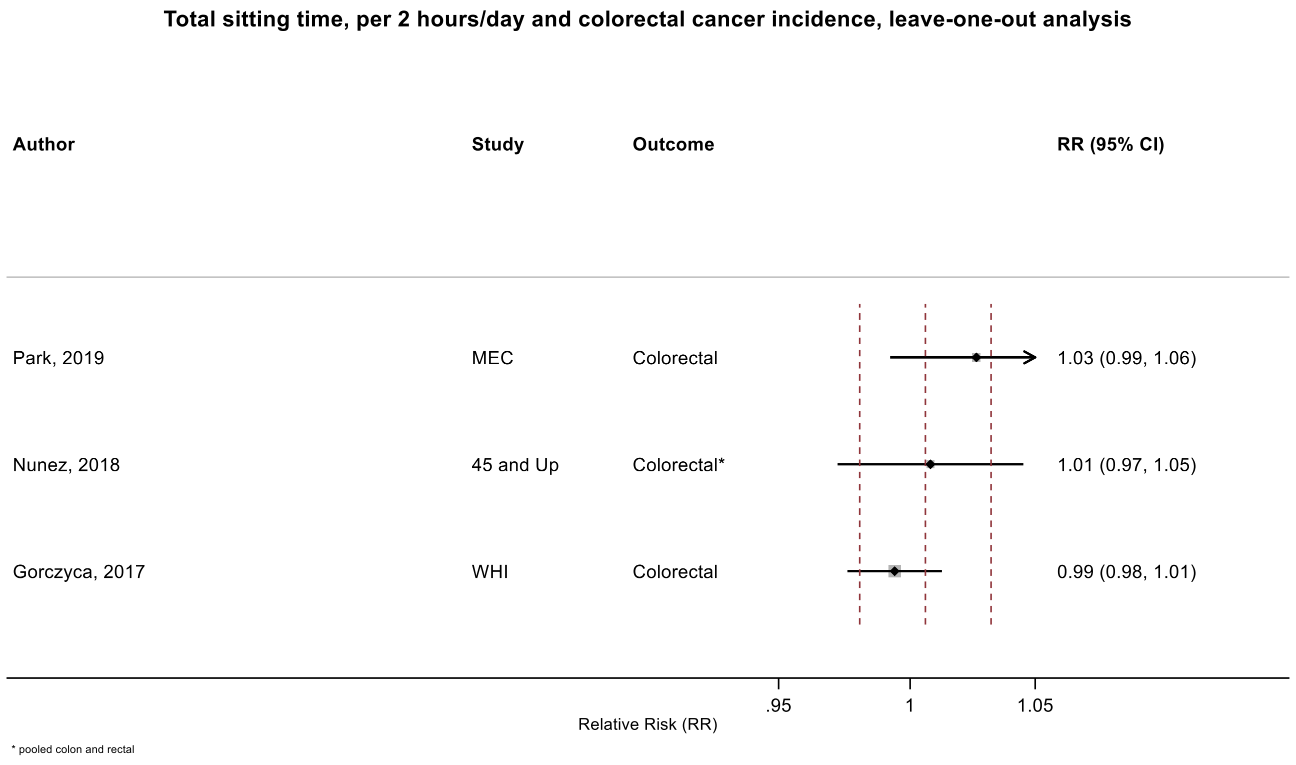

Figure legend: Forest plot shows the linear dose-response results (per 2 hours of sedentary time) from the inverse variance DerSimonian-Laird random-effects model after exclusion of each study from the meta-analysis. For each study, the square represents the summary relative risk (RR) estimate and the horizontal line across the square the corresponding 95% confidence interval (CI) computed from a meta-analysis excluding that study. The dashed vertical lines represent the results of the main meta-analysis.

Study abbreviations: MEC, Multiethnic Cohort Study; WHI, Women's Health Initiative

Park 2019 (30910780)[73]; Nunez 2018 (29510753)[66]; Gorczyca 2018 (28538039)[61]

#### Supplementary Figure 47: Linear dose-response meta-analysis of the association between total sitting time and lung cancer incidence, leave-one-out sensitivity analysis.

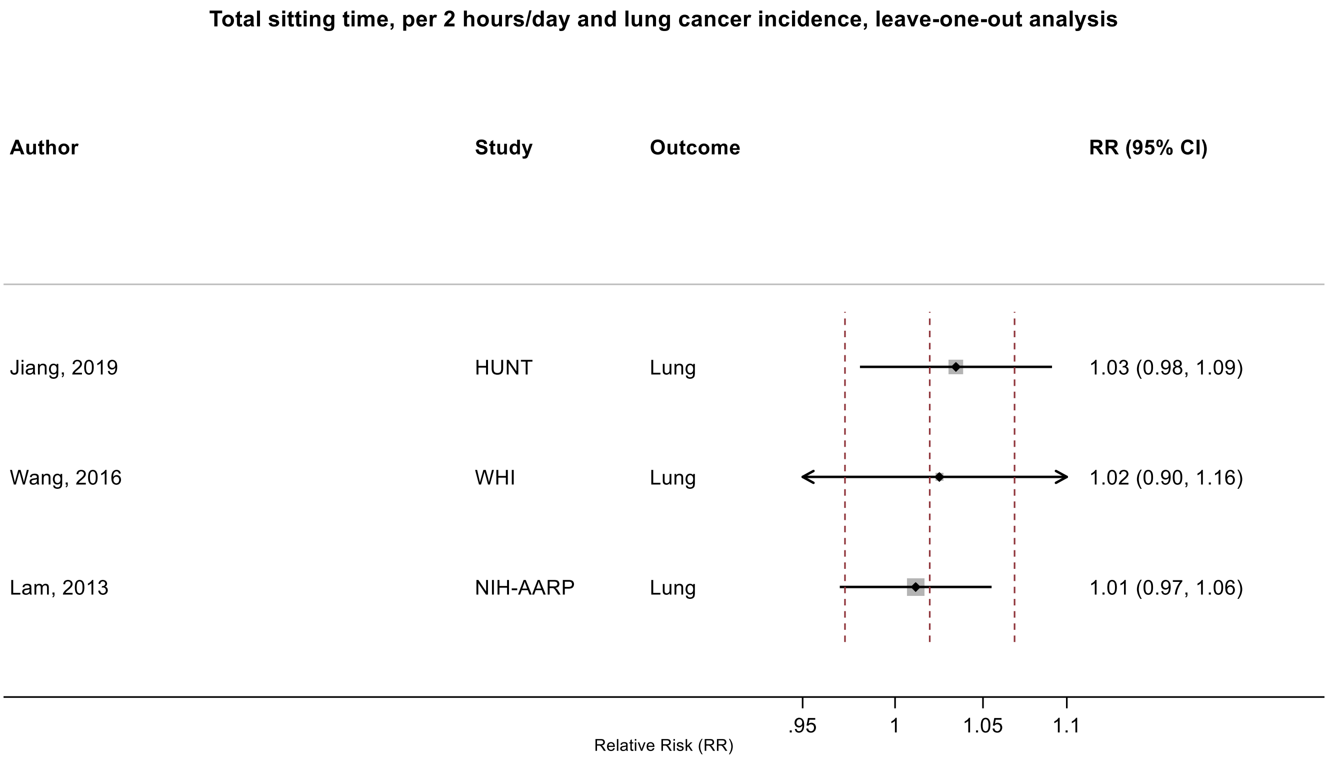

Figure legend: Forest plot shows the linear dose-response results (per 2 hours of sedentary time) from the inverse variance DerSimonian-Laird random-effects model after exclusion of each study from the meta-analysis. For each study, the square represents the summary relative risk (RR) estimate and the horizontal line across the square the corresponding 95% confidence interval (CI) computed from a meta-analysis excluding that study. The dashed vertical lines represent the results of the main meta-analysis.

Study abbreviations: HUNT, Trøndelag Health Study; WHI, Women’s Health Initiative, NIH-AARP, National Institute of Health + American Association of Retired Persons Diet and Health Study

Jiang 2019 (30859092)[72]; Wang 2016 (27439221)[59]; Lam 2013 (23940620)[48]

**Supplementary Figure 48: Linear dose-response meta-analysis of the association between total sitting time and breast cancer incidence, leave-one-out sensitivity analysis.**

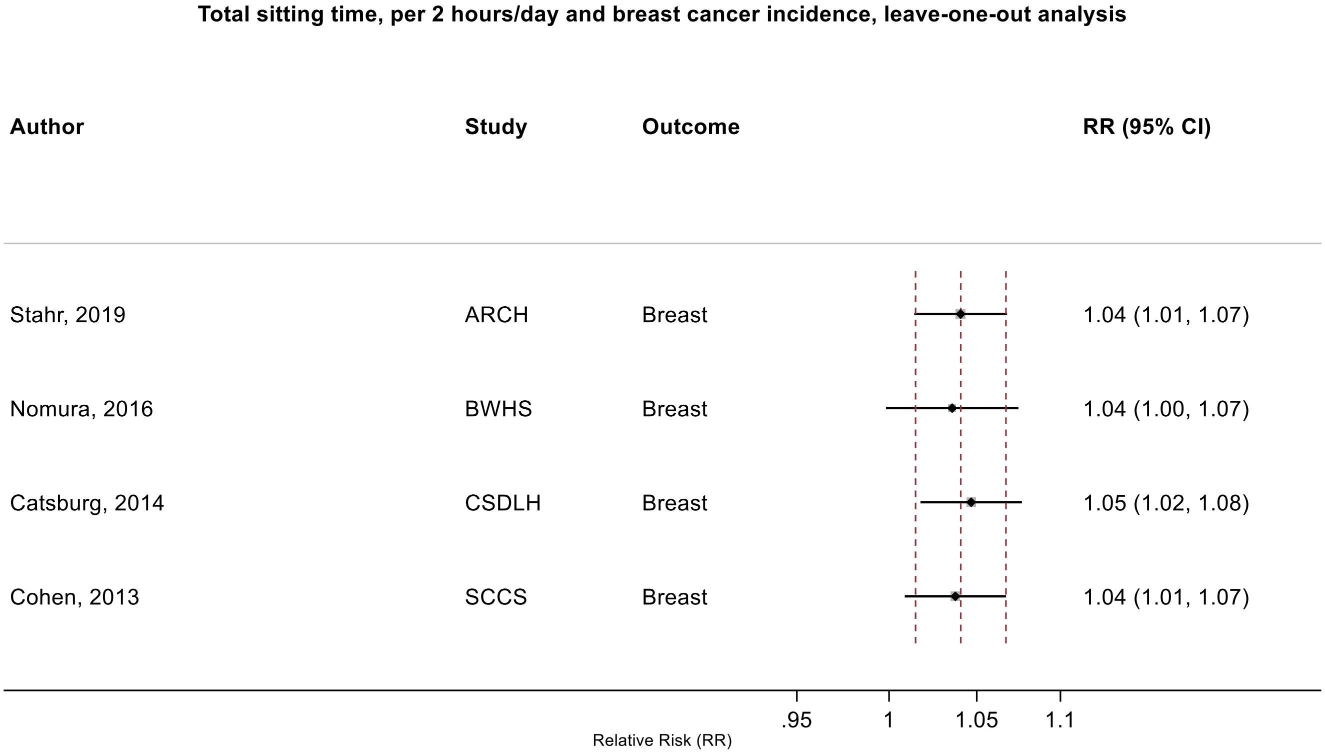

Figure legend: Forest plot shows the linear dose-response results (per 2 hours of sedentary time) from the inverse variance DerSimonian-Laird random-effects model after exclusion of each study from the meta-analysis. For each study, the square represents the summary relative risk (RR) estimate and the horizontal line across the square the corresponding 95% confidence interval (CI) computed from a meta-analysis excluding that study. The dashed vertical lines represent the results of the main meta-analysis.

Study abbreviations: ARCH, Arkansas Rural Community Health Study; BWHS, Black Women's Health Study; CSDLH, Canadian Study of Diet, Lifestyle and Health; SCCS, Southern Community Cohort Study

Stahr 2019 (35117114)[85]; Nomura 2016 (27632430)[60]; Catsburg 2014 (24781974)[55]; Cohen 2013 (23576427)[46]

#### Supplementary Figure 49: Linear dose-response meta-analysis of the association between occupational sitting time and colorectal cancer incidence, leave-one-out sensitivity analysis.

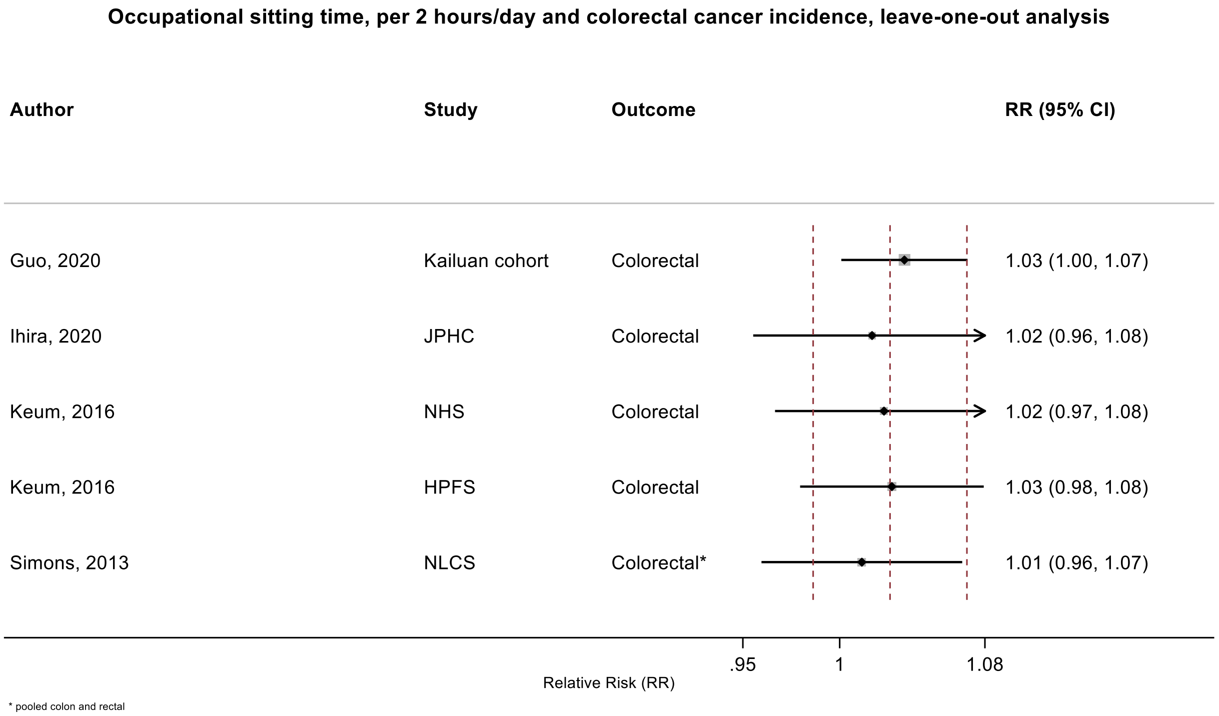

Figure legend: Forest plot shows the linear dose-response results (per 2 hours of sedentary time) from the inverse variance DerSimonian-Laird random-effects model after exclusion of each study from the meta-analysis. For each study, the square represents the summary relative risk (RR) estimate and the horizontal line across the square the corresponding 95% confidence interval (CI) computed from a meta-analysis excluding that study. The dashed vertical lines represent the results of the main meta-analysis.

Study abbreviations: JPHC, Japan Public Health Center-Based Prospective Study; NLCS, Netherlands Cohort Study on Diet and Cancer; HPFS, Health Professionals Follow-up Study; NHS, Nurses' Health Study

Guo 2020 (31773920)[76]; Ihira 2020 (31925977)[77]; Keum 2016 (26649988)[58]; Simons 2013 (23420352)[45]

#### Supplementary Figure 50: Linear dose-response meta-analysis of the association between occupational sitting time and pancreatic cancer incidence, leave-one-out sensitivity analysis.

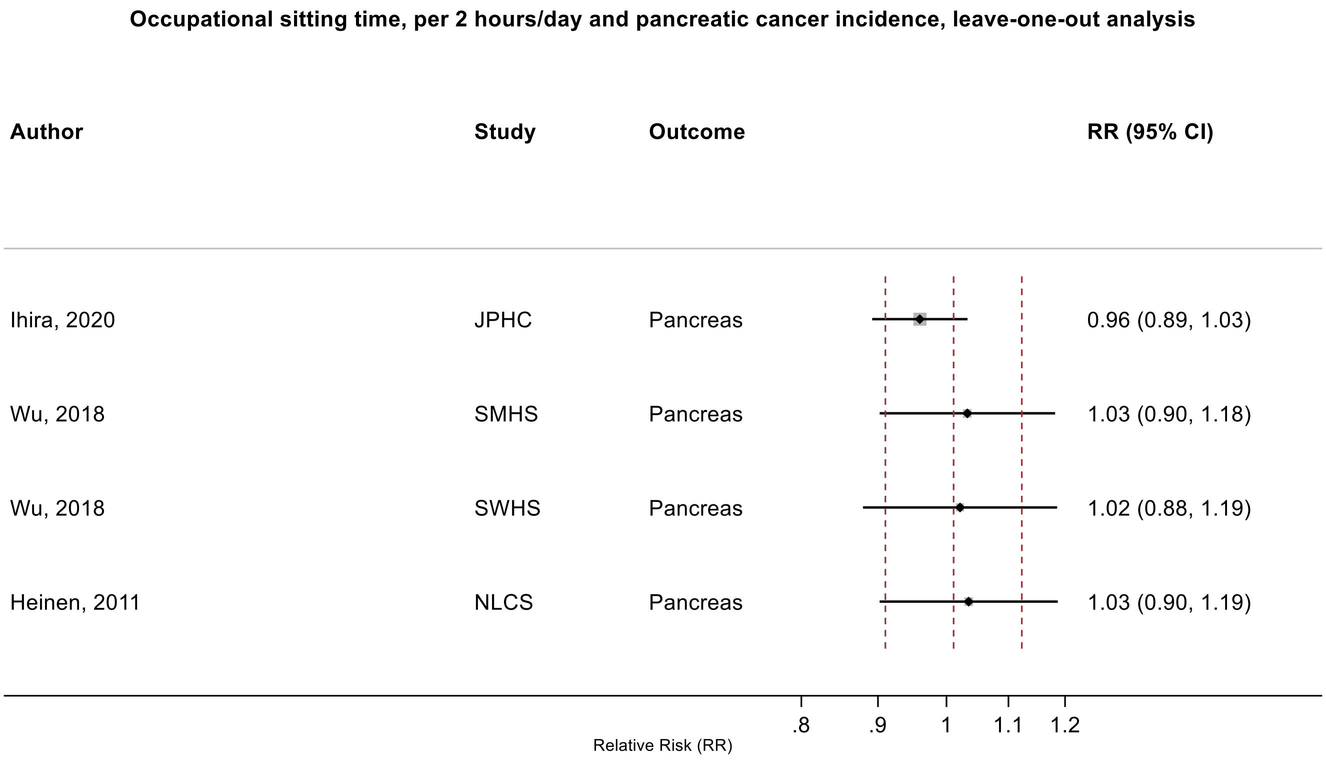

Figure legend: Forest plot shows the linear dose-response results (per 2 hours of sedentary time) from the inverse variance DerSimonian-Laird random-effects model after exclusion of each study from the meta-analysis. For each study, the square represents the summary relative risk (RR) estimate and the horizontal line across the square the corresponding 95% confidence interval (CI) computed from a meta-analysis excluding that study. The dashed vertical lines represent the results of the main meta-analysis.

Study abbreviations: JPHC, Japan Public Health Center-Based Prospective Study; NLCS, SWHS, Shanghai Women's Health Study; SMHS, SWHS, Shanghai Men's Health Study; NLCS, Netherlands Cohort Study on Diet and Cancer

Ihira 2020 (31925977)[77]; Wu 2018 (29475964)[65]; Heinen 2011 (21955648)[43]

**Supplementary Figure 51: Linear dose-response meta-analysis of the association between occupational sitting time and breast cancer incidence, leave-one-out sensitivity analysis.**

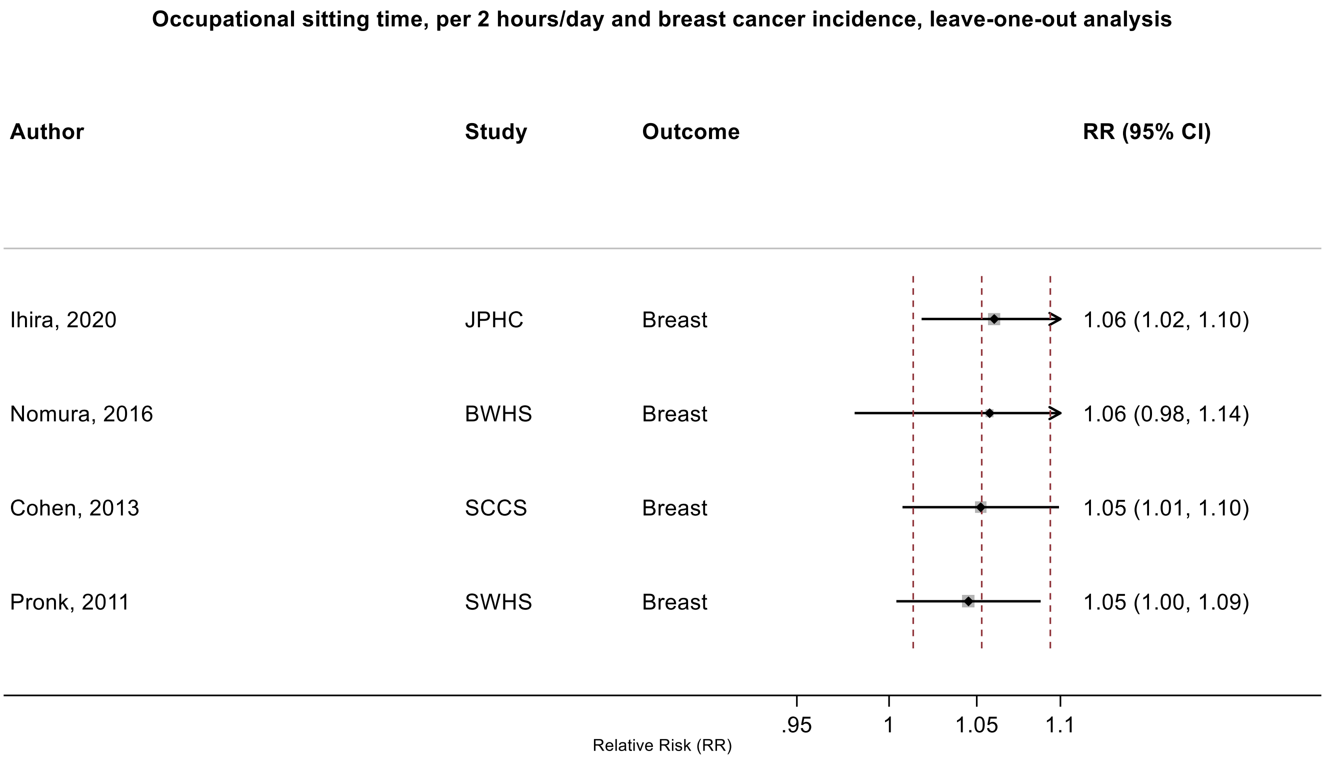

Figure legend: Forest plot shows the linear dose-response results (per 2 hours of sedentary time) from the inverse variance DerSimonian-Laird random-effects model after exclusion of each study from the meta-analysis. For each study, the square represents the summary relative risk (RR) estimate and the horizontal line across the square the corresponding 95% confidence interval (CI) computed from a meta-analysis excluding that study. The dashed vertical lines represent the results of the main meta-analysis.

Study abbreviations: JPHC, Japan Public Health Center-Based Prospective Study; BWHS, Black Women's Health Study; SCCS, Southern Community Cohort Study; SWHS, Shanghai Women’s Health Study

Ihira 2020 (31925977)[77]; Nomura 2016 (27632430)[60]; Cohen 2013 (23576427)[46]; Pronk 2011 (21934685)[42]

#### (B) Mixed definitions of sedentary, sitting, and screen time

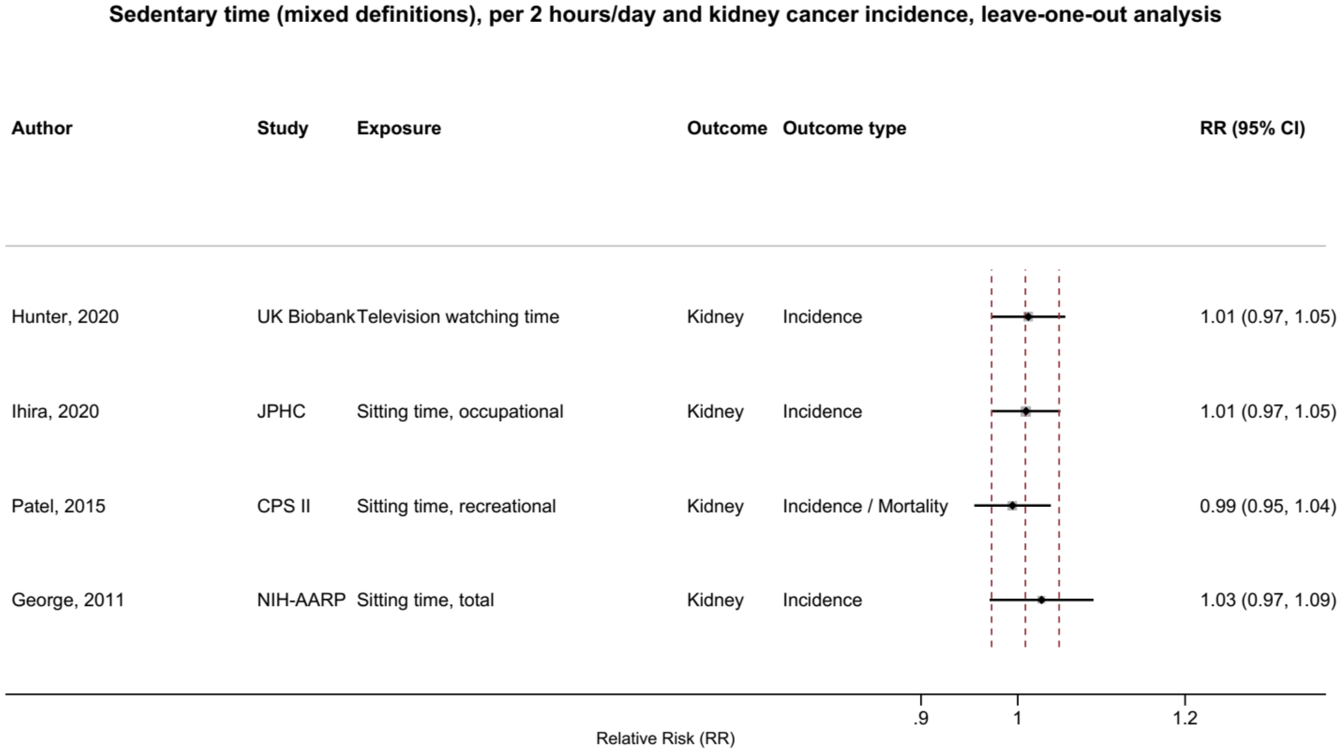

Figure legend: Forest plot shows the linear dose-response results (per 2 hours of sedentary time) from the inverse variance DerSimonian-Laird random-effects model after exclusion of each study from the meta-analysis. For each study, the square represents the summary relative risk (RR) estimate and the horizontal line across the square the corresponding 95% confidence interval (CI) computed from a meta-analysis excluding that study. The dashed vertical lines represent the results of the main meta-analysis.

Study abbreviations: CPS II, Cancer Prevention Study II; JPHC, Japan Public Health Center-Based Prospective Study; NIH-AARP, National Institute of Health – American Association for Retired Persons Diet and Health Study

George 2011 (21737302)[41]; Patel 2015 (26126627)[56]; Ihira 2020 (31925977)[77]; Hunter 2020 (32746843)[78]

#### Television watching time

##### Supplementary Figure 52: Linear dose-response meta-analysis of the association between television watching time and oesophageal cancer incidence.

**Television watching time, per 2 hours/day and oesophageal cancer incidence**

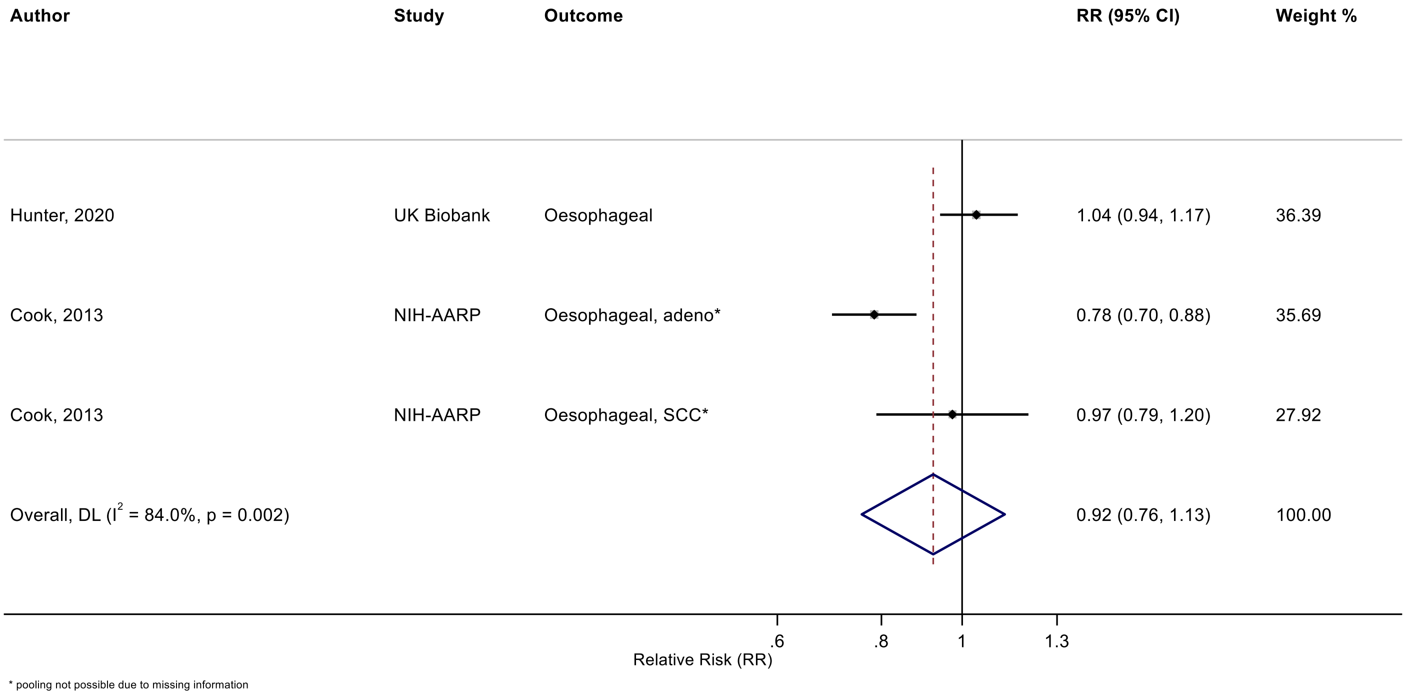

Figure legend: Forest plot shows the linear dose-response results (per 2 hours/day of sedentary time) from the inverse variance DerSimonian-Laird random-effects model. The diamond represents the summary relative risk (RR) estimate and the diamond's width the 95% confidence interval (CI). Each square and the horizontal line across the square represent the RR estimate of the individual study and its 95%CI.

Study abbreviations: NIH-AARP, National Institute of Health + American Association of Retired Persons Diet and Health Study

Hunter 2020 (32746843)[78]; Cook 2013 (24367697)[52]

#### Supplementary Figure 53: Linear dose-response meta-analysis of the association between television watching time and stomach cancer incidence.

**Television watching time, per 2 hours/day and stomach cancer incidence**

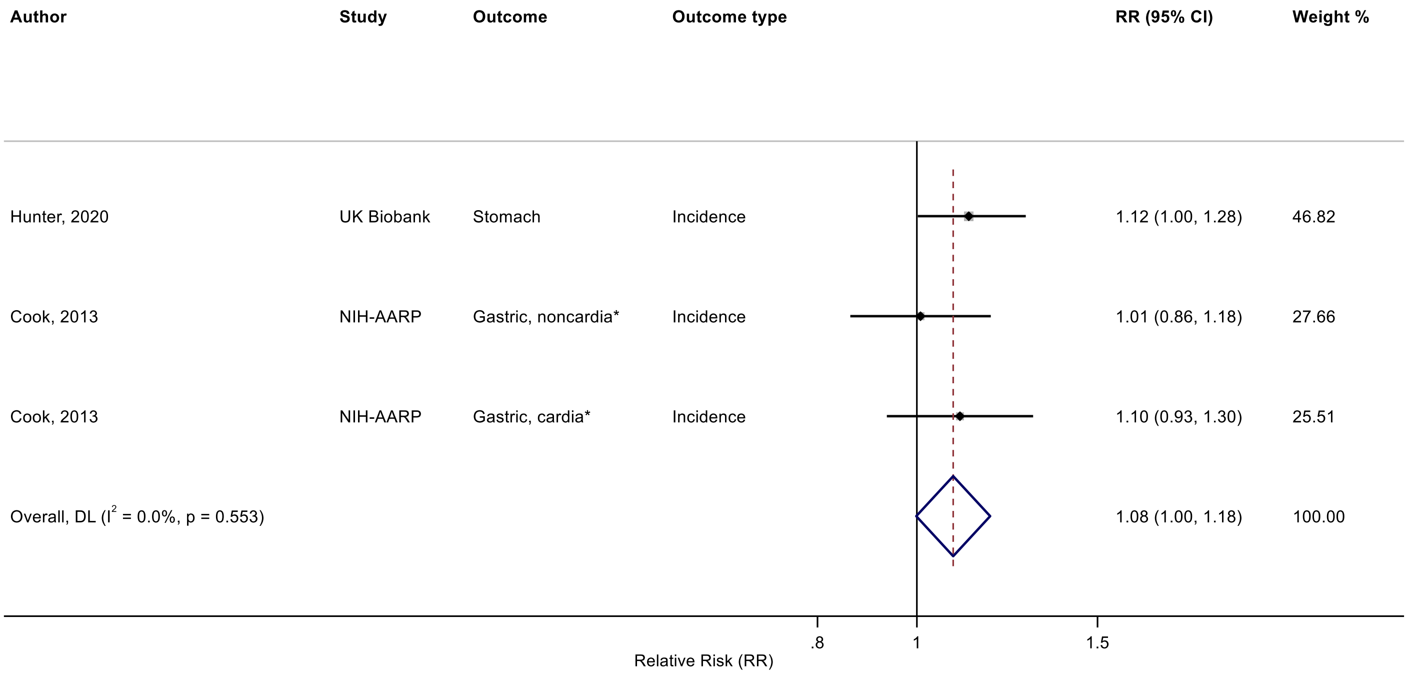

* pooling not possible due to missing information

Figure legend: Forest plot shows the linear dose-response results (per 2 hours/day of sedentary time) from the inverse variance DerSimonian-Laird random-effects model. The diamond represents the summary relative risk (RR) estimate and the diamond's width the 95% confidence interval (CI). Each square and the horizontal line across the square represent the RR estimate of the individual study and its 95%CI.

Study abbreviations: NIH-AARP, National Institute of Health + American Association of Retired Persons Diet and Health Study

Hunter 2020 (32746843)[78]; Cook 2013 (24367697)[52]

Supplementary Materials

#### Supplementary Figure 54: Linear dose-response meta-analysis of the association between television watching time and colorectal cancer incidence.

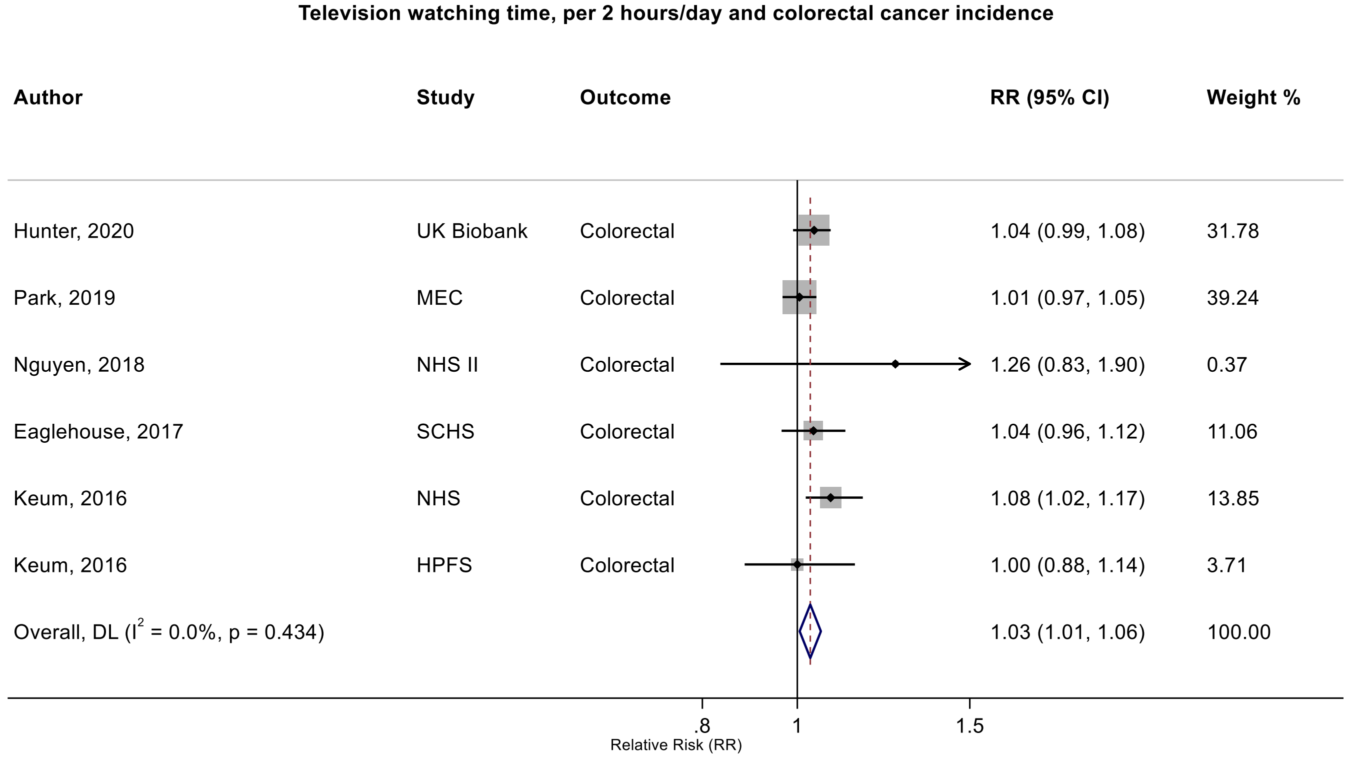

Figure legend: Forest plot shows the linear dose-response results (per 2 hours/day of sedentary time) from the inverse variance DerSimonian-Laird random-effects model. The diamond represents the summary relative risk (RR) estimate and the diamond's width the 95% confidence interval (CI). Each square and the horizontal line across the square represent the RR estimate of the individual study and its 95%CI.

Study abbreviations: MEC, Multiethnic Cohort Study; NHS, Nurses' Health Study; SCHS, Singapore Chinese Health study; HPFS, Health Professionals Follow-up Study

Hunter 2020 (32746843)[78]; Park 2019 (30910780)[73]; Nguyen 2018 (30740587)[71]; Eaglehouse 2017 (28542077)[62]; Keum 2016 (26649988)[58]

#### Supplementary Figure 55: Linear dose-response meta-analysis of the association between television watching time and lung cancer incidence.

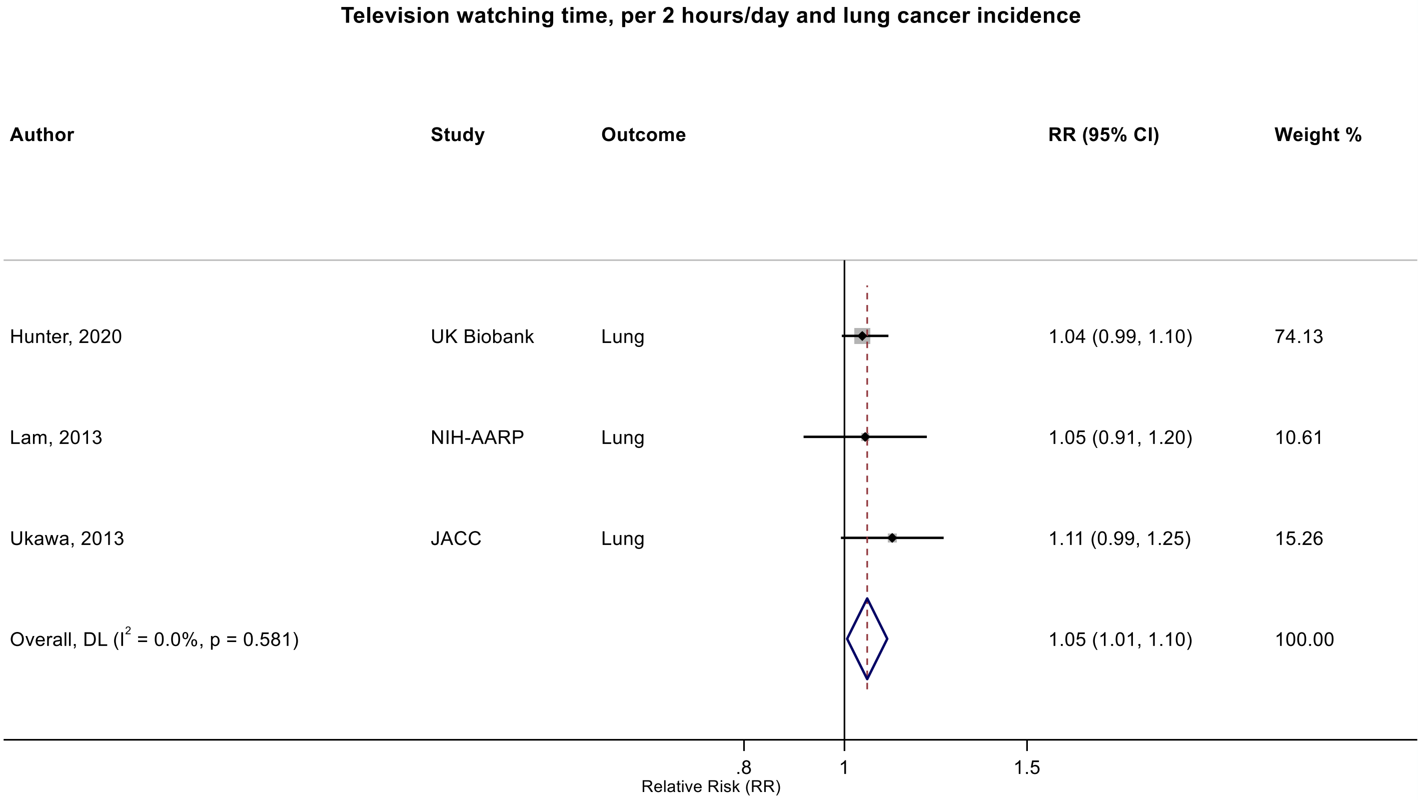

Figure legend: Forest plot shows the linear dose-response results (per 2 hours/day of sedentary time) from the inverse variance DerSimonian-Laird random-effects model. The diamond represents the summary relative risk (RR) estimate and the diamond's width the 95% confidence interval (CI). Each square and the horizontal line across the square represent the RR estimate of the individual study and its 95%CI.

Study abbreviations: NIH-AARP, National Institute of Health + American Association of Retired Persons Diet and Health Study; JACC, Japan Collaborative Cohort study

Hunter 2020 (32746843)[78]; Lam 2013 (23940620)[48]; Ukawa 2013 (23686441)[47]

**Supplementary Figure 56: Linear dose-response meta-analysis of the association between television watching time and breast cancer incidence.**

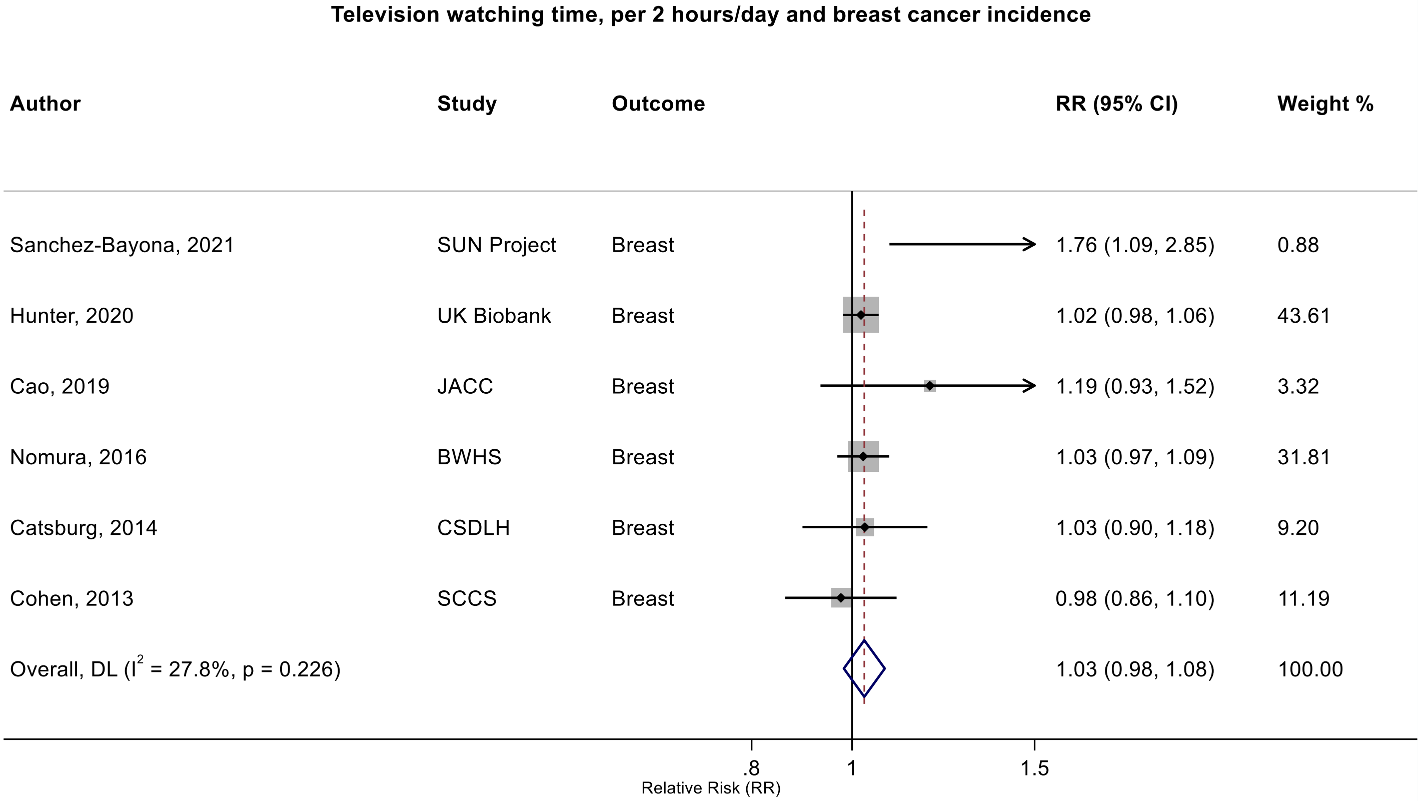

Figure legend: Forest plot shows the linear dose-response results (per 2 hours/day of sedentary time) from the inverse variance DerSimonian-Laird random-effects model. The diamond represents the summary relative risk (RR) estimate and the diamond's width the 95% confidence interval (CI). Each square and the horizontal line across the square represent the RR estimate of the individual study and its 95%CI.

Study abbreviations: SUN, Seguimiento Universidad de Navarra Cohort Study; JACC, Japan Collaborative Cohort study; BWHS, Black Women's Health Study; CSDLH, Canadian Study of Diet, Lifestyle and Health; SCCS, Southern Community Cohort Study

Sanchez-Bayona 2021 (33798533)[82]; Hunter 2020 (32746843)[78]; Cao 2019 (30913861)[74]; Nomura 2016 (27632430)[60]; Catsburg 2014 (24781974)[55]; Cohen 2013 (23576427)[46]

**Supplementary Figure 57: Linear dose-response meta-analysis of the association between television watching time and endometrial cancer incidence.**

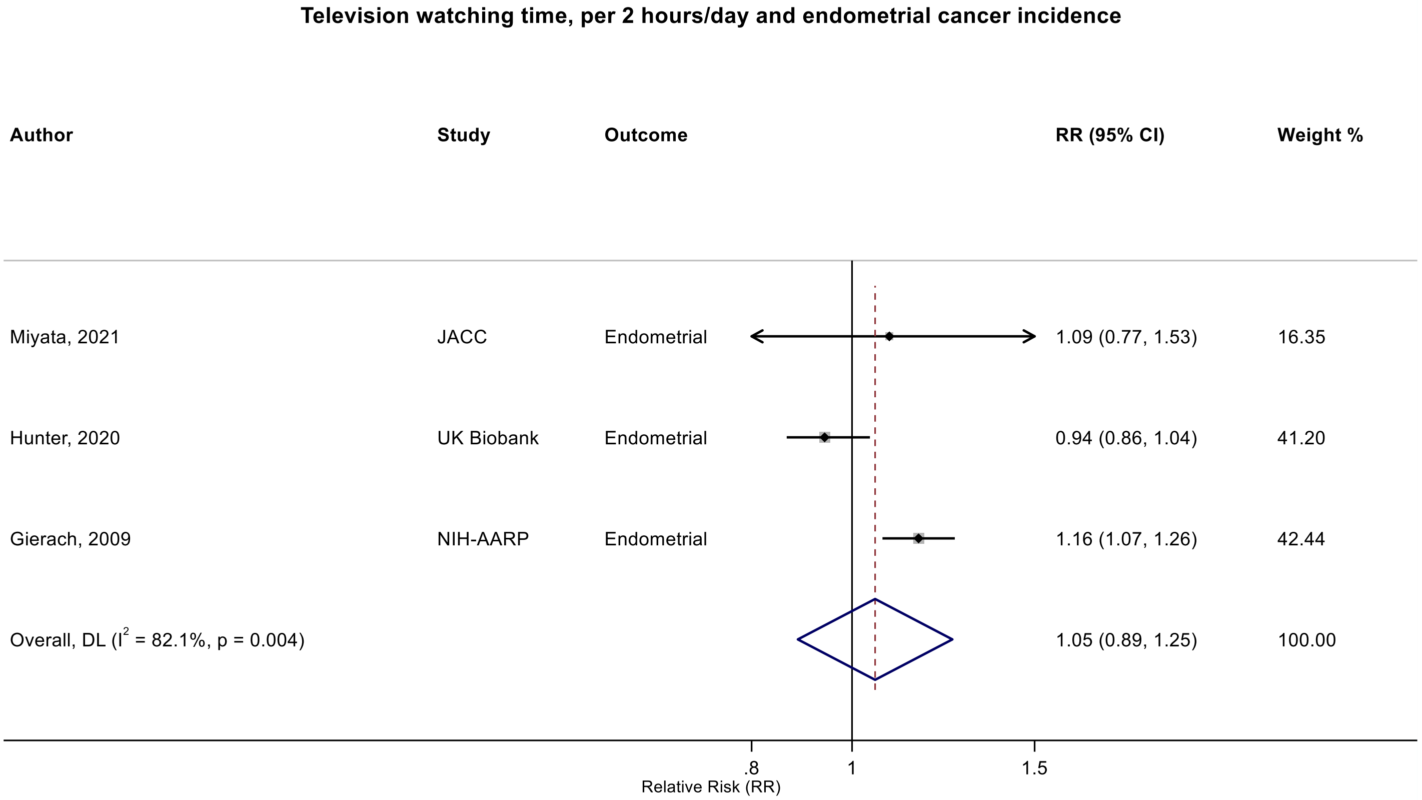

Figure legend: Forest plot shows the linear dose-response results (per 2 hours/day of sedentary time) from the inverse variance DerSimonian-Laird random-effects model. The diamond represents the summary relative risk (RR) estimate and the diamond's width the 95% confidence interval (CI). Each square and the horizontal line across the square represent the RR estimate of the individual study and its 95%CI.

Study abbreviations: JACC, Japan Collaborative Cohort study; NIH-AARP, National Institute of Health + American Association of Retired Persons Diet and Health Study

Miyata 2021 (32963209)[79]; Hunter 2020 (32746843)[78]; Gierach 2009 (19123463)[38]

#### Supplementary Figure 58: Linear dose-response meta-analysis of the association between television watching time and ovarian cancer incidence.

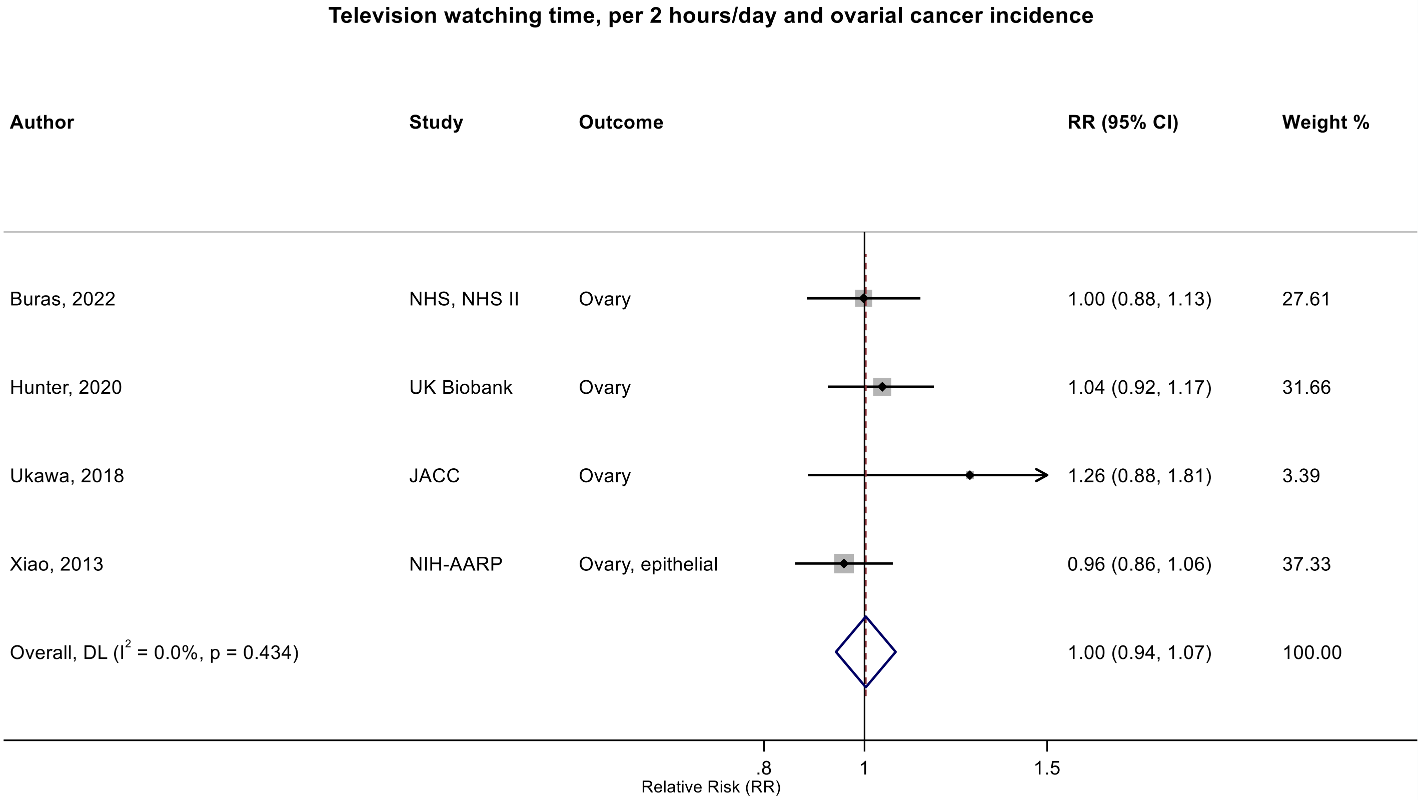

Figure legend: Forest plot shows the linear dose-response results (per 2 hours/day of sedentary time) from the inverse variance DerSimonian-Laird random-effects model. The diamond represents the summary relative risk (RR) estimate and the diamond's width the 95% confidence interval (CI). Each square and the horizontal line across the square represent the RR estimate of the individual study and its 95%CI.

Study abbreviations: NHS, Nurses' Health Study;  JACC, Japan Collaborative Cohort study; NIH-AARP, National Institute of Health + American Association of Retired Persons Diet and Health Study

Buras 2022 (35094053)[84]; Hunter 2020 (32746843)[78]; Ukawa 2018 (29340890)[64]; Xiao 2013 (23966580)[49]

#### Supplementary Figure 59: Linear dose-response meta-analysis of the association between television watching time and prostate cancer incidence.

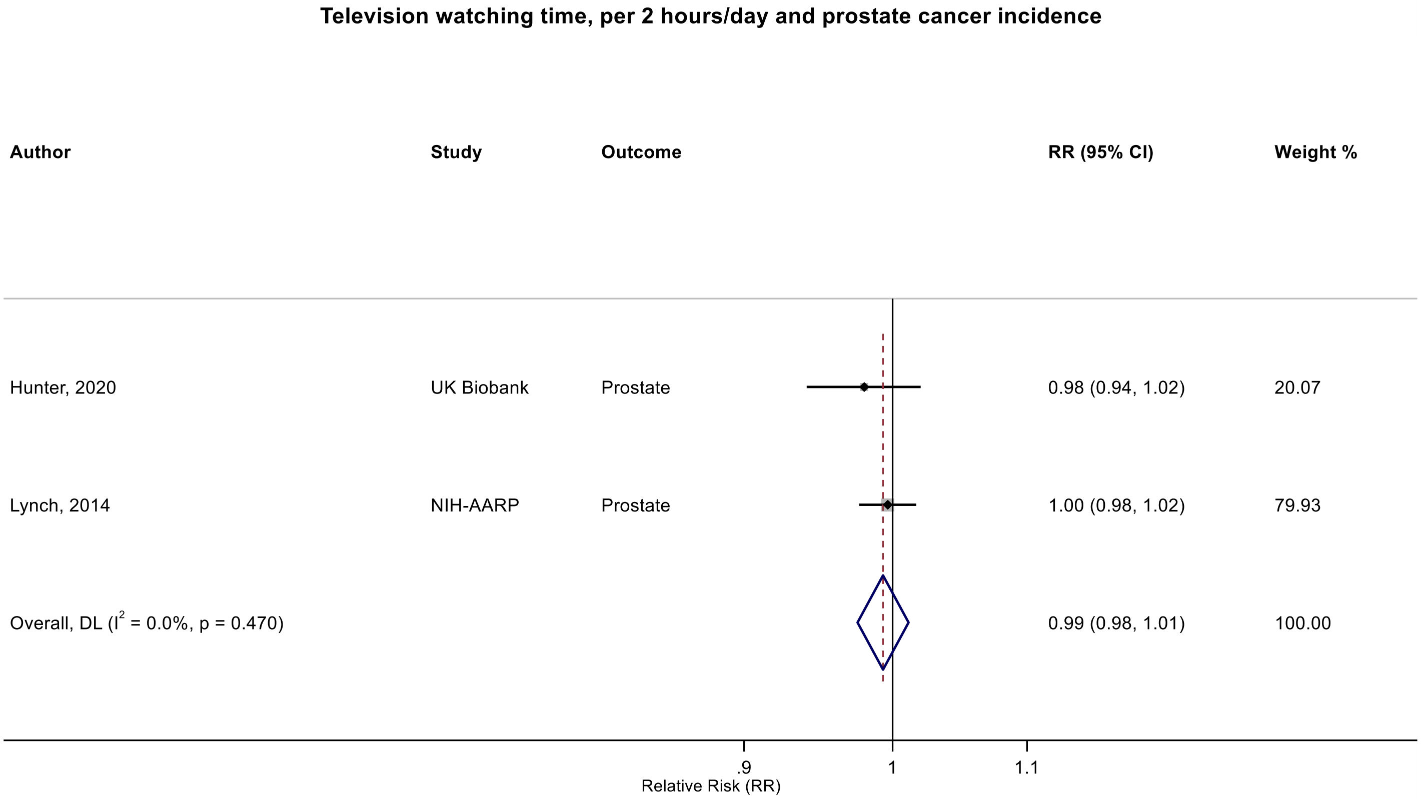

Figure legend: Forest plot shows the linear dose-response results (per 2 hours/day of sedentary time) from the inverse variance DerSimonian-Laird random-effects model. The diamond represents the summary relative risk (RR) estimate and the diamond's width the 95% confidence interval (CI). Each square and the horizontal line across the square represent the RR estimate of the individual study and its 95%CI.

Study abbreviations: NIH-AARP, National Institute of Health + American Association of Retired Persons Diet and Health Study

Hunter 2020 (32746843)[78]; Lynch 2014 (24526287)[53]

#### Supplementary Figure 60: Linear dose-response meta-analysis of the association between television watching time and kidney cancer incidence.

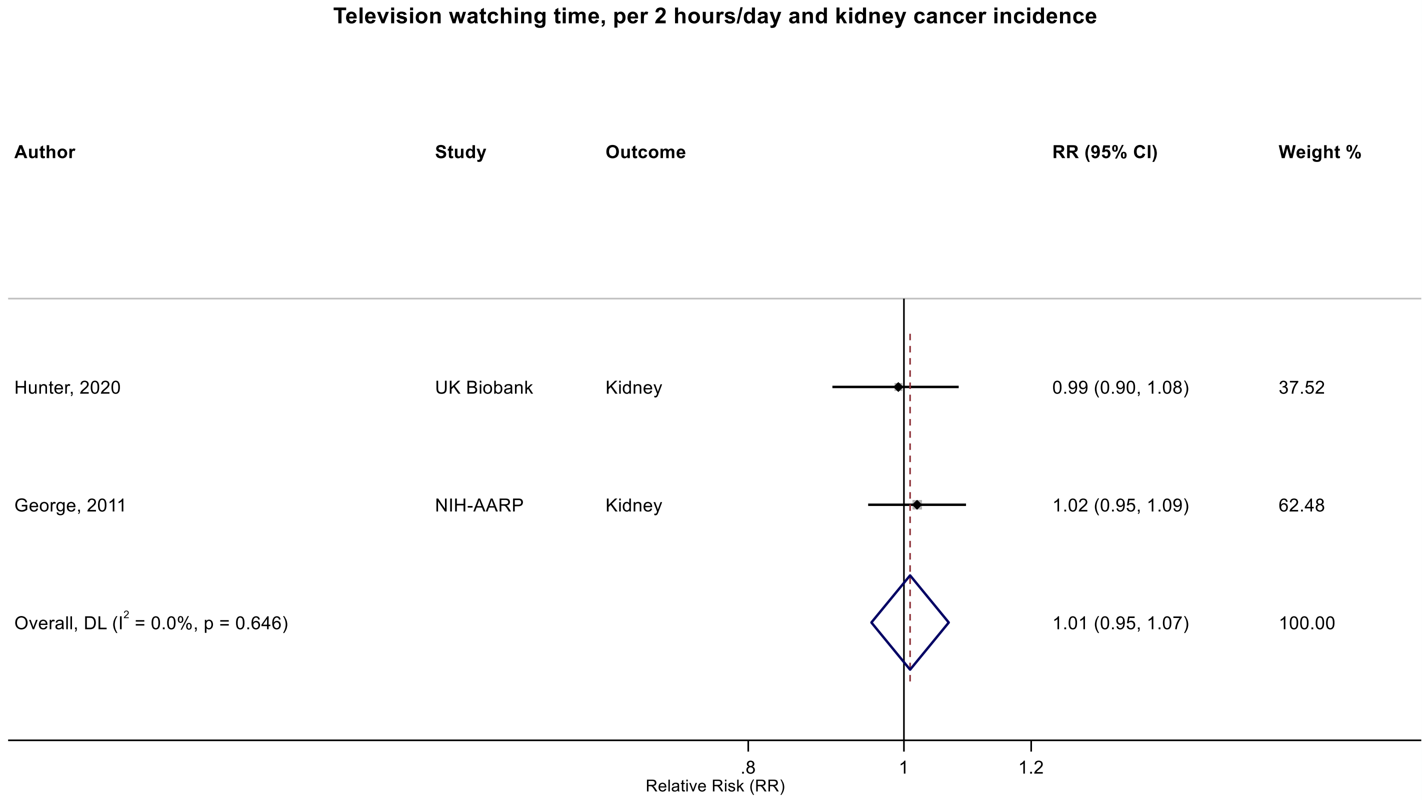

Figure legend: Forest plot shows the linear dose-response results (per 2 hours/day of sedentary time) from the inverse variance DerSimonian-Laird random-effects model. The diamond represents the summary relative risk (RR) estimate and the diamond's width the 95% confidence interval (CI). Each square and the horizontal line across the square represent the RR estimate of the individual study and its 95%CI.

Study abbreviations: NIH-AARP, National Institute of Health + American Association of Retired Persons Diet and Health Study

Hunter 2020 (32746843)[78]; George 2011 (21737302)[41]

**Supplementary Figure 61: Non-linear dose-response meta-analysis of the association between total television watching time and colon cancer incidence.**

The solid line represents the estimated summary dose-response relationship and the short-dashed line the 95% confidence intervals. The tick marks inside the x-axis indicate the total television watching time hours/days values for which relative risk estimate(s) were available. Non-linear curve was estimated using restricted cubic spline regressions with three knots placed at fixed percentiles (10%, 50%, and 90%) of the exposure distribution, which were pooled by fitting one-stage random-effects mixed models. 2 sitting hours/day was chosen as reference. The table shows selected television watching time hours/day values and their corresponding relative risk (95% confidence intervals) estimated in the non-linear dose-response meta-analysis.
Abbreviations: CI: confidence interval; RR, relative risk.

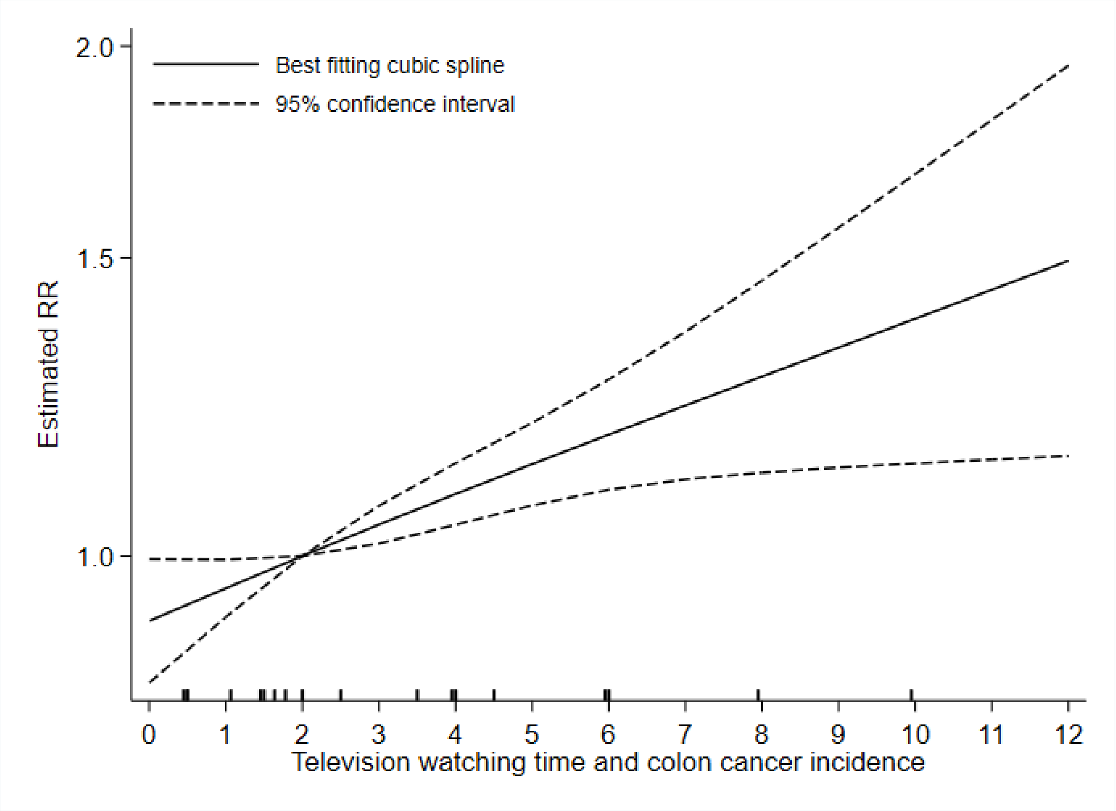

$P_{non\text{-}linearity}$ = 0.891

n = 5 studies (5 publications)

8,420 cases

| Television watching time (hours/day) | RR (95% CI) |
|---|---|
| 0 | 0.92 (0.84, 1.00) |
| 1 | 0.96 (0.92, 1.00) |
| 2 | 1 (Reference) |
| 3 | 1.04 (1.02, 1.07) |
| 4 | 1.09 (1.04, 1.13) |
| 5 | 1.13 (1.07, 1.20) |
| 6 | 1.18 (1.09, 1.27) |
| 7 | 1.23 (1.11, 1.36) |
| 8 | 1.28 (1.12, 1.45) |
| 9 | 1.33 (1.13, 1.56) |
| 10 | 1.38 (1.13, 1.68) |
| 11 | 1.44 (1.14, 1.81) |
| 12 | 1.49 (1.15, 1.95) |

**Supplementary Figure 62: Non-linear dose-response meta-analysis of the association between total television watching time and colorectal cancer incidence.**

The solid line represents the estimated summary dose-response relationship and the short-dashed line the 95% confidence intervals. The tick marks inside the x-axis indicate the total television watching time hours/days values for which relative risk estimate(s) were available. Non-linear curve was estimated using restricted cubic spline regressions with three knots placed at fixed percentiles (10%, 50%, and 90%) of the exposure distribution, which were pooled by fitting one-stage random-effects mixed models. 2 sitting hours/day was chosen as reference. The table shows selected television watching time hours/day values and their corresponding relative risk (95% confidence intervals) estimated in the non-linear dose-response meta-analysis. Abbreviations: CI: confidence interval; RR, relative risk.

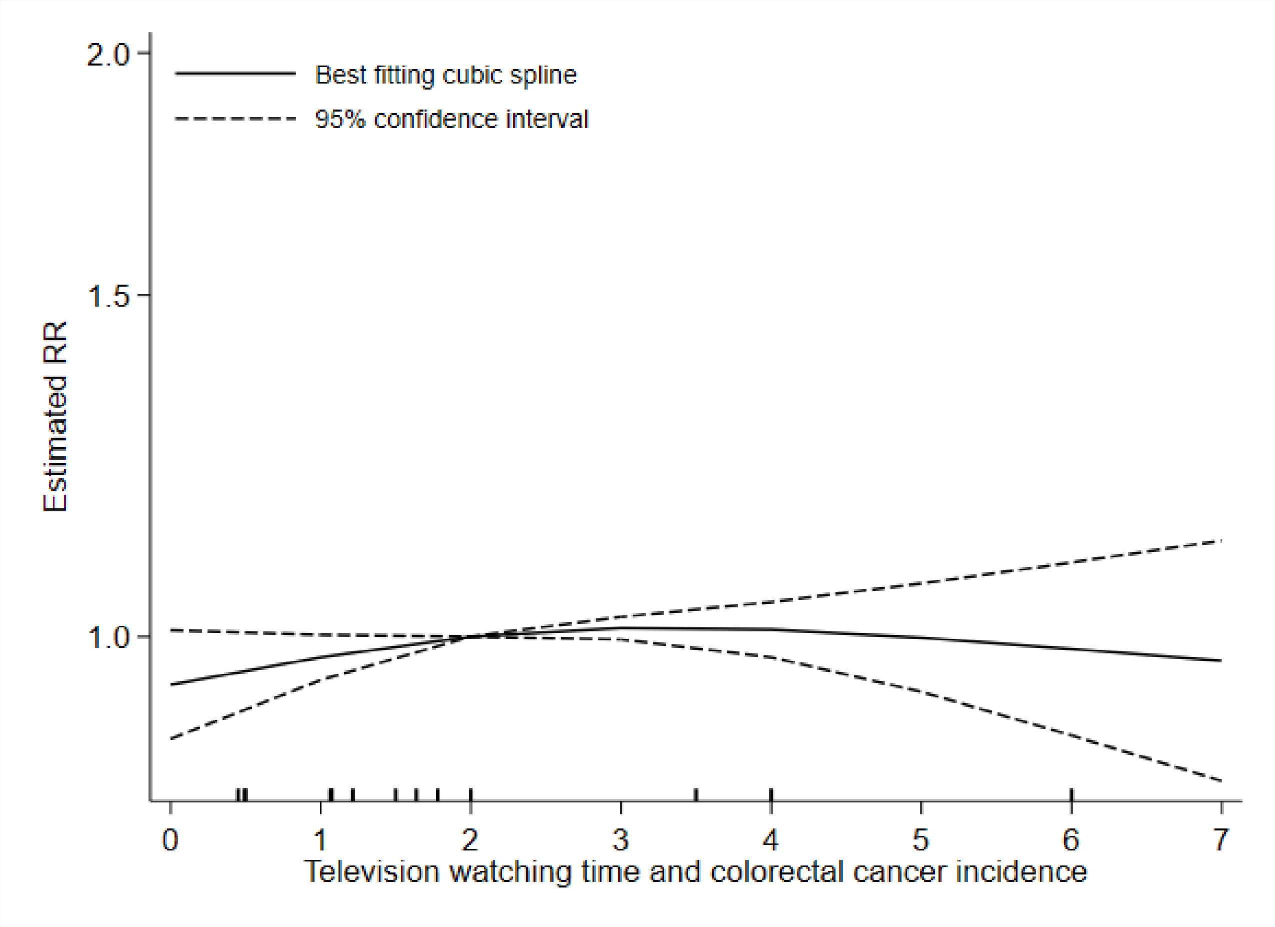

$P_{non\text{-}linearity}$ = 0.218

n = 6 studies (5 publications)

11,619 cases

| Television watching time (hours/day) | RR (95% CI) |
|---|---|
| 0 | 0.94 (0.89, 1.01) |
| 1 | 0.98 (0.95, 1.00) |
| 2 | 1 (Reference) |
| 3 | 1.01 (1.00, 1.02) |
| 4 | 1.01 (0.98, 1.04) |
| 5 | 1.00 (0.94, 1.07) |
| 6 | 0.99 (0.89, 1.09) |
| 7 | 0.97 (0.84, 1.12) |

**Supplementary Figure 63: Non-linear dose-response meta-analysis of the association between total television watching time and breast cancer incidence.**

The solid line represents the estimated summary dose-response relationship and the short-dashed line the 95% confidence intervals. The tick marks inside the x-axis indicate the total television watching time hours/days values for which relative risk estimate(s) were available. Non-linear curve was estimated using restricted cubic spline regressions with three knots placed at fixed percentiles (10%, 50%, and 90%) of the exposure distribution, which were pooled by fitting one-stage random-effects mixed models. 2 sitting hours/day was chosen as reference. The table shows selected television watching time hours/day values and their corresponding relative risk (95% confidence intervals) estimated in the non-linear dose-response meta-analysis.

Abbreviations: CI: confidence interval; RR, relative risk.

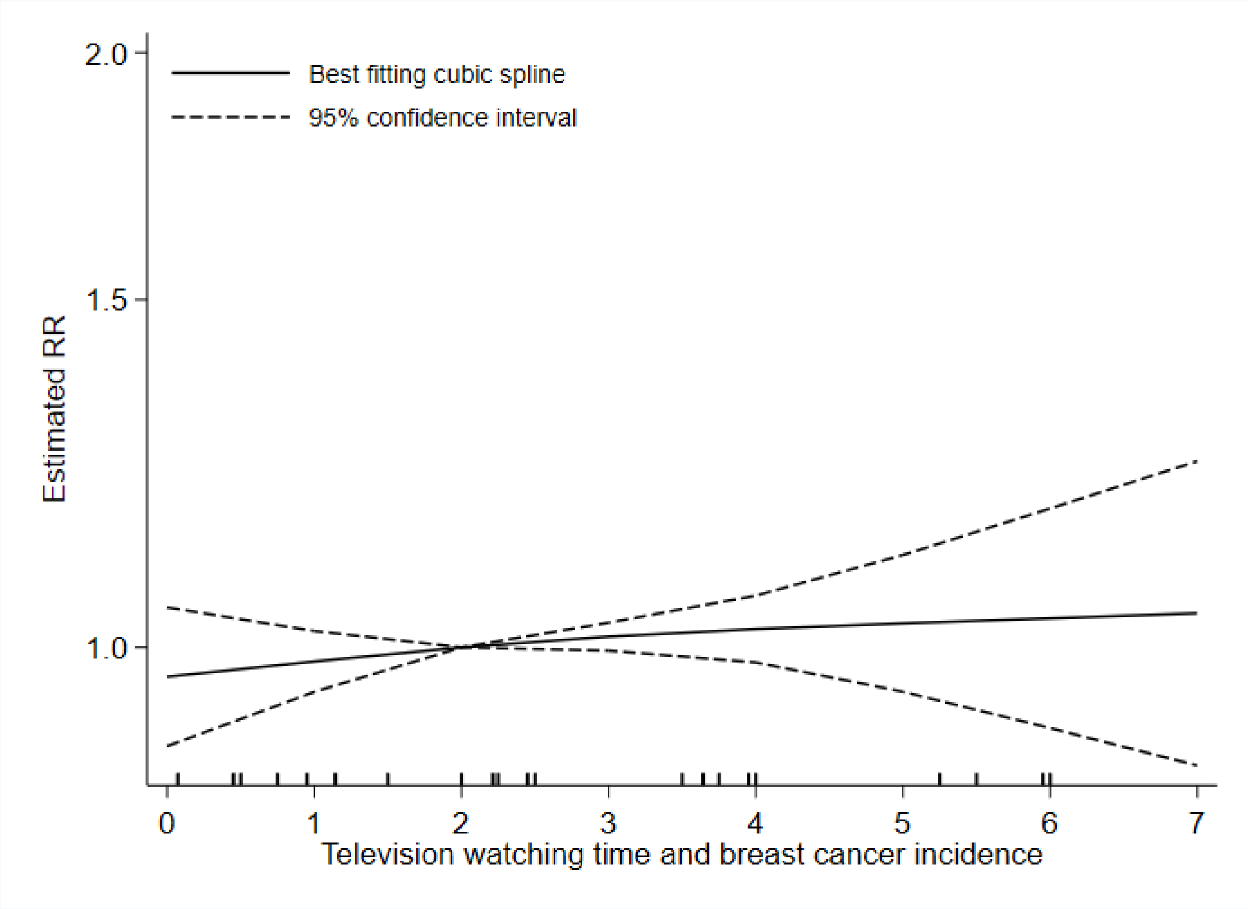

$P_{non\text{-}linearity}$ = 0.794

n = 6 studies (6 publications)

10,372 cases

| Television watching time (hours/day) | RR (95% CI) |
|---|---|
| 0 | 0.97 (0.89, 1.05) |
| 1 | 0.98 (0.95, 1.02) |
| 2 | 1 (Reference) |
| 3 | 1.01 (1.00, 1.03) |
| 4 | 1.02 (0.98, 1.06) |
| 5 | 1.03 (0.95, 1.11) |
| 6 | 1.03 (0.91, 1.18) |
| 7 | 1.04 (0.87, 1.24) |

#### Supplementary Figure 64: Categorical highest versus lowest meta-analysis of the association between television watching time and bladder cancer incidence.

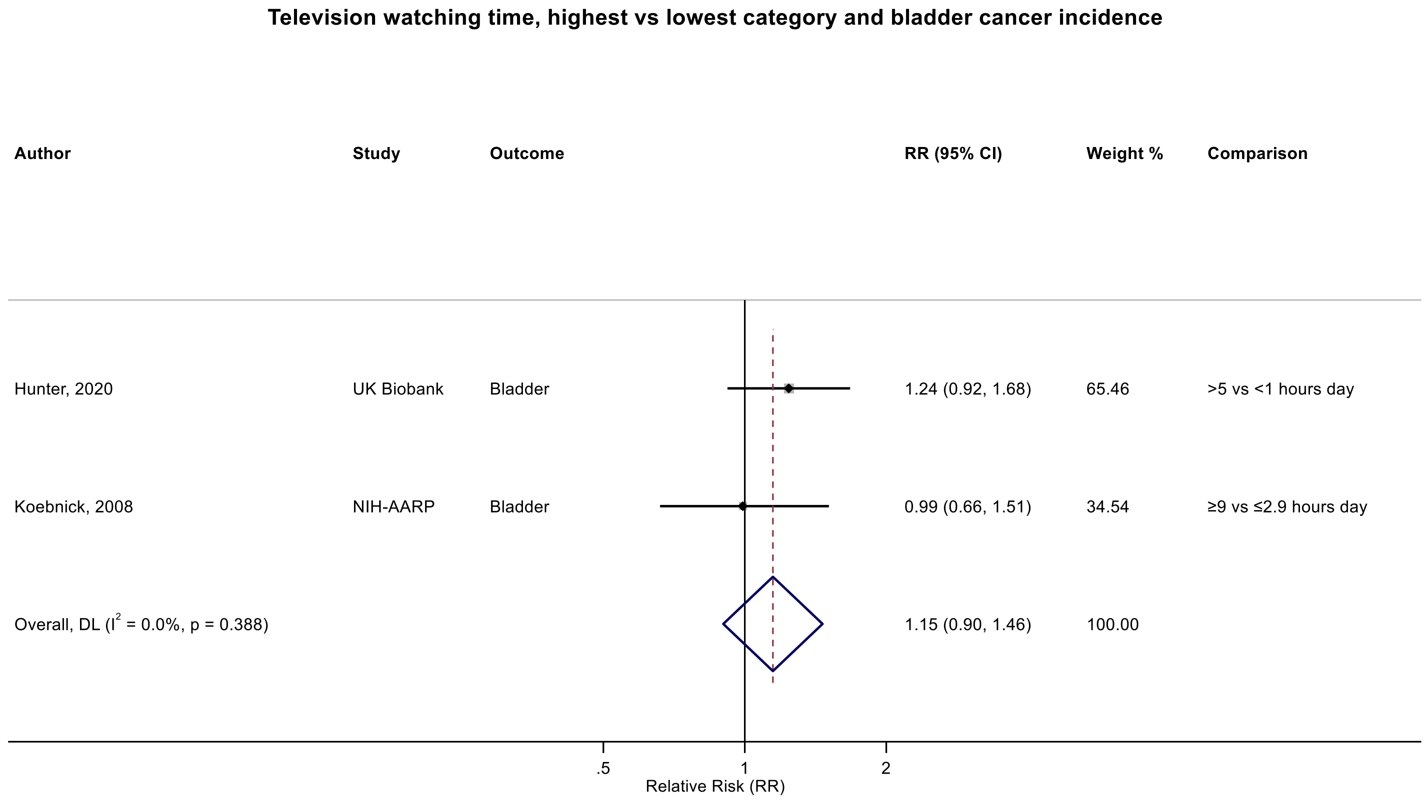

Figure legend: Forest plot shows the categorical comparison results from the individual studies for the associations with the largest contrast, as reported in the respective publication. Each square and the horizontal line across the square represent the relative risk (RR) estimate of the individual study and its 95% confidence interval (CI).

Study abbreviations: NIH-AARP, National Institute of Health + American Association of Retired Persons Diet and Health Study

Hunter 2020 (32746843)[78]; Koebnick 2008 (18483344)[37]

#### Supplementary Figure 65: Linear dose-response meta-analysis of the association between television watching time and colorectal cancer incidence, by anatomical subsite.

**Television watching time, per 2 hours/day and colorectal cancer incidence, by anatomical subsite**

| Author | Study | Outcome | RR (95% CI) | Weight % |
|---|---|---|---|---|
| **Colon** | | | | |
| Hunter, 2020 | UK Biobank | Colon | 1.08 (1.02, 1.12) | 36.17 |
| Eaglehouse, 2017 | SCHS | Colon | 1.04 (0.94, 1.14) | 8.76 |
| Keum, 2016 | NHS | Colon | 1.11 (0.98, 1.25) | 5.59 |
| Keum, 2016 | HPFS | Colon | 1.08 (0.91, 1.28) | 2.99 |
| Howard, 2008 | NIH-AARP | Colon | 1.08 (1.04, 1.13) | 46.49 |
| Subgroup, DL ($I^2$ = 0.0%, p = 0.938) | | | 1.08 (1.05, 1.11) | 100.00 |
| **Rectal** | | | | |
| Hunter, 2020 | UK Biobank | Rectal | 0.98 (0.90, 1.06) | 54.86 |
| Eaglehouse, 2017 | SCHS | Rectal | 1.04 (0.92, 1.17) | 24.37 |
| Keum, 2016 | NHS | Rectal | 1.08 (0.92, 1.25) | 15.09 |
| Keum, 2016 | HPFS | Rectal | 1.06 (0.83, 1.37) | 5.68 |
| Subgroup, DL ($I^2$ = 0.0%, p = 0.663) | | | 1.01 (0.95, 1.08) | 100.00 |
| Heterogeneity between groups: p = 0.063 | | | | |

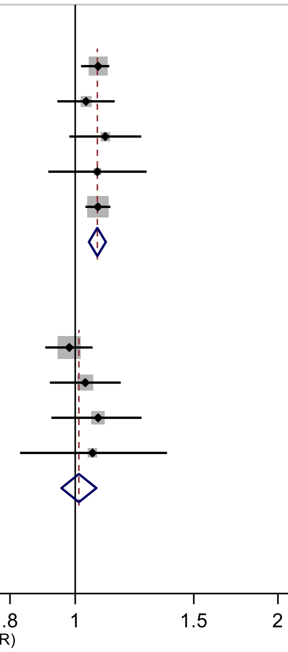

Figure legend: Forest plot shows the linear dose-response results (per 2 hours/day of sedentary time) from the inverse variance DerSimonian-Laird random-effects model. The diamond represents the summary relative risk (RR) estimate and the diamond's width the 95% confidence interval (CI). Each square and the horizontal line across the square represent the RR estimate of the individual study and its 95%CI.

Study abbreviations: SCHS, Singapore Chinese Health study; NHS, Nurses' Health Study, HPFS, Health Professional Follow-up Study; NIH-AARP, National Institute of Health + American Association of Retired Persons Diet and Health Study

Hunter 2020 (32746843)[78]; Eaglehouse 2017 (28542077)[62]; Keum 2016 (26649988)[58]; Howard 2008 (18437512)[36]

#### Supplementary Figure 66: Linear dose-response meta-analysis of the association between television watching time and colorectal cancer incidence, by physical activity adjustment.

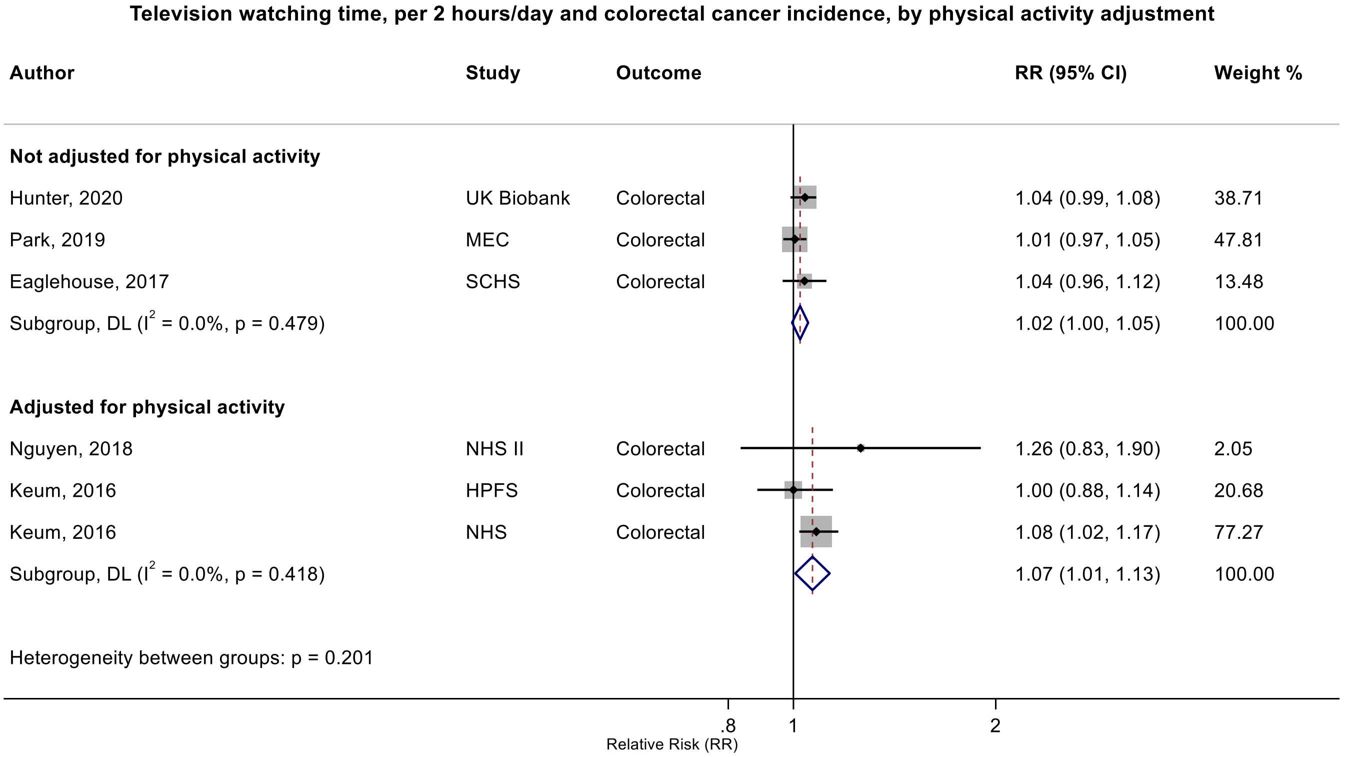

Figure legend: Forest plot shows the linear dose-response results (per 2 hours/day of sedentary time) from the inverse variance DerSimonian-Laird random-effects model. The diamond represents the summary relative risk (RR) estimate and the diamond's width the 95% confidence interval (CI). Each square and the horizontal line across the square represent the RR estimate of the individual study and its 95%CI.

Study abbreviations: MEC, Multiethnic Cohort Study; SCHS, Singapore Chinese Health study; NHS, Nurses' Health Study, HPFS, Health Professional Follow-up Study; NIH-AARP, National Institute of Health + American Association of Retired Persons Diet and Health Study

Hunter 2020 (32746843)[78]; Park 2019 (30910780)[73]; Eaglehouse 2017 (28542077)[62]; Nguyen 2018 (30740587)[71]; Keum 2016 (26649988)[58]

#### Supplementary Figure 67: Linear dose-response meta-analysis of the association between television watching time and colorectal cancer incidence, by adjustment for adiposity.

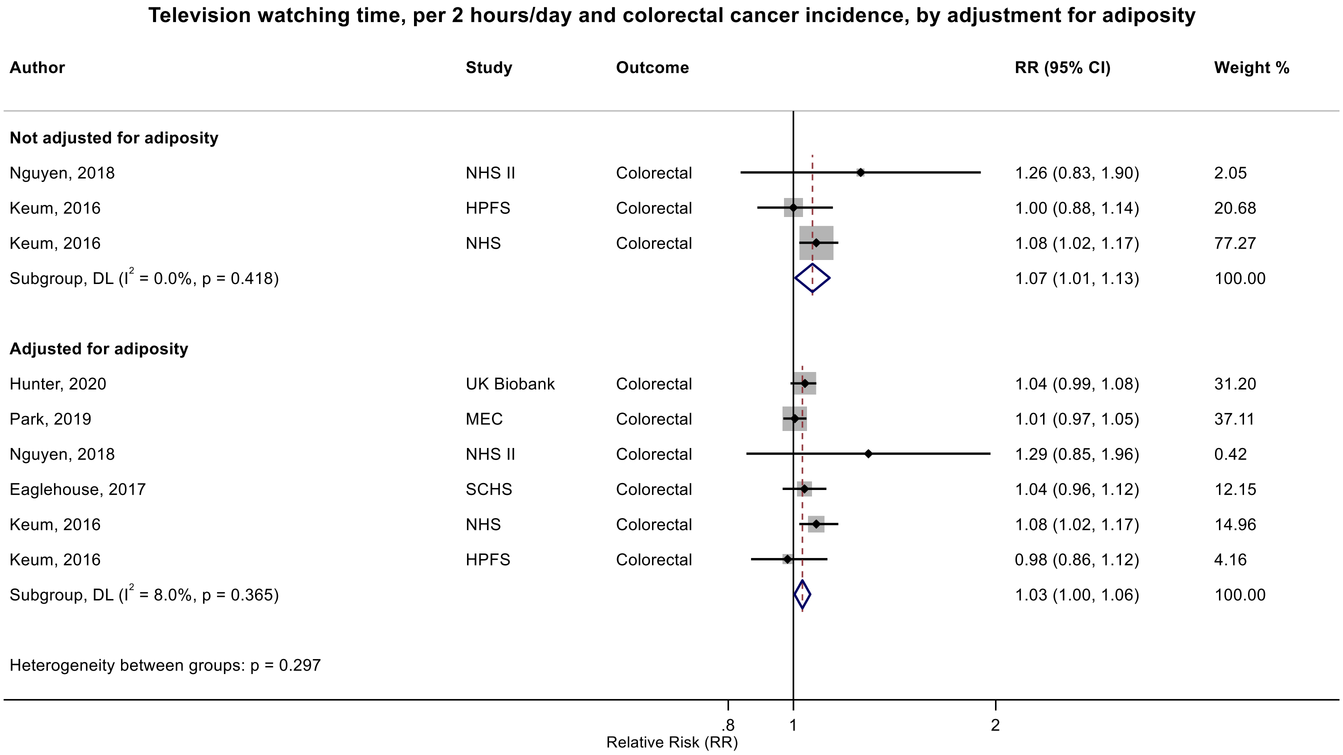

Figure legend: Forest plot shows the linear dose-response results (per 2 hours/day of sedentary time) from the inverse variance DerSimonian-Laird random-effects model. The diamond represents the summary relative risk (RR) estimate and the diamond's width the 95% confidence interval (CI). Each square and the horizontal line across the square represent the RR estimate of the individual study and its 95%CI.

Study abbreviations: MEC, Multiethnic Cohort Study; SCHS, Singapore Chinese Health study; NHS, Nurses' Health Study, HPFS, Health Professional Follow-up Study; NIH-AARP, National Institute of Health + American Association of Retired Persons Diet and Health Study

Nguyen 2018 (30740587)[71]; Keum 2016 (26649988)[58]; Hunter 2020 (32746843)[78]; Park 2019 (30910780)[73]; Eaglehouse 2017 (28542077)[62]

Supplementary Materials

#### Supplementary Figure 68: Linear dose-response meta-analysis of the association between television watching time and colorectal cancer incidence, by adjustment for adiposity and restricted to studies providing both adiposity-adjusted and unadjusted models.

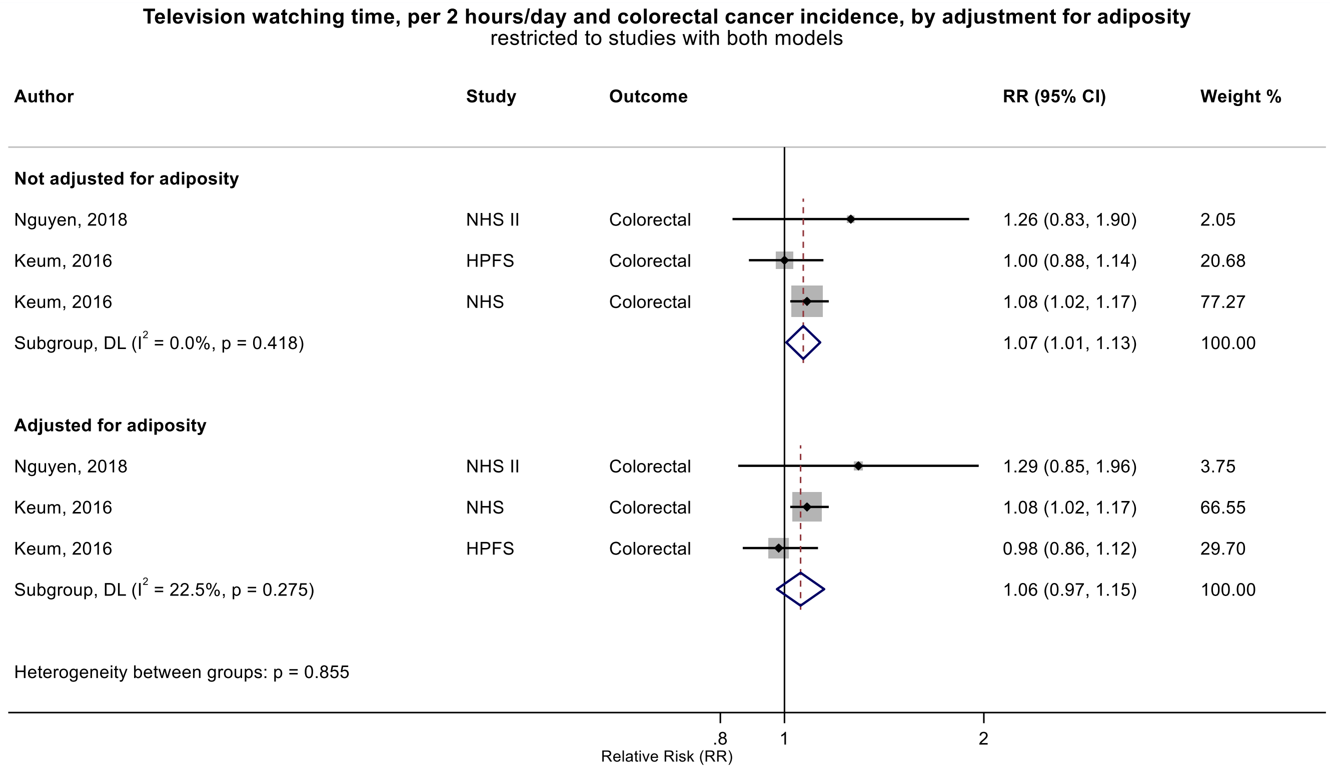

Figure legend: Forest plot shows the linear dose-response results (per 2 hours/day of sedentary time) from the inverse variance DerSimonian-Laird random-effects model. The diamond represents the summary relative risk (RR) estimate and the diamond's width the 95% confidence interval (CI). Each square and the horizontal line across the square represent the RR estimate of the individual study and its 95%CI.

Study abbreviations: NHS, Nurses' Health Study, HPFS, Health Professional Follow-up Study

Nguyen 2018 (30740587)[71]; Keum 2016 (26649988)[58]

### Supplementary Figure 69: Linear dose-response meta-analysis of the association between television watching time and colorectal cancer incidence, by sex.

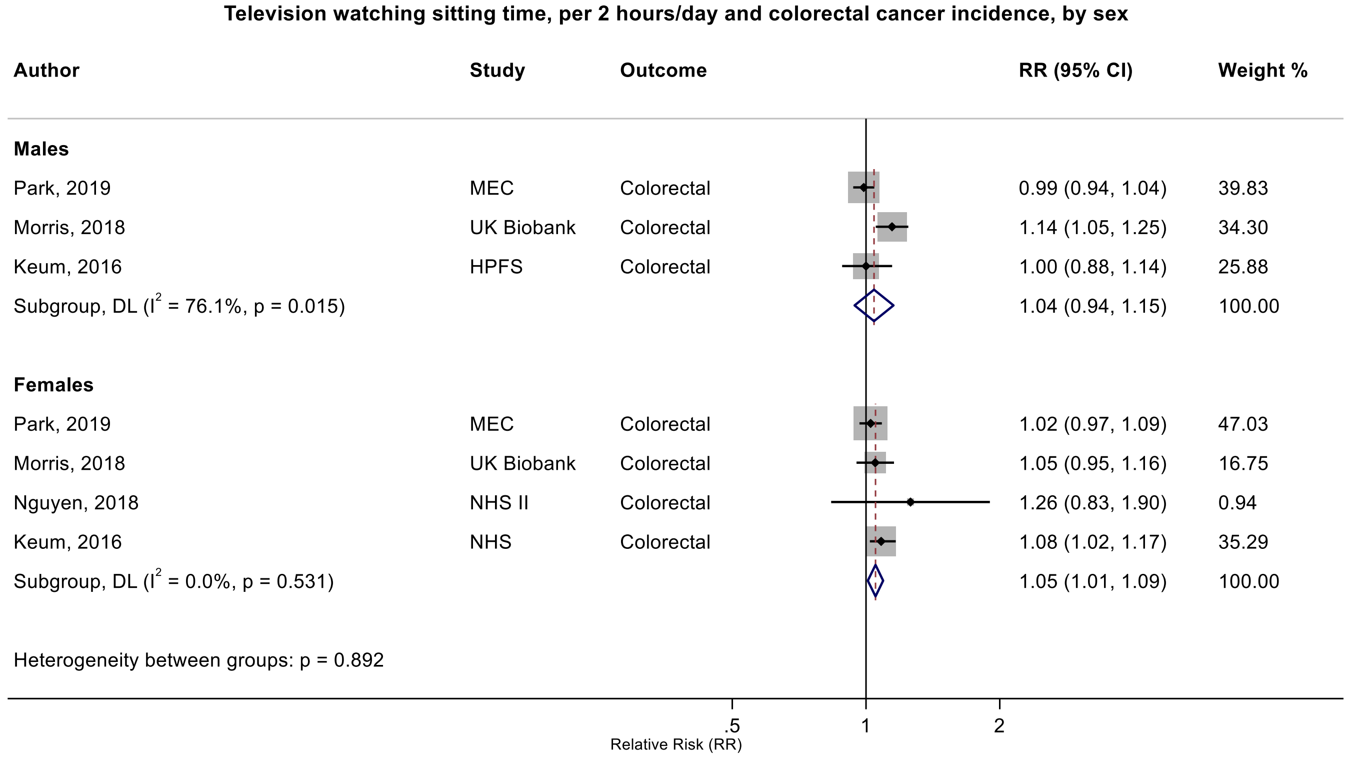

Figure legend: Forest plot shows the linear dose-response results (per 2 hours/day of sedentary time) from the inverse variance DerSimonian-Laird random-effects model. The diamond represents the summary relative risk (RR) estimate and the diamond's width the 95% confidence interval (CI). Each square and the horizontal line across the square represent the RR estimate of the individual study and its 95%CI.

Study abbreviations: MEC, Multiethnic Cohort Study; NHS, Nurses' Health Study, HPFS, Health Professional Follow-up Study

Park 2019 (30910780)[73]; Morris 2018 (29520109)[67]; Keum 2016 (26649988)[58]; Nguyen 2018 (30740587)[71]

**Supplementary Figure 70: Linear dose-response meta-analysis of the association between television watching time and colorectal cancer incidence, by risk of bias due to confounding (Domain 1).**

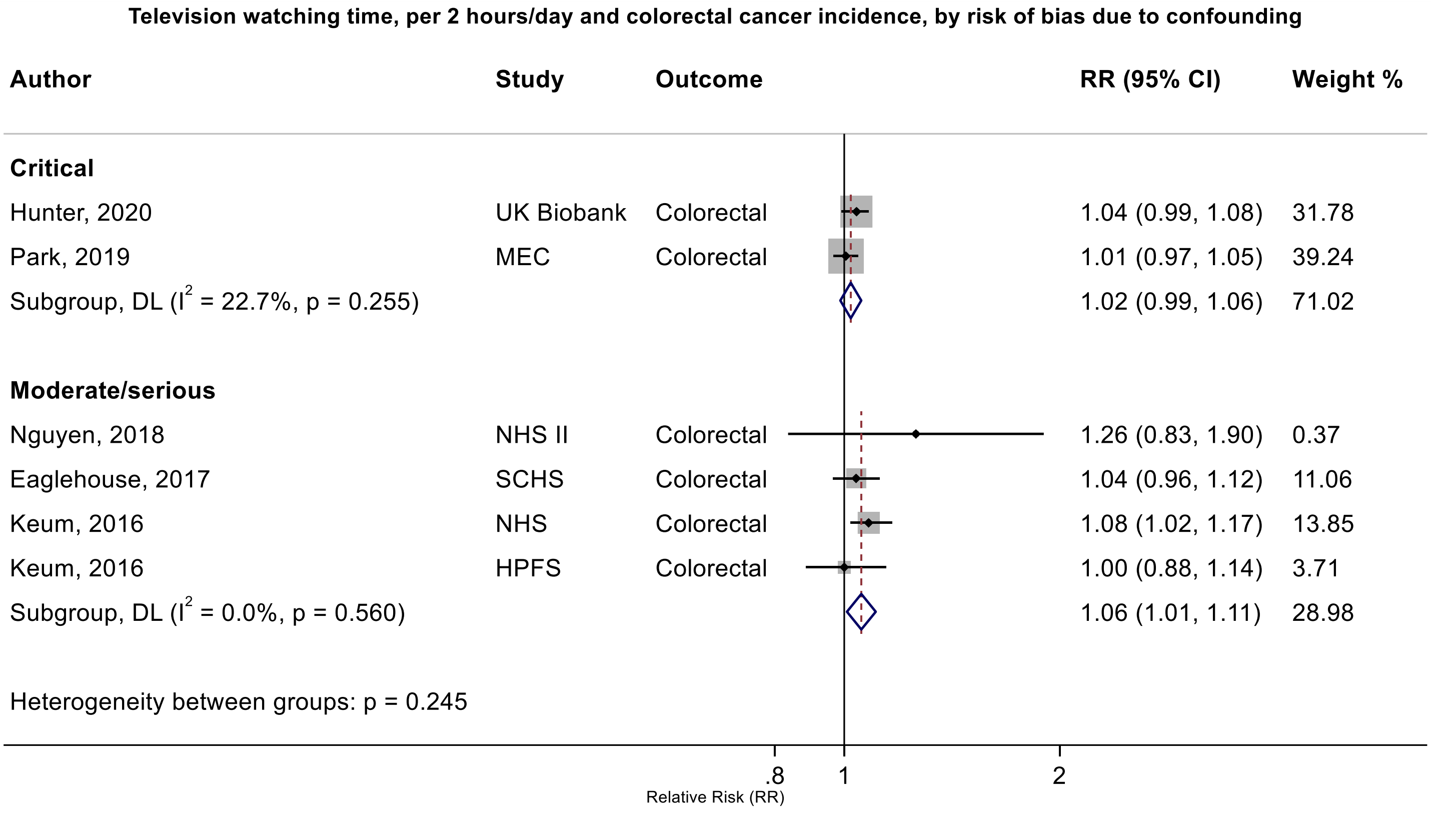

Figure legend: Forest plot shows the linear dose-response results (per 2 hours/day of sedentary time) from the inverse variance DerSimonian-Laird random-effects model. The diamond represents the summary relative risk (RR) estimate and the diamond's width the 95% confidence interval (CI). Each square and the horizontal line across the square represent the RR estimate of the individual study and its 95%CI.

Study abbreviations: MEC, Multiethnic Cohort Study; SCHS, Singapore Chinese Health study; NHS, Nurses' Health Study, HPFS, Health Professional Follow-up Study; NIH-AARP, National Institute of Health + American Association of Retired Persons Diet and Health Study

Hunter 2020 (32746843)[78]; Park 2019 (30910780)[73]; Eaglehouse 2017 (28542077)[62]; Nguyen 2018 (30740587)[71]; Keum 2016 (26649988)[58]

Supplementary Materials

#### Supplementary Figure 71: Linear dose-response meta-analysis of the association between television watching time and colon cancer incidence, by sex.

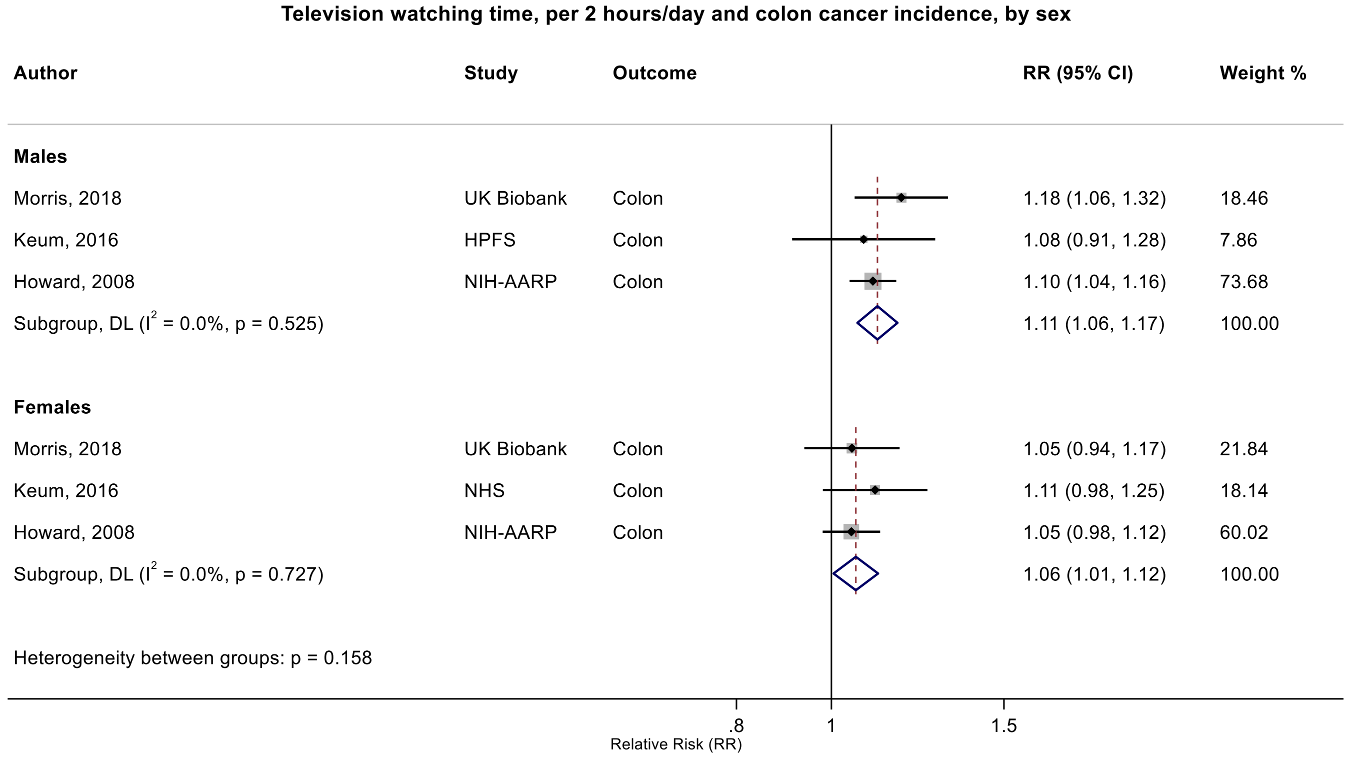

Figure legend: Forest plot shows the linear dose-response results (per 2 hours/day of sedentary time) from the inverse variance DerSimonian-Laird random-effects model. The diamond represents the summary relative risk (RR) estimate and the diamond's width the 95% confidence interval (CI). Each square and the horizontal line across the square represent the RR estimate of the individual study and its 95%CI.

Study abbreviations: NHS, Nurses' Health Study, HPFS, Health Professional Follow-up Study; NIH-AARP, National Institute of Health + American Association of Retired Persons Diet and Health Study

Morris 2018 (29520109)[67]; Keum 2016 (26649988)[58]; Howard 2008 (18437512)[36]

**Supplementary Figure 72: Linear dose-response meta-analysis of the association between television watching time and rectal cancer incidence, by sex.**

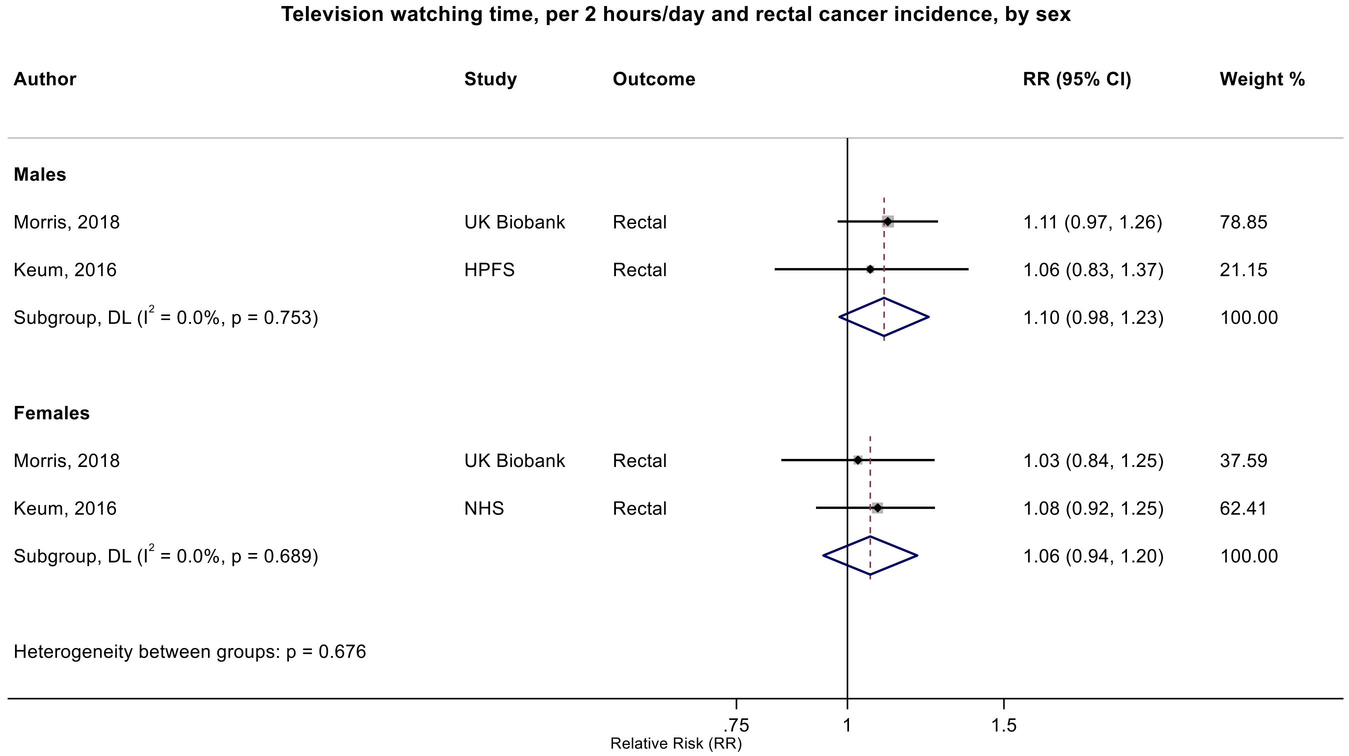

Figure legend: Forest plot shows the linear dose-response results (per 2 hours/day of sedentary time) from the inverse variance DerSimonian-Laird random-effects model. The diamond represents the summary relative risk (RR) estimate and the diamond's width the 95% confidence interval (CI). Each square and the horizontal line across the square represent the RR estimate of the individual study and its 95%CI.

Study abbreviations: NHS, Nurses' Health Study, HPFS, Health Professional Follow-up Study

Morris 2018 (29520109)[67]; Keum 2016 (26649988)[58]

**Supplementary Figure 73: Linear dose-response meta-analysis of the association between television watching time and breast cancer incidence, by physical activity adjustment.**

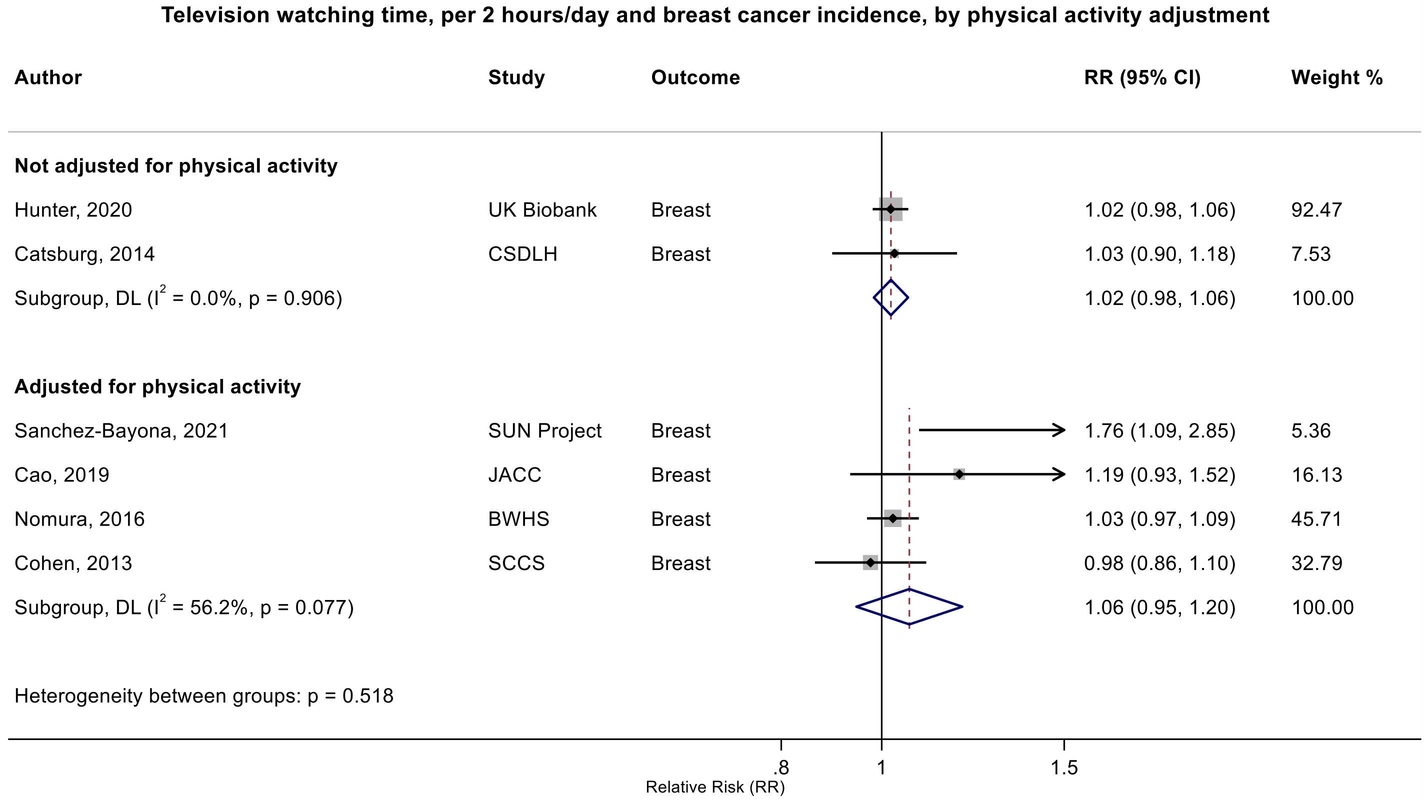

Figure legend: Forest plot shows the linear dose-response results (per 2 hours/day of sedentary time) from the inverse variance DerSimonian-Laird random-effects model. The diamond represents the summary relative risk (RR) estimate and the diamond's width the 95% confidence interval (CI). Each square and the horizontal line across the square represent the RR estimate of the individual study and its 95%CI.

Study abbreviations: CSDLH, Canadian Study of Diet, Lifestyle and Health; SUN, Seguimiento Universidad de Navarra Cohort Study; JACC, Japan Collaborative Cohort study; BWHS, Black Women Health Study; SCCS, Southern Community Cohort Study

Hunter 2020 (32746843)[78]; Catsburg 2014 (24781974)[55]; Sanchez-Bayona 2021 (33798533)[82]; Cao 2019 (30913861)[74]; Nomura 2016 (27632430)[60]; Cohen 2013 (23576427)[46]

#### Supplementary Figure 74: Linear dose-response meta-analysis of the association between television watching time and breast cancer incidence, by menopause status.

**Television watching time, per 2 hours/day and breast cancer incidence, by menopoause status**

| Author | Study | Outcome | RR (95% CI) | Weight % |
|---|---|---|---|---|
| **Premenopausal** | | | | |
| Sanchez-Bayona, 2021 | SUN Project | Breast | 2.40 (1.26, 4.56) | 2.60 |
| Hunter, 2020 | UK Biobank | Breast | 1.02 (0.94, 1.12) | 35.95 |
| Cao, 2019 | JACC | Breast | 1.03 (0.78, 1.36) | 11.10 |
| Nomura, 2016 | BWHS | Breast | 0.97 (0.88, 1.06) | 34.88 |
| Catsburg, 2014 | CSDLH | Breast | 0.95 (0.76, 1.19) | 15.48 |
| Subgroup, DL ($I^2$ = 49.9%, p = 0.092) | | | 1.01 (0.91, 1.13) | 100.00 |
| **Postmenopausal** | | | | |
| Sanchez-Bayona, 2021 | SUN Project | Breast | 1.07 (0.45, 2.57) | 0.10 |
| Hunter, 2020 | UK Biobank | Breast | 1.01 (0.96, 1.06) | 29.66 |
| Cao, 2019 | JACC | Breast | 1.65 (1.02, 2.65) | 0.32 |
| Nomura, 2016 | BWHS | Breast | 1.03 (0.95, 1.11) | 11.27 |
| Catsburg, 2014 | CSDLH | Breast | 1.10 (0.92, 1.31) | 2.29 |
| George, 2010 | NIH-AARP | Breast | 1.01 (0.98, 1.05) | 56.37 |
| Subgroup, DL ($I^2$ = 0.0%, p = 0.425) | | | 1.02 (0.99, 1.04) | 100.00 |
| Heterogeneity between groups: p = 0.983 | | | | |

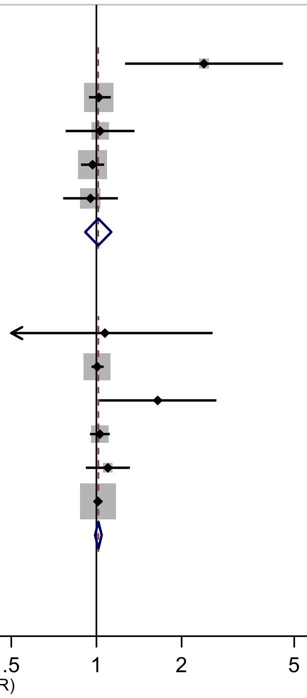

Figure legend: Forest plot shows the linear dose-response results (per 2 hours/day of sedentary time) from the inverse variance DerSimonian-Laird random-effects model. The diamond represents the summary relative risk (RR) estimate and the diamond's width the 95% confidence interval (CI). Each square and the horizontal line across the square represent the RR estimate of the individual study and its 95%CI.

Study abbreviations: CSDLH, Canadian Study of Diet, Lifestyle and Health; SUN, Seguimiento Universidad de Navarra Cohort Study; JACC, Japan Collaborative Cohort study; BWHS, Black Women Health Study; NIH-AARP, National Institute of Health + American Association of Retired Persons Diet and Health Study

Sanchez-Bayona 2021 (33798533)[82]; Hunter 2020 (32746843)[78]; Cao 2019 (30913861)[74]; Nomura 2016 (27632430)[60]; Catsburg 2014 (24781974)[55] ; George 2010 (20864719)[39]

#### Supplementary Figure 75: Linear dose-response meta-analysis of the association between television watching time and breast cancer incidence, by region.

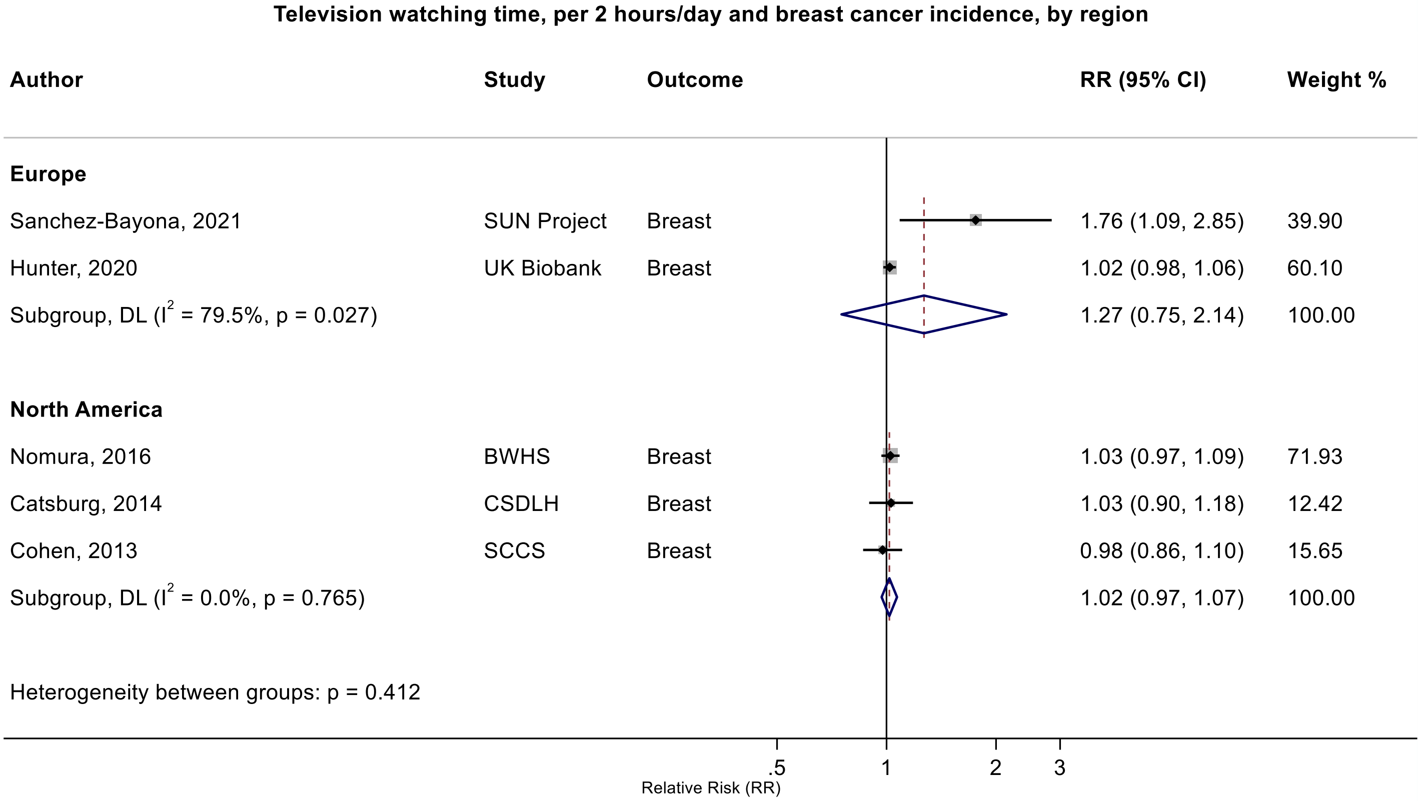

Figure legend: Forest plot shows the linear dose-response results (per 2 hours/day of sedentary time) from the inverse variance DerSimonian-Laird random-effects model. The diamond represents the summary relative risk (RR) estimate and the diamond's width the 95% confidence interval (CI). Each square and the horizontal line across the square represent the RR estimate of the individual study and its 95%CI.

Study abbreviations: CSDLH, Canadian Study of Diet, Lifestyle and Health; SUN, Seguimiento Universidad de Navarra Cohort Study; BWHS, Black Women Health Study; SCCS, Southern Community Cohort Study; NIH-AARP, National Institute of Health + American Association of Retired Persons Diet and Health Study

Sanchez-Bayona 2021 (33798533)[82]; Hunter 2020 (32746843)[78]; Nomura 2016 (27632430)[60]; Catsburg 2014 (24781974)[55]; Cohen 2013 (23576427)[46]

Supplementary Materials

**Supplementary Figure 76: Linear dose-response meta-analysis of the association between television watching time and breast cancer incidence, by risk of bias due to confounding (Domain 1).**

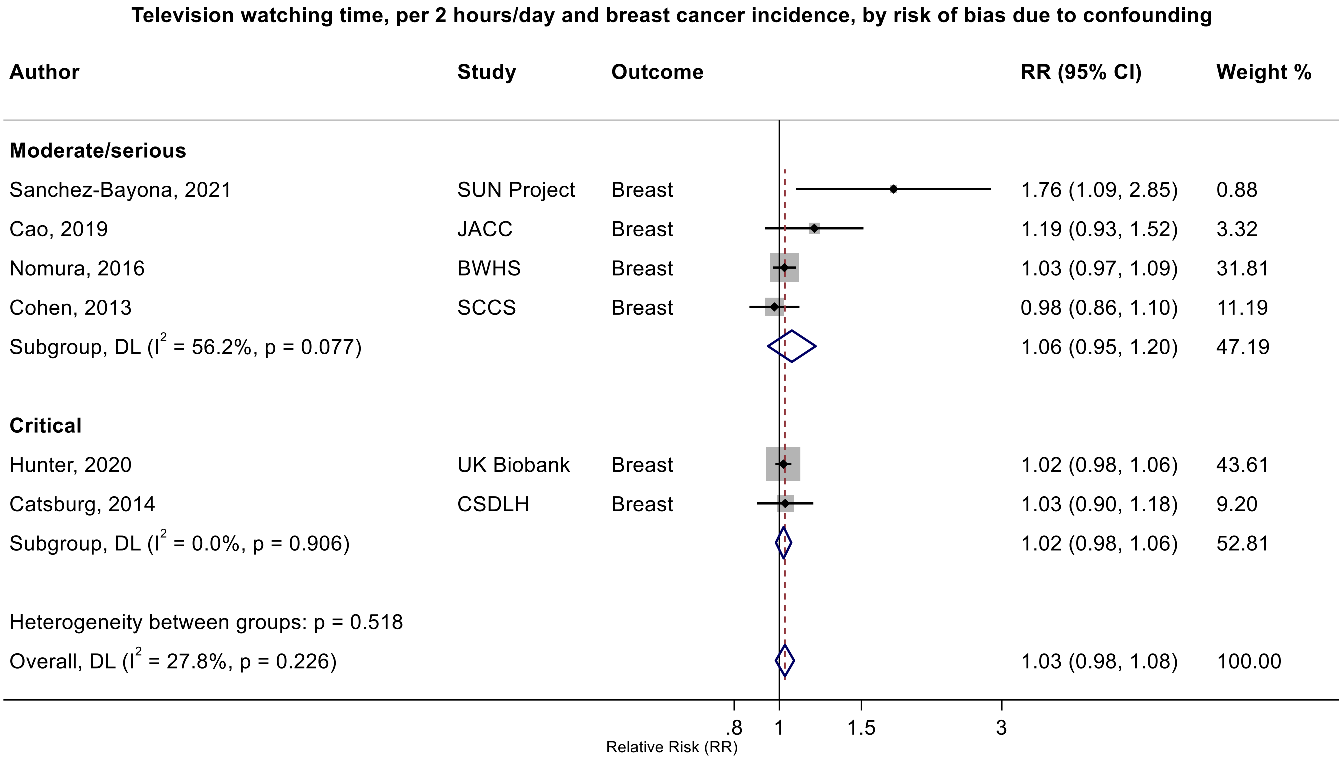

Figure legend: Forest plot shows the linear dose-response results (per 2 hours/day of sedentary time) from the inverse variance DerSimonian-Laird random-effects model. The diamond represents the summary relative risk (RR) estimate and the diamond's width the 95% confidence interval (CI). Each square and the horizontal line across the square represent the RR estimate of the individual study and its 95%CI.

Study abbreviations: CSDLH, Canadian Study of Diet, Lifestyle and Health; SUN, Seguimiento Universidad de Navarra Cohort Study; JACC, Japan Collaborative Cohort study; BWHS, Black Women Health Study; SCCS, Southern Community Cohort Study

Hunter 2020 (32746843)[78]; Catsburg 2014 (24781974)[55]; Sanchez-Bayona 2021 (33798533)[82]; Cao 2019 (30913861)[74]; Nomura 2016 (27632430)[60]; Cohen 2013 (23576427)[46]

#### Supplementary Figure 77: Linear dose-response meta-analysis of the association between television watching time and endometrial cancer incidence, by adjustment for adiposity.

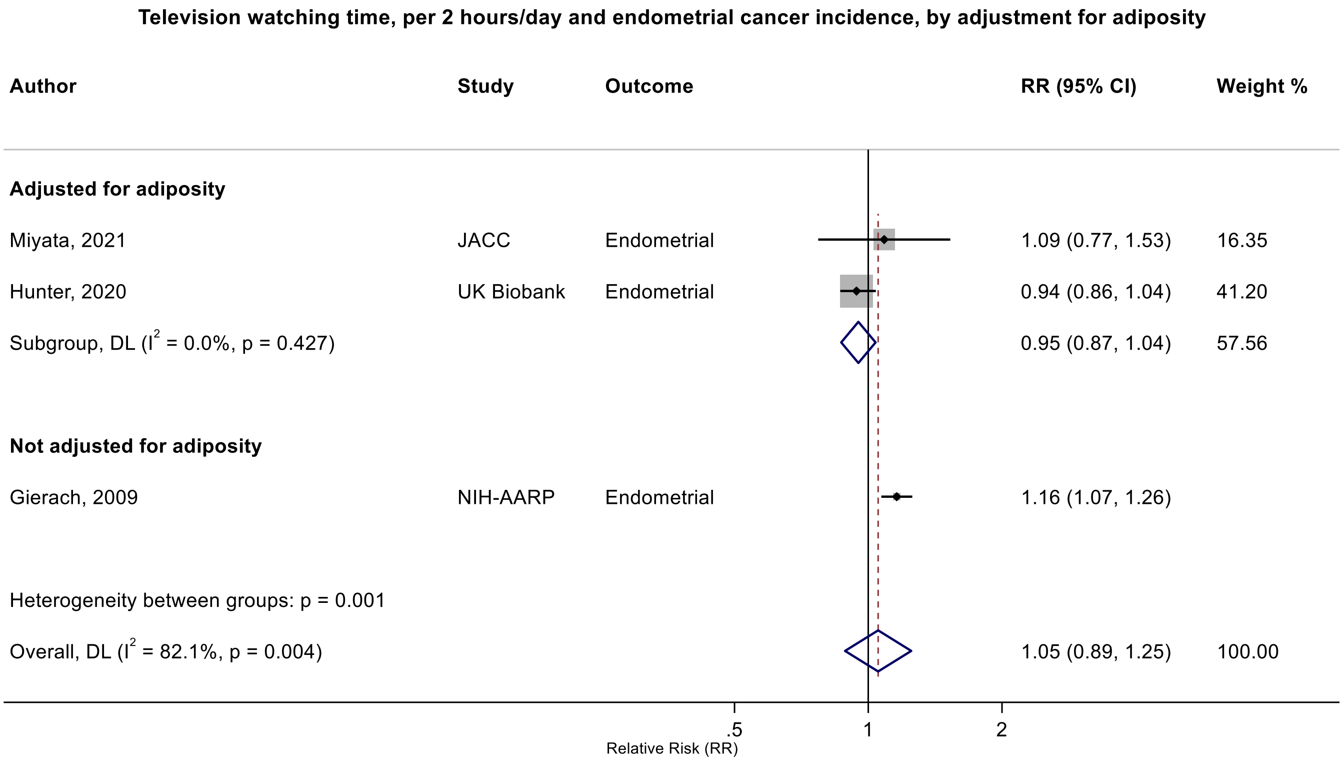

Figure legend: Forest plot shows the linear dose-response results (per 2 hours/day of sedentary time) from the inverse variance DerSimonian-Laird random-effects model. The diamond represents the summary relative risk (RR) estimate and the diamond's width the 95% confidence interval (CI). Each square and the horizontal line across the square represent the RR estimate of the individual study and its 95%CI.

Study abbreviations: JACC, Japan Collaborative Cohort study; NIH-AARP, National Institute of Health + American Association of Retired Persons Diet and Health Study

Miyata 2021 (32963209)[79]; Hunter 2020 (32746843)[78]; Gierach 2009 (19123463)[38]

**Supplementary Figure 78: Linear dose-response meta-analysis of the association between television watching time and ovarian cancer incidence, by physical activity adjustment.**

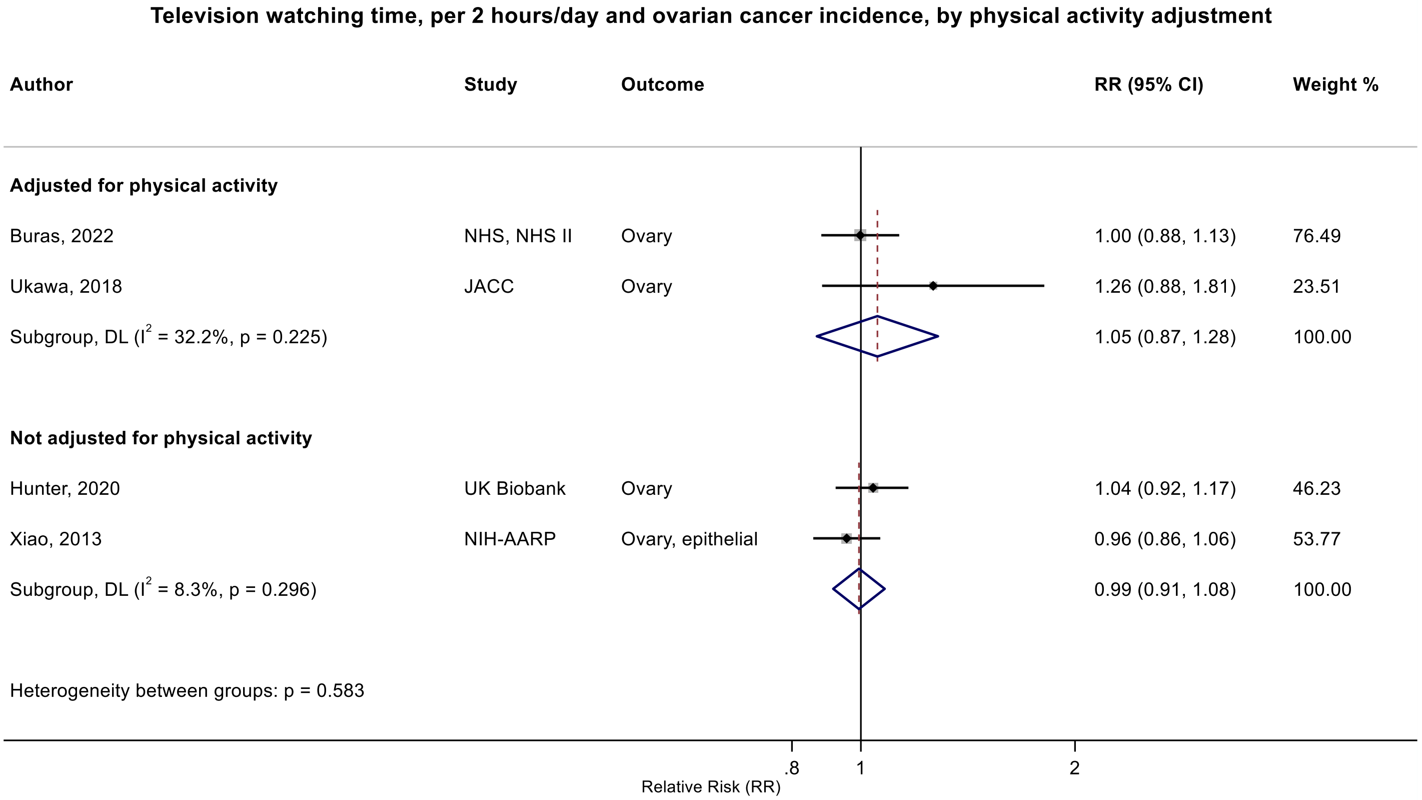

Figure legend: Forest plot shows the linear dose-response results (per 2 hours/day of sedentary time) from the inverse variance DerSimonian-Laird random-effects model. The diamond represents the summary relative risk (RR) estimate and the diamond's width the 95% confidence interval (CI). Each square and the horizontal line across the square represent the RR estimate of the individual study and its 95%CI.

Study abbreviations: NHS, Nurses' Health Study; JACC, Japan Collaborative Cohort study; NIH-AARP, National Institute of Health + American Association of Retired Persons Diet and Health Study

Buras 2022 (35094053)[84]; Ukawa 2018 (29340890)[64]; Hunter 2020 (32746843)[78]; Xiao 2013 (23966580)[49]

Supplementary Materials

#### Supplementary Figure 79: Linear dose-response meta-analysis of the association between television watching time and prostate cancer incidence by body mass index (BMI) category.

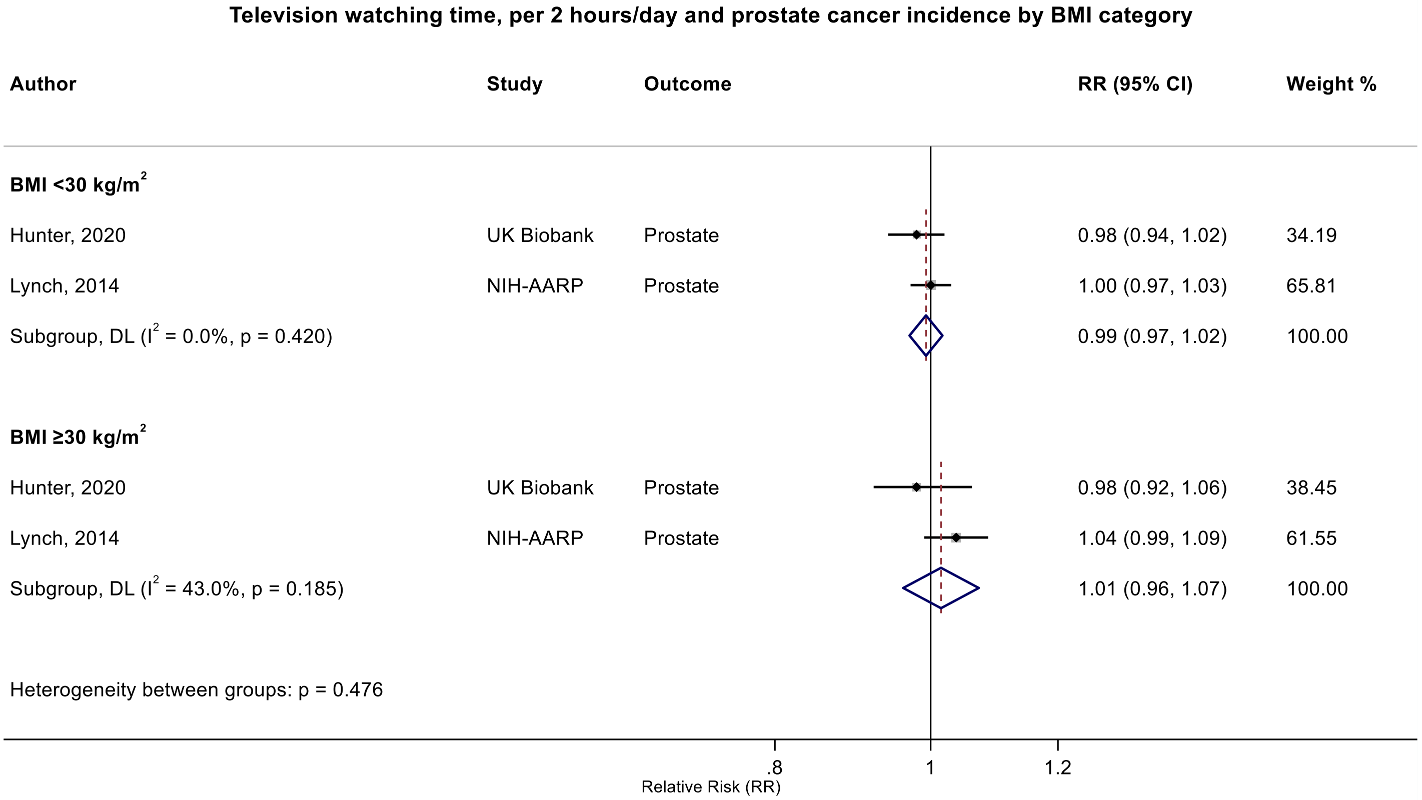

Figure legend: Forest plot shows the linear dose-response results (per 2 hours/day of sedentary time) from the inverse variance DerSimonian-Laird random-effects model. The diamond represents the summary relative risk (RR) estimate and the diamond's width the 95% confidence interval (CI). Each square and the horizontal line across the square represent the RR estimate of the individual study and its 95%CI.

Study abbreviations: NIH-AARP, National Institute of Health + American Association of Retired Persons Diet and Health Study

Hunter 2020 (32746843)[78]; Lynch 2014 (24526287)[53]

### Supplementary Figure 80: Linear dose-response meta-analysis of the association between television watching time and colorectal cancer incidence, sensitivity analysis, using the adiposity-adjusted estimates from studies reporting both adiposity-adjusted and unadjusted models.

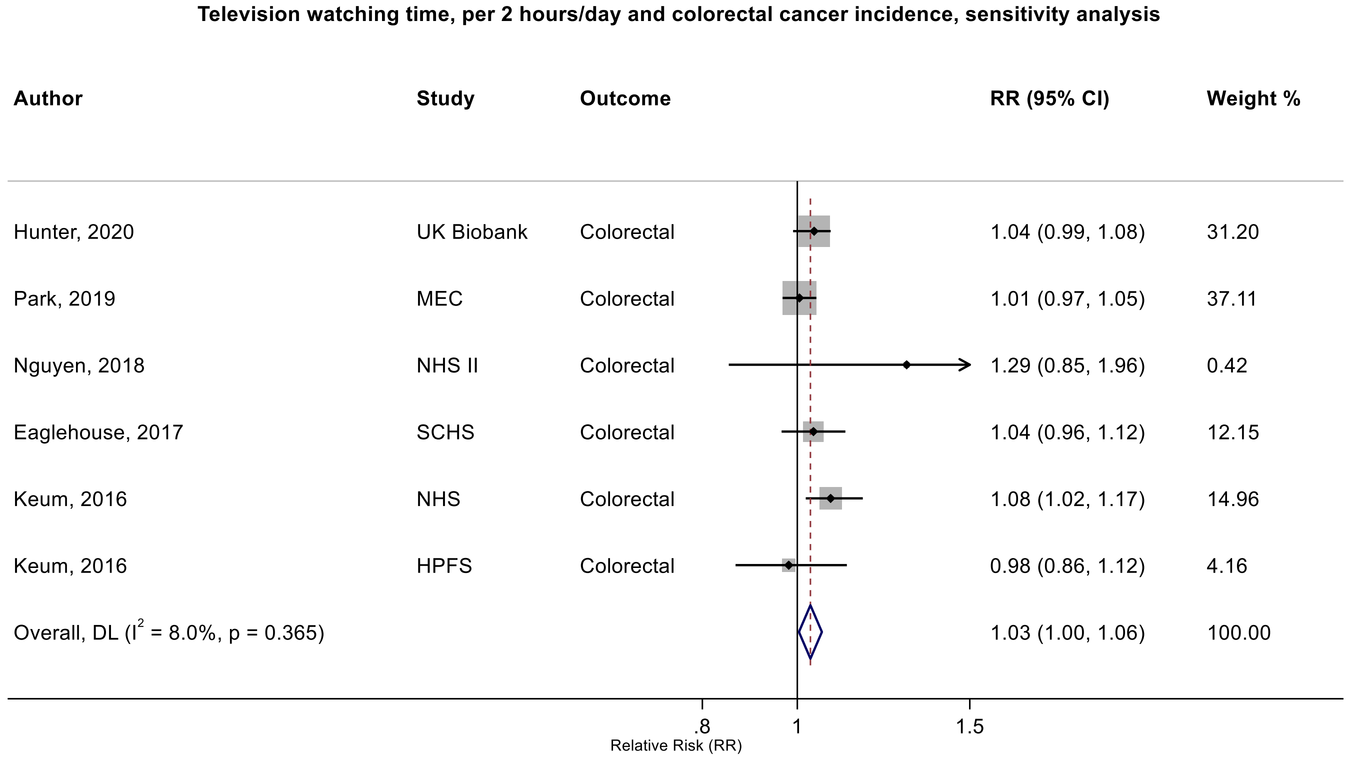

Figure legend: Forest plot shows the linear dose-response results (per 2 hours/day of sedentary time) from the inverse variance DerSimonian-Laird random-effects model. The diamond represents the summary relative risk (RR) estimate and the diamond's width the 95% confidence interval (CI). Each square and the horizontal line across the square represent the RR estimate of the individual study and its 95%CI.

Study abbreviations: MEC, Multiethnic Cohort Study; NHS, Nurses' Health Study, HPFS, Health Professional Follow-up Study; SCHS, Singapore Chinese Health study

Hunter 2020 (32746843)[78]; Park 2019 (30910780)[73]; Nguyen 2018 (30740587)[71]; Eaglehouse 2017 (28542077)[62]; Keum 2016 (26649988)[58]

**Supplementary Figure 81: Linear dose-response meta-analysis of the association between sedentary time and endometrial cancer incidence, sensitivity analysis, using the adiposity-adjusted estimates from studies reporting both adiposity-adjusted and unadjusted models.**

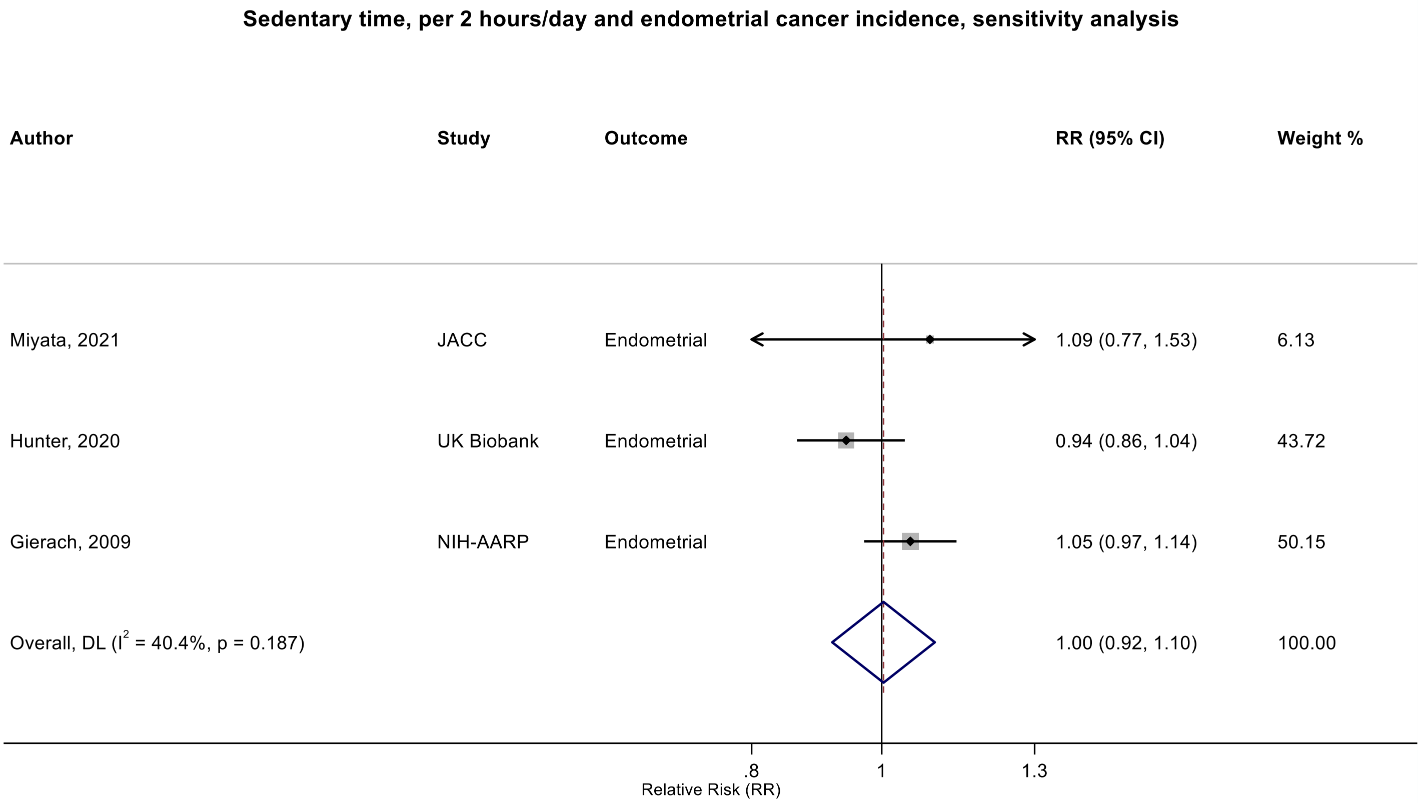

Figure legend: Forest plot shows the linear dose-response results (per 2 hours/day of sedentary time) from the inverse variance DerSimonian-Laird random-effects model. The diamond represents the summary relative risk (RR) estimate and the diamond's width the 95% confidence interval (CI). Each square and the horizontal line across the square represent the RR estimate of the individual study and its 95%CI.

Study abbreviations: JACC, Japan Collaborative Cohort study; NIH-AARP, National Institute of Health + American Association of Retired Persons Diet and Health Study

Miyata 2021 (32963209)[79]; Hunter 2020 (32746843)[78]; Gierach 2009 (19123463)[38]

#### Supplementary Figure 82: Categorical highest versus lowest meta-analysis of the association between television watching time and endometrial cancer.

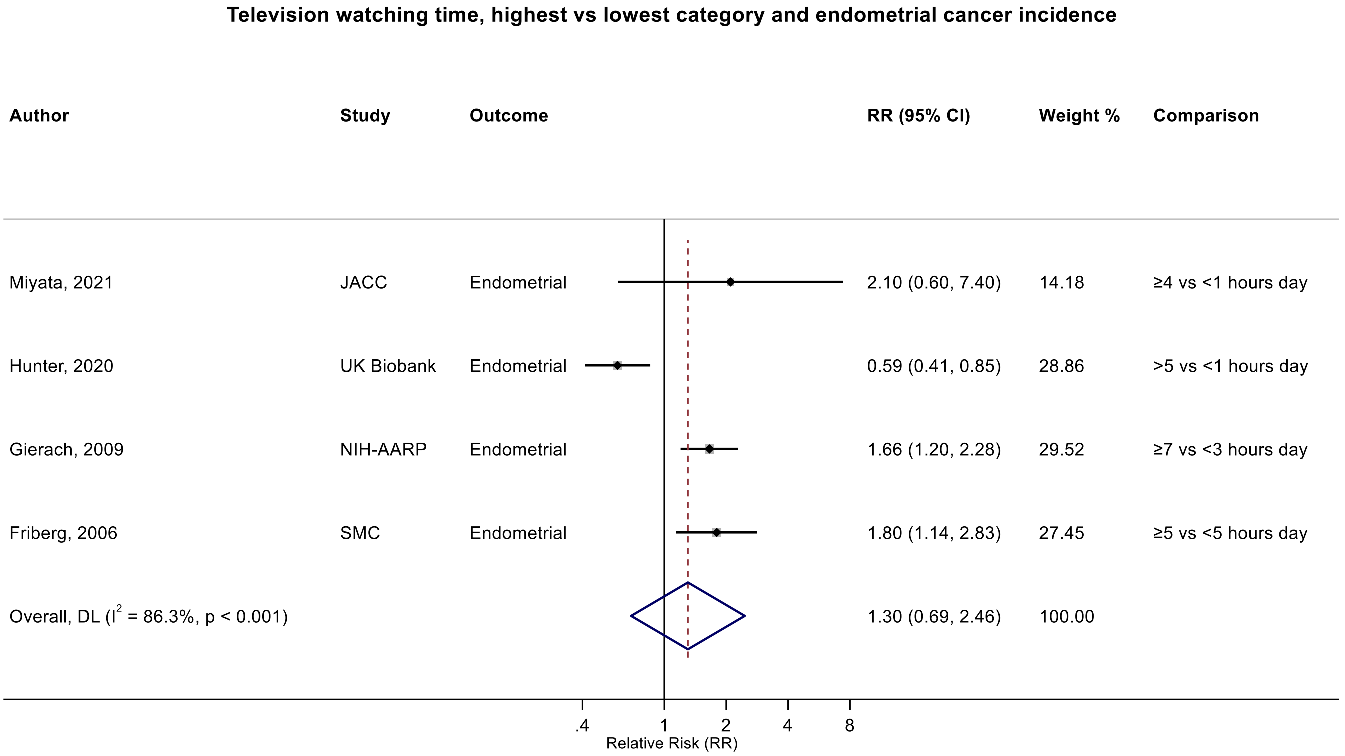

Figure legend: Forest plot shows the categorical comparison results from the individual studies for the associations with the largest contrast, as reported in the respective publication. Each square and the horizontal line across the square represent the relative risk (RR) estimate of the individual study and its 95% confidence interval (CI).

Study abbreviations: JACC, Japan Collaborative Cohort study; NIH-AARP, National Institute of Health + American Association of Retired Persons Diet and Health Study; SMC, Swedish Mammography Cohort

Miyata 2021 (32963209)[79]; Hunter 2020 (32746843)[78]; Gierach 2009 (19123463)[38]; Friberg 2006 (17057024)[35]

#### Supplementary Figure 83: Categorical highest versus lowest meta-analysis of the association between television watching time and lung cancer incidence.

**Television watching time, highest vs lowest category and lung cancer incidence**

| Author | Study | Outcome | RR (95% CI) | Weight % | Comparison |
|---|---|---|---|---|---|
| Bethea, 2022 | BWHS | Lung | 1.27 (0.72, 2.21) | 2.64 | ≥1 vs <1 hours week |
| Hunter, 2020 | UK Biobank | Lung | 1.28 (1.15, 1.43) | 73.67 | >5 vs <1 hours day |
| Lam, 2013 | NIH-AARP | Lung | 1.06 (0.77, 1.46) | 8.12 | ≥5 vs <3 hours day |
| Ukawa, 2013 | JACC | Lung | 1.25 (0.99, 1.58) | 15.57 | ≥4 vs <2 hours day |
| Overall, DL ($I^2$ = 0.0%, p = 0.746) | | | 1.26 (1.15, 1.38) | 100.00 | |

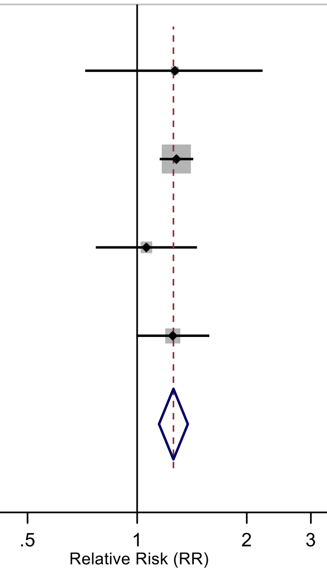

Figure legend: Forest plot shows the categorical comparison results from the individual studies for the associations with the largest contrast, as reported in the respective publication. Each square and the horizontal line across the square represent the relative risk (RR) estimate of the individual study and its 95% confidence interval (CI).

Study abbreviations: BWHS, Black Women Health Study; NIH-AARP, National Institute of Health + American Association of Retired Persons Diet and Health Study; JACC, Japan Collaborative Cohort study

Bethea 2022 (35325667)[87]; Hunter 2020 (32746843)[78]; Lam 2013 (23940620)[48]; Ukawa 2013 (23686441)[47]

#### Supplementary Figure 84: Categorical highest versus lowest meta-analysis of the association between television watching time and endometrial cancer, by adjustment for adiposity.

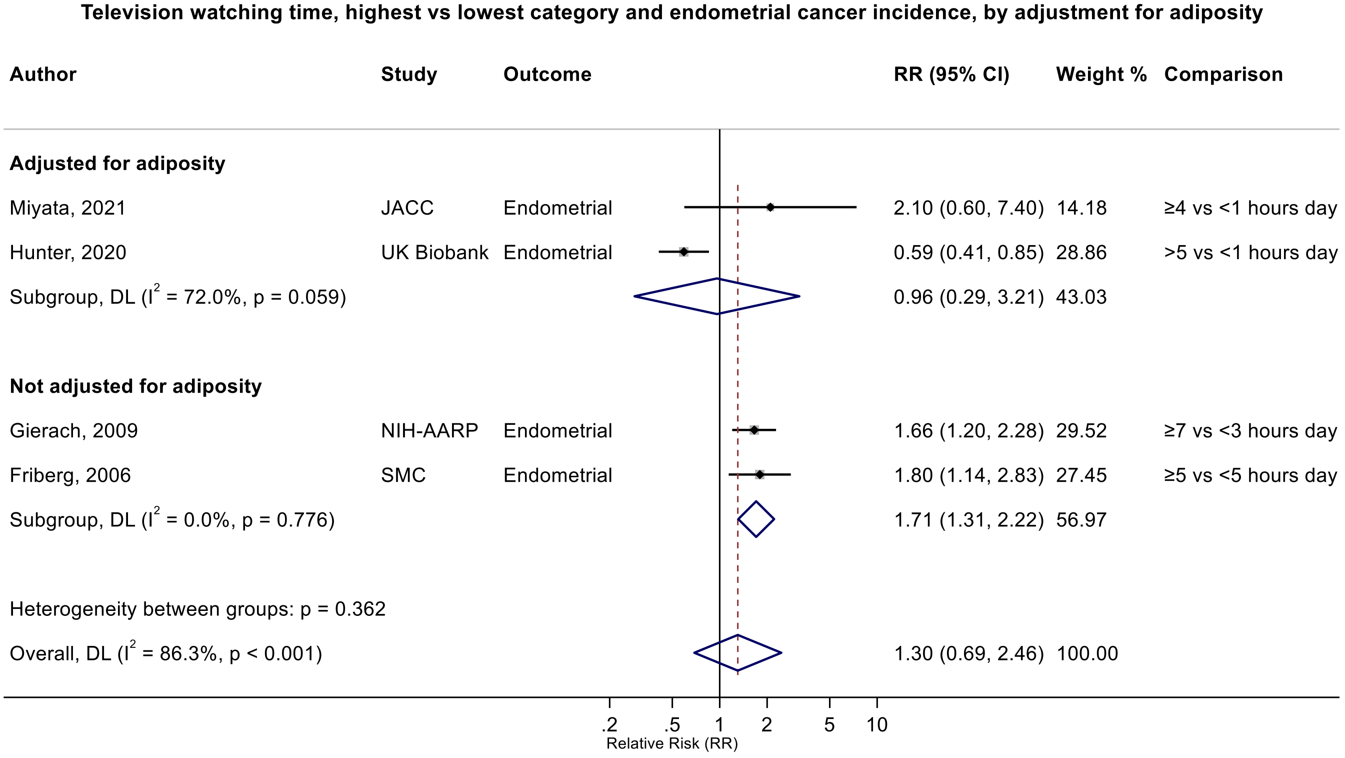

Figure legend: Forest plot shows the categorical comparison results from the individual studies for the associations with the largest contrast, as reported in the respective publication. Each square and the horizontal line across the square represent the relative risk (RR) estimate of the individual study and its 95% confidence interval (CI).

Study abbreviations: JACC, Japan Collaborative Cohort study; NIH-AARP, National Institute of Health + American Association of Retired Persons Diet and Health Study; SMC, Swedish Mammography Cohort

Miyata 2021 (32963209)[79]; Hunter 2020 (32746843)[78]; Gierach 2009 (19123463)[38]; Friberg 2006 (17057024)[35]

#### Supplementary Figure 85: Categorical highest versus lowest meta-analysis of the association between television watching time and lung cancer incidence, by risk of bias due to confounding (Domain 1).

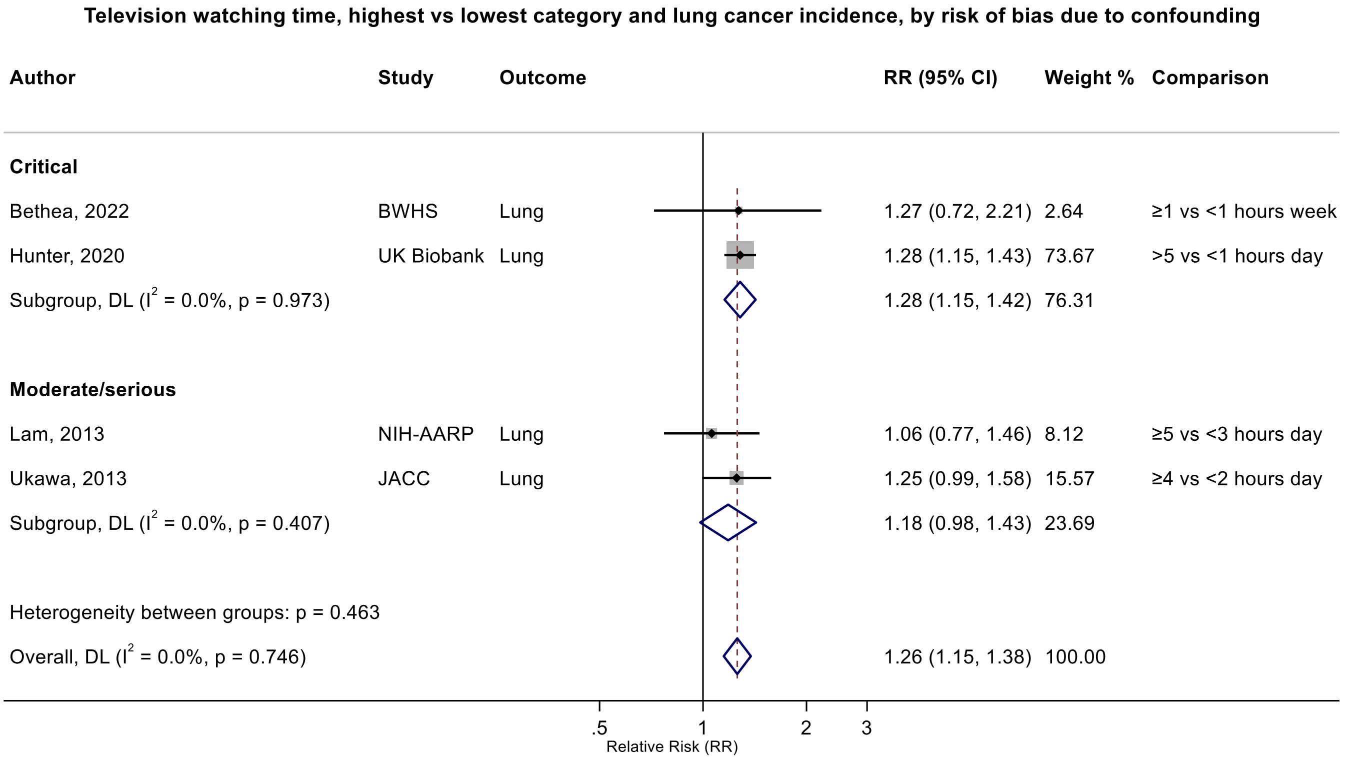

Figure legend: Forest plot shows the categorical comparison results from the individual studies for the associations with the largest contrast, as reported in the respective publication. Each square and the horizontal line across the square represent the relative risk (RR) estimate of the individual study and its 95% confidence interval (CI).

Study abbreviations: BWHS, Black Women Health Study; NIH-AARP, National Institute of Health + American Association of Retired Persons Diet and Health Study; JACC, Japan Collaborative Cohort study

Bethea 2022 (35325667)[87]; Hunter 2020 (32746843)[78]; Lam 2013 (23940620)[48]; Ukawa 2013 (23686441)[47]

**Supplementary Figure 86: Non-linear dose-response meta-analysis of the association between total television watching time and colon cancer incidence-sensitivity analysis, using the adiposity-adjusted estimates from studies reporting both adiposity-adjusted and unadjusted models.**

The solid line represents the estimated summary dose-response relationship and the short-dashed line the 95% confidence intervals. The tick marks inside the x-axis indicate the total television watching time hours/days values for which relative risk estimate(s) were available. Non-linear curve was estimated using restricted cubic spline regressions with three knots placed at fixed percentiles (10%, 50%, and 90%) of the exposure distribution, which were pooled by fitting one-stage random-effects mixed models. 2 sitting hours/day was chosen as reference. The table shows selected television watching time hours/day values and their corresponding relative risk (95% confidence intervals) estimated in the non-linear dose-response meta-analysis.

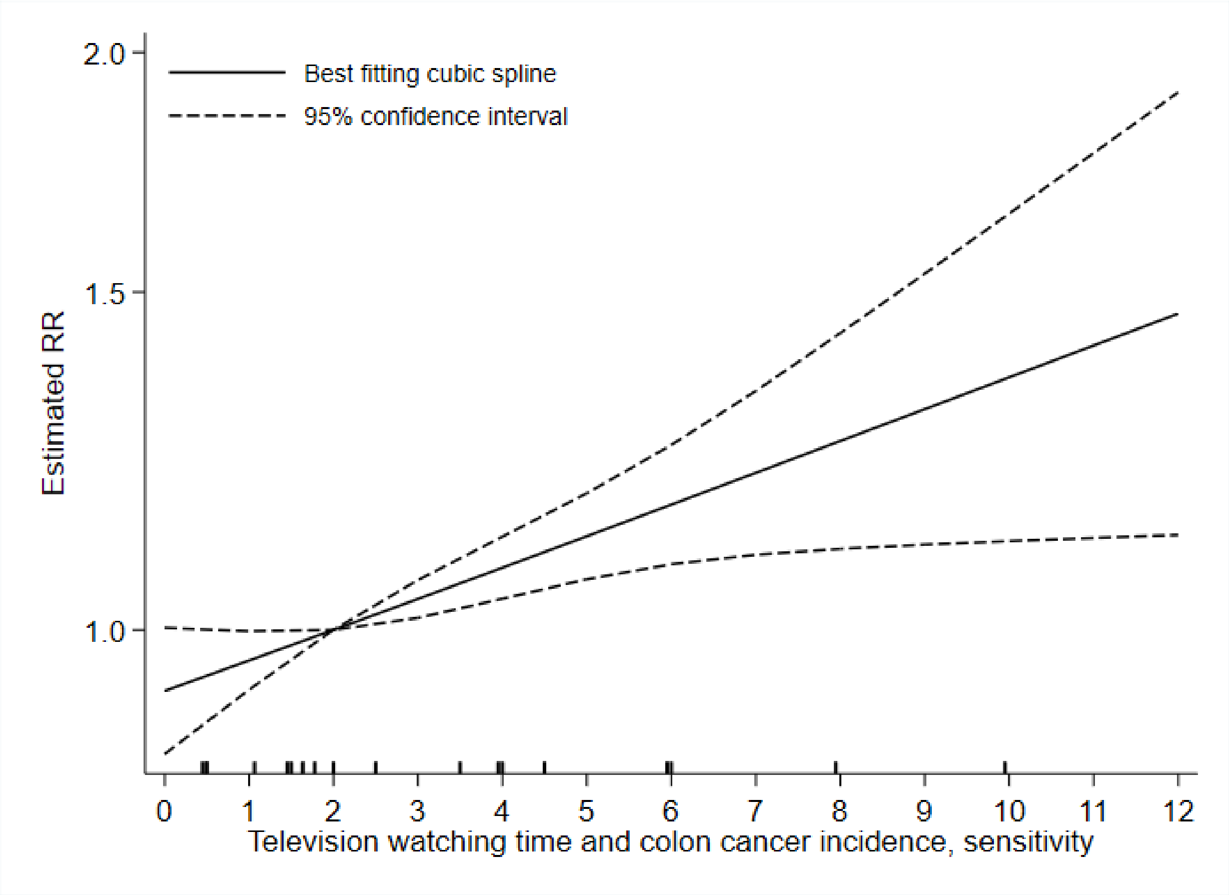

$P_{non\text{-}linearity}$ = 0.963

n = 5 studies (5 publications)

8,420 cases

| Television watching time (hours/day) | RR (95% CI) |
|---|---|
| 0 | 0.93 (0.86, 1.00) |
| 1 | 0.96 (0.93, 1.00) |
| 2 | 1 (Reference) |
| 3 | 1.04 (1.01, 1.06) |
| 4 | 1.08 (1.04, 1.12) |
| 5 | 1.12 (1.06, 1.18) |
| 6 | 1.16 (1.08, 1.25) |
| 7 | 1.21 (1.09, 1.33) |
| 8 | 1.25 (1.10, 1.43) |
| 9 | 1.30 (1.11, 1.53) |
| 10 | 1.35 (1.11, 1.65) |
| 11 | 1.41 (1.12, 1.77) |
| 12 | 1.46 (1.12, 1.91) |

**Supplementary Figure 87: Non-linear a dose-response meta-analysis of the association between total television watching time and colorectal cancer incidence-sensitivity analysis, using the adiposity-adjusted estimates from studies reporting both adiposity-adjusted and unadjusted models.**

The solid line represents the estimated summary dose-response relationship and the short-dashed line the 95% confidence intervals. The tick marks inside the x-axis indicate the total television watching time hours/days values for which relative risk estimate(s) were available. Non-linear curve was estimated using restricted cubic spline regressions with three knots placed at fixed percentiles (10%, 50%, and 90%) of the exposure distribution, which were pooled by fitting one-stage random-effects mixed models. 2 sitting hours/day was chosen as reference. The table shows selected television watching time hours/day values and their corresponding relative risk (95% confidence intervals) estimated in the non-linear dose-response meta-analysis.
Abbreviations: CI: confidence interval; RR, relative risk.

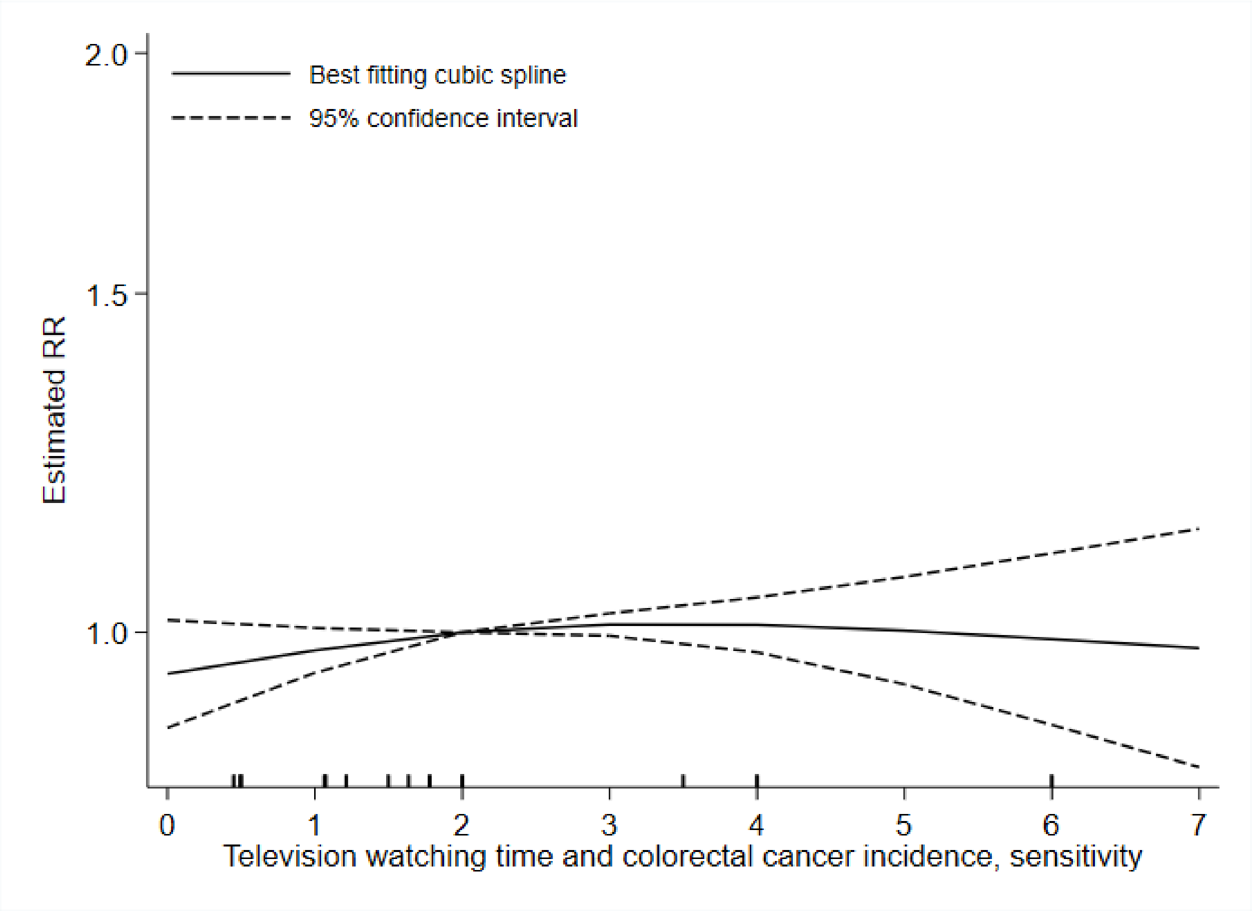

$P_{non\text{-}linearity}$ = 0.303

n = 6 studies (5 publications)

11,619 cases

| Television watching time (hours/day) | RR (95% CI) |
|---|---|
| 0 | 0.95 (0.89, 1.01) |
| 1 | 0.98 (0.95, 1.01) |
| 2 | 1 (Reference) |
| 3 | 1.01 (1.00, 1.02) |
| 4 | 1.01 (0.98, 1.04) |
| 5 | 1.00 (0.94, 1.07) |
| 6 | 0.99 (0.90, 1.10) |
| 7 | 0.98 (0.85, 1.13) |

#### Supplementary Figure 88: Linear dose-response meta-analysis of the association between television watching time and lung cancer incidence, leave-one-out sensitivity analysis.

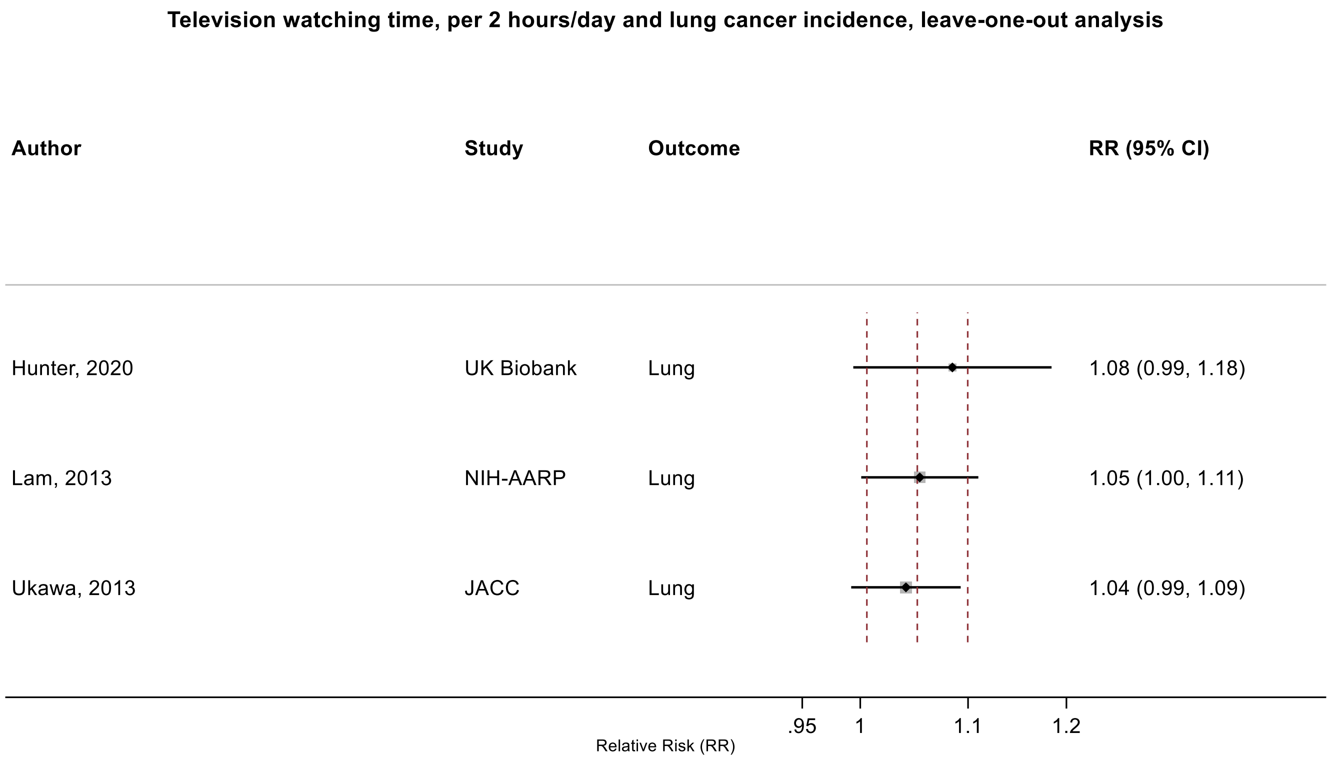

Figure legend: Forest plot shows the linear dose-response results (per 2 hours of sedentary time) from the inverse variance DerSimonian-Laird random-effects model after exclusion of each study from the meta-analysis. For each study, the square represents the summary relative risk (RR) estimate and the horizontal line across the square the corresponding 95% confidence interval (CI) computed from a meta-analysis excluding that study. The dashed vertical lines represent the results of the main meta-analysis.

Study abbreviations: NIH-AARP, National Institute of Health + American Association of Retired Persons Diet and Health Study; JACC, Japan Collaborative Cohort study

Hunter 2020 (32746843)[78]; Lam 2013 (23940620)[48]; Ukawa 2013 (23686441)[47]

**Supplementary Figure 89: Linear dose-response meta-analysis of the association between television watching time and colorectal cancer incidence, leave-one-out sensitivity analysis.**

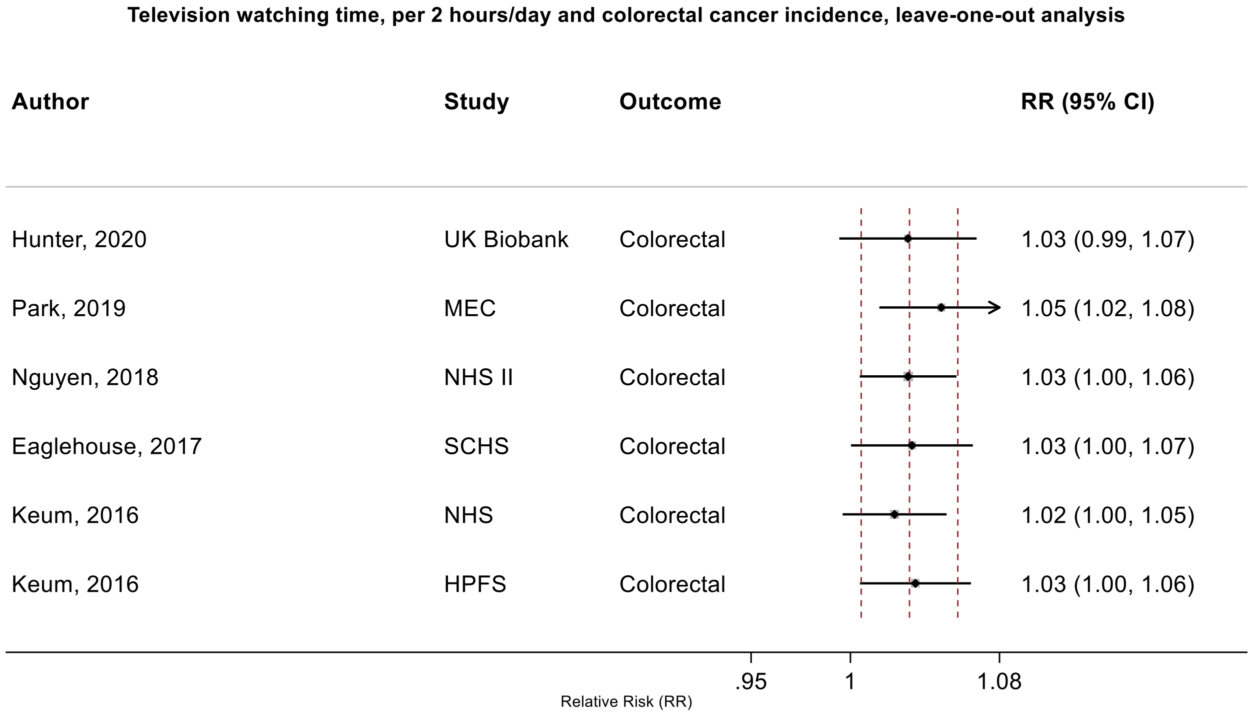

Figure legend: Forest plot shows the linear dose-response results (per 2 hours of sedentary time) from the inverse variance DerSimonian-Laird random-effects model after exclusion of each study from the meta-analysis. For each study, the square represents the summary relative risk (RR) estimate and the horizontal line across the square the corresponding 95% confidence interval (CI) computed from a meta-analysis excluding that study. The dashed vertical lines represent the results of the main meta-analysis.

Study abbreviations: MEC, Multiethnic Cohort Study; NHS, Nurses' Health Study, HPFS, Health Professional Follow-up Study; SCHS, Singapore Chinese Health study

Hunter 2020 (32746843)[78]; Park 2019 (30910780)[73]; Nguyen 2018 (30740587)[71]; Eaglehouse 2017 (28542077)[62]; Keum 2016 (26649988)[58]

#### Supplementary Figure 90: Linear dose-response meta-analysis of the association between television watching time and breast cancer incidence, leave-one-out sensitivity analysis.

**Television watching time, per 2 hours/day and breast cancer incidence, leave-one-out analysis**

| Author | Study | Outcome | RR (95% CI) |
|---|---|---|---|
| Sanchez-Bayona, 2021 | SUN Project | Breast | 1.02 (0.99, 1.05) |
| Hunter, 2020 | UK Biobank | Breast | 1.04 (0.96, 1.13) |
| Cao, 2019 | JACC | Breast | 1.02 (0.98, 1.07) |
| Nomura, 2016 | BWHS | Breast | 1.04 (0.96, 1.12) |
| Catsburg, 2014 | CSDLH | Breast | 1.03 (0.97, 1.09) |
| Cohen, 2013 | SCCS | Breast | 1.04 (0.98, 1.09) |

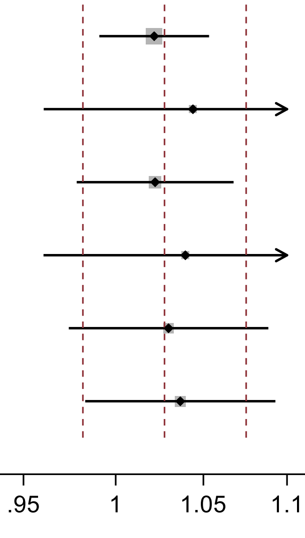

Figure legend: Forest plot shows the linear dose-response results (per 2 hours of sedentary time) from the inverse variance DerSimonian-Laird random-effects model after exclusion of each study from the meta-analysis. For each study, the square represents the summary relative risk (RR) estimate and the horizontal line across the square the corresponding 95% confidence interval (CI) computed from a meta-analysis excluding that study. The dashed vertical lines represent the results of the main meta-analysis.

Study abbreviations: CSDLH, Canadian Study of Diet, Lifestyle and Health; SUN, Seguimiento Universidad de Navarra Cohort Study; JACC, Japan Collaborative Cohort study; BWHS, Black Women Health Study; SCCS, Southern Community Cohort Study

Sanchez-Bayona 2021 (33798533)[82]; Hunter 2020 (32746843)[78]; Cao 2019 (30913861)[74]; Nomura 2016 (27632430)[60]; Catsburg 2014 (24781974)[55]; Cohen 2013 (23576427)[46]

#### Supplementary Figure 91: Linear dose-response meta-analysis of the association between television watching time and ovarian cancer incidence, leave-one-out sensitivity analysis.

**Television watching time, per 2 hours/day and ovarial cancer incidence, leave-one-out analysis**

| Author | Study | Outcome | RR (95% CI) |
|---|---|---|---|
| Buras, 2022 | NHS, NHS II | Ovary | 1.01 (0.92, 1.12) |
| Hunter, 2020 | UK Biobank | Ovary | 0.99 (0.91, 1.08) |
| Ukawa, 2018 | JACC | Ovary | 0.99 (0.93, 1.06) |
| Xiao, 2013 | NIH-AARP | Ovary, epithelial | 1.03 (0.95, 1.12) |

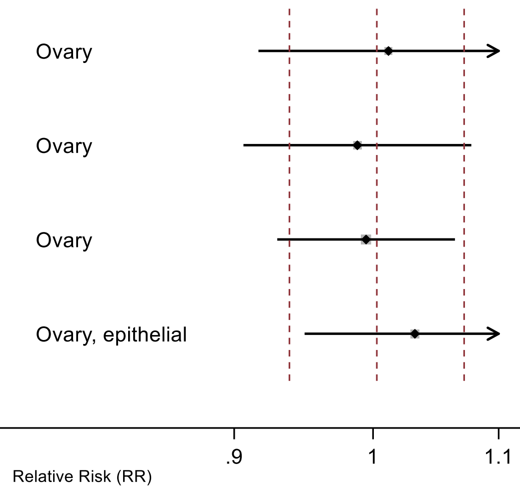

Figure legend: Forest plot shows the linear dose-response results (per 2 hours of sedentary time) from the inverse variance DerSimonian-Laird random-effects model after exclusion of each study from the meta-analysis. For each study, the square represents the summary relative risk (RR) estimate and the horizontal line across the square the corresponding 95% confidence interval (CI) computed from a meta-analysis excluding that study. The dashed vertical lines represent the results of the main meta-analysis.

Study abbreviations: NHS, Nurses' Health Study; JACC, Japan Collaborative Cohort study; NIH-AARP, National Institute of Health + American Association of Retired Persons Diet and Health Study

Buras 2022 (35094053)[84]; Hunter 2020 (32746843)[78]; Ukawa 2018 (29340890)[64]; Xiao 2013 (23966580)[49]

**Supplementary Figure 92: Linear dose-response meta-analysis of the association between television watching time and endometrial cancer incidence, leave-one-out sensitivity analysis.**

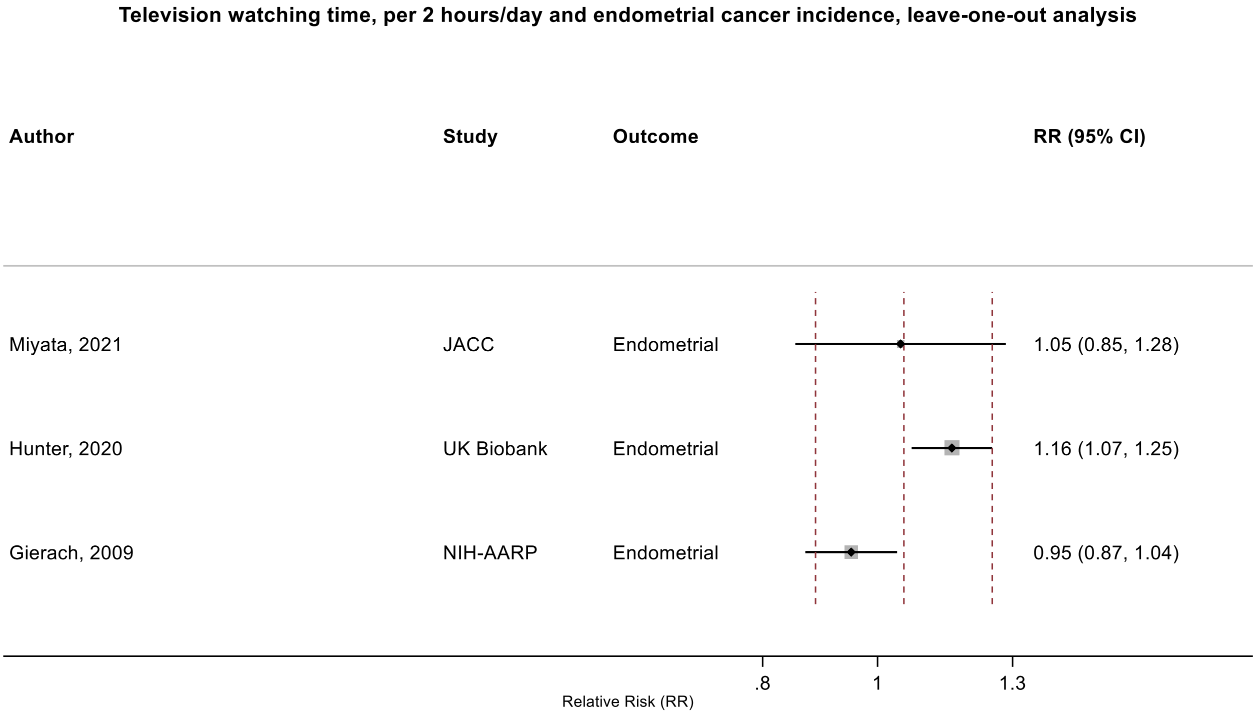

Figure legend: Forest plot shows the linear dose-response results (per 2 hours of sedentary time) from the inverse variance DerSimonian-Laird random-effects model after exclusion of each study from the meta-analysis. For each study, the square represents the summary relative risk (RR) estimate and the horizontal line across the square the corresponding 95% confidence interval (CI) computed from a meta-analysis excluding that study. The dashed vertical lines represent the results of the main meta-analysis.

Study abbreviations: JACC, Japan Collaborative Cohort study; NIH-AARP, National Institute of Health + American Association of Retired Persons Diet and Health Study

Miyata 2021 (32963209)[79]; Hunter 2020 (32746843)[78]; Gierach 2009 (19123463)[38]

### Supplementary Figure 93: Linear dose-response meta-analysis of the association between television watching time and colorectal cancer mortality.

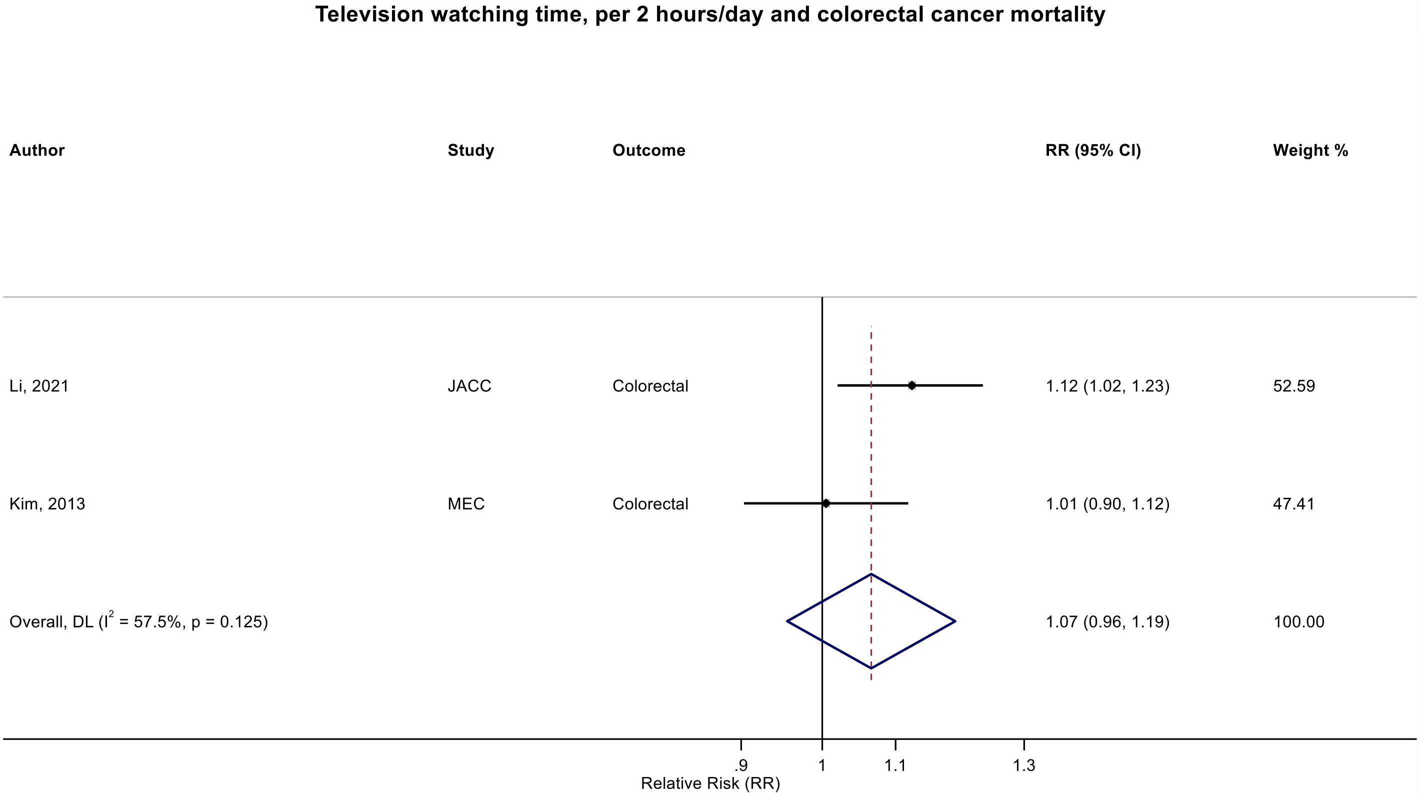

Figure legend: Forest plot shows the linear dose-response results (per 2 hours/day of sedentary time) from the inverse variance DerSimonian-Laird random-effects model. The diamond represents the summary relative risk (RR) estimate and the diamond's width the 95% confidence interval (CI). Each square and the horizontal line across the square represent the RR estimate of the individual study and its 95%CI.

Study abbreviations: JACC, Japan Collaborative Cohort study; MEC, Multiethnic Cohort Study

Li 2021 (33138348)[81]; Kim 2013 (24062293)[50]

Supplementary Materials

#### Supplementary Figure 94: Categorical highest versus lowest meta-analysis of the association between television watching time and breast cancer mortality.

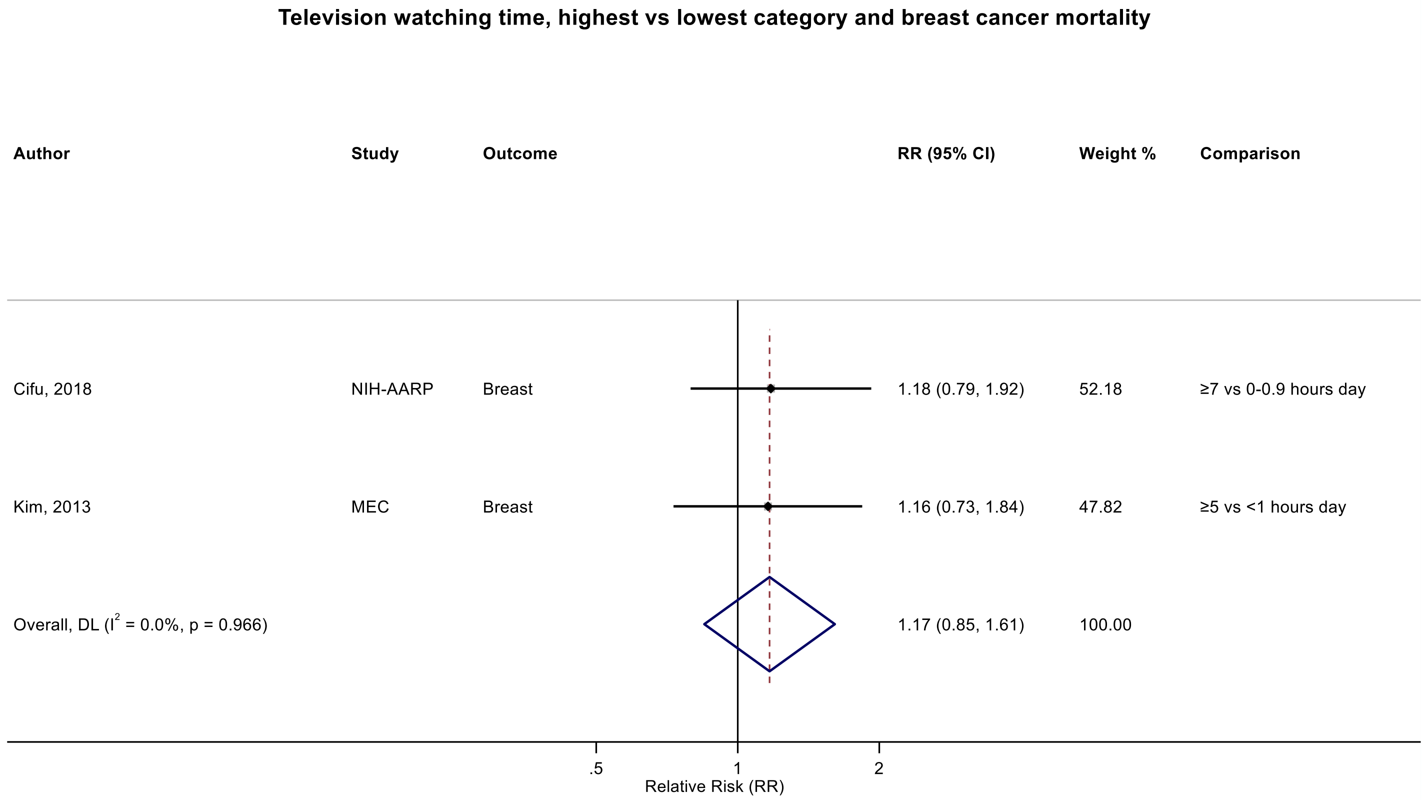

Figure legend: Forest plot shows the categorical comparison results from the individual studies for the associations with the largest contrast, as reported in the respective publication. Each square and the horizontal line across the square represent the relative risk (RR) estimate of the individual study and its 95% confidence interval (CI).

Study abbreviations: NIH-AARP, National Institute of Health + American Association of Retired Persons Diet and Health Study; MEC, Multiethnic Cohort Study

Cifu 2018 (30309689)[69]; Kim 2013 (24062293)[50]

#### Supplementary Figure 95: Linear dose-response meta-analysis of the association between television watching time and prostate cancer mortality.

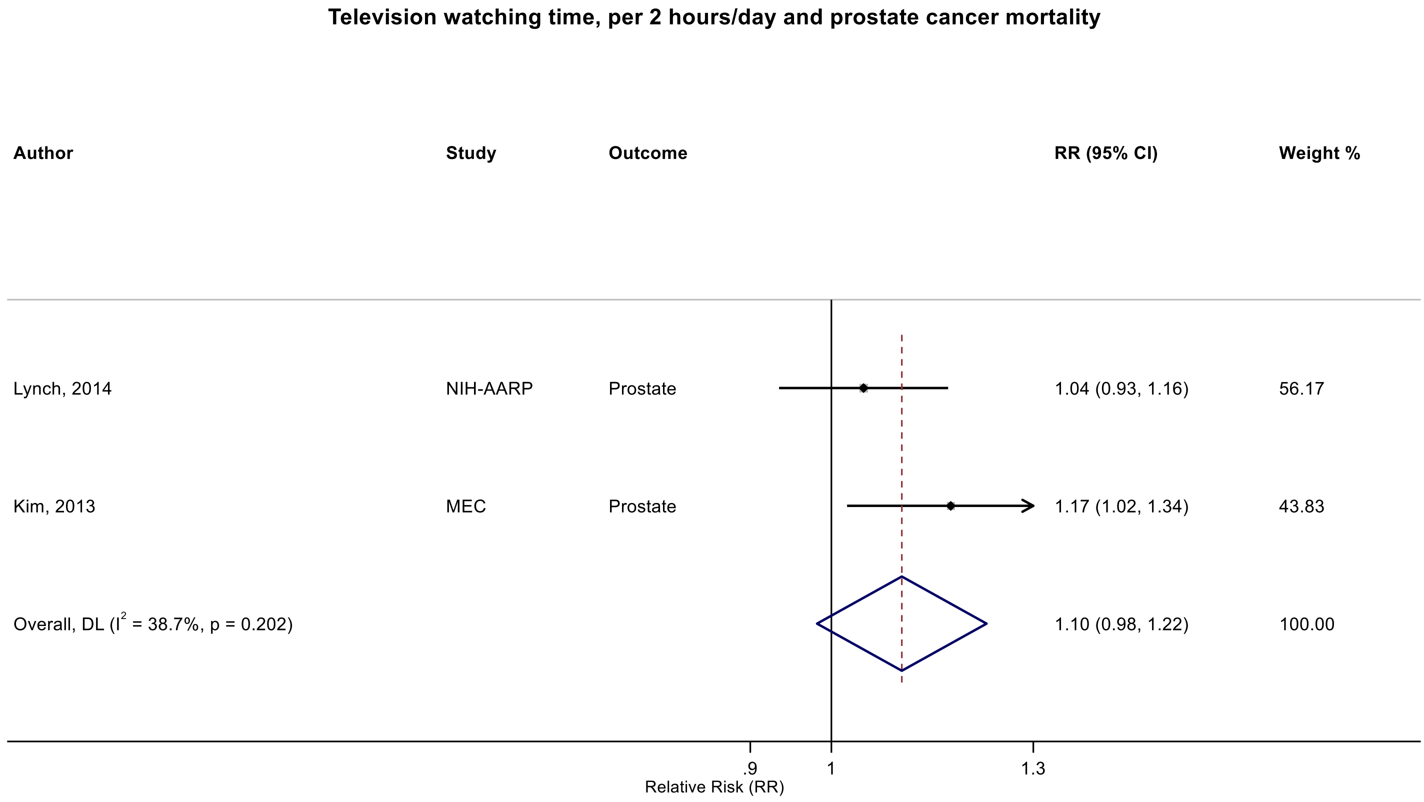

Figure legend: Forest plot shows the linear dose-response results (per 2 hours/day of sedentary time) from the inverse variance DerSimonian-Laird random-effects model. The diamond represents the summary relative risk (RR) estimate and the diamond's width the 95% confidence interval (CI). Each square and the horizontal line across the square represent the RR estimate of the individual study and its 95%CI.

Study abbreviations: NIH-AARP, National Institute of Health + American Association of Retired Persons Diet and Health Study; MEC, Multiethnic Cohort Study

Lynch 2014 (24526287)[53]; Kim 2013 (24062293)[50]

#### Mixed definitions of sedentary and sitting time

##### Supplementary Figure 96: Linear dose-response meta-analysis of the association between mixed definitions of sedentary time and oesophageal cancer incidence.

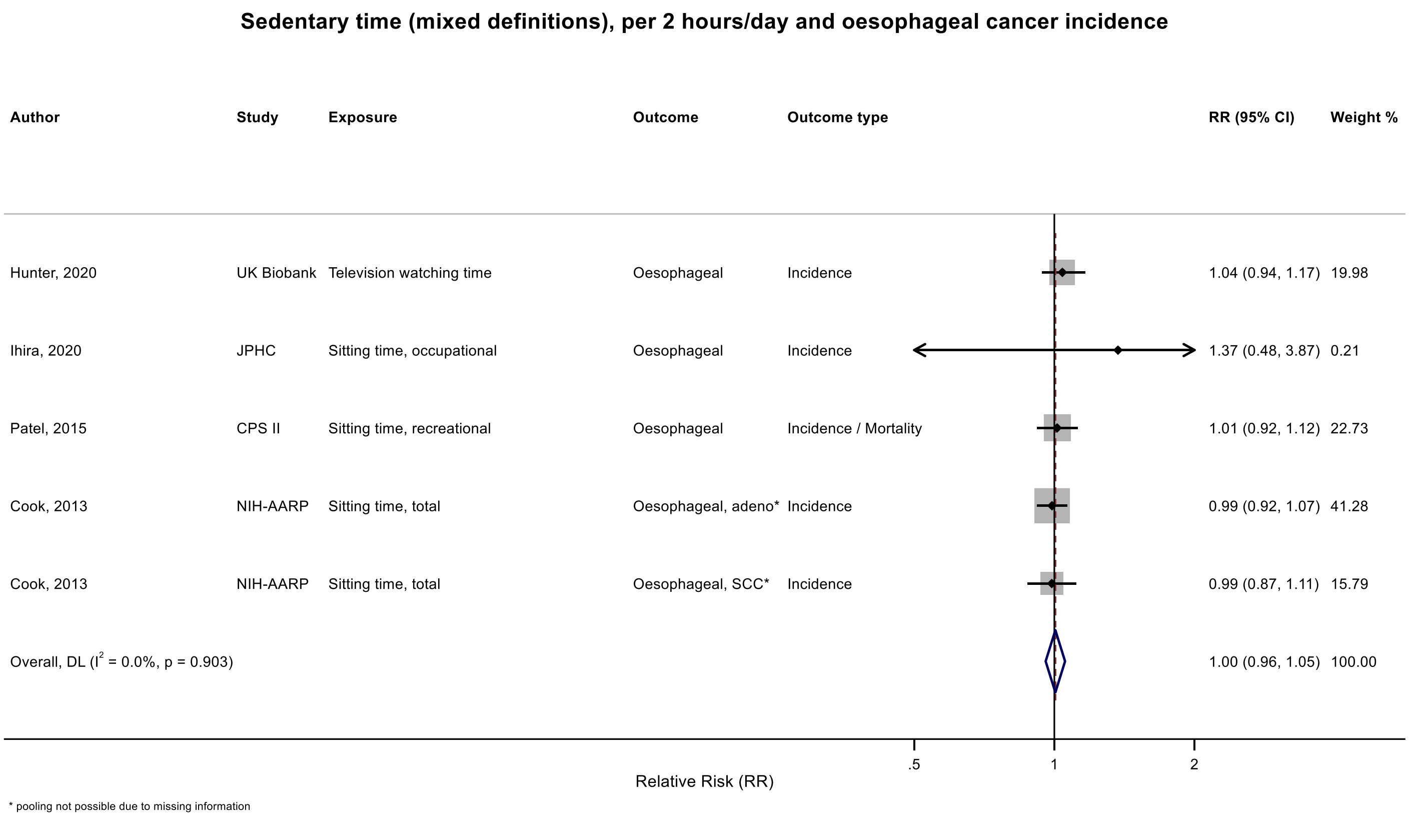

Figure legend: Forest plot shows the linear dose-response results (per 2 hours/day of sedentary time) from the inverse variance DerSimonian-Laird random-effects model. The diamond represents the summary relative risk (RR) estimate and the diamond's width the 95% confidence interval (CI). Each square and the horizontal line across the square represent the RR estimate of the individual study and its 95%CI.

Study abbreviations: CPS II, Cancer Prevention Study II; JPHC, Japan Public Health Center-Based Prospective Study; NIH-AARP, National Institute of Health – American Association for Retired Persons Diet and Health Study

Hunter 2020 (32746843)[78]; Ihira 2020 (31925977)[77]; Patel 2015 (26126627)[56]; Cook 2013 (24367697)[52]

### Supplementary Figure 97: Linear dose-response meta-analysis of the association between mixed definitions of sedentary time and stomach cancer incidence.

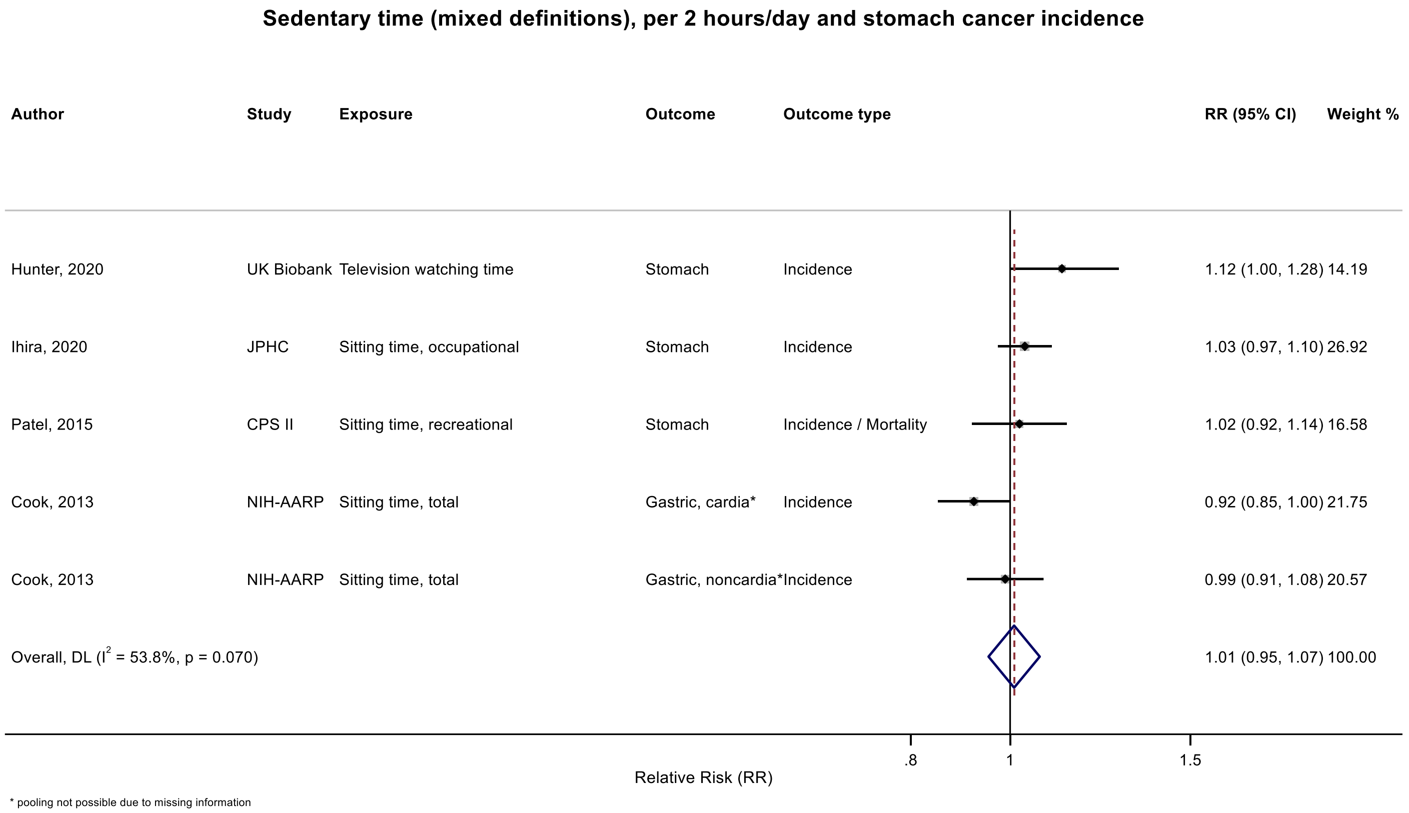

Figure legend: Forest plot shows the linear dose-response results (per 2 hours/day of sedentary time) from the inverse variance DerSimonian-Laird random-effects model. The diamond represents the summary relative risk (RR) estimate and the diamond's width the 95% confidence interval (CI). Each square and the horizontal line across the square represent the RR estimate of the individual study and its 95%CI.

Study abbreviations: CPS II, Cancer Prevention Study II; JPHC, Japan Public Health Center-Based Prospective Study; NIH-AARP, National Institute of Health – American Association for Retired Persons Diet and Health Study

Hunter 2020 (32746843)[78]; Ihira 2020 (31925977)[77]; Patel 2015 (26126627)[56]; Cook 2013 (24367697)[52]

Supplementary Materials

### Supplementary Figure 98: Linear dose-response meta-analysis of the association between mixed definitions of sedentary time and colorectal cancer incidence.

**Sedentary time (mixed definitions), per 2 hours/day and colorectal cancer incidence**

| Author | Study | Exposure | Outcome | Outcome type | RR (95% CI) | Weight % |
|---|---|---|---|---|---|---|
| Zhang, 2024 | UK Biobank | Screen time, recreational | Colorectal | Incidence | 1.05 (1.00, 1.11) | 10.13 |
| Guo, 2020 | Kailuan cohort | Sitting time, occupational | Colorectal | Incidence | 0.77 (0.59, 1.02) | 0.62 |
| Ihira, 2020 | JPHC | Sitting time, occupational | Colorectal | Incidence | 1.03 (0.98, 1.09) | 9.23 |
| Park, 2019 | MEC | Sitting time, total | Colorectal | Incidence | 0.99 (0.97, 1.01) | 20.05 |
| Nunez, 2018 | 45 and Up | Sitting time, total | Colorectal* | Incidence | 1.01 (0.94, 1.08) | 6.75 |
| Eaglehouse, 2017 | SCHS | Television watching time | Colorectal | Incidence | 1.04 (0.96, 1.12) | 6.23 |
| Gorczyca, 2017 | WHI | Sitting time, total | Colorectal | Incidence | 1.03 (0.99, 1.07) | 13.70 |
| Keum, 2016 | NHS | Television watching time | Colorectal | Incidence | 1.08 (1.02, 1.17) | 7.31 |
| Keum, 2016 | HPFS | Television watching time | Colorectal | Incidence | 1.00 (0.88, 1.14) | 2.53 |
| Patel, 2015 | CPS II | Sitting time, recreational | Colorectal | Incidence / Mortality | 0.99 (0.96, 1.03) | 14.63 |
| Simons, 2013 | NLCS | Sitting time, occupational | Colorectal* | Incidence | 1.06 (1.00, 1.12) | 8.83 |
| Overall, DL ($I^2$ = 46.5%, p = 0.044) | | | | | 1.02 (1.00, 1.04) | 100.00 |

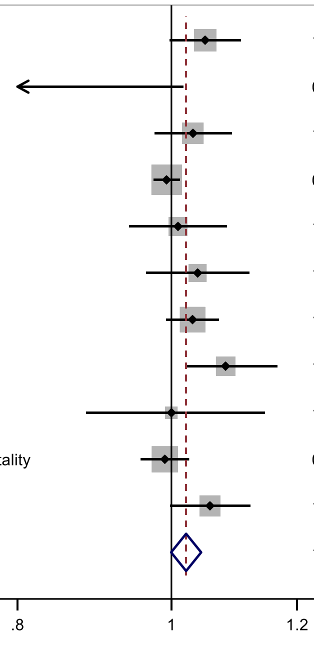

* pooled colon and rectal

Figure legend: Forest plot shows the linear dose-response results (per 2 hours/day of sedentary time) from the inverse variance DerSimonian-Laird random-effects model. The diamond represents the summary relative risk (RR) estimate and the diamond's width the 95% confidence interval (CI). Each square and the horizontal line across the square represent the RR estimate of the individual study and its 95%CI.

Study abbreviations: 45 and Up, Sax Institute's 45 and Up Study; CPS II, Cancer Prevention Study II; HPFS, Health Professionals Follow-Up Study; JPHC, Japan Public Health Center-Based Prospective Study; Kailuan cohort, Kailuan Cohort; MEC, Multiethnic Cohort Study; NHS, Nurses' Health Study; NLCS, The Netherlands Cohort Study; SCHS, Singapore Chinese Health Study; WHI, Women's Health Initiative Study

Simons 2013 (23420352)[45]; Patel 2015 (26126627)[56]; Keum 2016 (26649988)[58]; Gorczyca 2017 (28538039)[61]; Eaglehouse 2017 (28542077)[62]; Nunez 2018 (29510753)[66]; Park 2019 (30910780)[73]; Ihira 2020 (31925977)[77]; Guo 2020 (31773920)[76]; Zhang 2024 (38974470)[93]

#### Supplementary Figure 99: Linear dose-response meta-analysis of the association between mixed definitions of sedentary time and colorectal cancer incidence, by anatomical subsite.

**Sedentary time (mixed definitions), per 2 hours/day and colorectal cancer incidence, by anatomical subsite**

| Author | Study | Exposure | Outcome | Outcome type | RR (95% CI) | Weight % |
|---|---|---|---|---|---|---|
| **Colon** | | | | | | |
| Hunter, 2020 | UK Biobank | Television watching time | Colon | Incidence | 1.08 (1.02, 1.12) | 16.19 |
| Ihira, 2020 | JPHC | Sitting time, occupational | Colon | Incidence | 1.05 (0.98, 1.13) | 7.76 |
| Nunez, 2018 | 45 and Up | Sitting time, total | Colon | Incidence | 1.02 (0.96, 1.08) | 11.07 |
| Eaglehouse, 2017 | SCHS | Television watching time | Colon | Incidence | 1.04 (0.94, 1.14) | 3.92 |
| Gorczyca, 2017 | WHI | Sitting time, total | Colon | Incidence | 1.03 (0.99, 1.08) | 21.69 |
| Keum, 2016 | NHS | Television watching time | Colon | Incidence | 1.11 (0.98, 1.25) | 2.50 |
| Keum, 2016 | HPFS | Television watching time | Colon | Incidence | 1.08 (0.91, 1.28) | 1.34 |
| Simons, 2013 | NLCS | Sitting time, occupational | Colon | Incidence | 1.11 (1.04, 1.20) | 7.55 |
| Howard, 2008 | NIH-AARP | Sitting time, total | Colon | Incidence / Mortality | 1.04 (1.00, 1.08) | 27.98 |
| Subgroup, DL ($I^2$ = 0.0%, p = 0.538) | | | | | 1.05 (1.03, 1.07) | 100.00 |
| **Rectal** | | | | | | |
| Hunter, 2020 | UK Biobank | Television watching time | Rectal | Incidence | 0.98 (0.90, 1.06) | 20.53 |
| Ihira, 2020 | JPHC | Sitting time, occupational | Rectal | Incidence | 0.99 (0.90, 1.09) | 14.02 |
| Nunez, 2018 | 45 and Up | Sitting time, total | Rectal | Incidence | 0.98 (0.90, 1.07) | 17.38 |
| Eaglehouse, 2017 | SCHS | Television watching time | Rectal | Incidence | 1.04 (0.92, 1.17) | 9.12 |
| Gorczyca, 2017 | WHI | Sitting time, total | Rectal | Incidence | 1.00 (0.92, 1.10) | 16.47 |
| Keum, 2016 | HPFS | Television watching time | Rectal | Incidence | 1.06 (0.83, 1.37) | 2.12 |
| Keum, 2016 | NHS | Television watching time | Rectal | Incidence | 1.08 (0.92, 1.25) | 5.65 |
| Simons, 2013 | NLCS | Sitting time, occupational | Rectal | Incidence | 0.95 (0.87, 1.05) | 14.70 |
| Subgroup, DL ($I^2$ = 0.0%, p = 0.899) | | | | | 0.99 (0.96, 1.03) | 100.00 |

Heterogeneity between groups: p = 0.007

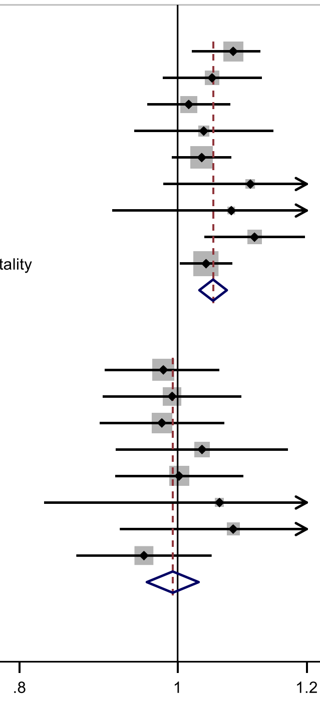

Figure legend: Forest plot shows the linear dose-response results (per 2 hours/day of sedentary time) from the inverse variance DerSimonian-Laird random-effects model. The diamond represents the summary relative risk (RR) estimate and the diamond's width the 95% confidence interval (CI). Each square and the horizontal line across the square represent the RR estimate of the individual study and its 95%CI.

Study abbreviations: 45 and Up, Sax Institute’s 45 and Up Study; HPFS, Health Professionals Follow-Up Study; JPHC, Japan Public Health Center-Based Prospective Study; NHS, Nurses' Health Study; NIH-AARP, National Institute of Health – American Association for Retired Persons Diet and Health Study; NLCS, The Netherlands Cohort Study; SCHS, Singapore Chinese Health Study; WHI, Women’s Health Initiative Study

Howard 2008 (18437512)[36]; Simons 2013 (23420352)[45]; Keum 2016 (26649988)[58]; Eaglehouse 2017 (28542077)[62]; Gorczyca 2017 (28538039)[61]; Nunez 2018 (29510753)[66]; Ihira 2020 (31925977)[77]; Hunter 2020 (32746843)[78]

#### Supplementary Figure 100: Linear dose-response meta-analysis of the association between mixed definitions of sedentary time and liver cancer risk.

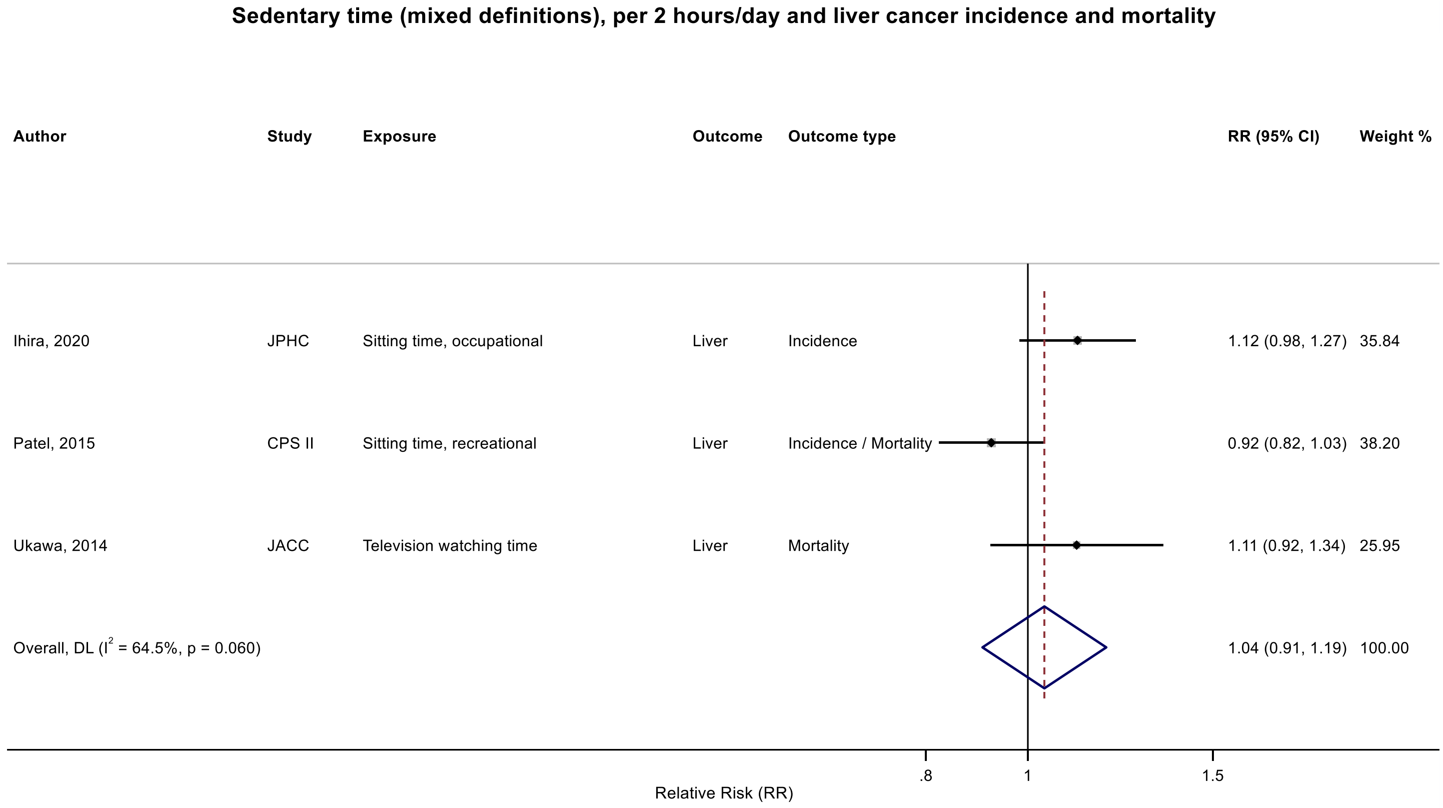

Figure legend: Forest plot shows the linear dose-response results (per 2 hours/day of sedentary time) from the inverse variance DerSimonian-Laird random-effects model. The diamond represents the summary relative risk (RR) estimate and the diamond's width the 95% confidence interval (CI). Each square and the horizontal line across the square represent the RR estimate of the individual study and its 95%CI.

Study abbreviations: CPS II, Cancer Prevention Study II; JACC, Japan Collaborative Cohort study; JPHC, Japan Public Health Center-Based Prospective Study

Ihira 2020 (31925977)[77]; Patel 2015 (26126627)[56]; Ukawa 2014 (24728669)[54]

### Supplementary Figure 101: Linear dose-response meta-analysis of the association between mixed definitions of sedentary time and pancreatic cancer incidence.

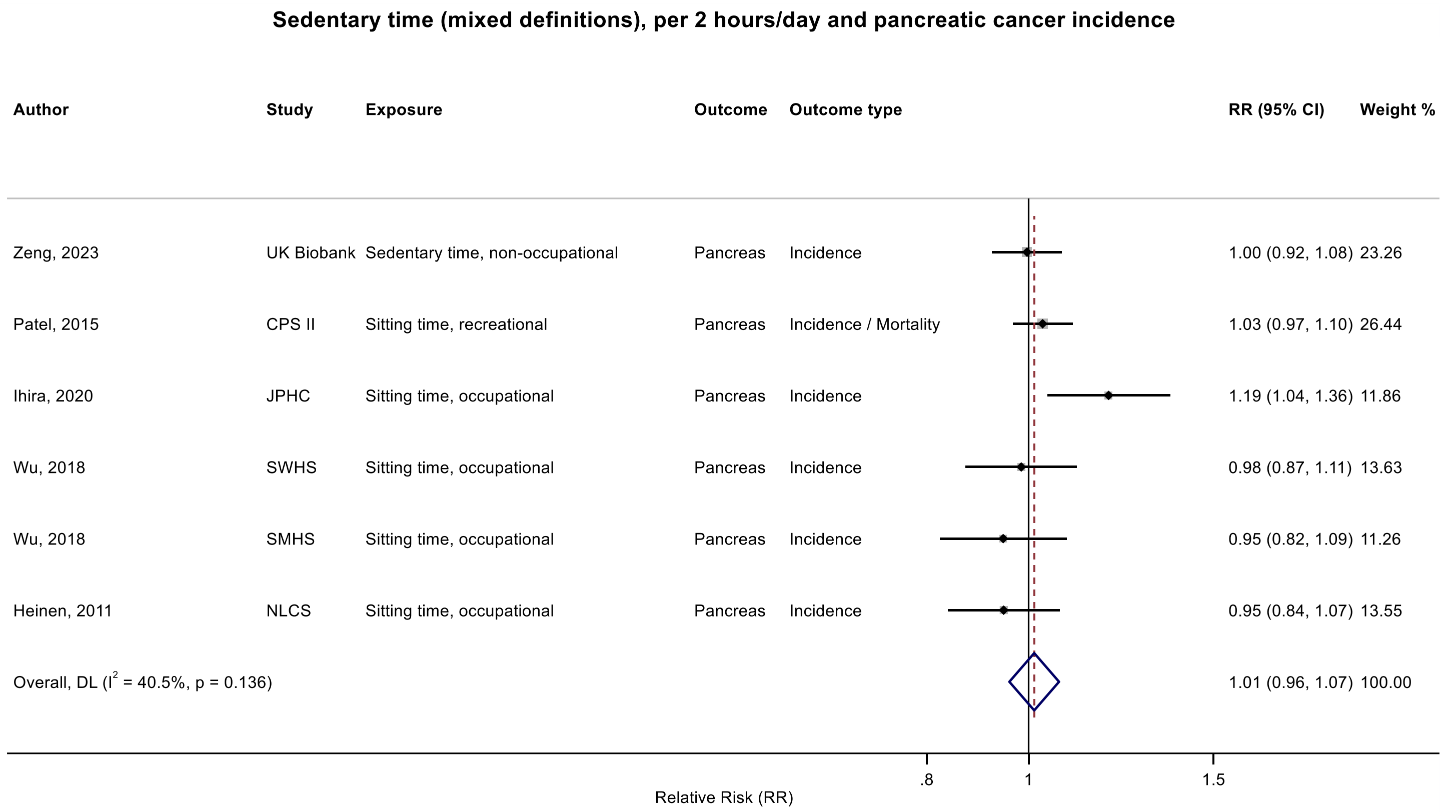

Figure legend: Forest plot shows the linear dose-response results (per 2 hours/day of sedentary time) from the inverse variance DerSimonian-Laird random-effects model. The diamond represents the summary relative risk (RR) estimate and the diamond's width the 95% confidence interval (CI). Each square and the horizontal line across the square represent the RR estimate of the individual study and its 95%CI.

Study abbreviations: CPS II, Cancer Prevention Study II; JPHC, Japan Public Health Center-Based Prospective Study; NLCS, The Netherlands Cohort Study; SMHS, Shanghai Men’s Health Study; SWHS, Shanghai Women’s Health Study; UK Biobank, UK Biobank

Heinen 2011 (21955648)[43]; Wu 2018 (29475964)[65]; Ihira 2020 (31925977)[77]; Patel 2015 (26126627)[56]; Zeng 2023 (38066552)[92]

#### Supplementary Figure 102: Linear dose-response meta-analysis of the association between mixed definitions of sedentary time and lung cancer incidence.

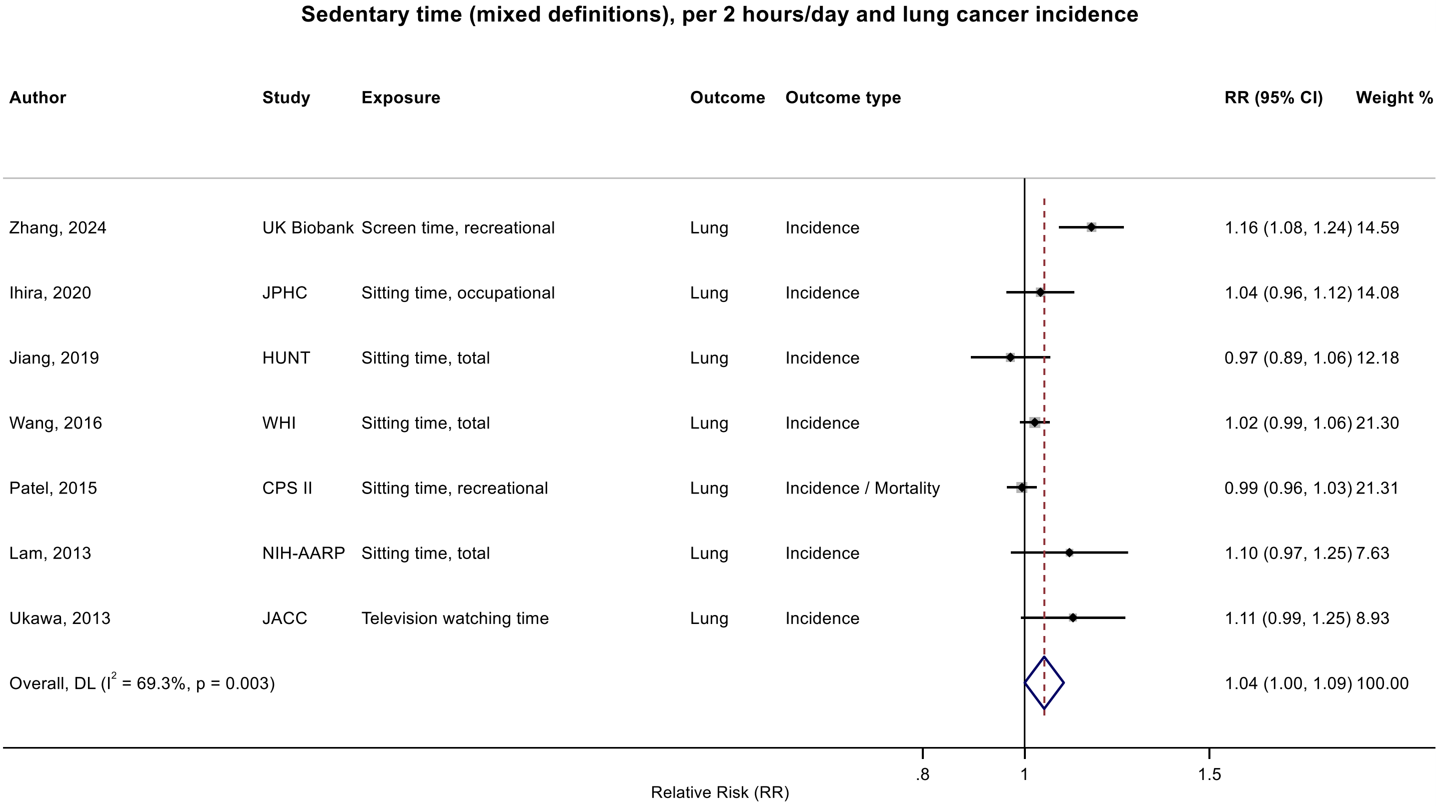

Figure legend: Forest plot shows the linear dose-response results (per 2 hours/day of sedentary time) from the inverse variance DerSimonian-Laird random-effects model. The diamond represents the summary relative risk (RR) estimate and the diamond's width the 95% confidence interval (CI). Each square and the horizontal line across the square represent the RR estimate of the individual study and its 95%CI.

Study abbreviations: CPS II, Cancer Prevention Study II; HUNT, The Trøndelag Health Study; JACC, Japan Collaborative Cohort study; JPHC, Japan Public Health Center-Based Prospective Study; NIH-AARP, National Institute of Health – American Association for Retired Persons Diet and Health Study; JACC, Japan Collaborative Cohort study; WHI, Women's Health Initiative Study

Ukawa 2013 (23686441)[47]; Lam 2013 (23940620)[48]; Patel 2015 (26126627)[56]; Wang 2016 (27439221)[59]; Jiang 2019 (30859092)[72]; Ihira 2020 (31925977)[77]; Zhang 2024 (38974470)[93]

#### Supplementary Figure 103: Linear dose-response meta-analysis of the association between mixed definitions of sedentary time and melanoma cancer incidence.

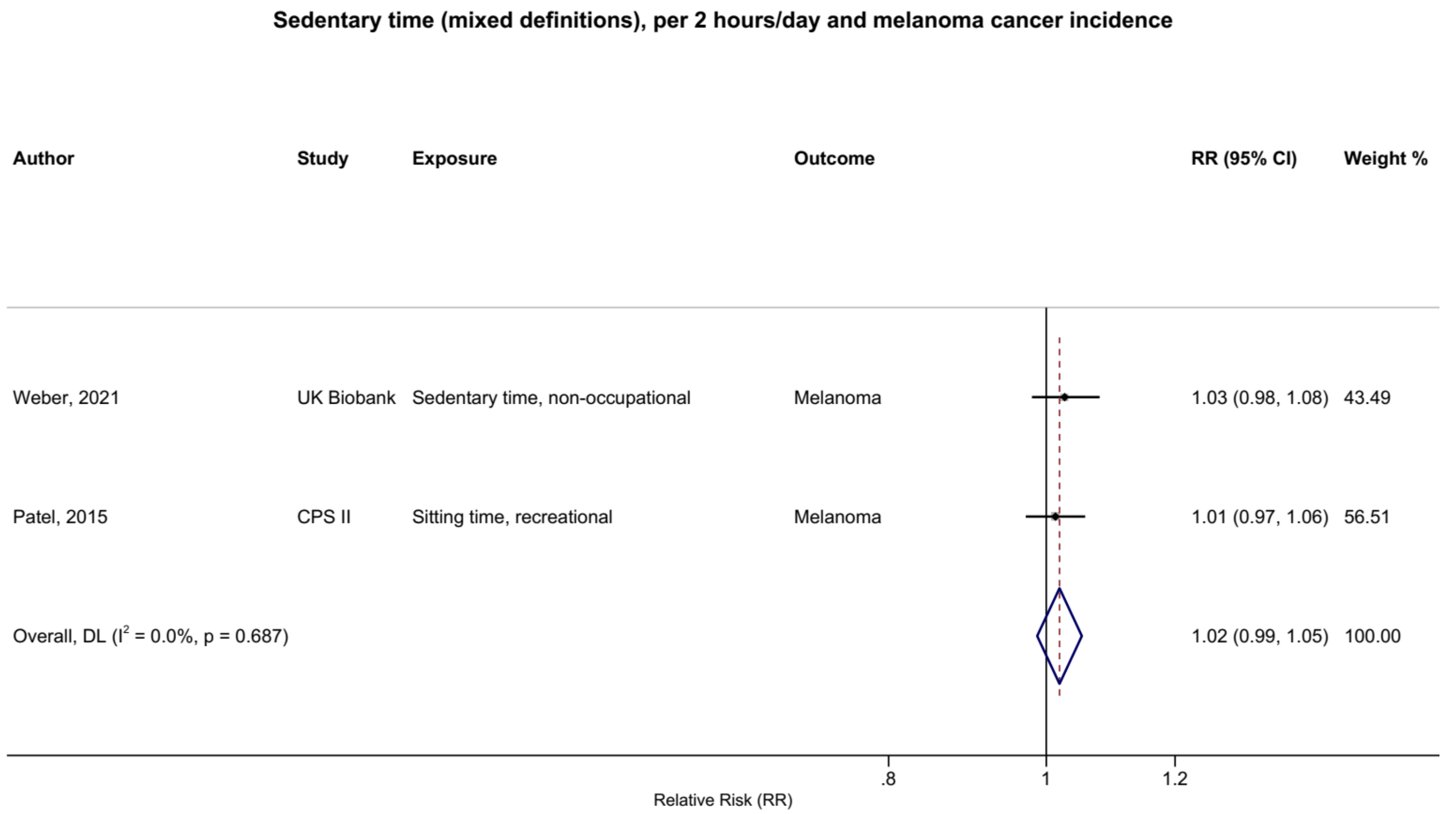

Figure legend: Forest plot shows the linear dose-response results (per 2 hours/day of sedentary time) from the inverse variance DerSimonian-Laird random-effects model. The diamond represents the summary relative risk (RR) estimate and the diamond's width the 95% confidence interval (CI). Each square and the horizontal line across the square represent the RR estimate of the individual study and its 95%CI.

Study abbreviations: CPS II, Cancer Prevention Study II; UK Biobank, UK Biobank

Patel 2015 (26126627)[56]; Weber 2021 (34059803)[83]

### Supplementary Figure 104: Linear dose-response meta-analysis of the association between mixed definitions of sedentary time and breast cancer incidence.

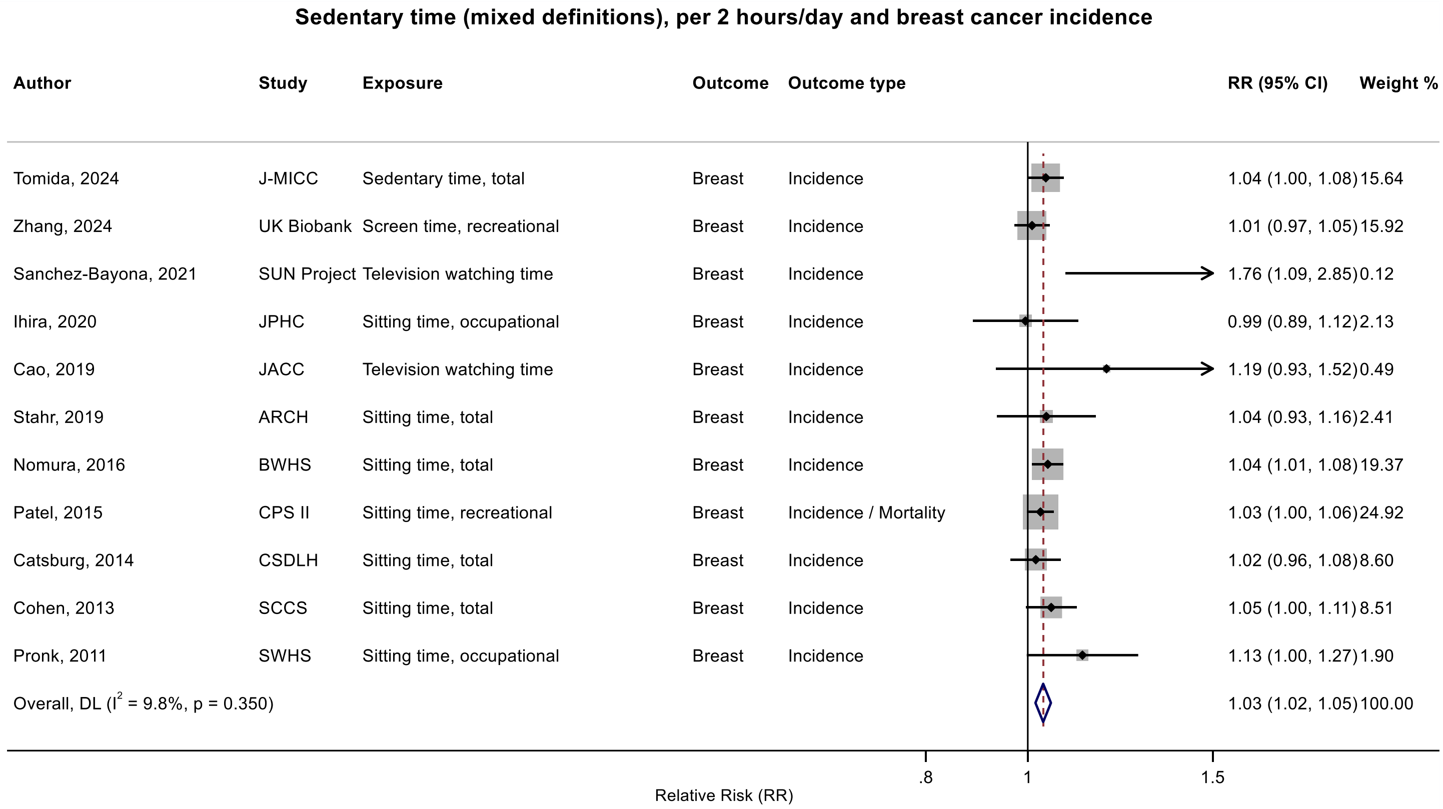

Figure legend: Forest plot shows the linear dose-response results (per 2 hours/day of sedentary time) from the inverse variance DerSimonian-Laird random-effects model. The diamond represents the summary relative risk (RR) estimate and the diamond's width the 95% confidence interval (CI). Each square and the horizontal line across the square represent the RR estimate of the individual study and its 95%CI.

Study abbreviations: ARCH, Arkansas Rural Community Health Study Cohort; BWHS, Black Women's Health Study; CPS II, Cancer Prevention Study II; CSDLH, Canadian Study of Diet, Lifestyle and Health; JACC, Japan Collaborative Cohort study; J-MICC, Japan Multi-Institutional Collaborative Cohort Study; JPHC, Japan Public Health Center-Based Prospective Study; SCCS, Southern Community Cohort Study; SWHS, Shanghai Women’s Health Study

Pronk 2011 (21934685)[42]; Cohen 2013 (23576427)[46]; Catsburg 2014 (24781974)[55]; Patel 2015 (26126627)[56]; Nomura 2016 (27632430)[60]; Cao 2019 (30913861)[74]; Stahr 2019 (35117114)[85]; Ihira 2020 (31925977)[77]; Sanchez-Bayona 2021 (33798533)[82]; Tomida 2024 (38041484)[91]; Zhang 2024 (38974470)[93]

#### Supplementary Figure 105: Linear dose-response meta-analysis of the association between mixed definitions of sedentary time and breast cancer incidence by menopausal status.

**Sedentary time (mixed definitions), per 2 hours/day and breast cancer incidence, by menopoause status**

| Author | Study | Exposure | Outcome | Outcome type | RR (95% CI) | Weight % |
|---|---|---|---|---|---|---|
| **Premenopausal** | | | | | | |
| Tomida, 2024 | J-MICC | Sedentary time, total | Breast | Incidence | 1.06 (1.00, 1.17) | 21.96 |
| Sanchez-Bayona, 2021 | SUN Project | Television watching time | Breast | Incidence | 2.40 (1.26, 4.56) | 0.45 |
| Hunter, 2020 | UK Biobank | Television watching time | Breast | Incidence | 1.02 (0.94, 1.12) | 17.85 |
| Cao, 2019 | JACC | Television watching time | Breast | Incidence | 1.03 (0.78, 1.36) | 2.30 |
| Stahr, 2019 | ARCH | Sitting time, total | Breast | Incidence | 1.04 (0.88, 1.24) | 5.61 |
| Nomura, 2016 | BWHS | Sitting time, total | Breast | Incidence | 1.02 (0.97, 1.08) | 31.97 |
| Catsburg, 2014 | CSDLH | Sitting time, total | Breast | Incidence | 1.01 (0.93, 1.10) | 19.85 |
| Subgroup, DL ($I^2$ = 20.1%, p = 0.276) | | | | | 1.03 (0.99, 1.08) | 100.00 |
| **Postmenopausal** | | | | | | |
| Tomida, 2024 | J-MICC | Sedentary time, total | Breast | Incidence | 1.02 (0.96, 1.08) | 7.33 |
| Sanchez-Bayona, 2021 | SUN Project | Television watching time | Breast | Incidence | 1.07 (0.45, 2.57) | 0.04 |
| Hunter, 2020 | UK Biobank | Television watching time | Breast | Incidence | 1.01 (0.96, 1.06) | 10.19 |
| Cao, 2019 | JACC | Television watching time | Breast | Incidence | 1.65 (1.02, 2.65) | 0.12 |
| Stahr, 2019 | ARCH | Sitting time, total | Breast | Incidence | 1.04 (0.91, 1.19) | 1.44 |
| Nomura, 2017 | WHI | Sitting time, total | Breast | Incidence | 1.00 (0.96, 1.04) | 15.07 |
| Nomura, 2016 | BWHS | Sitting time, total | Breast | Incidence | 1.05 (1.00, 1.10) | 10.88 |
| Catsburg, 2014 | CSDLH | Sitting time, total | Breast | Incidence | 1.03 (0.95, 1.11) | 4.35 |
| Hildebrand, 2013 | CPS II | Sitting time, recreational | Breast | Incidence / Mortality | 1.02 (0.99, 1.05) | 22.83 |
| George, 2010 | NIH-AARP | Sitting time, total | Breast | Incidence | 1.03 (1.00, 1.06) | 23.87 |
| Dirx, 2001 | NLCS | Sitting time, occupational | Breast | Incidence | 0.94 (0.86, 1.02) | 3.87 |
| Subgroup, DL ($I^2$ = 7.4%, p = 0.373) | | | | | 1.02 (1.00, 1.04) | 100.00 |

Heterogeneity between groups: p = 0.532

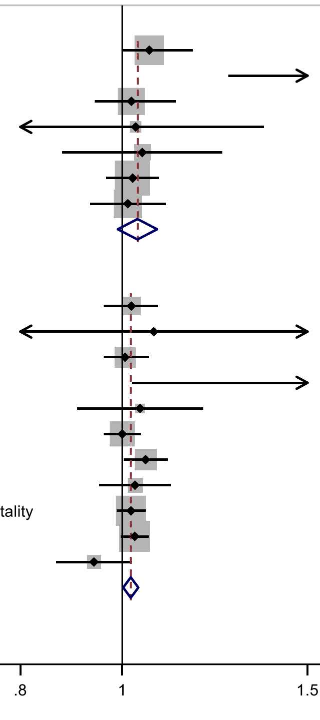

Figure legend: Forest plot shows the linear dose-response results (per 2 hours/day of sedentary time) from the inverse variance DerSimonian-Laird random-effects model. The diamond represents the summary relative risk (RR) estimate and the diamond's width the 95% confidence interval (CI). Each square and the horizontal line across the square represent the RR estimate of the individual study and its 95%CI.

Study abbreviations: ARCH, Arkansas Rural Community Health Study Cohort; BWHS, Black Women's Health Study; CPS II, Cancer Prevention Study II; CSDLH, Canadian Study of Diet, Lifestyle and Health; JACC, Japan Collaborative Cohort study; J-MICC, Japan Multi-Institutional Collaborative Cohort Study; NLCS, The Netherlands Cohort Study; WHI, Women's Health Initiative Study

Tomida 2024 (38041484)[91]; Sanchez-Bayona 2021 (33798533)[82]; Hunter 2020 (32746843)[78]; Cao 2019 (30913861)[74]; Stahr 2019 (35117114)[85]; Nomura 2017 (28975422)[63]; Nomura 2016 (27632430)[60]; Catsburg 2014 (24781974)[55]; Hildebrand 2013 (24097200)[51]; George 2010 (20864719)[39]; Dirx 2001 (11745243)[33]

#### Supplementary Figure 106: Linear dose-response meta-analysis of the association between mixed definitions of sedentary time and endometrial cancer incidence.

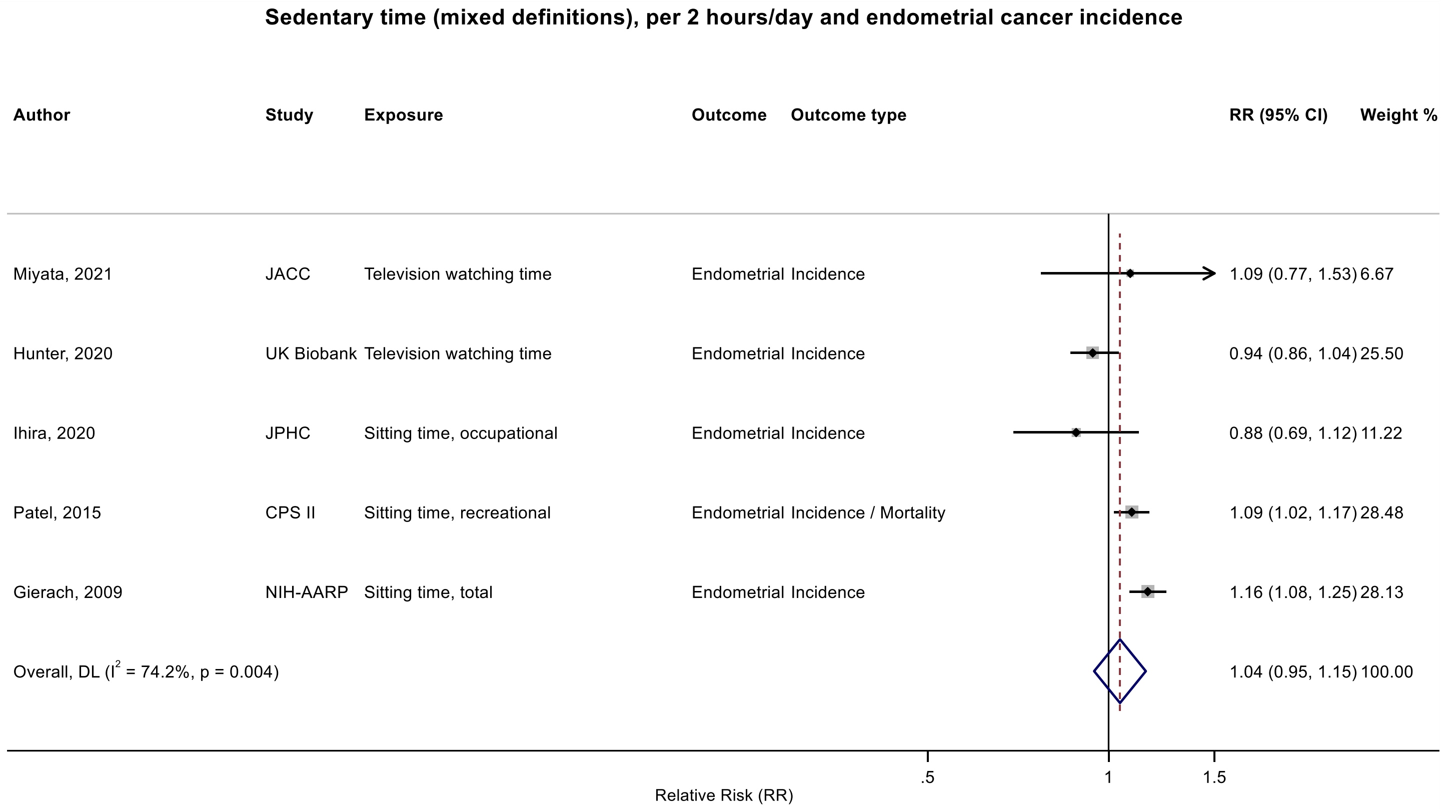

Figure legend: Forest plot shows the linear dose-response results (per 2 hours/day of sedentary time) from the inverse variance DerSimonian-Laird random-effects model. The diamond represents the summary relative risk (RR) estimate and the diamond's width the 95% confidence interval (CI). Each square and the horizontal line across the square represent the RR estimate of the individual study and its 95%CI.

Study abbreviations: CPS II, Cancer Prevention Study II; JACC, Japan Collaborative Cohort study; JPHC, Japan Public Health Center-Based Prospective Study; NIH-AARP, National Institute of Health – American Association for Retired Persons Diet and Health Study

Gierach 2009 (19123463)[38]; Patel 2015 (26126627)[56]; Ihira 2020 (31925977)[77]; Hunter 2020 (32746843)[78]; Miyata 2021 (32963209)[79]

#### Supplementary Figure 107: Linear dose-response meta-analysis of the association between mixed definitions of sedentary time and ovarian cancer incidence.

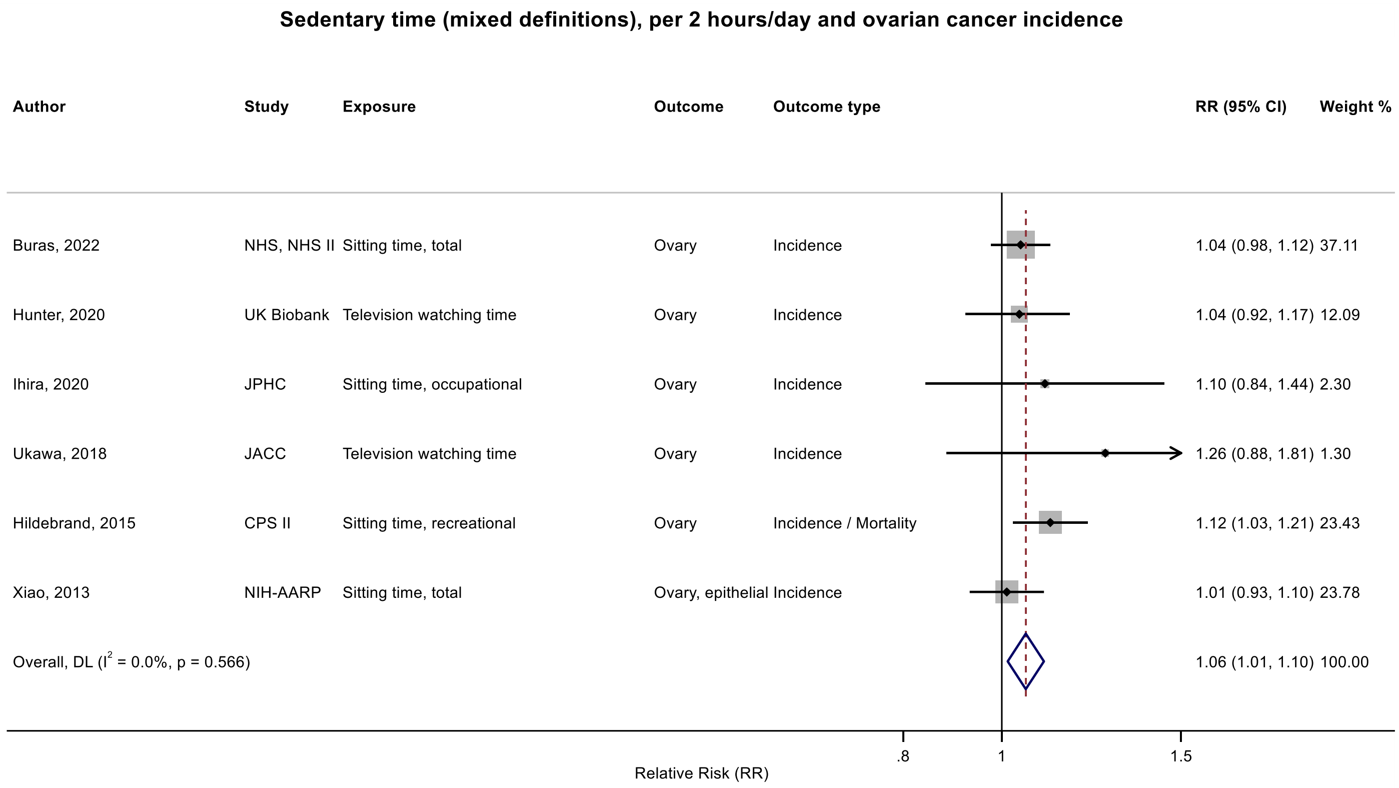

Figure legend: Forest plot shows the linear dose-response results (per 2 hours/day of sedentary time) from the inverse variance DerSimonian-Laird random-effects model. The diamond represents the summary relative risk (RR) estimate and the diamond's width the 95% confidence interval (CI). Each square and the horizontal line across the square represent the RR estimate of the individual study and its 95%CI.

Study abbreviations: CPS II, Cancer Prevention Study II; JACC, Japan Collaborative Cohort study;JPHC, Japan Public Health Center-Based Prospective Study; NHS, Nurses' Health Study; NHS II, Nurses' Health Study II; NIH-AARP, National Institute of Health – American Association for Retired Persons Diet and Health Study

Xiao 2013 (23966580)[49]; Hildebrand 2015 (26335264)[57]; Ukawa 2018 (29340890)[64]; Ihira 2020 (31925977)[77]; Buras 2022 (35094053)[84]; Hunter 2020 (32746843)[78]

#### Supplementary Figure 108: Linear dose-response meta-analysis of the association between mixed definitions of sedentary time and prostate cancer incidence.

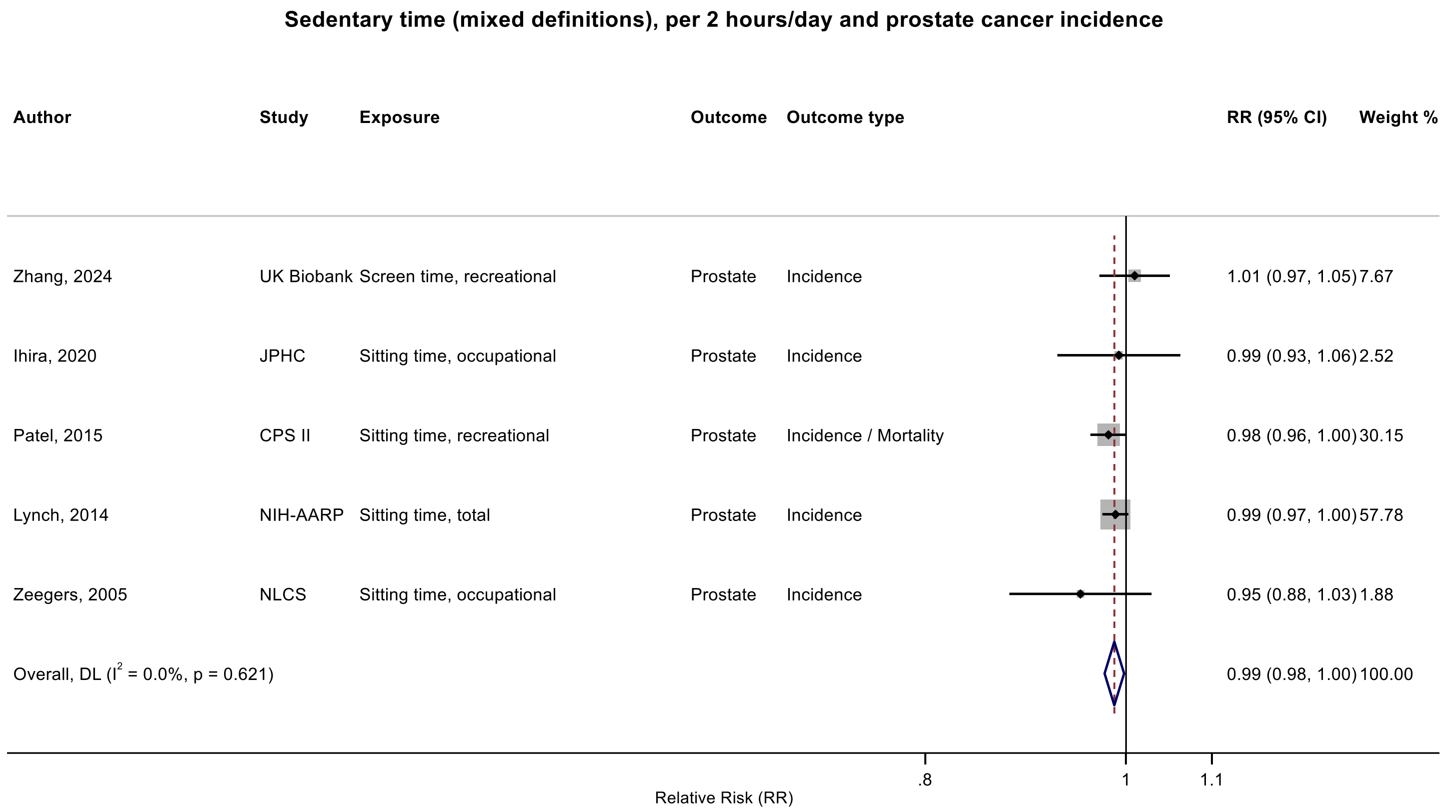

Figure legend: Forest plot shows the linear dose-response results (per 2 hours/day of sedentary time) from the inverse variance DerSimonian-Laird random-effects model. The diamond represents the summary relative risk (RR) estimate and the diamond's width the 95% confidence interval (CI). Each square and the horizontal line across the square represent the RR estimate of the individual study and its 95%CI.

Study abbreviations: CPS II, Cancer Prevention Study II; JPHC, Japan Public Health Center-Based Prospective Study; NIH-AARP, National Institute of Health – American Association for Retired Persons Diet and Health Study; NLCS, The Netherlands Cohort Study

Zeegers 2005 (15941961)[34]; Ihira 2020 (31925977)[77]; Patel 2015 (26126627)[56]; Lynch 2014 (24526287)[53]; Zhang 2024 (38974470)[93]

**Supplementary Figure 109: Linear dose-response meta-analysis of the association between mixed definitions of sedentary time and advanced prostate cancer incidence.**

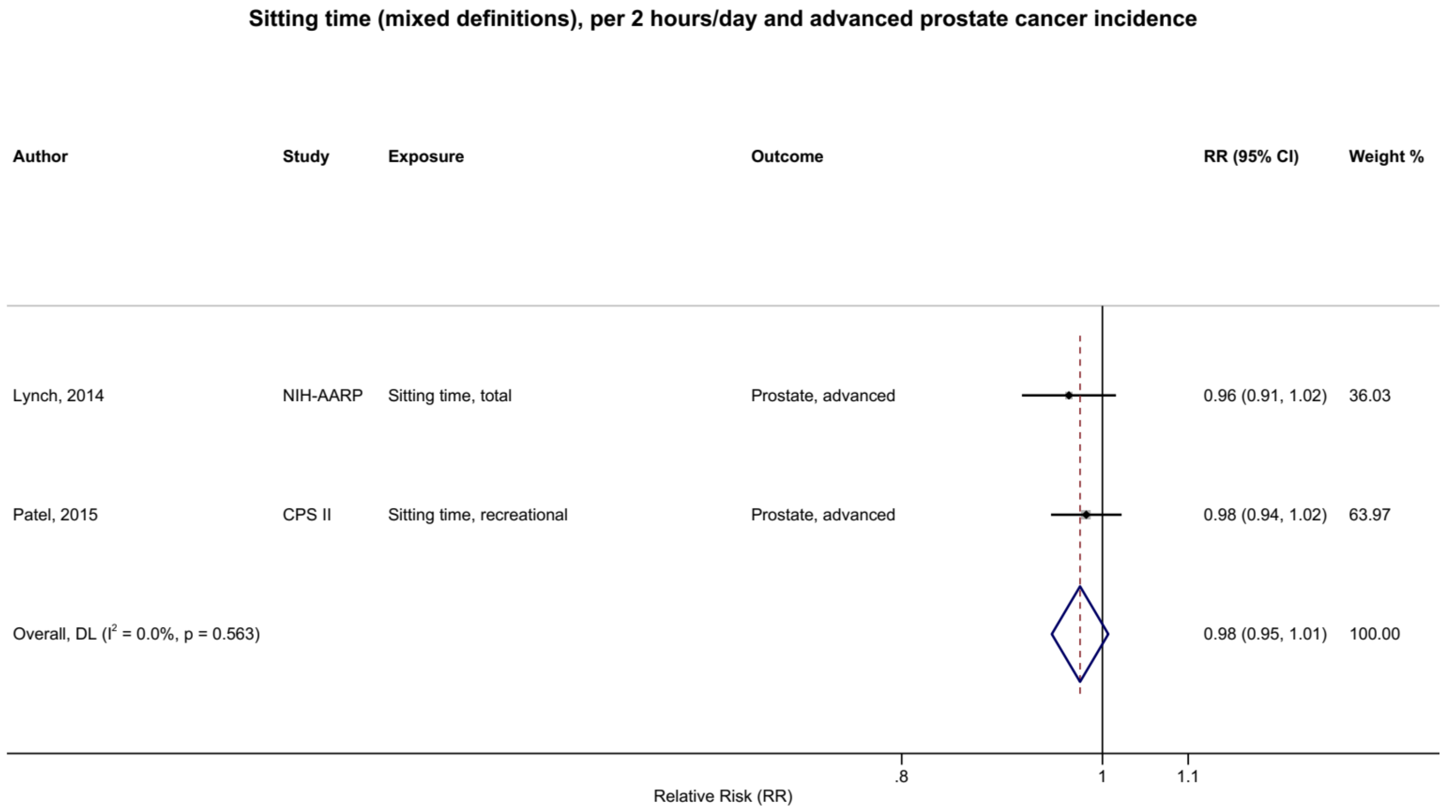

Figure legend: Forest plot shows the linear dose-response results (per 2 hours/day of sedentary time) from the inverse variance DerSimonian-Laird random-effects model. The diamond represents the summary relative risk (RR) estimate and the diamond's width the 95% confidence interval (CI). Each square and the horizontal line across the square represent the RR estimate of the individual study and its 95%CI.

Study abbreviations: CPS II, Cancer Prevention Study II; NIH-AARP, National Institute of Health – American Association for Retired Persons Diet and Health Study

Lynch 2014 (24526287)[53]; Patel 2015 (26126627)[56]

#### Supplementary Figure 110: Linear dose-response meta-analysis of the association between mixed definitions of sedentary time and kidney cancer incidence.

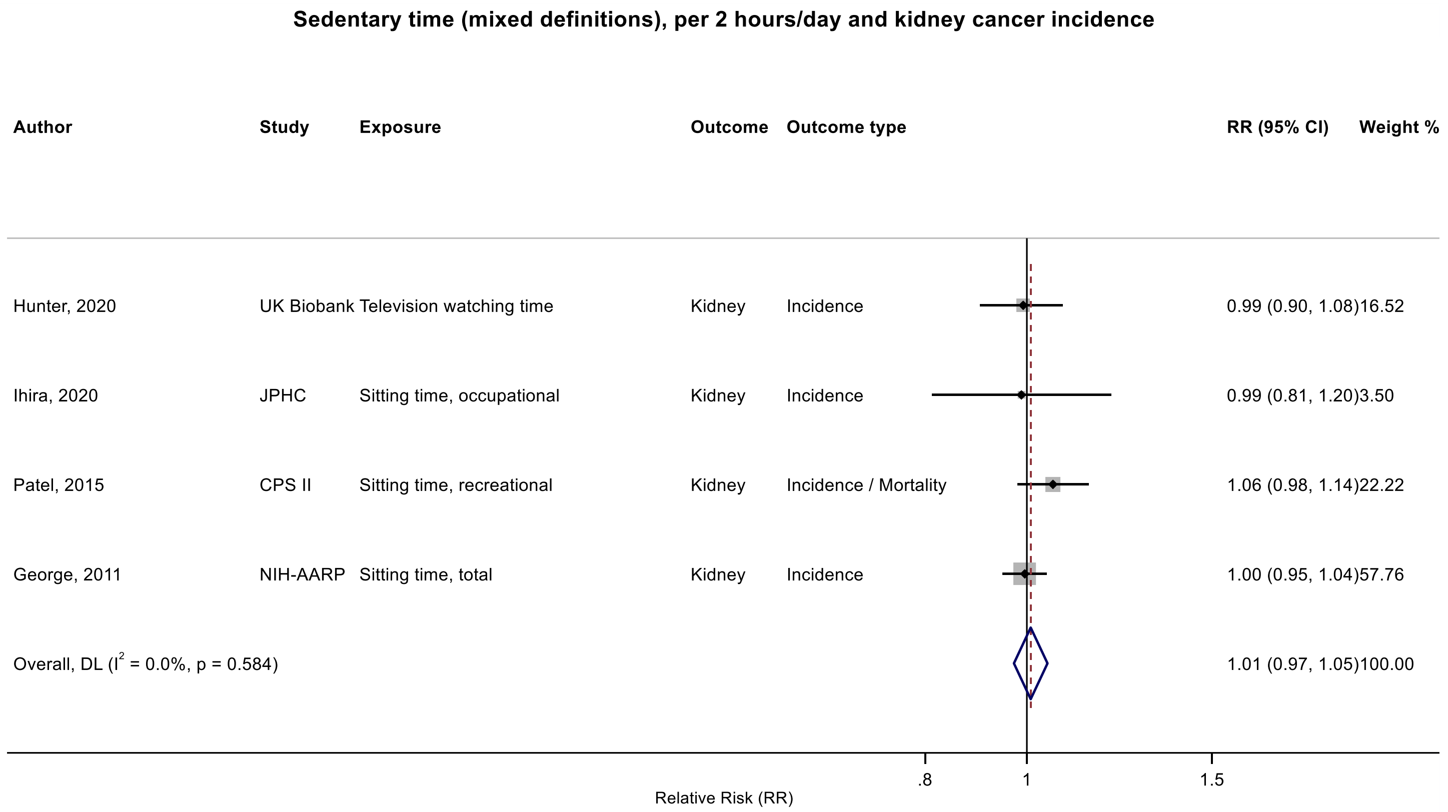

Figure legend: Forest plot shows the linear dose-response results (per 2 hours/day of sedentary time) from the inverse variance DerSimonian-Laird random-effects model. The diamond represents the summary relative risk (RR) estimate and the diamond's width the 95% confidence interval (CI). Each square and the horizontal line across the square represent the RR estimate of the individual study and its 95%CI.

Study abbreviations: CPS II, Cancer Prevention Study II; JPHC, Japan Public Health Center-Based Prospective Study; NIH-AARP, National Institute of Health – American Association for Retired Persons Diet and Health Study

George 2011 (21737302)[41]; Patel 2015 (26126627)[56]; Ihira 2020 (31925977)[77]; Hunter 2020 (32746843)[78]

### Supplementary Figure 111: Linear dose-response and categorical highest versus lowest meta-analysis of the association between mixed definitions of sedentary time and bladder cancer incidence.

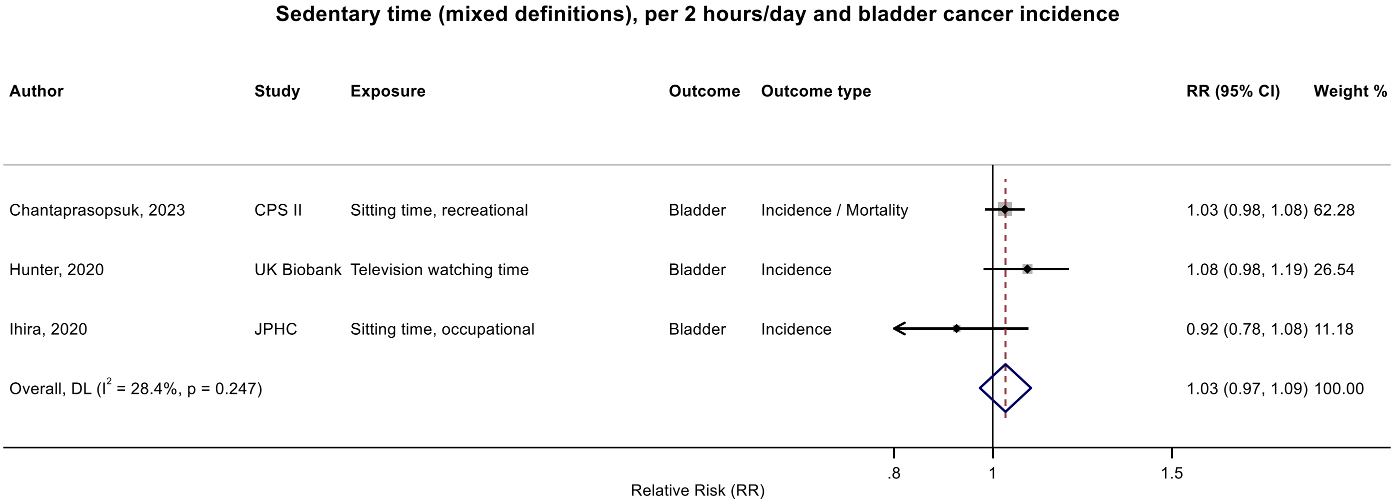

Figure legend: Forest plot shows the linear dose-response results (per 2 hours/day of sedentary time) from the inverse variance DerSimonian-Laird random-effects model. The diamond represents the summary relative risk (RR) estimate and the diamond's width the 95% confidence interval (CI). Each square and the horizontal line across the square represent the RR estimate of the individual study and its 95%CI.

Study abbreviations: CPS II, Cancer Prevention Study II; JPHC, Japan Public Health Center-Based Prospective Study

Chantaprasopsuk 2023 (37202564)[89]; Hunter 2020 (32746843)[78]; Ihira 2020 (31925977)[77]

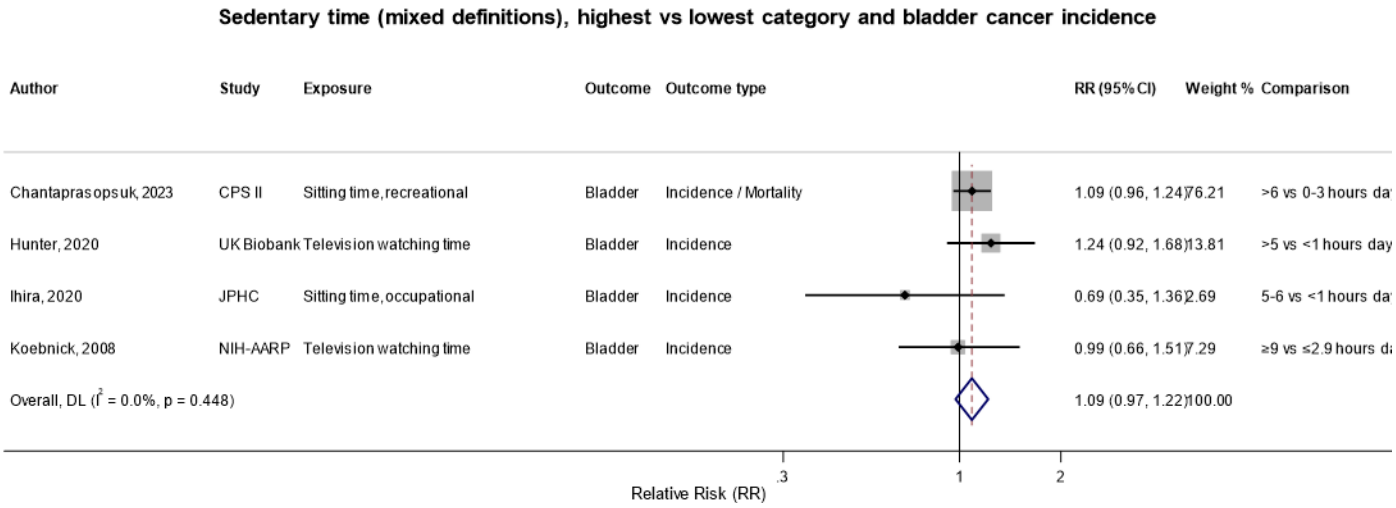

Figure legend: Forest plot shows the categorical comparison results from the individual studies for the associations with the largest contrast, as reported in the respective publication. Each square and the horizontal line across the square represent the relative risk (RR) estimate of the individual study and its 95% confidence interval (CI).

Study abbreviations: CPS II, Cancer Prevention Study II; JPHC, Japan Public Health Center-Based Prospective Study; NIH-AARP, National Institute of Health – American Association for Retired Persons Diet and Health Study

Chantaprasopsuk 2023 (37202564)[89]; Hunter 2020 (32746843)[78]; Ihira 2020 (31925977)[77]; Koebnick 2008 (18483344)[37]

**Supplementary Figure 112: Linear dose-response meta-analysis of the association between mixed definitions of sedentary time and oesophageal cancer incidence, leave-one-out sensitivity analysis.**

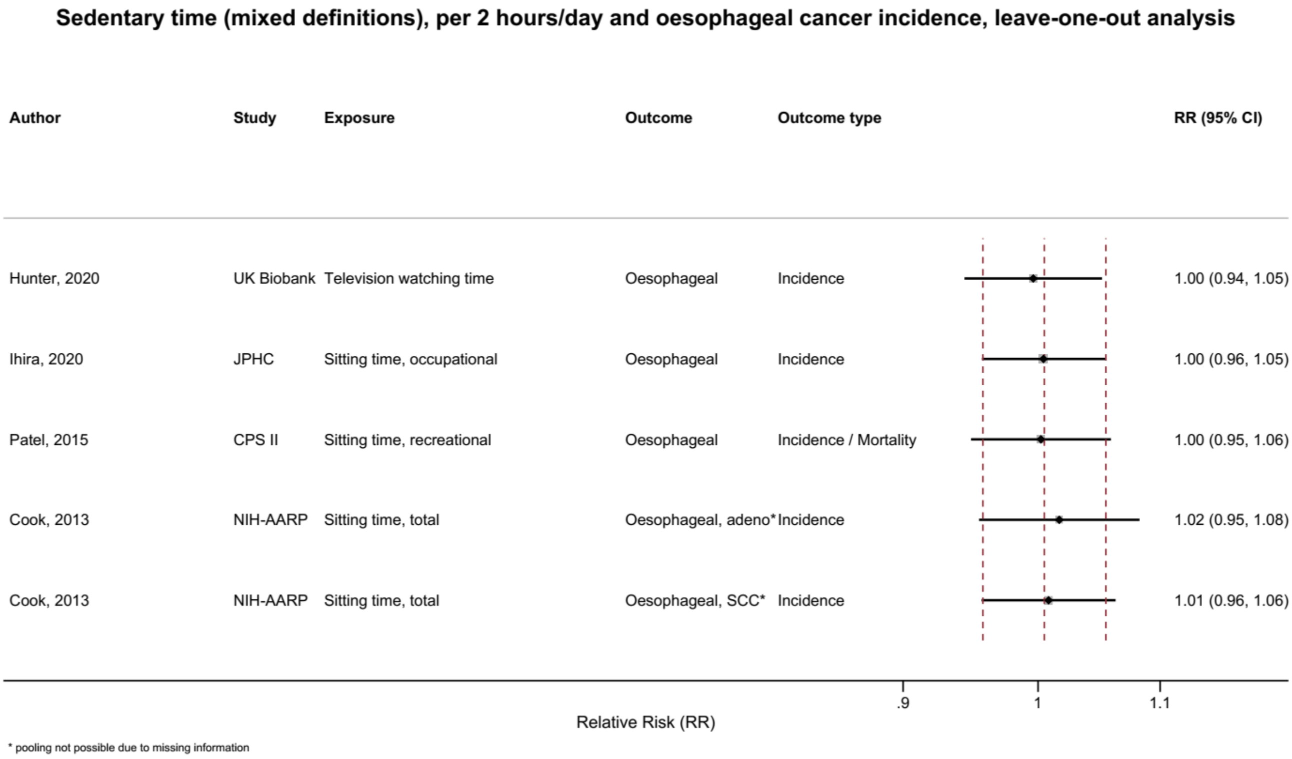

Figure legend: Forest plot shows the linear dose-response results (per 2 hours of sedentary time) from the inverse variance DerSimonian-Laird random-effects model after exclusion of each study from the meta-analysis. For each study, the square represents the summary relative risk (RR) estimate and the horizontal line across the square the corresponding 95% confidence interval (CI) computed from a meta-analysis excluding that study. The dashed vertical lines represent the results of the main meta-analysis.

Study abbreviations: CPS II, Cancer Prevention Study II; JPHC, Japan Public Health Center-Based Prospective Study; NIH-AARP, National Institute of Health – American Association for Retired Persons Diet and Health Study

Hunter 2020 (32746843)[78]; Ihira 2020 (31925977)[77]; Patel 2015 (26126627)[56]; Cook 2013 (24367697)[52]

**Supplementary Figure 113: Linear dose-response meta-analysis of the association between mixed definitions of sedentary time and stomach cancer incidence, leave-one-out sensitivity analysis.**

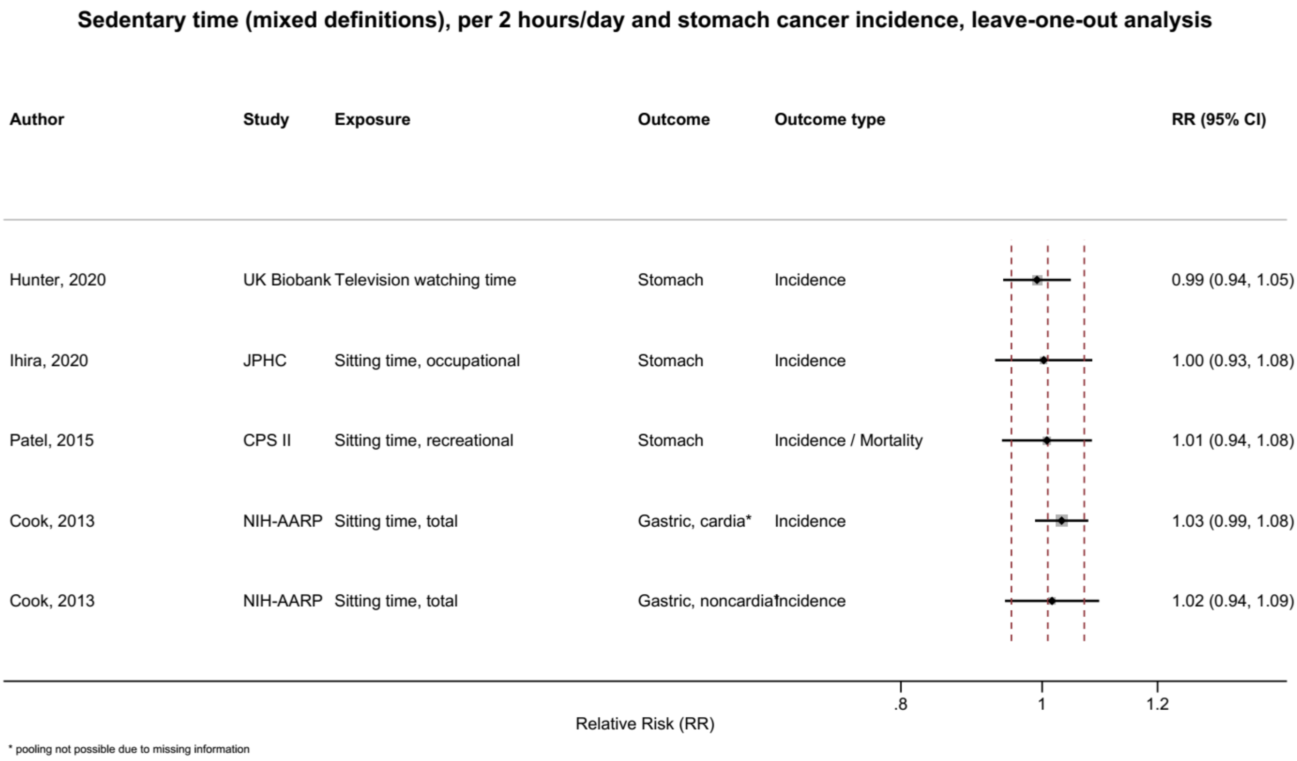

Figure legend: Forest plot shows the linear dose-response results (per 2 hours of sedentary time) from the inverse variance DerSimonian-Laird random-effects model after exclusion of each study from the meta-analysis. For each study, the square represents the summary relative risk (RR) estimate and the horizontal line across the square the corresponding 95% confidence interval (CI) computed from a meta-analysis excluding that study. The dashed vertical lines represent the results of the main meta-analysis.

**Supplementary Figure 114: Linear dose-response meta-analysis of the association between mixed definitions of sedentary time and colorectal cancer incidence, leave-one-out sensitivity analysis.**

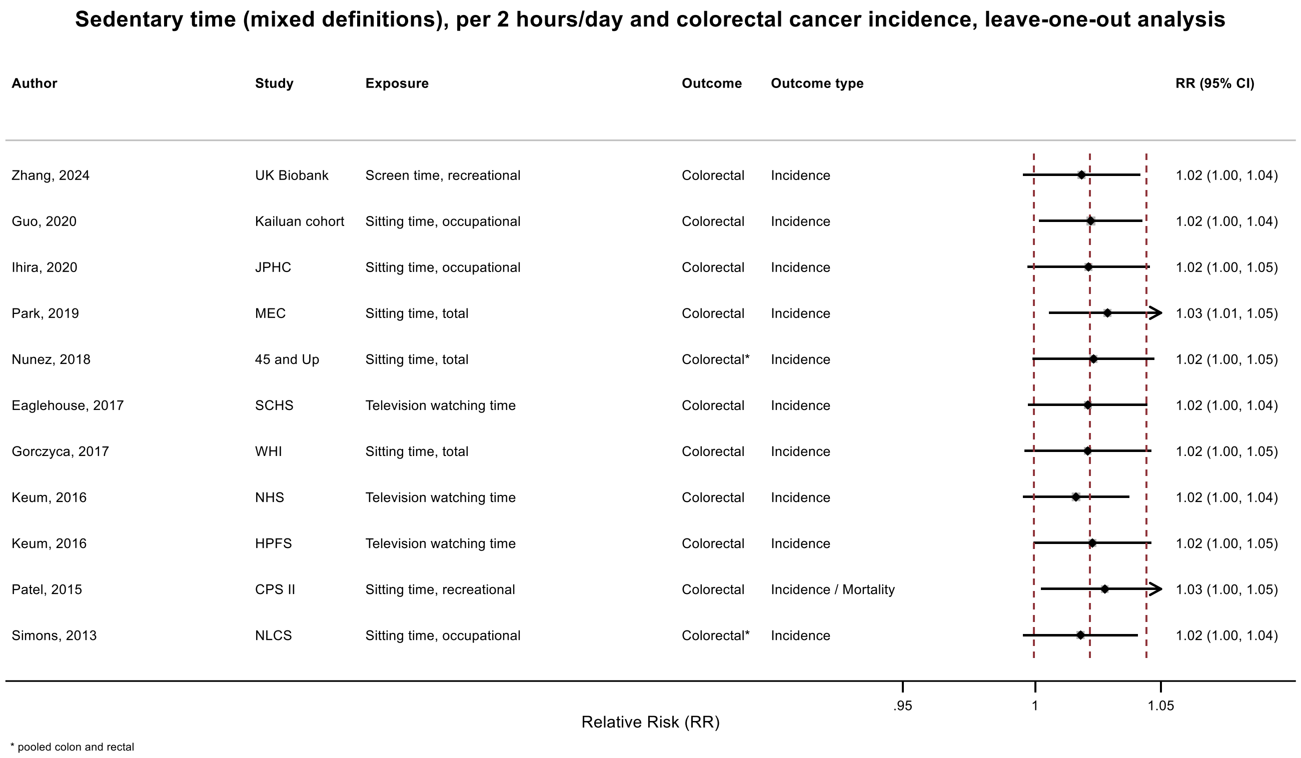

Figure legend: Forest plot shows the linear dose-response results (per 2 hours of sedentary time) from the inverse variance DerSimonian-Laird random-effects model after exclusion of each study from the meta-analysis. For each study, the square represents the summary relative risk (RR) estimate and the horizontal line across the square the corresponding 95% confidence interval (CI) computed from a meta-analysis excluding that study. The dashed vertical lines represent the results of the main meta-analysis.

Study abbreviations: 45 and Up, Sax Institute's 45 and Up Study; CPS II, Cancer Prevention Study II; HPFS, Health Professionals Follow-Up Study; JPHC, Japan Public Health Center-Based Prospective Study; Kailuan cohort, Kailuan Cohort; MEC, Multiethnic Cohort Study; NHS, Nurses' Health Study; NLCS, The Netherlands Cohort Study; SCHS, Singapore Chinese Health Study; WHI, Women's Health Initiative Study

Simons 2013 (23420352)[45]; Patel 2015 (26126627)[56]; Keum 2016 (26649988)[58]; Gorczyca 2017 (28538039)[61]; Eaglehouse 2017 (28542077)[62]; Nunez 2018 (29510753)[66]; Park 2019 (30910780)[73]; Ihira 2020 (31925977)[77]; Guo 2020 (31773920)[76]; Zhang 2024 (38974470)[93]

**Supplementary Figure 115: Linear dose-response meta-analysis of the association between mixed definitions of sedentary time and pancreatic cancer incidence, leave-one-out sensitivity analysis.**

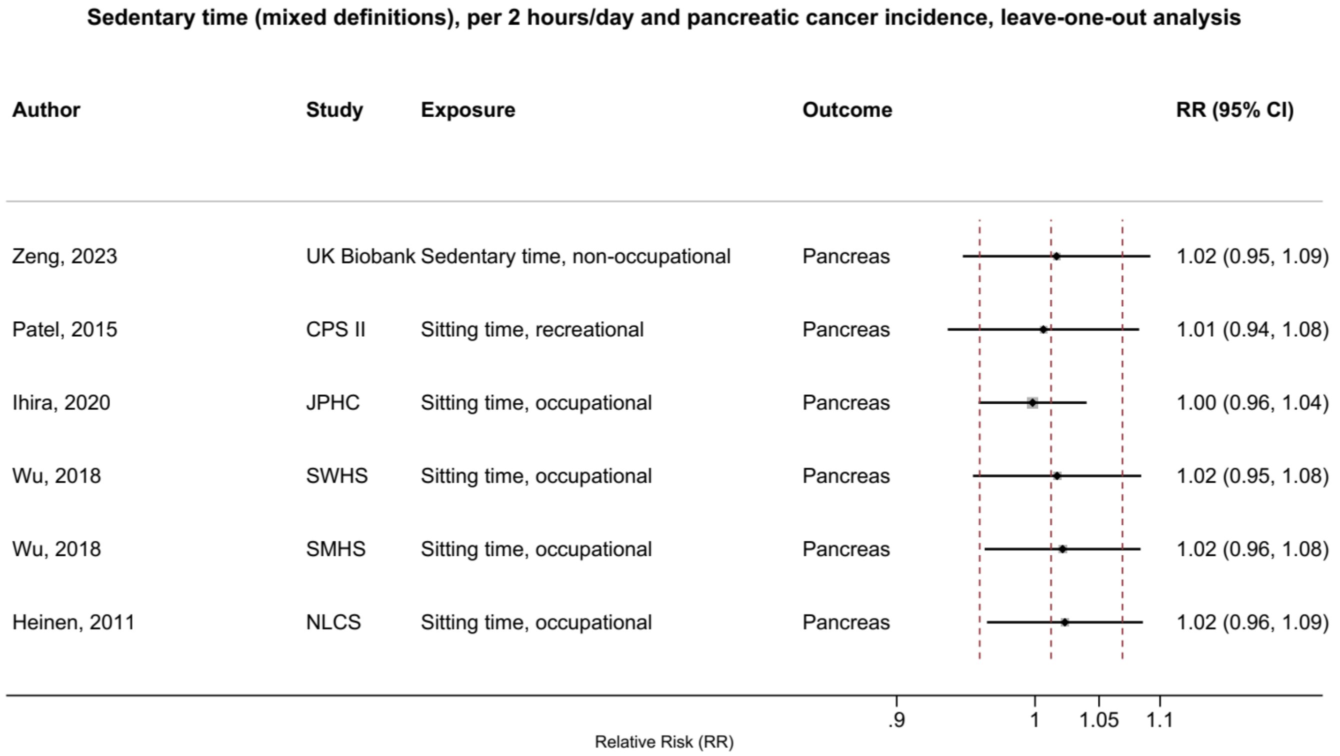

Figure legend: Forest plot shows the linear dose-response results (per 2 hours of sedentary time) from the inverse variance DerSimonian-Laird random-effects model after exclusion of each study from the meta-analysis. For each study, the square represents the summary relative risk (RR) estimate and the horizontal line across the square the corresponding 95% confidence interval (CI) computed from a meta-analysis excluding that study. The dashed vertical lines represent the results of the main meta-analysis.

Study abbreviations: CPS II, Cancer Prevention Study II; JPHC, Japan Public Health Center-Based Prospective Study; NLCS, The Netherlands Cohort Study; SMHS, Shanghai Men’s Health Study; SWHS, Shanghai Women’s Health Study; UK Biobank, UK Biobank

Heinen 2011 (21955648)[43]; Wu 2018 (29475964)[65]; Ihira 2020 (31925977)[77]; Patel 2015 (26126627)[56]; Zeng 2023 (38066552)[92]

**Supplementary Figure 116: Linear dose-response meta-analysis of the association between mixed definitions of sedentary time and lung cancer incidence, leave-one-out sensitivity analysis.**

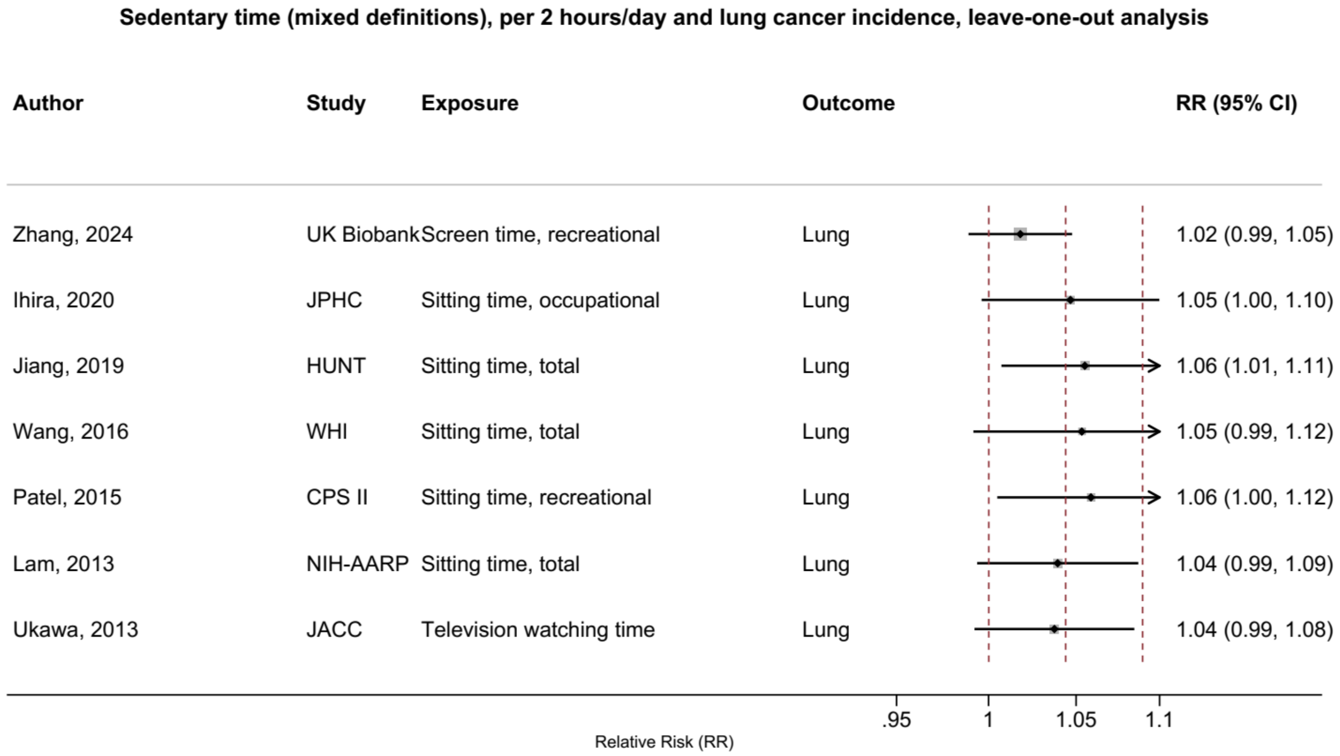

Figure legend: Forest plot shows the linear dose-response results (per 2 hours of sedentary time) from the inverse variance DerSimonian-Laird random-effects model after exclusion of each study from the meta-analysis. For each study, the square represents the summary relative risk (RR) estimate and the horizontal line across the square the corresponding 95% confidence interval (CI) computed from a meta-analysis excluding that study. The dashed vertical lines represent the results of the main meta-analysis.

Study abbreviations: CPS II, Cancer Prevention Study II; HUNT, The Trøndelag Health Study; JACC, Japan Collaborative Cohort study; JPHC, Japan Public Health Center-Based Prospective Study; NIH-AARP, National Institute of Health – American Association for Retired Persons Diet and Health Study; JACC, Japan Collaborative Cohort study; WHI, Women's Health Initiative Study

Ukawa 2013 (23686441)[47]; Lam 2013 (23940620)[48]; Patel 2015 (26126627)[56]; Wang 2016 (27439221)[59]; Jiang 2019 (30859092)[72]; Ihira 2020 (31925977)[77]; Zhang 2024 (38974470)[93];

**Supplementary Figure 117: Linear dose-response meta-analysis of the association between mixed definitions of sedentary time and breast cancer incidence, leave-one-out sensitivity analysis.**

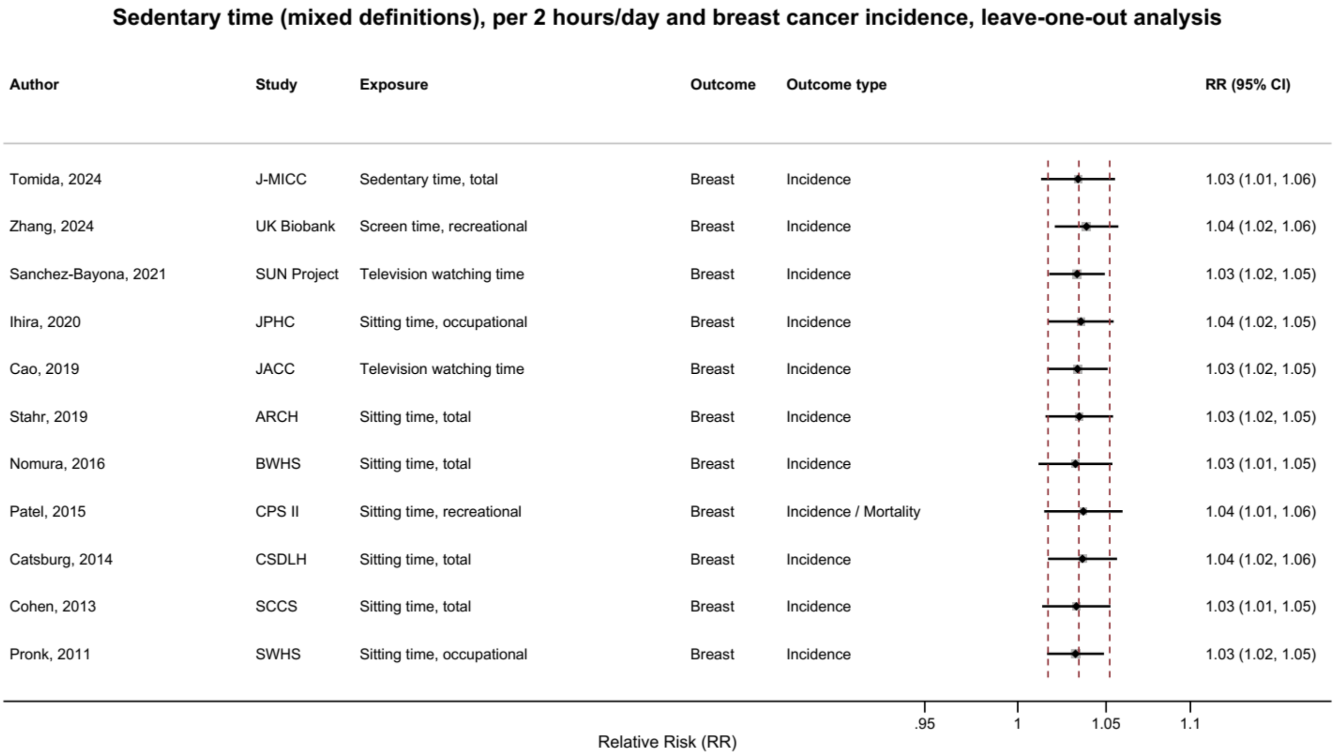

Figure legend: Forest plot shows the linear dose-response results (per 2 hours of sedentary time) from the inverse variance DerSimonian-Laird random-effects model after exclusion of each study from the meta-analysis. For each study, the square represents the summary relative risk (RR) estimate and the horizontal line across the square the corresponding 95% confidence interval (CI) computed from a meta-analysis excluding that study. The dashed vertical lines represent the results of the main meta-analysis.

Study abbreviations: ARCH, Arkansas Rural Community Health Study Cohort; BWHS, Black Women's Health Study; CPS II, Cancer Prevention Study II; CSDLH, Canadian Study of Diet, Lifestyle and Health; JACC, Japan Collaborative Cohort study; J-MICC, Japan Multi-Institutional Collaborative Cohort Study; JPHC, Japan Public Health Center-Based Prospective Study; SCCS, Southern Community Cohort Study; SWHS, Shanghai Women’s Health Study

Pronk 2011 (21934685)[42]; Cohen 2013 (23576427)[46]; Catsburg 2014 (24781974)[55]; Patel 2015 (26126627)[56]; Nomura 2016 (27632430)[60]; Cao 2019 (30913861)[74]; Stahr 2019 (35117114)[85]; Ihira 2020 (31925977)[77]; Sanchez-Bayona 2021 (33798533)[82]; Tomida 2024 (38041484)[91]; Zhang 2024 (38974470)[93]

**Supplementary Figure 118: Linear dose-response meta-analysis of the association between mixed definitions of sedentary time and endometrial cancer incidence, leave-one-out sensitivity analysis.**

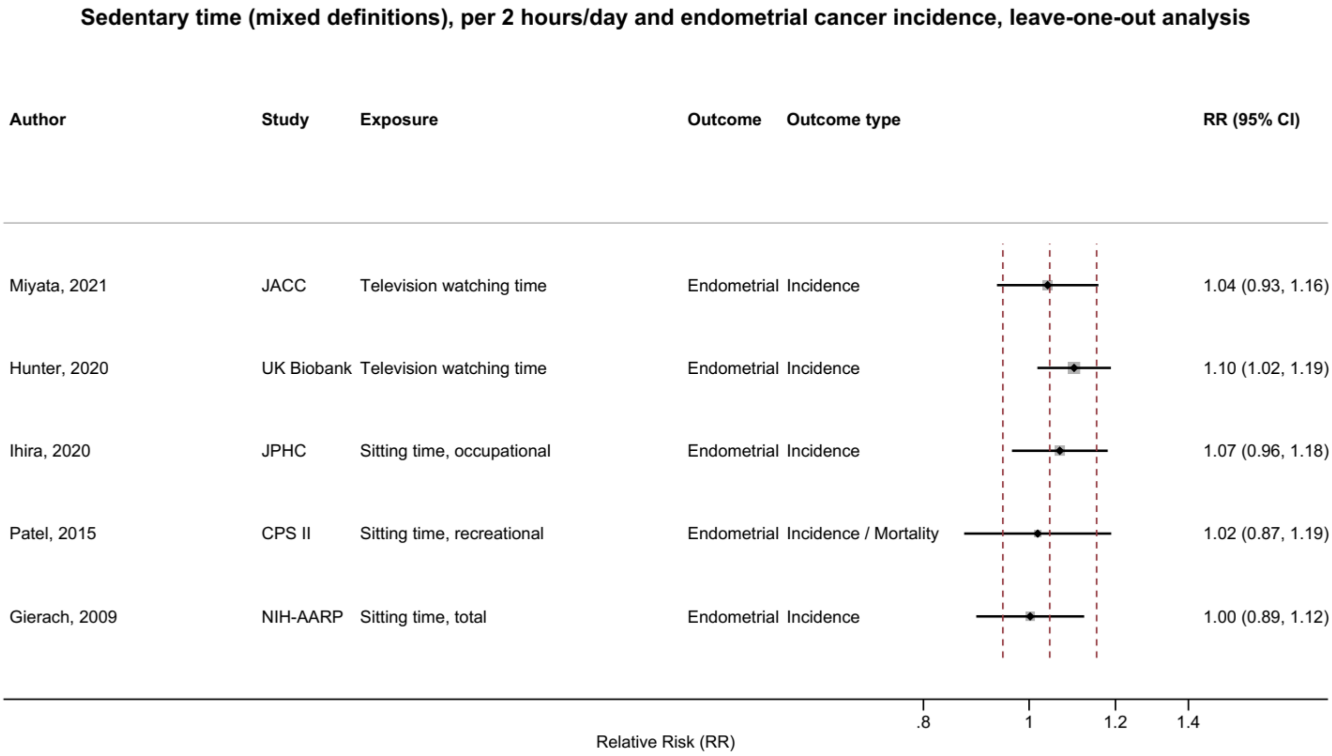

Figure legend: Forest plot shows the linear dose-response results (per 2 hours of sedentary time) from the inverse variance DerSimonian-Laird random-effects model after exclusion of each study from the meta-analysis. For each study, the square represents the summary relative risk (RR) estimate and the horizontal line across the square the corresponding 95% confidence interval (CI) computed from a meta-analysis excluding that study. The dashed vertical lines represent the results of the main meta-analysis.

Study abbreviations: CPS II, Cancer Prevention Study II; JACC, Japan Collaborative Cohort study; JPHC, Japan Public Health Center-Based Prospective Study; NIH-AARP, National Institute of Health – American Association for Retired Persons Diet and Health Study

Gierach 2009 (19123463)[38]; Patel 2015 (26126627)[56]; Ihira 2020 (31925977)[77]; Hunter 2020 (32746843)[78]; Miyata 2021 (32963209)[79]

### Supplementary Figure 119: Linear dose-response meta-analysis of the association between mixed definitions of sedentary time and ovarian cancer incidence, leave-one-out sensitivity analysis.

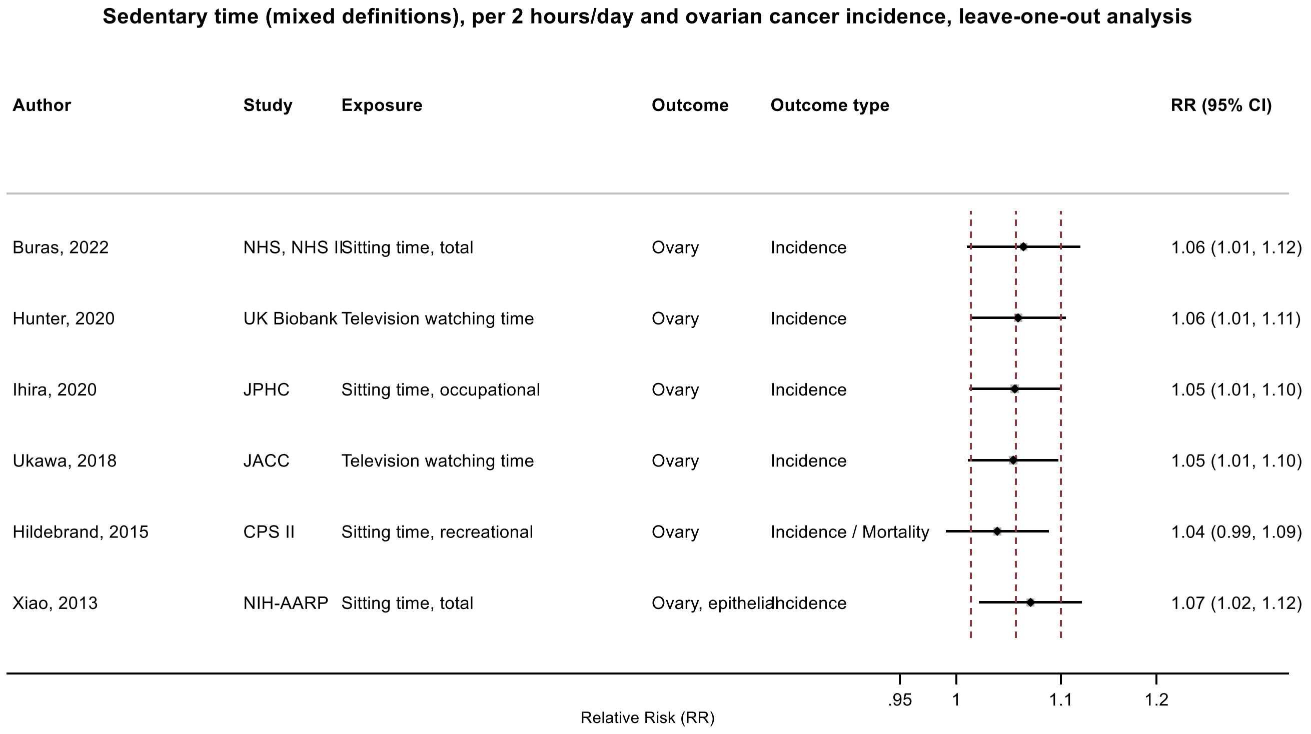

Figure legend: Forest plot shows the linear dose-response results (per 2 hours of sedentary time) from the inverse variance DerSimonian-Laird random-effects model after exclusion of each study from the meta-analysis. For each study, the square represents the summary relative risk (RR) estimate and the horizontal line across the square the corresponding 95% confidence interval (CI) computed from a meta-analysis excluding that study. The dashed vertical lines represent the results of the main meta-analysis.

Study abbreviations: CPS II, Cancer Prevention Study II; JACC, Japan Collaborative Cohort study;JPHC, Japan Public Health Center-Based Prospective Study; NHS, Nurses' Health Study; NHS II, Nurses' Health Study II; NIH-AARP, National Institute of Health – American Association for Retired Persons Diet and Health Study

Xiao 2013 (23966580)[49]; Hildebrand 2015 (26335264)[57]; Ukawa 2018 (29340890)[64]; Ihira 2020 (31925977)[77]; Buras 2022 (35094053)[84]; Hunter 2020 (32746843)[78]

**Supplementary Figure 120: Linear dose-response meta-analysis of the association between mixed definitions of sedentary time and prostate cancer incidence, leave-one-out sensitivity analysis.**

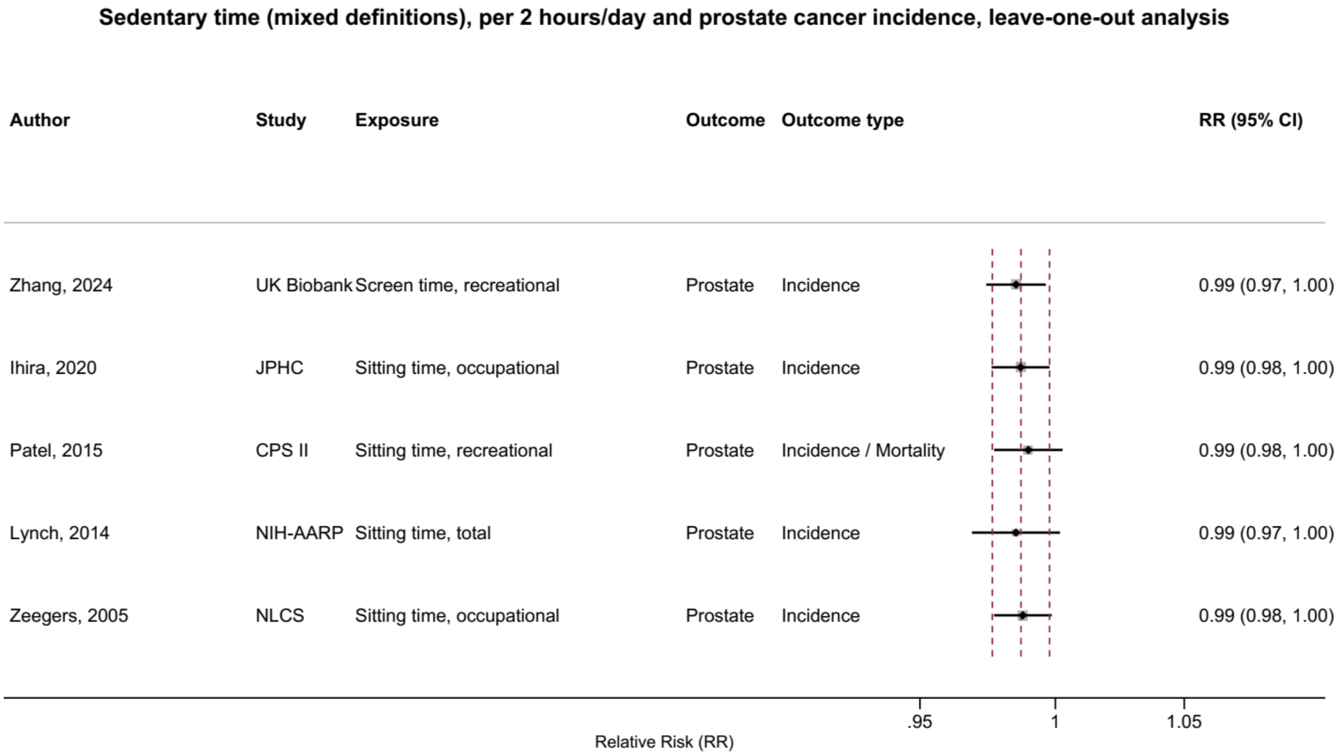

Figure legend: Forest plot shows the linear dose-response results (per 2 hours of sedentary time) from the inverse variance DerSimonian-Laird random-effects model after exclusion of each study from the meta-analysis. For each study, the square represents the summary relative risk (RR) estimate and the horizontal line across the square the corresponding 95% confidence interval (CI) computed from a meta-analysis excluding that study. The dashed vertical lines represent the results of the main meta-analysis.

Study abbreviations: CPS II, Cancer Prevention Study II; JPHC, Japan Public Health Center-Based Prospective Study; NIH-AARP, National Institute of Health – American Association for Retired Persons Diet and Health Study; NLCS, The Netherlands Cohort Study

Zeegers 2005 (15941961)[34]; Ihira 2020 (31925977)[77]; Patel 2015 (26126627)[56]; Lynch 2014 (24526287)[53]; Zhang 2024 (38974470)[93]

**Supplementary Figure 121: Linear dose-response meta-analysis of the association between mixed definitions of sedentary time and kidney cancer incidence, leave-one-out sensitivity analysis.**

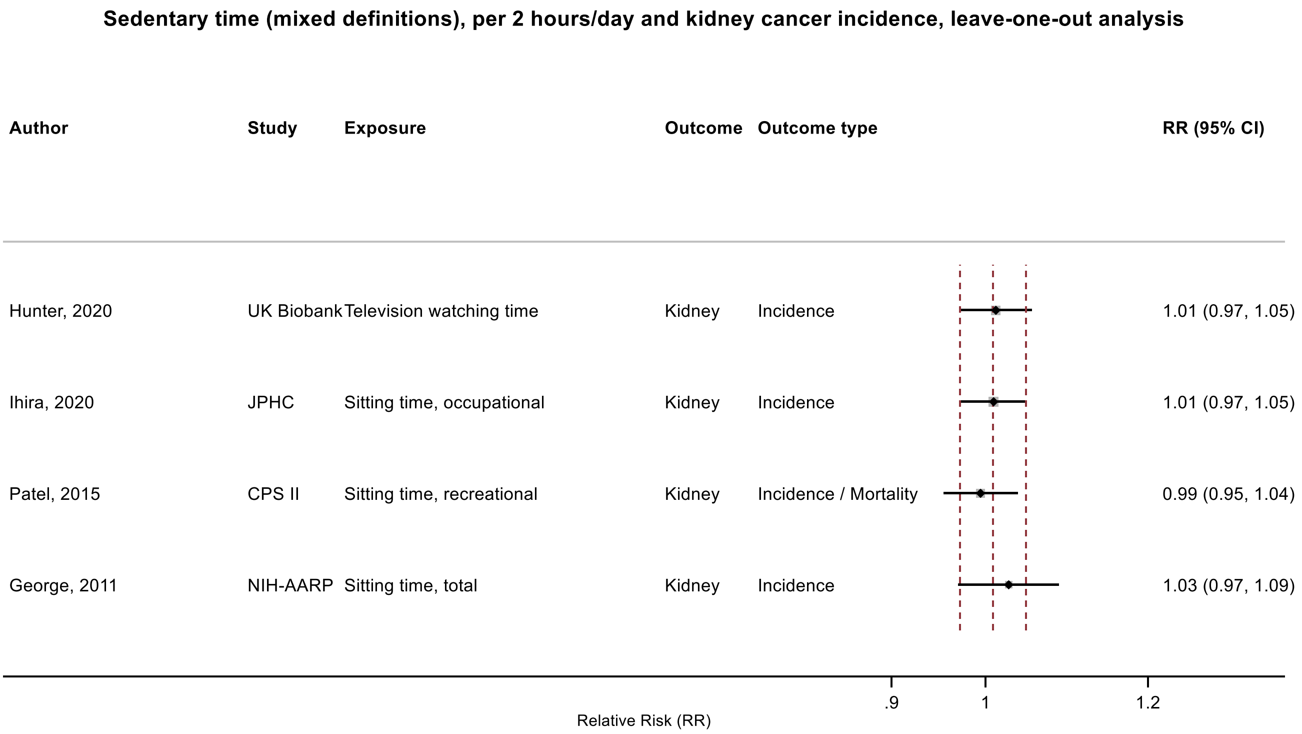

Figure legend: Forest plot shows the linear dose-response results (per 2 hours of sedentary time) from the inverse variance DerSimonian-Laird random-effects model after exclusion of each study from the meta-analysis. For each study, the square represents the summary relative risk (RR) estimate and the horizontal line across the square the corresponding 95% confidence interval (CI) computed from a meta-analysis excluding that study. The dashed vertical lines represent the results of the main meta-analysis.

Study abbreviations: CPS II, Cancer Prevention Study II; JPHC, Japan Public Health Center-Based Prospective Study; NIH-AARP, National Institute of Health – American Association for Retired Persons Diet and Health Study

George 2011 (21737302)[41]; Patel 2015 (26126627)[56]; Ihira 2020 (31925977)[77]

#### Risk of bias

##### Supplementary Figure 122: Risk of bias assessment of observational studies included in the meta-analyses on the association between sedentary behaviour and cancer risk

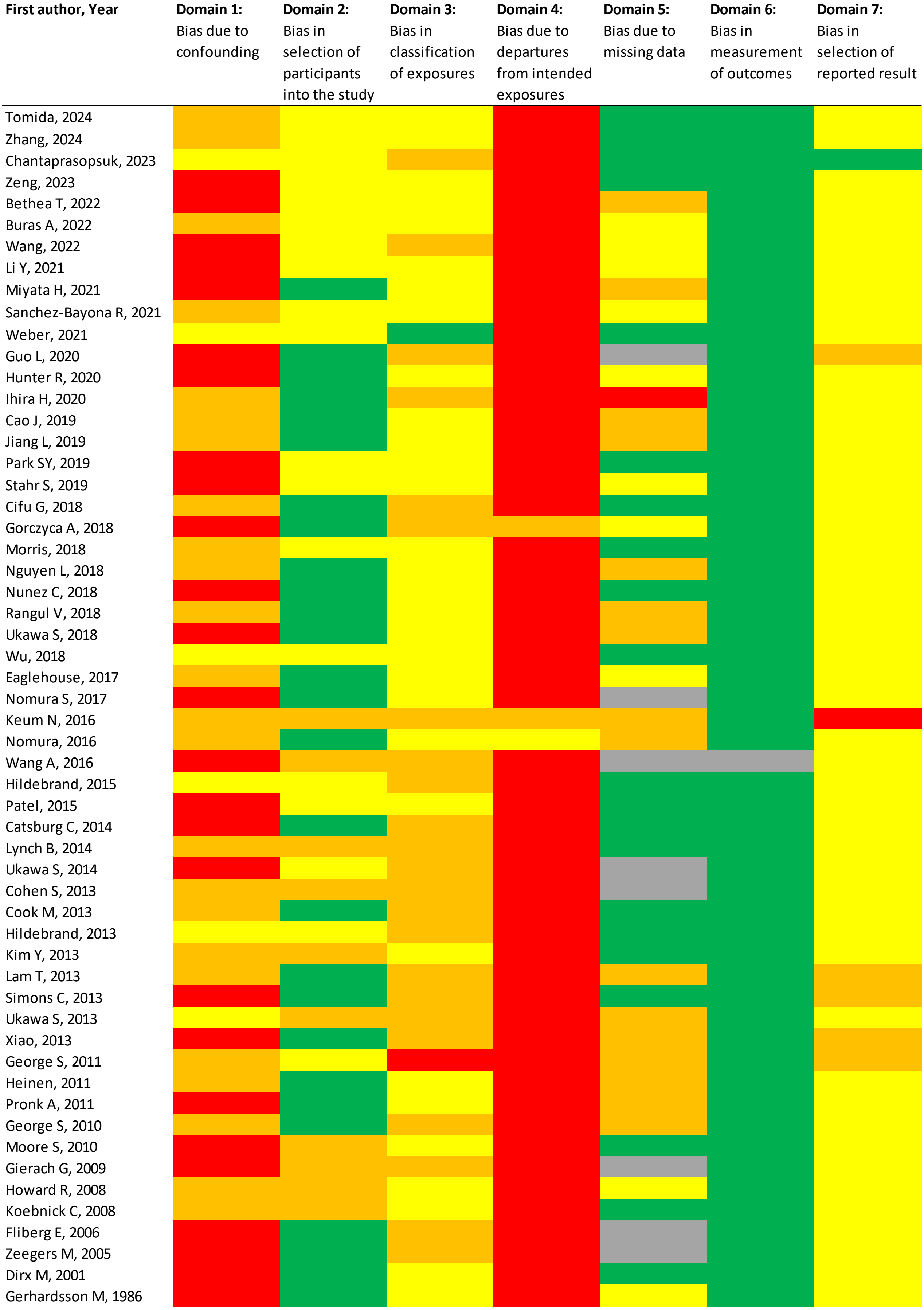

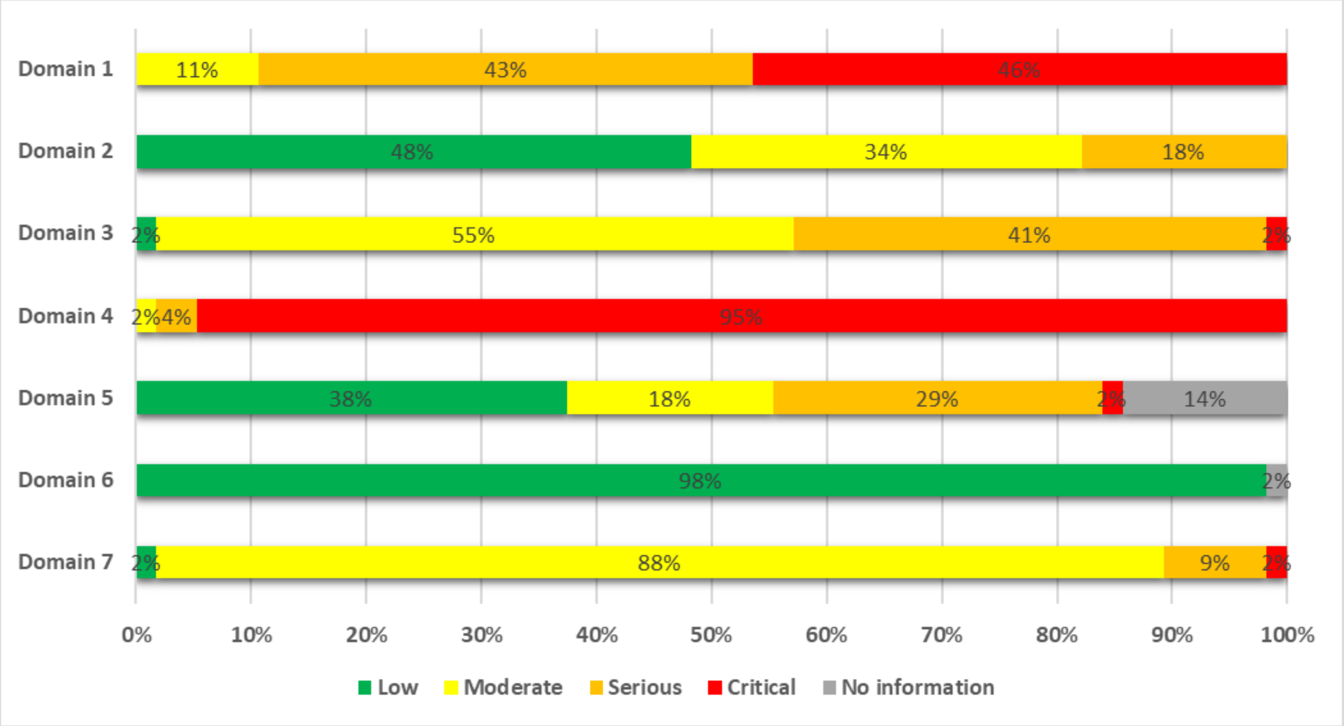

Tomida 2024 (38041484)[91], Zhang 2024 (38974470)[93]; Chantaprasopsuk 2023 (37202564)[89]; Zeng 2023 (38066552)[92]; Bethea 2022 (35325667)[87]; Buras 2022 (35094053)[84]; Wang 2022 (35159055)[86]; Li 2021 (33138348)[81]; Miyata 2021 (32963209)[79]; Sanchez-Bayona 2021 (33798533)[82]; Weber 2021 (34059803)[83]; Guo 2020 (31773920)[76]; Hunter 2020 (32746843)[78]; Ihira 2020 (31925977)[77]; Cao 2019 (30913861)[74]; Jiang 2019 (30859092)[72]; Park 2019 (30910780)[73]; Stahr 2019 (35117114)[85]; Cifu 2018 (30309689)[69]; Gorczyca 2018 (28538039)[61]; Morris 2018 (29520109)[67]; Nguyen 2018 (30740587)[71]; Nunez 2018 (29510753)[66]; Rangul 2018 (30352079)[70]; Ukawa 2018 (29340890)[64]; Wu 2018 (29475964)[65]; Eaglehouse 2017 (28542077)[62]; Nomura 2017 (28975422)[63]; Keum 2016 (26649988)[58]; Nomura 2016 (27632430)[60]; Wang 2016 (27439221)[59]; Hildebrand 2015 (26335264)[57]; Patel 2015 (26126627)[56]; Catsburg 2014 (24781974)[55]; Lynch 2014 (24526287)[53]; Ukawa 2014 (24728669)[54]; Cohen 2013 (23576427)[46]; Cook 2013 (24367697)[52]; Hildebrand 2013 (24097200)[51]; Kim 2013 (24062293)[50]; Lam 2013 (23940620)[48]; Simons 2013 (23420352)[45]; Ukawa 2013 (23686441)[47]; Xiao 2013 (23966580)[49]; George 2011 (21737302)[41]; Heinen 2011 (21955648)[43]; Pronk 2011 (21934685)[42]; George 2010 (20864719)[39]; Moore 2010 (20877336)[40]; Gierach 2009 (19123463)[38]; Howard 2008 (18437512)[36]; Koebnick 2008 (18483344)[37]; Friberg 2006 (17057024)[35]; Zeegers 2005 (15941961)[34]; Dirx 2001 (11745243)[33]; Gerhardsson 1986 (3962961)[94]
